# Evidence for Peroxisomal Dysfunction and Dysregulation of the CDP-Choline Pathway in Myalgic Encephalomyelitis/Chronic Fatigue Syndrome

**DOI:** 10.1101/2021.06.14.21258895

**Authors:** Xiaoyu Che, Christopher R. Brydges, Yuanzhi Yu, Adam Price, Shreyas Joshi, Ayan Roy, Bohyun Lee, Dinesh K. Barupal, Aaron Cheng, Dana March Palmer, Susan Levine, Daniel L. Peterson, Suzanne D. Vernon, Lucinda Bateman, Mady Hornig, Jose G. Montoya, Anthony L. Komaroff, Oliver Fiehn, W. Ian Lipkin

## Abstract

**Background:** Myalgic encephalomyelitis/chronic fatigue syndrome (ME/CFS) is a chronic and debilitating disease that is characterized by unexplained physical fatigue unrelieved by rest. Symptoms also include cognitive and sensory dysfunction, sleeping disturbances, orthostatic intolerance, and gastrointestinal problems. A syndrome clinically similar to ME/CFS has been reported following well-documented infections with the coronaviruses SARS-CoV and MERS-CoV. At least 10% of COVID-19 survivors develop post acute sequelae of SARS-CoV-2 infection (PASC). Although many individuals with PASC have evidence of structural organ damage, a subset have symptoms consistent with ME/CFS including fatigue, post exertional malaise, cognitive dysfunction, gastrointestinal disturbances, and postural orthostatic intolerance. These common features in ME/CFS and PASC suggest that insights into the pathogenesis of either may enrich our understanding of both syndromes, and could expedite the development of strategies for identifying those at risk and interventions that prevent or mitigate disease.

**Methods:** Using regression, Bayesian and enrichment analyses, we conducted targeted and untargeted metabolomic analysis of 888 metabolic analytes in plasma samples of 106 ME/CFS cases and 91 frequency-matched healthy controls.

**Results:** In ME/CFS cases, regression, Bayesian and enrichment analyses revealed evidence of peroxisomal dysfunction with decreased levels of plasmalogens. Other findings included decreased levels of several membrane lipids, including phosphatidylcholines and sphingomyelins, that may indicate dysregulation of the cytidine-5’-diphosphocholine pathway. Enrichment analyses revealed decreased levels of choline, ceramides and carnitines, and increased levels of long chain triglycerides (TG) and hydroxy-eicosapentaenoic acid. Elevated levels of dicarboxylic acids were consistent with abnormalities in the tricarboxylic acid cycle. Using machine learning algorithms with selected metabolites as predictors, we were able to differentiate female ME/CFS cases from female controls (highest AUC=0.794) and ME/CFS cases without self-reported irritable bowel syndrome (sr-IBS) from controls without sr-IBS (highest AUC=0.873).

**Conclusion:** Our findings are consistent with earlier ME/CFS work indicating compromised energy metabolism and redox imbalance, and highlight new abnormalities that may provide insights into the pathogenesis of ME/CFS.

**One Sentence Summary:** Plasma levels of plasmalogens are decreased in patients with myalgic encephalomyelitis/chronic fatigue syndrome suggesting peroxisome dysfunction.

## Background

Myalgic encephalomyelitis/chronic fatigue syndrome (ME/CFS) is a disease of unknown cause that is defined by impairment from fatigue lasting longer than six months, unrefreshing sleep, post-exertional malaise, and either cognitive dysfunction or orthostatic intolerance [1]. People with ME/CFS may also report gastrointestinal disturbances, influenza-like symptoms, and chronic pain [2]. It is estimated that ME/CFS affects between 0.4% to 2.5% of the global population, and 1.5 to 2.5 million people in the United States alone [1, 3]. There are no approved laboratory tests for ME/CFS. A diagnosis is based on medical history and the exclusion of other disorders that may result in chronic illness [4, 5].

Prior metabolomic studies of patients with ME/CFS have provided insights into the potential pathogenesis and course of the disease, demonstrating disturbances in energy, lipid, amino acid, and redox metabolism [6-16]. Metabolic dimensions of ME/CFS may be related to sex; women are disproportionately affected by ME/CFS [1, 17]. Naviaux et al. (2016) [15] found differences in metabolic pathway disturbances and altered metabolite levels when stratifying ME/CFS cases by sex. Others have also reported sex-specific differences in plasma biomarkers [14, 18, 19].

Comorbid gastrointestinal (GI) symptoms constitute a potential subtype in ME/CFS [10, 12, 14, 15, 18, 20-23]. Among those with ME/CFS, the presence or absence of self-reported irritable bowel syndrome (sr-IBS), in particular, has highlighted differences in the plasma proteome relating to immune dysregulation and altered levels of metabolites within the metabolome [14, 18]. In a fecal metagenomics study, Nagy-Szakal et al. (2017) identified eleven bacterial species delineating differences between ME/CFS patients with and without sr-IBS and found relations between bacterial taxa and symptoms relating to fatigue and pain [23].

In this study, we report targeted and untargeted analyses of 888 metabolic analytes comprising of primary metabolites, biogenic amines, complex lipids, and oxylipins in plasma of ME/CFS cases and controls. We identified altered metabolomic profiles between ME/CFS patients, controls, and subgroups within ME/CFS patients based on sex and sr-IBS.

## Methods

### Study population

Our starting population comprised 177 ME/CFS cases and 177 controls in ME/CFS clinics in Incline Village, NV; Miami FL; New York, NY; Salt Lake City, UT; and Palo Alto, CA. All ME/CFS cases met the 1994 CDC Fukuda [24] and Canadian consensus criteria for ME/CFS [25]. All ME/CFS cases completed standardized screening and assessment instruments including medical history and symptom rating scales as well as a physical examination. Controls were matched to cases on age, sex, race/ethnicity, geographic/clinical site, and date of sampling (±30 days). Based on screening criteria, we excluded 5 ME/CFS cases that met any exclusion criteria from the 1994 CDC Fukuda and/or Canadian consensus criteria for ME/CFS such as having chronic infections, rheumatic and chronic inflammatory diseases, neurological disorders, psychiatric conditions, or were taking any immunomodulatory medication. Controls underwent the same screening process as ME/CFS subjects and were excluded if they reported ME/CFS or other conditions deemed by the recruiting physician to be inconsistent with a healthy control population. Controls were also excluded if they had a history of substance abuse, psychiatric illness, antibiotics in the prior three months, immunomodulatory medications in the prior year, and clinically significant findings on physical exam or screening laboratory tests. One control was excluded after prescreening based on these criteria. Additionally, 21 participants were excluded prior to baseline due to withdrawal from the study (n=18), loss to follow-up (n=2), and enrollment capacity (n=1). The baseline questionnaire was completed by with 327 participants. During the study, an additional 63 participants were excluded for study protocol deviations (n=25), loss to follow-up (n=25), and withdrawal from the study (n=13), resulting in a total of 264 participants.

For the analysis reported here, a sub-cohort was established based on complete survey and biospecimen data (blood, saliva, and stool) at the first and last time points of the study and key demographic characteristics were frequency-matched to ensure that the nested cohort was similar to the full cohort. This sub-cohort consisted of 106 ME/CFS cases and 91 controls; the derivation of the sub-cohort is summarized in **Figure 1**. All participants provided informed written consent in accordance with protocols approved by the Institutional Review Board at Columbia University Irving Medical Center.

**Fig. 1.**
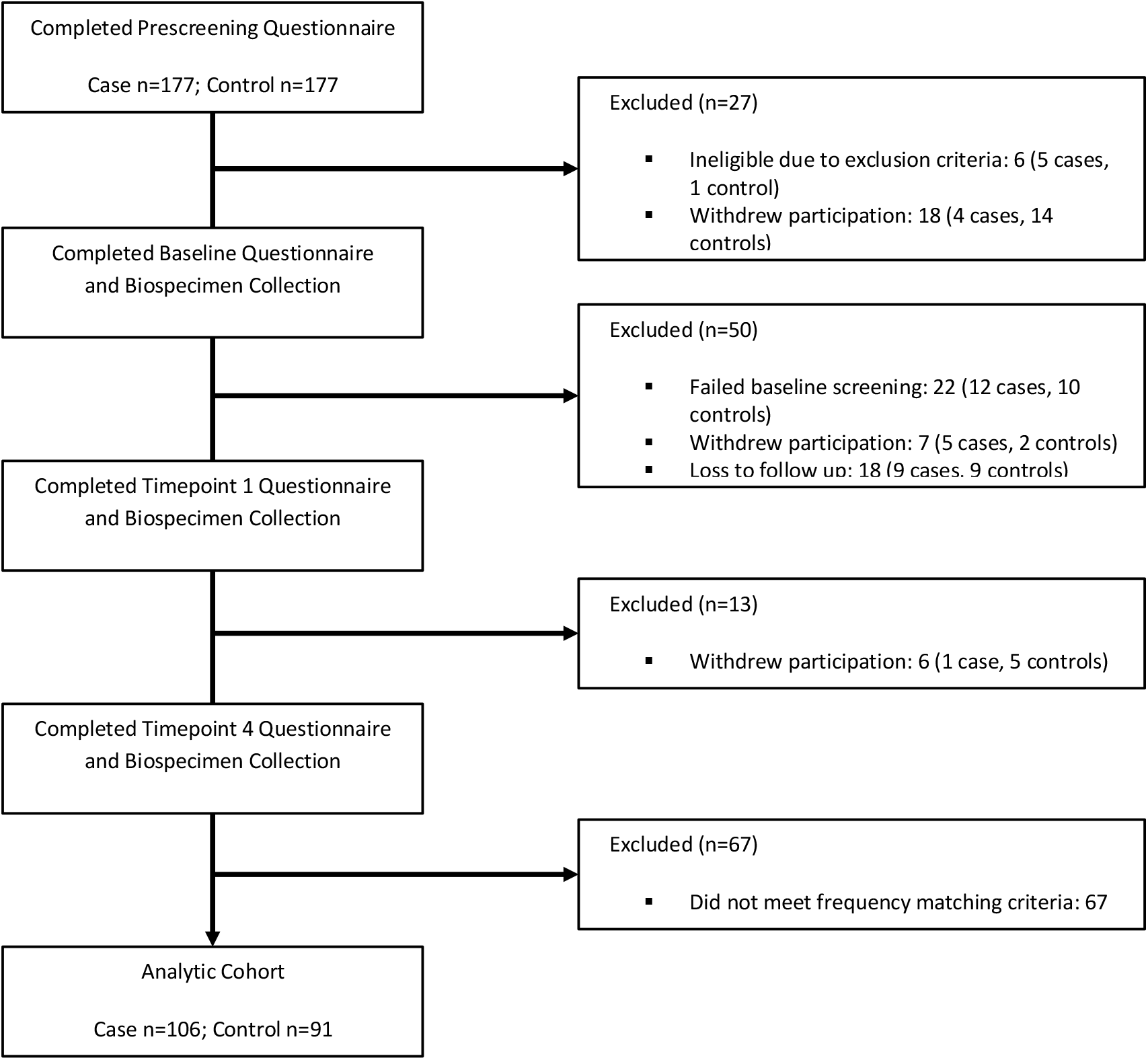
Pipeline for sample selection.

### Plasma collection

All participants fasted from midnight prior to the sample collection. Blood samples were collected into BD VacutainerTM Cell Preparation Tubes (CPT) with ethylenediaminetetraacetic acid (EDTA) anticoagulant between January 2016 and June 2016, and centrifuged to pellet red blood cells. The plasma was shipped to Columbia University at 4°C. After aliquoting, samples were stored at ™80°C until thawed for metabolomics analyses. All the samples were analyzed within two years of collection.

### Clinical assessment

Clinical symptoms and baseline health status were assessed on the day of physical examination and biological sample collection from both case and control subjects using the following instruments: the Short Form 36 Health Survey (SF-36), the Multidimensional Fatigue Inventory (MFI), DePaul Symptom Questionnaire (DSQ) [26], and Pittsburgh Sleep Quality Index (PSQI) [27]. The SF-36 includes the following subject-reported evaluations about current health status: physical and social functioning, physical and emotional limitations, vitality, pain, and general and mental health [28]. The MFI comprises of a 20-item self-reported questionnaire focused on general, physical and mental fatigue, reduced activity, and reduced motivation [29]. Cognitive function was tested based on the DSQ questionnaire data and was scored using a standard cognitive disturbance definition as well as a modified definition based on a subset of questionnaire variables. Sleeping disturbances linked to ME/CFS were tested and scored based on DSQ and PSQI questionnaire items. Each instrument was transformed into a 0–100 scale to facilitate combination and comparison wherein a score of 100 is equivalent to maximum disability or severity and a score of zero is equivalent to no disability or disturbance.

A diagnosis of sr-IBS was based on answers in the medical history form. Subjects were asked if they had received a previous IBS diagnosis by a physician and the date of that diagnosis. Of the 106 subjects with ME/CFS, 35 (33.0%) had sr-IBS. Of the 91 control subjects, 3 (3.3%) had sr-IBS.

### Metabolomics analysis

Samples were stored at -80°C before analysis. Untargeted metabolomics data were acquired using three chromatography/mass spectrometry-based assays (MS): (1) Primary metabolites such as mono- and disaccharides, hydroxyl- and amino acids were measured by gas chromatography/time-of-flight mass spectrometry (GC-TOF MS) [30] including data alignment and compound annotation using the BinBase database algorithm [31]. (2) Biogenic amines including microbial compounds such as trimethylamine N-oxide (TMAO), methylated and acetylated amino acids and short di- and tripeptides were measured by hydrophilic interaction liquid chromatography/quadrupole time-of-flight mass spectrometry (HILIC-QTOF MS). (3) Complex lipids including phosphoglycerolipids, triacylglycerides, sphingolipids, and free fatty acids were analyzed by liquid chromatography (LC)/quadrupole time-of-flight mass spectrometry (CSH-QTOF MS) [32]. Targeted bioactive oxylipin assay included thromboxanes, prostaglandins, and hdyroxy-, keto- and epoxy-lipins. All LC-MS/MS data included diverse sets of internal standards. LC-MS data were processed by MS-DIAL vs. 4.0 software [33], and the compounds were annotated based on accurate mass, retention time and MS/MS fragment matching using LipidBlast [34] and Massbank of North America libraries [35]. MS-FLO was used to remove erroneous peaks and reduce the false discovery rate in LC datasets [36]. A total of 821 known metabolites were annotated. Some complex lipids were annotated in both positive (ESI+) and negative (ESI-) ion modes, resulting in a total of 888 metabolic analytes that were included in our analysis. Data were normalized by SERRF [37]. Residual technical errors were assessed by coefficients of variation (CV) for known metabolites.

### Statistical analyses

For each metabolic analyte, zero values reflecting a measurement below the detection limit, were replaced with 50% of its smallest available value. In each of the four metabolomics panels, outliers were identified through principal component analysis (PCA). In primary metabolites (PM), 6 outliers (4 cases and 2 controls) were identified and removed; in complex lipids (CL), there were 5 outliers (3 cases and 2 controls); in oxylipins (OL), there was 1 outlier (1 case); in biogenic amines (BA), 4 outliers (3 cases and 1 control) were eliminated.

To compare the levels of each metabolite between ME/CFS cases and controls, we employed a variety of regression models with the metabolite level as the dependent variable and the binary case/control status as the independent variable, adjusting for all the matching variables (age, sex, race/ethnicity, geographic/clinical site, and season of sampling), body mass index (BMI) and sr-IBS. We considered two options for the dependent variable: 1) original metabolite levels, and 2) natural log-transformed metabolite levels. Before log-transformation, if necessary, all data points in metabolic analytes were multiplied by a minimal factor to keep the feature on a positive domain. Four regression models were considered: Gaussian regression with identity link, Gaussian regression with log link, lognormal regression and Gamma regression with log link. The Bayesian information criterion (BIC) was used to select the best fitting transformation/regression combination. We then calculated the estimated coefficient for the case/control status, together with its 95% confidence interval (95% CI) and p-value. Multiple comparisons over all metabolites were corrected using the Benjamini-Hochberg procedure [38] controlling the false discovery rate (FDR) at the 0.15 level. Additionally, chemical enrichment analyses were performed using ChemRICH [39] to determine chemical classes that were significantly altered between groups. ChemRICH does not rely upon background databases for statistical calculations and provides enrichment analysis based upon chemical structure, as opposed to defined pathways that can be inherently flawed [39].

For each metabolite, we also conducted Bayesian analysis with the best fitting transformation/regression combination using R packages “rstanarm” [40] and “bayestestR” [41]. Default (weakly informative) prior distributions from rstanarm were applied adjusting the scales of the priors internally. We then calculated the Bayes factors (BFs) and 95% highest density credible intervals (HDIs). The BF of a single parameter indicates the degree by which the mass of the posterior distribution has shifted further away from or closer to the null value (zero), relative to the prior distribution [42]. Hence, the BF measures the strength of evidence in favor of the alternative hypothesis (β≠0) over the null hypothesis (β=0). The 95% credible interval in the Bayesian framework is the range, within which the effect has 95% probability of falling, given the observed data. It has a different interpretation from the 95% confidence interval in the frequentist framework which instead signifies that with a large number of repeated samples, 95% of such calculated confidence intervals would include the true value of the parameter. We considered a metabolite significantly associated with ME/CFS if it satisfied the following criteria: 1) FDR adjusted p-value < 0.15, 2) BF > 3, and 3) 95% HDIs not covering 0. Jeffreys (1961) [43] suggested that the strength of evidence for the alternative hypothesis compared to the null hypothesis is regarded as noteworthy if BFs are above 3.

Naviaux et al. (2016) [15] showed that potential diagnostic metabolites for ME/CFS in targeted metabolomics are different between male and female subjects. Accordingly, we conducted sex-stratified analyses in addition to analyses with the whole cohort. In our previous work with a different cohort, sr-IBS comorbidity was identified as the strongest driving factor in the separation of topological networks based on fecal microbiome and plasma metabolic pathways [14, 23]. We subsequently found different patterns in the relationships between plasma proteomic profiling and ME/CFS when comparing ME/CFS with or without sr-IBS to healthy controls [18]. Given this precedent, we tested the hypothesis that sr-IBS subgroups in ME/CFS patients have altered metabolic profiles in a stratified analysis. As there were only 3 control subjects with sr-IBS, we focused on the comparison of ME/CFS subjects without sr-IBS versus controls without sr-IBS.

To explore the utility of the metabolomics assay as a biomarker tool for ME/CFS, we employed four machine learning algorithms: least absolute shrinkage and selection operator (Lasso) [44], adaptive Lasso (AdaLasso) [45], Random Forests (RF) [46] and XGBoost [47]. AdaLasso is different from Lasso in that AdaLasso has the oracle property that leads to consistent variable selection whereas Lasso is only consistent for variable selection under certain conditions on the shrinkage parameters and correlations [48]. However, neither outperforms the other consistently in predictions. For each of the algorithm, three sets of predictors were considered: 1) all metabolites, 2) metabolites with BF>1, and 3) metabolites with BF>3. The predictive models were first trained in the 80% randomly-selected training set using 10-fold cross-validation; the remaining 20% of the study population was used as the independent test set to validate model performance. We also applied the Bayesian Model Averaging (BMA) method [49] that combines the predictions of multiple models using weighted averages in which the weights are Bayesian posterior probabilities that the given model is the true model, conditional on the training data. The predictive performance of the 5 models (Lasso, AdaLasso, RF, XGBoost and Model Average) using the three sets of predictors in the test set was evaluated using Area under the Receiver Operating Characteristic curve (AUROC) values and Receiver Operating Characteristic (ROC) curves.

Data analyses were performed using MATLAB Statistics Toolbox R2013a (MathWorks, Inc., Natick, MA) and R version 3.6.3 (RStudio, Inc., Boston, MA). All p-values were 2-tailed.

## Results

### Study population characteristics

The study included plasma samples from 106 ME/CFS cases and 91 healthy controls recruited from five sites across the United States. Demographic and clinical characteristics of the study population are shown in **Table 1**. ME/CFS cases and controls were similar for all the frequency matching variables except season of collection (*Chi-squared p* = 0.004). We adjusted for all the matching variables, BMI and sr-IBS in our statistical analyses to account for confounding. All scales in SF-36 and MFI were significantly different between the two cohorts (*Wilcoxon rank-sum p* < 0.001). The study population is similar to the prescreened cohort that consisted of 177 ME/CFS cases and 177 controls in sex (*Chi-squared p*=0.60), race (*Chi-squared p*=0.66) and age (*Wilcoxon rank-sum p*=0.65).

**Table 1.**
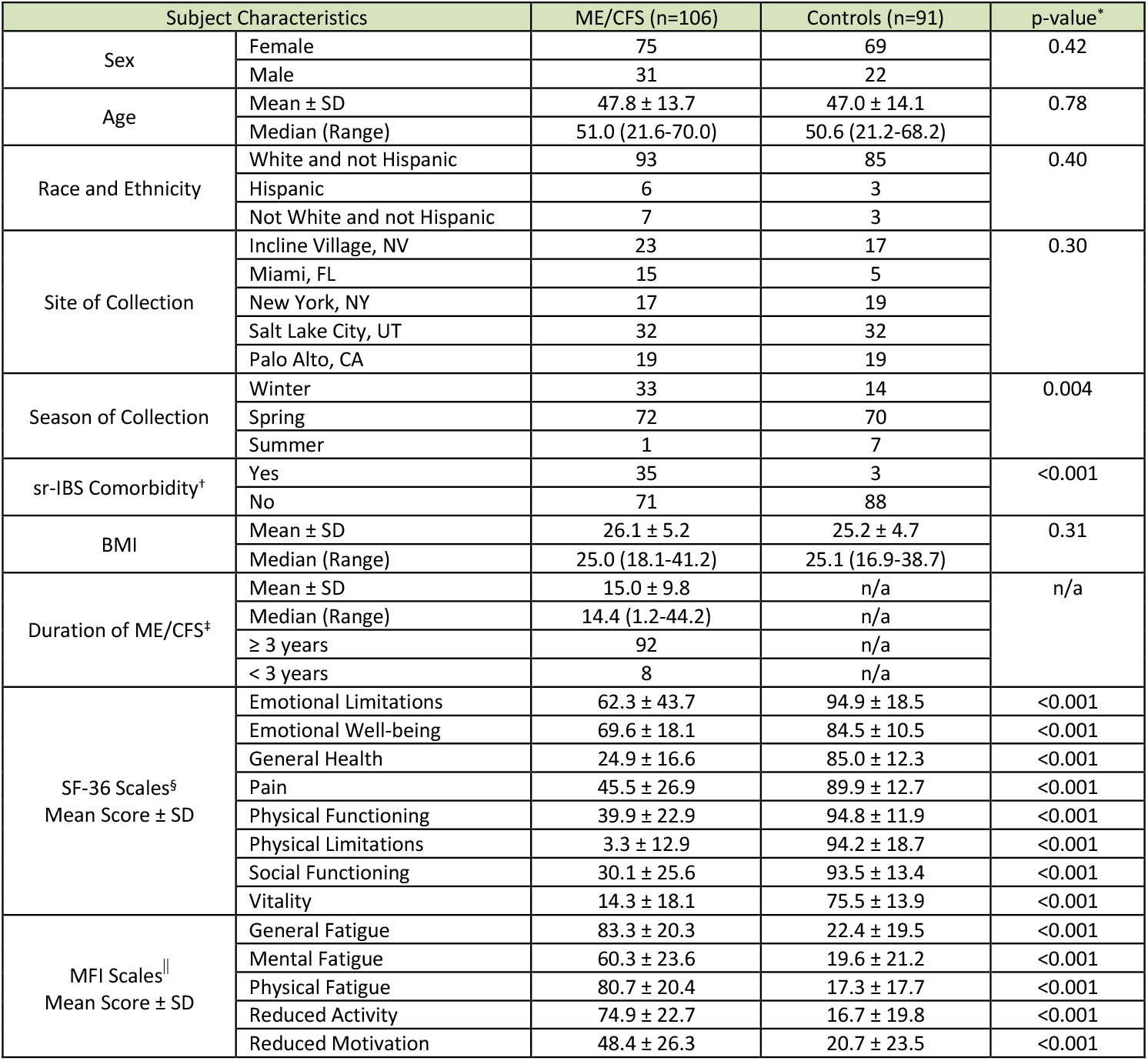
Subject characteristics. SD, standard deviation; ME/CFS, myalgic/encephalomyelitis/chronic fatigue syndrome. *For categorial variable, p-values were derived from Chi-squared tests; for continuous variables, p-values were derived from Wilcoxon rank-sum tests. †Prior physician diagnosed irritable bowel syndrome, self-reported on the questionnaire. ‡Only 90 responses were received for this item. §36-Item Short Form Health Survey; scored on a 0-100 scale with 0 = poor health status and 100 = excellent health status. ||Multidimensional Fatigue Inventory; scored on 0-100 scale with 0 = no fatigue and 100 = maximal fatigue.

| Subject Characteristics |  | ME/CFS (n=106) | Controls (n=91) | p-value* |
| --- | --- | --- | --- | --- |
| Sex | Female | 75 | 69 | 0.42 |
|  | Male | 31 | 22 |  |
| Age | Mean ± SD | 47.8 ± 13.7 | 47.0 ± 14.1 | 0.78 |
|  | Median (Range) | 51.0 (21.6-70.0) | 50.6 (21.2-68.2) |  |
| Race and Ethnicity | White and not Hispanic | 93 | 85 | 0.40 |
|  | Hispanic | 6 | 3 |  |
|  | Not White and not Hispanic | 7 | 3 |  |
| Site of Collection | Incline Village, NV | 23 | 17 | 0.30 |
|  | Miami, FL | 15 | 5 |  |
|  | New York, NY | 17 | 19 |  |
|  | Salt Lake City, UT | 32 | 32 |  |
|  | Palo Alto, CA | 19 | 19 |  |
| Season of Collection | Winter | 33 | 14 | 0.004 |
|  | Spring | 72 | 70 |  |
|  | Summer | 1 | 7 |  |
| sr-IBS Comorbidity† | Yes | 35 | 3 | <0.001 |
|  | No | 71 | 88 |  |
| BMI | Mean ± SD | 26.1 ± 5.2 | 25.2 ± 4.7 | 0.31 |
|  | Median (Range) | 25.0 (18.1-41.2) | 25.1 (16.9-38.7) |  |
| Duration of ME/CFS‡ | Mean ± SD | 15.0 ± 9.8 | n/a | n/a |
|  | Median (Range) | 14.4 (1.2-44.2) | n/a |  |
| | ≥ 3 years | 92 | n/a | |
|  | < 3 years | 8 | n/a |  |
| SF-36 Scales§<br>Mean Score ± SD | Emotional Limitations | 62.3 ± 43.7 | 94.9 ± 18.5 | <0.001 |
| | Emotional Well-being | 69.6 ± 18.1 | 84.5 ± 10.5 | <0.001 |
| | General Health | 24.9 ± 16.6 | 85.0 ± 12.3 | <0.001 |
| | Pain | 45.5 ± 26.9 | 89.9 ± 12.7 | <0.001 |
| | Physical Functioning | 39.9 ± 22.9 | 94.8 ± 11.9 | <0.001 |
| | Physical Limitations | 3.3 ± 12.9 | 94.2 ± 18.7 | <0.001 |
| | Social Functioning | 30.1 ± 25.6 | 93.5 ± 13.4 | <0.001 |
| | Vitality | 14.3 ± 18.1 | 75.5 ± 13.9 | <0.001 |
| MFI Scales\|\|<br>Mean Score ± SD | General Fatigue | 83.3 ± 20.3 | 22.4 ± 19.5 | <0.001 |
| | Mental Fatigue | 60.3 ± 23.6 | 19.6 ± 21.2 | <0.001 |
| | Physical Fatigue | 80.7 ± 20.4 | 17.3 ± 17.7 | <0.001 |
| | Reduced Activity | 74.9 ± 22.7 | 16.7 ± 19.8 | <0.001 |
| | Reduced Motivation | 48.4 ± 26.3 | 20.7 ± 23.5 | <0.001 |

### Metabolomic dataset

Targeted and untargeted mass spectrometry platforms yielded data for 888 metabolic analytes comprising 100 PM, 237 BA, 480 CL, and 71 OL. **Supplementary Table S1** shows the sample mean and the standard deviation (SD) of levels of each metabolite within all ME/CFS cases, all controls, female ME/CFS cases, female controls, male ME/CFS cases, male controls, ME/CFS cases without sr-IBS and controls without sr-IBS.

### ME/CFS associated with altered metabolomic profile

In PM, BA, and CL panels, lognormal regression models with log-transformed metabolite levels as dependent variables had the lowest BIC values and best fit the data; the estimated coefficients can be interpreted as the differences in the mean values of log-log transformation of metabolite levels between cases and controls. In OL panel, a mixture of lognormal and log-link Gamma regression models with original metabolite levels as dependent variables best fit the data. For lognormal regression models, the estimated coefficients are interpreted as the mean differences of log transformation of metabolite levels between two groups. For log-link Gamma regression models, the estimated coefficients are interpreted as the log of fold change between two groups.

We did not identify any metabolite as significantly associated with ME/CFS in the PM panel. In the BA panel, levels of acetaminophen were increased in ME/CFS cases compared to controls. In the CL panel, we found decreased levels of plasmalogens, unsaturated phospholipid ethers, unsaturated phosphatidylcholines (PC), an unsaturated sphingomyelin (SM), and an unsaturated lysophosphatidylcholines (LPC) in ME/CFS cases compared to controls. In the OL panel, decreased levels of Resolvin D1 were observed in ME/CFS cases compared to controls. **Table 2** shows the estimated coefficients in the regression models of these metabolites, their associated 95% CIs, p-values, FDR adjusted p-values and BFs. Because we used weakly informative priors in Bayesian analysis, the 95% HDIs were similar to the 95% CIs. We report estimations of HDIs in **Supplementary Table S2** where estimations for all metabolites are shown.

**Table 2.** Metabolites significantly associated with ME/CFS or ME/CFS subgroups. ME/CFS, myalgic encephalomyelitis/chronic fatigue syndrome; sr-IBS, self-reported irritable bowel syndrome; CI, confidence interval; FDR, false discovery rate adjusted p-value. For ME/CFS vs. controls, regression models were adjusted for age, sex, race/ethnicity, geographic/clinical site, season of sampling, body mass index, sr-IBS. In the sex-stratified comparisons, regression models were not adjusted for sex. In comparisons within subjects without sr-IBS, regression models were not adjusted for sr-IBS. For lognormal regression, estimated coefficients are interpreted as the differences in the mean values of log-log transformation of metabolite levels between cases and controls. For Gamma regression, estimated coefficients are interpreted as the log of fold change between two groups. Estimations in bold are significant in the corresponding comparisons. Criteria for significance: 1) FDR adjusted p-value from regression model < 0.15, 2) BayesFactor > 3, and 3) 95% highest density credible intervals not covering 0. The credible intervals were extremely similar to the confidence intervals and are shown in **Supplementary Table S2, S4**, and **S6**. No primary metabolites were found to be significantly associated with ME/CFS.

| Metabolite | Enrichment Cluster | Regression Model | ME/CFS vs. Control |  |  |  |  |
| --- | --- | --- | --- | --- | --- | --- | --- |
|  |  |  | Estimated Coefficient | 95% CI | p-value | FDR | BayesFactor |
| Biogenic Amines (BA) |  |  |  |  |  |  |  |
| Acetaminophen | drugs | Lognormal | 0.068 | (0.028, 0.108) | 0.001 | 0.103 | 3.035 |
| Alprazolam | drugs | Lognormal | 0.063 | (0.016, 0.109) | 0.009 | 0.178 | 0.963 |
| Acyclovir | drugs | Lognormal | 0.112 | (0.038, 0.185) | 0.003 | 0.139 | 2.674 |
| Complex Lipids (CL) |  |  |  |  |  |  |  |
| PE (p-36:2)/PE (o-36:3) - ESI(+) | plasmalogens | Lognormal | -0.028 | (-0.043, -0.013) | 0.000 | 0.074 | 20.935 |
| PE (p-34:2)/PE (o-34:3) | plasmalogens | Lognormal | -0.037 | (-0.060, -0.014) | 0.002 | 0.126 | 5.662 |
| SM (d40:3) | sphingomyelins | Lognormal | -0.025 | (-0.043, -0.007) | 0.007 | 0.173 | 1.222 |
| LPC (18:2) - ESI(-) | unsaturated lysophosphatidylcholines | Lognormal | -0.019 | (-0.032, -0.007) | 0.003 | 0.139 | 4.102 |
| PC (36:2) | unsaturated phosphatidylcholines | Lognormal | -0.007 | (-0.011, -0.003) | 0.000 | 0.074 | 11.241 |
| PC (36:4) A - ESI(+) | unsaturated phosphatidylcholines | Lognormal | -0.018 | (-0.028, -0.008) | 0.000 | 0.074 | 8.134 |
| PC (36:4) A - ESI(-) | unsaturated phosphatidylcholines | Lognormal | -0.019 | (-0.031, -0.008) | 0.001 | 0.103 | 4.032 |
| PC (32:2) - ESI(-) | unsaturated phosphatidylcholines | Lognormal | -0.027 | (-0.043, -0.010) | 0.002 | 0.135 | 7.389 |
| PC 34:4e | unsaturated phosphatidylcholines | Lognormal | -0.022 | (-0.036, -0.008) | 0.003 | 0.139 | 4.327 |
| PC (34:2) - ESI(+) | unsaturated phosphatidylcholines | Lognormal | -0.005 | (-0.008, -0.002) | 0.003 | 0.139 | 2.039 |
| PC (p-34:2)/PC (o-34:3) - ESI(+) | unsaturated phospholipid ethers | Lognormal | -0.018 | (-0.027, -0.009) | 0.000 | 0.062 | 44.620 |
| PC (p-34:1)/PC (o-34:2) | unsaturated phospholipid ethers | Lognormal | -0.021 | (-0.032, -0.010) | 0.000 | 0.062 | 178.678 |
| PC (p-36:1)/PC (o-36:2) | unsaturated phospholipid ethers | Lognormal | -0.055 | (-0.086, -0.024) | 0.001 | 0.074 | 11.555 |
| PC (p-34:2)/PC (o-34:3) - ESI(-) | unsaturated phospholipid ethers | Lognormal | -0.020 | (-0.032, -0.009) | 0.001 | 0.074 | 12.281 |
| PC (p-36:4)/PC (o-36:5) - ESI(-) | unsaturated phospholipid ethers | Lognormal | -0.021 | (-0.034, -0.009) | 0.001 | 0.103 | 7.046 |
| PC (p-34:1)/PC (o-34:2) A | unsaturated phospholipid ethers | Lognormal | -0.027 | (-0.044, -0.011) | 0.002 | 0.125 | 5.655 |
| Oxylipins (OL) |  |  |  |  |  |  |  |
| Resolvin D1 | OH-FA_22_6_1 | Gamma | -0.528 | (-0.846, -0.210) | 0.002 | 0.134 | 6.635 |
| Metabolite | Enrichment Cluster | Regression Model | Female ME/CFS vs. Female Control |  |  |  |  |
|  |  |  | Estimated Coefficient | 95% CI | p-value | FDR | BayesFactor |
| Biogenic Amines (BA) |  |  |  |  |  |  |  |
| Acetaminophen | drugs | Lognormal | 0.064 | (0.016, 0.113) | 0.010 | 0.211 | 1.172 |
| Alprazolam | drugs | Lognormal | 0.081 | (0.030, 0.132) | 0.002 | 0.121 | 3.486 |
| Acyclovir | drugs | Lognormal | 0.152 | (0.057, 0.247) | 0.002 | 0.121 | 3.179 |
| Complex Lipids (CL) |  |  |  |  |  |  |  |
| PE (p-36:2)/PE (o-36:3) - ESI(+) | plasmalogens | Lognormal | -0.033 | (-0.049, -0.017) | 0.000 | 0.048 | 24.602 |
| PE (p-34:2)/PE (o-34:3) | plasmalogens | Lognormal | -0.042 | (-0.066, -0.018) | 0.001 | 0.064 | 6.155 |
| SM (d40:3) | sphingomyelins | Lognormal | -0.035 | (-0.055, -0.014) | 0.001 | 0.064 | 6.392 |
| LPC (18:2) - ESI(-) | unsaturated lysophosphatidylcholines | Lognormal | -0.022 | (-0.036, -0.007) | 0.005 | 0.169 | 2.295 |
| PC (36:2) | unsaturated phosphatidylcholines | Lognormal | -0.009 | (-0.013, -0.004) | 0.000 | 0.054 | 14.972 |
| PC (36:4) A - ESI(+) | unsaturated phosphatidylcholines | Lognormal | -0.022 | (-0.033, -0.010) | 0.000 | 0.054 | 8.061 |
| PC (36:4) A - ESI(-) | unsaturated phosphatidylcholines | Lognormal | -0.025 | (-0.038, -0.011) | 0.000 | 0.054 | 11.432 |
| PC (32:2) - ESI(-) | unsaturated phosphatidylcholines | Lognormal | -0.023 | (-0.041, -0.006) | 0.011 | 0.218 | 0.909 |
| PC 34:4e | unsaturated phosphatidylcholines | Lognormal | -0.029 | (-0.045, -0.014) | 0.000 | 0.054 | 13.135 |
| PC (34:2) - ESI(+) | unsaturated phosphatidylcholines | Lognormal | -0.006 | (-0.010, -0.003) | 0.000 | 0.054 | 6.913 |
| PC (p-34:2)/PC (o-34:3) - ESI(+) | unsaturated phospholipid ethers | Lognormal | -0.022 | (-0.032, -0.011) | 0.000 | 0.048 | 36.107 |
| PC (p-34:1)/PC (o-34:2) | unsaturated phospholipid ethers | Lognormal | -0.025 | (-0.038, -0.012) | 0.000 | 0.054 | 26.013 |
| PC (p-36:1)/PC (o-36:2) | unsaturated phospholipid ethers | Lognormal | -0.067 | (-0.106, -0.029) | 0.001 | 0.064 | 5.458 |
| PC (p-34:2)/PC (o-34:3) - ESI(-) | unsaturated phospholipid ethers | Lognormal | -0.025 | (-0.038, -0.011) | 0.001 | 0.054 | 7.542 |
| PC (p-36:4)/PC (o-36:5) - ESI(-) | unsaturated phospholipid ethers | Lognormal | -0.025 | (-0.040, -0.011) | 0.001 | 0.064 | 7.044 |
| PC (p-34:1)/PC (o-34:2) A | unsaturated phospholipid ethers | Lognormal | -0.035 | (-0.055, -0.015) | 0.001 | 0.064 | 6.285 |
| Oxylipins (OL) |  |  |  |  |  |  |  |
| Resolvin D1 | OH-FA_22_6_1 | Gamma | -0.513 | (-0.871, -0.156) | 0.006 | 0.183 | 1.811 |
| Metabolite | Enrichment Cluster | Regression Model | Male ME/CFS vs. Male Control |  |  |  |  |
|  |  |  | Estimated Coefficient | 95% CI | p-value | FDR | BayesFactor |
| Biogenic Amines (BA) |  |  |  |  |  |  |  |
| Acetaminophen | drugs | Lognormal | 0.064 | (-0.020, 0.148) | 0.143 | 0.864 | 0.200 |
| Alprazolam | drugs | Lognormal | 0.017 | (-0.090, 0.123) | 0.761 | 0.956 | 0.068 |
| Acyclovir | drugs | Lognormal | 0.013 | (-0.093, 0.120) | 0.806 | 0.956 | 0.066 |
| Complex Lipids (CL) |  |  |  |  |  |  |  |
| PE (p-36:2)/PE (o-36:3) - ESI(+) | plasmalogens | Lognormal | -0.020 | (-0.059, 0.018) | 0.310 | 0.864 | 0.117 |
| PE (p-34:2)/PE (o-34:3) | plasmalogens | Lognormal | -0.032 | (-0.090, 0.027) | 0.291 | 0.864 | 0.119 |
| SM (d40:3) | sphingomyelins | Lognormal | -0.002 | (-0.040, 0.037) | 0.933 | 0.992 | 0.060 |
| LPC (18:2) - ESI(-) | unsaturated lysophosphatidylcholines | Lognormal | -0.022 | (-0.043, -0.001) | 0.048 | 0.864 | 0.426 |
| PC (36:2) | unsaturated phosphatidylcholines | Lognormal | -0.004 | (-0.011, 0.003) | 0.271 | 0.864 | 0.106 |
| PC (36:4) A - ESI(+) | unsaturated phosphatidylcholines | Lognormal | -0.015 | (-0.034, 0.004) | 0.128 | 0.864 | 0.174 |
| PC (36:4) A - ESI(-) | unsaturated phosphatidylcholines | Lognormal | -0.014 | (-0.035, 0.006) | 0.181 | 0.864 | 0.151 |
| PC (32:2) - ESI(-) | unsaturated phosphatidylcholines | Lognormal | -0.041 | (-0.084, 0.001) | 0.065 | 0.864 | 0.438 |
| PC 34:4e | unsaturated phosphatidylcholines | Lognormal | -0.007 | (-0.040, 0.025) | 0.666 | 0.928 | 0.073 |
| PC (34:2) - ESI(+) | unsaturated phosphatidylcholines | Lognormal | 0.000 | (-0.006, 0.006) | 0.900 | 0.976 | 0.061 |
| PC (p-34:2)/PC (o-34:3) - ESI(+) | unsaturated phospholipid ethers | Lognormal | -0.010 | (-0.031, 0.011) | 0.343 | 0.864 | 0.106 |
| PC (p-34:1)/PC (o-34:2) | unsaturated phospholipid ethers | Lognormal | -0.009 | (-0.028, 0.010) | 0.365 | 0.873 | 0.091 |
| PC (p-36:1)/PC (o-36:2) | unsaturated phospholipid ethers | Lognormal | -0.020 | (-0.061, 0.021) | 0.348 | 0.864 | 0.082 |
| PC (p-34:2)/PC (o-34:3) - ESI(-) | unsaturated phospholipid ethers | Lognormal | -0.016 | (-0.038, 0.005) | 0.144 | 0.864 | 0.169 |
| PC (p-36:4)/PC (o-36:5) - ESI(-) | unsaturated phospholipid ethers | Lognormal | -0.019 | (-0.046, 0.008) | 0.176 | 0.864 | 0.169 |
| PC (p-34:1)/PC (o-34:2) A | unsaturated phospholipid ethers | Lognormal | -0.013 | (-0.042, 0.015) | 0.363 | 0.873 | 0.107 |
| Oxylipins (OL) |  |  |  |  |  |  |  |
| Resolvin D1 | OH-FA_22_6_1 | Gamma | -0.282 | (-0.991, 0.426) | 0.439 | 0.881 | 0.099 |
| Metabolite | Enrichment Cluster |  | ME/CFS without sr-IBS vs. Control without sr-IBS |  |  |  |  |

|  |  | Regression Model | Estimated Coefficient | 95% CI | p-value | FDR | BayesFactor |
| --- | --- | --- | --- | --- | --- | --- | --- |
| Biogenic Amines (BA) |  |  |  |  |  |  |  |
| Acetaminophen | drugs | Lognormal | 0.066 | (0.023, 0.109) | 0.003 | 0.291 | 2.586 |
| Alprazolam | drugs | Lognormal | 0.068 | (0.018, 0.119) | 0.008 | 0.310 | 0.850 |
| Acyclovir | drugs | Lognormal | 0.104 | (0.031, 0.177) | 0.006 | 0.310 | 1.688 |
| Complex Lipids (CL) |  |  |  |  |  |  |  |
| PE (p-36:2)/PE (o-36:3) - ESI(+) | plasmalogens | Lognormal | -0.029 | (-0.045, -0.013) | 0.000 | 0.081 | 6.915 |
| PE (p-34:2)/PE (o-34:3) | plasmalogens | Lognormal | -0.036 | (-0.060, -0.012) | 0.004 | 0.310 | 2.620 |
| SM (d40:3) | sphingomyelins | Lognormal | -0.026 | (-0.046, -0.006) | 0.013 | 0.310 | 1.103 |
| LPC (18:2) - ESI(-) | unsaturated lysophosphatidylcholines | Lognormal | -0.017 | (-0.030, -0.003) | 0.016 | 0.310 | 0.720 |
| PC (36:2) | unsaturated phosphatidylcholines | Lognormal | -0.008 | (-0.012, -0.004) | 0.000 | 0.076 | 8.626 |
| PC (36:4) A - ESI(+) | unsaturated phosphatidylcholines | Lognormal | -0.017 | (-0.028, -0.007) | 0.002 | 0.218 | 3.025 |
| PC (36:4) A - ESI(-) | unsaturated phosphatidylcholines | Lognormal | -0.018 | (-0.030, -0.006) | 0.005 | 0.310 | 2.130 |
| PC (32:2) - ESI(-) | unsaturated phosphatidylcholines | Lognormal | -0.024 | (-0.040, -0.007) | 0.006 | 0.310 | 1.525 |
| PC 34:4e | unsaturated phosphatidylcholines | Lognormal | -0.023 | (-0.037, -0.008) | 0.003 | 0.248 | 3.559 |
| PC (34:2) - ESI(+) | unsaturated phosphatidylcholines | Lognormal | -0.004 | (-0.007, -0.001) | 0.021 | 0.327 | 0.553 |
| PC (p-34:2)/PC (o-34:3) - ESI(+) | unsaturated phospholipid ethers | Lognormal | -0.019 | (-0.028, -0.010) | 0.000 | 0.076 | 16.961 |
| PC (p-34:1)/PC (o-34:2) | unsaturated phospholipid ethers | Lognormal | -0.022 | (-0.033, -0.010) | 0.000 | 0.076 | 30.007 |
| PC (p-36:1)/PC (o-36:2) | unsaturated phospholipid ethers | Lognormal | -0.052 | (-0.084, -0.020) | 0.002 | 0.218 | 4.636 |
| PC (p-34:2)/PC (o-34:3) - ESI(-) | unsaturated phospholipid ethers | Lognormal | -0.019 | (-0.030, -0.007) | 0.002 | 0.248 | 3.502 |
| PC (p-36:4)/PC (o-36:5) - ESI(-) | unsaturated phospholipid ethers | Lognormal | -0.020 | (-0.032, -0.008) | 0.001 | 0.218 | 6.200 |
| PC (p-34:1)/PC (o-34:2) A | unsaturated phospholipid ethers | Lognormal | -0.025 | (-0.042, -0.008) | 0.005 | 0.310 | 1.458 |
| Oxylipins (OL) |  |  |  |  |  |  |  |
| Resolvin D1 | OH-FA_22_6_1 | Gamma | -0.379 | (-0.683, -0.073) | 0.016 | 0.310 | 0.597 |

Set enrichment analysis of the results from the regression models (**Figure 2A**) revealed that ME/CFS subjects had reduced levels of plasmalogens, sphingomyelins, unsaturated phospholipid ethers, unsaturated ceramides, carnitines, saturated lysophospholipids, unsaturated lysophosphoethanolamines, unsaturated lysophosphatidylcholines, saturated TG and prostaglandins. The majority of unsaturated phosphatidylcholines were also down-regulated in ME/CFS cases. Increased levels of hydroxy-eicosapentaenoic acid (HEPE), dicarboxylic acids, and the majority of unsaturated long chain TG were found in ME/CFS cases compared to controls. There were mixed directional alterations in the food exposome and epoxy fatty acids (EpODE). Complete data from ChemRICH enrichment analysis are provided in **Supplementary Table S3**. Data from compound-level enrichment analysis for the significantly altered metabolic clusters are illustrated in **Supplementary Table S4**. Levels of choline in food exposome were reduced in ME/CFS (estimated coefficient β=-0.009, p-value=0.004); levels of succinic acid (β=0.022, p-value=0.007) and alpha-ketoglutarate (β=0.016, p-value=0.048) in dicarboxylic acids were elevated in ME/CFS.

**Fig. 2.**
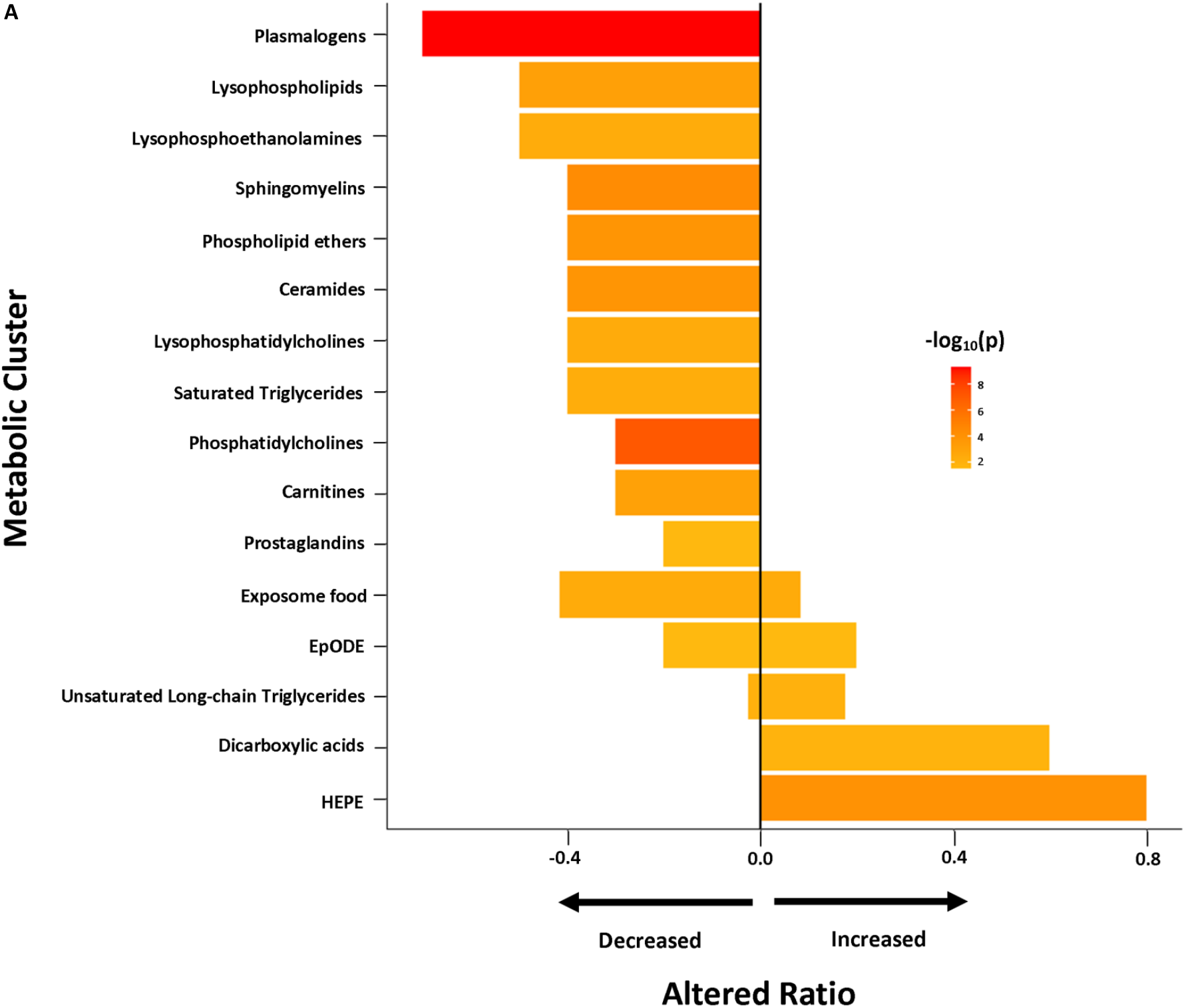
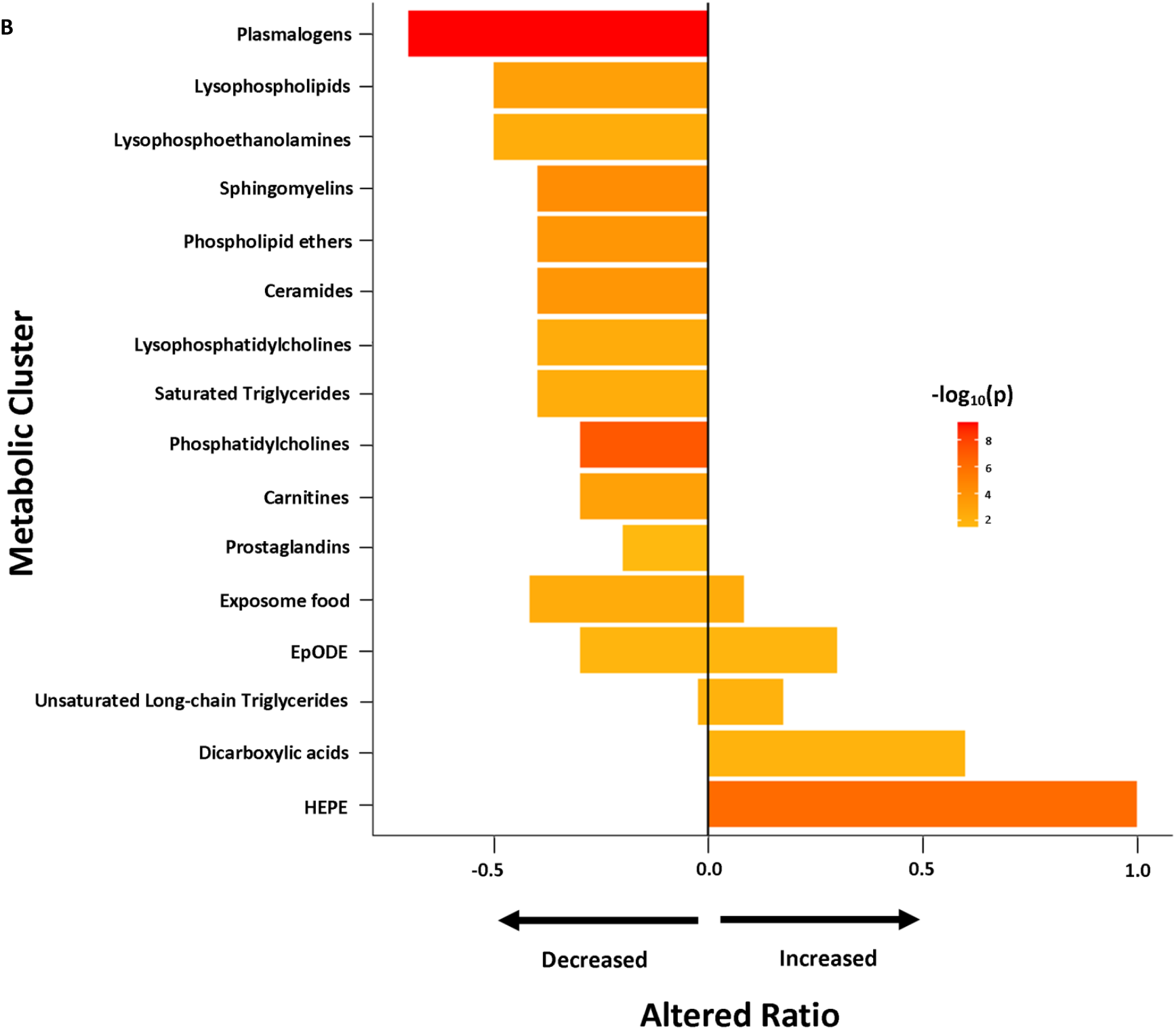
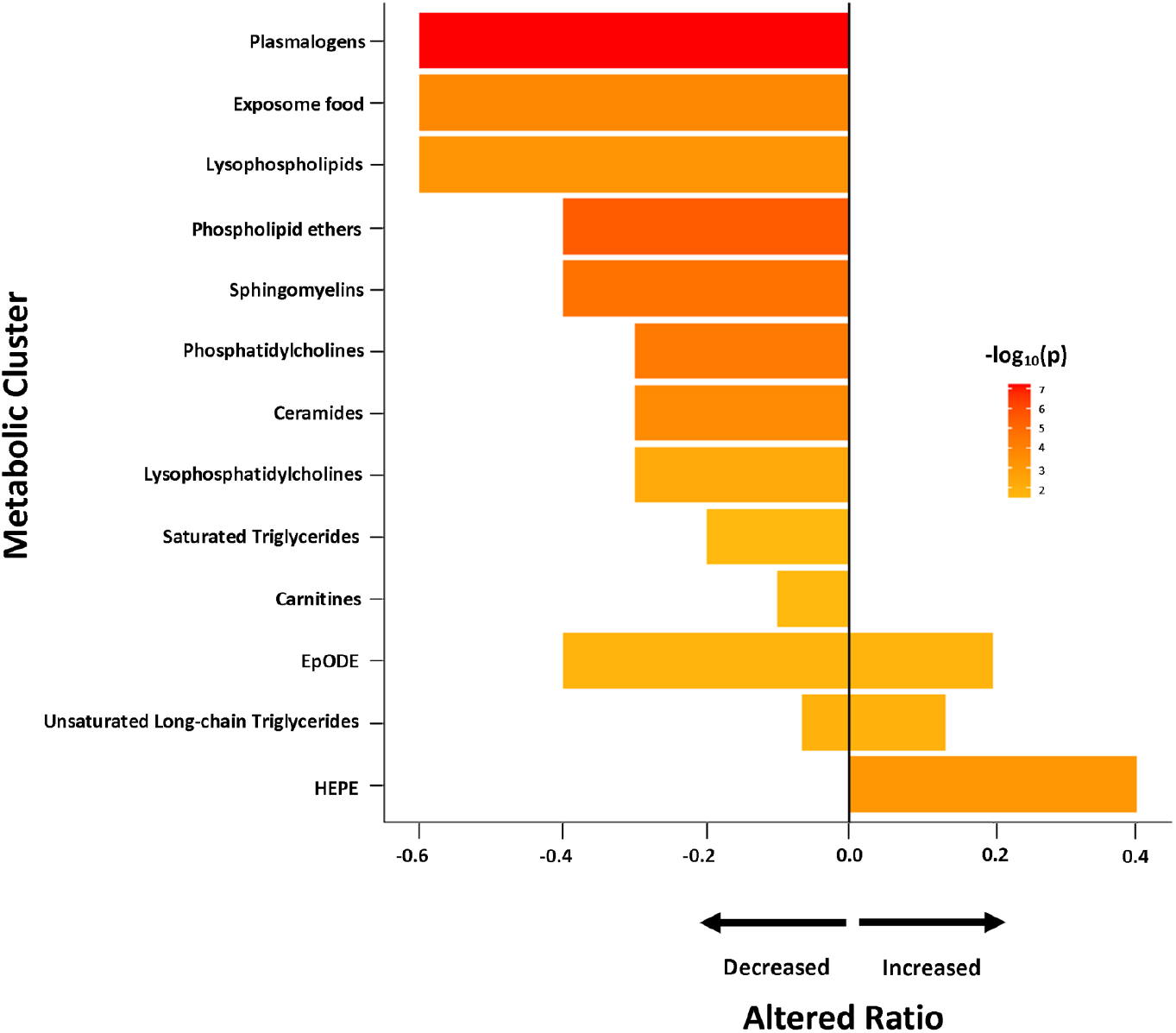
Chemical enrichment analyses using ChemRICH. HEPE, hydroxy eicosapentaenoic acid; ME/CFS, myalgic encephalomyelitis/chronic fatigue syndrome; sr-IBS, self-reported physician diagnosed irritable bowel syndrome. The length of the bar represents altered ratio for each metabolic cluster. A bar restricted to the left of the centered vertical line indicates a metabolic cluster that is lower in ME/CFS patients. A bar restricted to the right of the centered vertical line indicates a metabolic cluster that is higher in ME/CFS patients. A bar that crosses the vertical line indicates a metabolic cluster that is dysregulated in mixed directions. (**A**) All ME/CFS vs. controls. (**B**) Female ME/CFS vs. female controls. (**C**) ME/CFS without sr-IBS vs. controls without sr-IBS.

### Altered metabolomic profiles in female and male ME/CFS patients

Naviaux et al. (2016) [15] reported that women with ME/CFS, but not men, had disturbed fatty acid and endocannabinoid metabolism. Accordingly, we repeated separately the analyses in female and male cohorts in our study population.

In female subjects, regression and Bayesian analyses (**Table 2**) revealed that levels of PC, phosphatidylethanolamines (PE) and SM in the CL panel were decreased in ME/CFS patients compared to controls. In the BA panel, levels of two drug metabolites, alprazolam and acyclovir, were up-regulated in ME/CFS patients. We did not find the elevated levels of acetaminophen in female subjects that were observed in the entire ME/CFS (male and female population), presumably due to loss of power. Enrichment analysis in female subjects (**Figure 2B**) identified dysregulations in the same metabolic clusters as in the overall population. Complete data from enrichment analysis in female subjects are shown in **Supplementary Table S5**. In contrast, we did not find any metabolites significantly associated with risk of ME/CFS in male subjects. This may be due to limited sample size. **Supplementary Table S6** shows the regression and Bayesian estimations for all metabolites in male and female cohorts.

### Altered metabolomics profile in ME/CFS patients without sr-IBS

Due to the limited sample size of subjects with sr-IBS (35 ME/CFS cases and 3 controls), we only compared levels of metabolites between ME/CFS cases without sr-IBS and controls without sr-IBS. Levels of PC and PE were decreased in ME/CFS patients in this subgroup (**Table 2**). In the ChemRICH enrichment analysis, the dysregulations in metabolite clusters found to be dysregulated in the subgroup without sr-IBS (**Figure 2C**) were all identified in the overall population (**Figure 2A)**. Complete data pertaining to the regression, Bayesian and enrichment analyses are shown in **Supplementary Tables S7** and **S8**.

### Machine learning analyses

We considered three sets of metabolites as predictors to distinguish ME/CFS cases from controls, including all metabolites, metabolites with BF>1 and metabolites with BF>3. Each set of predictors was fitted in five different machine learning classifiers: Lasso, AdaLasso, RF, XGBoost, and BMA. The classifiers were first trained in the 80% randomly-selected training set and then validated in the remaining 20% test set. **Figure 3A-C** show the ROC curves and the AUROC values differentiating all ME/CFS cases from all controls, female ME/CFS from female controls, and ME/CFS without sr-IBS from controls without sr-IBS, respectively, in the test set. Although classifiers did not differentiate all ME/CFS from all controls, Lasso with BF>1 metabolites as predictors distinguished female ME/CFS patients from female controls with an AUROC value of 0.794 (95% CI: 0.612-0.976) and Lasso with BF>3 metabolites distinguished ME/CFS without sr-IBS from controls without sr-IBS with an AUROC value of 0.873 (95% CI: 0.747-0.999). The AUROC values and their associated 95% CIs of all the classifiers are shown in **Supplementary Table S9**.

**Fig. 3.**
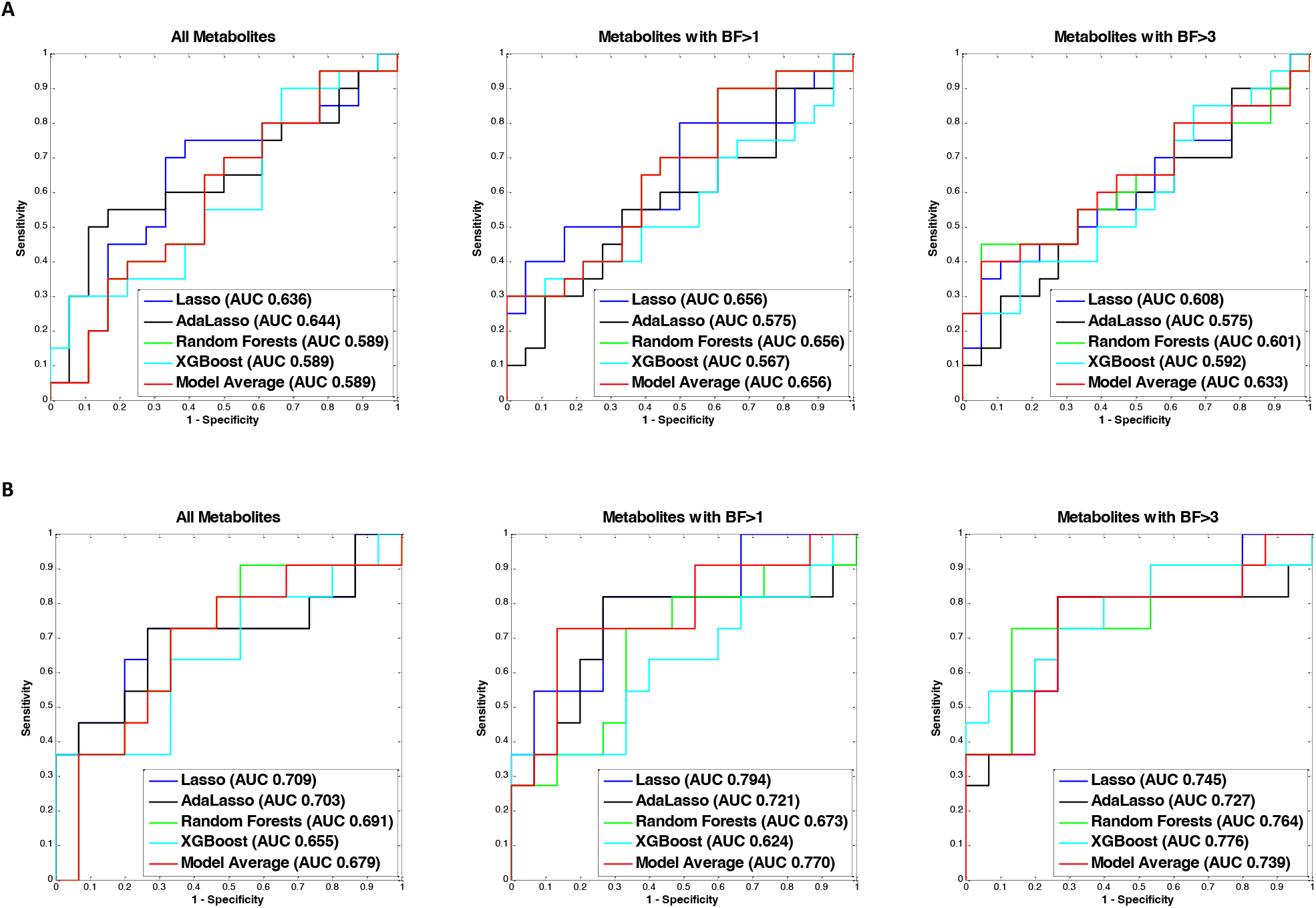
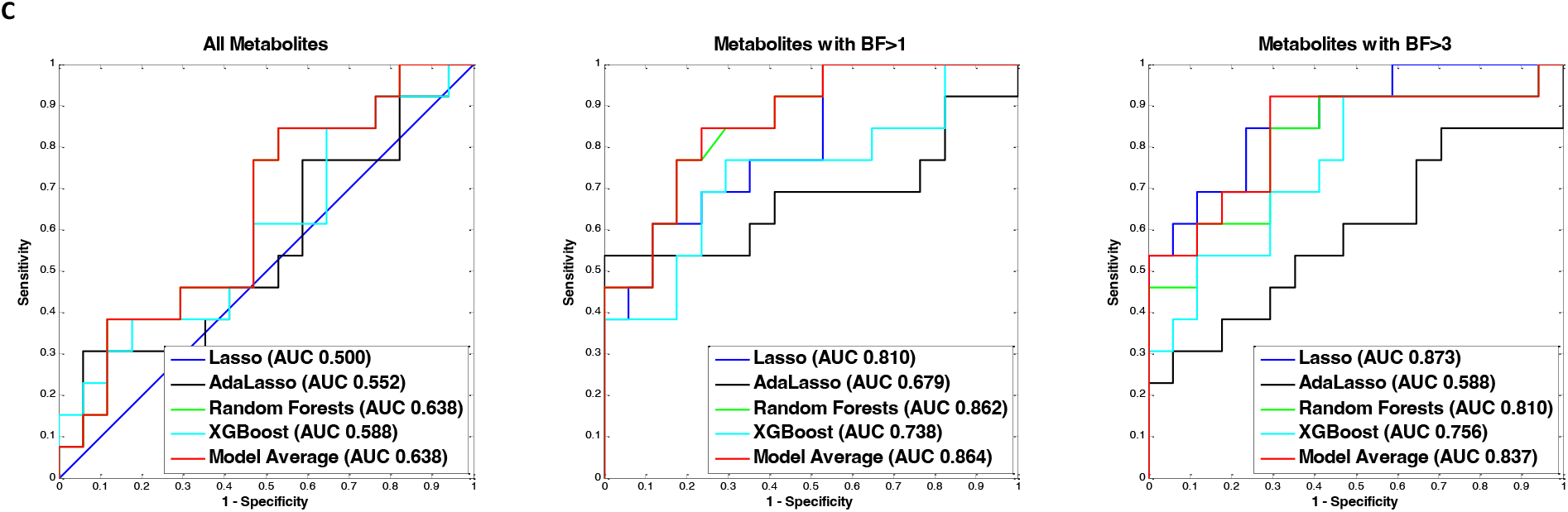
ME/CFS predictive modeling. ME/CFS, myalgic encephalomyelitis/chronic fatigue syndrome; sr-IBS, self-reported physician diagnosed irritable bowel syndrome; BF, BayesFactor; AUC, area under the receiver operating charactistic curve. To differentiate ME/CFS cases from healthy controls, we employed five machine learning algorithms: Lasso, adaptive Lasso (AdaLasso), Random Forests (RF), XGBoost, and Bayesian Model Averaging (Model average). For each algorithm, three sets of predictors were considered: 1) all metabolites, 2) metabolites with BayesFactor > 1, and 3) metabolites with BayesFactor > 3. The predictive models were first trained in the 80% randomly selected training set using 10-fold cross-validation, and the remaining 20% of the study population was used as the independent test set to validate model performance. (**A**) Overall population. (**B**) Women only. (**C**) No GI complaints.

### Correlations between metabolites and ME/CFS symptom severity scores

We investigated whether the plasma levels of metabolites in the metabolic clusters that were significantly altered in ME/CFS (bold in **Supplementary Table S4**) correlated with MFI scales using Spearman’s correlation tests. Heatmaps showing the correlation coefficients in all ME/CFS, all controls, female ME/CFS, female controls, male ME/CFS, and male controls are presented in **Figure 4A-C**.

**Fig. 4.**
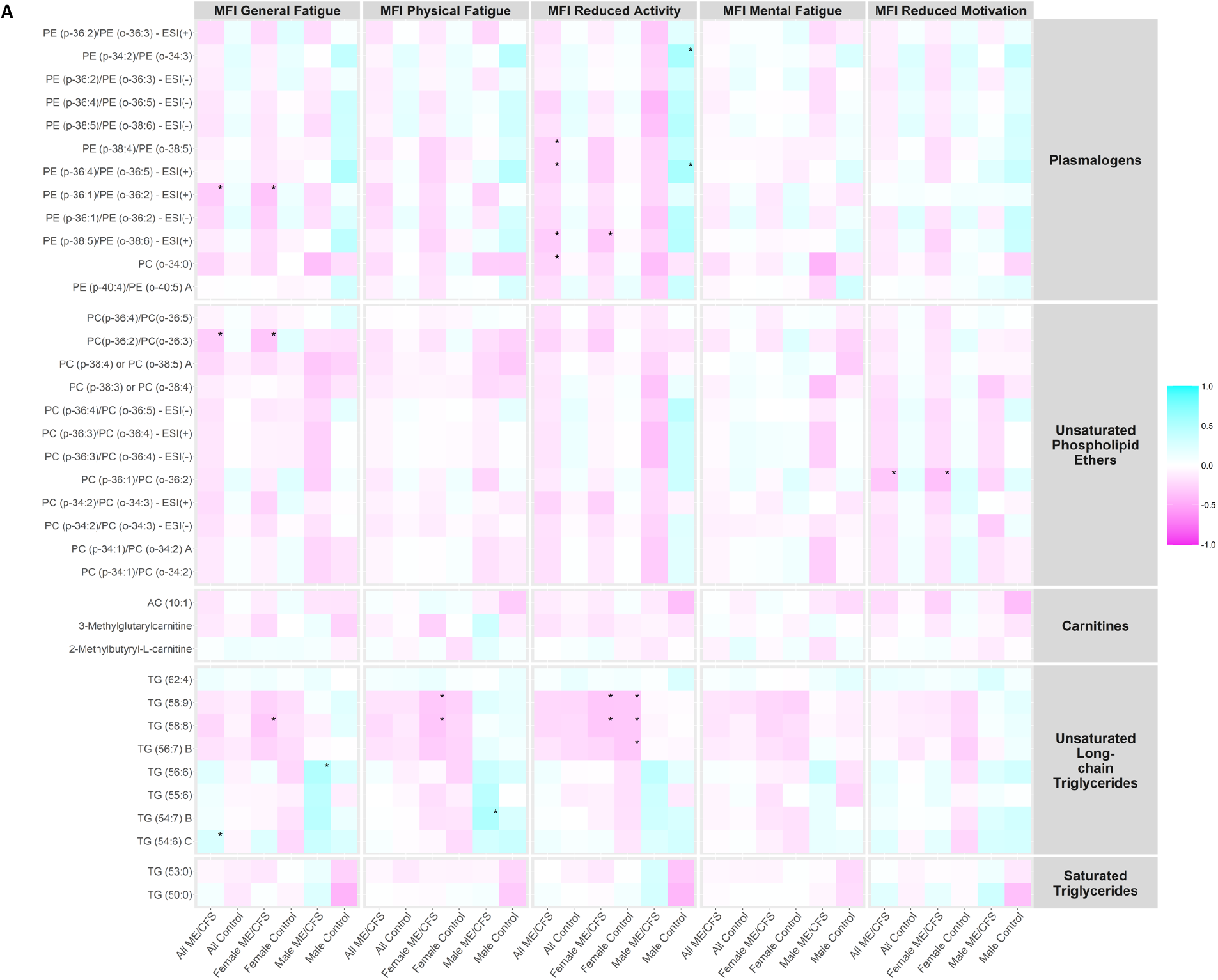
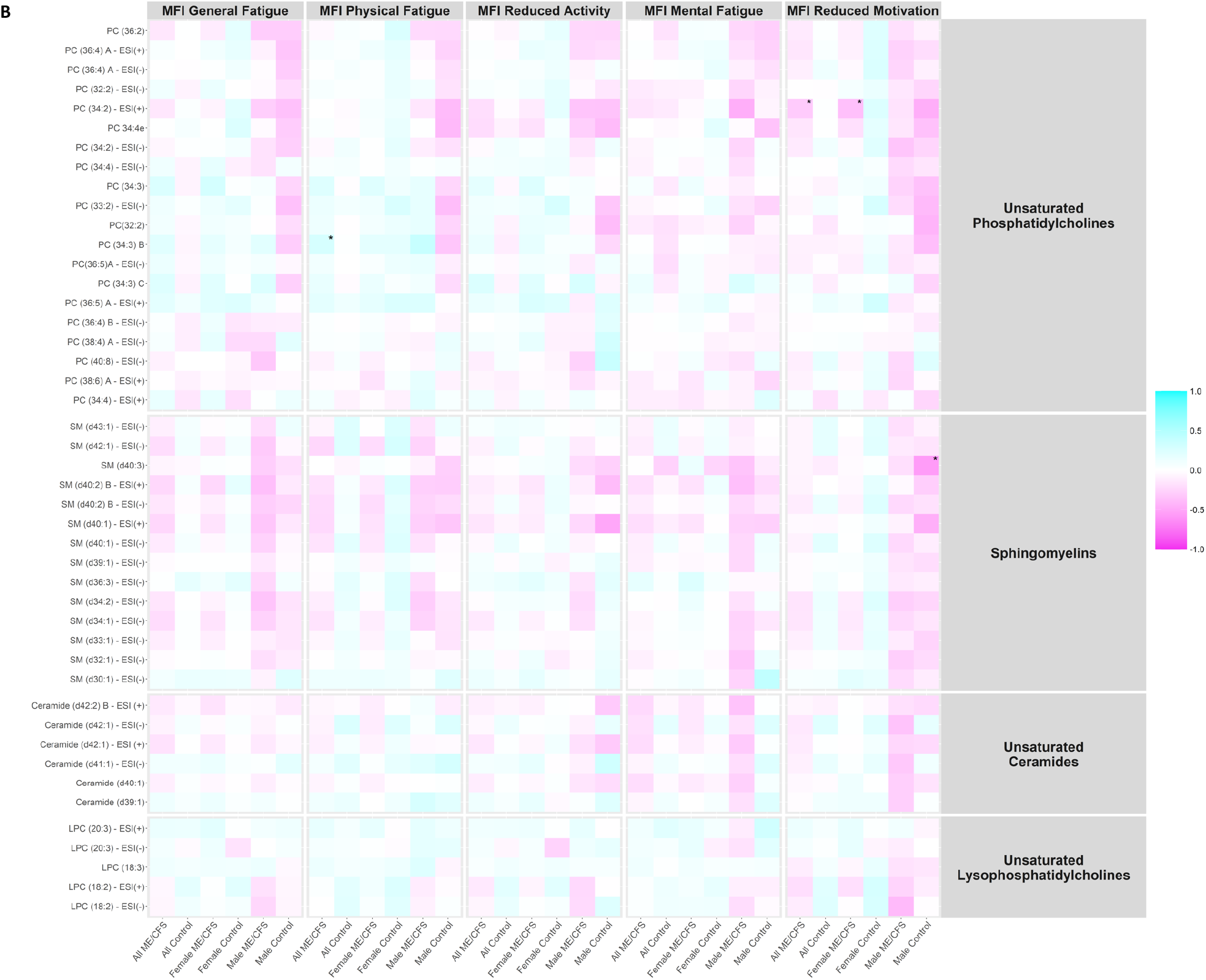
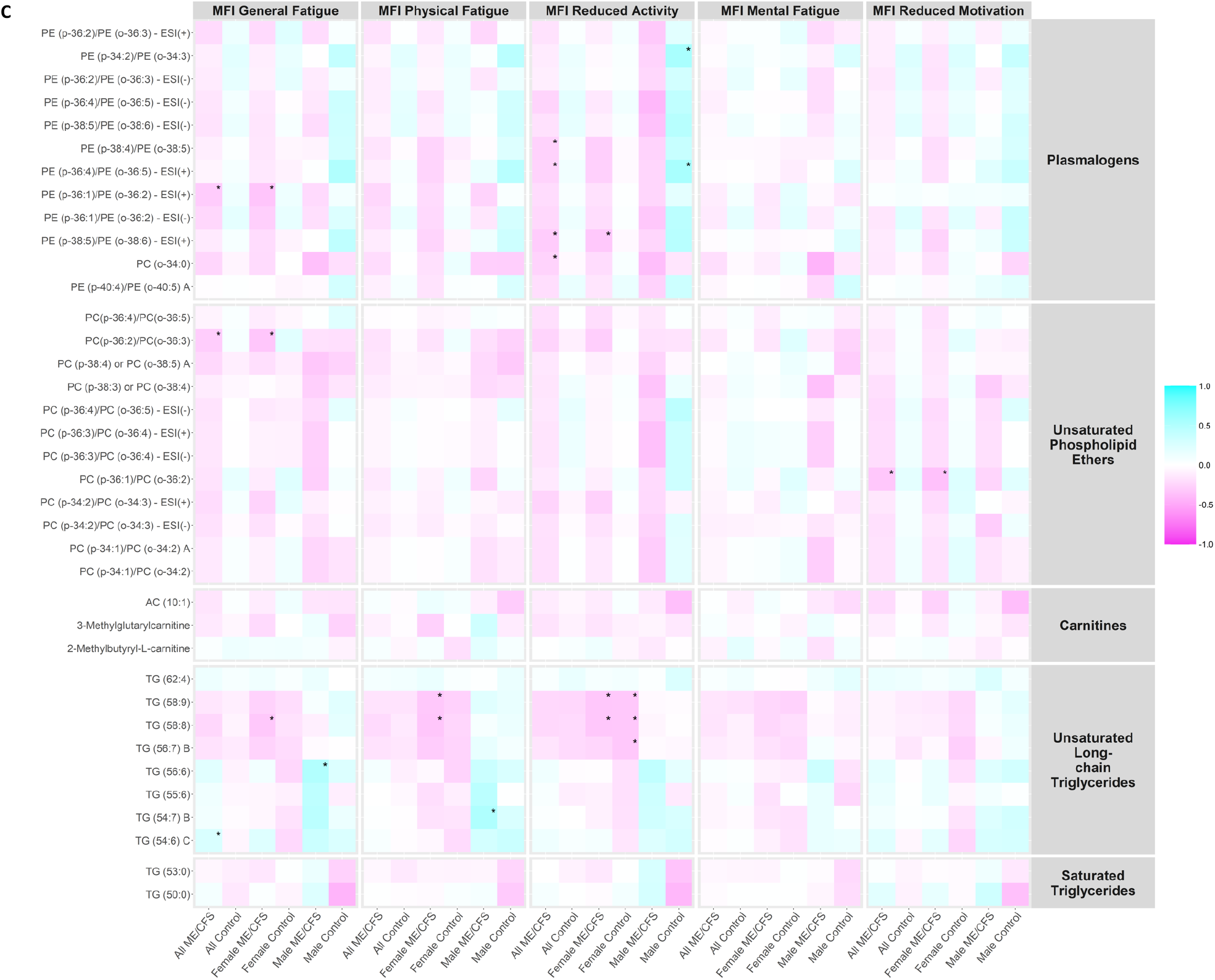
Correlation heatmap. MFI, Multidimensional Fatigue Inventory scored on 0-100 scale with 0 = no fatigue and 100 = maximal fatigue; ME/CFS, myalgic encephalomyelitis/chronic fatigue syndrome. *p-value < 0.01. Heatmap showing the correlation coefficients between the plasma levels of metabolites in the metabolic clusters that were significantly altered in ME/CFS (bold in **Supplementary Table S4**) and MFI scales using Spearman’s correlation tests in all ME/CFS, all controls, female ME/CFS, female controls, male ME/CFS, and male controls. (**A**) Correlation heatmap, part 1. (**B**) Correlation heatmap, part 2. (**C**) Correlation heatmap, part 3.

Notably, within male ME/CFS patients, levels of triglycerides (TG) (56:6) in unsaturated long-chain TG were positively correlated with MFI general fatigue scales (ρ=0.501, p-value=0.005), and levels of TG (54:7) B were positively correlated with MFI physical fatigue scales (ρ=0.519, p-value=0.003). Within male controls, levels of alpha-ketoglutarate in dicarboxylic acids were positively correlated with MFI physical fatigue (ρ=0.558, p-value=0.007), reduced activity (ρ=0.633, p-value=0.002) and reduced motivation (ρ=0.696, p-value<0.001) scales; more severe MFI reduced activity symptoms were associated with higher levels of two plasmalogens (PE (p-34:2)/PE (o-34:3): ρ=0.549, p-value=0.010; PE (p-36:4)/PE (o-36:5) - ESI(+): ρ=0.551, p-value=0.010); levels of SM (d40:3) in sphingomyelins were negatively correlated with MFI reduced activity scales (ρ=-0.557 p-value=0.009).

## Discussion

Since the first reports of large-scale metabolomic studies in people with ME/CFS were published in 2016 by Naviaux [15], several research teams, including Yamano [16], Fluge [9], Hoel [50], and our own [14], have reported metabolomic analyses of plasma. The common threads in the results from all these studies are decreased levels of phospholipids and metabolic dysregulation, suggesting abnormalities in mitochondrial activity that impair energy metabolism. In our study, we also observed decreased levels of phospholipids, especially phosphatidylcholine and sphingolipids. Furthermore, consistent with previous literature [6, 16, 50], our results suggest dysregulation of the cytidine-5’-diphosphocholine (CDP-choline) pathway and the tricarboxylic acid (TCA) cycle.

### Lipid metabolism abnormalities

Regression, Bayesian, and enrichment analyses revealed significant reduction in the levels of plasmalogens. Plasmalogens are abundant phospholipid ethers that protect phospholipids and lipoprotein particles from oxidative stress and associated damage [51, 52]. They are also responsible for maintaining the integrity of membrane structures [51]. Plasmalogen biosynthesis commences in the peroxisomes and is completed in the endoplasmic reticulum [53]. Maintenance of peroxisomal structure with functional enzymes is imperative for both plasmalogen biosynthesis and β-oxidation of very long-chain fatty acids [51]. Peroxisomal β-oxidation of very long-chain fatty acids leads to their breakdown into short-chain products that serve as substrates for mitochondrial β-oxidation [54]. We posit that this crosstalk between mitochondria and peroxisomes plays an important role in maintaining energy homeostasis, and that dysregulation contributes to the fatigue and cognitive dysfunction that are hallmarks of ME/CFS [55].

ME/CFS subjects had a significant reduction in levels of carnitines (**Figure 2A-C**). Carnitines regulate the cellular to mitochondrial ratio of free CoA to acyl-CoA, remove the unwanted acyl groups, and play a key role in the transport of long-chain fatty acids from cytoplasm to the mitochondrial matrix for oxidation [56]. Depletion of carnitines can threaten the integrity of cell and mitochondrial membranes, increase oxidative stress, and reduce the ability to counter inflammation [57]. We also observed increased levels of long-chain TG in ME/CFS. Depletion of carnitines leads to the accumulation of long-chain TG that become targets for lipid peroxidation by mitochondria [58]. The accumulation of toxic lipid peroxidation products can also lead to mitochondrial membrane damage [19]. Where carnitine is depleted and there is mitochondrial overload for fatty acid oxidation, peroxisomal β-oxidation has been reported to be a compensatory process that can produce carnitine as an intermediate product [59].

Peroxisomes regulate fatty acid metabolism through metabolic cross-talk with mitochondria [60]. Missailidis et al. (2021) [55] have reported potential dysregulation in mitochondrial β-oxidation in conjunction with dysregulation in peroxisomal processes in lymphoblasts of ME/CFS patients. Our findings of depleted levels of plasmalogens, unsaturated phospholipid ethers and carnitines are consistent with peroxisomal dysfunction. Peroxisomes also regulate the scavenging of reactive oxygen species (ROS). Redox imbalance is frequently seen in people with ME/CFS [61]; the peroxisomal dysfunction we observed could contribute to and/or reflect this redox imbalance. Finally, peroxisomes are critical in maintaining membrane integrity.

We found depleted levels of PCs in ME/CFS subjects. PCs are the most abundant phospholipids in the mitochondrial membranes [62, 63]. Most PCs are synthesized via the CDP-choline pathway [64]. PCs are essential to the formation of intermediate structures in membrane fusion and fission events, for stabilizing mem brane proteins into their optimal conformations, and for actin-filament disassembly in the end stage of cytokinesis [65-67]. One critical functional implication of reduced levels of PCs is impaired oxidative phosphorylation. PC depletion specifically affects the function and stability of the protein translocases of mitochondria, including the inner membrane translocase TIM23 complex [68] and the outer membrane sorting and assembly machinery (SAM) complex [68, 69]. The destabilization of TIM23 and SAM complexes lead to reduction in mitochondrial membrane potential and impair protein transport and respiratory chain activities [68].

ME/CFS patients were also found to have decreased levels of downstream products of PCs: ceramides, sphingomyelins, lysophosphatidylcholines, phospholipid ethers, prostaglandin D2 (PGD2) and prostaglandin F2α (PGF2α). Depleted levels of lysophosphatidylcholines and phospholipid ethers, as well as of PCs, can impede mitochondrial respiration [65]. Reduced synthesis of PGF2α and PGD2 in phospholipase A2γ-deficient mice induces mitochondrial dysfunction as well as oxidative stress that can contribute to further mitochondrial damage [70]. PCs and their downstream metabolites are important components of the lipid bilayer, and reduction in their levels dysregulate signal transduction across membranes. This alteration in the levels or conformation of membrane components can adversely affect the function of proteins embedded in the membranes such as G protein coupled receptors (GPCRs) [71]. Phospholipids can act as direct allosteric modulators of GPCR activity through the lipid head group that affect ligand binding (agonist and antagonist) and receptor activation [71]. In addition, PCs are precursors to many biologically active molecules that can act as second messengers. Prominent among them are diacylglycerol (DAG), fatty acids, phosphatidic acid, lysophosphatidic acid, N-arachidonylethanolamine, N-palmitoylethanolamine, N-steroylethanolamine and arachidonic acid [72-74]. Ceramides, are not only structural components of membranes, but can also act as second messengers in modulating a range of cellular signaling pathways [75].

Depletions in levels of choline approached but did not meet significance criteria (adjusted p=0.139, BF=2.75, 95% HDI=-0.015 ∼ -0.003). Choline is an essential nutrient; 95% of it is utilized in the synthesis of PCs via the CDP-choline pathway [64]. The remaining 5% exists as either free choline or is used in the synthesis of phosphocholine, glycerophosphocholine, CDP-choline, acetylcholine, and other choline-containing phospholipids like sphingomyelin, plasmalogens and lysophosphatidylcholine. Each of these compounds contributes to maintenance of the structure and signaling functionality of the plasma membrane [64, 72]. IgG autoantibodies that specifically target GPCRs have been reported, even in healthy individuals, but are more commonly found in ME/CFS [76, 77], particularly to autonomic nervous system targets including the M3 Acetylcholine receptor (M3AChR) and β2 Adrenergic receptor (β2AdR). Agonists for each of these receptors have choline precursors, acetylcholine (AC) and epinephrine (adrenaline), respectively. Choline also plays a role in the production of epinephrine, by donating the methyl group. Thus, choline deficiency could potentially lead to the autonomic dysfunction that is found in many people with ME/CFS, with reduced tissue blood flow and oxygen supply, leading to hypoxia, ischemia and fatigue [78]. Depletion in the levels of choline precursor, PCs and their downstream, including ceramides, sphingomyelins and lysophosphatidylcholines, suggest dysregulation of the CDP-choline pathway.

### TCA cycle and other abnormalities

Through enrichment analysis, we found significant elevations in the levels of dicarboxylic acids in ME/CFS subjects. The two TCA cycle intermediates, α-ketoglutarate (α-KG) and succinate, representing the dicarboxylic acids cluster, were elevated in ME/CFS. The TCA cycle is a conserved pathway in aerobic organisms through which the acetyl-CoA from carbohydrates, fats and proteins is converted into ATP [79]. Increased levels of α-KG have been reported previously in ME/CFS patients [12], although we are not aware of previous reports of elevated levels of succinate. Abnormal levels of TCA cycle intermediates suggest inefficiencies in ATP production that may contribute to the fatigue and post-exertional malaise reported in ME/CFS. Increases in α-KG levels have been reported to induce severe metabolic impairment of pyruvate oxidation in the tricarboxylic acid cycle, leading to cell death [79]. Succinate accumulation has been reported to induce HIF-1α stabilization as well as the transcriptional activation of the pro-inflammatory cytokine IL-1β [79]. Elevated succinate levels contribute to increased oxidative stress and neuronal degeneration in rat models [80]. Oxidative stress, in turn, augments nitrosative stress [81]. Nitrosative stress, which has been documented in people with ME/CFS [61, 82], can lead to the increased production of peroxynitrite and downregulate the function of both alpha-ketoglutarate dehydrogenase and succinate dehydrogenase [81-83]. Infection is a common cause of nitrosative stress. Many ME/CFS patients report symptoms consistent with systemic infection prior to the onset of the illness.

Our analyses also revealed reductions in levels of resolvin D1 in ME/CFS. Resolvin D1 is a derivative of docosahexanoic acid (DHA) and contributes to resolution of inflammation by targeting dead cells for clearance by macrophages [84]. Decreased levels of resolvin D1 in ME/CFS might be consistent with the possibilities of inflammatory damage associated with the disease [85, 86].

### Metabolomic findings as biomarkers of disease and of disease severity

To identify biomarkers for ME/CFS, we explored three sets of predictors and five different machine learning models. None of the classifiers differentiated all ME/CFS subjects from controls; however, they did differentiate female ME/CFS patients from female controls, and ME/CFS patients without sr-IBS from controls without sr-IBS, especially when using metabolites with BF>1 or BF>3 as predictors. We further tested their predictive capacities with an independent cohort whose metabolomics profiling we previously explored [14]. The metabolomics assay of the validation set was matched with our current assay, resulting in 630 metabolites in common. The metabolites that overlapped with the three sets of predictors (all, BF>1, BF>3) were fitted into the same machine learning models, and the predictive performance was evaluated using 10-fold cross-validation in the validation dataset. The AUROC values for distinguishing all ME/CFS patients from all controls ranged between 0.514 and 0.738. For differentiating female ME/CFS subjects from female controls, the AUROC values were between 0.616 and 0.784; for differentiating ME/CFS patients without sr-IBS from controls without sr-IBS, the AUROC values ranged between 0.614 and 0.828. The predictive performance was similar in the validation set as observed in the current study (**Supplementary Table S9**), and models with larger BFs as predictors also performed better than those fitted with all metabolites.

From **Figure 4A**, we observed generally higher correlation coefficients in all peroxisome-related metabolites (plasmalogen, phospholipid ethers, carnitines, and long-chain TGs) with energy MFI scales (General Fatigue, Physical Fatigue, and Reduced Activity) than with mental MFI scales (Mental Fatigue and Reduced Motivation). Plasmalogens and phospholipid ethers, in particular, are directly linked to peroxisomal disorders, and were generally negatively correlated with energy MFI scales in ME/CFS and ME/CFS subgroups. This is consistent with the findings in the statistical analyses that levels of these compounds were depleted in ME/CFS compared to controls, and the stronger correlations in the energy MFI scales provided further support to the notion that dysregulated interactions between mitochondria and peroxisomes contribute to fatigue in ME/CFS [55]. The magnitudes of the correlations were more conspicuous in the male cohort possibly by chance due to the smaller sample size.

### Strengths and limitations

The strength of this study lies in the quality of patient characterization, robust metabolomic analysis involving a comprehensive set of compounds, and complete and cautious statistical approaches. Our subgroup analyses focusing on female subjects and subjects without sr-IBS did not reveal dysregulated metabolic clusters different from those in the overall study population. These analyses were limited by small sample sizes in the subgroups of male subjects and subjects with sr-IBS.

## Conclusion

Our findings indicate a series of interconnected metabolic alterations in people with ME/CFS that may contribute to the pathogenesis of ME/CFS: (i) reduced levels of plasmalogens, unsaturated phospholipid ethers, and carnitines suggest peroxisomal dysfunction; (ii) reductions in levels of PCs and their downstream products indicate dysregulation of CDP-choline pathway, and (iii) elevations in the levels of dicarboxylic acids, particularly the TCA cycle intermediates α-ketoglutarate and succinate, are consistent with an impairment in the TCA cycle that may contribute to physical and cognitive fatigue.

## Supporting information

All Supplemental Tables

## Data Availability

All data produced in the present work are contained in the manuscript.

## List of Abbreviations

α -KG: α-ketoglutarate
β2AdR: β2 Adrenergic receptor
AC: acetylcholine
AdaLasso: adaptive Lasso
AUROC: Area under the Receiver Operating Characteristic curve
BA: biogenic amines
BF: Bayes factors
BIC: Bayesian information criterion
BMA: Bayesian Model Average
BMI: body mass index
CDP-choline: cytidine-5’-diphosphocholine
CI: confidence interval
CL: complex lipids
CSH-QTOF MS: quadrupole time-of-flight mass spectrometry
CV: coefficients of variation
DAG: diacylglycerol
DHA: docosahexanoic acid
DSQ: DePaul Symptom Questionnaire
EpODE: ePoxy fatty acids
FDR: false discovery rate
GC-ToF MS: gas chromatography/time-of-flight mass spectrometry
GI: gastrointestinal
GPCR: G protein coupled receptors
HDI: highest density credible interval
HEPE: hydroxy-eicosapentaenoic acid
HILIC-QTOF-MS: hydrophilic interaction liquid chromatography/quadrupole time-of-flight mass spectrometry
Lasso: least absolute shrinkage and selection operator
LC: liquid chromatography
LPC: lysophophatidycholines
M3AChR: M3 Acetylcholine receptor
ME/CFS: myalgic encephalomyelitis/chronic fatigue syndrome
MFI: Multidimensional Fatigue Inventory
MS: mass spectrometry-based assays
OL: oxylipins
PASC: post acute sequelae of SARS-CoV-2 infection
PCA: principal component analysis
PC: phosphatidycholines
PE: phosphatidylethanolamines
PGD2: prostaglandin D2
PGF2α: prostaglandin F2 α
PM: primary metabolites
PSQI: Pittsburgh Sleep Quality Index
RF: Random Forests
ROC: Receiving Operating Characteristic
ROS: reactive oxygen species
SAM: sorting and assembly machinery
SD: standard deviation
SF-36: Short Form 36 Health Survey
SM: sphingomyelin
sr-IBS: self-reported irritable bowel syndrome
TCA: tricarboxylic acid
TG: triglycerides
TMAO: trimethylamine N-oxide

## Declarations

### Ethics approval and consent to participate

This study was approved by the Columbia University Irving Medical Center (CUIMC) Institutional Review Board. All necessary patient/participant consent has been obtained and the appropriate institutional forms have been archived. All necessary patient/participant consent has been obtained and the appropriate institutional forms have been archived.

### Consent for publication

Not applicable.

### Availability of data and materials

All data associated with this study are present in the paper or the Supplementary Materials.

## Author Contributions

Experimental design: XC, BL, DKB, OF, WIL

Case and control recruitment and characterization: SL, DLP, SDV, LB, JGM

Biostatistical analysis: XC, CRB, YY, BL, AC, DMP

Bioinformatics analysis: XC, CRB, BL, DKB, AC

Data interpretation and functional analysis: XC, ARP, SJ, AR, ALK, WIL

Writing and review of manuscript before submission for publication: XC, CRB, ARP, YY, SJ, AR, BL, DKB, AC, DMP, SL, DLP, SDV, LB, MH, JGM, ALK, OF, WIL

All authors read and approved the final manuscript, with the exception of BL, who unfortunately passed away prior to the final submission of this manuscript.

## Acknowledgements

We are grateful to Kelly Magnus of the Center for Infection and Immunity at Columbia University for assistance with manuscript preparation, to Kelly Paglia of the West Coast Metabolomics Center at University of California, Davis for her support and coordination, to the Chronic Fatigue Initiative of the Hutchins Family Foundation and the ME/CFS patients who provided the samples and inspiration that enabled our work. We dedicate this paper to the memory of Bohyun Lee and her contributions to research in ME/CFS.

## References

1. Committee on the Diagnostic Criteria for Myalgic Encephalomyelitis/Chronic Fatigue Syndrome; Board on the Health of Select Populations; Institute of Medicine. Beyond Myalgic Encephalomyelitis/Chronic Fatigue Syndrome: Redefining an Illness. Washington (DC): National Academies Press (US); 2015.

2. Carruthers BM, van de Sande MI, De Meirleir KL, Klimas NG, Broderick G, Mitchell T, et al. Myalgic encephalomyelitis: International Consensus Criteria. J Intern Med. 2011;270(4):327–338.

3. Jason LA, Mirin AA. Updating the National Academy of Medicine ME/CFS prevalence and economic impact figures to account for population growth and inflation. Fatigue: Biomedicine, Health & Behavior. 2021;9(1):9–13.

4. Haney E, Smith ME, McDonagh M, Pappas M, Daeges M, Wasson N, et al. Diagnostic Methods for Myalgic Encephalomyelitis/Chronic Fatigue Syndrome: A Systematic Review for a National Institutes of Health Pathways to Prevention Workshop. Ann Intern Med. 2015;162(12):834–840.

5. Scheibenbogen C, Freitag H, Blanco J, Capelli E, Lacerda E, Authier J, et al. The European ME/CFS Biomarker Landscape project: an initiative of the European network EUROMENE. J Transl Med. 2017;15(1):162.

6. Armstrong CW, McGregor NR, Lewis DP, Butt HL, Gooley PR. Metabolic profiling reveals anomalous energy metabolism and oxidative stress pathways in chronic fatigue syndrome patients. Metabolomics. 2015;11:1626–1639.

7. Armstrong CW, McGregor NR, Lewis DP, Butt HL, Gooley PR. The association of fecal microbiota and fecal, blood serum and urine metabolites in myalgic encephalomyelitis/chronic fatigue syndrome. Metabolomics. 2017;13(8).

8. Armstrong CW, McGregor NR, Sheedy JR, Buttfield I, Butt HL, Gooley PR. NMR metabolic profiling of serum identifies amino acid disturbances in chronic fatigue syndrome. Clin Chim Acta. 2012;413(19-20):1525–1531.

9. Fluge O, Mella O, Bruland O, Risa K, Dyrstad SE, Alme K, et al. Metabolic profiling indicates impaired pyruvate dehydrogenase function in myalgic encephalopathy/chronic fatigue syndrome. JCI Insight. 2016;1(21):e89376.

10. Germain A, Barupal DK, Levine SM, Hanson MR. Comprehensive Circulatory Metabolomics in ME/CFS Reveals Disrupted Metabolism of Acyl Lipids and Steroids. Metabolites. 2020;10(1):34.

11. Germain A, Ruppert D, Levine SM, Hanson MR. Metabolic profiling of a myalgic encephalomyelitis/chronic fatigue syndrome discovery cohort reveals disturbances in fatty acid and lipid metabolism. Mol Biosyst. 2017;13(2):371–379.

12. Germain A, Ruppert D, Levine SM, Hanson MR. Prospective Biomarkers from Plasma Metabolomics of Myalgic Encephalomyelitis/Chronic Fatigue Syndrome Implicate Redox Imbalance in Disease Symptomatology. Metabolites. 2018;8(4):90.

13. McGregor NR, Armstrong CW, Lewis DP, Gooley PR. Post-Exertional Malaise Is Associated with Hypermetabolism, Hypoacetylation and Purine Metabolism Deregulation in ME/CFS Cases. Diagnostics (Basel). 2019;9(3):70.

14. Nagy-Szakal D, Barupal DK, Lee B, Che X, Williams BL, Kahn EJR, et al: Insights into myalgic encephalomyelitis/chronic fatigue syndrome phenotypes through comprehensive metabolomics. Sci Rep. 2018;8(1):10056.

15. Naviaux RK, Naviaux JC, Li K, Bright AT, Alaynick WA, Wang L, et al. Metabolic features of chronic fatigue syndrome. Proc Natl Acad Sci U S A. 2016;113(37):E5472–5480.

16. Yamano E, Sugimoto M, Hirayama A, Kume S, Yamato M, Jin G, et al. Index markers of chronic fatigue syndrome with dysfunction of TCA and urea cycles. Sci Rep. 2016;6:34990.

17. Valdez AR, Hancock EE, Adebayo S, Kiernicki DJ, Proskauer D, Attewell JR, et al. Estimating Prevalence, Demographics, and Costs of ME/CFS Using Large Scale Medical Claims Data and Machine Learning. Front Pediatr. 2018;6:412.

18. Milivojevic M, Che X, Bateman L, Cheng A, Garcia BA, Hornig M, et al: Plasma proteomic profiling suggests an association between antigen driven clonal B cell expansion and ME/CFS. PLoS One 2020;15(7):e0236148.

19. Tomic S, Brkic S, Maric D, Mikic AN. Lipid and protein oxidation in female patients with chronic fatigue syndrome. Arch Med Sci. 2012;8(5):886–891.

20. Aaron LA, Herrell R, Ashton S, Belcourt M, Schmaling K, Goldberg J, et al. Comorbid clinical conditions in chronic fatigue: a co-twin control study. J Gen Intern Med. 2001;16(1):24–31.

21. Giloteaux L, Goodrich JK, Walters WA, Levine SM, Ley RE, Hanson MR. Reduced diversity and altered composition of the gut microbiome in individuals with myalgic encephalomyelitis/chronic fatigue syndrome. Microbiome. 2016;4(1):30.

22. Maes M, Bosmans E, Kubera M. Increased expression of activation antigens on CD8+ T lymphocytes in Myalgic Encephalomyelitis/chronic fatigue syndrome: inverse associations with lowered CD19+ expression and CD4+/CD8+ ratio, but no associations with (auto)immune, leaky gut, oxidative and nitrosative stress biomarkers. Neuro Endocrinol Lett. 2015;36(5):439–446.

23. Nagy-Szakal D, Williams BL, Mishra N, Che X, Lee B, Bateman L, et al. Fecal metagenomic profiles in subgroups of patients with myalgic encephalomyelitis/chronic fatigue syndrome. Microbiome 2017;5(1):44.

24. Fukuda K, Straus SE, Hickie I, Sharpe MC, Dobbins JG, Komaroff A. The chronic fatigue syndrome: a comprehensive approach to its definition and study. International Chronic Fatigue Syndrome Study Group. Ann Intern Med. 1994;121(12):953–959.

25. Carruthers BM, Jain AK, De Meirleir KL, Peterson DL, Klimas NG, Lerner AM, et al. Myalgic Encephalomyelitis/Chronic Fatigue Syndrome. J Chronic Fatigue Syndr. 2003;11(1):7–115.

26. Jason LA, Evans M, Porter N, Brown M, Brown A, Hunnell J, et al. The Development of a Revised Canadian Myalgic Encephalomyelitis Chronic Fatigue Syndrome Case Definition. Am J Biochem Biotechnol. 2010;6(2):120–135.

27. Buysse DJ, Reynolds CF 3rd, Monk TH, Berman SR, Kupfer DJ. The Pittsburgh Sleep Quality Index: a new instrument for psychiatric practice and research. Psychiatry Res. 1989;28(2):193–213.

28. Ware JE Jr., Sherbourne CD. The MOS 36-item short-form health survey (SF-36). I. Conceptual framework and item selection. Med Care. 1992;30(6):473–483.

29. Smets EM, Garssen B, Bonke B, De Haes JC. The Multidimensional Fatigue Inventory (MFI) psychometric qualities of an instrument to assess fatigue. J Psychosom Res. 1995;39(3):315–325.

30. Fiehn O. Metabolomics by Gas Chromatography-Mass Spectrometry: Combined Targeted and Untargeted Profiling. Curr Protoc Mol Biol. 2016;114:30.4.1-30.4.32.

31. Kind T, Wohlgemuth G, Lee DY, Lu Y, Palazoglu M, Shahbaz S, et al. FiehnLib: mass spectral and retention index libraries for metabolomics based on quadrupole and time-of-flight gas chromatography/mass spectrometry. Anal Chem. 2009;81(24):10038–10048.

32. Cajka T, Smilowitz JT, Fiehn O. Validating Quantitative Untargeted Lipidomics Across Nine Liquid Chromatography-High-Resolution Mass Spectrometry Platforms. Anal Chem. 2017;89(22):12360–12368.

33. Tsugawa H, Cajka T, Kind T, Ma Y, Higgins B, Ikeda K, et al. MS-DIAL: data-independent MS/MS deconvolution for comprehensive metabolome analysis. Nat Methods. 2015;12(6):523–526.

34. Kind T, Liu KH, Lee DY, DeFelice B, Meissen JK, Fiehn O. LipidBlast in silico tandem mass spectrometry database for lipid identification. Nat Methods. 2013;10(8):755–758.

35. Bakovic M, Fullerton MD, Michel V. Metabolic and molecular aspects of ethanolamine phospholipid biosynthesis: the role of CTP:phosphoethanolamine cytidylyltransferase (Pcyt2). Biochem Cell Biol. 2007;85(3):283–300.

36. DeFelice BC, Mehta SS, Samra S, Cajka T, Wancewicz B, Fahrmann JF, et al. Mass Spectral Feature List Optimizer (MS-FLO): A Tool To Minimize False Positive Peak Reports in Untargeted Liquid Chromatography-Mass Spectroscopy (LC-MS) Data Processing. Anal Chem. 2017;89(6):3250–3255.

37. Fan S, Kind T, Cajka T, Hazen SL, Tang WHW, Kaddurah-Daouk R, et al. Systematic Error Removal Using Random Forest for Normalizing Large-Scale Untargeted Lipidomics Data. Anal Chem. 2019;91(5):3590–3596.

38. Benjamini Y, Hochberg Y. Controlling the False Discovery Rate - a Practical and Powerful Approach to Multiple Testing. J R Stat Soc Series B Stat Methodol. 1995;57(1):289–300.

39. Barupal DK, Fiehn O. Chemical Similarity Enrichment Analysis (ChemRICH) as alternative to biochemical pathway mapping for metabolomic datasets. Sci Rep. 2017;7(1):14567.

40. Goodrich B, Gabry J, Ali I, Brilleman S. rstanarm: Bayesian applied regression modeling via Stan. 2020. https://mc-stan.org/rstanarm.

41. Makowski D, Ben-Shachar MS, Lüdecke D. bayestestR: Describing Effects and their Uncertainty, Existence and Significance within the Bayseian Framework. J Open Source Softw. 2019;4(40):1541.

42. Makowski D, Ben-Shachar MS, Chen SHA, Ludecke D. Indices of Effect Existence and Significance in the Bayesian Framework. Front Psychol. 2019;10:2767.

43. Jeffreys H. Theory of Probability. 3rd edn. Oxford: Clarendon Press; 1961.

44. Tibshirani R. Regression Shrinkage and Selection Via the Lasso. J R Stat Soc Series B Stat Methodol. 1996;58(1):267–288.

45. Zou H. The Adaptive Lasso and Its Oracle Properties. J Am Stat Assoc. 2012;101(476):1418–1429.

46. Breiman L. Random Forests. Machine Learning. 2001;45:5–32.

47. Chen T, Guestrin C. XGBoost: A Scalable Tree Boosting System. In: 22nd ACM SIGKDD International Conference on Knowledge Discovery and Data Mining. San Francisco, CA. 2016; 785–794.

48. Fan J, Li R: Variable Selection via Nonconcave Penalized Likelihood and its Oracle Properties. J Am Stat Assoc. 2001;96(456):1348–1360.

49. Hoeting JA, Madigan D, Raftery AE, Volinsky CT. Bayesian Model Averaging: A Tutorial. Statistical Science 1999;14(4):382–401.

50. Hoel F, Hoel A, Pettersen IK, Rekeland IG, Risa K, Alme K, et al. A map of metabolic phenotypes in patients with myalgic encephalomyelitis/chronic fatigue syndrome. JCI Insight. 2021;6(16).

51. Messias MCF, Mecatti GC, Priolli DG, de Oliveira Carvalho P. Plasmalogen lipids: functional mechanism and their involvement in gastrointestinal cancer. Lipids Health Dis. 2018;17(1):41.

52. Wanders RJ, Poll-The BT. Role of peroxisomes in human lipid metabolism and its importance for neurological development. Neurosci Lett. 2017;637:11–17.

53. Honsho M, Fujiki Y. Plasmalogen homeostasis - regulation of plasmalogen biosynthesis and its physiological consequence in mammals. FEBS Lett. 2017;591(18):2720–2729.

54. Wanders RJ, Waterham HR, Ferdinandusse S. Metabolic Interplay between Peroxisomes and Other Subcellular Organelles Including Mitochondria and the Endoplasmic Reticulum. Front Cell Dev Biol. 2016;3:83.

55. Missailidis D, Sanislav O, Allan CY, Smith PK, Annesley SJ, Fisher PR. Dysregulated Provision of Oxidisable Substrates to the Mitochondria in ME/CFS Lymphoblasts. Int J Mol Sci 2021;22(4):2046.

56. Flanagan JL, Simmons PA, Vehige J, Willcox MD, Garrett Q. Role of carnitine in disease. Nutr Metab (Lond). 2010;7:30.

57. Li JL, Wang QY, Luan HY, Kang ZC, Wang CB. Effects of L-carnitine against oxidative stress in human hepatocytes: involvement of peroxisome proliferator-activated receptor alpha. J Biomed Sci. 2012;19:32.

58. Vacha GM, Giorcelli G, Siliprandi N, Corsi M. Favorable effects of L-carnitine treatment on hypertriglyceridemia in hemodialysis patients: decisive role of low levels of high-density lipoprotein-cholesterol. Am J Clin Nutr. 1983;38(4):532–540.

59. Violante S, Ijlst L, Te Brinke H, Koster J, Tavares de Almeida I, Wanders RJ, et al. Peroxisomes contribute to the acylcarnitine production when the carnitine shuttle is deficient. Biochim Biophys Acta. 2013;1831(9):1467–1474.

60. Demarquoy J, Le Borgne F. Crosstalk between mitochondria and peroxisomes. World J Biol Chem. 2015;6(4):301–309.

61. Paul BD, Lemle MD, Komaroff AL, Snyder SH. Redox imbalance links COVID-19 and myalgic encephalomyelitis/chronic fatigue syndrome. Proc Natl Acad Sci U S A. 2021;118(34).

62. Sperka-Gottlieb CD, Hermetter A, Paltauf F, Daum G. Lipid topology and physical properties of the outer mitochondrial membrane of the yeast, Saccharomyces cerevisiae. Biochim Biophys Acta. 1988;946(2):227–234.

63. Zinser E, Sperka-Gottlieb CD, Fasch EV, Kohlwein SD, Paltauf F, Daum G. Phospholipid synthesis and lipid composition of subcellular membranes in the unicellular eukaryote Saccharomyces cerevisiae. J Bacteriol. 1991;173(6):2026–2034.

64. Gibellini F, Smith TK. The Kennedy pathway--De novo synthesis of phosphatidylethanolamine and phosphatidylcholine. IUBMB Life. 2010;62(6):414–428.

65. Birner R, Burgermeister M, Schneiter R, Daum G. Roles of phosphatidylethanolamine and of its several biosynthetic pathways in Saccharomyces cerevisiae. Mol Biol Cell. 2001;12(4):997–1007.

66. Dowhan W, Bogdanov M. Lipid-dependent membrane protein topogenesis. Annu Rev Biochem. 2009;78:515–540.

67. Furt F, Moreau P. Importance of lipid metabolism for intracellular and mitochondrial membrane fusion/fission processes. Int J Biochem Cell Biol. 2009;41(10):1828–1836.

68. Schuler MH, Di Bartolomeo F, Martensson CU, Daum G, Becker T. Phosphatidylcholine Affects Inner Membrane Protein Translocases of Mitochondria. J Biol Chem. 2016;291(36):18718–18729.

69. Schuler MH, Di Bartolomeo F, Bottinger L, Horvath SE, Wenz LS, Daum G, et al. Phosphatidylcholine affects the role of the sorting and assembly machinery in the biogenesis of mitochondrial beta-barrel proteins. J Biol Chem. 2015;290(44):26523–26532.

70. Yoda E, Hachisu K, Taketomi Y, Yoshida K, Nakamura M, Ikeda K, et al. Mitochondrial dysfunction and reduced prostaglandin synthesis in skeletal muscle of Group VIB Ca2+-independent phospholipase A2gamma-deficient mice. J Lipid Res. 2010;51(10):3003–3015.

71. Dawaliby R, Trubbia C, Delporte C, Masureel M, Van Antwerpen P, Kobilka BK, et al. Allosteric regulation of G protein-coupled receptor activity by phospholipids. Nat Chem Biol. 2016;12(1):35–39.

72. Li Z, Vance DE. Phosphatidylcholine and choline homeostasis. J Lipid Res. 2008;49(6):1187–1194.

73. Momchilova A, Markovska T. Phosphatidylethanolamine and phosphatidylcholine are sources of diacylglycerol in ras-transformed NIH 3T3 fibroblasts. Int J Biochem Cell Biol. 1999;31(2):311–318.

74. Okamoto Y, Morishita J, Tsuboi K, Tonai T, Ueda N. Molecular characterization of a phospholipase D generating anandamide and its congeners. J Biol Chem. 2004;279(7):5298–5305.

75. Mathias S, Kolesnick R. Ceramide: a novel second messenger. Adv Lipid Res. 1993;25:65–90.

76. Cabral-Marques O, Marques A, Giil LM, De Vito R, Rademacher J, Gunther J, et al. GPCR-specific autoantibody signatures are associated with physiological and pathological immune homeostasis. Nat Commun. 2018;9(1):5224.

77. Loebel M, Grabowski P, Heidecke H, Bauer S, Hanitsch LG, Wittke K, et al. Antibodies to beta adrenergic and muscarinic cholinergic receptors in patients with Chronic Fatigue Syndrome. Brain Behav Immun. 2016;52:32–39.

78. Wirth K, Scheibenbogen C. A Unifying Hypothesis of the Pathophysiology of Myalgic Encephalomyelitis/Chronic Fatigue Syndrome (ME/CFS): Recognitions from the finding of autoantibodies against ss2-adrenergic receptors. Autoimmun Rev. 2020;19(6):102527.

79. Martinez-Reyes I, Chandel NS. Mitochondrial TCA cycle metabolites control physiology and disease. Nat Commun. 2020;11(1):102.

80. Zhang Y, Zhang M, Zhu W, Yu J, Wang Q, Zhang J, et al. Succinate accumulation induces mitochondrial reactive oxygen species generation and promotes status epilepticus in the kainic acid rat model. Redox Biol. 2020;28:101365.

81. Palmieri EM, Gonzalez-Cotto M, Baseler WA, Davies LC, Ghesquiere B, Maio N, et al. Nitric oxide orchestrates metabolic rewiring in M1 macrophages by targeting aconitase 2 and pyruvate dehydrogenase. Nat Commun. 2020;11(1):698.

82. Morris G, Berk M, Klein H, Walder K, Galecki P, Maes M. Nitrosative Stress, Hypernitrosylation, and Autoimmune Responses to Nitrosylated Proteins: New Pathways in Neuroprogressive Disorders Including Depression and Chronic Fatigue Syndrome. Mol Neurobiol. 2017;54(6):4271–4291.

83. Morris G, Maes M. Mitochondrial dysfunctions in myalgic encephalomyelitis/chronic fatigue syndrome explained by activated immuno-inflammatory, oxidative and nitrosative stress pathways. Metab Brain Dis. 2014; 29(1):19–36.

84. Gerlach BD, Marinello M, Heinz J, Rymut N, Sansbury BE, Riley CO, et al. Resolvin D1 promotes the targeting and clearance of necroptotic cells. Cell Death Differ. 2020;27(2):525–539.

85. Komaroff AL. Inflammation correlates with symptoms in chronic fatigue syndrome. Proc Natl Acad Sci U S A. 2017;114(34):8914–8916.

86. Nakatomi Y, Kuratsune H, Watanabe Y. Neuroinflammation in the Brain of Patients with Myalgic Encephalomyelitis/Chronic Fatigue Syndrome. Brain Nerve. 2018;70(1):19–25.

