## Supplementary material for "Evidence for Peroxisomal Dysfunction and Dysregulation of the CDP-Choline Pathway in Myalgic Encephalomyelitis/Chronic Fatigue Syndrome": All Supplemental Tables

**Supplementary Table S1.** Sample mean and standard deviations of metabolites in ME/CFS patients, controls, and their subgroups.

| Metabolite | ME/CFS (n=106) | Control (n=91) | Female ME/CFS (n=75) | Female Control (n=69) |
|---|---|---|---|---|
| | mean (SD) | mean (SD) | mean (SD) | mean (SD) |
| **Primary Metabolites (PM)** | | | | |
| 1,2,4-benzenetriol | 1079.549(1400.131) | 1012.135(991.038) | 1219.986(1622.134) | 1003.791(974.943) |
| 1-methylgalactose NIST | 10405.216(25620.129) | 6749.551(12633.562) | 9280.083(19819.358) | 7326.91(14383.141) |
| 2-aminobutyric acid | 6778.127(2312.798) | 6819.584(2333.931) | 6538.875(2122.79) | 6681.09(2450.474) |
| 2-deoxytetronic acid | 3802(1871.5) | 3811.933(1810.183) | 3405.014(1336.781) | 3496.373(1349.925) |
| 2-hydroxybutanoic acid | 37828.971(19975.01) | 35735.831(20029.106) | 35074.056(19703.304) | 33061.194(17514.433) |
| 2-hydroxyvaleric acid | 4237.382(2334.07) | 4287.303(2132.918) | 4003.222(1997.14) | 4230.507(2200.495) |
| 2-ketoisocaproic acid | 18813.069(5556.745) | 19149.258(6078.798) | 17882.208(5701.625) | 18095.209(5592.465) |
| 3-hydroxybutyric acid | 39212.265(52464.62) | 34152(42085.017) | 40661.653(58622.991) | 35564.567(37757.331) |
| 4-hydroxybutyric acid | 2256.422(736.812) | 2269.843(849.082) | 2271.958(777.963) | 2256.269(835.419) |
| adipic acid | 2895.686(1583.539) | 2847.101(1343.166) | 3036.819(1790.081) | 2859.149(1326.626) |
| alanine | 73245.52(30295.714) | 75399.281(33101.158) | 73297.403(29228.707) | 76971.896(35025.025) |
| alloxanoic acid | 2006.5(1375.468) | 1837.64(1056.367) | 1910.056(1352.394) | 1729.866(1008.286) |
| alpha-ketoglutarate | 1657.627(531.612) | 1527.562(557.531) | 1568.597(485.248) | 1502.254(536.214) |
| aminomalonate | 3619.186(1973.13) | 2911.888(1665.318) | 3724.167(2146.781) | 3059.761(1777.82) |
| arachidic acid | 4924.814(2338.338) | 4808.124(2355.798) | 4992.903(2595.024) | 4769.925(2184.92) |
| behenic acid | 2886.951(836.07) | 3031.506(992.687) | 2889.347(920.857) | 3044.09(951.682) |
| benzoic acid | 17834.373(6150.942) | 17061.82(6244.429) | 17735.486(6186.192) | 17418.731(6079.543) |
| beta-alanine | 1309.833(728.789) | 1377.112(890.61) | 1245.958(556.16) | 1391.507(984.314) |
| capric acid | 1629.931(794.154) | 1873.809(1125.953) | 1659.306(869.109) | 1926.597(1199.193) |
| caprylic acid | 5704.039(1875.061) | 5854.764(2939.166) | 5768.153(1874.782) | 5987.373(3013.043) |
| citric acid | 41473.245(20630.652) | 41135.135(19465.808) | 42914.653(22662.022) | 43440.642(20818.822) |
| citrulline | 4478.147(1507.009) | 4592.506(1393.12) | 4498.722(1671.082) | 4685.075(1358.663) |
| conduritol-beta-expoxide | 1309.863(794.473) | 1296.146(815.46) | 1439.264(869.208) | 1299.627(854.864) |
| creatine | 7813.647(4623.762) | 7540.27(4234.565) | 7484.681(4610.221) | 7697.672(4194.764) |
| creatinine | 8228.716(4668.248) | 8192.91(4583.811) | 8071.514(4793.075) | 8733.493(4284.52) |
| erythritol | 7881.353(18212.112) | 8811.472(35942.446) | 7359.583(16455.919) | 8897.06(40653.41) |
| fructose | 2597.775(3830.73) | 2215.82(2366.76) | 2047.014(2353.022) | 2276.851(2639.361) |
| fumaric acid | 9386.029(3097.763) | 9434.101(2971.299) | 9300.639(3300.011) | 9383.06(3025.01) |
| gluconic acid | 783.971(293.026) | 776.438(203.662) | 808.403(326.755) | 812.418(190.038) |
| glucose | 388539.039(113173.178) | 378196.978(110227.014) | 381069.236(112778.529) | 386519.015(103390.2) |
| glucose-1-phosphate | 2858.431(1035.667) | 2901.236(1183.342) | 2806.708(931.937) | 2919.687(1297.013) |
| glucuronic acid | 1283.441(842.594) | 1067.326(518.171) | 1182.639(562.989) | 1090.776(561.713) |
| glutamic acid | 8913.157(3993.644) | 8273.382(4085.076) | 8536.792(3830.733) | 8214.761(4266.633) |
| glutamine | 30937.275(21124.794) | 32325.416(15805.25) | 31662.556(22695.996) | 34107.239(16505.65) |
| glutaric acid | 568.461(206.452) | 532.449(209.713) | 573.208(220.796) | 535.06(202.663) |
| glyceric acid | 10747.598(4862.652) | 9509.18(4092.259) | 10589.486(4595.623) | 9543.403(4305.927) |
| glycerol | 94812.804(37180.866) | 95386.449(35827.521) | 100344.306(37690.486) | 99697.985(36613.08) |
| glycerol-alpha-phosphate | 970.676(374.848) | 952.978(388.821) | 991.222(373.441) | 959.343(403.965) |
| glycine | 68370.147(24574.146) | 66183.809(20178.354) | 73137.5(26791.978) | 69909.254(20799.518) |
| glycolic acid | 6131.461(2349.621) | 6039.528(2582.552) | 6211.681(2417.159) | 6016.627(2773.779) |
| heptadecanoic acid | 4659.657(1498.193) | 4722.045(1568.799) | 4736.75(1623.265) | 4662.478(1570.389) |
| hydroxycarbamate NIST | 9159.343(4227.978) | 9278.079(4002.355) | 9397.375(4421.922) | 9357.597(4099.154) |
| indole-3-acetate | 2888.461(1569.254) | 2930.517(2502.67) | 2828.514(1675.344) | 3057.955(2831.206) |
| indole-3-lactate | 1920.853(610.853) | 2047.753(765.819) | 1834.125(531.593) | 1942.627(629.712) |
| indole-3-propionic acid | 1758.569(1386.393) | 2146.528(1481.229) | 1732.056(994.58) | 2217.716(1584.021) |
| isoleucine | 42005.147(16265.503) | 42337.787(14176.603) | 39405.694(14415.12) | 41216.806(14003.215) |
| isopropylbenzene | 16849.167(16235.754) | 16249.494(18003.148) | 18402.083(17917.397) | 16790.97(20151.387) |

**Supplementary Table S1.** Sample mean and standard deviations of metabolites in ME/CFS patients, controls, and their subgroups.

| Metabolite | ME/CFS (n=106) | Control (n=91) | Female ME/CFS (n=75) | Female Control (n=69) |
|---|---|---|---|---|
| | mean (SD) | mean (SD) | mean (SD) | mean (SD) |
| isothreonic acid | 1867.461(718.975) | 1810.944(701.346) | 1844.194(735.944) | 1898.612(674.425) |
| lactic acid | 192717.833(119615.393) | 173890.775(103094.546) | 180763.528(111652.076) | 162160.896(91976.988) |
| lauric acid | 8363.078(12244.741) | 10000.079(13605.883) | 9131(14384.754) | 11053.09(15461.218) |
| leucine | 70058.255(22610.471) | 77074.056(24098.618) | 67000.458(22724.459) | 75390(24758.076) |
| levoglucosan | 1659.137(1216.05) | 1870.697(1782.358) | 1726.236(1329.44) | 1800.09(1389.732) |
| linoleic acid | 1164.52(386.052) | 1194.708(500.78) | 1203.625(364.979) | 1207.209(520.374) |
| lysine | 15344.127(10811.81) | 15204.719(7437.624) | 15836.986(10789.329) | 15994.418(7552.007) |
| lysine | 18427.5(13565.891) | 18658(9668.614) | 19624.056(13111.026) | 19306.612(9738.677) |
| lyxitol | 3098.794(1031.473) | 3248.989(1189.283) | 3096.708(970.26) | 3127.582(1000.34) |
| maleic acid | 878.186(349.551) | 788.416(361.008) | 874.556(374.467) | 809.09(349.401) |
| maleimide | 8053.353(10208.802) | 6862.888(5737.055) | 8891.208(11955.832) | 6344.701(3004.902) |
| malic acid | 1120.422(375.058) | 1096.719(407.251) | 1105.236(362.52) | 1111.716(361.939) |
| maltose | 1438.294(1117.335) | 1611.483(3326.737) | 1404.944(780.681) | 1754.448(3794.843) |
| mannitol | 7384.294(34427.962) | 2531.82(2424.356) | 9210.819(40878.375) | 2354.657(1910.561) |
| mannose | 23546.755(8923.898) | 30536.393(70206.191) | 22653.167(9106.751) | 33367.104(80635.606) |
| methionine | 2349.51(1025.929) | 2403.135(919.059) | 2337.778(1119.461) | 2507.567(850.817) |
| myo-inositol | 21222.363(6524.406) | 20873.449(7257.918) | 21375.847(6329.61) | 20822.343(7312.203) |
| myristic acid | 2466.971(764.975) | 2620.674(814.52) | 2514.778(810.279) | 2671.358(814.573) |
| N-acetylornithine | 2287.627(630.487) | 2420.708(709.39) | 2296.653(679.83) | 2434.06(749.283) |
| N-acetylputrescine | 2335.549(2188.271) | 2392.753(2148.301) | 2130.389(1024.341) | 2557.403(2426.534) |
| nicotinic acid | 22012.588(31343.256) | 23151.326(37119.635) | 22864.819(32433.543) | 26788.985(40997.646) |
| ornithine | 22444.167(16004.833) | 23350.124(11618.996) | 22788.375(16240.437) | 24179.194(11856.253) |
| oxalic acid | 38138.853(34967.284) | 31155.685(23852.651) | 35563.861(35006.662) | 30009.03(24888.703) |
| oxoproline | 148843.422(35478.319) | 149227.169(31627.972) | 147583.903(40185.571) | 149267.433(32922.76) |
| palmitic acid | 83826.745(26436.792) | 87530.236(29254.435) | 85898.806(28513.631) | 85002.239(28878.676) |
| palmitoleic acid | 620.618(273.823) | 620.933(267.671) | 654.528(303.431) | 620.821(255.343) |
| pelargonic acid | 29902.833(10040.315) | 30430.506(13250.889) | 30961.722(10552.778) | 30450.269(12658.324) |
| pentadecanoic acid | 10300.402(2114.862) | 10098.079(2516.636) | 10283.056(2261.178) | 10342.313(2451.259) |
| phenylalanine | 21140.618(6021.369) | 22473.629(6495.431) | 20492.139(5953.159) | 22896.507(6363.749) |
| phosphate | 62360.137(25668.008) | 60891.899(24903.713) | 67682.625(26025.372) | 64793.806(25638.342) |
| phthalic acid | 6103.99(2526.222) | 6071.337(3064.102) | 6283.014(2757.299) | 6236.478(3381.886) |
| proline | 10358.696(7248.838) | 10114.865(6954.499) | 10278.653(7595.157) | 9602.463(6087.033) |
| pseudo uridine | 2775.873(653.848) | 2855.146(716.503) | 2839.042(677.462) | 2909.299(750.373) |
| pyrrole-2-carboxylic acid | 6415.049(4040.235) | 5793.022(3390.2) | 6070.722(3862.072) | 5839.91(3286.061) |
| pyruvic acid | 11489.667(4788.685) | 10820.404(3860.672) | 11843.722(4927.602) | 10898.537(3981.009) |
| quinic acid | 1272.48(1447.647) | 1600.753(1769.131) | 1207.264(1395.694) | 1627.761(1726.716) |
| ribonic acid | 873.931(245.494) | 868.337(240.982) | 878.722(246.588) | 867.343(256.395) |
| ribose | 2038.549(602.463) | 2037.371(644.28) | 1952.694(588.947) | 2069.731(671.325) |
| salicylic acid | 4293.333(14512.238) | 2853.427(9490.954) | 4624.556(16288.869) | 3082.567(10830.814) |
| serine | 37492.01(17259.952) | 38555.775(15218.55) | 39316.236(19150.177) | 40813.806(15539.561) |
| stearic acid | 378059.412(154306.997) | 405425.36(162088.008) | 392137.014(173120.537) | 389522.836(144375.349) |
| succinic acid | 2428.686(828.589) | 2106.888(729.61) | 2445.069(869.246) | 2112.403(771.778) |
| sucrose | 671.637(698.395) | 613.989(381.592) | 744.056(789.835) | 635.194(402.983) |
| tagatose | 4063.725(4123.989) | 3589.213(2553.245) | 3440.903(2751.873) | 3661.881(2813.897) |
| threitol | 1452.196(807.611) | 1395.584(582.087) | 1511.361(901.417) | 1392.925(611.212) |
| threonic acid | 8560.99(4195.661) | 7285.618(3051.907) | 8602.167(4062.708) | 7531(3287.101) |
| threonine | 13282.686(4817.033) | 14237.438(5929.372) | 13235.931(5140.448) | 15006.119(6365.817) |
| trans-4-hydroxyproline | 1700(1224.851) | 1418.764(658.035) | 1794.542(1357.079) | 1371.925(553.495) |

**Supplementary Table S1.** Sample mean and standard deviations of metabolites in ME/CFS patients, controls, and their subgroups.

| Metabolite | ME/CFS (n=106) | Control (n=91) | Female ME/CFS (n=75) | Female Control (n=69) |
|---|---|---|---|---|
| | mean (SD) | mean (SD) | mean (SD) | mean (SD) |
| tryptophan | 21152.706(10249.693) | 21495.079(8899.268) | 20616.319(9846.47) | 21355.015(8793.98) |
| tyrosine | 42719.343(12721.061) | 44217.124(10941.659) | 41085.653(12684.991) | 44149.269(9748.327) |
| urea | 785114.216(265741.232) | 831665.056(279983.579) | 754943.208(248424.764) | 838122.119(274463.33) |
| uric acid | 32108.833(16219.644) | 33489.944(17068.328) | 29210.222(15439.048) | 29761.881(13946.09) |
| valine | 79110.618(21747.997) | 82792.506(32138.457) | 75807.153(20507.272) | 81418.313(34557.102) |
| **Biogenic Amines (BA)** | | | | |
| (2R)-3-Hydroxyisovaleroylcarnitine | 1880.687(1046.064) | 1825.6(555.898) | 1736.25(759.31) | 1762.654(565.708) |
| (3-Carboxypropyl)trimethylammonium cation | 16403.7(5302.415) | 17555.878(5550.678) | 15580.716(5251.357) | 16940.727(5053.444) |
| (R)-Butyrylcarnitine | 34078.718(15321.062) | 37105.876(16533.628) | 34088.945(16762.288) | 36207.842(16171.868) |
| .beta.-Phenyl-.gamma.-aminobutyric acid | 869.907(2440.04) | 709.92(241.154) | 643.618(301.777) | 721.387(263.381) |
| .epsilon.-Caprolactam | 87755.933(68018.05) | 89742.427(68532.183) | 86328.183(70251.29) | 85416.594(66739.158) |
| 1,2-Dimethylimidazole | 1819.366(2845.633) | 1694.49(1032.734) | 2037.18(3377.863) | 1762.948(1162.502) |
| 1-Acetyl-3-piperidinamine | 1839.3(818.798) | 1738.972(605.781) | 1852.274(813.83) | 1703.792(523.936) |
| 1-Acetyl-4-piperidinamine | 2047.585(2838.897) | 1533.704(815.512) | 1979.034(2088.427) | 1536.323(805.703) |
| 1-Methyladenosine A | 4344.676(1038.43) | 4455.103(1138.778) | 4429.169(1077.875) | 4439.09(1166.254) |
| 1-Methyl-L-histidine | 22958.602(14651.539) | 23284.637(14336.386) | 22354.704(14663.536) | 23572.374(14325.805) |
| 1-Methylnicotinamide | 14470.114(11101.857) | 14251.823(7589.804) | 14418.734(9495.696) | 14476.406(7942.219) |
| 1-Monostearin | 2115.305(1451.321) | 2830.121(4236.602) | 2177.064(1526.767) | 2601.232(2151.589) |
| 1-Oleoyl-2-acetyl-sn-glycerol | 9191.209(8932.606) | 14964.193(39026.124) | 9841.135(10395.606) | 16928.701(44698.374) |
| 1-Oleoyl-sn-glycero-3-phosphoethanolamine | 6355.005(5324.408) | 6525.509(3772.709) | 6095.835(5637.756) | 6338.457(3307.175) |
| 1-Palmitoyl-2-hydroxy-sn-glycero-3-phosphoethanolamine | 6410.996(4032.932) | 6606.582(3360.162) | 6517.585(4263.178) | 6682.955(3226.653) |
| 1-Phenylpyrrolidine | 143.457(290.638) | 117.902(39.385) | 160.128(346.32) | 120.76(41.362) |
| 1-Stearoyl-2-arachidonoyl-sn-glycero-3-phospho-(1'-myo-inositol) | 1329.25(881.122) | 1346.97(825.295) | 1412.649(948.673) | 1375.1(847.811) |
| 1-Stearoyl-2-arachidonoyl-sn-glycero-3-phosphoserine | 1894.838(2682.073) | 1778.287(2056.277) | 1856.246(2611.087) | 1935.426(2038.887) |
| 1-Stearoyl-2-hydroxy-sn-glycero-3-phosphoethanolamine | 6094.917(4604.091) | 5803.43(2687.326) | 6235.181(4995.225) | 5778.688(2431.299) |
| 2,2',2''-Nitrilotriethanol | 4650.025(6102.206) | 5725.159(8642.651) | 5320.019(7154.417) | 6576.946(9784.759) |
| 2,2-Bishydroxymethyl]-2,2',2''-nitrilotriethanol | 661.365(1011.212) | 683.907(582.601) | 729.049(1200.747) | 729.169(650.841) |
| 2,6-Diaminopimelic acid | 658.783(580.92) | 497.274(446.965) | 678.564(585.353) | 508.356(456.288) |
| 2-Amino-1-phenylethanol | 50064.106(14796.501) | 51939.812(12195.207) | 48749.621(14343.952) | 52299.921(13191.029) |
| 2-Hydroxyibuprofen | 3646.161(11203.95) | 2794.402(8390.204) | 2716.27(5046.864) | 2665.938(9448.507) |
| 2-Indolinone | 22360.698(11782.007) | 25707.648(13967.826) | 23360.713(12417.222) | 25395.462(14495.492) |
| 2-Methylbutyryl-L-carnitine | 15742.272(6758.335) | 19766.912(10894.625) | 14167.581(6470.11) | 17337.175(7188.976) |
| 3-(1-Pyrazolyl)-alanine | 358.412(1009.805) | 338.461(595.673) | 321.331(714.353) | 400.74(672.044) |
| 3,4-Dimethoxybenzaldehyde | 2111.85(1817.258) | 2433.796(3874.558) | 2231.388(2013.467) | 2447.356(4335.605) |
| 3,5-Dihydroxyphenylglycine | 1521.258(1937.934) | 1263.783(1225.374) | 1742.967(2202.951) | 1342.855(1314.494) |
| 3-Amino-1-propanol | 14759.177(17371.398) | 11356.849(7175.122) | 15972.716(19546.631) | 10627.843(6034.141) |
| 3-Aminoquinoline | 2768.697(3284.974) | 3229.232(3720.884) | 2992.714(3269.222) | 3676.498(3966.221) |
| 3-Cysteinylacetaminophen | 85.628(278.461) | 31.56(22.915) | 86.812(304.878) | 32.108(22.791) |
| 3-Dehydrocarnitine | 2713.916(1241.044) | 2870.766(1231.212) | 2693.857(1322.487) | 2599.336(925.909) |
| 3-Hydroxybutyrylcarnitine | 3061.608(2849.628) | 2805.221(2351.317) | 3310.658(3136.113) | 2772.17(1938.867) |
| 3-Hydroxyoleylcarnitine | 1499.787(2022.956) | 1253.212(1804.396) | 1372.661(1923.433) | 1130.92(1716.403) |
| 3-Hydroxypyridine | 1971.739(3569.602) | 3209.464(4987.632) | 1482.697(2804.788) | 3347.393(5367.51) |
| 3-Methylglutarylcarnitine | 575.563(258.16) | 1052.615(1965.769) | 572.823(284.547) | 659.9(336.824) |
| 3-Methylxanthine | 555.738(650.872) | 522.536(446.424) | 602.522(690.615) | 568.763(477.615) |
| 3-Pyridinemthanol | 1549.304(1050.899) | 1769.148(2316.24) | 1582.528(1146.057) | 1835.908(2625.598) |
| 4,5,7-Trihydroxyisoflavone | 507.397(2564.172) | 124.958(731.854) | 561.595(2843.781) | 153.911(841.29) |
| 4-Acetamidobutyric acid | 3471.712(2342.69) | 3727.521(2854.822) | 3615.048(2473.068) | 3939.109(3184.595) |
| 4-Aminomethylcyclohexanecarboxylic acid; | 1589.572(3239.52) | 1326.667(747.566) | 1401.408(1985.592) | 1323.682(729.624) |

**Supplementary Table S1.** Sample mean and standard deviations of metabolites in ME/CFS patients, controls, and their subgroups.

| Metabolite | ME/CFS (n=106) | Control (n=91) | Female ME/CFS (n=75) | Female Control (n=69) |
|---|---|---|---|---|
| | mean (SD) | mean (SD) | mean (SD) | mean (SD) |
| 4-Fluoro-.alpha.-pyrrolidinobutiophenone | 532.638(610.097) | 458.298(230.627) | 554.359(649.079) | 459.133(227.638) |
| 4'-Methyl-N-methylhexanophenone | 2337.883(2599.719) | 2483.552(2368.085) | 2417.654(2633.99) | 2642.039(2537.353) |
| 4-Pyridoxic acid; | 9506.692(32185.74) | 3611.761(6927.589) | 10060.296(36968.279) | 3804.289(7433.485) |
| 5'-S-Methyl-5'-thioadenosine | 2295.417(2441.494) | 2541.641(1775.543) | 2469.835(2761.965) | 2655.655(1832.667) |
| 5'-S-Methylthioadenosine | 2232.692(2448.886) | 2396.981(1900.293) | 2370.237(2777.789) | 2481.076(1983.511) |
| 6-Hydroxyflavone | 1191.845(6678.29) | 403.876(528.261) | 1481.81(7954.915) | 353.353(409.453) |
| 6-Methoxynaphthaleneacetic acid | 7862.951(2449.945) | 9327.172(12784.412) | 8185.531(2515.823) | 8105.686(2065.758) |
| 7.alpha.-Hydroxy-3-oxo-4-cholestenoic acid | 7832.128(4085.847) | 7971.016(3525.274) | 7988.535(4451.445) | 7856.561(3289.14) |
| 7-Hydroxywarfarin | 256.276(1326.793) | 125.152(55.26) | 316.435(1586.026) | 120.883(52.118) |
| 7-Methylguanosine | 2894.162(982.806) | 3126.746(1088.991) | 2980.301(1011.499) | 3171.08(1170.041) |
| Acetazolamide | 111.881(1032.576) | 10.68(5.05) | 10.574(6.06) | 10.878(4.95) |
| Acetominophen | 8598.401(25412.26) | 1888.819(3095.431) | 8664.723(27961.761) | 1954.635(3481.327) |
| Acetyl-DL-carnitine | 4040.595(1781.273) | 3886.498(1192.6) | 4161.284(1862.636) | 3932.342(1212.884) |
| Acyclovir | 10042.642(38991.986) | 102.244(109.125) | 13378.101(45628.802) | 99.14(108.798) |
| Adenosine | 476.693(252.498) | 554.001(724.574) | 477.541(212.691) | 611.275(821.588) |
| Ala-Ile | 1117.937(531.714) | 1022.708(379.234) | 1153.66(538.781) | 1022.593(369.487) |
| Albendazole | 1404.595(9285.314) | 342.564(133.753) | 1876.415(11095.377) | 355.058(141.429) |
| Albendazole sulfoxide | 556.474(1732.254) | 706.284(5840.411) | 658.421(2024.354) | 911.792(6718.083) |
| alpha-Methylhistidine; | 18823.314(15559.922) | 19428.048(15625.399) | 18017.263(14601.37) | 20347.68(16173.525) |
| Alprazolam | 219.224(700.569) | 79.36(42.975) | 209.565(755.994) | 76.592(43.888) |
| Aminodiphenylmethane | 10035.44(39641.241) | 4831.419(12787.502) | 7128.186(14760.998) | 5206.398(14422.701) |
| Androstan-3-ol-17-one 3-glucuronide | 1018.393(816.112) | 997.46(700.178) | 954.271(739.508) | 898.382(668.913) |
| Arginine | 66971.722(30497.565) | 63686.741(31676.786) | 70541.746(33026.077) | 66753.473(27292.861) |
| Atenolol | 2831.445(26482.196) | 235.929(152.586) | 228.711(127.871) | 242.031(156.639) |
| Atorvastatin | 310.188(170.392) | 307.876(96.997) | 329.376(192.18) | 309.451(103.349) |
| Avobenzone | 1620.013(901.713) | 2204.034(6233.272) | 1625.707(809.631) | 1517.537(583.705) |
| Benthiavalicarb-isopropyl | 397.898(475.323) | 338.666(425.872) | 387.129(452.525) | 343.498(481.04) |
| Benzophenone-3 | 79.42(118.399) | 152.05(480.887) | 94.023(138.386) | 176.503(548.803) |
| Betaine | 1396162.001(400978.201) | 1465520.587(366608.787) | 1356785.522(423707.652) | 1417694.595(341114.75) |
| Betaine aldehyde cation | 14682.235(107622.215) | 56.189(19.083) | 20851.707(128492.149) | 56.738(19.839) |
| Betonicine | 5018.889(9595.989) | 5425.561(9356.727) | 4242.1(6533.329) | 6202.06(10533.221) |
| Biliverden | 37882.387(29682.943) | 38990.463(24851.237) | 35273.646(28537.043) | 35664.215(20507.162) |
| Borrelidin | 1800.784(4109.912) | 1698.556(4228.984) | 1876.809(4290.433) | 1452.819(3956.247) |
| Bradykinin | 2350.694(4905.075) | 2102.181(4523.581) | 2055.799(4093.04) | 2277.815(4790.793) |
| Caffeine | 46358.68(97388.926) | 86219.943(115602.167) | 41207.184(95909.44) | 92115.413(118321.704) |
| Carbamazepine | 424.616(1432.071) | 11883.366(108750.28) | 375.298(1107.091) | 15626.683(125106.825) |
| Carnitine | 1001164.333(283021.215) | 1045982.351(310270.412) | 970340.809(286347.389) | 1001183.088(275670.844) |
| Choline cation | 254611.526(57164.648) | 282373.342(63913.774) | 256045.629(58199.308) | 280565.281(62246.483) |
| cis-10,11-Dihydroxy-10,11-dihydrocarbamazepine | 525.109(153.075) | 959.717(3818.829) | 547.058(164.975) | 1094.893(4391.718) |
| Citrulline | 19901.677(6196.281) | 21504.169(6882.343) | 20450.559(6314.125) | 22463.956(7168.386) |
| Codeine-6-.beta.-D-glucuronide | 774.993(5522.859) | 232.532(91.583) | 1020.604(6604.132) | 235.304(100.647) |
| Coniferylaldehyde | 765.911(462.54) | 787.668(824.323) | 768.803(350.473) | 835.32(940.869) |
| Cotinine N-.beta.-D-glucuronide | 76.449(283.47) | 46.197(159.887) | 57.743(224.767) | 31.424(55.891) |
| Creatine | 217981.51(118303.317) | 203650.285(97913.88) | 239445.001(122895.866) | 224296.739(95276.546) |
| Creatinine | 1006729.671(287698.034) | 1029213.304(254200.686) | 968014.1(297487.798) | 1005188.696(241774.03) |
| Cyclo(Leu-Pro) | 2228.564(1706.261) | 2027.347(1104.514) | 2361.372(1874.821) | 2002.407(1032.619) |
| D-.alpha.-Cyclohexylglycine | 6326.782(6206.867) | 8327.262(8211.34) | 6963.999(6970.157) | 8208.176(8697.467) |
| Decanoyl-L-carnitine | 42146.5(48589.406) | 42071.952(30458.853) | 41378.73(53905.48) | 44599.02(33313.09) |

**Supplementary Table S1.** Sample mean and standard deviations of metabolites in ME/CFS patients, controls, and their subgroups.

| Metabolite | ME/CFS (n=106) | Control (n=91) | Female ME/CFS (n=75) | Female Control (n=69) |
|---|---|---|---|---|
| | mean (SD) | mean (SD) | mean (SD) | mean (SD) |
| D-erythro-Sphingosine-1-phosphate | 7295.174(3378.972) | 7543.128(3687.008) | 7215.842(3142.458) | 8064.372(3531.397) |
| Dexpanthenol | 1546.048(2341.347) | 2108.653(3911.294) | 1339.362(633.825) | 2386.132(4460.192) |
| D-Fructose | 34526.336(8214.427) | 34866.41(9180.115) | 35258.4(7984.551) | 35466.161(9360.386) |
| Diazepam | 370.721(2360.704) | 46.476(76.296) | 354.102(2509.34) | 48.869(87.424) |
| Dinor-12-oxophytodienoic acid | 525.23(862.885) | 416.42(194.711) | 602.168(1019.314) | 425.507(195.769) |
| Diphenhydramine | 2585.349(6127.955) | 3178.366(14715.458) | 2336.057(3983.41) | 3707.339(16925.362) |
| DL-Indole-3-lactic acid | 37199.448(18913.594) | 35604.136(12065.446) | 37363.886(20753.868) | 35327.001(12981.249) |
| D-Pyroglutamic acid | 121612.569(22366.016) | 123013.056(24064.978) | 123418.191(24064.122) | 125690.461(24817.147) |
| D-Turanose | 525.908(351.452) | 631.074(581.896) | 520.138(265.379) | 609.634(488.419) |
| Ergothioneine | 767.24(1494.771) | 719.663(431.749) | 641.187(444.426) | 742.568(450.052) |
| Esomeprazole | 2660.51(10721.074) | 1307.945(4540.326) | 3400.906(12758.474) | 1477.047(5219.404) |
| Ethiolat | 11862.296(20760.107) | 9779.989(18228.881) | 11694.874(22626.1) | 10043.125(20657.08) |
| Ethyldiethanolamine | 734.979(766.768) | 727.196(595.857) | 782.081(892.314) | 784.328(667.115) |
| Ethylenediaminetetraacetic acid | 663091.766(891136.229) | 496339.244(788805.165) | 707188.787(939588.255) | 525798.856(819400.236) |
| Ezetimibe | 223.611(238.157) | 234.24(306.197) | 237.685(276.619) | 238.293(346.126) |
| Fexofenadine | 450.355(238.197) | 841.091(3125.726) | 445.615(146.548) | 979.139(3591.011) |
| Gabapentin | 62080.105(201779.015) | 5287.117(32734.04) | 65249.893(197458.731) | 1736.245(1006.505) |
| Glaucine | 860.63(2849.169) | 574.959(169.559) | 1011.561(3402.675) | 575.815(174.676) |
| Glutamic acid | 7933.975(3124.287) | 7180.498(3426.266) | 7685.959(2912.252) | 6790.446(2759.596) |
| Glutamine | 101505.785(19190.477) | 102907.791(20574.24) | 103011.688(20506.373) | 105238.609(21074.201) |
| Glycocholic acid | 4874.144(5532.195) | 3707.728(3340.534) | 4835.094(5666.975) | 3839.337(3220.547) |
| Glycodeoxycholic acid | 16574.334(17400.385) | 14780.84(19064.118) | 15665.2(16876.362) | 14124.624(16898.648) |
| Gly-Pro-Arg | 586.043(2198.483) | 428.808(1170.147) | 394.694(2029.309) | 440.56(1188.759) |
| Guanidine | 5493.87(8781.142) | 4475.798(4190.121) | 6162.546(10314.276) | 4770.688(4754.17) |
| Guanine | 6226.314(23441.739) | 279.003(1082.414) | 8246.388(27387.081) | 169.027(155.108) |
| Heptadecasphing-4-enine | 5557.437(4651.009) | 5538.189(4468.342) | 5527.139(4907.465) | 5161.482(4445.015) |
| Hexanoyl-L-carnitine | 8996.499(13556.537) | 8356.705(5002.169) | 9402.717(15859.832) | 8656.44(5504.128) |
| H-gamma-glutamyl-glutamine | 2339.192(1792.988) | 2006.339(1305.502) | 2544.089(1995.762) | 2084.667(1260.692) |
| Histidine | 49555.943(23320.509) | 53178.919(21745.875) | 49244.588(24505.38) | 54934.704(20736.674) |
| Homoarginine; | 3381.815(2097.316) | 3026.593(2160.863) | 3437.227(2173.842) | 3116.108(1905.262) |
| H-Pro-Hyp-OH | 2228.15(1753.581) | 1756.456(871.446) | 2360.853(1986.558) | 1739.039(885.645) |
| Hydroxybupropion | 884.813(2016.341) | 1883.832(8028.514) | 966.47(2225.998) | 1326.55(4137.981) |
| Hypoxanthine | 31281.328(19491.261) | 30486.398(22288.554) | 31017.305(20197.579) | 28804.254(15185.823) |
| Ile-Glu-Arg | 365.728(1522.667) | 320.225(998.515) | 286.888(1615.263) | 371.056(1124.937) |
| Indole-3-propionic acid | 1220.413(1004.075) | 1438.512(994.93) | 1294.439(896.394) | 1450.489(959.941) |
| Irbesartan | 34111.466(330125.328) | 1632.597(1008.364) | 48237.142(394834.653) | 1665.958(1152.085) |
| Isoleucine | 15404.33(5244.539) | 15607.18(5393.939) | 14827.325(5014.375) | 15178.898(5537.463) |
| Isopentenyladenine | 1014642.651(399845.125) | 1046922.522(370983.691) | 1031908.219(421699.096) | 1056127.36(345968.422) |
| Isopropylamine | 4749.851(1414.044) | 5211.39(1333.561) | 4813.12(1502.012) | 5179.224(1250.988) |
| Kynurenine | 2125.376(754.54) | 2313.805(749.317) | 2055.529(729.308) | 2285.971(712.949) |
| Lamotrigine; | 2244.641(22565.331) | 18.464(16.389) | 3201.952(26989.449) | 18.54(17.344) |
| Lansoprazole | 736.475(4048.63) | 2665.031(13708.313) | 987.707(4814.561) | 2829.094(14792.141) |
| Lauric acid diethanolamide | 413.111(567.669) | 345.335(216.443) | 448.33(666.531) | 327.439(204.242) |
| Lauroyl-L-carnitine | 5784.303(4911.391) | 6763.368(3908.375) | 5731.635(5283.284) | 6696.194(3998.479) |
| L-Carnitine | 16079.975(4595.436) | 15890.768(4307.224) | 15793.384(4678.999) | 15559.285(4194.778) |
| L-Citrulline | 20161.291(5985.134) | 21758.529(6402.073) | 20674.288(6096.622) | 22617.585(6585.22) |
| L-Cysteine-glutathione disulfide | 1116.696(1162.972) | 1224.484(1444.167) | 1216.194(1224.408) | 1353.062(1496.972) |
| L-Cystine | 2137.312(1969.911) | 2271.747(2210.344) | 2310.258(1847.501) | 2449.457(2214.124) |

**Supplementary Table S1.** Sample mean and standard deviations of metabolites in ME/CFS patients, controls, and their subgroups.

| Metabolite | ME/CFS (n=106) | Control (n=91) | Female ME/CFS (n=75) | Female Control (n=69) |
|---|---|---|---|---|
| | mean (SD) | mean (SD) | mean (SD) | mean (SD) |
| Levocetirizine; | 9007.024(38087.67) | 5404.405(22513.101) | 9755.004(42513.045) | 6447.278(25353.913) |
| Linoleoylcarnitine | 6028.057(4694.504) | 6937.757(5240.418) | 5890.895(4970.782) | 7324.182(4782.704) |
| L-Leucine, methyl ester | 936.172(1279.128) | 734.385(675.979) | 844.02(959.174) | 727.712(723.551) |
| Losartan | 1124.145(6579.061) | 2289.546(19221.441) | 255.822(125.143) | 2942.867(22113.406) |
| L-Threonine | 6399.383(2197.795) | 6434.59(1928.87) | 6569.693(2432.17) | 6582.749(1953.44) |
| L-Tyrosine | 3214.55(1382.508) | 3345.219(1261.579) | 3191.419(1464.291) | 3283.667(1355.218) |
| Lysine | 24048.561(14481.217) | 23142.646(14133.037) | 26085.381(16010.536) | 24289.949(13450.24) |
| Matrine | 2325.219(12579.91) | 500.431(207.181) | 3144.599(15002.584) | 513.737(206.412) |
| Mefenorex | 622.656(716.718) | 3807.946(26441.59) | 634.16(803.491) | 668.885(826.358) |
| Meloxicam | 28306.258(195269.072) | 12851.881(121675.611) | 17836.666(151075.883) | 17003.109(139981.187) |
| Meprobamate | 1573.996(3579.443) | 932.297(1488.818) | 1271.328(2301.477) | 921.028(1674.405) |
| Metformin | 7135.099(48045.124) | 3475.203(23828.338) | 9863.786(57367.973) | 4323.549(27408.303) |
| Methacholine cation | 56501.078(33409.327) | 67485.057(42836.21) | 59647.284(34908.691) | 64433.74(23560.197) |
| Methionine | 6161.544(1634.068) | 6223.87(1770.355) | 6036.444(1672.884) | 6246.537(1851.937) |
| Methioninesulfoxide | 753.827(372.4) | 693.081(379.811) | 768.805(401.695) | 731.508(415.592) |
| Methylgallate | 139.383(458.151) | 67.958(251.269) | 159.529(526.421) | 72.307(280.953) |
| Metoprolol | 897.413(4862.805) | 234.563(653.184) | 1006.345(5713.033) | 260.193(750.546) |
| Metoprolol acid | 7786.566(33910.371) | 535.074(2767.371) | 5313.31(28575.468) | 635.6(3182.653) |
| Milnacipran | 276.882(1331.735) | 148.067(53.863) | 339.641(1591.763) | 151.333(53.12) |
| Modafinil | 1075.48(6709.26) | 71.168(24.162) | 856.222(5843.393) | 70.136(24.567) |
| Modafinil acid | 131.611(721.079) | 24.78(13.35) | 89.453(410.063) | 23.701(13.311) |
| Montelukast-1,2-diol | 75.941(301.129) | 84.645(479.266) | 68.948(274.135) | 98.954(550.468) |
| Moxonidine | 3500.133(2726.862) | 3040.75(1987.891) | 3494.77(2782.516) | 3129.874(1890.894) |
| N-(3-(Aminomethyl)benzyl)acetamidine | 20921.024(28034.826) | 18034.449(15522.733) | 21426.169(26692.226) | 18342.407(16707.634) |
| N-.alpha.-Acetyl-L-arginine | 3082.187(1181.981) | 3213.461(1045.445) | 3045.215(1184.893) | 3232.865(1091.37) |
| N.alpha.-Methyl-L-lysine | 29306.073(114182.748) | 13134.602(36620.029) | 27616.816(101350.581) | 9580.597(6849.304) |
| N.epsilon.-Acetyl-L-lysine | 2453.841(2107.986) | 2280.92(1269.094) | 2602.365(2447.778) | 2358.717(1430.008) |
| N.epsilon.-Methyl-L-lysine | 8975.039(6927.428) | 8793.019(6711.352) | 9268.888(7015.526) | 9284.578(7186.447) |
| N8-Acetylspermidine | 3001.558(1211.178) | 2900.086(896.957) | 2881.329(1046.705) | 2851.314(833.99) |
| N-Acetylalanine | 707.391(950.515) | 582.675(496.092) | 792.246(1091.701) | 619.1(511.521) |
| N-Acetyl-D-norleucine | 3757.634(3749.095) | 3860.092(3382.148) | 4143.693(3978.725) | 4142.792(3707.786) |
| N-Acetylhistidine | 1008.892(648.298) | 1039.722(610.487) | 1029.937(698.38) | 1051.991(674.644) |
| N-Acetyl-L-carnosine | 2954.051(1639.621) | 2926.55(1628.741) | 2355.911(950.184) | 2451.763(1167.01) |
| Naproxen | 3476.065(13446.579) | 4450.949(22153.769) | 2032.557(7784.078) | 5543.465(25430.423) |
| NEPSILON,NEPSILON,NEPSILON-TRIMETHYLLYSINE | 13358.236(4821.32) | 15219.514(11953.152) | 12581.416(4428.925) | 13549.071(6450.745) |
| N-Methylhistidine | 35730.75(37645.645) | 33537.29(29498.297) | 30230.148(33903.282) | 32150.208(28181.793) |
| N-Methylproline | 11656.687(9296.226) | 11510.001(9904.91) | 11753.456(9333.458) | 11857.717(10588.637) |
| Norleucine | 15444.661(6060.568) | 15678.34(4128.345) | 14637.073(4788.935) | 15362.688(4038.533) |
| Nudifloramide | 64111.911(56284.553) | 63703.328(37012.76) | 58887.733(33435.743) | 63376.868(34799.728) |
| Octanoylcarnitine | 46353.596(55972.637) | 47100.775(29066.033) | 47144.841(64211.055) | 49577.747(31693.746) |
| Oleoyl-L-carnitine | 10823.414(8463.672) | 11676.244(8434.192) | 10729.659(8590.403) | 12310.139(8440.117) |
| Omeprazole sulfone | 4929.053(23929.418) | 1843.877(13957.834) | 6301.035(28062.735) | 2096.193(15907.146) |
| Omeprazole sulfone N-oxide | 1165.746(5101.882) | 421.937(2441.309) | 1400.94(5908.665) | 450.787(2744.995) |
| Ondansetron | 365.61(1271.909) | 240.115(62.33) | 430.734(1519.455) | 243.04(68.123) |
| Ornithine | 3959.933(2287.816) | 4022.624(2665.327) | 4128.265(2438.596) | 4207.488(2640.386) |
| p-Acetamidophenyl .beta.-D-glucuronide | 766.028(3177.057) | 171.088(73.211) | 854.182(3679.025) | 171.801(71.797) |
| Palmitamide | 3270.611(17777.123) | 1962.253(8761.313) | 4316.593(21213.234) | 2315.319(10069.12) |
| Pantoprazole | 898.088(8152.763) | 16270.845(118825.791) | 1249.917(9750.392) | 15650.814(128427.894) |

**Supplementary Table S1.** Sample mean and standard deviations of metabolites in ME/CFS patients, controls, and their subgroups.

| Metabolite | ME/CFS (n=106) | Control (n=91) | Female ME/CFS (n=75) | Female Control (n=69) |
|---|---|---|---|---|
| | mean (SD) | mean (SD) | mean (SD) | mean (SD) |
| Pantothenic acid | 10030.786(7968.504) | 8465.694(5284.919) | 10126.233(8359.344) | 8677.218(5145.136) |
| Penciclovir | 615.263(2184.586) | 280.151(80.439) | 747.499(2600.119) | 284.949(82.361) |
| Phenylacetylglutamine | 31218.152(22442.043) | 34681.21(38768.962) | 32853.447(24164.403) | 36131.129(43000.834) |
| Phenylacetyl-L-glutamine | 14884.784(10785.176) | 16607.075(17719.928) | 15603.194(11631.592) | 17446.503(19744.39) |
| Phenylalanine | 55973.85(16422.42) | 58120.533(13302.726) | 54542.869(15990.617) | 58589.269(14424.404) |
| Pipecolic acid | 425.09(586.911) | 426.157(508.716) | 422.322(411.915) | 466.253(576.586) |
| Piperine | 5718.294(8337.056) | 7720.583(10963.746) | 5437.646(7527.174) | 6841.21(7360.472) |
| Prazepam | 288.862(1542.242) | 137.608(93.414) | 357.083(1843.726) | 135.971(95.239) |
| Proline | 14486.098(6367.337) | 14482.522(5997.29) | 14411.268(7109.856) | 13721.924(5754.6) |
| Propionylcarnitine | 68344.79(23209.488) | 71041.55(26326.498) | 65475.827(22787.033) | 65045.673(20218.858) |
| Pyrantel | 372.256(111.716) | 1129.226(7044.272) | 384.923(119.14) | 1372.434(8103.556) |
| Pyridoxal | 1642.179(3991.664) | 946.29(438.171) | 1926.346(4738.724) | 966.128(487.166) |
| Pyridoxine; | 1022.88(4388.483) | 435.057(382.972) | 1291.631(5232.132) | 402.065(337.933) |
| Quetiapine | 369.86(1339.4) | 222.872(66.167) | 255.71(234.5) | 224.248(71.778) |
| Quetiapine sulfoxide | 650.477(4802.113) | 136.563(42.808) | 205.765(560.74) | 136.638(46.732) |
| R-(-)-O-Desmethylvenlafaxine | 592.445(3164.213) | 410.727(1880.497) | 790.269(3775.065) | 263.649(936.258) |
| rac-4-Sulfoxypropranolol | 202.948(514.821) | 151.335(57.243) | 162.535(82.087) | 153.516(60.927) |
| Ranitidine | 2507.731(13468.136) | 205.008(53.061) | 2017.312(11803.676) | 206.538(53.841) |
| Ranitidine N-oxide | 713.593(4219.564) | 62.623(17.85) | 390.958(1801.266) | 62.105(18.285) |
| Ranitidine-S-oxide | 213.634(515.841) | 132.212(90.781) | 145.129(175.128) | 133.881(98.329) |
| Scopoletin | 591.68(2509.742) | 178.452(414.272) | 688.798(2864.176) | 192.277(467.346) |
| SDMA | 9500.368(2703.971) | 10370.308(2765.049) | 9808.401(2760.319) | 10599.213(2673.464) |
| Serotonin | 18189.163(41222.489) | 14668.474(23684.703) | 14353.675(19998.499) | 13363.044(17156.067) |
| Ser-Tyr-Lys | 193.809(988.093) | 91.613(175.965) | 184.445(1115.822) | 104.226(200.316) |
| Stachydrine | 563959.448(691819.57) | 612164.26(685817.817) | 587518.508(706098.216) | 653922.988(726897.281) |
| Sulfamethoxazole | 41.919(48.984) | 4622.278(43440.039) | 41.835(43.623) | 6101.074(49975.795) |
| Tapentadol-.beta.-D-glucuronide | 221.214(1394.74) | 127.85(761.689) | 41.736(24.102) | 147.58(875.506) |
| Tauroursodeoxycholic acid | 2706.846(6111.732) | 1545.619(2380.983) | 2678.948(5486.303) | 1274.586(1130.138) |
| Telmisartan | 4699.894(41335.897) | 3856.6(30518.071) | 6481.819(49436.552) | 4898.127(35109.147) |
| Temazepam | 279.199(1061.022) | 153.782(115.023) | 342.847(1265.834) | 146.924(65.02) |
| Testosterone | 3622.17(4140.278) | 4445.438(4912.789) | 3663.498(4396.175) | 4590.956(5246.58) |
| Theanine; | 1819.454(2742.739) | 1366.781(1424.619) | 2138.05(3146.371) | 1493.761(1527.77) |
| Theobromine | 43840.022(50239.28) | 50274.237(39008.573) | 42746.307(46884.552) | 53954.352(39621.193) |
| threo-Dihydrobupropion | 959.671(1263.114) | 899.034(1318.122) | 970.564(1208.202) | 898.166(1350.257) |
| Thr-Ile-Arg | 334.018(1580.162) | 174.397(391.703) | 299.942(1746.529) | 188.59(438.92) |
| Ticlopidine | 228.217(1202.183) | 52.712(180.862) | 277.805(1397.262) | 45.125(168.628) |
| Topiramate | 883.512(3767.87) | 360.884(470.487) | 1085.88(4482.725) | 366.228(537.766) |
| Toradol | 356.538(2066.064) | 150.603(104.488) | 455.463(2469.352) | 148.593(102.035) |
| trans-3'-Hydroxycotinine | 2268.615(10601.908) | 1686.299(5203.465) | 1233.236(3119.976) | 1359.612(4030.731) |
| Trazodone | 2672.469(13819.619) | 206.455(55.025) | 3739.443(16447.77) | 208.643(57.472) |
| Tri-2-ethylhexyl trimellitate | 6954.814(58249.323) | 2150.885(4550.116) | 1221.862(1696.585) | 2565.286(5161.634) |
| Trigonelline | 62593.319(128404.523) | 87574.388(113723.624) | 66818.372(143147.881) | 94052.01(122519.557) |
| Trileptal | 378.242(124.735) | 1737.665(13037.79) | 392.867(128.704) | 2189.462(14998.322) |
| Trimethoprim | 234.855(71.982) | 2512.23(21564.074) | 240.288(76.487) | 3245.027(24808.634) |
| Trimethylamine-N-oxide | 22496.762(34083.237) | 15784.326(13954.105) | 25028.914(38657.129) | 14414.784(10594.282) |
| Triptolide | 572.071(359.723) | 709.428(1341.517) | 608.313(410.976) | 757.077(1539.512) |
| Tryptophan | 15719.222(4762.968) | 15591.457(4927.165) | 15508.626(4851.561) | 15048.978(5005.799) |
| Tyrosine | 6484.429(2346.848) | 6649.41(2125.156) | 6372.675(2355.791) | 6620.619(2272.411) |

**Supplementary Table S1.** Sample mean and standard deviations of metabolites in ME/CFS patients, controls, and their subgroups.

| Metabolite | ME/CFS (n=106) | Control (n=91) | Female ME/CFS (n=75) | Female Control (n=69) |
|---|---|---|---|---|
| | mean (SD) | mean (SD) | mean (SD) | mean (SD) |
| Urea | 128763.489(48161.656) | 135736.88(41188.088) | 128773.302(50486.088) | 132146.779(38578.177) |
| Urocanic acid; | 20103.21(25401.881) | 20113.659(16158.372) | 21750.5(29758.063) | 21866.059(18012.708) |
| Usnic acid | 6421.074(38109.236) | 5909.508(26905.817) | 2771.907(1725.232) | 6767.755(30846.452) |
| **Complex Lipids (CL)** | | | | |
| AC (10:0) | 14300.667(14495.722) | 14592.903(12632.497) | 14299.18(15986.705) | 15304.621(13912.854) |
| AC (10:1) | 7852.356(5661.808) | 8700.584(5463.897) | 7704.051(6028.18) | 8895.645(5924.822) |
| AC (12:0) | 6801.104(4854.843) | 7472.708(4034.354) | 6652.962(5029.517) | 7433.406(3978.211) |
| AC (12:1) | 5342.259(2975.805) | 5782.079(2495.997) | 5250.448(3005.47) | 5779.664(2466.414) |
| AC (14:1) | 7653.305(5643.002) | 7865.258(4525.933) | 7670.8(5957.272) | 7876.844(4540.047) |
| AC (14:2) | 4537.778(3261.62) | 5105.633(2880.79) | 4533.971(3556.58) | 5111.986(2806.317) |
| AC (16:0) | 10907.214(3516.947) | 10958.197(3206.646) | 10240.456(3053.947) | 10630.098(2958.93) |
| AC (18:0) | 4409.46(1523.433) | 4658.607(1398.628) | 4241.255(1498.351) | 4527.743(1288.803) |
| AC (18:1) | 15169.411(6563.264) | 14716.482(4589.592) | 14552.69(6291.98) | 14539.384(4595.967) |
| AC (18:2) | 9086.774(4258.756) | 9507.785(3723.066) | 8473.128(4008.707) | 9215.757(3414.944) |
| AC (8:0) | 5424.664(7636.621) | 5324.659(5489.183) | 5578.054(8810.926) | 5661.698(6126.403) |
| CE (14:0) | 483.105(328.13) | 550.26(299.965) | 517.754(354.457) | 566.25(294.07) |
| CE (16:0) | 6284.196(5322.107) | 5971.764(4027.806) | 6640.605(5705.632) | 6052.502(3655.343) |
| CE (16:1) | 44723.031(22333.233) | 45240.669(19130.581) | 46466.245(21911.29) | 44961.974(18550.491) |
| CE (18:0) | 3286.85(1793.581) | 3220.682(1502.875) | 3248.724(1768.19) | 3255.714(1566.484) |
| CE (18:1) | 32176.362(13035.556) | 33019.476(10977.599) | 33329.958(13182.182) | 33803.223(10306.817) |
| CE (18:2) | 360370.43(102450.354) | 395504.729(97524.484) | 365905.916(104757.775) | 408420.872(96560.058) |
| CE (18:3) | 13226.28(5296.356) | 16158.621(6188.509) | 13538.692(5308.083) | 16026.716(5476.892) |
| CE (20:2) | 7353.518(4456.574) | 8560.232(3729.559) | 7480.067(4723.572) | 8876.919(3620.926) |
| CE (20:3) | 65342.381(23690.393) | 72249.761(24364.326) | 66916.365(23589.777) | 74083.382(23946.671) |
| CE (20:4) | 760763.777(278873.716) | 769431.808(251914.063) | 774727.643(270809.091) | 762789.124(215866.426) |
| CE (20:5) | 17407.549(19917.453) | 12423.777(8256.741) | 19104.609(22573.844) | 12828.237(8956.276) |
| CE (22:2) | 686.936(506.637) | 714.698(520.067) | 676.49(502.562) | 745.29(543.744) |
| CE (22:6) | 132582.468(60465.938) | 114384.853(39714.847) | 139199.071(65068.192) | 118510.435(39055.205) |
| Ceramide (d34:1) - ESI(+) | 7991.308(2279.1) | 7701.808(1766.753) | 7687.199(2250.215) | 7834.529(1868.036) |
| Ceramide (d36:1) - ESI(+) | 5055.887(1639.5) | 4781.199(1246.336) | 4945.072(1449.706) | 4913.929(1227.46) |
| Ceramide (d38:1) - ESI(+) | 6696.998(1780.221) | 6981.944(1458.36) | 6664.704(1627.166) | 7112.106(1389.354) |
| Ceramide (d40:1) | 21686.844(5740.739) | 23489.285(4735.334) | 21422.451(5095.414) | 23401.997(4615.503) |
| Ceramide (d41:1) - ESI (+) | 6918.214(1876.08) | 7517.217(1659.079) | 6992.447(1758.751) | 7645.828(1704.017) |
| Ceramide (d42:1) - ESI (+) | 71508.239(18019.881) | 78197.952(14457.978) | 70141.421(16527.622) | 77355.897(13980.101) |
| Ceramide (d42:2) A - ESI (+) | 19273.129(4975.246) | 18114.439(3325.542) | 18658.838(4645.216) | 18167.611(3294.727) |
| Ceramide (d42:2) B - ESI (+) | 14981.161(4149.123) | 17174.124(4245.274) | 15071.338(4164.235) | 17288.064(4089.868) |
| Ceramide (d32:1) | 3595.683(1323.517) | 3755.63(977.806) | 3473.329(1135.936) | 3749.235(876.773) |
| Ceramide (d33:1) | 2916.634(935.773) | 3016.705(807.728) | 2905.891(954.228) | 3011.126(830.753) |
| Ceramide (d34:0) | 4886.997(2053.157) | 4687(1716.579) | 4705.485(1904.271) | 4578.351(1702.528) |
| Ceramide (d34:1) - ESI(-) | 18074.154(4880.408) | 17694.653(3936.635) | 16797.099(3886.53) | 17358.862(3879.255) |
| Ceramide (d34:2) | 3094.181(847.888) | 3030.248(713.119) | 3059.406(823.881) | 3103.748(732.199) |
| Ceramide (d36:1) - ESI(-) | 1637.345(721.82) | 1539.002(608.946) | 1547.936(631.269) | 1516.191(611.004) |
| Ceramide (d38:1) - ESI(-) | 11993.384(4407.452) | 12156.053(3738.061) | 11661.638(4066.525) | 12037.971(3600.075) |
| Ceramide (d39:1) | 6299.483(2739.989) | 6979.275(2440.541) | 6280.654(2642.502) | 6928.753(2492.845) |
| Ceramide (d40:0) | 1765.181(1227.484) | 1564.678(901.085) | 1775.733(1264.199) | 1479.739(772.332) |
| Ceramide (d40:2) | 6057.125(4179.286) | 6235.898(4315.18) | 5995.39(4515.358) | 6101.231(4353.501) |
| Ceramide (d41:1) - ESI(-) | 42041.811(15059.383) | 46380.145(13427.707) | 41060.244(14002.301) | 45628.819(13367.535) |
| Ceramide (d42:0) | 11948.609(7113.944) | 11294.104(4976.297) | 11987.369(7448.888) | 10658.27(4389.341) |

**Supplementary Table S1.** Sample mean and standard deviations of metabolites in ME/CFS patients, controls, and their subgroups.

| Metabolite | ME/CFS (n=106) | Control (n=91) | Female ME/CFS (n=75) | Female Control (n=69) |
|---|---|---|---|---|
| | mean (SD) | mean (SD) | mean (SD) | mean (SD) |
| Ceramide (d42:1) - ESI(-) | 136674.648(44023.767) | 149770.415(39576.94) | 131275.247(38296.961) | 146093.73(38714.63) |
| Ceramide (d42:2) A - ESI(-) | 49252.166(17827.101) | 47220.491(13180.04) | 45881.62(14014.478) | 45775.601(12303.101) |
| Ceramide (d42:2) B - ESI(-) | 37317.754(16468.604) | 35789.863(11255.565) | 34603.652(14003.87) | 34888.751(10464.998) |
| Ceramide (d43:1) | 2528.23(1370.14) | 2789.752(1196.521) | 2524.053(1366.934) | 2658.272(1055.678) |
| Ceramide (d44:1) | 846.904(541.153) | 822.512(399.283) | 766.114(383.948) | 766.223(335.297) |
| Cholesterol | 266952.992(63543.111) | 283483.733(67507.559) | 259126.592(62668.048) | 282785.483(70992.096) |
| DG (32:0) | 2029.32(748.253) | 1982.392(914.732) | 1876.907(663.523) | 1834.096(743.093) |
| DG (32:1) | 5251.897(3457.449) | 4947.61(2880.584) | 4839.272(3261.309) | 4417.438(2663.105) |
| DG (34:1) | 1041.038(494.395) | 999.382(487.017) | 957.669(477.567) | 926.246(447.834) |
| DG (34:2) | 32528.392(13807.47) | 30539.842(13070.979) | 29395.286(12112.58) | 28013.19(12155.434) |
| DG (34:3) | 5185.782(3045.124) | 4572.847(2077.17) | 4934.302(2970.925) | 4487.862(2240.726) |
| DG (36:1) | 6350.769(3167.505) | 6121.406(3281.803) | 5774.621(2635.876) | 5434.807(2715.326) |
| DG (36:2) | 55205.662(20357.43) | 49677.788(17704.742) | 52703.881(20071.649) | 46096.408(15020.654) |
| DG (36:3) | 70925.903(27194.121) | 65340.022(22354.965) | 66885.539(26880.219) | 62128.554(20637.751) |
| DG (36:4) A | 16838.454(8807.583) | 16004.561(7408.839) | 15646.998(8185.164) | 15562.018(6792.317) |
| DG (36:4) B | 4169.876(3637.695) | 3516.148(2613.304) | 3898.521(3366.237) | 3094.96(2394.278) |
| DG (36:5) | 2206.156(1211.632) | 2086.253(902.788) | 2013.999(1107.932) | 2061.556(899.951) |
| DG (38:5) | 10566.304(4064.74) | 10089.518(3680.913) | 9992.912(3910.752) | 9028.535(2964.831) |
| DG (38:6) | 6169.284(3104.712) | 5407.065(2535.955) | 5872.225(2936.044) | 4956.003(2363.195) |
| FA (10:0) (capric acid) | 1604.441(1050.232) | 1407.005(1073.102) | 1560.414(987.235) | 1389.12(1089.533) |
| FA (12:0) (lauric acid) | 8080.776(5386.504) | 8269.26(5625.99) | 7817.814(5729.118) | 8256.763(6226.421) |
| FA (14:0) (myristic acid) | 46315.443(13985.33) | 46285.78(10922.906) | 47112.249(14211.236) | 46853.296(11340.191) |
| FA (14:1) (physeteric acid) | 5158.99(3232.701) | 4681.581(1944.979) | 5675.279(3405.73) | 4939.859(1863.019) |
| FA (15:0) (pentadecylic acid) | 10544.8(2976.688) | 10266.457(2239.062) | 10445.753(2875.65) | 10166.415(2277.093) |
| FA (16:1) (palmitoleic acid) | 87362.569(45359.632) | 76933.028(35142.058) | 94190.824(42663.898) | 81229.717(35498.966) |
| FA (17:0) (margaric acid) | 22117.789(4474.162) | 21235.84(3887.35) | 21808.81(4648.64) | 21042.005(4049.927) |
| FA (18:1) (oleic acid) | 1454192.087(480456.365) | 1349284.059(394857.372) | 1514940.428(454631.23) | 1371889.761(394848.646) |
| FA (18:2) (linoleic acid) | 573453.977(197151.585) | 559405.212(162215.142) | 585206.405(180559.916) | 569154.315(153284.183) |
| FA (18:3) (linolenic acid) | 52408.007(25261.774) | 49568.813(16119.839) | 51000.998(19313.505) | 51302.963(15495.082) |
| FA (20:1) (eicosenoic acid) | 18300.811(5932.05) | 17239.139(4691.897) | 18874.442(6071.433) | 17346.67(4535.503) |
| FA (20:2) (eicosadienoic acid) | 9414.386(2783.824) | 9398.564(2656.075) | 9649.009(2661.535) | 9583.691(2575.188) |
| FA (20:3) (eicosatrienoic acid) | 2064.852(782.798) | 2106.53(696.017) | 2092.753(748.261) | 2033.302(662.201) |
| FA (20:3) (homo-gamma-linolenic acid) | 9019.322(2564.168) | 9232.52(2388.112) | 9003.087(2432.207) | 9053.404(2381.869) |
| FA (20:4) (arachidonic acid) | 43084.156(11307.632) | 45485.009(12124.954) | 41194.285(10661.025) | 43026.798(9467.557) |
| FA (20:5) (eicosapentaenoic acid) | 7210.305(8172.735) | 5310.154(4416.295) | 6853.569(8590.614) | 5238.719(4843.886) |
| FA (22:0) (behenic acid) | 4796.588(1883.561) | 4776.335(2235.782) | 4558.918(1650.069) | 4565.138(1731.25) |
| FA (22:6) (docosahexaenoic acid) | 35785.158(23581.553) | 31506.346(14537.092) | 36424.522(25779.108) | 31812.444(15865.045) |
| FA (24:0) (lignoceric acid) | 9002.74(3316.901) | 9144.823(3857.725) | 8443.659(2750.451) | 8653.491(3064.249) |
| FA (24:1) (nervonic acid) | 3404.095(1062.601) | 3216.118(871.277) | 3222.997(823.848) | 3107.598(848.043) |
| Gal-Gal-Cer (d18:1/16:0)/Lactosylceramide (d18:1/16:0) | 37250.999(11809.088) | 37236.749(10987.775) | 36441.158(12453.56) | 37090.618(11161.394) |
| GlcCer (d38:1) | 3730.008(1287.974) | 3948.513(1065.45) | 3672.435(1216.352) | 3940.785(1040.384) |
| GlcCer (d40:1) - ESI(-) | 22073.89(7846.193) | 22731.523(6665.458) | 21197.084(7708.24) | 22296.642(6309.151) |
| GlcCer (d41:1) | 12088.153(4455.943) | 12743.895(4333.197) | 11654.937(4140.417) | 12390.181(3955.261) |
| GlcCer (d42:1) - ESI(-) | 25575.096(9070.161) | 26263.818(7929.532) | 24105.616(8565.355) | 25349.459(6949.024) |
| GlcCer (d42:2) - ESI(-) | 19888.456(7749.181) | 17973.465(6288.748) | 18678.346(6996.248) | 17368.677(5282.948) |
| GlcCer (d14:1(4E)/20:0(2OH)) | 6302.067(3492.07) | 6460.128(3125.454) | 6498.705(3668.466) | 6436.079(3158.302) |
| GlcCer (d34:1) | 7468.445(3751.415) | 8251.003(4419.557) | 6935.992(3609.829) | 8118.449(4638.314) |
| GlcCer (d40:1) - ESI(+) | 4100.143(1480.243) | 4234.688(1219.049) | 4066.633(1476.06) | 4231.376(1212.092) |

**Supplementary Table S1.** Sample mean and standard deviations of metabolites in ME/CFS patients, controls, and their subgroups.

| Metabolite | ME/CFS (n=106) | Control (n=91) | Female ME/CFS (n=75) | Female Control (n=69) |
|---|---|---|---|---|
| | mean (SD) | mean (SD) | mean (SD) | mean (SD) |
| GlcCer (d42:1) - ESI(+) | 17983.989(6028.82) | 18240.869(5395.163) | 17508.557(6212.084) | 17779.495(4934.506) |
| GlcCer (d42:2) - ESI(+) | 13729.159(5608.295) | 12051.673(4176.986) | 13382.165(5620) | 12032.955(3833.784) |
| Lactosylceramide (d18:1/24:1(15Z)) | 2755.733(1184.633) | 2537.724(815.608) | 2706.015(1061.845) | 2538.341(810.176) |
| LPC (14:0) - ESI(-) | 1816.114(1007.334) | 2045.03(925.463) | 1806.548(974.751) | 1983.11(915.949) |
| LPC (16:0) - ESI(-) | 224092.722(72172.597) | 240419.333(71704.173) | 210628.888(62974.918) | 229102.383(66473.46) |
| LPC (16:1) - ESI(-) | 3256.883(1286.541) | 3465.425(1194.712) | 3230.526(1270.007) | 3355.22(1189.375) |
| LPC (17:1) | 2907.335(879.847) | 2996.073(1096.874) | 2853.821(865.494) | 2872.984(1044.8) |
| LPC (18:0) A - ESI(-) | 7418.389(3009.415) | 8077.783(2590.056) | 7053.492(2810.519) | 7812.018(2599.199) |
| LPC (18:0) B - ESI(-) | 99494.905(36134.374) | 108242.985(32910.725) | 94247.037(33404.908) | 104492.539(33092.3) |
| LPC (18:1) - ESI(-) | 47798.748(19375.801) | 51591.653(19211.667) | 45171.37(18891.253) | 49476.855(18392.004) |
| LPC (18:2) - ESI(-) | 60906.166(28020.858) | 71522.652(31446.609) | 55570.296(24563.184) | 70307.404(30731.739) |
| LPC (20:0) | 2602.173(1120.102) | 2667.429(891.909) | 2537.269(1076.668) | 2657.007(901.485) |
| LPC (20:1) - ESI(-) | 3013.633(1298.505) | 2917.096(1050.301) | 2949.314(1165.903) | 2882.011(1092.87) |
| LPC (20:1) - ESI(+) | 713.456(419.647) | 654.494(268.413) | 667.442(370.369) | 640.514(265.426) |
| LPC (20:2) - ESI(-) | 4059.133(1281.292) | 4450.809(1469.606) | 3913.556(1120.135) | 4415.508(1530.112) |
| LPC (20:2) - ESI(+) | 518.937(285.076) | 598.846(314.265) | 489.388(255.973) | 591.929(320.009) |
| LPC (20:3) - ESI(-) | 3582.326(1705.034) | 4021.253(1564.565) | 3377.067(1587.043) | 3778.861(1520.097) |
| LPC (22:4) | 1706.729(976.366) | 1894.898(1004.387) | 1482.79(851.308) | 1675.62(753.05) |
| LPC (22:5) - ESI(-) | 675.167(447.702) | 664.823(383.062) | 588.229(365.946) | 625.645(378.889) |
| LPC (22:6) | 18296.637(8748.95) | 17120.89(7390.865) | 17892.205(8795.155) | 17059.345(7938.417) |
| LPC (o-16:0) | 7726.959(3135.329) | 8366.675(3289.673) | 6972.434(2368.186) | 8002.555(3166.715) |
| LPC (p-16:0)/LPC (o-16:1) | 10752.235(4110.82) | 11778.247(4172.532) | 9871.584(3425.003) | 11229.396(3803.267) |
| LPC (p-18:0)/LPC (o-18:1) | 5609.615(1996.592) | 5649.593(2083.157) | 5320.569(1820.383) | 5448.102(2139.783) |
| LPC (14:0) - ESI(+) | 12225.274(5163.537) | 14568.798(5062.405) | 12327.159(4956.072) | 14298.088(5037.991) |
| LPC (15:0) | 12084.127(4819.914) | 12943.535(4361.419) | 11797.995(4420.944) | 12914.474(4391.999) |
| LPC (16:0) - ESI(+) | 1926106.937(412946.468) | 2025455.264(509547.986) | 1845303.056(386878.744) | 1953940.844(488040.87) |
| LPC (16:1) - ESI(+) | 18357.864(6537.325) | 19256.04(6089.078) | 18102.21(5252.964) | 19005.283(6187.711) |
| LPC (18:0) - ESI(+) | 597884.618(199525.827) | 642665.246(221367.761) | 575304.178(190861.694) | 627007.336(232772.768) |
| LPC (18:1) - ESI(+) | 267553.679(104625.2) | 280752.212(113394.873) | 250815.703(90364.26) | 265656.896(100100.885) |
| LPC (18:2) - ESI(+) | 453109.903(200728.084) | 547483.468(267853.354) | 411754.083(175401.553) | 530079.324(250479.673) |
| LPC (18:3) | 3781.745(1556.029) | 4762.878(2448.244) | 3586.936(1415.53) | 4529.316(1778.483) |
| LPC (20:3) - ESI(+) | 18658.988(7504.447) | 20830.978(7040.033) | 17363.957(6484.747) | 19646.263(5900.561) |
| LPC (20:4) | 92588.56(39171.901) | 99801.287(39143.693) | 83115.248(29224.178) | 91499.708(29535.203) |
| LPC (20:5) | 8554.056(10259.538) | 6693.724(5097.151) | 8602.432(11112.606) | 6635.493(5583.273) |
| LPC (22:5) - ESI(+) | 4133.499(2034.537) | 4006.158(1626.825) | 3743.911(1786.148) | 3766.871(1528.189) |
| LPE (16:0) | 3007.892(1492.945) | 3345.916(1755.537) | 2828.742(1357.831) | 3246.499(1691.23) |
| LPE (18:2) - ESI(-) | 4521.977(2417.216) | 5517.915(3099.215) | 4201.716(2091.519) | 5649.956(3201.503) |
| LPE (20:4) - ESI(-) | 3411.332(1579.491) | 3835.167(1358.006) | 3242.724(1471.892) | 3781.322(1408.073) |
| LPE (22:6) | 2511.314(1103.103) | 2584.869(1142.272) | 2513.448(1167.059) | 2639.765(1167.794) |
| LPE (18:0) | 2976.4(935.468) | 3330.697(987.882) | 2976.72(914.575) | 3291.538(1032.858) |
| LPE (18:2) - ESI(+) | 3940.782(1604.285) | 4738.299(2407.019) | 3705.225(1546.128) | 4828.722(2527.474) |
| LPE (20:4) - ESI(+) | 3205.181(1241.82) | 3617.274(1320.76) | 3131.703(1232.432) | 3563.497(1232.696) |
| PC (16:0/9:0(CHO)) | 4327.064(2722.879) | 4282.307(2603.353) | 4063.458(2415.238) | 4088.188(2455.075) |
| PC (32:0) - ESI(-) | 14490.961(3964.7) | 14558.291(3478.332) | 14173.518(3865.137) | 14514.44(3524.546) |
| PC (32:1) - ESI(-) | 14447.376(7522.552) | 14662.134(6394.392) | 15141.45(7626.112) | 14641.654(6148.462) |
| PC (32:2) - ESI(-) | 13463.361(6508.264) | 16204.369(5624.383) | 14050.917(5985.079) | 16666.589(5393.249) |
| PC (33:1) - ESI(-) | 2353.684(1083.204) | 2374.083(853.644) | 2445.074(1104.637) | 2361.716(832.373) |
| PC (33:2) - ESI(-) | 3860.198(1605.855) | 4336.55(1397.757) | 4062.384(1595.616) | 4497.506(1396.348) |

**Supplementary Table S1.** Sample mean and standard deviations of metabolites in ME/CFS patients, controls, and their subgroups.

| Metabolite | ME/CFS (n=106) | Control (n=91) | Female ME/CFS (n=75) | Female Control (n=69) |
|---|---|---|---|---|
| | mean (SD) | mean (SD) | mean (SD) | mean (SD) |
| PC (34:0) - ESI(-) | 8251.523(2184.754) | 8469.609(1811.655) | 8053.581(2009.851) | 8465.187(1814.817) |
| PC (34:2) - ESI(-) | 240630.097(60351.691) | 258867.547(56457.148) | 237975.775(58150.543) | 260900.337(57213.603) |
| PC (34:3) | 16244.273(6359.067) | 18088.554(6094.852) | 16619.89(6500.646) | 18691.548(6145.103) |
| PC (34:4) - ESI(-) | 3988.556(2296.794) | 4598.624(1731.583) | 4259(2366.311) | 4641.965(1743.509) |
| PC (35:1) - ESI(-) | 3093.959(1098.086) | 3051.722(977.026) | 3114.587(1111.442) | 2990.347(962.299) |
| PC (35:2) | 9207.837(2691.891) | 9716.991(2633.897) | 9074.375(2713.072) | 9977.682(2615.637) |
| PC (35:4) - ESI(-) | 2567.664(1136.268) | 2622.666(936.586) | 2598.481(1176.03) | 2585.304(775.291) |
| PC (36:1) - ESI(-) | 60700.134(17787.723) | 61602.252(15986.223) | 60443.364(17987.433) | 61466.942(16301.303) |
| PC (36:3) A - ESI(-) | 78001.315(26248.654) | 80601.946(20165.252) | 76566.024(25173.844) | 80141.523(19892.495) |
| PC (36:3) B - ESI(-) | 80122.729(25240.712) | 84455.963(19138.885) | 79575.825(24715.727) | 83766.047(19227.98) |
| PC (36:4) A - ESI(-) | 50001.555(19418.961) | 58881.412(19057.435) | 49070.262(18583.73) | 60741.165(19396.955) |
| PC (36:4) B - ESI(-) | 165608.498(45603.524) | 172562.505(36378.869) | 161991.938(43674.505) | 170395.971(35833.334) |
| PC (36:5) A | 2138.31(1339.816) | 2567.216(1605.787) | 2117.074(1258.51) | 2799.54(1649.863) |
| PC (36:5) B | 41857.83(35378.719) | 33407.87(22725.882) | 39725.734(31438.644) | 33188.832(24114.183) |
| PC (37:2) - ESI(-) | 1427.867(571.478) | 1549.892(656.208) | 1435.635(578.748) | 1615.775(681.224) |
| PC (37:3) | 19429.669(8642.077) | 20766.774(7555.42) | 19976.601(8593.197) | 21666.748(7553.164) |
| PC (37:4) - ESI(-) | 3610.43(1262.482) | 3600.739(1164.94) | 3505.437(1289.633) | 3567.025(1181.887) |
| PC (38:2) | 4724.469(1540.569) | 4816.371(1178.537) | 4783.461(1545.779) | 4933.012(1212.134) |
| PC (38:3) - ESI(-) | 29648.655(10260.002) | 31432.948(7745.999) | 29658.427(10093.354) | 30947.485(7542.327) |
| PC (38:4) A - ESI(-) | 57986.619(17566.019) | 60883.346(16516.825) | 57004.694(17456.97) | 60297.426(17078.597) |
| PC (38:5) A - ESI(-) | 19182.481(6029.043) | 19166.914(4819.422) | 18676.833(5687.059) | 18966.356(4798.877) |
| PC (38:5) B - ESI(-) | 12329.808(8869.88) | 10448.295(4369.029) | 12064.361(8160.002) | 10504.881(4619.341) |
| PC (38:6) - ESI(-) | 75564.438(34561.384) | 68827.018(26086.774) | 75541.752(33241.58) | 68594.619(26922.203) |
| PC (39:6) | 18191.534(9715.309) | 16532.441(6454.175) | 20136.065(10343.655) | 17559.66(6586.754) |
| PC (40:4) - ESI(-) | 3651.767(1838.864) | 3888.447(1301.818) | 3545.995(1888.218) | 3778.623(1260.441) |
| PC (40:5) A - ESI(-) | 4364.652(1888.948) | 4187.943(1433.041) | 4197.852(1807.296) | 4159.711(1422.71) |
| PC (40:5) B - ESI(-) | 1949.867(1019.491) | 2160.184(912.295) | 1965.854(1071.325) | 2061.666(780.046) |
| PC (40:6) B | 20657.541(8269.246) | 18554.405(6543.511) | 20500.278(7688.845) | 18505.132(6694.103) |
| PC (40:7) - ESI(-) | 2669.718(1393.762) | 2433.114(925.635) | 2687.754(1387.038) | 2469.947(915.054) |
| PC (40:8) - ESI(-) | 2630.788(1139.643) | 2734.932(900.85) | 2529.097(1022.741) | 2689.323(903.241) |
| PC (42:5) | 3139.766(1127.172) | 3024.694(1140.962) | 3350.413(1008.909) | 3183.849(1178.252) |
| PC (42:6) | 2237.693(1216.735) | 2297.879(1311.502) | 2395.646(1254.694) | 2328.609(1331.271) |
| PC (o-32:0) - ESI(-) | 2806.219(868.036) | 2847.252(794.141) | 2704.901(854.074) | 2839.836(808.911) |
| PC (p-32:0)/PC (o-32:1) - ESI(-) | 2338.644(927.886) | 2365.854(804.708) | 2198.427(840.691) | 2327.428(708.201) |
| PC (p-32:0)/PC (o-32:1) - ESI(+) | 29152.037(7872.188) | 30771.998(7341.47) | 28791.902(7290.532) | 31452.394(7114.976) |
| PC (p-34:1)/PC (o-34:2) A | 4520.84(2258.236) | 5300.943(2014.065) | 4423.379(2326.169) | 5437.238(2111.38) |
| PC (p-34:1)/PC (o-34:2) B | 2874.162(1241.363) | 2975.868(1271.407) | 2840.858(1220.775) | 3051.915(1202.753) |
| PC (p-34:2)/PC (o-34:3) - ESI(-) | 13080.94(4655.151) | 15147.291(4618.811) | 12892.841(4865.648) | 15380.58(4804.902) |
| PC (p-36:1)/PC (o-36:2) | 1579.393(1039.658) | 1842.803(957.813) | 1526.708(1086.379) | 1895.22(1014.876) |
| PC (p-36:3)/PC (o-36:4) - ESI(-) | 21432.678(7961.51) | 23318.595(6892.306) | 20094.728(7800.44) | 22439.049(5928.081) |
| PC (p-36:3)/PC (o-36:4) - ESI(+) | 227889.598(84207.01) | 254677.838(68833.936) | 218750.994(79336.03) | 250260.239(66263.843) |
| PC (p-36:4)/PC (o-36:5) - ESI(-) | 12136.93(5243.46) | 13380.29(4590.962) | 11209.419(4598.339) | 12873.218(4180.114) |
| PC (p-38:3)/PC (o-38:4) - ESI(-) | 7180.199(2581.694) | 8011.26(2278.842) | 6875.31(2520.292) | 7818.047(2159.075) |
| PC (p-38:4)/PC (o-38:5) A | 16408.903(5205.505) | 17597.828(4431.343) | 15828.761(5468.838) | 17257.046(4153.423) |
| PC (p-38:4)/PC (o-38:5) B | 2559.33(1097.47) | 2769.763(1152.827) | 2456.79(1084.768) | 2618.333(1025.333) |
| PC (p-38:5)/PC (o-38:6) | 1297.362(880.416) | 1291.415(783.351) | 1279.173(752.065) | 1286.904(829.333) |
| PC (p-40:1)/PC (o-40:2) | 1485.155(608.109) | 1292.088(473.435) | 1494.473(632.47) | 1309.365(478.286) |
| PC (p-40:3)/PC (o-40:4) | 1750.816(685.913) | 1868.47(638.755) | 1709.895(678.64) | 1814.465(639.48) |

**Supplementary Table S1.** Sample mean and standard deviations of metabolites in ME/CFS patients, controls, and their subgroups.

| Metabolite | ME/CFS (n=106) | Control (n=91) | Female ME/CFS (n=75) | Female Control (n=69) |
|---|---|---|---|---|
| | mean (SD) | mean (SD) | mean (SD) | mean (SD) |
| PC (p-40:4)/PC (o-40:5) - ESI(-) | 1855.515(641.097) | 1784.499(529.196) | 1816.722(663.909) | 1767.847(538.828) |
| PC (p-40:7)/PC (o-40:8) | 12784.77(9315.863) | 11294.973(6789.332) | 14104.589(8777.856) | 12457.324(6433.103) |
| PC (p-42:3)/PC (o-42:4) | 4785.842(1784.509) | 5178.399(1630.148) | 4717.303(1763.415) | 5127.617(1512.806) |
| PC (p-42:4)/PC (o-42:5) - ESI(-) | 2268.307(909.076) | 2270.498(870.57) | 2194.447(934.078) | 2241.394(850.209) |
| PC (p-42:5)/PC (o-42:6) | 618.687(355.575) | 610.366(274.685) | 607.345(377.235) | 581.2(268.483) |
| PC (p-44:4)/PC (o-44:5) - ESI(-) | 2845.694(1045.311) | 2893.365(908.463) | 2771.075(1020.627) | 2855.548(932.176) |
| PC (p-44:5)/PC (o-44:6) | 3310.494(1275.999) | 3174.484(993.019) | 3381.333(1340.399) | 3219.029(969.884) |
| PC 34:4e | 6443.081(2515.863) | 7999.075(2933.128) | 6516.076(2412.817) | 8349.359(2908.212) |
| PC 38:7e | 16964.282(13526.498) | 15342.968(9446.371) | 17136.82(13560.53) | 15811.988(10420.609) |
| PC 40:5e | 12165.024(3482.196) | 12677.876(3009.874) | 12168.625(3744.067) | 12594.987(2717.054) |
| PC 40:6e | 22316.948(7430.313) | 21474.846(6189.986) | 22072.534(7682.321) | 21356.415(5856.757) |
| PC (28:0) | 3246.565(4662.849) | 3239.16(2348.658) | 3627.569(5348.607) | 3183.558(2184.545) |
| PC (30:0) | 158777.616(83572.094) | 179096.713(65617.314) | 168743.529(90634.233) | 178533.126(59846.758) |
| PC (31:0) | 10922.351(4521.628) | 11042.588(3052.892) | 11579.689(4762.557) | 11515.423(3114.079) |
| PC (31:1) | 1677.788(1455.465) | 1660.939(915.401) | 1878.221(1600.447) | 1764.88(950.338) |
| PC (32:0) - ESI(+) | 256268.716(58170.605) | 250135.791(53964.518) | 257737.991(58248.811) | 252341.298(49340.284) |
| PC (32:1) - ESI(+) | 239846.485(124170.713) | 249536.641(113064.066) | 256265.278(131558.123) | 254543.335(118029.568) |
| PC (32:2) - ESI(+) | 56756.644(28246.729) | 67703.831(25937.146) | 61281.052(25930.591) | 72298.473(25007.78) |
| PC (33:0) | 10952.596(5641.864) | 11592.978(3878.492) | 11587.264(5855.338) | 11988.418(3933.779) |
| PC (33:1) - ESI(+) | 53543.632(20928.978) | 53553.558(13337.174) | 57061.868(21666.224) | 55115.812(13475.168) |
| PC (33:2) - ESI(+) | 78531.369(27527.116) | 87270.676(25504.3) | 83522.469(27977.154) | 93784.229(23994.517) |
| PC (34:0) - ESI(+) | 56673.124(14226.497) | 56575.078(11243.201) | 56978.355(13113.206) | 57245.93(10988.946) |
| PC (34:1) | 4013338.964(720952.007) | 4067426.422(727466.087) | 4097795.071(686391.117) | 4072387.886(668154.082) |
| PC (34:2) - ESI(+) | 8298954.78(1436737.757) | 8880671.15(1333757.007) | 8418099.742(1517545.767) | 9131469.985(1282337.625) |
| PC (34:3) A | 97711.278(31702.813) | 106631.972(39043.209) | 103128.289(28956.558) | 112048.788(37578.688) |
| PC (34:3) B | 108771.059(36242.03) | 128990.918(46188.814) | 116006.756(36977.04) | 137512.831(45224.383) |
| PC (34:3) C | 86598.612(39702.386) | 108697.965(49708.994) | 90974.287(44406.274) | 112006.398(49285.341) |
| PC (34:4) - ESI(+) | 17218.357(9083.701) | 20014.966(7594.249) | 18765.38(9434.486) | 20817.07(7544.067) |
| PC (35:1) - ESI(+) | 46139.315(13080.522) | 45638.931(9232.36) | 48062.7(13479.237) | 46489.793(9024.083) |
| PC (35:2) A | 109536.799(36351.893) | 103212.896(30675.48) | 115910.298(36351.794) | 104178.68(32765.995) |
| PC (35:2) B | 131176.614(32124.982) | 137458.977(30973.043) | 136153.641(31409.779) | 144620.016(29535.737) |
| PC (35:3) | 40372.126(10564.132) | 44343.372(11336.707) | 42224.005(10290.336) | 46715.552(10847.772) |
| PC (35:4) - ESI(+) | 22778.31(8961.192) | 23974.006(6756.722) | 24298.305(9278.648) | 25055.909(6516.302) |
| PC (36:1) - ESI(+) | 656088.196(185659.526) | 660392.494(148989.452) | 684314.568(191032.324) | 670588.694(154080.891) |
| PC (36:2) | 4529060.48(909760.507) | 4996033.975(881985.183) | 4659485.5(885858.543) | 5202361.202(878003.988) |
| PC (36:3) A - ESI(+) | 1277000.345(445850.9) | 1366182.069(370360.325) | 1306783.59(413360.001) | 1413698.006(377268.745) |
| PC (36:3) B - ESI(+) | 1639032.954(502617.113) | 1716418.049(393778.108) | 1656786.979(497457.461) | 1754675.994(417736.798) |
| PC (36:4) A - ESI(+) | 344228.244(141889.724) | 433150.27(175314.159) | 352156.763(135378.468) | 460327.086(179157.48) |
| PC (36:4) B - ESI(+) | 254789.338(417790.093) | 206974.331(69274.833) | 290186.954(490075.248) | 211942.437(72849.794) |
| PC (36:4) C - ESI(+) | 3945315.162(980469.466) | 4037185.749(932394.375) | 4017345.587(950229.435) | 4041263.953(904180.707) |
| PC (36:5) C | 63676.772(293144.05) | 37777.207(75598.623) | 75066.456(344291.891) | 36454.358(71241.705) |
| PC (36:5) D | 466335.725(583640.353) | 328794.138(256617.495) | 522920.29(662773.132) | 349071.021(285083.822) |
| PC (36:5)A | 10087.21(5302.765) | 12082.737(6355.171) | 10537.385(5204.716) | 13178.345(6561.16) |
| PC (36:6) | 7517.927(4687.721) | 7514.303(3539.182) | 8522.707(4868.049) | 8177.996(3691.057) |
| PC (37:2) - ESI(+) | 12948.098(3497.278) | 14134.187(3647.086) | 13382.403(3420.515) | 14766.724(3764.208) |
| PC (37:4) - ESI(+) | 51981.159(19322.358) | 49132.105(16397.252) | 55140.179(19721.563) | 49171.954(15891.326) |
| PC (37:5) | 36908.954(34432.55) | 28967.669(14940.073) | 40739.105(38835.732) | 30400.788(16224.117) |
| PC (37:6) | 9891.772(5465.158) | 8696.041(4047.719) | 11125.227(5788.227) | 9452.384(4154.234) |

**Supplementary Table S1.** Sample mean and standard deviations of metabolites in ME/CFS patients, controls, and their subgroups.

| Metabolite | ME/CFS (n=106) | Control (n=91) | Female ME/CFS (n=75) | Female Control (n=69) |
|---|---|---|---|---|
| | mean (SD) | mean (SD) | mean (SD) | mean (SD) |
| PC (38:3) - ESI(+) | 545962.223(199887.214) | 581406.422(170804.445) | 563669.536(203120.777) | 586858.299(174007.685) |
| PC (38:4) A - ESI(+) | 129827.943(52570.47) | 130649.431(33178.708) | 136588.525(56450.574) | 134821.392(33538.443) |
| PC (38:4) B - ESI(+) | 138945.853(49865.612) | 150899.97(43419.782) | 139468.52(48409.776) | 148559.57(42347.703) |
| PC (38:4) C - ESI(+) | 2313829.998(565304.201) | 2391882.015(574272.595) | 2369459.874(544941.787) | 2417067.78(566241.852) |
| PC (38:5) A | 732724.247(176405.92) | 721011.877(165292.616) | 749250.346(174332.641) | 726361.694(164347.65) |
| PC (38:5) B - ESI(+) | 385628.57(330057.265) | 324021.431(136699.566) | 421681.473(375952.014) | 336941.192(149353.949) |
| PC (38:6) A - ESI(+) | 122703.865(38145.153) | 135842.09(35120.916) | 127714.075(38815.023) | 139468.196(35495.718) |
| PC (38:6) B - ESI(+) | 3683483.214(1066200.996) | 3373471.275(825187.655) | 3837736.421(1126227.192) | 3485281.204(837697.48) |
| PC (38:7) | 9067.129(4522.378) | 8494.519(3671.831) | 10063.09(4613.949) | 9197.56(3668.259) |
| PC (39:4) | 4951.364(1392.551) | 4980.401(1209.519) | 5161.287(1412.986) | 5084.381(1196.739) |
| PC (40:4) - ESI(+) | 40710.749(17107.156) | 43768.486(14761.039) | 39908.33(17452.64) | 42287.995(13584.315) |
| PC (40:5) A - ESI(+) | 160984.496(51414.716) | 153215.298(48927.121) | 163977.645(53826.665) | 154048.446(52495.588) |
| PC (40:5) B - ESI(+) | 39290.673(16831.51) | 43514.802(16159.023) | 39626.637(17297.008) | 43357.103(15760.681) |
| PC (40:6) A | 26324.155(8352.257) | 26286.775(7120.512) | 27429.399(8246.52) | 27065.817(7139.141) |
| PC (40:6)B | 523343.738(235330.416) | 448154.607(162581.361) | 562604.084(254764.122) | 467455.297(167059.509) |
| PC (40:7) A - ESI(+) | 14123.954(4521.843) | 15295.102(4113.716) | 14194.433(4112.738) | 15377.362(4186.091) |
| PC (40:7) B - ESI(+) | 294687.493(123648.862) | 260986.662(85720.284) | 310043.863(123498.525) | 274436.004(85433.318) |
| PC (40:8) - ESI(+) | 15239.14(4848.028) | 15297.801(4079.753) | 15552.704(4225.795) | 15705.297(4292.469) |
| PC (42:10) | 2726.931(1374.175) | 2509.831(1360.724) | 2807.197(1358.449) | 2557.702(1471.573) |
| PC (o-32:0) - ESI(+) | 33066.939(8835.009) | 34463.696(8408.236) | 32634.257(8847.324) | 35119.871(8361.956) |
| PC (o-34:0) | 4253.752(1544.24) | 4625.498(1473.237) | 4254.556(1486.614) | 4787.885(1516.673) |
| PC (p-32:1)/PC (o-32:2) | 7732.065(2525.44) | 8442.017(2078.799) | 7658.733(2098.63) | 8519.511(2085.491) |
| PC (p-34:1)/PC (o-34:2) | 90514.308(41518.544) | 112732.145(42037.713) | 90362.3(43840.811) | 118159.371(43883.945) |
| PC (p-34:2)/PC (o-34:3) - ESI(+) | 134516.098(48579.028) | 167161.547(53529.226) | 136366.181(49093.247) | 173386.082(53513.378) |
| PC (p-36:1)/PC (o-36:2) B | 4576.41(1860.937) | 4990.913(1761.819) | 4686.759(1871.805) | 5235.944(1736.014) |
| PC (p-36:2)/PC (o-36:3) | 18401.743(7473.236) | 22260.671(8493.763) | 18848.555(7606.071) | 23242.285(8689.103) |
| PC (p-36:4)/PC (o-36:5) - ESI(+) | 222255.821(73873.841) | 244403.223(62140.636) | 213961.102(65308.437) | 240147.53(55991.321) |
| PC (p-36:5)/PC (o-36:6) | 43163.98(47010.214) | 35793.982(29043.548) | 43196.944(47713.695) | 35753.832(31191.928) |
| PC (p-38:3)/PC (o-38:4) A - ESI(+) | 23415.263(7747.924) | 26268.221(7020.386) | 23132.94(7893.985) | 25693.398(6251.518) |
| PC (p-38:3)/PC (o-38:4) B - ESI(+) | 60681.832(24417.325) | 68924.225(21887.47) | 59401.39(24899.037) | 68257.213(21742.315) |
| PC (p-38:4)/PC (o-38:5) A | 244736.068(73932.864) | 253186.358(52211.505) | 241258.162(73545.028) | 251216.834(48379.253) |
| PC (p-38:4)/PC (o-38:5) B | 46348.51(17350.154) | 52251.201(14810.562) | 45822.847(17680.613) | 51798.501(13880.981) |
| PC (p-38:5)/PC (o-38:6) A | 47602.588(24906.045) | 44907.468(17837.495) | 48165.106(23970.811) | 46178.478(18332.548) |
| PC (p-38:5)/PC (o-38:6) B | 60693.148(18465.69) | 63136.031(15203.868) | 61419.069(17813.941) | 64316.344(14030.843) |
| PC (p-38:6)/PC (o-38:7) | 38202.501(19743.59) | 36368.564(13760.086) | 38853.179(19547.248) | 37243.933(14318.613) |
| PC (p-40:4)/PC (o-40:5) - ESI(+) | 17184.561(5389.491) | 17672.249(4621.409) | 17091.52(5725.53) | 17547.731(4356.749) |
| PC (p-40:6)/PC (o-40:7) A | 40200.774(19957.817) | 35888.168(13738.726) | 41534.712(18886.27) | 37069.642(14321.062) |
| PC (p-40:6)/PC (o-40:7) B | 13510.106(6889.457) | 13100.064(5544.826) | 13963.862(7043.192) | 13685.693(5731.919) |
| PC (p-42:4)/PC (o-42:5) - ESI(+) | 12900.718(4496.163) | 12896.037(3917.285) | 12960.829(4885.885) | 12882.917(3559.204) |
| PC (p-42:5)/PC (o-42:6) A | 10237.461(3726.694) | 10713.657(3758.542) | 10263.989(3735.885) | 10641.522(3407.914) |
| PC (p-42:5)/PC (o-42:6) B | 3974.188(1415.292) | 3956.085(1237.45) | 4073.113(1509.236) | 4103.264(1204.198) |
| PC (p-44:4)/PC (o-44:5) - ESI(+) | 14453.858(5455.893) | 14223.731(4628.827) | 14264.516(5559.845) | 14295.85(4388.886) |
| PE (34:1) | 1935.048(1193.975) | 1997.27(1266.931) | 1940.194(1173.534) | 1969.02(1276.846) |
| PE (34:2) - ESI(-) | 2982.653(1876.385) | 3405.593(2151.45) | 3029.793(1764.276) | 3459.196(2123.934) |
| PE (36:1) - ESI(-) | 418.862(181.396) | 445.435(181.825) | 428.886(176.713) | 435.757(175.15) |
| PE (36:2) | 7905.774(4135.38) | 8803.532(4225.603) | 7732.053(3516.886) | 8864.748(4103.972) |
| PE (36:3) | 2026.786(1248.378) | 2279.738(1430.26) | 1955.058(975.455) | 2312.145(1448.017) |
| PE (36:4) - ESI(-) | 2941.499(1644.752) | 3164.472(1950.982) | 3029.575(1641.817) | 3130.314(1951.074) |

**Supplementary Table S1.** Sample mean and standard deviations of metabolites in ME/CFS patients, controls, and their subgroups.

| Metabolite | ME/CFS (n=106) | Control (n=91) | Female ME/CFS (n=75) | Female Control (n=69) |
|---|---|---|---|---|
| | mean (SD) | mean (SD) | mean (SD) | mean (SD) |
| PE (38:2) | 629.602(222.186) | 682.787(206.229) | 631.364(228.296) | 669.438(196.116) |
| PE (38:6) - ESI(-) | 7437.759(5142.646) | 7356.489(4624.489) | 8012.329(5306.57) | 7427.759(4778.33) |
| PE (p-34:1)/PE (o-34:2) - ESI(-) | 2025.04(785.489) | 2051.91(695.945) | 1973.582(738.517) | 2059.683(685.308) |
| PE (p-34:2)/PE (o-34:3) | 3040.334(1576.283) | 3609.46(1747.103) | 2882.224(1449.324) | 3661.438(1859.658) |
| PE (p-36:1)/PE (o-36:2) - ESI(-) | 2760.373(1320.05) | 3068.12(1351.566) | 2635.577(1273.389) | 3006.465(1281.886) |
| PE (p-36:2)/PE (o-36:3) - ESI(-) | 5957.994(2773.012) | 6972.062(2936.918) | 5688.236(2599.885) | 6988.788(3104.579) |
| PE (p-36:4)/PE (o-36:5) - ESI(-) | 15261.909(9626.399) | 15928.908(8581.977) | 13935.692(7710.994) | 15318.456(7991.015) |
| PE (p-36:5)/PE (o-36:6) | 2102.097(2962.828) | 1485.236(2041.939) | 1789.331(2236.012) | 1462.848(2221.901) |
| PE (p-38:3)/PE (o-38:4) | 1340.345(687.934) | 1513.805(782.147) | 1283.123(668.687) | 1460.129(771.765) |
| PE (p-38:5)/PE (o-38:6) - ESI(-) | 11786.825(6628.023) | 12149.489(5341.659) | 11040.293(5570.985) | 11907.516(5099.043) |
| PE (p-38:6)/PE (o-38:7) | 7532.92(4292.66) | 7188.888(3364.912) | 7276.368(3883.07) | 7078.837(3513.041) |
| PE (p-40:4)/PE (o-40:5) | 3399.555(1897.801) | 3410.027(1799.437) | 3305.616(1812.216) | 3332.351(1602.38) |
| PE (34:2) - ESI(+) | 8757.929(6410.569) | 9879.682(7371.827) | 9220.507(6694.552) | 9966.166(7816.693) |
| PE (36:1) - ESI(+) | 3379.71(1611.842) | 3316.561(1592.325) | 3263.487(1446.003) | 3237.931(1493.842) |
| PE (36:4) - ESI(+) | 14739.008(7468.332) | 15255.178(7705.241) | 15753.285(7583.178) | 15504.53(7845.412) |
| PE (38:4) | 39570.697(19154.594) | 40390.086(15772.1) | 41121.491(18723.161) | 41293.918(15461.624) |
| PE (38:6) - ESI(+) | 20257.66(12022.688) | 19884.696(11866.824) | 23154.655(12574.001) | 20815.727(12437.645) |
| PE (p-34:1)/PE (o-34:2) - ESI(+) | 2742.254(1098.637) | 2862.174(974.659) | 2672.766(988.62) | 2874.238(962.614) |
| PE (p-36:1)/PE (o-36:2) - ESI(+) | 2454.413(1024.933) | 2762.367(992.448) | 2409.249(954.259) | 2777.374(951.171) |
| PE (p-36:2)/PE (o-36:3) - ESI(+) | 8991.903(3876.705) | 11041.406(4021.531) | 8785.477(3533.158) | 11426.737(4221.509) |
| PE (p-36:4)/PE (o-36:5) - ESI(+) | 29345.071(18055.185) | 30532.635(14508.762) | 26863.725(13494.708) | 30216.432(14178.163) |
| PE (p-38:4)/PE (o-38:5) | 42614.164(20321.576) | 46103.9(17411.189) | 40822.867(18224.537) | 45964.889(16706.401) |
| PE (p-38:5)/PE (o-38:6) - ESI(+) | 24747.834(11095.999) | 26262.641(9101.812) | 23571.321(8663.977) | 26241.401(8898.027) |
| PE (p-40:4)/PE (o-40:5) A | 2896.209(2724.733) | 2989.973(2175.181) | 2689.895(2373.18) | 2967.712(1955.17) |
| PE (p-40:4)/PE (o-40:5) B | 3708.007(1951.993) | 3692.493(1528.458) | 3694.774(1901.318) | 3668.946(1409.857) |
| PE (p-40:5)/PE (o-40:6) | 6740.897(2550.286) | 6806.706(1987.436) | 6474.732(2366.006) | 6819.497(1911.385) |
| PG (34:0)/PG (17:0/17:0) | 76853.368(21921.357) | 85280.313(20224.124) | 75357.297(22903.419) | 85157.322(20327.055) |
| PI (38:4)/PI (18:0-20:4) | 120817.481(39400.359) | 124008.066(31642.823) | 120581.869(37338.394) | 122380.788(30748.13) |
| SM (d30:1) - ESI(-) | 1704.845(1134.453) | 1947.17(774.108) | 1741.938(1093.709) | 1972.955(715.8) |
| SM (d32:0) - ESI(-) | 1032.341(809.55) | 1064.49(657.551) | 1053.879(858.514) | 1017.411(603.003) |
| SM (d32:1) - ESI(-) | 44492.705(15861.723) | 49754.267(12913.605) | 44727.747(15016.049) | 49684.296(12615.847) |
| SM (d32:2) - ESI(-) | 3030.003(1334.652) | 3230.614(1251.27) | 3226.202(1351.425) | 3409.244(1299.318) |
| SM (d33:1) - ESI(-) | 22975.489(8246.125) | 25365.753(7028.514) | 22644.729(8358.501) | 25229.246(7270.357) |
| SM (d34:0) - ESI(-) | 4927.976(1784.951) | 5032.83(1672.907) | 4794.344(1692.618) | 5037.227(1704.116) |
| SM (d34:1) - ESI(-) | 155715.48(39317.352) | 163369.808(34296.98) | 150308.85(36641.578) | 162587.859(33725.445) |
| SM (d34:2) - ESI(-) | 57475.881(14166.69) | 60371.724(13403.338) | 57244.429(13620.13) | 61368.681(13998.656) |
| SM (d36:0) - ESI(-) | 1066.803(653.242) | 879.715(483.034) | 1014.529(570.255) | 793.512(407.436) |
| SM (d36:1) - ESI(-) | 36132.26(10486.124) | 36986.044(8862.176) | 34995.042(8656.887) | 36417.919(8481.388) |
| SM (d36:2) - ESI(-) | 5807.249(1805.715) | 5799.331(1630.41) | 5775.03(1738.188) | 5779.314(1601.4) |
| SM (d36:3) - ESI(-) | 1713.798(653.047) | 1804.421(563.54) | 1757.144(685.763) | 1812.821(597.69) |
| SM (d37:1) | 2411.418(1019.95) | 2404.494(818.507) | 2444.066(950.289) | 2409.022(763.946) |
| SM (d38:0) | 696.441(811.576) | 522.617(447.127) | 700.134(856.544) | 478.008(462.61) |
| SM (d38:2) - ESI(-) | 4790.522(1380.077) | 4974.002(1318.512) | 4778.59(1309.009) | 5107.688(1365.518) |
| SM (d39:1) - ESI(-) | 12841.047(4278.992) | 14353.601(3784.665) | 13016.383(4211.156) | 14492.537(3819.146) |
| SM (d39:2) | 12449.201(3511.981) | 12954.634(3279.589) | 13477.133(3201.714) | 13842.164(2925.889) |
| SM (d40:1) - ESI(-) | 46897.381(13275.565) | 50082.888(10695.858) | 46399.113(12523.495) | 49650.664(10908.118) |
| SM (d40:2) A - ESI(-) | 12273.795(4442.543) | 12319.223(4118.364) | 11967.871(3692.789) | 12241.65(3901.865) |
| SM (d40:2) B - ESI(-) | 17613.206(5158.79) | 18864.027(4350.959) | 17746.067(4989.101) | 19207.058(4516.62) |

**Supplementary Table S1.** Sample mean and standard deviations of metabolites in ME/CFS patients, controls, and their subgroups.

| Metabolite | ME/CFS (n=106) | Control (n=91) | Female ME/CFS (n=75) | Female Control (n=69) |
|---|---|---|---|---|
| | mean (SD) | mean (SD) | mean (SD) | mean (SD) |
| SM (d40:3) | 1079.194(398.684) | 1151.905(387.017) | 1050.49(391.197) | 1191.378(407.458) |
| SM (d41:1) - ESI(-) | 11075.677(11704.719) | 13579.641(12424.293) | 10616.592(11274.003) | 13795.881(11975.701) |
| SM (d41:2) - ESI(-) | 10157.654(3759.068) | 10335.005(4166.592) | 10404.338(3672.889) | 10507.896(4378.528) |
| SM (d42:0) - ESI(+) | 631.117(319.733) | 572.32(282.695) | 622.379(298.174) | 537.076(274.555) |
| SM (d42:0) - ESI(-) | 2435.513(802.742) | 2325.825(740.385) | 2470.148(793.653) | 2287.991(787.949) |
| SM (d42:1) - ESI(-) | 74113.422(22000.912) | 76751.556(18783.673) | 72184.156(20435.816) | 74908.652(18692.96) |
| SM (d42:3) - ESI(-) | 45369.2(13437.893) | 44941.003(10551.682) | 44198.136(12142.453) | 45334.226(10354.542) |
| SM (d43:1) - ESI(-) | 1610.475(804.178) | 1785.517(763.855) | 1618.018(814.733) | 1667.998(676) |
| SM (d43:2) - ESI(-) | 6637.877(3329.003) | 6677.809(2804.014) | 6613.861(3155.233) | 6397.93(2458.095) |
| SM (d44:2) | 3401.174(1320.178) | 2954.684(961.942) | 3395.111(1198.217) | 2970.944(987.86) |
| SM (d41:3) | 17542.807(4858.342) | 18495.055(4033.934) | 17960.03(4846.253) | 18787.425(3742.284) |
| SM (d30:1) - ESI(+) | 4627.855(3294.462) | 5081.766(2205.718) | 5028.808(3462.684) | 5291.701(2009.151) |
| SM (d32:0) - ESI(+) | 4527.417(2257.972) | 4461.163(1515.584) | 4874.777(2374.938) | 4638.532(1513.107) |
| SM (d32:1) - ESI(+) | 127487.665(50390.527) | 141586.567(42157.93) | 136372.342(49199.774) | 147059.917(42153.968) |
| SM (d32:2) - ESI(+) | 8419.355(3072.987) | 8850.662(2752.191) | 9261.809(3018.86) | 9547.077(2585.789) |
| SM (d33:1) - ESI(+) | 78897.543(30030.413) | 81545.929(24155.104) | 83598.798(30549.918) | 85232.023(24689.826) |
| SM (d34:0) - ESI(+) | 77128.042(21734.185) | 77407.547(19252.704) | 79148.787(20897.091) | 80066.77(18722.585) |
| SM (d34:1) - ESI(+) | 1815521.625(427328.564) | 1839477.475(326129.156) | 1828329.791(428344.536) | 1881662.975(305967.847) |
| SM (d34:2) - ESI(+) | 169414.754(41285.046) | 172230.478(39764.32) | 177731.172(39450.959) | 181137.089(38632.856) |
| SM (d36:0) - ESI(+) | 18743.046(8907.96) | 15332.243(6070.249) | 18843.91(8196.485) | 15312.391(6112.929) |
| SM (d36:1) - ESI(+) | 282139.394(75455.508) | 277277.338(58280.773) | 286717.136(68048.084) | 283125.157(50994.077) |
| SM (d36:2) - ESI(+) | 637435.743(182402.554) | 634824.46(155709.619) | 648968.61(164665.017) | 658143.625(148372.072) |
| SM (d36:3) - ESI(+) | 6491.66(2413.088) | 6629.303(2061.528) | 6979.774(2511.247) | 7017.571(2071.407) |
| SM (d38:1) | 224074.217(52840.443) | 238184.289(46405.144) | 233237.845(47492.938) | 244612.794(44373.49) |
| SM (d38:2) - ESI(+) | 102238.769(23254.677) | 103697.728(23384.632) | 106659.856(22123.4) | 109042.026(21335.321) |
| SM (d39:1) - ESI(+) | 55126.946(19385.078) | 60979.282(17058.805) | 59429.032(18852.674) | 63790.017(17030.253) |
| SM (d40:0) | 8519.977(3597.278) | 7725.238(2745.348) | 8866.396(3724.585) | 7636.915(2899.059) |
| SM (d40:1) - ESI(+) | 357194.17(87098.783) | 373303.076(69403.177) | 363600.727(79860.724) | 376764.167(65965.404) |
| SM (d40:2) A - ESI(+) | 192003.867(74381.653) | 206939.521(72862.228) | 197726.627(74937.129) | 215299.681(75251.437) |
| SM (d40:2) B - ESI(+) | 235158.05(59785.384) | 255298.203(57750.75) | 245271.672(56923.439) | 266399.439(54255.607) |
| SM (d41:1) - ESI(+) | 136526.853(38416.919) | 143873.721(36214.448) | 143360.216(37159.026) | 147377.748(35202.988) |
| SM (d41:2) A - ESI(+) | 41382.821(14921.589) | 40584.006(12105.041) | 43803.632(15241.692) | 42626.808(11718.251) |
| SM (d41:2) B - ESI(+) | 65492.772(20330.14) | 72077.791(21032.207) | 70731.196(19417.519) | 77074.321(20042.703) |
| SM (d42:1) - ESI(+) | 227228.943(65564.52) | 235888.054(53178.336) | 229198.281(66687.672) | 234400.426(53184.15) |
| SM (d42:2) | 552276.944(187216.547) | 490026.241(121909.07) | 554709.294(182209.049) | 499090.359(116944.768) |
| SM (d42:3) - ESI(+) | 360096.856(99212.364) | 348735.256(79546.208) | 369329.425(97539.146) | 363893.997(72446.491) |
| SM (d43:1) - ESI(+) | 6015.056(2273.428) | 6198.991(2041.865) | 6204.102(2326.495) | 6117.228(1928.598) |
| SM (d43:2) - ESI(+) | 14891.052(6807.646) | 13945.815(5754.744) | 15587.392(7127.791) | 14090.14(5552.322) |
| TG (55:6) | 10089.48(6268.964) | 8140.342(3509.246) | 10618.261(7123.437) | 7813.759(3404.847) |
| TAG (58:7)/TAG (18:1/18:1/22:5) | 17350.361(7968.934) | 16648.95(5845.672) | 17642.412(8739.651) | 16064.8(5748.474) |
| TG (60:12) | 6594.761(13689.356) | 3710.704(4070.071) | 7831.422(15989.851) | 3920.165(4474.049) |
| TG (53:5) | 10973.54(5251.608) | 10286.984(3942.7) | 10685.391(5107.823) | 10116.072(3418.881) |
| TG (60:11) | 8428.304(11674.572) | 5497.847(3845.608) | 9546.81(13530.976) | 5506.312(3912.515) |
| TG (40:0) | 6969.903(22773.737) | 5975.792(11932.991) | 5366.541(12887.212) | 6134.296(13304.798) |
| TG (40:1) | 1931.434(4239.428) | 2087.013(3783.783) | 1582.02(2660.445) | 2140.263(4173.486) |
| TG (42:0) | 9234.522(21761.184) | 8844.978(15901.608) | 7560.81(14465.31) | 9004.003(17596.082) |
| TG (42:1) | 6797.568(17520.658) | 7739.975(16319.178) | 5616.372(10985.062) | 7542.464(16998.656) |
| TG (42:2) | 2571.971(5933.309) | 3829.423(10795.773) | 2203.121(3842.725) | 3854.495(11684.619) |

**Supplementary Table S1.** Sample mean and standard deviations of metabolites in ME/CFS patients, controls, and their subgroups.

| Metabolite | ME/CFS (n=106) | Control (n=91) | Female ME/CFS (n=75) | Female Control (n=69) |
|---|---|---|---|---|
| | mean (SD) | mean (SD) | mean (SD) | mean (SD) |
| TG (42:3) | 591.158(714.913) | 671.443(879.22) | 543.039(624.657) | 667.857(910.252) |
| TG (44:0) | 19087.048(32238.546) | 17886.319(25507.344) | 16866.3(27378.177) | 16999.68(26581.255) |
| TG (44:1) | 18788.092(29782.097) | 19478.318(28152.179) | 16881.987(23775.499) | 18816.535(29572.076) |
| TG (44:2) | 8060.679(14384.219) | 10064.354(23367.775) | 6958.338(11163.511) | 10584.424(26138.23) |
| TG (46:0) | 2913.683(2889.773) | 3210.711(2865.495) | 2633.088(2657.701) | 3022.861(2847.843) |
| TG (46:1) | 78988.174(99974.002) | 74842.169(86950.751) | 73527.42(94024.564) | 67406.911(82723.716) |
| TG (46:2) | 34399.438(37849.21) | 37666.18(47712.032) | 32184.39(35761.877) | 36569.847(51554.153) |
| TG (46:3) A | 601.482(563.369) | 733.247(1466.526) | 541.566(491.284) | 783.319(1666.273) |
| TG (46:3) B | 5520.005(7775.947) | 5107.884(7131.762) | 4751.972(6081.917) | 5340.125(7936.867) |
| TG (46:4) A | 1736.197(1706.675) | 2452.533(7382.425) | 1571.274(1434.583) | 2689.604(8429.38) |
| TG (46:4) B | 1520.942(1603.93) | 2160.803(6838.834) | 1363.537(1358.418) | 2381.213(7803.968) |
| TG (46:5) | 350.027(264.398) | 426.476(1213.469) | 326.444(238.678) | 470.062(1385.206) |
| TG (48:0) | 59633.947(40059.321) | 72255.424(51804.827) | 56646.378(39295.717) | 70024.672(52229.989) |
| TG (48:1) | 445353.036(404942.517) | 421062.403(327886.505) | 412944.557(383735.571) | 379546.827(287273.446) |
| TG (48:2) | 232753.491(212433.045) | 215309.555(160839.721) | 223287.825(211409.198) | 198900.58(150604.766) |
| TG (48:3) | 66257.783(45379.776) | 68608.011(52545.382) | 63809.422(43231.229) | 65745.352(52409.463) |
| TG (48:4) A | 14636.937(12229.537) | 16489.544(19911.708) | 13975.86(11673.51) | 16013.507(20807.177) |
| TG (48:4) B | 10569.309(10549.743) | 12439.978(18504.13) | 9774.202(10123.516) | 11658.077(19127.757) |
| TG (48:5) | 2394.932(2124.793) | 2607.435(2613.644) | 2321.835(2169.799) | 2554.014(2691.21) |
| TG (48:6) | 731.456(341.644) | 1120.169(4094.332) | 735.053(362.762) | 1248.354(4683.687) |
| TG (49:0) | 5475.884(3437.407) | 5547.724(2851.467) | 5219.237(3275.43) | 5313.1(2932.212) |
| TG (49:1) | 38039.35(31682.085) | 32431.609(18199.476) | 35461.279(30090.794) | 30042.222(16962.334) |
| TG (49:2) | 5347.71(2994.124) | 4908.392(3180.31) | 5284.331(2880.868) | 4910.13(3450.395) |
| TG (49:3) | 8782.798(5619.053) | 8058.518(3765.588) | 8602.413(5371.364) | 8056.122(3929.513) |
| TG (50:0) | 28294.36(17765.307) | 35864.991(22391.136) | 26737.748(17017.073) | 33243.52(20485.763) |
| TG (50:1) | 295740.881(378872.38) | 268871.222(259991.973) | 283860.844(350257.099) | 241553.582(258746.168) |
| TG (50:2) | 1473341.655(765417.549) | 1337335.537(606243.723) | 1396824.909(729547.499) | 1257985.029(574862.635) |
| TG (50:3) A | 599554.098(373888.427) | 538070.653(271710.362) | 578201.298(358127.789) | 515201.921(266481.825) |
| TG (50:3) B | 1463.989(2654.267) | 1979.068(6822.777) | 1490.21(2747.56) | 2269.534(7783.442) |
| TG (50:4) | 113705.603(61749.855) | 111373.426(49803.428) | 108601.458(55033.535) | 110330.359(48106.23) |
| TG (50:5) | 18164.523(11639.543) | 17487.784(9234.201) | 17437.444(10448.935) | 17542.815(8919.388) |
| TG (50:6) | 2907.741(1946.11) | 2679.302(1733.966) | 2917.163(1969.149) | 2626.251(1822.35) |
| TG (51:2) | 92658.55(49837.791) | 81298.301(33345.525) | 88644.76(46745.452) | 76313.994(30285.369) |
| TG (51:3) | 73828.122(33989.875) | 68061.653(24068.889) | 70084.162(29641.3) | 65950.35(22204.849) |
| TG (51:4) | 20600.271(10073.4) | 19733.727(8062.558) | 19502.825(8653.773) | 19752.067(7808.805) |
| TG (51:5) | 3397.515(1704.001) | 3356.461(1275.262) | 3291.237(1492.907) | 3360.513(1180.21) |
| TG (52:1) | 273344.812(258320.644) | 254133.337(220416.288) | 227065.38(199122.882) | 215185.613(194263.196) |
| TG (52:2) | 3924538.986(1182333.569) | 3636556.102(1122279.122) | 3768364.854(1099628.234) | 3436434.472(1007005.155) |
| TG (52:3) | 4416231.615(1252804.656) | 4180937.847(1101985.633) | 4199674.991(1183957.428) | 4057679.469(1019804.465) |
| TG (52:4) | 133068.448(44192.17) | 130479.658(39344.454) | 126352.449(42083.066) | 129251.411(37900.358) |
| TG (52:5) | 197791.574(102954.234) | 174359.995(84000.915) | 188217.153(97375.415) | 170162.202(67877.07) |
| TG (52:6) | 25178.432(15261.735) | 23719.028(10920.524) | 24662.199(13887.622) | 22725.563(10345.977) |
| TG (53:0) | 1583.982(681.915) | 1815.471(661.05) | 1594.348(752.808) | 1804.08(672.071) |
| TG (53:1) | 5936.774(4668.413) | 5776.819(3671.696) | 5445.202(4000.772) | 5170.883(3349.695) |
| TG (53:2) | 55792.745(24993.696) | 51317.499(21651.56) | 53435.252(23073.739) | 47192.778(17319.808) |
| TG (53:3) | 73056.718(28445.208) | 67703.814(23631.61) | 70038.964(26227.57) | 64558.692(18876.147) |
| TG (53:4) | 46389.39(19729.068) | 43856.196(15698.609) | 45059.64(18992.915) | 42639.927(13631.342) |
| TG (54:1) | 29942.881(33162.778) | 29095.13(22321.153) | 23372.449(14886.461) | 25893.09(19302.326) |

**Supplementary Table S1.** Sample mean and standard deviations of metabolites in ME/CFS patients, controls, and their subgroups.

| Metabolite | ME/CFS (n=106) | Control (n=91) | Female ME/CFS (n=75) | Female Control (n=69) |
|---|---|---|---|---|
| | mean (SD) | mean (SD) | mean (SD) | mean (SD) |
| TG (54:2) | 321515.5(253928.808) | 286249.499(201970.532) | 272775.075(155956.486) | 248260.886(151416.82) |
| TG (54:3) | 1412729.685(654867.912) | 1276837.39(524744.236) | 1326143.056(537873.167) | 1201773.944(430758.739) |
| TG (54:4) | 1525311.494(686168.793) | 1377217.291(578506.358) | 1428692.291(658070.149) | 1310971.798(477314.851) |
| TG (54:5) A | 886580.788(456096.063) | 802522.834(352287.756) | 829669.398(458056.817) | 770422.494(295087.83) |
| TG (54:5) B | 262000.181(111804.806) | 243734.88(92887.319) | 246470.56(97593.421) | 220769.995(79245.363) |
| TG (54:6) A | 199405.641(130496.852) | 176401.555(94248.554) | 188477.062(133217.914) | 171595.941(78375.698) |
| TG (54:6) B | 1358.531(998.464) | 1341.842(782.147) | 1271.477(992.345) | 1356.734(732.422) |
| TG (54:6) C | 156170.234(80514.484) | 137719.361(62448.4) | 149032.02(69075.362) | 126494.779(50961.587) |
| TG (54:7) A | 49122.361(41128.683) | 44915.907(27315.049) | 46528.908(41940.085) | 44917.947(25113.962) |
| TG (54:7) B | 43061.59(27306.858) | 34198.973(17471.92) | 43732.939(29760.111) | 32296.045(16095.635) |
| TG (54:8) | 7077.455(4680.131) | 6400.128(3571.251) | 7293.169(4891.851) | 6104.244(3149.695) |
| TG (54:9) | 965.383(952.873) | 800.15(740.888) | 1118.466(1027.977) | 833.562(828.393) |
| TG (55:1) | 2381.308(1476.992) | 2293.733(1193.428) | 2159.599(1208.603) | 2084.107(1008.765) |
| TG (55:2) | 3209.3(1454.698) | 3026.046(1288.231) | 3034.168(1311.937) | 2784.835(1101.838) |
| TG (56:1) | 8425.454(9238.656) | 8724.83(13724.801) | 7159.574(8184.624) | 6856.677(5945.118) |
| TG (56:10) | 3107.113(3323.17) | 2681.061(1812.641) | 3522.679(3729.117) | 2724.639(1890.743) |
| TG (56:2) | 12956.986(10569.613) | 12163.693(13180.75) | 11547.417(9211.255) | 10159.27(7087.405) |
| TG (56:3) | 30313.86(19743.679) | 27075.315(20697.58) | 27887.522(18193.363) | 23433.262(13258.936) |
| TG (56:4) | 52216.171(23781.081) | 49073.426(20687.965) | 50726.706(24292.856) | 46878.52(18714.874) |
| TG (56:5) A | 53791.601(25940.471) | 48027.297(18998.273) | 50499.723(24773.705) | 43962.011(15967.169) |
| TG (56:5) B | 56496.764(28288.186) | 50801.304(20794.293) | 52700.992(25050.493) | 45780.962(15711.276) |
| TG (56:5) C | 113050.899(43777.393) | 107344.31(43874.117) | 110759.221(46991.808) | 102525.692(40194.12) |
| TG (56:6) | 257540.767(82209.876) | 233475.447(74511.483) | 247768.718(77523.926) | 216721.854(59967.74) |
| TG (56:7) A | 847.239(514.565) | 783.159(401.886) | 809.886(444.189) | 737.903(404.859) |
| TG (56:7) B | 187264.223(134086.996) | 142003.825(96398.78) | 199255.168(150216.195) | 136094.6(95356.661) |
| TG (56:8) A | 48927.531(26901.231) | 42661.135(18087.989) | 48460.43(28330.769) | 41241.651(17049.738) |
| TG (56:8) B | 88914.046(72816.031) | 66492.66(53152.85) | 95506.578(81848.551) | 64933.259(53859.705) |
| TG (56:9) | 16134.849(11093.145) | 13921.157(6903.286) | 17113.136(12041.655) | 13653.569(6683.489) |
| TG (57:1) | 1925.193(905.225) | 1863.322(943.086) | 1799.662(828.566) | 1695.476(767.306) |
| TG (57:2) | 2561.985(1153.018) | 2479.474(1287.033) | 2354.363(992.929) | 2217.816(949.567) |
| TG (58:1) | 6517.314(7345.499) | 6747.301(10399.217) | 5867.012(7243.153) | 5136.303(4122.358) |
| TG (58:10) | 19441.101(14120.814) | 16598.368(8410.925) | 20720.386(15648.293) | 16503.089(8469.218) |
| TG (58:2) | 10622.347(16674.782) | 10834.266(23253.565) | 9540.797(16741.937) | 7660.389(9265.011) |
| TG (58:3) | 6147.029(9675.465) | 5932.174(10911.709) | 5656.27(10073.707) | 4465.516(4635.348) |
| TG (58:4) | 3243.624(2529.139) | 2899.892(2258.659) | 3126.268(2695.788) | 2655.21(2009.437) |
| TG (58:5) | 3750.517(1804.936) | 3824.305(1507.07) | 3735.783(1894.391) | 3688.34(1418.742) |
| TG (58:6) | 14653.583(5134.288) | 13533.44(3981.36) | 14281.308(4839.397) | 12750.519(3470.825) |
| TG (58:8) | 61621.544(53618.106) | 44303.591(28847.455) | 64812.49(56697.219) | 42434.62(24878.919) |
| TG (58:9) | 45461.573(35467.981) | 34450.345(20298.576) | 48212.448(38602.355) | 33687.789(18971.409) |
| TG (59:2) | 1491.779(1048.687) | 1415.968(980.975) | 1335.284(749.673) | 1244.29(609.866) |
| TG (59:3) | 1482.159(789.741) | 1388.449(810.7) | 1407.147(698.83) | 1273.949(610.407) |
| TG (60:1) | 2548.677(2775.351) | 2609.795(3662.099) | 2380.665(2872.92) | 2089.792(1817.214) |
| TG (60:2) | 5106.678(6459.935) | 5391.177(12127.075) | 4637.255(5088.102) | 4114.618(5785.42) |
| TG (60:3) | 3356.443(4588.274) | 3498.227(6192.138) | 3132.217(4805.77) | 2631.123(2498.163) |
| TG (60:4) | 1479.004(1459.393) | 1489.448(1601.775) | 1415.878(1531.614) | 1264.185(1005.741) |
| TG (60:6) | 1058.884(479.188) | 956.092(409.713) | 1043.772(455.724) | 921.05(418.144) |
| TG (62:1) | 1025.712(1380.888) | 981.286(1057.831) | 994.782(1496.735) | 859.475(551.843) |
| TG (62:2) | 1814.769(2212.815) | 1781.426(2417.302) | 1782.514(2388.615) | 1471.359(1405.363) |

**Supplementary Table S1.** Sample mean and standard deviations of metabolites in ME/CFS patients, controls, and their subgroups.

| Metabolite | ME/CFS (n=106) | Control (n=91) | Female ME/CFS (n=75) | Female Control (n=69) |
|---|---|---|---|---|
| | mean (SD) | mean (SD) | mean (SD) | mean (SD) |
| TG (62:3) | 571.887(485.151) | 559.893(537.266) | 599.577(541.896) | 519.154(519.164) |
| TG (62:4) | 547.101(334.64) | 613.733(374.856) | 537.383(359.013) | 571.537(297.461) |
| TG (64:2) | 864.662(732.038) | 883.063(767.604) | 852.086(716.207) | 760.911(506.986) |
| TG (64:3) | 552.787(234.996) | 552.569(261.395) | 538.92(231.867) | 541.498(241.639) |
| **Oxylipins (OL)** | | | | |
| 10-nitrolinoleic acid | 0.186(0.36) | 0.149(0.221) | 0.159(0.327) | 0.141(0.219) |
| 10-nitrooleic acid | 5.022(4.039) | 4.734(2.95) | 5.063(4.384) | 4.444(2.475) |
| 11(12)-epoxy-5,8,14,17- eicosatetraenoic acid | 0.045(0.051) | 0.046(0.062) | 0.052(0.054) | 0.043(0.062) |
| 11,12-Dihydroxyicosa-5,8,14-trienoic acid | 0.729(0.365) | 0.798(0.415) | 0.688(0.328) | 0.748(0.37) |
| 11,12-Epoxyeicosa-5,8,14-trienoic acid | 0.298(0.235) | 0.25(0.225) | 0.273(0.221) | 0.235(0.213) |
| 11-Hydroxy-14,15-epoxyeicosatrienoic acid | 0.137(0.271) | 0.236(0.486) | 0.145(0.302) | 0.208(0.493) |
| 11-Hydroxy-arachidonic acid | 0.954(0.559) | 0.918(0.459) | 0.987(0.593) | 0.932(0.429) |
| 12(13)-epoxy-9,15-octadecadienoic acid | 0.131(0.14) | 0.155(0.255) | 0.093(0.079) | 0.159(0.284) |
| 12,13-dihydroxyoctadec-9-enoic acid | 3.714(2.623) | 4.151(2.308) | 3.648(2.615) | 4.042(1.846) |
| 12,13-dihydroxyoctadeca-9,15-dienoic acid | 0.107(0.148) | 0.111(0.152) | 0.12(0.157) | 0.127(0.167) |
| 12,13-epoxy-9-octadecenoic acid | 2.678(3.381) | 3.027(3.742) | 2.24(2.216) | 2.801(3.104) |
| 12-Hydroxy-5,8,10,14,17-eicosapentaenoic acid | 1.953(2.755) | 1.117(1.211) | 2.132(2.957) | 1.226(1.308) |
| 12-Hydroxy-5,8,10,14-eicosatetraenoic acid | 17.204(15.381) | 15.752(12.397) | 17.655(15.309) | 16.334(11.497) |
| 13-hydroxyoctadeca-9,11,15-trienoic acid | 0.935(0.885) | 0.869(0.593) | 0.804(0.536) | 0.9(0.626) |
| 13-Hydroxyoctadecadienoic acid | 19.586(7.961) | 19.825(7.897) | 19.549(8.022) | 19.806(7.067) |
| 13-ketooctadeca-9,11-dienoic acid | 11.779(7.152) | 12.915(7.243) | 11.241(7.005) | 12.197(7.172) |
| 14(15)-epoxy-5,8,11,17-eicosatetraenoic acid | 0.234(0.427) | 0.211(0.435) | 0.185(0.259) | 0.222(0.481) |
| 14,15-dihydroxyeicosa-5,8,11,17-tetraenoic acid | 0.445(0.453) | 0.503(0.551) | 0.44(0.469) | 0.439(0.394) |
| 14,15-dihydroxyeicosa-5,8,11-trienoic acid | 0.816(0.427) | 0.868(0.394) | 0.768(0.397) | 0.812(0.344) |
| 14-hydroxydocosa-4,7,10,12,16,19-hexaenoic acid | 13.05(14.814) | 10.01(9.899) | 14.349(15.523) | 10.753(9.705) |
| 15(16)-epoxy-9,12-octadecadienoic acid | 0.221(0.211) | 0.382(0.521) | 0.212(0.191) | 0.392(0.543) |
| 15,16-dihydroxyoctadeca-9,12-dienoic acid | 11.678(8.843) | 12.862(6.666) | 11.008(7.506) | 13.369(6.958) |
| 15-Deoxy-delta-12,14-Prostaglandin J2 | 0.043(0.036) | 0.05(0.053) | 0.041(0.03) | 0.05(0.055) |
| 15-hydroxyeicosa-5,8,11,13,17-pentaenoic acid | 0.387(0.417) | 0.288(0.303) | 0.405(0.453) | 0.256(0.293) |
| 15-hydroxyeicosa-5,8,11,13-tetraenoic acid | 3.04(1.536) | 2.905(1.282) | 3.08(1.537) | 2.967(1.282) |
| 15-ketoeicosa-5,8,11,13-tetraenoic acid | 0.159(0.211) | 0.167(0.276) | 0.165(0.197) | 0.166(0.296) |
| 15-Keto-prostaglandin E2 | 0.319(0.368) | 0.405(0.635) | 0.345(0.405) | 0.364(0.637) |
| 16(17)-epoxy-4,7,10,13,19-docosapentaenoic acid | 0.189(0.199) | 0.159(0.206) | 0.168(0.185) | 0.138(0.134) |
| 17,18-dihydroxyeicosa-5,8,11,14-tetraenoic acid | 6.057(5.778) | 4.785(3.299) | 6.143(6.338) | 4.677(3.081) |
| 17,18-Epoxy-5,8,11,14-eicosatetraenoic acid | 0.09(0.188) | 0.122(0.18) | 0.079(0.152) | 0.106(0.148) |
| 17-hydroxy-4,7,10,13,15,19-docosahexaenoic acid | 1.382(0.957) | 1.263(1.094) | 1.371(0.977) | 1.199(0.823) |
| 18-(3-ethyloxiran-2-yl)octadeca-4,7,10,13,16-pentaenoic acid | 0.406(0.383) | 0.403(0.512) | 0.376(0.277) | 0.402(0.553) |
| 19,20-dihydroxydocosa-4,7,10,13,16-pentaenoic acid | 1.929(1.019) | 1.91(1) | 1.977(1.018) | 1.94(0.997) |
| 20-Hydroxyarachidonic acid | 4.005(8.301) | 3.874(6.762) | 4.335(9.226) | 4.269(7.105) |
| 4-hydroxydocosa-5,7,10,13,16,19-hexaenoic acid | 0.763(0.83) | 0.673(0.657) | 0.798(0.865) | 0.66(0.67) |
| 5,15-dihydroxyeicosa-6,8,11,13-tetraenoic acid | 0.038(0.055) | 0.053(0.095) | 0.039(0.055) | 0.059(0.106) |
| 5,6,15-trihydroxyeicosa-7,9,11,13-tetraenoic acid | 0.157(0.194) | 0.207(0.32) | 0.152(0.189) | 0.197(0.309) |
| 5,6-dihydroxyeicosa-8,11,14-trienoic acid | 0.665(0.513) | 0.709(0.521) | 0.59(0.446) | 0.596(0.354) |
| 5-Hydroxy-6,8,11,14,17-eicosapentaenoic acid | 0.954(0.728) | 0.874(0.675) | 1.002(0.759) | 0.874(0.721) |
| 5-Hydroxy-6,8,11,14-eicosatetraenoic acid | 3.249(2.155) | 3.575(2.057) | 3.286(2.245) | 3.461(1.99) |
| 5-ketoeicosa-6,8,11,14-tetraenoic acid | 0.116(0.104) | 0.137(0.127) | 0.121(0.108) | 0.128(0.117) |
| 6-Ketoprostaglandin F1 alpha | 0.111(0.191) | 0.091(0.14) | 0.117(0.178) | 0.087(0.139) |
| 6-trans-Leukotriene B4 | 0.046(0.066) | 0.051(0.099) | 0.045(0.071) | 0.056(0.11) |

**Supplementary Table S1.** Sample mean and standard deviations of metabolites in ME/CFS patients, controls, and their subgroups.

| Metabolite | ME/CFS (n=106) | Control (n=91) | Female ME/CFS (n=75) | Female Control (n=69) |
|---|---|---|---|---|
| | mean (SD) | mean (SD) | mean (SD) | mean (SD) |
| 8,15-dihydroxyeicosa-5,9,11,13-tetraenoic acid | 0.121(0.272) | 0.115(0.202) | 0.128(0.312) | 0.109(0.191) |
| 8,9-dihydroxyeicosa-5,11,14-trienoic acid | 0.391(0.311) | 0.373(0.243) | 0.35(0.264) | 0.337(0.197) |
| 8,9-Epoxyeicosa-5,11,14-trienoic acid | 0.627(0.483) | 0.848(0.762) | 0.563(0.448) | 0.783(0.635) |
| 8-hydroxyeicosa-5,9,11,14-tetraenoic acid | 0.707(0.521) | 0.693(0.505) | 0.744(0.563) | 0.648(0.43) |
| 9(10)-epoxy-12,15-octadecadienoic acid | 1.238(2.543) | 1.495(3.452) | 1.472(2.98) | 1.232(2.343) |
| 9(10)-epoxy-12Z-octadecenoic acid | 0.778(0.586) | 0.789(0.455) | 0.689(0.388) | 0.808(0.465) |
| 9,10-dihydroxyoctadec-12-enoic acid | 3.994(2.996) | 4.527(2.771) | 3.862(3.199) | 4.57(2.863) |
| 9,10-dihydroxyoctadeca-12,15-dienoic acid | 0.195(0.127) | 0.233(0.156) | 0.18(0.123) | 0.253(0.164) |
| 9,10-Epoxystearic acid | 2.482(2.294) | 2.68(2.248) | 2.416(2.099) | 2.472(2.177) |
| 9,11,15-trihydroxy-5,13,1Z-prostatrienoic acid | 0.022(0.082) | 0.033(0.09) | 0.023(0.096) | 0.031(0.086) |
| 9,12,13-trihydroxyoctadec-10-enoic acid | 6.746(3.013) | 7.161(3.062) | 6.492(2.67) | 7.125(3.057) |
| 9-Hydroxy-5,7,11,14,17-icosapentaenoic acid | 0.259(0.225) | 0.218(0.148) | 0.259(0.222) | 0.221(0.149) |
| 9-hydroxyeicosa-5,7,11,14-tetraenoic acid | 0.366(0.266) | 0.386(0.267) | 0.347(0.258) | 0.354(0.23) |
| 9-Hydroxylinoleic acid | 9.759(4.207) | 10.407(3.998) | 9.884(4.493) | 10.535(3.833) |
| 9-hydroxyoctadeca-10,12,15-trienoic acid | 0.602(0.484) | 0.715(0.595) | 0.553(0.466) | 0.77(0.639) |
| 9-ketooctadeca-10,12-dienoic acid | 1.218(0.86) | 1.249(0.847) | 1.184(0.863) | 1.15(0.836) |
| 9-nitrooleic acid | 0.611(0.737) | 0.692(0.676) | 0.667(0.837) | 0.591(0.564) |
| 9S,10R-dihydroxy-stearic acid | 2.561(1.966) | 2.883(2.166) | 2.623(2.19) | 2.929(2.17) |
| Leukotriene B4 | 0.079(0.051) | 0.099(0.087) | 0.083(0.053) | 0.086(0.05) |
| Leukotriene B5 | 0.01(0.015) | 0.012(0.018) | 0.01(0.016) | 0.012(0.018) |
| Prostaglandin D2 | 0.336(0.565) | 0.54(0.959) | 0.285(0.479) | 0.593(0.945) |
| Prostaglandin E1 | 0.02(0.021) | 0.019(0.026) | 0.018(0.021) | 0.019(0.027) |
| Prostaglandin E2 | 0.057(0.126) | 0.052(0.08) | 0.064(0.146) | 0.05(0.084) |
| Prostaglandin E3 | 0.185(0.427) | 0.14(0.232) | 0.146(0.26) | 0.159(0.256) |
| Prostaglandin F2a | 0.83(0.921) | 0.952(0.717) | 0.802(0.998) | 0.912(0.611) |
| Resolvin D1 | 0.147(0.362) | 0.162(0.194) | 0.113(0.158) | 0.149(0.178) |
| Thromboxane B2 | 1.893(2.013) | 2.178(4.62) | 1.952(2.115) | 2.372(5.278) |
| trans-12,13-epoxy-11-oxo-trans-9-octadecenoic acid | 1.085(0.917) | 1.082(0.66) | 0.973(0.585) | 1.072(0.653) |

**Supplementary Table S1.** Sample mean and standard deviations of metabolites in ME/CFS patients, controls, and their subgroups.

| Metabolite | ME/CFS (n=106) | Control (n=91) | Male ME/CFS (n=31) | Male Control (n=22) |
|---|---|---|---|---|
| | mean (SD) | mean (SD) | mean (SD) | mean (SD) |
| **Primary Metabolites (PM)** | | | | |
| 1,2,4-benzenetriol | 1079.549(1400.131) | 1012.135(991.038) | 742.5(467.772) | 1037.545(1061.824) |
| 1-methylgalactose NIST | 10405.216(25620.129) | 6749.551(12633.562) | 13105.533(36244.524) | 4991.227(3787.963) |
| 2-aminobutyric acid | 6778.127(2312.798) | 6819.584(2333.931) | 7352.333(2667.15) | 7241.364(1925.267) |
| 2-deoxytetronic acid | 3802(1871.5) | 3811.933(1810.183) | 4754.767(2548.144) | 4772.955(2592.052) |
| 2-hydroxybutanoic acid | 37828.971(19975.01) | 35735.831(20029.106) | 44440.767(19367.184) | 43881.318(24993.048) |
| 2-hydroxyvaleric acid | 4237.382(2334.07) | 4287.303(2132.918) | 4799.367(2957.312) | 4460.273(1950.384) |
| 2-ketoisocaproic acid | 18813.069(5556.745) | 19149.258(6078.798) | 21047.133(4542.494) | 22359.318(6496.982) |
| 3-hydroxybutyric acid | 39212.265(52464.62) | 34152(42085.017) | 35733.733(33982.447) | 29850.091(53997.176) |
| 4-hydroxybutyric acid | 2256.422(736.812) | 2269.843(849.082) | 2219.133(637.934) | 2311.182(908.418) |
| adipic acid | 2895.686(1583.539) | 2847.101(1343.166) | 2556.967(848.514) | 2810.409(1423.696) |
| alanine | 73245.52(30295.714) | 75399.281(33101.158) | 73121(33240.851) | 70609.955(26533.394) |
| alloxanoic acid | 2006.5(1375.468) | 1837.64(1056.367) | 2237.967(1425.74) | 2165.864(1153.744) |
| alpha-ketoglutarate | 1657.627(531.612) | 1527.562(557.531) | 1871.3(583.844) | 1604.636(625.02) |
| aminomalonate | 3619.186(1973.13) | 2911.888(1665.318) | 3367.233(1477.467) | 2461.545(1185.624) |
| arachidic acid | 4924.814(2338.338) | 4808.124(2355.798) | 4761.4(1586.479) | 4924.455(2869.459) |
| behenic acid | 2886.951(836.07) | 3031.506(992.687) | 2881.2(598.633) | 2993.182(1131.761) |
| benzoic acid | 17834.373(6150.942) | 17061.82(6244.429) | 18071.7(6163.721) | 15974.864(6752.184) |
| beta-alanine | 1309.833(728.789) | 1377.112(890.61) | 1463.133(1028.63) | 1333.273(525.477) |
| capric acid | 1629.931(794.154) | 1873.809(1125.953) | 1559.433(583.025) | 1713.045(870.035) |
| caprylic acid | 5704.039(1875.061) | 5854.764(2939.166) | 5550.167(1898.669) | 5450.909(2727.835) |
| citric acid | 41473.245(20630.652) | 41135.135(19465.808) | 38013.867(14403.206) | 34113.818(12532.075) |
| citrulline | 4478.147(1507.009) | 4592.506(1393.12) | 4428.767(1034.018) | 4310.591(1490.17) |
| conduritol-beta-expoxide | 1309.863(794.473) | 1296.146(815.46) | 999.3(455.193) | 1285.545(699.734) |
| creatine | 7813.647(4623.762) | 7540.27(4234.565) | 8603.167(4637.781) | 7060.909(4418.151) |
| creatinine | 8228.716(4668.248) | 8192.91(4583.811) | 8606(4409.523) | 6546.591(5155.764) |
| erythritol | 7881.353(18212.112) | 8811.472(35942.446) | 9133.6(22133.243) | 8550.818(14805.823) |
| fructose | 2597.775(3830.73) | 2215.82(2366.76) | 3919.6(5915.381) | 2029.955(1237.433) |
| fumaric acid | 9386.029(3097.763) | 9434.101(2971.299) | 9590.967(2587.965) | 9589.545(2864.12) |
| gluconic acid | 783.971(293.026) | 776.438(203.662) | 725.333(180.568) | 666.864(208.811) |
| glucose | 388539.039(113173.178) | 378196.978(110227.014) | 406466.567(114004.975) | 352852.591(128158.886) |
| glucose-1-phosphate | 2858.431(1035.667) | 2901.236(1183.342) | 2982.567(1259.644) | 2845.045(759.257) |
| glucuronic acid | 1283.441(842.594) | 1067.326(518.171) | 1525.367(1269.199) | 995.909(355.541) |
| glutamic acid | 8913.157(3993.644) | 8273.382(4085.076) | 9816.433(4292.34) | 8451.909(3559.859) |
| glutamine | 30937.275(21124.794) | 32325.416(15805.25) | 29196.6(16989.341) | 26898.955(12231.057) |
| glutaric acid | 568.461(206.452) | 532.449(209.713) | 557.067(169.993) | 524.5(234.781) |
| glyceric acid | 10747.598(4862.652) | 9509.18(4092.259) | 11127.067(5516.625) | 9404.955(3448.089) |
| glycerol | 94812.804(37180.866) | 95386.449(35827.521) | 81537.2(32838.773) | 82255.864(30429.52) |
| glycerol-alpha-phosphate | 970.676(374.848) | 952.978(388.821) | 921.367(379.96) | 933.591(346.587) |
| glycine | 68370.147(24574.146) | 66183.809(20178.354) | 56928.5(12407.6) | 54838.136(12939.173) |
| glycolic acid | 6131.461(2349.621) | 6039.528(2582.552) | 5938.933(2206.481) | 6109.273(1939.393) |
| heptadecanoic acid | 4659.657(1498.193) | 4722.045(1568.799) | 4474.633(1147.165) | 4903.455(1586.455) |
| hydroxycarbamate NIST | 9159.343(4227.978) | 9278.079(4002.355) | 8588.067(3729.171) | 9035.909(3772.975) |
| indole-3-acetate | 2888.461(1569.254) | 2930.517(2502.67) | 3032.333(1293.983) | 2542.409(918.984) |
| indole-3-lactate | 1920.853(610.853) | 2047.753(765.819) | 2129(737.703) | 2367.909(1033.791) |
| indole-3-propionic acid | 1758.569(1386.393) | 2146.528(1481.229) | 1822.2(2065.53) | 1929.727(1114.829) |
| isoleucine | 42005.147(16265.503) | 42337.787(14176.603) | 48243.833(18858.439) | 45751.682(14480.461) |
| isopropylbenzene | 16849.167(16235.754) | 16249.494(18003.148) | 13122.167(10569.752) | 14600.455(8840.735) |

**Supplementary Table S1.** Sample mean and standard deviations of metabolites in ME/CFS patients, controls, and their subgroups.

| Metabolite | ME/CFS (n=106) | Control (n=91) | Male ME/CFS (n=31) | Male Control (n=22) |
|---|---|---|---|---|
| | mean (SD) | mean (SD) | mean (SD) | mean (SD) |
| isothreonic acid | 1867.461(718.975) | 1810.944(701.346) | 1923.3(685.372) | 1543.955(729.734) |
| lactic acid | 192717.833(119615.393) | 173890.775(103094.546) | 221408.167(134550.285) | 209613.591(127179.843) |
| lauric acid | 8363.078(12244.741) | 10000.079(13605.883) | 6520.067(3256.506) | 6793.182(3183.059) |
| leucine | 70058.255(22610.471) | 77074.056(24098.618) | 77396.967(20911.301) | 82202.773(21691.79) |
| levoglucosan | 1659.137(1216.05) | 1870.697(1782.358) | 1498.1(886.062) | 2085.727(2679.176) |
| linoleic acid | 1164.52(386.052) | 1194.708(500.78) | 1070.667(424.282) | 1156.636(444.772) |
| lysine | 15344.127(10811.81) | 15204.719(7437.624) | 14161.267(10957.394) | 12799.727(6671.938) |
| lysine | 18427.5(13565.891) | 18658(9668.614) | 15555.767(14422.268) | 16682.682(9393.094) |
| lyxitol | 3098.794(1031.473) | 3248.989(1189.283) | 3103.8(1183.462) | 3618.727(1609.89) |
| maleic acid | 878.186(349.551) | 788.416(361.008) | 886.9(286.57) | 725.455(396.146) |
| maleimide | 8053.353(10208.802) | 6862.888(5737.055) | 6042.5(2661.724) | 8441(10299.541) |
| malic acid | 1120.422(375.058) | 1096.719(407.251) | 1156.867(407.694) | 1051.045(529.516) |
| maltose | 1438.294(1117.335) | 1611.483(3326.737) | 1518.333(1687.153) | 1176.091(923.678) |
| mannitol | 7384.294(34427.962) | 2531.82(2424.356) | 3000.633(2954.186) | 3071.364(3571.028) |
| mannose | 23546.755(8923.898) | 30536.393(70206.191) | 25691.367(8220.081) | 21915.591(10764.775) |
| methionine | 2349.51(1025.929) | 2403.135(919.059) | 2377.667(772.261) | 2085.091(1060.058) |
| myo-inositol | 21222.363(6524.406) | 20873.449(7257.918) | 20854(7068.743) | 21029.091(7257.152) |
| myristic acid | 2466.971(764.975) | 2620.674(814.52) | 2352.233(641.368) | 2466.318(813.394) |
| N-acetylornithine | 2287.627(630.487) | 2420.708(709.39) | 2265.967(502.235) | 2380.045(584.82) |
| N-acetylputrescine | 2335.549(2188.271) | 2392.753(2148.301) | 2827.933(3708.515) | 1891.318(696.145) |
| nicotinic acid | 22012.588(31343.256) | 23151.326(37119.635) | 19967.233(28981.09) | 12073(17905.092) |
| ornithine | 22444.167(16004.833) | 23350.124(11618.996) | 21618.067(15664.852) | 20825.227(10726.293) |
| oxalic acid | 38138.853(34967.284) | 31155.685(23852.651) | 44318.833(34672.125) | 34647.773(20502.725) |
| oxoproline | 148843.422(35478.319) | 149227.169(31627.972) | 151866.267(20413.814) | 149104.545(28022.56) |
| palmitic acid | 83826.745(26436.792) | 87530.236(29254.435) | 78853.8(20183.079) | 95229.136(29710.987) |
| palmitoleic acid | 620.618(273.823) | 620.933(267.671) | 539.233(161.287) | 621.273(308.745) |
| pelargonic acid | 29902.833(10040.315) | 30430.506(13250.889) | 27361.5(8305.543) | 30370.318(15237.921) |
| pentadecanoic acid | 10300.402(2114.862) | 10098.079(2516.636) | 10342.033(1748.35) | 9354.273(2624.098) |
| phenylalanine | 21140.618(6021.369) | 22473.629(6495.431) | 22696.967(5996.417) | 21185.773(6871.203) |
| phosphate | 62360.137(25668.008) | 60891.899(24903.713) | 49586.167(19929.948) | 49008.818(18344.709) |
| phthalic acid | 6103.99(2526.222) | 6071.337(3064.102) | 5674.333(1828.173) | 5568.409(1745.246) |
| proline | 10358.696(7248.838) | 10114.865(6954.499) | 10550.8(6458.906) | 11675.364(9101.36) |
| pseudo uridine | 2775.873(653.848) | 2855.146(716.503) | 2624.267(575.854) | 2690.227(586.367) |
| pyrrole-2-carboxylic acid | 6415.049(4040.235) | 5793.022(3390.2) | 7241.433(4396.871) | 5650.227(3767.956) |
| pyruvic acid | 11489.667(4788.685) | 10820.404(3860.672) | 10639.933(4399.941) | 10582.455(3545.42) |
| quinic acid | 1272.48(1447.647) | 1600.753(1769.131) | 1429(1579.146) | 1518.5(1932.734) |
| ribonic acid | 873.931(245.494) | 868.337(240.982) | 862.433(246.645) | 871.364(191.653) |
| ribose | 2038.549(602.463) | 2037.371(644.28) | 2244.6(593.871) | 1938.818(556.346) |
| salicylic acid | 4293.333(14512.238) | 2853.427(9490.954) | 3498.4(9108.574) | 2155.591(2848.722) |
| serine | 37492.01(17259.952) | 38555.775(15218.55) | 33113.867(10563.558) | 31679.045(12074.469) |
| stearic acid | 378059.412(154306.997) | 405425.36(162088.008) | 344273.167(88753.876) | 453855.773(203272.896) |
| succinic acid | 2428.686(828.589) | 2106.888(729.61) | 2389.367(734.142) | 2090.091(598.59) |
| sucrose | 671.637(698.395) | 613.989(381.592) | 497.833(356.559) | 549.409(306.587) |
| tagatose | 4063.725(4123.989) | 3589.213(2553.245) | 5558.5(6116.983) | 3367.909(1537.732) |
| threitol | 1452.196(807.611) | 1395.584(582.087) | 1310.2(502.671) | 1403.682(495.623) |
| threonic acid | 8560.99(4195.661) | 7285.618(3051.907) | 8462.167(4569.948) | 6538.318(2072.396) |
| threonine | 13282.686(4817.033) | 14237.438(5929.372) | 13394.9(4012.612) | 11896.455(3512.857) |
| trans-4-hydroxyproline | 1700(1224.851) | 1418.764(658.035) | 1473.1(800.435) | 1561.409(907.394) |

**Supplementary Table S1.** Sample mean and standard deviations of metabolites in ME/CFS patients, controls, and their subgroups.

| Metabolite | ME/CFS (n=106) | Control (n=91) | Male ME/CFS (n=31) | Male Control (n=22) |
|---|---|---|---|---|
| | mean (SD) | mean (SD) | mean (SD) | mean (SD) |
| tryptophan | 21152.706(10249.693) | 21495.079(8899.268) | 22440.033(11228.94) | 21921.636(9411.156) |
| tyrosine | 42719.343(12721.061) | 44217.124(10941.659) | 46640.2(12129.286) | 44423.773(14246.354) |
| urea | 785114.216(265741.232) | 831665.056(279983.579) | 857524.633(295241.831) | 812000.364(302001.423) |
| uric acid | 32108.833(16219.644) | 33489.944(17068.328) | 39065.5(16178.005) | 44843.591(20740.081) |
| valine | 79110.618(21747.997) | 82792.506(32138.457) | 87038.933(22924.252) | 86977.545(23467.177) |
| **Biogenic Amines (BA)** | | | | |
| (2R)-3-Hydroxyisovaleroylcarnitine | 1880.687(1046.064) | 1825.6(555.898) | 2216.153(1479.725) | 2020.16(485.95) |
| (3-Carboxypropyl)trimethylammonium cation | 16403.7(5302.415) | 17555.878(5550.678) | 18315.149(4992.706) | 19457.256(6639.8) |
| (R)-Butyrylcarnitine | 34078.718(15321.062) | 37105.876(16533.628) | 34054.963(11538.003) | 39881.617(17704.327) |
| .beta.-Phenyl-.gamma.-aminobutyric acid | 869.907(2440.04) | 709.92(241.154) | 1395.481(4429.338) | 674.478(152.987) |
| .epsilon.-Caprolactam | 87755.933(68018.05) | 89742.427(68532.183) | 91071.996(63510.75) | 103113.184(73798.78) |
| 1,2-Dimethylimidazole | 1819.366(2845.633) | 1694.49(1032.734) | 1313.473(387.35) | 1482.894(382.59) |
| 1-Acetyl-3-piperidinamine | 1839.3(818.798) | 1738.972(605.781) | 1809.168(842.991) | 1847.71(814.279) |
| 1-Acetyl-4-piperidinamine | 2047.585(2838.897) | 1533.704(815.512) | 2206.801(4128.195) | 1525.61(864.511) |
| 1-Methyladenosine A | 4344.676(1038.43) | 4455.103(1138.778) | 4148.434(927.249) | 4504.597(1073.83) |
| 1-Methyl-L-histidine | 22958.602(14651.539) | 23284.637(14336.386) | 24361.203(14767.623) | 22395.266(14669.359) |
| 1-Methylnicotinamide | 14470.114(11101.857) | 14251.823(7589.804) | 14589.447(14339.977) | 13557.656(6497.407) |
| 1-Monostearin | 2115.305(1451.321) | 2830.121(4236.602) | 1971.864(1270.58) | 3537.597(7784.916) |
| 1-Oleoyl-2-acetyl-sn-glycerol | 9191.209(8932.606) | 14964.193(39026.124) | 7681.703(3487.146) | 8892.076(5409.579) |
| 1-Oleoyl-sn-glycero-3-phosphoethanolamine | 6355.005(5324.408) | 6525.509(3772.709) | 6956.948(4541.941) | 7103.671(4996.319) |
| 1-Palmitoyl-2-hydroxy-sn-glycero-3-phosphoethanolamine | 6410.996(4032.932) | 6606.582(3360.162) | 6163.435(3492.196) | 6370.52(3815.333) |
| 1-Phenylpyrrolidine | 143.457(290.638) | 117.902(39.385) | 104.738(33.627) | 109.069(31.742) |
| 1-Stearoyl-2-arachidonoyl-sn-glycero-3-phospho-(1'-myo-inositol) | 1329.25(881.122) | 1346.97(825.295) | 1135.548(673.987) | 1260.022(763.463) |
| 1-Stearoyl-2-arachidonoyl-sn-glycero-3-phosphoserine | 1894.838(2682.073) | 1778.287(2056.277) | 1984.471(2882.826) | 1292.586(2080.799) |
| 1-Stearoyl-2-hydroxy-sn-glycero-3-phosphoethanolamine | 6094.917(4604.091) | 5803.43(2687.326) | 5769.144(3586.285) | 5879.906(3426.172) |
| 2,2',2''-Nitrilotriethanol | 4650.025(6102.206) | 5725.159(8642.651) | 3093.909(1373.507) | 3092.362(1222.395) |
| 2,2-Bishydroxymethyl]-2,2',2''-nitrilotriethanol | 661.365(1011.212) | 683.907(582.601) | 504.164(167.032) | 544.009(244.754) |
| 2,6-Diaminopimelic acid | 658.783(580.92) | 497.274(446.965) | 612.84(577.376) | 463.019(425.204) |
| 2-Amino-1-phenylethanol | 50064.106(14796.501) | 51939.812(12195.207) | 53117.104(15609.813) | 50826.745(8569.221) |
| 2-Hydroxyibuprofen | 3646.161(11203.95) | 2794.402(8390.204) | 5805.907(18963.666) | 3191.473(3646.479) |
| 2-Indolinone | 22360.698(11782.007) | 25707.648(13967.826) | 20038.083(9954.332) | 26672.588(12457.185) |
| 2-Methylbutyryl-L-carnitine | 15742.272(6758.335) | 19766.912(10894.625) | 19399.619(6037.212) | 27277.009(16122.643) |
| 3-(1-Pyrazolyl)-alanine | 358.412(1009.805) | 338.461(595.673) | 444.536(1499.442) | 145.964(107.017) |
| 3,4-Dimethoxybenzaldehyde | 2111.85(1817.258) | 2433.796(3874.558) | 1834.215(1232.77) | 2391.884(1909.925) |
| 3,5-Dihydroxyphenylglycine | 1521.258(1937.934) | 1263.783(1225.374) | 1006.321(944.252) | 1019.377(876.383) |
| 3-Amino-1-propanol | 14759.177(17371.398) | 11356.849(7175.122) | 11940.636(10489.283) | 13610.142(9745.729) |
| 3-Aminoquinoline | 2768.697(3284.974) | 3229.232(3720.884) | 2248.4(3315.861) | 1846.773(2416.041) |
| 3-Cysteinylacetaminophen | 85.628(278.461) | 31.56(22.915) | 82.88(208.911) | 29.864(23.752) |
| 3-Dehydrocarnitine | 2713.916(1241.044) | 2870.766(1231.212) | 2760.503(1046.042) | 3709.729(1647.21) |
| 3-Hydroxybutyrylcarnitine | 3061.608(2849.628) | 2805.221(2351.317) | 2483.169(1959.092) | 2907.379(3379.792) |
| 3-Hydroxyoleylcarnitine | 1499.787(2022.956) | 1253.212(1804.396) | 1795.047(2242.635) | 1631.204(2049.684) |
| 3-Hydroxypyridine | 1971.739(3569.602) | 3209.464(4987.632) | 3107.58(4774.692) | 2783.136(3641.25) |
| 3-Methylglutarylcarnitine | 575.563(258.16) | 1052.615(1965.769) | 581.926(186.861) | 2266.461(3737.928) |
| 3-Methylxanthine | 555.738(650.872) | 522.536(446.424) | 447.077(542.331) | 379.653(297.53) |
| 3-Pyridinemthanol | 1549.304(1050.899) | 1769.148(2316.24) | 1472.138(798.518) | 1562.799(826.931) |
| 4,5,7-Trihydroxyisoflavone | 507.397(2564.172) | 124.958(731.854) | 381.52(1786.633) | 35.467(27.31) |
| 4-Acetamidobutyric acid | 3471.712(2342.69) | 3727.521(2854.822) | 3138.805(2005.327) | 3073.521(1261.264) |
| 4-Aminomethylcyclohexanecarboxylic acid; | 1589.572(3239.52) | 1326.667(747.566) | 2026.6(5105.699) | 1335.892(818.481) |

**Supplementary Table S1.** Sample mean and standard deviations of metabolites in ME/CFS patients, controls, and their subgroups.

| Metabolite | ME/CFS (n=106) | Control (n=91) | Male ME/CFS (n=31) | Male Control (n=22) |
|---|---|---|---|---|
| | mean (SD) | mean (SD) | mean (SD) | mean (SD) |
| 4-Fluoro-.alpha.-pyrrolidinobutiophenone | 532.638(610.097) | 458.298(230.627) | 482.19(514.486) | 455.718(245.117) |
| 4'-Methyl-N-methylhexanophenone | 2337.883(2599.719) | 2483.552(2368.085) | 2152.606(2551.192) | 1993.683(1700.874) |
| 4-Pyridoxic acid; | 9506.692(32185.74) | 3611.761(6927.589) | 8220.902(16890.138) | 3016.675(5158.128) |
| 5'-S-Methyl-5'-thioadenosine | 2295.417(2441.494) | 2541.641(1775.543) | 1890.319(1403.725) | 2189.236(1572.54) |
| 5'-S-Methylthioadenosine | 2232.692(2448.886) | 2396.981(1900.293) | 1913.232(1406.276) | 2137.052(1630.408) |
| 6-Hydroxyflavone | 1191.845(6678.29) | 403.876(528.261) | 518.379(1097.162) | 560.039(783.568) |
| 6-Methoxynaphthaleneacetic acid | 7862.951(2449.945) | 9327.172(12784.412) | 7113.733(2144.366) | 13102.675(25676.836) |
| 7.alpha.-Hydroxy-3-oxo-4-cholestenoic acid | 7832.128(4085.847) | 7971.016(3525.274) | 7468.86(3109.447) | 8324.788(4240.242) |
| 7-Hydroxywarfarin | 256.276(1326.793) | 125.152(55.26) | 116.55(56.018) | 138.344(63.514) |
| 7-Methylguanosine | 2894.162(982.806) | 3126.746(1088.991) | 2694.095(896.384) | 2989.712(795.094) |
| Acetazolamide | 111.881(1032.576) | 10.68(5.05) | 347.176(1882.338) | 10.069(5.423) |
| Acetominophen | 8598.401(25412.26) | 1888.819(3095.431) | 8444.362(18580.227) | 1685.388(1372.342) |
| Acetyl-DL-carnitine | 4040.595(1781.273) | 3886.498(1192.6) | 3760.285(1568.718) | 3744.798(1143.026) |
| Acyclovir | 10042.642(38991.986) | 102.244(109.125) | 2295.767(12376.951) | 111.839(112.139) |
| Adenosine | 476.693(252.498) | 554.001(724.574) | 474.724(331.211) | 376.975(167.302) |
| Ala-Ile | 1117.937(531.714) | 1022.708(379.234) | 1034.968(513.869) | 1023.066(417.073) |
| Albendazole | 1404.595(9285.314) | 342.564(133.753) | 308.753(93.125) | 303.945(99.669) |
| Albendazole sulfoxide | 556.474(1732.254) | 706.284(5840.411) | 319.695(648.762) | 71.077(95.162) |
| alpha-Methylhistidine; | 18823.314(15559.922) | 19428.048(15625.399) | 20695.432(17703.776) | 16585.549(13746.526) |
| Alprazolam | 219.224(700.569) | 79.36(42.975) | 241.657(561.562) | 87.916(39.751) |
| Aminodiphenylmethane | 10035.44(39641.241) | 4831.419(12787.502) | 16787.772(68991.233) | 3672.393(5242.794) |
| Androstan-3-ol-17-one 3-glucuronide | 1018.393(816.112) | 997.46(700.178) | 1167.323(968.235) | 1303.702(721.195) |
| Arginine | 66971.722(30497.565) | 63686.741(31676.786) | 58680.054(21893.801) | 54207.753(41849.9) |
| Atenolol | 2831.445(26482.196) | 235.929(152.586) | 8876.504(48274.063) | 217.071(141.065) |
| Atorvastatin | 310.188(170.392) | 307.876(96.997) | 265.621(91.484) | 303.006(75.916) |
| Avobenzone | 1620.013(901.713) | 2204.034(6233.272) | 1606.788(1101.309) | 4325.933(12543.351) |
| Benthiavalicarb-isopropyl | 397.898(475.323) | 338.666(425.872) | 422.91(531.603) | 323.732(173.396) |
| Benzophenone-3 | 79.42(118.399) | 152.05(480.887) | 45.504(25.274) | 76.466(105.947) |
| Betaine | 1396162.001(400978.201) | 1465520.587(366608.787) | 1487617.048(330781.755) | 1613346.38(409960.194) |
| Betaine aldehyde cation | 14682.235(107622.215) | 56.189(19.083) | 353.14(1704.805) | 54.492(16.838) |
| Betonicine | 5018.889(9595.989) | 5425.561(9356.727) | 6823.043(14396.209) | 3025.475(3011.925) |
| Biliverden | 37882.387(29682.943) | 38990.463(24851.237) | 43941.398(31844.437) | 49271.595(33601.845) |
| Borrelidin | 1800.784(4109.912) | 1698.556(4228.984) | 1624.211(3717.448) | 2458.108(5005.844) |
| Bradykinin | 2350.694(4905.075) | 2102.181(4523.581) | 3035.61(6439.022) | 1559.314(3617.7) |
| Caffeine | 46358.68(97388.926) | 86219.943(115602.167) | 58323.446(101321.084) | 67997.582(107285.396) |
| Carbamazepine | 424.616(1432.071) | 11883.366(108750.28) | 539.159(2013.135) | 313.112(551.69) |
| Carnitine | 1001164.333(283021.215) | 1045982.351(310270.412) | 1072754.451(265918.791) | 1184452.8(372756.681) |
| Choline cation | 254611.526(57164.648) | 282373.342(63913.774) | 251280.707(55478.214) | 287961.893(70052.064) |
| cis-10,11-Dihydroxy-10,11-dihydrocarbamazepine | 525.109(153.075) | 959.717(3818.829) | 474.129(106.838) | 541.899(169.458) |
| Citrulline | 19901.677(6196.281) | 21504.169(6882.343) | 18626.854(5812.245) | 18537.558(4959.491) |
| Codeine-6-.beta.-D-glucuronide | 774.993(5522.859) | 232.532(91.583) | 204.544(67.41) | 223.963(55.908) |
| Coniferylaldehyde | 765.911(462.54) | 787.668(824.323) | 759.192(660.787) | 640.38(159.462) |
| Cotinine N-.beta.-D-glucuronide | 76.449(283.47) | 46.197(159.887) | 119.894(388.398) | 91.857(309.006) |
| Creatine | 217981.51(118303.317) | 203650.285(97913.88) | 168130.821(90371.637) | 139833.974(77604.487) |
| Creatinine | 1006729.671(287698.034) | 1029213.304(254200.686) | 1096649.707(244986.626) | 1103471.185(282335.148) |
| Cyclo(Leu-Pro) | 2228.564(1706.261) | 2027.347(1104.514) | 1920.108(1199.64) | 2104.435(1326.657) |
| D-.alpha.-Cyclohexylglycine | 6326.782(6206.867) | 8327.262(8211.34) | 4846.795(3573.209) | 8695.348(6650.099) |
| Decanoyl-L-carnitine | 42146.5(48589.406) | 42071.952(30458.853) | 43929.706(33843.773) | 34261.014(17509.83) |

**Supplementary Table S1.** Sample mean and standard deviations of metabolites in ME/CFS patients, controls, and their subgroups.

| Metabolite | ME/CFS (n=106) | Control (n=91) | Male ME/CFS (n=31) | Male Control (n=22) |
|---|---|---|---|---|
| | mean (SD) | mean (SD) | mean (SD) | mean (SD) |
| D-erythro-Sphingosine-1-phosphate | 7295.174(3378.972) | 7543.128(3687.008) | 7479.43(3924.057) | 5932.01(3771.74) |
| Dexpanthenol | 1546.048(2341.347) | 2108.653(3911.294) | 2026.092(4164.98) | 1250.989(588.431) |
| D-Fructose | 34526.336(8214.427) | 34866.41(9180.115) | 32826.06(8617.741) | 33012.635(8535.76) |
| Diazepam | 370.721(2360.704) | 46.476(76.296) | 409.32(2010.804) | 39.081(14.492) |
| Dinor-12-oxophytodienoic acid | 525.23(862.885) | 416.42(194.711) | 346.537(159.282) | 388.334(193.145) |
| Diphenhydramine | 2585.349(6127.955) | 3178.366(14715.458) | 3164.351(9467.176) | 1543.357(247.978) |
| DL-Indole-3-lactic acid | 37199.448(18913.594) | 35604.136(12065.446) | 36817.529(14023.849) | 36460.735(8849.131) |
| D-Pyroglutamic acid | 121612.569(22366.016) | 123013.056(24064.978) | 117418.865(17444.755) | 114737.442(19860.385) |
| D-Turanose | 525.908(351.452) | 631.074(581.896) | 539.308(503.012) | 697.345(817.216) |
| Ergothioneine | 767.24(1494.771) | 719.663(431.749) | 1060.01(2646.243) | 648.864(369.92) |
| Esomeprazole | 2660.51(10721.074) | 1307.945(4540.326) | 940.882(1089.34) | 785.266(268.594) |
| Ethiolat | 11862.296(20760.107) | 9779.989(18228.881) | 12251.146(15922.411) | 8966.66(6778.225) |
| Ethyldiethanolamine | 734.979(766.768) | 727.196(595.857) | 625.582(311.265) | 550.606(203.917) |
| Ethylenediaminetetraacetic acid | 663091.766(891136.229) | 496339.244(788805.165) | 560672.877(771465.86) | 405282.259(695244.459) |
| Ezetimibe | 223.611(238.157) | 234.24(306.197) | 190.923(100.856) | 221.715(122.08) |
| Fexofenadine | 450.355(238.197) | 841.091(3125.726) | 461.362(376.699) | 414.397(109.942) |
| Gabapentin | 62080.105(201779.015) | 5287.117(32734.04) | 54718.015(214648.278) | 16262.541(66113.057) |
| Glaucine | 860.63(2849.169) | 574.959(169.559) | 510.08(130.422) | 572.313(156.493) |
| Glutamic acid | 7933.975(3124.287) | 7180.498(3426.266) | 8510.012(3553.203) | 8386.114(4841.512) |
| Glutamine | 101505.785(19190.477) | 102907.791(20574.24) | 98008.205(15454.429) | 95703.445(17465.874) |
| Glycocholic acid | 4874.144(5532.195) | 3707.728(3340.534) | 4964.843(5295.367) | 3300.934(3738.019) |
| Glycodeoxycholic acid | 16574.334(17400.385) | 14780.84(19064.118) | 18685.87(18675.775) | 16809.144(24970.091) |
| Gly-Pro-Arg | 586.043(2198.483) | 428.808(1170.147) | 1030.467(2528.87) | 392.486(1136.9) |
| Guanidine | 5493.87(8781.142) | 4475.798(4190.121) | 3940.816(2612.878) | 3564.317(1070.149) |
| Guanine | 6226.314(23441.739) | 279.003(1082.414) | 1534.53(7790.221) | 618.926(2174.504) |
| Heptadecasphing-4-enine | 5557.437(4651.009) | 5538.189(4468.342) | 5627.805(4067.451) | 6702.555(4438.514) |
| Hexanoyl-L-carnitine | 8996.499(13556.537) | 8356.705(5002.169) | 8053.024(5313.882) | 7430.25(2863.103) |
| H-gamma-glutamyl-glutamine | 2339.192(1792.988) | 2006.339(1305.502) | 1863.303(1081.189) | 1764.231(1439.13) |
| Histidine | 49555.943(23320.509) | 53178.919(21745.875) | 50279.091(20666.114) | 47751.945(24317.713) |
| Homoarginine; | 3381.815(2097.316) | 3026.593(2160.863) | 3253.114(1935.796) | 2749.911(2846.298) |
| H-Pro-Hyp-OH | 2228.15(1753.581) | 1756.456(871.446) | 1919.935(987.366) | 1810.289(843.778) |
| Hydroxybupropion | 884.813(2016.341) | 1883.832(8028.514) | 695.159(1429.329) | 3606.341(14643.461) |
| Hypoxanthine | 31281.328(19491.261) | 30486.398(22288.554) | 31894.543(18046.395) | 35685.752(36498.809) |
| Ile-Glu-Arg | 365.728(1522.667) | 320.225(998.515) | 548.839(1287.857) | 163.108(392.16) |
| Indole-3-propionic acid | 1220.413(1004.075) | 1438.512(994.93) | 1048.48(1217.539) | 1401.494(1119.532) |
| Irbesartan | 34111.466(330125.328) | 1632.597(1008.364) | 1303.444(269.683) | 1529.481(244.582) |
| Isoleucine | 15404.33(5244.539) | 15607.18(5393.939) | 16744.473(5599.589) | 16930.963(4800.484) |
| Isopentenyladenine | 1014642.651(399845.125) | 1046922.522(370983.691) | 974541.979(346896.356) | 1018471.203(447527.278) |
| Isopropylamine | 4749.851(1414.044) | 5211.39(1333.561) | 4602.906(1194.634) | 5310.815(1590.676) |
| Kynurenine | 2125.376(754.54) | 2313.805(749.317) | 2287.602(798.757) | 2399.838(864.647) |
| Lamotrigine; | 2244.641(22565.331) | 18.464(16.389) | 21.207(17.446) | 18.228(13.363) |
| Lansoprazole | 736.475(4048.63) | 2665.031(13708.313) | 152.966(606.74) | 2157.927(9897.333) |
| Lauric acid diethanolamide | 413.111(567.669) | 345.335(216.443) | 331.313(185.283) | 400.649(247.413) |
| Lauroyl-L-carnitine | 5784.303(4911.391) | 6763.368(3908.375) | 5906.628(3991.35) | 6970.996(3697.297) |
| L-Carnitine | 16079.975(4595.436) | 15890.768(4307.224) | 16745.605(4396.915) | 16915.351(4585.871) |
| L-Citrulline | 20161.291(5985.134) | 21758.529(6402.073) | 18969.814(5632.898) | 19103.265(5057.045) |
| L-Cysteine-glutathione disulfide | 1116.696(1162.972) | 1224.484(1444.167) | 885.604(985.658) | 827.062(1212.613) |
| L-Cystine | 2137.312(1969.911) | 2271.747(2210.344) | 1735.631(2208.461) | 1722.462(2155.587) |

**Supplementary Table S1.** Sample mean and standard deviations of metabolites in ME/CFS patients, controls, and their subgroups.

| Metabolite | ME/CFS (n=106) | Control (n=91) | Male ME/CFS (n=31) | Male Control (n=22) |
|---|---|---|---|---|
| | mean (SD) | mean (SD) | mean (SD) | mean (SD) |
| Levocetirizine; | 9007.024(38087.67) | 5404.405(22513.101) | 7269.781(25502.991) | 2180.98(9095.237) |
| Linoleoylcarnitine | 6028.057(4694.504) | 6937.757(5240.418) | 6346.627(4037.73) | 5743.351(6436.512) |
| L-Leucine, methyl ester | 936.172(1279.128) | 734.385(675.979) | 1150.202(1821.508) | 755.012(515.461) |
| Losartan | 1124.145(6579.061) | 2289.546(19221.441) | 3140.895(11879.237) | 270.189(131.167) |
| L-Threonine | 6399.383(2197.795) | 6434.59(1928.87) | 6003.824(1480.482) | 5976.646(1817.305) |
| L-Tyrosine | 3214.55(1382.508) | 3345.219(1261.579) | 3268.273(1191.538) | 3535.471(914.006) |
| Lysine | 24048.561(14481.217) | 23142.646(14133.037) | 19317.883(8558.758) | 19596.435(15871.555) |
| Matrine | 2325.219(12579.91) | 500.431(207.181) | 422.143(164.006) | 459.304(208.897) |
| Mefenorex | 622.656(716.718) | 3807.946(26441.59) | 595.937(466.432) | 13510.497(53201.397) |
| Meloxicam | 28306.258(195269.072) | 12851.881(121675.611) | 52622.731(273406.929) | 20.812(20.718) |
| Meprobamate | 1573.996(3579.443) | 932.297(1488.818) | 2276.967(5504.175) | 967.128(668.927) |
| Metformin | 7135.099(48045.124) | 3475.203(23828.338) | 797.504(248.925) | 853.039(281.844) |
| Methacholine cation | 56501.078(33409.327) | 67485.057(42836.21) | 49193.762(28844.926) | 76916.401(76696.421) |
| Methionine | 6161.544(1634.068) | 6223.87(1770.355) | 6452.099(1526.628) | 6153.806(1527.667) |
| Methioninesulfoxide | 753.827(372.4) | 693.081(379.811) | 719.037(296.389) | 574.306(201.898) |
| Methylgallate | 139.383(458.151) | 67.958(251.269) | 92.591(233.625) | 54.513(124.446) |
| Metoprolol | 897.413(4862.805) | 234.563(653.184) | 644.411(1749.197) | 155.342(47.167) |
| Metoprolol acid | 7786.566(33910.371) | 535.074(2767.371) | 13530.901(43913.642) | 224.357(75.422) |
| Milnacipran | 276.882(1331.735) | 148.067(53.863) | 131.117(45.911) | 137.972(56.141) |
| Modafinil | 1075.48(6709.26) | 71.168(24.162) | 1584.725(8476.699) | 74.36(23.117) |
| Modafinil acid | 131.611(721.079) | 24.78(13.35) | 229.527(1164.351) | 28.115(13.217) |
| Montelukast-1,2-diol | 75.941(301.129) | 84.645(479.266) | 92.183(360.641) | 40.416(63.232) |
| Moxonidine | 3500.133(2726.862) | 3040.75(1987.891) | 3512.588(2637.755) | 2765.278(2288.019) |
| N-(3-(Aminomethyl)benzyl)acetamidine | 20921.024(28034.826) | 18034.449(15522.733) | 19747.783(31368.924) | 17082.579(11372.308) |
| N-.alpha.-Acetyl-L-arginine | 3082.187(1181.981) | 3213.461(1045.445) | 3168.06(1190.142) | 3153.484(909.352) |
| N.alpha.-Methyl-L-lysine | 29306.073(114182.748) | 13134.602(36620.029) | 33229.507(141404.266) | 24119.706(73255.747) |
| N.epsilon.-Acetyl-L-lysine | 2453.841(2107.986) | 2280.92(1269.094) | 2108.882(867.284) | 2040.458(470.561) |
| N.epsilon.-Methyl-L-lysine | 8975.039(6927.428) | 8793.019(6711.352) | 8292.549(6781.832) | 7273.656(4787.515) |
| N8-Acetylspermidine | 3001.558(1211.178) | 2900.086(896.957) | 3280.799(1509.793) | 3050.834(1076.603) |
| N-Acetylalanine | 707.391(950.515) | 582.675(496.092) | 510.307(440.215) | 470.09(436.636) |
| N-Acetyl-D-norleucine | 3757.634(3749.095) | 3860.092(3382.148) | 2860.983(3022.584) | 2986.29(1886.514) |
| N-Acetylhistidine | 1008.892(648.298) | 1039.722(610.487) | 960.013(520.722) | 1001.798(354.107) |
| N-Acetyl-L-carnosine | 2954.051(1639.621) | 2926.55(1628.741) | 4343.277(2037.336) | 4394.072(1977.762) |
| Naproxen | 3476.065(13446.579) | 4450.949(22153.769) | 6828.729(21324.608) | 1074.083(948.624) |
| NEPSILON,NEPSILON,NEPSILON-TRIMETHYLLYSINE | 13358.236(4821.32) | 15219.514(11953.152) | 15162.463(5272.414) | 20382.701(20875.919) |
| N-Methylhistidine | 35730.75(37645.645) | 33537.29(29498.297) | 48506.34(43091.365) | 37824.633(33591.229) |
| N-Methylproline | 11656.687(9296.226) | 11510.001(9904.91) | 11431.935(9358.647) | 10435.242(7514.82) |
| Norleucine | 15444.661(6060.568) | 15678.34(4128.345) | 17320.35(8087.398) | 16653.99(4344.55) |
| Nudifloramide | 64111.911(56284.553) | 63703.328(37012.76) | 76245.487(88924.622) | 64712.387(44054.616) |
| Octanoylcarnitine | 46353.596(55972.637) | 47100.775(29066.033) | 44515.865(29817.486) | 39444.681(17158.127) |
| Oleoyl-L-carnitine | 10823.414(8463.672) | 11676.244(8434.192) | 11041.169(8296.771) | 9716.931(8299.466) |
| Omeprazole sulfone | 4929.053(23929.418) | 1843.877(13957.834) | 1742.513(8252.115) | 1063.989(4185.411) |
| Omeprazole sulfone N-oxide | 1165.746(5101.882) | 421.937(2441.309) | 619.489(2330.734) | 332.765(1098.976) |
| Ondansetron | 365.61(1271.909) | 240.115(62.33) | 214.354(50.086) | 231.076(39.312) |
| Ornithine | 3959.933(2287.816) | 4022.624(2665.327) | 3568.97(1869.763) | 3451.224(2722.453) |
| p-Acetamidophenyl .beta.-D-glucuronide | 766.028(3177.057) | 171.088(73.211) | 561.283(1491.047) | 168.884(79.134) |
| Palmitamide | 3270.611(17777.123) | 1962.253(8761.313) | 841.234(872.513) | 870.961(440.581) |
| Pantoprazole | 898.088(8152.763) | 16270.845(118825.791) | 80.935(57.003) | 18187.304(84924.38) |

**Supplementary Table S1.** Sample mean and standard deviations of metabolites in ME/CFS patients, controls, and their subgroups.

| Metabolite | ME/CFS (n=106) | Control (n=91) | Male ME/CFS (n=31) | Male Control (n=22) |
|---|---|---|---|---|
| | mean (SD) | mean (SD) | mean (SD) | mean (SD) |
| Pantothenic acid | 10030.786(7968.504) | 8465.694(5284.919) | 9809.103(7101.967) | 7811.894(5772.281) |
| Penciclovir | 615.263(2184.586) | 280.151(80.439) | 308.135(294.352) | 265.322(73.998) |
| Phenylacetylglutamine | 31218.152(22442.043) | 34681.21(38768.962) | 27420.047(17582.075) | 30199.641(21041.081) |
| Phenylacetyl-L-glutamine | 14884.784(10785.176) | 16607.075(17719.928) | 13216.22(8436.638) | 14012.479(8810.894) |
| Phenylalanine | 55973.85(16422.42) | 58120.533(13302.726) | 59297.418(17189.557) | 56671.711(9124.259) |
| Pipecolic acid | 425.09(586.911) | 426.157(508.716) | 431.518(877.242) | 302.227(121.702) |
| Piperine | 5718.294(8337.056) | 7720.583(10963.746) | 6370.124(10079.811) | 10438.647(18064.942) |
| Prazepam | 288.862(1542.242) | 137.608(93.414) | 130.413(68.935) | 142.665(89.484) |
| Proline | 14486.098(6367.337) | 14482.522(5997.29) | 14659.894(4262.202) | 16833.462(6254.299) |
| Propionylcarnitine | 68344.79(23209.488) | 71041.55(26326.498) | 75008.185(23173.021) | 89574.261(34012.594) |
| Pyrantel | 372.256(111.716) | 1129.226(7044.272) | 342.836(86.954) | 377.493(85.041) |
| Pyridoxal | 1642.179(3991.664) | 946.29(438.171) | 982.176(620.401) | 884.975(226.448) |
| Pyridoxine; | 1022.88(4388.483) | 435.057(382.972) | 398.684(340.458) | 537.032(492.771) |
| Quetiapine | 369.86(1339.4) | 222.872(66.167) | 634.982(2421.885) | 218.621(45.738) |
| Quetiapine sulfoxide | 650.477(4802.113) | 136.563(42.808) | 1683.355(8722.602) | 136.332(28.26) |
| R-(-)-O-Desmethylvenlafaxine | 592.445(3164.213) | 410.727(1880.497) | 132.984(43.917) | 865.333(3450.186) |
| rac-4-Sulfoxypropranolol | 202.948(514.821) | 151.335(57.243) | 296.812(933.899) | 144.592(44.507) |
| Ranitidine | 2507.731(13468.136) | 205.008(53.061) | 3646.768(16884.028) | 200.279(51.505) |
| Ranitidine N-oxide | 713.593(4219.564) | 62.623(17.85) | 1462.938(7212.987) | 64.225(16.737) |
| Ranitidine-S-oxide | 213.634(515.841) | 132.212(90.781) | 372.742(891.461) | 127.054(63.583) |
| Scopoletin | 591.68(2509.742) | 178.452(414.272) | 366.114(1387.719) | 135.721(167.27) |
| SDMA | 9500.368(2703.971) | 10370.308(2765.049) | 8784.935(2463.706) | 9662.784(2984.059) |
| Serotonin | 18189.163(41222.489) | 14668.474(23684.703) | 27097.392(68656.868) | 18703.44(37627.043) |
| Ser-Tyr-Lys | 193.809(988.093) | 91.613(175.965) | 215.559(610.064) | 52.627(33.116) |
| Stachydrine | 563959.448(691819.57) | 612164.26(685817.817) | 509241.63(665509.328) | 483091.828(533375.278) |
| Sulfamethoxazole | 41.919(48.984) | 4622.278(43440.039) | 42.114(60.452) | 51.453(64.423) |
| Tapentadol-.beta.-D-glucuronide | 221.214(1394.74) | 127.85(761.689) | 638.067(2521.067) | 66.867(90.138) |
| Tauroursodeoxycholic acid | 2706.846(6111.732) | 1545.619(2380.983) | 2771.642(7467.22) | 2383.357(4356.385) |
| Telmisartan | 4699.894(41335.897) | 3856.6(30518.071) | 561.231(207.047) | 637.336(209.82) |
| Temazepam | 279.199(1061.022) | 153.782(115.023) | 131.371(55.789) | 174.98(204.841) |
| Testosterone | 3622.17(4140.278) | 4445.438(4912.789) | 3526.184(3539.749) | 3995.656(3766.305) |
| Theanine; | 1819.454(2742.739) | 1366.781(1424.619) | 1079.488(1156.863) | 974.3(970.032) |
| Theobromine | 43840.022(50239.28) | 50274.237(39008.573) | 46380.263(58049.085) | 38899.333(35511.114) |
| threo-Dihydrobupropion | 959.671(1263.114) | 899.034(1318.122) | 934.373(1403.161) | 901.716(1243.618) |
| Thr-Ile-Arg | 334.018(1580.162) | 174.397(391.703) | 413.161(1122.966) | 130.527(181.492) |
| Ticlopidine | 228.217(1202.183) | 52.712(180.862) | 113.045(523.142) | 76.163(217.135) |
| Topiramate | 883.512(3767.87) | 360.884(470.487) | 413.497(620.452) | 344.367(122.878) |
| Toradol | 356.538(2066.064) | 150.603(104.488) | 126.775(63.646) | 156.815(114.02) |
| trans-3'-Hydroxycotinine | 2268.615(10601.908) | 1686.299(5203.465) | 4673.366(18723.658) | 2696.057(7842.329) |
| Trazodone | 2672.469(13819.619) | 206.455(55.025) | 194.336(89.539) | 199.693(47.225) |
| Tri-2-ethylhexyl trimellitate | 6954.814(58249.323) | 2150.885(4550.116) | 20270.058(106147.287) | 870.01(683.104) |
| Trigonelline | 62593.319(128404.523) | 87574.388(113723.624) | 52780.293(86138.21) | 67552.649(79770.613) |
| Trileptal | 378.242(124.735) | 1737.665(13037.79) | 344.273(109.505) | 341.202(87.944) |
| Trimethoprim | 234.855(71.982) | 2512.23(21564.074) | 222.236(59.463) | 247.22(56.674) |
| Trimethylamine-N-oxide | 22496.762(34083.237) | 15784.326(13954.105) | 16615.634(19022.479) | 20017.457(21030.643) |
| Triptolide | 572.071(359.723) | 709.428(1341.517) | 487.897(172.496) | 562.15(187.996) |
| Tryptophan | 15719.222(4762.968) | 15591.457(4927.165) | 16208.348(4590.48) | 17268.208(4363.797) |
| Tyrosine | 6484.429(2346.848) | 6649.41(2125.156) | 6743.986(2343.539) | 6738.398(1629.213) |

**Supplementary Table S1.** Sample mean and standard deviations of metabolites in ME/CFS patients, controls, and their subgroups.

| Metabolite | ME/CFS (n=106) | Control (n=91) | Male ME/CFS (n=31) | Male Control (n=22) |
|---|---|---|---|---|
| | mean (SD) | mean (SD) | mean (SD) | mean (SD) |
| Urea | 128763.489(48161.656) | 135736.88(41188.088) | 128740.698(43060.26) | 146833.554(47651.964) |
| Urocanic acid; | 20103.21(25401.881) | 20113.659(16158.372) | 16277.246(8743.394) | 14697.149(5539.213) |
| Usnic acid | 6421.074(38109.236) | 5909.508(26905.817) | 14896.56(69459.594) | 3256.747(4748.431) |
| **Complex Lipids (CL)** | | | | |
| AC (10:0) | 14300.667(14495.722) | 14592.903(12632.497) | 14304.284(10224.154) | 12288.294(6811.885) |
| AC (10:1) | 7852.356(5661.808) | 8700.584(5463.897) | 8213.231(4726.31) | 8068.959(3635.011) |
| AC (12:0) | 6801.104(4854.843) | 7472.708(4034.354) | 7161.583(4461.614) | 7599.974(4309.822) |
| AC (12:1) | 5342.259(2975.805) | 5782.079(2495.997) | 5565.666(2940.647) | 5789.9(2652.02) |
| AC (14:1) | 7653.305(5643.002) | 7865.258(4525.933) | 7610.733(4887.504) | 7827.739(4591.044) |
| AC (14:2) | 4537.778(3261.62) | 5105.633(2880.79) | 4547.041(2451.901) | 5085.062(3183.104) |
| AC (16:0) | 10907.214(3516.947) | 10958.197(3206.646) | 12529.657(4062.832) | 12020.615(3789.727) |
| AC (18:0) | 4409.46(1523.433) | 4658.607(1398.628) | 4818.758(1531.186) | 5082.356(1672.108) |
| AC (18:1) | 15169.411(6563.264) | 14716.482(4589.592) | 16670.1(7066.335) | 15289.944(4633.514) |
| AC (18:2) | 9086.774(4258.756) | 9507.785(3723.066) | 10579.98(4543.181) | 10453.403(4548.995) |
| AC (8:0) | 5424.664(7636.621) | 5324.659(5489.183) | 5051.415(3488.917) | 4233.297(2281.491) |
| CE (14:0) | 483.105(328.13) | 550.26(299.965) | 398.792(237.464) | 498.482(320.195) |
| CE (16:0) | 6284.196(5322.107) | 5971.764(4027.806) | 5416.936(4207.504) | 5710.329(5150.448) |
| CE (16:1) | 44723.031(22333.233) | 45240.669(19130.581) | 40481.212(23152.954) | 46143.111(21363.158) |
| CE (18:0) | 3286.85(1793.581) | 3220.682(1502.875) | 3379.625(1881.442) | 3107.244(1303.777) |
| CE (18:1) | 32176.362(13035.556) | 33019.476(10977.599) | 29369.279(12439.441) | 30481.628(12865.088) |
| CE (18:2) | 360370.43(102450.354) | 395504.729(97524.484) | 346900.748(96985.256) | 353681.03(90607.595) |
| CE (18:3) | 13226.28(5296.356) | 16158.621(6188.509) | 12466.078(5278.865) | 16585.743(8232.324) |
| CE (20:2) | 7353.518(4456.574) | 8560.232(3729.559) | 7045.581(3784.452) | 7534.769(3979.312) |
| CE (20:3) | 65342.381(23690.393) | 72249.761(24364.326) | 61512.354(23895.339) | 66312.322(25346.564) |
| CE (20:4) | 760763.777(278873.716) | 769431.808(251914.063) | 726785.037(299621.635) | 790941.454(349981.263) |
| CE (20:5) | 17407.549(19917.453) | 12423.777(8256.741) | 13278.036(10259.439) | 11114.096(5374.717) |
| CE (22:2) | 686.936(506.637) | 714.698(520.067) | 712.355(524.213) | 615.637(431.426) |
| CE (22:6) | 132582.468(60465.938) | 114384.853(39714.847) | 116482.07(44378.479) | 101025.826(39811.332) |
| Ceramide (d34:1) - ESI(+) | 7991.308(2279.1) | 7701.808(1766.753) | 8731.306(2213.355) | 7272.043(1338.049) |
| Ceramide (d36:1) - ESI(+) | 5055.887(1639.5) | 4781.199(1246.336) | 5325.535(2032.287) | 4351.407(1238.385) |
| Ceramide (d38:1) - ESI(+) | 6696.998(1780.221) | 6981.944(1458.36) | 6775.581(2136.414) | 6560.467(1627.061) |
| Ceramide (d40:1) | 21686.844(5740.739) | 23489.285(4735.334) | 22330.201(7130.91) | 23771.933(5214.261) |
| Ceramide (d41:1) - ESI (+) | 6918.214(1876.08) | 7517.217(1659.079) | 6737.579(2156.907) | 7100.762(1464.764) |
| Ceramide (d42:1) - ESI (+) | 71508.239(18019.881) | 78197.952(14457.978) | 74834.162(21160.412) | 80924.605(15962.261) |
| Ceramide (d42:2) A - ESI (+) | 19273.129(4975.246) | 18114.439(3325.542) | 20767.903(5498.018) | 17942.265(3500.693) |
| Ceramide (d42:2) B - ESI (+) | 14981.161(4149.123) | 17174.124(4245.274) | 14761.732(4174.526) | 16805.176(4803.723) |
| Ceramide (d32:1) | 3595.683(1323.517) | 3755.63(977.806) | 3893.41(1681.704) | 3776.339(1277.115) |
| Ceramide (d33:1) | 2916.634(935.773) | 3016.705(807.728) | 2942.778(904.581) | 3034.769(747.137) |
| Ceramide (d34:0) | 4886.997(2053.157) | 4687(1716.579) | 5328.675(2353.493) | 5038.817(1756.365) |
| Ceramide (d34:1) - ESI(-) | 18074.154(4880.408) | 17694.653(3936.635) | 21181.657(5672.547) | 18781.979(4018.654) |
| Ceramide (d34:2) | 3094.181(847.888) | 3030.248(713.119) | 3178.801(912.631) | 2792.248(603.107) |
| Ceramide (d36:1) - ESI(-) | 1637.345(721.82) | 1539.002(608.946) | 1854.908(879.827) | 1612.868(611.104) |
| Ceramide (d38:1) - ESI(-) | 11993.384(4407.452) | 12156.053(3738.061) | 12800.634(5130.013) | 12538.413(4226.451) |
| Ceramide (d39:1) | 6299.483(2739.989) | 6979.275(2440.541) | 6345.302(3011) | 7142.867(2313.623) |
| Ceramide (d40:0) | 1765.181(1227.484) | 1564.678(901.085) | 1739.503(1153.507) | 1839.717(1212.59) |
| Ceramide (d40:2) | 6057.125(4179.286) | 6235.898(4315.18) | 6207.346(3283.457) | 6671.962(4263.527) |
| Ceramide (d41:1) - ESI(-) | 42041.811(15059.383) | 46380.145(13427.707) | 44430.29(17393.967) | 48813.011(13659.64) |
| Ceramide (d42:0) | 11948.609(7113.944) | 11294.104(4976.297) | 11854.292(6342.721) | 13352.996(6212.231) |

**Supplementary Table S1.** Sample mean and standard deviations of metabolites in ME/CFS patients, controls, and their subgroups.

| Metabolite | ME/CFS (n=106) | Control (n=91) | Male ME/CFS (n=31) | Male Control (n=22) |
|---|---|---|---|---|
| | mean (SD) | mean (SD) | mean (SD) | mean (SD) |
| Ceramide (d42:1) - ESI(-) | 136674.648(44023.767) | 149770.415(39576.94) | 149813.191(54068.674) | 161675.872(40939.304) |
| Ceramide (d42:2) A - ESI(-) | 49252.166(17827.101) | 47220.491(13180.04) | 57453.828(23064.842) | 51899.184(15072.465) |
| Ceramide (d42:2) B - ESI(-) | 37317.754(16468.604) | 35789.863(11255.565) | 43922.069(20084.189) | 38707.75(13373.343) |
| Ceramide (d43:1) | 2528.23(1370.14) | 2789.752(1196.521) | 2538.392(1401.305) | 3215.499(1522.097) |
| Ceramide (d44:1) | 846.904(541.153) | 822.512(399.283) | 1043.492(779.49) | 1004.782(528.397) |
| Cholesterol | 266952.992(63543.111) | 283483.733(67507.559) | 285997.23(62624.153) | 285744.734(56226.102) |
| DG (32:0) | 2029.32(748.253) | 1982.392(914.732) | 2400.192(821.835) | 2462.589(1230.819) |
| DG (32:1) | 5251.897(3457.449) | 4947.61(2880.584) | 6255.949(3763.84) | 6664.355(2949.793) |
| DG (34:1) | 1041.038(494.395) | 999.382(487.017) | 1243.903(483.107) | 1236.201(542.847) |
| DG (34:2) | 32528.392(13807.47) | 30539.842(13070.979) | 40152.283(14881.321) | 38721.383(12836.446) |
| DG (34:3) | 5185.782(3045.124) | 4572.847(2077.17) | 5797.716(3186.568) | 4848.035(1435.431) |
| DG (36:1) | 6350.769(3167.505) | 6121.406(3281.803) | 7752.728(3894.885) | 8344.677(3987.05) |
| DG (36:2) | 55205.662(20357.43) | 49677.788(17704.742) | 61293.329(20082.532) | 61274.639(20941.988) |
| DG (36:3) | 70925.903(27194.121) | 65340.022(22354.965) | 80757.457(25808.489) | 75739.062(24968.809) |
| DG (36:4) A | 16838.454(8807.583) | 16004.561(7408.839) | 19737.663(9707.678) | 17437.56(9172.99) |
| DG (36:4) B | 4169.876(3637.695) | 3516.148(2613.304) | 4830.172(4215.821) | 4879.996(2879.032) |
| DG (36:5) | 2206.156(1211.632) | 2086.253(902.788) | 2673.737(1340.431) | 2166.224(929.58) |
| DG (38:5) | 10566.304(4064.74) | 10089.518(3680.913) | 11961.559(4159.269) | 13525.081(3734.743) |
| DG (38:6) | 6169.284(3104.712) | 5407.065(2535.955) | 6892.129(3426.167) | 6867.647(2579.977) |
| FA (10:0) (capric acid) | 1604.441(1050.232) | 1407.005(1073.102) | 1711.573(1201.227) | 1464.917(1041.863) |
| FA (12:0) (lauric acid) | 8080.776(5386.504) | 8269.26(5625.99) | 8720.65(4467.889) | 8309.724(3064.579) |
| FA (14:0) (myristic acid) | 46315.443(13985.33) | 46285.78(10922.906) | 44376.55(13454.821) | 44448.11(9461.115) |
| FA (14:1) (physeteric acid) | 5158.99(3232.701) | 4681.581(1944.979) | 3902.687(2378.069) | 3845.25(2014.051) |
| FA (15:0) (pentadecylic acid) | 10544.8(2976.688) | 10266.457(2239.062) | 10785.814(3247.999) | 10590.402(2131.781) |
| FA (16:1) (palmitoleic acid) | 87362.569(45359.632) | 76933.028(35142.058) | 70747.149(48110.445) | 63019.939(30760.838) |
| FA (17:0) (margaric acid) | 22117.789(4474.162) | 21235.84(3887.35) | 22869.638(3991.423) | 21863.498(3317.04) |
| FA (18:1) (oleic acid) | 1454192.087(480456.365) | 1349284.059(394857.372) | 1306371.124(516584.708) | 1276084.64(395433.334) |
| FA (18:2) (linoleic acid) | 573453.977(197151.585) | 559405.212(162215.142) | 544856.4(233611.766) | 527836.688(188942.371) |
| FA (18:3) (linolenic acid) | 52408.007(25261.774) | 49568.813(16119.839) | 55831.729(36074.175) | 43953.469(17195.239) |
| FA (20:1) (eicosenoic acid) | 18300.811(5932.05) | 17239.139(4691.897) | 16904.974(5422.633) | 16890.944(5270.901) |
| FA (20:2) (eicosadienoic acid) | 9414.386(2783.824) | 9398.564(2656.075) | 8843.47(3032.234) | 8799.106(2886.371) |
| FA (20:3) (eicosatrienoic acid) | 2064.852(782.798) | 2106.53(696.017) | 1996.96(870.894) | 2343.651(765.021) |
| FA (20:3) (homo-gamma-linolenic acid) | 9019.322(2564.168) | 9232.52(2388.112) | 9058.827(2904.539) | 9812.517(2371.855) |
| FA (20:4) (arachidonic acid) | 43084.156(11307.632) | 45485.009(12124.954) | 47682.842(11690.665) | 53444.932(16109.434) |
| FA (20:5) (eicosapentaenoic acid) | 7210.305(8172.735) | 5310.154(4416.295) | 8078.364(7113.727) | 5541.467(2672.217) |
| FA (22:0) (behenic acid) | 4796.588(1883.561) | 4776.335(2235.782) | 5374.917(2287.009) | 5460.211(3363.174) |
| FA (22:6) (docosahexaenoic acid) | 35785.158(23581.553) | 31506.346(14537.092) | 34229.371(17390.242) | 30515.171(9235.535) |
| FA (24:0) (lignoceric acid) | 9002.74(3316.901) | 9144.823(3857.725) | 10363.172(4148.816) | 10735.801(5526.953) |
| FA (24:1) (nervonic acid) | 3404.095(1062.601) | 3216.118(871.277) | 3844.766(1415.216) | 3567.518(872.473) |
| Gal-Gal-Cer (d18:1/16:0)/Lactosylceramide (d18:1/16:0) | 37250.999(11809.088) | 37236.749(10987.775) | 39221.61(9988.658) | 37709.938(10657.275) |
| GlcCer (d38:1) | 3730.008(1287.974) | 3948.513(1065.45) | 3870.099(1460.396) | 3973.536(1169.578) |
| GlcCer (d40:1) - ESI(-) | 22073.89(7846.193) | 22731.523(6665.458) | 24207.449(7897.39) | 24139.711(7707.882) |
| GlcCer (d41:1) | 12088.153(4455.943) | 12743.895(4333.197) | 13142.311(5064.807) | 13889.254(5329.775) |
| GlcCer (d42:1) - ESI(-) | 25575.096(9070.161) | 26263.818(7929.532) | 29150.83(9409.836) | 29224.6(10141.294) |
| GlcCer (d42:2) - ESI(-) | 19888.456(7749.181) | 17973.465(6288.748) | 22833.056(8776.62) | 19931.828(8674.401) |
| GlcCer (d14:1(4E)/20:0(2OH)) | 6302.067(3492.07) | 6460.128(3125.454) | 5823.58(3024.058) | 6538.001(3091.453) |
| GlcCer (d34:1) | 7468.445(3751.415) | 8251.003(4419.557) | 8764.08(3833.514) | 8680.226(3690.26) |
| GlcCer (d40:1) - ESI(+) | 4100.143(1480.243) | 4234.688(1219.049) | 4181.685(1512.505) | 4245.413(1271.567) |

**Supplementary Table S1.** Sample mean and standard deviations of metabolites in ME/CFS patients, controls, and their subgroups.

| Metabolite | ME/CFS (n=106) | Control (n=91) | Male ME/CFS (n=31) | Male Control (n=22) |
|---|---|---|---|---|
| | mean (SD) | mean (SD) | mean (SD) | mean (SD) |
| GlcCer (d42:1) - ESI(+) | 17983.989(6028.82) | 18240.869(5395.163) | 19140.873(5484.236) | 19734.842(6590.648) |
| GlcCer (d42:2) - ESI(+) | 13729.159(5608.295) | 12051.673(4176.986) | 14573.512(5583.067) | 12112.285(5246.402) |
| Lactosylceramide (d18:1/24:1(15Z)) | 2755.733(1184.633) | 2537.724(815.608) | 2876.713(1454.383) | 2535.727(853.264) |
| LPC (14:0) - ESI(-) | 1816.114(1007.334) | 2045.03(925.463) | 1839.392(1099.666) | 2245.535(950.128) |
| LPC (16:0) - ESI(-) | 224092.722(72172.597) | 240419.333(71704.173) | 256854.719(83114.403) | 277064.698(77293.971) |
| LPC (16:1) - ESI(-) | 3256.883(1286.541) | 3465.425(1194.712) | 3321.019(1345.811) | 3822.279(1168.9) |
| LPC (17:1) | 2907.335(879.847) | 2996.073(1096.874) | 3037.551(915.572) | 3394.649(1191.042) |
| LPC (18:0) A - ESI(-) | 7418.389(3009.415) | 8077.783(2590.056) | 8306.305(3330.475) | 8938.356(2422.212) |
| LPC (18:0) B - ESI(-) | 99494.905(36134.374) | 108242.985(32910.725) | 112264.719(39798.755) | 120387.288(29907.213) |
| LPC (18:1) - ESI(-) | 47798.748(19375.801) | 51591.653(19211.667) | 54192.035(19358.272) | 58439.57(20648.312) |
| LPC (18:2) - ESI(-) | 60906.166(28020.858) | 71522.652(31446.609) | 73890.117(31899.667) | 75457.741(34146.137) |
| LPC (20:0) | 2602.173(1120.102) | 2667.429(891.909) | 2760.108(1224.082) | 2701.178(881.013) |
| LPC (20:1) - ESI(-) | 3013.633(1298.505) | 2917.096(1050.301) | 3170.145(1587.396) | 3030.706(913.737) |
| LPC (20:1) - ESI(+) | 713.456(419.647) | 654.494(268.413) | 825.423(510.424) | 699.765(279.592) |
| LPC (20:2) - ESI(-) | 4059.133(1281.292) | 4450.809(1469.606) | 4413.372(1573.531) | 4565.117(1281.303) |
| LPC (20:2) - ESI(+) | 518.937(285.076) | 598.846(314.265) | 590.84(340.026) | 621.246(301.34) |
| LPC (20:3) - ESI(-) | 3582.326(1705.034) | 4021.253(1564.565) | 4081.79(1899.384) | 4806.142(1477.554) |
| LPC (22:4) | 1706.729(976.366) | 1894.898(1004.387) | 2251.648(1058.406) | 2604.942(1358.714) |
| LPC (22:5) - ESI(-) | 675.167(447.702) | 664.823(383.062) | 886.715(554.24) | 791.686(377.634) |
| LPC (22:6) | 18296.637(8748.95) | 17120.89(7390.865) | 19280.755(8703.872) | 17320.179(5402.133) |
| LPC (o-16:0) | 7726.959(3135.329) | 8366.675(3289.673) | 9562.972(3966.252) | 9545.73(3480.232) |
| LPC (p-16:0)/LPC (o-16:1) | 10752.235(4110.82) | 11778.247(4172.532) | 12895.152(4859.04) | 13555.48(4879.159) |
| LPC (p-18:0)/LPC (o-18:1) | 5609.615(1996.592) | 5649.593(2083.157) | 6312.961(2252.03) | 6302.042(1780.572) |
| LPC (14:0) - ESI(+) | 12225.274(5163.537) | 14568.798(5062.405) | 11977.356(5718.762) | 15445.385(5165.213) |
| LPC (15:0) | 12084.127(4819.914) | 12943.535(4361.419) | 12780.381(5698.997) | 13037.637(4366.256) |
| LPC (16:0) - ESI(+) | 1926106.937(412946.468) | 2025455.264(509547.986) | 2122729.714(414415.433) | 2257025.766(520385.84) |
| LPC (16:1) - ESI(+) | 18357.864(6537.325) | 19256.04(6089.078) | 18979.953(9013.425) | 20068.018(5828.219) |
| LPC (18:0) - ESI(+) | 597884.618(199525.827) | 642665.246(221367.761) | 652830.354(212542.515) | 693367.052(174842.328) |
| LPC (18:1) - ESI(+) | 267553.679(104625.2) | 280752.212(113394.873) | 308282.753(125723.885) | 329632.285(140448.722) |
| LPC (18:2) - ESI(+) | 453109.903(200728.084) | 547483.468(267853.354) | 553742.399(224834.804) | 603839.746(318018.732) |
| LPC (18:3) | 3781.745(1556.029) | 4762.878(2448.244) | 4255.782(1792.563) | 5519.175(3871.826) |
| LPC (20:3) - ESI(+) | 18658.988(7504.447) | 20830.978(7040.033) | 21810.228(8898.33) | 24667.198(9011.821) |
| LPC (20:4) | 92588.56(39171.901) | 99801.287(39143.693) | 115640.286(50009.696) | 126682.589(53164.432) |
| LPC (20:5) | 8554.056(10259.538) | 6693.724(5097.151) | 8436.341(7975.092) | 6882.284(3136.558) |
| LPC (22:5) - ESI(+) | 4133.499(2034.537) | 4006.158(1626.825) | 5081.496(2307.922) | 4780.993(1730.992) |
| LPE (16:0) | 3007.892(1492.945) | 3345.916(1755.537) | 3443.824(1727.622) | 3667.836(1958.6) |
| LPE (18:2) - ESI(-) | 4521.977(2417.216) | 5517.915(3099.215) | 5301.279(2967.128) | 5090.353(2770.4) |
| LPE (20:4) - ESI(-) | 3411.332(1579.491) | 3835.167(1358.006) | 3821.613(1774.902) | 4009.519(1196.108) |
| LPE (22:6) | 2511.314(1103.103) | 2584.869(1142.272) | 2506.121(947.775) | 2407.111(1062.591) |
| LPE (18:0) | 2976.4(935.468) | 3330.697(987.882) | 2975.619(1000.622) | 3457.5(835.551) |
| LPE (18:2) - ESI(+) | 3940.782(1604.285) | 4738.299(2407.019) | 4513.971(1624.136) | 4445.501(1993.594) |
| LPE (20:4) - ESI(+) | 3205.181(1241.82) | 3617.274(1320.76) | 3383.978(1267.399) | 3791.41(1594.759) |
| PC (16:0/9:0(CHO)) | 4327.064(2722.879) | 4282.307(2603.353) | 4968.504(3315.67) | 4910.879(3014.302) |
| PC (32:0) - ESI(-) | 14490.961(3964.7) | 14558.291(3478.332) | 15263.408(4162.383) | 14700.287(3404.682) |
| PC (32:1) - ESI(-) | 14447.376(7522.552) | 14662.134(6394.392) | 12758.462(7105.05) | 14728.451(7297.961) |
| PC (32:2) - ESI(-) | 13463.361(6508.264) | 16204.369(5624.383) | 12033.643(7553.981) | 14707.656(6218.337) |
| PC (33:1) - ESI(-) | 2353.684(1083.204) | 2374.083(853.644) | 2131.3(1012.513) | 2414.131(939.724) |
| PC (33:2) - ESI(-) | 3860.198(1605.855) | 4336.55(1397.757) | 3368.212(1547.831) | 3815.359(1300.496) |

**Supplementary Table S1.** Sample mean and standard deviations of metabolites in ME/CFS patients, controls, and their subgroups.

| Metabolite | ME/CFS (n=106) | Control (n=91) | Male ME/CFS (n=31) | Male Control (n=22) |
|---|---|---|---|---|
| | mean (SD) | mean (SD) | mean (SD) | mean (SD) |
| PC (34:0) - ESI(-) | 8251.523(2184.754) | 8469.609(1811.655) | 8733.18(2533.887) | 8483.927(1845.942) |
| PC (34:2) - ESI(-) | 240630.097(60351.691) | 258867.547(56457.148) | 247088.946(65989.688) | 252285.178(54764.773) |
| PC (34:3) | 16244.273(6359.067) | 18088.554(6094.852) | 15330.272(6007.704) | 16136.002(5630.699) |
| PC (34:4) - ESI(-) | 3988.556(2296.794) | 4598.624(1731.583) | 3330.478(2005.054) | 4458.283(1726.963) |
| PC (35:1) - ESI(-) | 3093.959(1098.086) | 3051.722(977.026) | 3043.764(1081.863) | 3250.46(1021.62) |
| PC (35:2) | 9207.837(2691.891) | 9716.991(2633.897) | 9532.595(2656.684) | 8872.848(2574.126) |
| PC (35:4) - ESI(-) | 2567.664(1136.268) | 2622.666(936.586) | 2492.677(1048.406) | 2743.646(1351.27) |
| PC (36:1) - ESI(-) | 60700.134(17787.723) | 61602.252(15986.223) | 61324.941(17578.546) | 62040.398(15296.844) |
| PC (36:3) A - ESI(-) | 78001.315(26248.654) | 80601.946(20165.252) | 81493.858(28847.298) | 82092.84(21459.655) |
| PC (36:3) B - ESI(-) | 80122.729(25240.712) | 84455.963(19138.885) | 81453.529(26862.449) | 86689.976(19139.009) |
| PC (36:4) A - ESI(-) | 50001.555(19418.961) | 58881.412(19057.435) | 52267.702(21480.43) | 52859.354(16963.772) |
| PC (36:4) B - ESI(-) | 165608.498(45603.524) | 172562.505(36378.869) | 174408.792(49658.16) | 179577.946(38130.404) |
| PC (36:5) A | 2138.31(1339.816) | 2567.216(1605.787) | 2189.983(1541.952) | 1814.926(1203.75) |
| PC (36:5) B | 41857.83(35378.719) | 33407.87(22725.882) | 47045.93(43693.834) | 34117.138(17993.151) |
| PC (37:2) - ESI(-) | 1427.867(571.478) | 1549.892(656.208) | 1408.964(562.641) | 1336.555(526.791) |
| PC (37:3) | 19429.669(8642.077) | 20766.774(7555.42) | 18098.801(8761.691) | 17852.572(6955.697) |
| PC (37:4) - ESI(-) | 3610.43(1262.482) | 3600.739(1164.94) | 3865.914(1175.379) | 3709.906(1129.303) |
| PC (38:2) | 4724.469(1540.569) | 4816.371(1178.537) | 4580.92(1544.406) | 4438.677(996.639) |
| PC (38:3) - ESI(-) | 29648.655(10260.002) | 31432.948(7745.999) | 29624.875(10831.337) | 33004.923(8368.738) |
| PC (38:4) A - ESI(-) | 57986.619(17566.019) | 60883.346(16516.825) | 60375.971(17898.448) | 62780.612(14774.114) |
| PC (38:5) A - ESI(-) | 19182.481(6029.043) | 19166.914(4819.422) | 20412.89(6733.556) | 19816.339(4946.775) |
| PC (38:5) B - ESI(-) | 12329.808(8869.88) | 10448.295(4369.029) | 12975.729(10525.834) | 10265.063(3529.805) |
| PC (38:6) - ESI(-) | 75564.438(34561.384) | 68827.018(26086.774) | 75619.639(38181.774) | 69579.548(23778.393) |
| PC (39:6) | 18191.534(9715.309) | 16532.441(6454.175) | 13459.844(5802.603) | 13206.209(4768.915) |
| PC (40:4) - ESI(-) | 3651.767(1838.864) | 3888.447(1301.818) | 3909.143(1715.993) | 4244.068(1400.298) |
| PC (40:5) A - ESI(-) | 4364.652(1888.948) | 4187.943(1433.041) | 4770.531(2049.396) | 4279.36(1497.875) |
| PC (40:5) B - ESI(-) | 1949.867(1019.491) | 2160.184(912.295) | 1910.966(896.617) | 2479.194(1218.116) |
| PC (40:6) B | 20657.541(8269.246) | 18554.405(6543.511) | 21040.215(9670.584) | 18713.955(6184.276) |
| PC (40:7) - ESI(-) | 2669.718(1393.762) | 2433.114(925.635) | 2625.831(1432.89) | 2313.846(972.287) |
| PC (40:8) - ESI(-) | 2630.788(1139.643) | 2734.932(900.85) | 2878.238(1371.79) | 2882.618(898.713) |
| PC (42:5) | 3139.766(1127.172) | 3024.694(1140.962) | 2627.191(1248.213) | 2509.332(843.894) |
| PC (42:6) | 2237.693(1216.735) | 2297.879(1311.502) | 1853.339(1040.649) | 2198.375(1271.775) |
| PC (o-32:0) - ESI(-) | 2806.219(868.036) | 2847.252(794.141) | 3052.76(866.282) | 2871.266(762.942) |
| PC (p-32:0)/PC (o-32:1) - ESI(-) | 2338.644(927.886) | 2365.854(804.708) | 2679.84(1050.529) | 2490.279(1071.344) |
| PC (p-32:0)/PC (o-32:1) - ESI(+) | 29152.037(7872.188) | 30771.998(7341.47) | 30028.365(9213.253) | 28568.812(7803.221) |
| PC (p-34:1)/PC (o-34:2) A | 4520.84(2258.236) | 5300.943(2014.065) | 4757.993(2102.423) | 4859.606(1626.867) |
| PC (p-34:1)/PC (o-34:2) B | 2874.162(1241.363) | 2975.868(1271.407) | 2955.203(1307.817) | 2729.62(1477.495) |
| PC (p-34:2)/PC (o-34:3) - ESI(-) | 13080.94(4655.151) | 15147.291(4618.811) | 13538.65(4139.591) | 14391.877(3967.502) |
| PC (p-36:1)/PC (o-36:2) | 1579.393(1039.658) | 1842.803(957.813) | 1707.593(920.629) | 1673.072(739.313) |
| PC (p-36:3)/PC (o-36:4) - ESI(-) | 21432.678(7961.51) | 23318.595(6892.306) | 24688.357(7510.221) | 26166.647(8952.3) |
| PC (p-36:3)/PC (o-36:4) - ESI(+) | 227889.598(84207.01) | 254677.838(68833.936) | 250126.868(92689.597) | 268982.446(76530.895) |
| PC (p-36:4)/PC (o-36:5) - ESI(-) | 12136.93(5243.46) | 13380.29(4590.962) | 14393.873(6063.86) | 15022.239(5522.478) |
| PC (p-38:3)/PC (o-38:4) - ESI(-) | 7180.199(2581.694) | 8011.26(2278.842) | 7922.095(2620.948) | 8636.903(2587.55) |
| PC (p-38:4)/PC (o-38:5) A | 16408.903(5205.505) | 17597.828(4431.343) | 17820.582(4259.592) | 18701.309(5190.175) |
| PC (p-38:4)/PC (o-38:5) B | 2559.33(1097.47) | 2769.763(1152.827) | 2808.843(1106.312) | 3260.105(1412.566) |
| PC (p-38:5)/PC (o-38:6) | 1297.362(880.416) | 1291.415(783.351) | 1341.621(1148.57) | 1306.021(628.98) |
| PC (p-40:1)/PC (o-40:2) | 1485.155(608.109) | 1292.088(473.435) | 1462.481(553.862) | 1236.145(464.308) |
| PC (p-40:3)/PC (o-40:4) | 1750.816(685.913) | 1868.47(638.755) | 1850.389(704.887) | 2043.343(619.095) |

**Supplementary Table S1.** Sample mean and standard deviations of metabolites in ME/CFS patients, controls, and their subgroups.

| Metabolite | ME/CFS (n=106) | Control (n=91) | Male ME/CFS (n=31) | Male Control (n=22) |
|---|---|---|---|---|
| | mean (SD) | mean (SD) | mean (SD) | mean (SD) |
| PC (p-40:4)/PC (o-40:5) - ESI(-) | 1855.515(641.097) | 1784.499(529.196) | 1949.912(581.6) | 1838.42(505.562) |
| PC (p-40:7)/PC (o-40:8) | 12784.77(9315.863) | 11294.973(6789.332) | 9573.21(9944.436) | 7531.17(6686.616) |
| PC (p-42:3)/PC (o-42:4) | 4785.842(1784.509) | 5178.399(1630.148) | 4952.621(1854.587) | 5342.838(1997.141) |
| PC (p-42:4)/PC (o-42:5) - ESI(-) | 2268.307(909.076) | 2270.498(870.57) | 2448.033(832.681) | 2364.737(949.186) |
| PC (p-42:5)/PC (o-42:6) | 618.687(355.575) | 610.366(274.685) | 646.286(300.455) | 704.809(279.735) |
| PC (p-44:4)/PC (o-44:5) - ESI(-) | 2845.694(1045.311) | 2893.365(908.463) | 3027.265(1099.475) | 3015.819(836.508) |
| PC (p-44:5)/PC (o-44:6) | 3310.494(1275.999) | 3174.484(993.019) | 3138.119(1105.716) | 3030.243(1076.533) |
| PC 34:4e | 6443.081(2515.863) | 7999.075(2933.128) | 6265.46(2786.167) | 6864.822(2784.41) |
| PC 38:7e | 16964.282(13526.498) | 15342.968(9446.371) | 16544.441(13664.894) | 13824.238(5068.143) |
| PC 40:5e | 12165.024(3482.196) | 12677.876(3009.874) | 12156.263(2800.976) | 12946.278(3876.998) |
| PC 40:6e | 22316.948(7430.313) | 21474.846(6189.986) | 22911.69(6865.962) | 21858.337(7312.84) |
| PC (28:0) | 3246.565(4662.849) | 3239.16(2348.658) | 2319.456(2047.457) | 3419.203(2870.489) |
| PC (30:0) | 158777.616(83572.094) | 179096.713(65617.314) | 134527.229(57552.582) | 180921.663(83317.035) |
| PC (31:0) | 10922.351(4521.628) | 11042.588(3052.892) | 9322.83(3444.162) | 9511.503(2302.278) |
| PC (31:1) | 1677.788(1455.465) | 1660.939(915.401) | 1190.067(862.691) | 1324.367(711.207) |
| PC (32:0) - ESI(+) | 256268.716(58170.605) | 250135.791(53964.518) | 252693.482(58815.308) | 242994.15(67734.773) |
| PC (32:1) - ESI(+) | 239846.485(124170.713) | 249536.641(113064.066) | 199894.088(94497.866) | 233324.489(96006.991) |
| PC (32:2) - ESI(+) | 56756.644(28246.729) | 67703.831(25937.146) | 45747.251(30984.152) | 52825.941(23680.951) |
| PC (33:0) | 10952.596(5641.864) | 11592.978(3878.492) | 9408.237(4832.541) | 10312.507(3477.739) |
| PC (33:1) - ESI(+) | 53543.632(20928.978) | 53553.558(13337.174) | 44982.592(16376.287) | 48494.832(11798.772) |
| PC (33:2) - ESI(+) | 78531.369(27527.116) | 87270.676(25504.3) | 66386.357(22506.876) | 66179.171(17944.435) |
| PC (34:0) - ESI(+) | 56673.124(14226.497) | 56575.078(11243.201) | 55930.398(16856.304) | 54402.796(12049.128) |
| PC (34:1) | 4013338.964(720952.007) | 4067426.422(727466.087) | 3807829.103(772533.633) | 4051360.726(912477.724) |
| PC (34:2) - ESI(+) | 8298954.78(1436737.757) | 8880671.15(1333757.007) | 8009035.373(1191638.206) | 8068560.636(1188330.124) |
| PC (34:3) A | 97711.278(31702.813) | 106631.972(39043.209) | 84529.885(34636.865) | 89091.807(39417.08) |
| PC (34:3) B | 108771.059(36242.03) | 128990.918(46188.814) | 91164.199(27797.142) | 101396.149(38586.988) |
| PC (34:3) C | 86598.612(39702.386) | 108697.965(49708.994) | 75951.137(21974.678) | 97984.941(50767.182) |
| PC (34:4) - ESI(+) | 17218.357(9083.701) | 20014.966(7594.249) | 13453.933(6967.576) | 17417.677(7336.924) |
| PC (35:1) - ESI(+) | 46139.315(13080.522) | 45638.931(9232.36) | 41459.078(10896.604) | 42883.759(9581.456) |
| PC (35:2) A | 109536.799(36351.893) | 103212.896(30675.48) | 94027.949(31874.332) | 100085.598(23028.225) |
| PC (35:2) B | 131176.614(32124.982) | 137458.977(30973.043) | 119065.848(31085.946) | 114270.85(23658.334) |
| PC (35:3) | 40372.126(10564.132) | 44343.372(11336.707) | 35865.887(9999.311) | 36662.028(9497.255) |
| PC (35:4) - ESI(+) | 22778.31(8961.192) | 23974.006(6756.722) | 19079.655(6980.574) | 20470.702(6462.17) |
| PC (36:1) - ESI(+) | 656088.196(185659.526) | 660392.494(148989.452) | 587404.023(154102.201) | 627376.228(128997.811) |
| PC (36:2) | 4529060.48(909760.507) | 4996033.975(881985.183) | 4211692.931(903186.615) | 4327926.764(476276.167) |
| PC (36:3) A - ESI(+) | 1277000.345(445850.9) | 1366182.069(370360.325) | 1204527.783(516992.597) | 1212320.94(306902.701) |
| PC (36:3) B - ESI(+) | 1639032.954(502617.113) | 1716418.049(393778.108) | 1595831.492(520981.85) | 1592535.179(276748.936) |
| PC (36:4) A - ESI(+) | 344228.244(141889.724) | 433150.27(175314.159) | 324935.514(157371.936) | 345149.151(130634.465) |
| PC (36:4) B - ESI(+) | 254789.338(417790.093) | 206974.331(69274.833) | 168655.141(82504.342) | 190887.131(54600.311) |
| PC (36:4) C - ESI(+) | 3945315.162(980469.466) | 4037185.749(932394.375) | 3770041.129(1046223.193) | 4023980.133(1042196.2) |
| PC (36:5) C | 63676.772(293144.05) | 37777.207(75598.623) | 35961.874(82637.482) | 42060.718(90105.09) |
| PC (36:5) D | 466335.725(583640.353) | 328794.138(256617.495) | 328646.617(282542.953) | 263135.658(107530.626) |
| PC (36:5)A | 10087.21(5302.765) | 12082.737(6355.171) | 8991.784(5467.663) | 8535.056(4024.666) |
| PC (36:6) | 7517.927(4687.721) | 7514.303(3539.182) | 5072.96(3119.115) | 5365.199(1768.116) |
| PC (37:2) - ESI(+) | 12948.098(3497.278) | 14134.187(3647.086) | 11891.289(3512.971) | 12085.972(2300.68) |
| PC (37:4) - ESI(+) | 51981.159(19322.358) | 49132.105(16397.252) | 44294.212(16164.247) | 49003.07(18357.963) |
| PC (37:5) | 36908.954(34432.55) | 28967.669(14940.073) | 27588.92(17283.673) | 24327.094(8409.348) |
| PC (37:6) | 9891.772(5465.158) | 8696.041(4047.719) | 6890.365(2953.421) | 6246.927(2456.317) |

**Supplementary Table S1.** Sample mean and standard deviations of metabolites in ME/CFS patients, controls, and their subgroups.

| Metabolite | ME/CFS (n=106) | Control (n=91) | Male ME/CFS (n=31) | Male Control (n=22) |
|---|---|---|---|---|
| | mean (SD) | mean (SD) | mean (SD) | mean (SD) |
| PC (38:3) - ESI(+) | 545962.223(199887.214) | 581406.422(170804.445) | 502874.429(188115.354) | 563752.727(162801.855) |
| PC (38:4) A - ESI(+) | 129827.943(52570.47) | 130649.431(33178.708) | 113377.194(37599.238) | 117140.224(28716.98) |
| PC (38:4) B - ESI(+) | 138945.853(49865.612) | 150899.97(43419.782) | 137674.029(54084.911) | 158478.41(46996.215) |
| PC (38:4) C - ESI(+) | 2313829.998(565304.201) | 2391882.015(574272.595) | 2178463.964(599975.838) | 2310328.11(606482.831) |
| PC (38:5) A | 732724.247(176405.92) | 721011.877(165292.616) | 692510.741(177868.353) | 703688.662(171227.462) |
| PC (38:5) B - ESI(+) | 385628.57(330057.265) | 324021.431(136699.566) | 297899.839(144961.97) | 282186.016(71340.288) |
| PC (38:6) A - ESI(+) | 122703.865(38145.153) | 135842.09(35120.916) | 110512.355(34062.971) | 124100.415(31890.776) |
| PC (38:6) B - ESI(+) | 3683483.214(1066200.996) | 3373471.275(825187.655) | 3308133.746(802249.627) | 3011420.074(682021.451) |
| PC (38:7) | 9067.129(4522.378) | 8494.519(3671.831) | 6643.623(3241.473) | 6218.005(2668.734) |
| PC (39:4) | 4951.364(1392.551) | 4980.401(1209.519) | 4440.55(1217.731) | 4643.705(1217.911) |
| PC (40:4) - ESI(+) | 40710.749(17107.156) | 43768.486(14761.039) | 42663.302(16356.582) | 48562.457(17576.609) |
| PC (40:5) A - ESI(+) | 160984.496(51414.716) | 153215.298(48927.121) | 153701.168(45022.035) | 150517.483(35932.021) |
| PC (40:5) B - ESI(+) | 39290.673(16831.51) | 43514.802(16159.023) | 38473.161(15894.973) | 44025.445(17787.819) |
| PC (40:6) A | 26324.155(8352.257) | 26286.775(7120.512) | 23634.728(8121.937) | 23764.161(6603.153) |
| PC (40:6)B | 523343.738(235330.416) | 448154.607(162581.361) | 427810.229(142555.352) | 385657.136(132065.673) |
| PC (40:7) A - ESI(+) | 14123.954(4521.843) | 15295.102(4113.716) | 13952.454(5466.208) | 15028.736(3957.139) |
| PC (40:7) B - ESI(+) | 294687.493(123648.862) | 260986.662(85720.284) | 257320.328(117772.802) | 217436.412(72618.439) |
| PC (40:8) - ESI(+) | 15239.14(4848.028) | 15297.801(4079.753) | 14476.135(6122.24) | 13978.291(3019.574) |
| PC (42:10) | 2726.931(1374.175) | 2509.831(1360.724) | 2531.617(1415.798) | 2354.819(927.021) |
| PC (o-32:0) - ESI(+) | 33066.939(8835.009) | 34463.696(8408.236) | 34119.797(8865.124) | 32338.938(8404.113) |
| PC (o-34:0) | 4253.752(1544.24) | 4625.498(1473.237) | 4251.794(1703.095) | 4099.673(1209.919) |
| PC (p-32:1)/PC (o-32:2) | 7732.065(2525.44) | 8442.017(2078.799) | 7910.504(3383.972) | 8191.083(2087.48) |
| PC (p-34:1)/PC (o-34:2) | 90514.308(41518.544) | 112732.145(42037.713) | 90884.196(35928.824) | 95158.27(29994.724) |
| PC (p-34:2)/PC (o-34:3) - ESI(+) | 134516.098(48579.028) | 167161.547(53529.226) | 130014.23(47822.804) | 147005.911(49558.404) |
| PC (p-36:1)/PC (o-36:2) B | 4576.41(1860.937) | 4990.913(1761.819) | 4307.895(1837.538) | 4197.481(1642.081) |
| PC (p-36:2)/PC (o-36:3) | 18401.743(7473.236) | 22260.671(8493.763) | 17314.501(7146.789) | 19082.113(7114.945) |
| PC (p-36:4)/PC (o-36:5) - ESI(+) | 222255.821(73873.841) | 244403.223(62140.636) | 242439.639(89502.675) | 258183.564(78912.006) |
| PC (p-36:5)/PC (o-36:6) | 43163.98(47010.214) | 35793.982(29043.548) | 43083.765(46051.211) | 35923.993(21264.091) |
| PC (p-38:3)/PC (o-38:4) A - ESI(+) | 23415.263(7747.924) | 26268.221(7020.386) | 24102.249(7465.869) | 28129.553(9009.63) |
| PC (p-38:3)/PC (o-38:4) B - ESI(+) | 60681.832(24417.325) | 68924.225(21887.47) | 63797.576(23315.526) | 71084.075(22755.644) |
| PC (p-38:4)/PC (o-38:5) A | 244736.068(73932.864) | 253186.358(52211.505) | 253198.973(75445.49) | 259563.862(64014.349) |
| PC (p-38:4)/PC (o-38:5) B | 46348.51(17350.154) | 52251.201(14810.562) | 47627.623(16741.534) | 53717.086(17796.507) |
| PC (p-38:5)/PC (o-38:6) A | 47602.588(24906.045) | 44907.468(17837.495) | 46233.794(27430.939) | 40791.817(15837.344) |
| PC (p-38:5)/PC (o-38:6) B | 60693.148(18465.69) | 63136.031(15203.868) | 58926.741(20171.535) | 59314.065(18371.793) |
| PC (p-38:6)/PC (o-38:7) | 38202.501(19743.59) | 36368.564(13760.086) | 36619.184(20463.261) | 33534.038(11628.77) |
| PC (p-40:4)/PC (o-40:5) - ESI(+) | 17184.561(5389.491) | 17672.249(4621.409) | 17410.96(4549.737) | 18075.452(5491.996) |
| PC (p-40:6)/PC (o-40:7) A | 40200.774(19957.817) | 35888.168(13738.726) | 36954.859(22360.938) | 32062.44(11105.695) |
| PC (p-40:6)/PC (o-40:7) B | 13510.106(6889.457) | 13100.064(5544.826) | 12405.966(6481.044) | 11203.745(4502.511) |
| PC (p-42:4)/PC (o-42:5) - ESI(+) | 12900.718(4496.163) | 12896.037(3917.285) | 12754.45(3435.607) | 12938.521(5007.842) |
| PC (p-42:5)/PC (o-42:6) A | 10237.461(3726.694) | 10713.657(3758.542) | 10172.91(3767.066) | 10947.237(4814.118) |
| PC (p-42:5)/PC (o-42:6) B | 3974.188(1415.292) | 3956.085(1237.45) | 3733.472(1142.554) | 3479.506(1252.076) |
| PC (p-44:4)/PC (o-44:5) - ESI(+) | 14453.858(5455.893) | 14223.731(4628.827) | 14914.59(5257.409) | 13990.202(5447.096) |
| PE (34:1) | 1935.048(1193.975) | 1997.27(1266.931) | 1922.526(1262.798) | 2088.749(1260.706) |
| PE (34:2) - ESI(-) | 2982.653(1876.385) | 3405.593(2151.45) | 2867.947(2153.222) | 3232.022(2283.173) |
| PE (36:1) - ESI(-) | 418.862(181.396) | 445.435(181.825) | 394.47(193.221) | 476.773(203.336) |
| PE (36:2) | 7905.774(4135.38) | 8803.532(4225.603) | 8328.494(5401.93) | 8605.309(4699.84) |
| PE (36:3) | 2026.786(1248.378) | 2279.738(1430.26) | 2201.323(1753.452) | 2174.804(1400.564) |
| PE (36:4) - ESI(-) | 2941.499(1644.752) | 3164.472(1950.982) | 2727.181(1659.941) | 3275.078(1994.651) |

**Supplementary Table S1.** Sample mean and standard deviations of metabolites in ME/CFS patients, controls, and their subgroups.

| Metabolite | ME/CFS (n=106) | Control (n=91) | Male ME/CFS (n=31) | Male Control (n=22) |
|---|---|---|---|---|
| | mean (SD) | mean (SD) | mean (SD) | mean (SD) |
| PE (38:2) | 629.602(222.186) | 682.787(206.229) | 625.315(210.258) | 726.012(236.051) |
| PE (38:6) - ESI(-) | 7437.759(5142.646) | 7356.489(4624.489) | 6039.64(4500.327) | 7125.711(4187.612) |
| PE (p-34:1)/PE (o-34:2) - ESI(-) | 2025.04(785.489) | 2051.91(695.945) | 2150.254(890.571) | 2026.74(746.259) |
| PE (p-34:2)/PE (o-34:3) | 3040.334(1576.283) | 3609.46(1747.103) | 3425.067(1818.788) | 3441.15(1343.908) |
| PE (p-36:1)/PE (o-36:2) - ESI(-) | 2760.373(1320.05) | 3068.12(1351.566) | 3064.044(1403.021) | 3267.766(1574.172) |
| PE (p-36:2)/PE (o-36:3) - ESI(-) | 5957.994(2773.012) | 6972.062(2936.918) | 6614.407(3104.076) | 6917.904(2378.957) |
| PE (p-36:4)/PE (o-36:5) - ESI(-) | 15261.909(9626.399) | 15928.908(8581.977) | 18489.037(12771.445) | 17905.612(10235.869) |
| PE (p-36:5)/PE (o-36:6) | 2102.097(2962.828) | 1485.236(2041.939) | 2863.162(4197.256) | 1557.732(1341.719) |
| PE (p-38:3)/PE (o-38:4) | 1340.345(687.934) | 1513.805(782.147) | 1479.584(725.327) | 1687.613(809.239) |
| PE (p-38:5)/PE (o-38:6) - ESI(-) | 11786.825(6628.023) | 12149.489(5341.659) | 13603.388(8523.127) | 12933.022(6132.057) |
| PE (p-38:6)/PE (o-38:7) | 7532.92(4292.66) | 7188.888(3364.912) | 8157.198(5177.597) | 7545.245(2881.175) |
| PE (p-40:4)/PE (o-40:5) | 3399.555(1897.801) | 3410.027(1799.437) | 3628.141(2106.633) | 3661.55(2357.67) |
| PE (34:2) - ESI(+) | 8757.929(6410.569) | 9879.682(7371.827) | 7632.324(5605.642) | 9599.64(5858.139) |
| PE (36:1) - ESI(+) | 3379.71(1611.842) | 3316.561(1592.325) | 3662.52(1957.017) | 3571.172(1895.091) |
| PE (36:4) - ESI(+) | 14739.008(7468.332) | 15255.178(7705.241) | 12270.933(6672.071) | 14447.752(7358.067) |
| PE (38:4) | 39570.697(19154.594) | 40390.086(15772.1) | 35797.097(19983.436) | 37463.39(16790.23) |
| PE (38:6) - ESI(+) | 20257.66(12022.688) | 19884.696(11866.824) | 13208.306(6582.466) | 16869.926(9428.463) |
| PE (p-34:1)/PE (o-34:2) - ESI(+) | 2742.254(1098.637) | 2862.174(974.659) | 2911.341(1333.048) | 2823.109(1036.114) |
| PE (p-36:1)/PE (o-36:2) - ESI(+) | 2454.413(1024.933) | 2762.367(992.448) | 2564.311(1190.111) | 2713.77(1140.048) |
| PE (p-36:2)/PE (o-36:3) - ESI(+) | 8991.903(3876.705) | 11041.406(4021.531) | 9494.206(4636.704) | 9793.668(3052.8) |
| PE (p-36:4)/PE (o-36:5) - ESI(+) | 29345.071(18055.185) | 30532.635(14508.762) | 35383.013(25322.79) | 31556.53(15854.325) |
| PE (p-38:4)/PE (o-38:5) | 42614.164(20321.576) | 46103.9(17411.189) | 46972.987(24498.228) | 46554.029(19964.508) |
| PE (p-38:5)/PE (o-38:6) - ESI(+) | 24747.834(11095.999) | 26262.641(9101.812) | 27610.681(15320.481) | 26331.421(9963.269) |
| PE (p-40:4)/PE (o-40:5) A | 2896.209(2724.733) | 2989.973(2175.181) | 3398.239(3429.56) | 3062.06(2829.321) |
| PE (p-40:4)/PE (o-40:5) B | 3708.007(1951.993) | 3692.493(1528.458) | 3740.207(2103.555) | 3768.74(1900.642) |
| PE (p-40:5)/PE (o-40:6) | 6740.897(2550.286) | 6806.706(1987.436) | 7388.568(2892.277) | 6765.288(2266.793) |
| PG (34:0)/PG (17:0/17:0) | 76853.368(21921.357) | 85280.313(20224.124) | 80493.808(19195.823) | 85678.567(20378.055) |
| PI (38:4)/PI (18:0-20:4) | 120817.481(39400.359) | 124008.066(31642.823) | 121390.805(44702.266) | 129277.347(34643.533) |
| SM (d30:1) - ESI(-) | 1704.845(1134.453) | 1947.17(774.108) | 1614.585(1242.928) | 1863.674(954.282) |
| SM (d32:0) - ESI(-) | 1032.341(809.55) | 1064.49(657.551) | 979.933(686.425) | 1216.938(807.716) |
| SM (d32:1) - ESI(-) | 44492.705(15861.723) | 49754.267(12913.605) | 43920.769(18017.314) | 49980.838(14159.591) |
| SM (d32:2) - ESI(-) | 3030.003(1334.652) | 3230.614(1251.27) | 2552.585(1182.446) | 2652.193(879.559) |
| SM (d33:1) - ESI(-) | 22975.489(8246.125) | 25365.753(7028.514) | 23780.337(8047.678) | 25807.776(6325.893) |
| SM (d34:0) - ESI(-) | 4927.976(1784.951) | 5032.83(1672.907) | 5253.149(1984.636) | 5018.593(1607.857) |
| SM (d34:1) - ESI(-) | 155715.48(39317.352) | 163369.808(34296.98) | 168871.615(43024.9) | 165901.833(36831.034) |
| SM (d34:2) - ESI(-) | 57475.881(14166.69) | 60371.724(13403.338) | 58039.082(15647.963) | 57143.485(10939.005) |
| SM (d36:0) - ESI(-) | 1066.803(653.242) | 879.715(483.034) | 1194.004(818.479) | 1158.849(602.847) |
| SM (d36:1) - ESI(-) | 36132.26(10486.124) | 36986.044(8862.176) | 38899.489(13766.352) | 38825.686(9996.914) |
| SM (d36:2) - ESI(-) | 5807.249(1805.715) | 5799.331(1630.41) | 5885.647(1989.529) | 5864.148(1760.527) |
| SM (d36:3) - ESI(-) | 1713.798(653.047) | 1804.421(563.54) | 1608.324(562.31) | 1777.223(446.756) |
| SM (d37:1) | 2411.418(1019.95) | 2404.494(818.507) | 2331.972(1186.471) | 2389.831(996.191) |
| SM (d38:0) | 696.441(811.576) | 522.617(447.127) | 687.455(703.572) | 667.066(366.14) |
| SM (d38:2) - ESI(-) | 4790.522(1380.077) | 4974.002(1318.512) | 4819.556(1563.18) | 4541.111(1070.145) |
| SM (d39:1) - ESI(-) | 12841.047(4278.992) | 14353.601(3784.665) | 12414.395(4483.9) | 13903.713(3726.042) |
| SM (d39:2) | 12449.201(3511.981) | 12954.634(3279.589) | 9947.898(2966.294) | 10080.729(2701.081) |
| SM (d40:1) - ESI(-) | 46897.381(13275.565) | 50082.888(10695.858) | 48109.832(15111.068) | 51482.472(10102.847) |
| SM (d40:2) A - ESI(-) | 12273.795(4442.543) | 12319.223(4118.364) | 13018.212(5895.037) | 12570.412(4851.714) |
| SM (d40:2) B - ESI(-) | 17613.206(5158.79) | 18864.027(4350.959) | 17289.91(5626.146) | 17753.258(3641.515) |

**Supplementary Table S1.** Sample mean and standard deviations of metabolites in ME/CFS patients, controls, and their subgroups.

| Metabolite | ME/CFS (n=106) | Control (n=91) | Male ME/CFS (n=31) | Male Control (n=22) |
|---|---|---|---|---|
| | mean (SD) | mean (SD) | mean (SD) | mean (SD) |
| SM (d40:3) | 1079.194(398.684) | 1151.905(387.017) | 1149.041(414.719) | 1024.09(283.58) |
| SM (d41:1) - ESI(-) | 11075.677(11704.719) | 13579.641(12424.293) | 12192.786(12824.811) | 12879.432(14073.923) |
| SM (d41:2) - ESI(-) | 10157.654(3759.068) | 10335.005(4166.592) | 9557.389(3960.061) | 9775.169(3425.035) |
| SM (d42:0) - ESI(+) | 631.117(319.733) | 572.32(282.695) | 652.379(371.709) | 686.442(284.971) |
| SM (d42:0) - ESI(-) | 2435.513(802.742) | 2325.825(740.385) | 2351.234(832.029) | 2448.334(558.062) |
| SM (d42:1) - ESI(-) | 74113.422(22000.912) | 76751.556(18783.673) | 78807.969(25168.668) | 82719.057(18246.025) |
| SM (d42:3) - ESI(-) | 45369.2(13437.893) | 44941.003(10551.682) | 48218.789(16038.226) | 43667.711(11335.064) |
| SM (d43:1) - ESI(-) | 1610.475(804.178) | 1785.517(763.855) | 1592.12(791.256) | 2166.055(915.104) |
| SM (d43:2) - ESI(-) | 6637.877(3329.003) | 6677.809(2804.014) | 6696.314(3775.834) | 7584.084(3636.587) |
| SM (d44:2) | 3401.174(1320.178) | 2954.684(961.942) | 3415.928(1601.631) | 2902.033(893.587) |
| SM (d41:3) | 17542.807(4858.342) | 18495.055(4033.934) | 16527.565(4817.077) | 17548.333(4842.736) |
| SM (d30:1) - ESI(+) | 4627.855(3294.462) | 5081.766(2205.718) | 3652.203(2648.807) | 4401.976(2692.374) |
| SM (d32:0) - ESI(+) | 4527.417(2257.972) | 4461.163(1515.584) | 3682.177(1698.839) | 3886.828(1408.425) |
| SM (d32:1) - ESI(+) | 127487.665(50390.527) | 141586.567(42157.93) | 105868.284(47317.742) | 123863.338(37889.321) |
| SM (d32:2) - ESI(+) | 8419.355(3072.987) | 8850.662(2752.191) | 6369.383(2110.37) | 6595.601(1985.036) |
| SM (d33:1) - ESI(+) | 78897.543(30030.413) | 81545.929(24155.104) | 67457.824(25763.753) | 69610.008(18148.263) |
| SM (d34:0) - ESI(+) | 77128.042(21734.185) | 77407.547(19252.704) | 72210.897(23280.32) | 68796.729(18834.64) |
| SM (d34:1) - ESI(+) | 1815521.625(427328.564) | 1839477.475(326129.156) | 1784355.088(430500.205) | 1702876.808(358785.295) |
| SM (d34:2) - ESI(+) | 169414.754(41285.046) | 172230.478(39764.32) | 149178.135(39155.163) | 143390.023(28536.708) |
| SM (d36:0) - ESI(+) | 18743.046(8907.96) | 15332.243(6070.249) | 18497.609(10593.099) | 15396.526(6078.054) |
| SM (d36:1) - ESI(+) | 282139.394(75455.508) | 277277.338(58280.773) | 271000.224(91366.868) | 258341.543(75770.475) |
| SM (d36:2) - ESI(+) | 637435.743(182402.554) | 634824.46(155709.619) | 609372.433(220346.568) | 559314.781(158419.266) |
| SM (d36:3) - ESI(+) | 6491.66(2413.088) | 6629.303(2061.528) | 5303.916(1662.699) | 5372.054(1467.429) |
| SM (d38:1) | 224074.217(52840.443) | 238184.289(46405.144) | 201776.054(59116.355) | 217368.176(47785.659) |
| SM (d38:2) - ESI(+) | 102238.769(23254.677) | 103697.728(23384.632) | 91480.79(22758.592) | 86392.382(21671.092) |
| SM (d39:1) - ESI(+) | 55126.946(19385.078) | 60979.282(17058.805) | 44658.537(16713.1) | 51877.857(13963.345) |
| SM (d40:0) | 8519.977(3597.278) | 7725.238(2745.348) | 7677.025(3167.843) | 8011.238(2212.435) |
| SM (d40:1) - ESI(+) | 357194.17(87098.783) | 373303.076(69403.177) | 341604.881(102437.243) | 362095.736(80274.56) |
| SM (d40:2) A - ESI(+) | 192003.867(74381.653) | 206939.521(72862.228) | 178078.484(72349.431) | 179868.527(58152.864) |
| SM (d40:2) B - ESI(+) | 235158.05(59785.384) | 255298.203(57750.75) | 210548.235(60355.748) | 219351.346(55114.116) |
| SM (d41:1) - ESI(+) | 136526.853(38416.919) | 143873.721(36214.448) | 119899.005(36867.559) | 132527.346(37975.354) |
| SM (d41:2) A - ESI(+) | 41382.821(14921.589) | 40584.006(12105.041) | 35492.179(12478.446) | 33969.219(11162.26) |
| SM (d41:2) B - ESI(+) | 65492.772(20330.14) | 72077.791(21032.207) | 52745.939(16747.156) | 55898.553(15520.632) |
| SM (d42:1) - ESI(+) | 227228.943(65564.52) | 235888.054(53178.336) | 222436.887(63597.422) | 240705.135(54179.07) |
| SM (d42:2) | 552276.944(187216.547) | 490026.241(121909.07) | 546358.226(201991.029) | 460675.764(135621.859) |
| SM (d42:3) - ESI(+) | 360096.856(99212.364) | 348735.256(79546.208) | 337630.936(101307.266) | 299649.811(83353.79) |
| SM (d43:1) - ESI(+) | 6015.056(2273.428) | 6198.991(2041.865) | 5555.044(2105.178) | 6463.747(2405.808) |
| SM (d43:2) - ESI(+) | 14891.052(6807.646) | 13945.815(5754.744) | 13196.624(5716.2) | 13478.476(6491.555) |
| TG (55:6) | 10089.48(6268.964) | 8140.342(3509.246) | 8802.778(3134.839) | 9197.851(3716.421) |
| TAG (58:7)/TAG (18:1/18:1/22:5) | 17350.361(7968.934) | 16648.95(5845.672) | 16639.702(5743.219) | 18540.485(5893.952) |
| TG (60:12) | 6594.761(13689.356) | 3710.704(4070.071) | 3585.552(3335.897) | 3032.451(2280.041) |
| TG (53:5) | 10973.54(5251.608) | 10286.984(3942.7) | 11674.703(5613.463) | 10840.413(5368.36) |
| TG (60:11) | 8428.304(11674.572) | 5497.847(3845.608) | 5706.605(3743.089) | 5470.435(3713.245) |
| TG (40:0) | 6969.903(22773.737) | 5975.792(11932.991) | 10871.416(37277.846) | 5462.54(5759.558) |
| TG (40:1) | 1931.434(4239.428) | 2087.013(3783.783) | 2781.675(6677.307) | 1914.582(2145.639) |
| TG (42:0) | 9234.522(21761.184) | 8844.978(15901.608) | 13307.221(33494.34) | 8330.04(8659.705) |
| TG (42:1) | 6797.568(17520.658) | 7739.975(16319.178) | 9671.813(27713.589) | 8379.533(14255.854) |
| TG (42:2) | 2571.971(5933.309) | 3829.423(10795.773) | 3469.506(9272.753) | 3748.236(7445.047) |

**Supplementary Table S1.** Sample mean and standard deviations of metabolites in ME/CFS patients, controls, and their subgroups.

| Metabolite | ME/CFS (n=106) | Control (n=91) | Male ME/CFS (n=31) | Male Control (n=22) |
|---|---|---|---|---|
| | mean (SD) | mean (SD) | mean (SD) | mean (SD) |
| TG (42:3) | 591.158(714.913) | 671.443(879.22) | 708.25(899.383) | 683.053(790.865) |
| TG (44:0) | 19087.048(32238.546) | 17886.319(25507.344) | 24490.87(41856.21) | 20757.338(22009.867) |
| TG (44:1) | 18788.092(29782.097) | 19478.318(28152.179) | 23426.281(41047.033) | 21621.234(23479.509) |
| TG (44:2) | 8060.679(14384.219) | 10064.354(23367.775) | 10743.044(20194.658) | 8380.317(10487.693) |
| TG (46:0) | 2913.683(2889.773) | 3210.711(2865.495) | 3596.463(3339.855) | 3818.989(2907.059) |
| TG (46:1) | 78988.174(99974.002) | 74842.169(86950.751) | 92276.008(113786.144) | 98918.242(97696.155) |
| TG (46:2) | 34399.438(37849.21) | 37666.18(47712.032) | 39789.39(42673.591) | 41216.21(33094.939) |
| TG (46:3) A | 601.482(563.369) | 733.247(1466.526) | 747.28(697.175) | 571.11(354.662) |
| TG (46:3) B | 5520.005(7775.947) | 5107.884(7131.762) | 7388.887(10758.099) | 4355.864(3462.121) |
| TG (46:4) A | 1736.197(1706.675) | 2452.533(7382.425) | 2137.511(2213.628) | 1684.875(979.075) |
| TG (46:4) B | 1520.942(1603.93) | 2160.803(6838.834) | 1903.962(2062.248) | 1447.092(1031.978) |
| TG (46:5) | 350.027(264.398) | 426.476(1213.469) | 407.412(315.649) | 285.338(153.895) |
| TG (48:0) | 59633.947(40059.321) | 72255.424(51804.827) | 66903.697(41634.01) | 79478.814(50971.025) |
| TG (48:1) | 445353.036(404942.517) | 421062.403(327886.505) | 524213.67(449533.987) | 555493.791(414420.584) |
| TG (48:2) | 232753.491(212433.045) | 215309.555(160839.721) | 255786.613(216764.761) | 268443.377(184286.697) |
| TG (48:3) | 66257.783(45379.776) | 68608.011(52545.382) | 72215.461(50509.48) | 77877.575(53186.217) |
| TG (48:4) A | 14636.937(12229.537) | 16489.544(19911.708) | 16245.558(13562.432) | 18030.998(17055.356) |
| TG (48:4) B | 10569.309(10549.743) | 12439.978(18504.13) | 12504.071(11469.438) | 14971.849(16495.296) |
| TG (48:5) | 2394.932(2124.793) | 2607.435(2613.644) | 2572.8(2035.771) | 2780.416(2398.568) |
| TG (48:6) | 731.456(341.644) | 1120.169(4094.332) | 722.702(289.312) | 705.093(184.872) |
| TG (49:0) | 5475.884(3437.407) | 5547.724(2851.467) | 6100.391(3788.603) | 6307.46(2485.905) |
| TG (49:1) | 38039.35(31682.085) | 32431.609(18199.476) | 44312.657(34999.664) | 40168.672(20279.081) |
| TG (49:2) | 5347.71(2994.124) | 4908.392(3180.31) | 5501.932(3300.177) | 4902.763(2149.578) |
| TG (49:3) | 8782.798(5619.053) | 8058.518(3765.588) | 9221.733(6256.187) | 8066.278(3265.39) |
| TG (50:0) | 28294.36(17765.307) | 35864.991(22391.136) | 32082.118(19239.608) | 44353.561(26478.113) |
| TG (50:1) | 295740.881(378872.38) | 268871.222(259991.973) | 324648.971(446178.44) | 357328.342(249774.674) |
| TG (50:2) | 1473341.655(765417.549) | 1337335.537(606243.723) | 1659532.403(829820.745) | 1594280.037(647567.882) |
| TG (50:3) A | 599554.098(373888.427) | 538070.653(271710.362) | 651512.576(411480.486) | 612121.785(281795.546) |
| TG (50:3) B | 1463.989(2654.267) | 1979.068(6822.777) | 1400.186(2455.795) | 1038.515(809.914) |
| TG (50:4) | 113705.603(61749.855) | 111373.426(49803.428) | 126125.687(75277.714) | 114750.977(56083.861) |
| TG (50:5) | 18164.523(11639.543) | 17487.784(9234.201) | 19933.75(14173.046) | 17309.587(10422.837) |
| TG (50:6) | 2907.741(1946.11) | 2679.302(1733.966) | 2884.817(1921.776) | 2851.086(1436.465) |
| TG (51:2) | 92658.55(49837.791) | 81298.301(33345.525) | 102425.44(56318.197) | 97437.964(38234.191) |
| TG (51:3) | 73828.122(33989.875) | 68061.653(24068.889) | 82938.425(41964.237) | 74898.254(28861.793) |
| TG (51:4) | 20600.271(10073.4) | 19733.727(8062.558) | 23270.722(12671.588) | 19674.341(9041.132) |
| TG (51:5) | 3397.515(1704.001) | 3356.461(1275.262) | 3656.126(2140.47) | 3343.341(1577.736) |
| TG (52:1) | 273344.812(258320.644) | 254133.337(220416.288) | 385958.096(343150.582) | 380249.778(255901.526) |
| TG (52:2) | 3924538.986(1182333.569) | 3636556.102(1122279.122) | 4304562.707(1305332.999) | 4284568.998(1252064.19) |
| TG (52:3) | 4416231.615(1252804.656) | 4180937.847(1101985.633) | 4943186.066(1278613.496) | 4580060.216(1280748.182) |
| TG (52:4) | 133068.448(44192.17) | 130479.658(39344.454) | 149410.712(45631.61) | 134456.837(44467.248) |
| TG (52:5) | 197791.574(102954.234) | 174359.995(84000.915) | 221089.331(113787.339) | 187952.848(123930.287) |
| TG (52:6) | 25178.432(15261.735) | 23719.028(10920.524) | 26434.601(18387.355) | 26935.963(12326.041) |
| TG (53:0) | 1583.982(681.915) | 1815.471(661.05) | 1558.757(477.059) | 1852.357(638.551) |
| TG (53:1) | 5936.774(4668.413) | 5776.819(3671.696) | 7132.934(5901.455) | 7738.895(4054.487) |
| TG (53:2) | 55792.745(24993.696) | 51317.499(21651.56) | 61529.31(28763.169) | 64673.739(28506.202) |
| TG (53:3) | 73056.718(28445.208) | 67703.814(23631.61) | 80399.918(32547.588) | 77888.018(33481.67) |
| TG (53:4) | 46389.39(19729.068) | 43856.196(15698.609) | 49625.115(21404.389) | 47794.59(20989.819) |
| TG (54:1) | 29942.881(33162.778) | 29095.13(22321.153) | 45930.932(54266.538) | 39463.641(28219.587) |

**Supplementary Table S1.** Sample mean and standard deviations of metabolites in ME/CFS patients, controls, and their subgroups.

| Metabolite | ME/CFS (n=106) | Control (n=91) | Male ME/CFS (n=31) | Male Control (n=22) |
|---|---|---|---|---|
| | mean (SD) | mean (SD) | mean (SD) | mean (SD) |
| TG (54:2) | 321515.5(253928.808) | 286249.499(201970.532) | 440117.203(381933.248) | 409260.248(286155.043) |
| TG (54:3) | 1412729.685(654867.912) | 1276837.39(524744.236) | 1623423.817(851645.459) | 1519899.979(713284.681) |
| TG (54:4) | 1525311.494(686168.793) | 1377217.291(578506.358) | 1760418.221(707219.698) | 1591726.506(803792.174) |
| TG (54:5) A | 886580.788(456096.063) | 802522.834(352287.756) | 1025065.173(427497.763) | 906466.793(489401.587) |
| TG (54:5) B | 262000.181(111804.806) | 243734.88(92887.319) | 299788.926(135038.285) | 318097.363(96574.507) |
| TG (54:6) A | 199405.641(130496.852) | 176401.555(94248.554) | 225998.516(121666.969) | 191962.588(134808.348) |
| TG (54:6) B | 1358.531(998.464) | 1341.842(782.147) | 1570.362(998.026) | 1293.618(944.161) |
| TG (54:6) C | 156170.234(80514.484) | 137719.361(62448.4) | 173539.888(102538.702) | 174065.629(81507.228) |
| TG (54:7) A | 49122.361(41128.683) | 44915.907(27315.049) | 55433.098(39043.891) | 44909.3(34205.411) |
| TG (54:7) B | 43061.59(27306.858) | 34198.973(17471.92) | 41427.972(20491.371) | 40360.834(20569.824) |
| TG (54:8) | 7077.455(4680.131) | 6400.128(3571.251) | 6552.552(4150.332) | 7358.226(4649.884) |
| TG (54:9) | 965.383(952.873) | 800.15(740.888) | 592.879(606.119) | 691.956(316.62) |
| TG (55:1) | 2381.308(1476.992) | 2293.733(1193.428) | 2920.799(1903.01) | 2972.521(1491.511) |
| TG (55:2) | 3209.3(1454.698) | 3026.046(1288.231) | 3635.453(1704.304) | 3807.112(1548.074) |
| TG (56:1) | 8425.454(9238.656) | 8724.83(13724.801) | 11505.762(10956.361) | 14774.086(25693.106) |
| TG (56:10) | 3107.113(3323.17) | 2681.061(1812.641) | 2095.903(1680.467) | 2539.949(1566.405) |
| TG (56:2) | 12956.986(10569.613) | 12163.693(13180.75) | 16386.937(12849.427) | 18654.205(23200.288) |
| TG (56:3) | 30313.86(19743.679) | 27075.315(20697.58) | 36217.951(22324.807) | 38868.629(33239.305) |
| TG (56:4) | 52216.171(23781.081) | 49073.426(20687.965) | 55840.537(22467.404) | 56180.741(25306.504) |
| TG (56:5) A | 53791.601(25940.471) | 48027.297(18998.273) | 61801.837(27374.552) | 61191.078(22268.531) |
| TG (56:5) B | 56496.764(28288.186) | 50801.304(20794.293) | 65733.144(33645.995) | 67057.648(26692.101) |
| TG (56:5) C | 113050.899(43777.393) | 107344.31(43874.117) | 118627.317(34824.732) | 122947.456(52182.386) |
| TG (56:6) | 257540.767(82209.876) | 233475.447(74511.483) | 281319.421(89579.965) | 287725.177(91307.651) |
| TG (56:7) A | 847.239(514.565) | 783.159(401.886) | 938.132(655.262) | 929.7(363.376) |
| TG (56:7) B | 187264.223(134086.996) | 142003.825(96398.78) | 158086.258(77277.145) | 161138.457(99617.435) |
| TG (56:8) A | 48927.531(26901.231) | 42661.135(18087.989) | 50064.144(23467.331) | 47257.557(20897.699) |
| TG (56:8) B | 88914.046(72816.031) | 66492.66(53152.85) | 72872.217(40508.64) | 71542.152(51749.829) |
| TG (56:9) | 16134.849(11093.145) | 13921.157(6903.286) | 13754.351(8034.258) | 14787.636(7681.834) |
| TG (57:1) | 1925.193(905.225) | 1863.322(943.086) | 2230.651(1020.533) | 2406.822(1238.998) |
| TG (57:2) | 2561.985(1153.018) | 2479.474(1287.033) | 3067.197(1362.234) | 3326.747(1811.417) |
| TG (58:1) | 6517.314(7345.499) | 6747.301(10399.217) | 8099.715(7474.558) | 11963.869(19532.172) |
| TG (58:10) | 19441.101(14120.814) | 16598.368(8410.925) | 16328.172(8901.342) | 16906.89(8417.462) |
| TG (58:2) | 10622.347(16674.782) | 10834.266(23253.565) | 13254.119(16491.06) | 21111.582(44118.983) |
| TG (58:3) | 6147.029(9675.465) | 5932.174(10911.709) | 7341.209(8673.779) | 10681.352(20516.14) |
| TG (58:4) | 3243.624(2529.139) | 2899.892(2258.659) | 3529.189(2082.371) | 3692.194(2838.546) |
| TG (58:5) | 3750.517(1804.936) | 3824.305(1507.07) | 3786.371(1595.824) | 4264.572(1727.481) |
| TG (58:6) | 14653.583(5134.288) | 13533.44(3981.36) | 15559.451(5777.052) | 16068.613(4533.944) |
| TG (58:8) | 61621.544(53618.106) | 44303.591(28847.455) | 53856.909(45196.344) | 50355.497(39213.793) |
| TG (58:9) | 45461.573(35467.981) | 34450.345(20298.576) | 38767.777(25681.665) | 36919.574(24471.402) |
| TG (59:2) | 1491.779(1048.687) | 1415.968(980.975) | 1872.583(1503.685) | 1971.88(1601.089) |
| TG (59:3) | 1482.159(789.741) | 1388.449(810.7) | 1664.691(965.689) | 1759.21(1206.118) |
| TG (60:1) | 2548.677(2775.351) | 2609.795(3662.099) | 2957.506(2521.105) | 4293.616(6636.964) |
| TG (60:2) | 5106.678(6459.935) | 5391.177(12127.075) | 6248.941(8977.588) | 9524.796(22615.929) |
| TG (60:3) | 3356.443(4588.274) | 3498.227(6192.138) | 3902.058(4033.732) | 6305.992(11703.269) |
| TG (60:4) | 1479.004(1459.393) | 1489.448(1601.775) | 1632.611(1277.701) | 2218.869(2677.546) |
| TG (60:6) | 1058.884(479.188) | 956.092(409.713) | 1095.657(538.542) | 1069.564(367.672) |
| TG (62:1) | 1025.712(1380.888) | 981.286(1057.831) | 1100.973(1066.149) | 1375.722(1920.844) |
| TG (62:2) | 1814.769(2212.815) | 1781.426(2417.302) | 1893.257(1745.858) | 2785.453(4208.202) |

**Supplementary Table S1.** Sample mean and standard deviations of metabolites in ME/CFS patients, controls, and their subgroups.

| Metabolite | ME/CFS (n=106) | Control (n=91) | Male ME/CFS (n=31) | Male Control (n=22) |
|---|---|---|---|---|
| | mean (SD) | mean (SD) | mean (SD) | mean (SD) |
| TG (62:3) | 571.887(485.151) | 559.893(537.266) | 504.508(303.591) | 691.811(585.866) |
| TG (62:4) | 547.101(334.64) | 613.733(374.856) | 570.749(270.285) | 750.37(544.243) |
| TG (64:2) | 864.662(732.038) | 883.063(767.604) | 895.264(780.971) | 1278.603(1231.454) |
| TG (64:3) | 552.787(234.996) | 552.569(261.395) | 586.53(243.089) | 588.42(321.354) |
| **Oxylipins (OL)** | | | | |
| 10-nitrolinoleic acid | 0.186(0.36) | 0.149(0.221) | 0.251(0.429) | 0.172(0.231) |
| 10-nitrooleic acid | 5.022(4.039) | 4.734(2.95) | 4.925(3.125) | 5.642(4.04) |
| 11(12)-epoxy-5,8,14,17- eicosatetraenoic acid | 0.045(0.051) | 0.046(0.062) | 0.026(0.033) | 0.057(0.062) |
| 11,12-Dihydroxyicosa-5,8,14-trienoic acid | 0.729(0.365) | 0.798(0.415) | 0.828(0.43) | 0.956(0.51) |
| 11,12-Epoxyeicosa-5,8,14-trienoic acid | 0.298(0.235) | 0.25(0.225) | 0.357(0.26) | 0.298(0.257) |
| 11-Hydroxy-14,15-epoxyeicosatrienoic acid | 0.137(0.271) | 0.236(0.486) | 0.119(0.181) | 0.327(0.463) |
| 11-Hydroxy-arachidonic acid | 0.954(0.559) | 0.918(0.459) | 0.874(0.468) | 0.873(0.552) |
| 12(13)-epoxy-9,15-octadecadienoic acid | 0.131(0.14) | 0.155(0.255) | 0.224(0.2) | 0.146(0.135) |
| 12,13-dihydroxyoctadec-9-enoic acid | 3.714(2.623) | 4.151(2.308) | 3.873(2.679) | 4.493(3.412) |
| 12,13-dihydroxyoctadeca-9,15-dienoic acid | 0.107(0.148) | 0.111(0.152) | 0.077(0.119) | 0.06(0.077) |
| 12,13-epoxy-9-octadecenoic acid | 2.678(3.381) | 3.027(3.742) | 3.723(5.106) | 3.736(5.303) |
| 12-Hydroxy-5,8,10,14,17-eicosapentaenoic acid | 1.953(2.755) | 1.117(1.211) | 1.527(2.182) | 0.773(0.762) |
| 12-Hydroxy-5,8,10,14-eicosatetraenoic acid | 17.204(15.381) | 15.752(12.397) | 16.128(15.755) | 13.926(15.033) |
| 13-hydroxyoctadeca-9,11,15-trienoic acid | 0.935(0.885) | 0.869(0.593) | 1.247(1.37) | 0.772(0.473) |
| 13-Hydroxyoctadecadienoic acid | 19.586(7.961) | 19.825(7.897) | 19.673(7.942) | 19.885(10.275) |
| 13-ketooctadeca-9,11-dienoic acid | 11.779(7.152) | 12.915(7.243) | 13.064(7.45) | 15.166(7.158) |
| 14(15)-epoxy-5,8,11,17-eicosatetraenoic acid | 0.234(0.427) | 0.211(0.435) | 0.353(0.67) | 0.178(0.248) |
| 14,15-dihydroxyeicosa-5,8,11,17-tetraenoic acid | 0.445(0.453) | 0.503(0.551) | 0.455(0.42) | 0.705(0.862) |
| 14,15-dihydroxyeicosa-5,8,11-trienoic acid | 0.816(0.427) | 0.868(0.394) | 0.931(0.478) | 1.042(0.491) |
| 14-hydroxydocosa-4,7,10,12,16,19-hexaenoic acid | 13.05(14.814) | 10.01(9.899) | 9.949(12.661) | 7.68(10.364) |
| 15(16)-epoxy-9,12-octadecadienoic acid | 0.221(0.211) | 0.382(0.521) | 0.242(0.256) | 0.351(0.455) |
| 15,16-dihydroxyoctadeca-9,12-dienoic acid | 11.678(8.843) | 12.862(6.666) | 13.275(11.413) | 11.275(5.498) |
| 15-Deoxy-delta-12,14-Prostaglandin J2 | 0.043(0.036) | 0.05(0.053) | 0.049(0.048) | 0.047(0.049) |
| 15-hydroxyeicosa-5,8,11,13,17-pentaenoic acid | 0.387(0.417) | 0.288(0.303) | 0.344(0.317) | 0.387(0.317) |
| 15-hydroxyeicosa-5,8,11,13-tetraenoic acid | 3.04(1.536) | 2.905(1.282) | 2.942(1.555) | 2.711(1.289) |
| 15-ketoeicosa-5,8,11,13-tetraenoic acid | 0.159(0.211) | 0.167(0.276) | 0.145(0.244) | 0.17(0.202) |
| 15-Keto-prostaglandin E2 | 0.319(0.368) | 0.405(0.635) | 0.258(0.253) | 0.531(0.627) |
| 16(17)-epoxy-4,7,10,13,19-docosapentaenoic acid | 0.189(0.199) | 0.159(0.206) | 0.239(0.226) | 0.224(0.343) |
| 17,18-dihydroxyeicosa-5,8,11,14-tetraenoic acid | 6.057(5.778) | 4.785(3.299) | 5.853(4.234) | 5.125(3.967) |
| 17,18-Epoxy-5,8,11,14-eicosatetraenoic acid | 0.09(0.188) | 0.122(0.18) | 0.114(0.256) | 0.173(0.255) |
| 17-hydroxy-4,7,10,13,15,19-docosahexaenoic acid | 1.382(0.957) | 1.263(1.094) | 1.408(0.922) | 1.463(1.698) |
| 18-(3-ethyloxiran-2-yl)octadeca-4,7,10,13,16-pentaenoic acid | 0.406(0.383) | 0.403(0.512) | 0.476(0.562) | 0.406(0.366) |
| 19,20-dihydroxydocosa-4,7,10,13,16-pentaenoic acid | 1.929(1.019) | 1.91(1) | 1.814(1.027) | 1.815(1.027) |
| 20-Hydroxyarachidonic acid | 4.005(8.301) | 3.874(6.762) | 3.217(5.556) | 2.633(5.511) |
| 4-hydroxydocosa-5,7,10,13,16,19-hexaenoic acid | 0.763(0.83) | 0.673(0.657) | 0.679(0.748) | 0.714(0.627) |
| 5,15-dihydroxyeicosa-6,8,11,13-tetraenoic acid | 0.038(0.055) | 0.053(0.095) | 0.037(0.056) | 0.033(0.044) |
| 5,6,15-trihydroxyeicosa-7,9,11,13-tetraenoic acid | 0.157(0.194) | 0.207(0.32) | 0.169(0.207) | 0.239(0.359) |
| 5,6-dihydroxyeicosa-8,11,14-trienoic acid | 0.665(0.513) | 0.709(0.521) | 0.845(0.618) | 1.064(0.765) |
| 5-Hydroxy-6,8,11,14,17-eicosapentaenoic acid | 0.954(0.728) | 0.874(0.675) | 0.84(0.643) | 0.875(0.519) |
| 5-Hydroxy-6,8,11,14-eicosatetraenoic acid | 3.249(2.155) | 3.575(2.057) | 3.162(1.957) | 3.932(2.264) |
| 5-ketoeicosa-6,8,11,14-tetraenoic acid | 0.116(0.104) | 0.137(0.127) | 0.105(0.095) | 0.166(0.154) |
| 6-Ketoprostaglandin F1 alpha | 0.111(0.191) | 0.091(0.14) | 0.095(0.221) | 0.104(0.149) |
| 6-trans-Leukotriene B4 | 0.046(0.066) | 0.051(0.099) | 0.048(0.053) | 0.036(0.053) |

**Supplementary Table S1.** Sample mean and standard deviations of metabolites in ME/CFS patients, controls, and their subgroups.

| Metabolite | ME/CFS (n=106) | Control (n=91) | Male ME/CFS (n=31) | Male Control (n=22) |
|---|---|---|---|---|
| | mean (SD) | mean (SD) | mean (SD) | mean (SD) |
| 8,15-dihydroxyeicosa-5,9,11,13-tetraenoic acid | 0.121(0.272) | 0.115(0.202) | 0.103(0.142) | 0.137(0.236) |
| 8,9-dihydroxyeicosa-5,11,14-trienoic acid | 0.391(0.311) | 0.373(0.243) | 0.49(0.39) | 0.484(0.334) |
| 8,9-Epoxyeicosa-5,11,14-trienoic acid | 0.627(0.483) | 0.848(0.762) | 0.781(0.535) | 1.054(1.059) |
| 8-hydroxyeicosa-5,9,11,14-tetraenoic acid | 0.707(0.521) | 0.693(0.505) | 0.617(0.398) | 0.837(0.683) |
| 9(10)-epoxy-12,15-octadecadienoic acid | 1.238(2.543) | 1.495(3.452) | 0.68(0.596) | 2.319(5.687) |
| 9(10)-epoxy-12Z-octadecenoic acid | 0.778(0.586) | 0.789(0.455) | 0.991(0.871) | 0.73(0.429) |
| 9,10-dihydroxyoctadec-12-enoic acid | 3.994(2.996) | 4.527(2.771) | 4.31(2.465) | 4.392(2.521) |
| 9,10-dihydroxyoctadeca-12,15-dienoic acid | 0.195(0.127) | 0.233(0.156) | 0.23(0.131) | 0.172(0.109) |
| 9,10-Epoxystearic acid | 2.482(2.294) | 2.68(2.248) | 2.639(2.737) | 3.33(2.391) |
| 9,11,15-trihydroxy-5,13,1Z-prostatrienoic acid | 0.022(0.082) | 0.033(0.09) | 0.018(0.026) | 0.039(0.105) |
| 9,12,13-trihydroxyoctadec-10-enoic acid | 6.746(3.013) | 7.161(3.062) | 7.35(3.685) | 7.274(3.146) |
| 9-Hydroxy-5,7,11,14,17-icosapentaenoic acid | 0.259(0.225) | 0.218(0.148) | 0.26(0.235) | 0.209(0.147) |
| 9-hydroxyeicosa-5,7,11,14-tetraenoic acid | 0.366(0.266) | 0.386(0.267) | 0.412(0.284) | 0.489(0.346) |
| 9-Hydroxylinoleic acid | 9.759(4.207) | 10.407(3.998) | 9.461(3.48) | 10.005(4.55) |
| 9-hydroxyoctadeca-10,12,15-trienoic acid | 0.602(0.484) | 0.715(0.595) | 0.721(0.513) | 0.539(0.387) |
| 9-ketooctadeca-10,12-dienoic acid | 1.218(0.86) | 1.249(0.847) | 1.297(0.861) | 1.561(0.823) |
| 9-nitrooleic acid | 0.611(0.737) | 0.692(0.676) | 0.479(0.391) | 1.009(0.889) |
| 9S,10R-dihydroxy-stearic acid | 2.561(1.966) | 2.883(2.166) | 2.414(1.3) | 2.741(2.201) |
| Leukotriene B4 | 0.079(0.051) | 0.099(0.087) | 0.069(0.047) | 0.139(0.148) |
| Leukotriene B5 | 0.01(0.015) | 0.012(0.018) | 0.01(0.013) | 0.009(0.017) |
| Prostaglandin D2 | 0.336(0.565) | 0.54(0.959) | 0.459(0.726) | 0.374(1.005) |
| Prostaglandin E1 | 0.02(0.021) | 0.019(0.026) | 0.023(0.022) | 0.019(0.023) |
| Prostaglandin E2 | 0.057(0.126) | 0.052(0.08) | 0.038(0.05) | 0.058(0.066) |
| Prostaglandin E3 | 0.185(0.427) | 0.14(0.232) | 0.28(0.673) | 0.079(0.115) |
| Prostaglandin F2a | 0.83(0.921) | 0.952(0.717) | 0.896(0.714) | 1.078(0.987) |
| Resolvin D1 | 0.147(0.362) | 0.162(0.194) | 0.228(0.619) | 0.204(0.237) |
| Thromboxane B2 | 1.893(2.013) | 2.178(4.62) | 1.753(1.77) | 1.569(0.863) |
| trans-12,13-epoxy-11-oxo-trans-9-octadecenoic acid | 1.085(0.917) | 1.082(0.66) | 1.353(1.407) | 1.114(0.694) |

**Supplementary Table S1.** Sample mean and standard deviations of metabolites in ME/CFS patients, controls, and their subgroups.

| Metabolite | ME/CFS (n=106) | Control (n=91) | ME/CFS without sr-IBS (n-71) | Control without sr-IBS (n-88) |
|---|---|---|---|---|
| | mean (SD) | mean (SD) | mean (SD) | mean (SD) |
| Primary Metabolites (PM) | | | | |
| 1,2,4-benzenetriol | 1079.549(1400.131) | 1012.135(991.038) | 1002.294(1446.43) | 1013(996.75) |
| 1-methylgalactose NIST | 10405.216(25620.129) | 6749.551(12633.562) | 9639.456(25578.02) | 6826.651(12840.65) |
| 2-aminobutyric acid | 6778.127(2312.798) | 6819.584(2333.931) | 6947.426(2457.48) | 6853.965(2362.536) |
| 2-deoxytetronic acid | 3802(1871.5) | 3811.933(1810.183) | 4136.647(1892.756) | 3851.767(1815.416) |
| 2-hydroxybutanoic acid | 37828.971(19975.01) | 35735.831(20029.106) | 40403.824(21082.852) | 35550.186(20212.41) |
| 2-hydroxyvaleric acid | 4237.382(2334.07) | 4287.303(2132.918) | 4376(2466.058) | 4307.965(2155.877) |
| 2-ketoisocaproic acid | 18813.069(5556.745) | 19149.258(6078.798) | 19437.824(5839.105) | 19111.605(6066.511) |
| 3-hydroxybutyric acid | 39212.265(52464.62) | 34152(42085.017) | 47151.324(61640.069) | 34644.686(42734.988) |
| 4-hydroxybutyric acid | 2256.422(736.812) | 2269.843(849.082) | 2329.868(748.029) | 2256.988(857.976) |
| adipic acid | 2895.686(1583.539) | 2847.101(1343.166) | 2937.044(1688.932) | 2856.442(1357.957) |
| alanine | 73245.52(30295.714) | 75399.281(33101.158) | 69742.426(30239.563) | 74847.57(33516.509) |
| alloxanoic acid | 2006.5(1375.468) | 1837.64(1056.367) | 2001.853(1303.393) | 1832.198(1059.848) |
| alpha-ketoglutarate | 1657.627(531.612) | 1527.562(557.531) | 1660.176(533.451) | 1544.233(552.207) |
| aminomalonate | 3619.186(1973.13) | 2911.888(1665.318) | 3531.279(1803.841) | 2949.116(1674.386) |
| arachidic acid | 4924.814(2338.338) | 4808.124(2355.798) | 4976.015(2288.031) | 4799.756(2320.373) |
| behenic acid | 2886.951(836.07) | 3031.506(992.687) | 2896.779(911.452) | 3051.674(1002.272) |
| benzoic acid | 17834.373(6150.942) | 17061.82(6244.429) | 17951.088(6555.996) | 17204.547(6268.506) |
| beta-alanine | 1309.833(728.789) | 1377.112(890.61) | 1354.279(751.62) | 1378.314(898.302) |
| capric acid | 1629.931(794.154) | 1873.809(1125.953) | 1717.574(849.397) | 1863.744(1136.127) |
| caprylic acid | 5704.039(1875.061) | 5854.764(2939.166) | 5700.471(1999.195) | 5865.64(2982.179) |
| citric acid | 41473.245(20630.652) | 41135.135(19465.808) | 41179.353(18335.064) | 41220.093(19676.169) |
| citrulline | 4478.147(1507.009) | 4592.506(1393.12) | 4419.206(1367.037) | 4624.5(1405.977) |
| conduritol-beta-expoxide | 1309.863(794.473) | 1296.146(815.46) | 1272.441(740.555) | 1309.209(826.609) |
| creatine | 7813.647(4623.762) | 7540.27(4234.565) | 7800.397(4490.2) | 7663.07(4255.072) |
| creatinine | 8228.716(4668.248) | 8192.91(4583.811) | 8038.088(4635.892) | 8315.093(4603.393) |
| erythritol | 7881.353(18212.112) | 8811.472(35942.446) | 8728.588(20951.725) | 8912.953(36563.641) |
| fructose | 2597.775(3830.73) | 2215.82(2366.76) | 2974.118(4455.038) | 2048.233(1719.954) |
| fumaric acid | 9386.029(3097.763) | 9434.101(2971.299) | 9307.059(3078.13) | 9442.756(3017.205) |
| gluconic acid | 783.971(293.026) | 776.438(203.662) | 788.294(322.547) | 779.826(202.775) |
| glucose | 388539.039(113173.178) | 378196.978(110227.014) | 372105.956(111571.311) | 375136.081(110684.535) |
| glucose-1-phosphate | 2858.431(1035.667) | 2901.236(1183.342) | 2823.221(1092.204) | 2892.035(1190.876) |
| glucuronic acid | 1283.441(842.594) | 1067.326(518.171) | 1246.574(910.65) | 1032.86(424.542) |
| glutamic acid | 8913.157(3993.644) | 8273.382(4085.076) | 8891.029(4135.891) | 8300.256(4125.786) |
| glutamine | 30937.275(21124.794) | 32325.416(15805.25) | 27801.838(15542.15) | 32439.128(15970.123) |
| glutaric acid | 568.461(206.452) | 532.449(209.713) | 574.647(209.789) | 535.547(212.303) |
| glyceric acid | 10747.598(4862.652) | 9509.18(4092.259) | 10877.706(4945.546) | 9548.802(4138.782) |
| glycerol | 94812.804(37180.866) | 95386.449(35827.521) | 94768.426(39627.514) | 95732.535(36317.421) |
| glycerol-alpha-phosphate | 970.676(374.848) | 952.978(388.821) | 983.456(371.538) | 953.663(394.123) |
| glycine | 68370.147(24574.146) | 66183.809(20178.354) | 65419.279(21264.336) | 66699.453(20297.528) |
| glycolic acid | 6131.461(2349.621) | 6039.528(2582.552) | 5853.515(2358.401) | 6002.709(2618.469) |
| heptadecanoic acid | 4659.657(1498.193) | 4722.045(1568.799) | 4730.838(1554.602) | 4738.151(1582.092) |
| hydroxycarbamate NIST | 9159.343(4227.978) | 9278.079(4002.355) | 9382.147(4686.494) | 9319.419(4063.919) |
| indole-3-acetate | 2888.461(1569.254) | 2930.517(2502.67) | 2886.25(1656.91) | 2953.919(2541.819) |
| indole-3-lactate | 1920.853(610.853) | 2047.753(765.819) | 1928.029(667.814) | 2070.14(765.945) |
| indole-3-propionic acid | 1758.569(1386.393) | 2146.528(1481.229) | 1827.75(1593.418) | 2183.977(1491.28) |
| isoleucine | 42005.147(16265.503) | 42337.787(14176.603) | 43098.574(18307.065) | 42542.116(14223.111) |
| isopropylbenzene | 16849.167(16235.754) | 16249.494(18003.148) | 18262.765(17708.733) | 16298.965(18277.12) |

**Supplementary Table S1.** Sample mean and standard deviations of metabolites in ME/CFS patients, controls, and their subgroups.

| Metabolite | ME/CFS (n=106) | Control (n=91) | ME/CFS without sr-IBS (n-71) | Control without sr-IBS (n-88) |
|---|---|---|---|---|
| | mean (SD) | mean (SD) | mean (SD) | mean (SD) |
| isothreonic acid | 1867.461(718.975) | 1810.944(701.346) | 1772.471(715.529) | 1826.884(701.858) |
| lactic acid | 192717.833(119615.393) | 173890.775(103094.546) | 192678.868(114457.944) | 171758.884(103576.056) |
| lauric acid | 8363.078(12244.741) | 10000.079(13605.883) | 9314.75(14848.838) | 9955.012(13819.512) |
| leucine | 70058.255(22610.471) | 77074.056(24098.618) | 71820(24550.985) | 77269.186(24091.818) |
| levoglucosan | 1659.137(1216.05) | 1870.697(1782.358) | 1529.176(894.134) | 1870.988(1812.067) |
| linoleic acid | 1164.52(386.052) | 1194.708(500.78) | 1198.294(396.716) | 1194.977(505.334) |
| lysine | 15344.127(10811.81) | 15204.719(7437.624) | 14990.118(11169.501) | 15319.256(7540.723) |
| lysine | 18427.5(13565.891) | 18658(9668.614) | 17582.191(13501.045) | 18648.163(9819.033) |
| lyxitol | 3098.794(1031.473) | 3248.989(1189.283) | 3132.412(1034.536) | 3225.488(1186.925) |
| maleic acid | 878.186(349.551) | 788.416(361.008) | 851.632(350.688) | 785.826(366.892) |
| maleimide | 8053.353(10208.802) | 6862.888(5737.055) | 6768.412(3769.233) | 6916.012(5828.894) |
| malic acid | 1120.422(375.058) | 1096.719(407.251) | 1138.147(353.636) | 1090.058(409.804) |
| maltose | 1438.294(1117.335) | 1611.483(3326.737) | 1522.544(1310.21) | 1619.837(3383.48) |
| mannitol | 7384.294(34427.962) | 2531.82(2424.356) | 9382.632(42047.894) | 2403.279(2037.218) |
| mannose | 23546.755(8923.898) | 30536.393(70206.191) | 22654.794(9218.45) | 30986.453(71369.263) |
| methionine | 2349.51(1025.929) | 2403.135(919.059) | 2337.412(1031.868) | 2422.814(918.995) |
| myo-inositol | 21222.363(6524.406) | 20873.449(7257.918) | 20795.441(6090.007) | 21064.256(7295.825) |
| myristic acid | 2466.971(764.975) | 2620.674(814.52) | 2537.25(804.074) | 2624.453(818.158) |
| N-acetylornithine | 2287.627(630.487) | 2420.708(709.39) | 2301.544(647.472) | 2421.628(716.318) |
| N-acetylputrescine | 2335.549(2188.271) | 2392.753(2148.301) | 2347.221(2525.656) | 2411.477(2180.955) |
| nicotinic acid | 22012.588(31343.256) | 23151.326(37119.635) | 20393.426(29744.69) | 23701.186(37639.783) |
| ornithine | 22444.167(16004.833) | 23350.124(11618.996) | 21495.647(14697.704) | 23422.43(11691.106) |
| oxalic acid | 38138.853(34967.284) | 31155.685(23852.651) | 38951.441(30778.027) | 31253.128(23836.053) |
| oxoproline | 148843.422(35478.319) | 149227.169(31627.972) | 148538.309(40405.122) | 149327.919(31936.87) |
| palmitic acid | 83826.745(26436.792) | 87530.236(29254.435) | 84827.985(28657.204) | 87770.872(29640.081) |
| palmitoleic acid | 620.618(273.823) | 620.933(267.671) | 593.971(269.69) | 625.977(270.778) |
| pelargonic acid | 29902.833(10040.315) | 30430.506(13250.889) | 30018.147(10233.407) | 30625.291(13371.786) |
| pentadecanoic acid | 10300.402(2114.862) | 10098.079(2516.636) | 10086.853(2173.19) | 10036.256(2497.45) |
| phenylalanine | 21140.618(6021.369) | 22473.629(6495.431) | 21719.412(6158.625) | 22616.035(6532.526) |
| phosphate | 62360.137(25668.008) | 60891.899(24903.713) | 61856.309(24654.349) | 61628.128(24999.471) |
| phthalic acid | 6103.99(2526.222) | 6071.337(3064.102) | 6055.574(2780.118) | 6088.884(3112.388) |
| proline | 10358.696(7248.838) | 10114.865(6954.499) | 9822.294(6694.674) | 10273.558(7021.455) |
| pseudo uridine | 2775.873(653.848) | 2855.146(716.503) | 2704.059(685.11) | 2871.849(722.867) |
| pyrrole-2-carboxylic acid | 6415.049(4040.235) | 5793.022(3390.2) | 6453.632(4334.966) | 5733.477(3358.36) |
| pyruvic acid | 11489.667(4788.685) | 10820.404(3860.672) | 10829.544(4837.382) | 10764.674(3883.488) |
| quinic acid | 1272.48(1447.647) | 1600.753(1769.131) | 1426.118(1615.364) | 1578.547(1745.589) |
| ribonic acid | 873.931(245.494) | 868.337(240.982) | 871.985(256.98) | 866.93(244) |
| ribose | 2038.549(602.463) | 2037.371(644.28) | 2063.721(569.019) | 2019.605(644.323) |
| salicylic acid | 4293.333(14512.238) | 2853.427(9490.954) | 4636.882(17195.631) | 2906.756(9652.561) |
| serine | 37492.01(17259.952) | 38555.775(15218.55) | 37454.265(17223.185) | 38798.081(15359.179) |
| stearic acid | 378059.412(154306.997) | 405425.36(162088.008) | 387684.956(176535.7) | 405699.663(163820.179) |
| succinic acid | 2428.686(828.589) | 2106.888(729.61) | 2470.044(851.573) | 2121.686(737.847) |
| sucrose | 671.637(698.395) | 613.989(381.592) | 661.397(755.413) | 616.337(387.753) |
| tagatose | 4063.725(4123.989) | 3589.213(2553.245) | 4486.853(4726.886) | 3405.826(1874.604) |
| threitol | 1452.196(807.611) | 1395.584(582.087) | 1448.044(828.449) | 1399.756(591.753) |
| threonic acid | 8560.99(4195.661) | 7285.618(3051.907) | 8697.882(4347.523) | 7371.233(3044.807) |
| threonine | 13282.686(4817.033) | 14237.438(5929.372) | 13059.941(5172.206) | 14384.919(5972.952) |
| trans-4-hydroxyproline | 1700(1224.851) | 1418.764(658.035) | 1583.5(1075.776) | 1433.93(663.804) |

**Supplementary Table S1.** Sample mean and standard deviations of metabolites in ME/CFS patients, controls, and their subgroups.

| Metabolite | ME/CFS (n=106) | Control (n=91) | ME/CFS without sr-IBS (n-71) | Control without sr-IBS (n-88) |
|---|---|---|---|---|
| | mean (SD) | mean (SD) | mean (SD) | mean (SD) |
| tryptophan | 21152.706(10249.693) | 21495.079(8899.268) | 20456.412(10101.724) | 21642.988(8927.827) |
| tyrosine | 42719.343(12721.061) | 44217.124(10941.659) | 42160.985(11955.971) | 44092.942(11061.251) |
| urea | 785114.216(265741.232) | 831665.056(279983.579) | 773192.765(255076.842) | 835214.047(283057.568) |
| uric acid | 32108.833(16219.644) | 33489.944(17068.328) | 33592.912(17269.352) | 33287.047(16813.956) |
| valine | 79110.618(21747.997) | 82792.506(32138.457) | 81631.765(22525.532) | 82766.407(32568.183) |
| **Biogenic Amines (BA)** | | | | |
| (2R)-3-Hydroxyisovaleroylcarnitine | 1880.687(1046.064) | 1825.6(555.898) | 1964.082(1111.925) | 1823.274(546.373) |
| (3-Carboxypropyl)trimethylammonium cation | 16403.7(5302.415) | 17555.878(5550.678) | 17082.591(5521.617) | 17650.284(5558.043) |
| (R)-Butyrylcarnitine | 34078.718(15321.062) | 37105.876(16533.628) | 34942.84(16189.473) | 37122.349(16792.039) |
| .beta.-Phenyl-.gamma.-aminobutyric acid | 869.907(2440.04) | 709.92(241.154) | 1010.528(2952.619) | 703.772(235.316) |
| .epsilon.-Caprolactam | 87755.933(68018.05) | 89742.427(68532.183) | 97864.372(72253.16) | 88548.606(69039.186) |
| 1,2-Dimethylimidazole | 1819.366(2845.633) | 1694.49(1032.734) | 1913.487(3270.852) | 1665.473(999.874) |
| 1-Acetyl-3-piperidinamine | 1839.3(818.798) | 1738.972(605.781) | 1869.535(789.474) | 1741.517(610.102) |
| 1-Acetyl-4-piperidinamine | 2047.585(2838.897) | 1533.704(815.512) | 2030.882(3074.765) | 1526.143(826.822) |
| 1-Methyladenosine A | 4344.676(1038.43) | 4455.103(1138.778) | 4247.068(954.61) | 4438.3(1139.104) |
| 1-Methyl-L-histidine | 22958.602(14651.539) | 23284.637(14336.386) | 21813.926(12773.707) | 23553.407(14488.922) |
| 1-Methylnicotinamide | 14470.114(11101.857) | 14251.823(7589.804) | 14151.955(11178.5) | 14172.234(7654.143) |
| 1-Monostearin | 2115.305(1451.321) | 2830.121(4236.602) | 2214.838(1447.189) | 2838.533(4308.542) |
| 1-Oleoyl-2-acetyl-sn-glycerol | 9191.209(8932.606) | 14964.193(39026.124) | 9675.634(10096.748) | 15001.706(39686.906) |
| 1-Oleoyl-sn-glycero-3-phosphoethanolamine | 6355.005(5324.408) | 6525.509(3772.709) | 6705.173(4473.497) | 6383.607(3732.415) |
| 1-Palmitoyl-2-hydroxy-sn-glycero-3-phosphoethanolamine | 6410.996(4032.932) | 6606.582(3360.162) | 7007.096(4148.859) | 6538.955(3387.552) |
| 1-Phenylpyrrolidine | 143.457(290.638) | 117.902(39.385) | 119.987(38.857) | 117.753(40.034) |
| 1-Stearoyl-2-arachidonoyl-sn-glycero-3-phospho-(1'-myo-inositol) | 1329.25(881.122) | 1346.97(825.295) | 1450.035(901.658) | 1341.422(838.428) |
| 1-Stearoyl-2-arachidonoyl-sn-glycero-3-phosphoserine | 1894.838(2682.073) | 1778.287(2056.277) | 2260.31(3019.281) | 1784.815(2076.621) |
| 1-Stearoyl-2-hydroxy-sn-glycero-3-phosphoethanolamine | 6094.917(4604.091) | 5803.43(2687.326) | 6849.569(4993.056) | 5777.603(2728.394) |
| 2,2',2''-Nitrilotriethanol | 4650.025(6102.206) | 5725.159(8642.651) | 5000.241(7220.226) | 5778.639(8785.674) |
| 2,2-Bishydroxymethyl]-2,2',2''-nitrilotriethanol | 661.365(1011.212) | 683.907(582.601) | 740.988(1215.03) | 687.713(591.539) |
| 2,6-Diaminopimelic acid | 658.783(580.92) | 497.274(446.965) | 597.387(510.284) | 503.577(452.301) |
| 2-Amino-1-phenylethanol | 50064.106(14796.501) | 51939.812(12195.207) | 51385.965(15583.684) | 51976.379(12402.231) |
| 2-Hydroxyibuprofen | 3646.161(11203.95) | 2794.402(8390.204) | 3815.493(12987.705) | 2822.803(8533.437) |
| 2-Indolinone | 22360.698(11782.007) | 25707.648(13967.826) | 21831.879(11194.974) | 25783.918(14012.867) |
| 2-Methylbutyryl-L-carnitine | 15742.272(6758.335) | 19766.912(10894.625) | 16474.913(7150.175) | 19684.529(10960.024) |
| 3-(1-Pyrazolyl)-alanine | 358.412(1009.805) | 338.461(595.673) | 445.471(1212.875) | 346.941(604.154) |
| 3,4-Dimethoxybenzaldehyde | 2111.85(1817.258) | 2433.796(3874.558) | 2365.347(1960.417) | 2414.933(3935.923) |
| 3,5-Dihydroxyphenylglycine | 1521.258(1937.934) | 1263.783(1225.374) | 1207.551(1146.119) | 1289.513(1238.172) |
| 3-Amino-1-propanol | 14759.177(17371.398) | 11356.849(7175.122) | 14919.726(18496.61) | 11469.922(7268.15) |
| 3-Aminoquinoline | 2768.697(3284.974) | 3229.232(3720.884) | 2775.351(3446.016) | 3309.084(3758.856) |
| 3-Cysteinylacetaminophen | 85.628(278.461) | 31.56(22.915) | 94.786(309.643) | 31.33(22.996) |
| 3-Dehydrocarnitine | 2713.916(1241.044) | 2870.766(1231.212) | 2716.877(1166.048) | 2807.342(1127.746) |
| 3-Hydroxybutyrylcarnitine | 3061.608(2849.628) | 2805.221(2351.317) | 3173.922(3045.39) | 2827.079(2388.429) |
| 3-Hydroxyoleylcarnitine | 1499.787(2022.956) | 1253.212(1804.396) | 1701.127(2109.072) | 1243.896(1798.729) |
| 3-Hydroxypyridine | 1971.739(3569.602) | 3209.464(4987.632) | 2499.475(4090.079) | 3107.744(4861.246) |
| 3-Methylglutarylcarnitine | 575.563(258.16) | 1052.615(1965.769) | 573.297(187.677) | 1064.058(1998.76) |
| 3-Methylxanthine | 555.738(650.872) | 522.536(446.424) | 537.007(604.597) | 513.71(445.158) |
| 3-Pyridinemthanol | 1549.304(1050.899) | 1769.148(2316.24) | 1653.049(1204.888) | 1766.567(2354.01) |
| 4,5,7-Trihydroxyisoflavone | 507.397(2564.172) | 124.958(731.854) | 721.161(3093.783) | 127.992(744.321) |
| 4-Acetamidobutyric acid | 3471.712(2342.69) | 3727.521(2854.822) | 3098.373(1702.956) | 3756.798(2896.733) |
| 4-Aminomethylcyclohexanecarboxylic acid; | 1589.572(3239.52) | 1326.667(747.566) | 1479.649(2000.114) | 1310.013(751.776) |

**Supplementary Table S1.** Sample mean and standard deviations of metabolites in ME/CFS patients, controls, and their subgroups.

| Metabolite | ME/CFS (n=106) | Control (n=91) | ME/CFS without sr-IBS (n-71) | Control without sr-IBS (n-88) |
|---|---|---|---|---|
| | mean (SD) | mean (SD) | mean (SD) | mean (SD) |
| 4-Fluoro-.alpha.-pyrrolidinobutiophenone | 532.638(610.097) | 458.298(230.627) | 557.186(655.283) | 458.501(234.515) |
| 4'-Methyl-N-methylhexanophenone | 2337.883(2599.719) | 2483.552(2368.085) | 2164.285(2224.075) | 2511.922(2393.071) |
| 4-Pyridoxic acid; | 9506.692(32185.74) | 3611.761(6927.589) | 9769.983(37038.8) | 3519.903(6939.346) |
| 5'-S-Methyl-5'-thioadenosine | 2295.417(2441.494) | 2541.641(1775.543) | 2262.935(1824.192) | 2495.25(1746.721) |
| 5'-S-Methylthioadenosine | 2232.692(2448.886) | 2396.981(1900.293) | 2231.378(1881.53) | 2343.903(1871.579) |
| 6-Hydroxyflavone | 1191.845(6678.29) | 403.876(528.261) | 1347.613(7984.023) | 405.891(537.138) |
| 6-Methoxynaphthaleneacetic acid | 7862.951(2449.945) | 9327.172(12784.412) | 8028.875(2352.943) | 9343.602(13002.803) |
| 7.alpha.-Hydroxy-3-oxo-4-cholestenoic acid | 7832.128(4085.847) | 7971.016(3525.274) | 7885.787(4371.77) | 7902.631(3519.163) |
| 7-Hydroxywarfarin | 256.276(1326.793) | 125.152(55.26) | 316.493(1609.047) | 124.8(55.977) |
| 7-Methylguanosine | 2894.162(982.806) | 3126.746(1088.991) | 2769.782(932.698) | 3119.694(1106.789) |
| Acetazolamide | 111.881(1032.576) | 10.68(5.05) | 9.858(5.071) | 10.672(5.112) |
| Acetominophen | 8598.401(25412.26) | 1888.819(3095.431) | 11005.405(30203.075) | 1911.304(3146.42) |
| Acetyl-DL-carnitine | 4040.595(1781.273) | 3886.498(1192.6) | 4095.347(1885.905) | 3904.943(1204.502) |
| Acyclovir | 10042.642(38991.986) | 102.244(109.125) | 11367.848(39695.969) | 103.086(110.886) |
| Adenosine | 476.693(252.498) | 554.001(724.574) | 493.273(252.142) | 559.269(736.49) |
| Ala-Ile | 1117.937(531.714) | 1022.708(379.234) | 1120.486(516.01) | 1031.695(382.119) |
| Albendazole | 1404.595(9285.314) | 342.564(133.753) | 347.851(119.357) | 336.326(123.991) |
| Albendazole sulfoxide | 556.474(1732.254) | 706.284(5840.411) | 556.283(1593.204) | 729.06(5940.08) |
| alpha-Methylhistidine; | 18823.314(15559.922) | 19428.048(15625.399) | 16855.378(14043.866) | 19790.576(15733.801) |
| Alprazolam | 219.224(700.569) | 79.36(42.975) | 281.874(843.18) | 79.176(43.537) |
| Aminodiphenylmethane | 10035.44(39641.241) | 4831.419(12787.502) | 12822.363(47422.83) | 4871.615(13000.8) |
| Androstan-3-ol-17-one 3-glucuronide | 1018.393(816.112) | 997.46(700.178) | 1061.494(856.214) | 987.382(709.035) |
| Arginine | 66971.722(30497.565) | 63686.741(31676.786) | 63685.04(25678.028) | 64501.784(31805.912) |
| Atenolol | 2831.445(26482.196) | 235.929(152.586) | 232.143(116.582) | 234.526(152.882) |
| Atorvastatin | 310.188(170.392) | 307.876(96.997) | 321.126(194.431) | 306.747(97.525) |
| Avobenzone | 1620.013(901.713) | 2204.034(6233.272) | 1758.853(946.147) | 2217.883(6339.81) |
| Benthiavalicarb-isopropyl | 397.898(475.323) | 338.666(425.872) | 398.161(445.499) | 335.46(432.386) |
| Benzophenone-3 | 79.42(118.399) | 152.05(480.887) | 72.709(70.367) | 154.79(488.954) |
| Betaine | 1396162.001(400978.201) | 1465520.587(366608.787) | 1391194.829(379473.219) | 1467580.291(372092.124) |
| Betaine aldehyde cation | 14682.235(107622.215) | 56.189(19.083) | 7983.037(65174.742) | 55.811(19.183) |
| Betonicine | 5018.889(9595.989) | 5425.561(9356.727) | 5418.853(11064.457) | 5521.092(9493.804) |
| Biliverden | 37882.387(29682.943) | 38990.463(24851.237) | 39249.673(27018.447) | 39623.023(25017.01) |
| Borrelidin | 1800.784(4109.912) | 1698.556(4228.984) | 1443.631(3762.587) | 1739.927(4296.017) |
| Bradykinin | 2350.694(4905.075) | 2102.181(4523.581) | 2167.847(4312.877) | 2154.829(4592.393) |
| Caffeine | 46358.68(97388.926) | 86219.943(115602.167) | 50554.399(106716.969) | 83627.982(115088.639) |
| Carbamazepine | 424.616(1432.071) | 11883.366(108750.28) | 403.365(1360.899) | 12286.104(110608.577) |
| Carnitine | 1001164.333(283021.215) | 1045982.351(310270.412) | 1025126.234(295143.84) | 1042090.348(313096.555) |
| Choline cation | 254611.526(57164.648) | 282373.342(63913.774) | 252335.789(56953.937) | 281693.168(63982.361) |
| cis-10,11-Dihydroxy-10,11-dihydrocarbamazepine | 525.109(153.075) | 959.717(3818.829) | 523.748(156.925) | 972.976(3884.166) |
| Citrulline | 19901.677(6196.281) | 21504.169(6882.343) | 19304.822(5922.577) | 21774.49(6785.432) |
| Codeine-6-.beta.-D-glucuronide | 774.993(5522.859) | 232.532(91.583) | 1031.562(6698.825) | 231.962(93.078) |
| Coniferylaldehyde | 765.911(462.54) | 787.668(824.323) | 784.989(491.256) | 789.407(838.498) |
| Cotinine N-.beta.-D-glucuronide | 76.449(283.47) | 46.197(159.887) | 101.112(341.741) | 47.042(162.585) |
| Creatine | 217981.51(118303.317) | 203650.285(97913.88) | 211908.405(121174.486) | 205858.724(98348.005) |
| Creatinine | 1006729.671(287698.034) | 1029213.304(254200.686) | 1033881.328(295858.663) | 1022055.057(251490.929) |
| Cyclo(Leu-Pro) | 2228.564(1706.261) | 2027.347(1104.514) | 2193.953(1508.134) | 1964.97(1012.064) |
| D-.alpha.-Cyclohexylglycine | 6326.782(6206.867) | 8327.262(8211.34) | 6294.91(5326.197) | 8250.983(8285.554) |
| Decanoyl-L-carnitine | 42146.5(48589.406) | 42071.952(30458.853) | 46740.83(55984.725) | 42126.188(30821.977) |

**Supplementary Table S1.** Sample mean and standard deviations of metabolites in ME/CFS patients, controls, and their subgroups.

| Metabolite | ME/CFS (n=106) | Control (n=91) | ME/CFS without sr-IBS (n-71) | Control without sr-IBS (n-88) |
|---|---|---|---|---|
| | mean (SD) | mean (SD) | mean (SD) | mean (SD) |
| D-erythro-Sphingosine-1-phosphate | 7295.174(3378.972) | 7543.128(3687.008) | 7149.919(3292.013) | 7567.496(3729.923) |
| Dexpanthenol | 1546.048(2341.347) | 2108.653(3911.294) | 1667.173(2793.252) | 2123.987(3977.94) |
| D-Fructose | 34526.336(8214.427) | 34866.41(9180.115) | 33495.541(7851.696) | 34735.272(9281.089) |
| Diazepam | 370.721(2360.704) | 46.476(76.296) | 526.068(2856.845) | 46.208(77.458) |
| Dinor-12-oxophytodienoic acid | 525.23(862.885) | 416.42(194.711) | 557.78(959.241) | 414.892(197.743) |
| Diphenhydramine | 2585.349(6127.955) | 3178.366(14715.458) | 2824.194(7289.56) | 3227.817(14967.412) |
| DL-Indole-3-lactic acid | 37199.448(18913.594) | 35604.136(12065.446) | 38487.416(21614.263) | 35443.174(12139.329) |
| D-Pyroglutamic acid | 121612.569(22366.016) | 123013.056(24064.978) | 120213.273(22046.967) | 123457.112(24079.229) |
| D-Turanose | 525.908(351.452) | 631.074(581.896) | 511.203(333.272) | 630.228(590.162) |
| Ergothioneine | 767.24(1494.771) | 719.663(431.749) | 852.919(1799.119) | 724.995(438.079) |
| Esomeprazole | 2660.51(10721.074) | 1307.945(4540.326) | 2883.769(11843.121) | 1322.815(4618.105) |
| Ethiolat | 11862.296(20760.107) | 9779.989(18228.881) | 11778.576(20175.243) | 9744.616(18528.037) |
| Ethyldiethanolamine | 734.979(766.768) | 727.196(595.857) | 697.378(488.565) | 719.253(595.658) |
| Ethylenediaminetetraacetic acid | 663091.766(891136.229) | 496339.244(788805.165) | 602816.808(882295.101) | 507817.899(798840.96) |
| Ezetimibe | 223.611(238.157) | 234.24(306.197) | 246.134(270.897) | 234.272(311.216) |
| Fexofenadine | 450.355(238.197) | 841.091(3125.726) | 440.082(123.375) | 853.737(3178.997) |
| Gabapentin | 62080.105(201779.015) | 5287.117(32734.04) | 42184.573(179451.146) | 5380.885(33296.054) |
| Glaucine | 860.63(2849.169) | 574.959(169.559) | 1020.539(3449.8) | 572.998(170.928) |
| Glutamic acid | 7933.975(3124.287) | 7180.498(3426.266) | 8006.862(3234.641) | 7257.869(3456.197) |
| Glutamine | 101505.785(19190.477) | 102907.791(20574.24) | 100918.678(18945.978) | 103180.337(20602.324) |
| Glycocholic acid | 4874.144(5532.195) | 3707.728(3340.534) | 4264.406(4640.264) | 3686.655(3372.994) |
| Glycodeoxycholic acid | 16574.334(17400.385) | 14780.84(19064.118) | 14727.616(15895.011) | 14697.086(19278.505) |
| Gly-Pro-Arg | 586.043(2198.483) | 428.808(1170.147) | 536.568(2176.596) | 440.366(1188.677) |
| Guanidine | 5493.87(8781.142) | 4475.798(4190.121) | 4774.838(3439.928) | 4491.344(4261.485) |
| Guanine | 6226.314(23441.739) | 279.003(1082.414) | 7109.677(24227.455) | 284.1(1100.765) |
| Heptadecasphing-4-enine | 5557.437(4651.009) | 5538.189(4468.342) | 6346.584(4544.059) | 5406.862(4462.629) |
| Hexanoyl-L-carnitine | 8996.499(13556.537) | 8356.705(5002.169) | 10094.335(16137.172) | 8326.454(5023.511) |
| H-gamma-glutamyl-glutamine | 2339.192(1792.988) | 2006.339(1305.502) | 2063.811(1241.507) | 2042.336(1310.079) |
| Histidine | 49555.943(23320.509) | 53178.919(21745.875) | 48156.01(21266.441) | 53685.785(21575.736) |
| Homoarginine; | 3381.815(2097.316) | 3026.593(2160.863) | 3192.592(2022.598) | 3054.736(2192.018) |
| H-Pro-Hyp-OH | 2228.15(1753.581) | 1756.456(871.446) | 2137.306(1824.059) | 1786.547(866.895) |
| Hydroxybupropion | 884.813(2016.341) | 1883.832(8028.514) | 599.334(949.899) | 1142.921(3669.234) |
| Hypoxanthine | 31281.328(19491.261) | 30486.398(22288.554) | 33840.545(21057.089) | 30357.187(22599.14) |
| Ile-Glu-Arg | 365.728(1522.667) | 320.225(998.515) | 363.02(1672.624) | 328.745(1014.695) |
| Indole-3-propionic acid | 1220.413(1004.075) | 1438.512(994.93) | 1286.035(1140.982) | 1464.082(1001.703) |
| Irbesartan | 34111.466(330125.328) | 1632.597(1008.364) | 49418.34(400452.975) | 1628.175(1023.2) |
| Isoleucine | 15404.33(5244.539) | 15607.18(5393.939) | 15857.795(5470.903) | 15551.535(5413.814) |
| Isopentenyladenine | 1014642.651(399845.125) | 1046922.522(370983.691) | 1040237.761(443711.938) | 1051035.2(375459.37) |
| Isopropylamine | 4749.851(1414.044) | 5211.39(1333.561) | 4703.129(1377.742) | 5212.833(1349.979) |
| Kynurenine | 2125.376(754.54) | 2313.805(749.317) | 2134.227(784.952) | 2315.98(759.339) |
| Lamotrigine; | 2244.641(22565.331) | 18.464(16.389) | 3293.018(27372.274) | 18.308(16.126) |
| Lansoprazole | 736.475(4048.63) | 2665.031(13708.313) | 874.922(4660.598) | 2217.891(13107.924) |
| Lauric acid diethanolamide | 413.111(567.669) | 345.335(216.443) | 384.371(227.102) | 344.743(217.118) |
| Lauroyl-L-carnitine | 5784.303(4911.391) | 6763.368(3908.375) | 6324.233(5264.912) | 6730.446(3968.773) |
| L-Carnitine | 16079.975(4595.436) | 15890.768(4307.224) | 15854.424(4409.427) | 15894.437(4362.249) |
| L-Citrulline | 20161.291(5985.134) | 21758.529(6402.073) | 19548.345(5692.582) | 22022.422(6292.986) |
| L-Cysteine-glutathione disulfide | 1116.696(1162.972) | 1224.484(1444.167) | 993.135(1007.08) | 1252.166(1460.802) |
| L-Cystine | 2137.312(1969.911) | 2271.747(2210.344) | 1920.176(1901.115) | 2313.903(2235.856) |

**Supplementary Table S1.** Sample mean and standard deviations of metabolites in ME/CFS patients, controls, and their subgroups.

| Metabolite | ME/CFS (n=106) | Control (n=91) | ME/CFS without sr-IBS (n-71) | Control without sr-IBS (n-88) |
|---|---|---|---|---|
| | mean (SD) | mean (SD) | mean (SD) | mean (SD) |
| Levocetirizine; | 9007.024(38087.67) | 5404.405(22513.101) | 5961.791(22780.063) | 5582.386(22881.407) |
| Linoleoylcarnitine | 6028.057(4694.504) | 6937.757(5240.418) | 5446.701(3980.575) | 6937.188(5311.453) |
| L-Leucine, methyl ester | 936.172(1279.128) | 734.385(675.979) | 1016.061(1338.561) | 744.236(684.933) |
| Losartan | 1124.145(6579.061) | 2289.546(19221.441) | 1542.5(7964.007) | 263.914(134.048) |
| L-Threonine | 6399.383(2197.795) | 6434.59(1928.87) | 6121.688(1937.236) | 6477.306(1940.987) |
| L-Tyrosine | 3214.55(1382.508) | 3345.219(1261.579) | 3149.322(1298.755) | 3360.462(1280.179) |
| Lysine | 24048.561(14481.217) | 23142.646(14133.037) | 22328.548(12990.746) | 23472.954(14233.908) |
| Matrine | 2325.219(12579.91) | 500.431(207.181) | 1979.721(11338.092) | 497.761(210.046) |
| Mefenorex | 622.656(716.718) | 3807.946(26441.59) | 717.441(840.228) | 3922.235(26891.457) |
| Meloxicam | 28306.258(195269.072) | 12851.881(121675.611) | 40080.013(236137.948) | 13294.223(123755.678) |
| Meprobamate | 1573.996(3579.443) | 932.297(1488.818) | 1420.676(3236.736) | 936.585(1514.249) |
| Metformin | 7135.099(48045.124) | 3475.203(23828.338) | 9721.006(58168.254) | 963.019(505.131) |
| Methacholine cation | 56501.078(33409.327) | 67485.057(42836.21) | 60371.886(33845.977) | 67825.387(43486.302) |
| Methionine | 6161.544(1634.068) | 6223.87(1770.355) | 6127.603(1616.691) | 6241.36(1792.871) |
| Methioninesulfoxide | 753.827(372.4) | 693.081(379.811) | 736.68(312.772) | 696.836(381.164) |
| Methylgallate | 139.383(458.151) | 67.958(251.269) | 124.443(429.276) | 62.674(248.786) |
| Metoprolol | 897.413(4862.805) | 234.563(653.184) | 1030.256(5729.681) | 236.384(664.401) |
| Metoprolol acid | 7786.566(33910.371) | 535.074(2767.371) | 7101.932(33547.471) | 544.392(2814.749) |
| Milnacipran | 276.882(1331.735) | 148.067(53.863) | 344.499(1614.036) | 147.503(54.686) |
| Modafinil | 1075.48(6709.26) | 71.168(24.162) | 1446.432(8084.443) | 71.001(24.362) |
| Modafinil acid | 131.611(721.079) | 24.78(13.35) | 164.487(862.058) | 24.762(13.498) |
| Montelukast-1,2-diol | 75.941(301.129) | 84.645(479.266) | 94.825(363.323) | 86.966(487.386) |
| Moxonidine | 3500.133(2726.862) | 3040.75(1987.891) | 3269.679(2397.851) | 3035.903(2015.855) |
| N-(3-(Aminomethyl)benzyl)acetamidine | 20921.024(28034.826) | 18034.449(15522.733) | 21990.895(31106.262) | 17947.809(15636.204) |
| N-.alpha.-Acetyl-L-arginine | 3082.187(1181.981) | 3213.461(1045.445) | 2974.499(1008.387) | 3239.071(1053.579) |
| N.alpha.-Methyl-L-lysine | 29306.073(114182.748) | 13134.602(36620.029) | 38981.267(137655.472) | 13367.14(37228.844) |
| N.epsilon.-Acetyl-L-lysine | 2453.841(2107.986) | 2280.92(1269.094) | 2214.053(870.202) | 2287.94(1287.955) |
| N.epsilon.-Methyl-L-lysine | 8975.039(6927.428) | 8793.019(6711.352) | 8847.949(6185.507) | 8950.23(6772.251) |
| N8-Acetylspermidine | 3001.558(1211.178) | 2900.086(896.957) | 2933.751(1133.346) | 2921.058(892.561) |
| N-Acetylalanine | 707.391(950.515) | 582.675(496.092) | 687.736(1072.698) | 585.421(501.727) |
| N-Acetyl-D-norleucine | 3757.634(3749.095) | 3860.092(3382.148) | 3762.306(3311.768) | 3925.62(3421.079) |
| N-Acetylhistidine | 1008.892(648.298) | 1039.722(610.487) | 910.498(431.301) | 1046.649(618.025) |
| N-Acetyl-L-carnosine | 2954.051(1639.621) | 2926.55(1628.741) | 2790.698(1423.746) | 2923.049(1604.241) |
| Naproxen | 3476.065(13446.579) | 4450.949(22153.769) | 3552.838(13562.626) | 2410.542(10255.922) |
| NEPSILON,NEPSILON,NEPSILON-TRIMETHYLLYSINE | 13358.236(4821.32) | 15219.514(11953.152) | 12738.5(4616.782) | 15324.205(12109.067) |
| N-Methylhistidine | 35730.75(37645.645) | 33537.29(29498.297) | 34522.756(31691.234) | 33848.774(29950.778) |
| N-Methylproline | 11656.687(9296.226) | 11510.001(9904.91) | 11144.479(7784.066) | 11702.239(10011.173) |
| Norleucine | 15444.661(6060.568) | 15678.34(4128.345) | 16027.945(6891.363) | 15607.118(4128.386) |
| Nudifloramide | 64111.911(56284.553) | 63703.328(37012.76) | 65381.674(63955.93) | 62610.205(35407.751) |
| Octanoylcarnitine | 46353.596(55972.637) | 47100.775(29066.033) | 50793.521(65993.487) | 46929.062(29425.169) |
| Oleoyl-L-carnitine | 10823.414(8463.672) | 11676.244(8434.192) | 10434.869(7958.94) | 11562.747(8454.363) |
| Omeprazole sulfone | 4929.053(23929.418) | 1843.877(13957.834) | 4682.463(25395.639) | 1902.955(14195.467) |
| Omeprazole sulfone N-oxide | 1165.746(5101.882) | 421.937(2441.309) | 1203.344(5637.807) | 433.673(2482.683) |
| Ondansetron | 365.61(1271.909) | 240.115(62.33) | 240.038(61.294) | 239.359(63.023) |
| Ornithine | 3959.933(2287.816) | 4022.624(2665.327) | 3719.599(2105.118) | 4077.565(2687.759) |
| p-Acetamidophenyl .beta.-D-glucuronide | 766.028(3177.057) | 171.088(73.211) | 922.587(3756.171) | 171.036(74.362) |
| Palmitamide | 3270.611(17777.123) | 1962.253(8761.313) | 2255.543(11239.789) | 1905.623(8878.473) |
| Pantoprazole | 898.088(8152.763) | 16270.845(118825.791) | 1274.501(9889.644) | 12249.767(113541.365) |

**Supplementary Table S1.** Sample mean and standard deviations of metabolites in ME/CFS patients, controls, and their subgroups.

| Metabolite | ME/CFS (n=106) | Control (n=91) | ME/CFS without sr-IBS (n-71) | Control without sr-IBS (n-88) |
|---|---|---|---|---|
| | mean (SD) | mean (SD) | mean (SD) | mean (SD) |
| Pantothenic acid | 10030.786(7968.504) | 8465.694(5284.919) | 9300.128(6599.677) | 8462.982(5355.706) |
| Penciclovir | 615.263(2184.586) | 280.151(80.439) | 333.415(306.869) | 280.24(81.822) |
| Phenylacetylglutamine | 31218.152(22442.043) | 34681.21(38768.962) | 29540.297(23675.457) | 34874.898(39323.491) |
| Phenylacetyl-L-glutamine | 14884.784(10785.176) | 16607.075(17719.928) | 14176.705(11371.852) | 16687.101(17973.17) |
| Phenylalanine | 55973.85(16422.42) | 58120.533(13302.726) | 57474.874(17249.079) | 58127.78(13527.3) |
| Pipecolic acid | 425.09(586.911) | 426.157(508.716) | 464.872(675.825) | 429.944(516.87) |
| Piperine | 5718.294(8337.056) | 7720.583(10963.746) | 4754.636(5578) | 7636.219(11059.26) |
| Prazepam | 288.862(1542.242) | 137.608(93.414) | 148.414(91.439) | 137.501(94.513) |
| Proline | 14486.098(6367.337) | 14482.522(5997.29) | 15009.419(6447.947) | 14418.197(6063.209) |
| Propionylcarnitine | 68344.79(23209.488) | 71041.55(26326.498) | 67631.021(23042.474) | 71003.485(26698.093) |
| Pyrantel | 372.256(111.716) | 1129.226(7044.272) | 383.177(96.807) | 1153.099(7164.864) |
| Pyridoxal | 1642.179(3991.664) | 946.29(438.171) | 1554.326(4323.843) | 942.404(443.038) |
| Pyridoxine; | 1022.88(4388.483) | 435.057(382.972) | 1120.681(5181.253) | 436.614(389.41) |
| Quetiapine | 369.86(1339.4) | 222.872(66.167) | 446.395(1621.698) | 222.739(67.065) |
| Quetiapine sulfoxide | 650.477(4802.113) | 136.563(42.808) | 895.536(5822.128) | 136.001(43.418) |
| R-(-)-O-Desmethylvenlafaxine | 592.445(3164.213) | 410.727(1880.497) | 542.382(3282.244) | 419.039(1912.463) |
| rac-4-Sulfoxypropranolol | 202.948(514.821) | 151.335(57.243) | 232.419(621.83) | 146.544(45.621) |
| Ranitidine | 2507.731(13468.136) | 205.008(53.061) | 3294.597(16111.517) | 204.252(53.547) |
| Ranitidine N-oxide | 713.593(4219.564) | 62.623(17.85) | 961.933(5087.309) | 62.584(18.092) |
| Ranitidine-S-oxide | 213.634(515.841) | 132.212(90.781) | 246.652(618.406) | 133.197(91.771) |
| Scopoletin | 591.68(2509.742) | 178.452(414.272) | 434.817(2045.369) | 176.338(420.311) |
| SDMA | 9500.368(2703.971) | 10370.308(2765.049) | 9042.982(2572.939) | 10476.979(2721.616) |
| Serotonin | 18189.163(41222.489) | 14668.474(23684.703) | 22486.287(49447.018) | 14792.877(24081.722) |
| Ser-Tyr-Lys | 193.809(988.093) | 91.613(175.965) | 213.48(1141.475) | 92.741(178.835) |
| Stachydrine | 563959.448(691819.57) | 612164.26(685817.817) | 511672.999(618599.937) | 621464.731(693246.546) |
| Sulfamethoxazole | 41.919(48.984) | 4622.278(43440.039) | 45.834(55.18) | 4780.702(44182.6) |
| Tapentadol-.beta.-D-glucuronide | 221.214(1394.74) | 127.85(761.689) | 223.307(1554.834) | 130.896(774.678) |
| Tauroursodeoxycholic acid | 2706.846(6111.732) | 1545.619(2380.983) | 2178.484(4947.103) | 1558.752(2419.545) |
| Telmisartan | 4699.894(41335.897) | 3856.6(30518.071) | 6649.958(50137.588) | 3969.104(31039.611) |
| Temazepam | 279.199(1061.022) | 153.782(115.023) | 351.216(1283.164) | 153.133(116.734) |
| Testosterone | 3622.17(4140.278) | 4445.438(4912.789) | 3150.597(2338.175) | 4523.775(4977.94) |
| Theanine; | 1819.454(2742.739) | 1366.781(1424.619) | 1492.438(1917.573) | 1396.886(1439.654) |
| Theobromine | 43840.022(50239.28) | 50274.237(39008.573) | 45494.606(50281.908) | 48151.022(37511.801) |
| threo-Dihydrobupropion | 959.671(1263.114) | 899.034(1318.122) | 768.344(397.79) | 838.135(1199.449) |
| Thr-Ile-Arg | 334.018(1580.162) | 174.397(391.703) | 328.627(1774.274) | 177.059(398.201) |
| Ticlopidine | 228.217(1202.183) | 52.712(180.862) | 211.436(1291.966) | 53.867(183.874) |
| Topiramate | 883.512(3767.87) | 360.884(470.487) | 385.991(426.722) | 361.833(478.507) |
| Toradol | 356.538(2066.064) | 150.603(104.488) | 452.906(2504.906) | 151.592(105.936) |
| trans-3'-Hydroxycotinine | 2268.615(10601.908) | 1686.299(5203.465) | 3048.199(12814.502) | 1720.57(5290.065) |
| Trazodone | 2672.469(13819.619) | 206.455(55.025) | 1697.31(9177.841) | 206.049(55.555) |
| Tri-2-ethylhexyl trimellitate | 6954.814(58249.323) | 2150.885(4550.116) | 1351.522(2489.591) | 2182.813(4622.634) |
| Trigonelline | 62593.319(128404.523) | 87574.388(113723.624) | 74835.97(150041.321) | 84879.579(111042.199) |
| Trileptal | 378.242(124.735) | 1737.665(13037.79) | 385.504(132.649) | 1782.652(13260.894) |
| Trimethoprim | 234.855(71.982) | 2512.23(21564.074) | 238.109(68.142) | 2590.361(21932.743) |
| Trimethylamine-N-oxide | 22496.762(34083.237) | 15784.326(13954.105) | 23261.064(38326.745) | 15894.929(14167.11) |
| Triptolide | 572.071(359.723) | 709.428(1341.517) | 552.552(251.866) | 716.691(1364.097) |
| Tryptophan | 15719.222(4762.968) | 15591.457(4927.165) | 15909.637(4590.351) | 15635.127(4890.189) |
| Tyrosine | 6484.429(2346.848) | 6649.41(2125.156) | 6424.944(2350.708) | 6679.864(2155) |

**Supplementary Table S1.** Sample mean and standard deviations of metabolites in ME/CFS patients, controls, and their subgroups.

| Metabolite | ME/CFS (n=106) | Control (n=91) | ME/CFS without sr-IBS (n-71) | Control without sr-IBS (n-88) |
|---|---|---|---|---|
| | mean (SD) | mean (SD) | mean (SD) | mean (SD) |
| Urea | 128763.489(48161.656) | 135736.88(41188.088) | 132639.649(47795.604) | 135287.137(41375.423) |
| Urocanic acid; | 20103.21(25401.881) | 20113.659(16158.372) | 20730.595(29343.465) | 20194.811(16339.206) |
| Usnic acid | 6421.074(38109.236) | 5909.508(26905.817) | 2675.635(1396.189) | 3084.644(2776.796) |
| **Complex Lipids (CL)** | | | | |
| AC (10:0) | 14300.667(14495.722) | 14592.903(12632.497) | 14605.269(15294.075) | 14549.52(12733.997) |
| AC (10:1) | 7852.356(5661.808) | 8700.584(5463.897) | 7807.902(6119.907) | 8702.836(5489.19) |
| AC (12:0) | 6801.104(4854.843) | 7472.708(4034.354) | 7024.789(5213.572) | 7500.509(4071.683) |
| AC (12:1) | 5342.259(2975.805) | 5782.079(2495.997) | 5381.782(2875.004) | 5793.067(2518.137) |
| AC (14:1) | 7653.305(5643.002) | 7865.258(4525.933) | 7885.232(6002.322) | 7914.837(4558.016) |
| AC (14:2) | 4537.778(3261.62) | 5105.633(2880.79) | 4577.987(3375.946) | 5138.183(2902.62) |
| AC (16:0) | 10907.214(3516.947) | 10958.197(3206.646) | 11163.093(3655.045) | 10942.32(3232.673) |
| AC (18:0) | 4409.46(1523.433) | 4658.607(1398.628) | 4531.472(1619.583) | 4656.223(1412.475) |
| AC (18:1) | 15169.411(6563.264) | 14716.482(4589.592) | 15085.89(6843.601) | 14640.462(4609.767) |
| AC (18:2) | 9086.774(4258.756) | 9507.785(3723.066) | 8897.388(4404.011) | 9460.75(3748.879) |
| AC (8:0) | 5424.664(7636.621) | 5324.659(5489.183) | 5206.69(6508.227) | 5274.749(5514.915) |
| CE (14:0) | 483.105(328.13) | 550.26(299.965) | 518.33(358.842) | 552.164(302.75) |
| CE (16:0) | 6284.196(5322.107) | 5971.764(4027.806) | 6581.319(5737.272) | 5931.384(4031.777) |
| CE (16:1) | 44723.031(22333.233) | 45240.669(19130.581) | 46447.98(24469.213) | 45051.908(19198.96) |
| CE (18:0) | 3286.85(1793.581) | 3220.682(1502.875) | 3526.574(1877.186) | 3200.334(1513.806) |
| CE (18:1) | 32176.362(13035.556) | 33019.476(10977.599) | 32674.823(13317.324) | 33024.184(11098.589) |
| CE (18:2) | 360370.43(102450.354) | 395504.729(97524.484) | 360426.024(111369.215) | 396258.771(98429.215) |
| CE (18:3) | 13226.28(5296.356) | 16158.621(6188.509) | 13354.326(5702.323) | 16259.704(6222.806) |
| CE (20:2) | 7353.518(4456.574) | 8560.232(3729.559) | 7123.948(4300.333) | 8629.842(3731.012) |
| CE (20:3) | 65342.381(23690.393) | 72249.761(24364.326) | 65336.374(23908.2) | 72329.291(24626.098) |
| CE (20:4) | 760763.777(278873.716) | 769431.808(251914.063) | 776905.931(292801.179) | 773402.023(253390.857) |
| CE (20:5) | 17407.549(19917.453) | 12423.777(8256.741) | 17943.006(21644.577) | 12547.557(8309.52) |
| CE (22:2) | 686.936(506.637) | 714.698(520.067) | 770.278(535.802) | 714.697(525.222) |
| CE (22:6) | 132582.468(60465.938) | 114384.853(39714.847) | 130693.301(63530.758) | 114958.904(39983.681) |
| Ceramide (d34:1) - ESI(+) | 7991.308(2279.1) | 7701.808(1766.753) | 8163.665(2522.072) | 7680.806(1778.694) |
| Ceramide (d36:1) - ESI(+) | 5055.887(1639.5) | 4781.199(1246.336) | 5109.903(1645.787) | 4787.826(1258.971) |
| Ceramide (d38:1) - ESI(+) | 6696.998(1780.221) | 6981.944(1458.36) | 6733.486(1844.048) | 6980.003(1432.779) |
| Ceramide (d40:1) | 21686.844(5740.739) | 23489.285(4735.334) | 21904.442(6142.908) | 23491.446(4760.333) |
| Ceramide (d41:1) - ESI (+) | 6918.214(1876.08) | 7517.217(1659.079) | 7010.178(1921.702) | 7513.459(1678.067) |
| Ceramide (d42:1) - ESI (+) | 71508.239(18019.881) | 78197.952(14457.978) | 71968.394(19122.953) | 78266.18(14617.875) |
| Ceramide (d42:2) A - ESI (+) | 19273.129(4975.246) | 18114.439(3325.542) | 18973.318(5067.804) | 18081.507(3355.413) |
| Ceramide (d42:2) B - ESI (+) | 14981.161(4149.123) | 17174.124(4245.274) | 15181.345(4134.712) | 17196.409(4290.779) |
| Ceramide (d32:1) | 3595.683(1323.517) | 3755.63(977.806) | 3630.861(1393.167) | 3750.835(988.134) |
| Ceramide (d33:1) | 2916.634(935.773) | 3016.705(807.728) | 2879.441(921.486) | 3015.656(806.98) |
| Ceramide (d34:0) | 4886.997(2053.157) | 4687(1716.579) | 4742.189(2020.416) | 4687.849(1736.201) |
| Ceramide (d34:1) - ESI(-) | 18074.154(4880.408) | 17694.653(3936.635) | 17805.341(5062.865) | 17578.908(3825.734) |
| Ceramide (d34:2) | 3094.181(847.888) | 3030.248(713.119) | 3022.781(901.367) | 3028.889(721.295) |
| Ceramide (d36:1) - ESI(-) | 1637.345(721.82) | 1539.002(608.946) | 1614.569(707.754) | 1523.585(587.181) |
| Ceramide (d38:1) - ESI(-) | 11993.384(4407.452) | 12156.053(3738.061) | 11894.289(4485.06) | 12116.782(3647.169) |
| Ceramide (d39:1) | 6299.483(2739.989) | 6979.275(2440.541) | 6330.75(2703.721) | 6939.846(2399.521) |
| Ceramide (d40:0) | 1765.181(1227.484) | 1564.678(901.085) | 1693.591(1280.901) | 1560.751(910.01) |
| Ceramide (d40:2) | 6057.125(4179.286) | 6235.898(4315.18) | 6154.521(4337.808) | 6241.05(4338.496) |
| Ceramide (d41:1) - ESI(-) | 42041.811(15059.383) | 46380.145(13427.707) | 41883.777(14693.073) | 46114.893(13216.731) |
| Ceramide (d42:0) | 11948.609(7113.944) | 11294.104(4976.297) | 11573.042(7705.79) | 11311.311(4978.851) |

**Supplementary Table S1.** Sample mean and standard deviations of metabolites in ME/CFS patients, controls, and their subgroups.

| Metabolite | ME/CFS (n=106) | Control (n=91) | ME/CFS without sr-IBS (n-71) | Control without sr-IBS (n-88) |
|---|---|---|---|---|
| | mean (SD) | mean (SD) | mean (SD) | mean (SD) |
| Ceramide (d42:1) - ESI(-) | 136674.648(44023.767) | 149770.415(39576.94) | 136190.726(44092.942) | 149392.901(39527.405) |
| Ceramide (d42:2) A - ESI(-) | 49252.166(17827.101) | 47220.491(13180.04) | 47781.287(19009.221) | 46967.429(13005.976) |
| Ceramide (d42:2) B - ESI(-) | 37317.754(16468.604) | 35789.863(11255.565) | 36310.405(16665.687) | 35510.767(11062.936) |
| Ceramide (d43:1) | 2528.23(1370.14) | 2789.752(1196.521) | 2528.074(1295.411) | 2781.344(1188.393) |
| Ceramide (d44:1) | 846.904(541.153) | 822.512(399.283) | 844.843(612.712) | 824.67(403.621) |
| Cholesterol | 266952.992(63543.111) | 283483.733(67507.559) | 270735.61(69029.172) | 283563.583(68251.393) |
| DG (32:0) | 2029.32(748.253) | 1982.392(914.732) | 2129.17(771.564) | 1966.058(917.989) |
| DG (32:1) | 5251.897(3457.449) | 4947.61(2880.584) | 5363.106(3601.043) | 4879.383(2877.177) |
| DG (34:1) | 1041.038(494.395) | 999.382(487.017) | 1067.266(507.496) | 991.249(487.464) |
| DG (34:2) | 32528.392(13807.47) | 30539.842(13070.979) | 32862.579(13681.52) | 30129.214(12756.43) |
| DG (34:3) | 5185.782(3045.124) | 4572.847(2077.17) | 5015.399(3044.733) | 4519.506(2066.811) |
| DG (36:1) | 6350.769(3167.505) | 6121.406(3281.803) | 6677.517(3265.854) | 6040.81(3269.019) |
| DG (36:2) | 55205.662(20357.43) | 49677.788(17704.742) | 54727.262(20725.423) | 49332.853(17712.748) |
| DG (36:3) | 70925.903(27194.121) | 65340.022(22354.965) | 69414.597(26559.636) | 64824.688(22056.768) |
| DG (36:4) A | 16838.454(8807.583) | 16004.561(7408.839) | 16138.078(8196.396) | 15872.433(7304.423) |
| DG (36:4) B | 4169.876(3637.695) | 3516.148(2613.304) | 4115.961(3435.276) | 3573.376(2615.064) |
| DG (36:5) | 2206.156(1211.632) | 2086.253(902.788) | 2190.083(1269.996) | 2083.453(899.178) |
| DG (38:5) | 10566.304(4064.74) | 10089.518(3680.913) | 11132.338(4362.87) | 10071.225(3668.467) |
| DG (38:6) | 6169.284(3104.712) | 5407.065(2535.955) | 6485.037(3285.409) | 5393.998(2542.726) |
| FA (10:0) (capric acid) | 1604.441(1050.232) | 1407.005(1073.102) | 1565.562(941.764) | 1317.378(758.378) |
| FA (12:0) (lauric acid) | 8080.776(5386.504) | 8269.26(5625.99) | 8644.72(6130.857) | 7781.091(4038.99) |
| FA (14:0) (myristic acid) | 46315.443(13985.33) | 46285.78(10922.906) | 48601.306(14763.115) | 46064.933(10864.49) |
| FA (14:1) (physeteric acid) | 5158.99(3232.701) | 4681.581(1944.979) | 5562.146(3597.282) | 4714.16(1950.248) |
| FA (15:0) (pentadecylic acid) | 10544.8(2976.688) | 10266.457(2239.062) | 10666.906(2745.41) | 10279.316(2247.903) |
| FA (16:1) (palmitoleic acid) | 87362.569(45359.632) | 76933.028(35142.058) | 88715.774(46306.12) | 77631.404(35224.576) |
| FA (17:0) (margaric acid) | 22117.789(4474.162) | 21235.84(3887.35) | 22367.807(4051.958) | 21187.509(3901.147) |
| FA (18:1) (oleic acid) | 1454192.087(480456.365) | 1349284.059(394857.372) | 1484484.426(491581.065) | 1355919.32(392066.033) |
| FA (18:2) (linoleic acid) | 573453.977(197151.585) | 559405.212(162215.142) | 588596.244(203846.628) | 560450.802(160694.115) |
| FA (18:3) (linolenic acid) | 52408.007(25261.774) | 49568.813(16119.839) | 53212.903(28367.993) | 49968.942(15968.286) |
| FA (20:1) (eicosenoic acid) | 18300.811(5932.05) | 17239.139(4691.897) | 18599.422(6446.265) | 17284.272(4728.359) |
| FA (20:2) (eicosadienoic acid) | 9414.386(2783.824) | 9398.564(2656.075) | 9545.485(2990.685) | 9424.846(2651.935) |
| FA (20:3) (eicosatrienoic acid) | 2064.852(782.798) | 2106.53(696.017) | 2055.843(802.71) | 2122.482(695.112) |
| FA (20:3) (homo-gamma-linolenic acid) | 9019.322(2564.168) | 9232.52(2388.112) | 9207.061(2815.945) | 9251.423(2411.537) |
| FA (20:4) (arachidonic acid) | 43084.156(11307.632) | 45485.009(12124.954) | 42956.129(11563.455) | 45697.144(12175.132) |
| FA (20:5) (eicosapentaenoic acid) | 7210.305(8172.735) | 5310.154(4416.295) | 7322.744(9333.703) | 5362.462(4452.664) |
| FA (22:0) (behenic acid) | 4796.588(1883.561) | 4776.335(2235.782) | 4695.661(1816.385) | 4742.038(2208.438) |
| FA (22:6) (docosahexaenoic acid) | 35785.158(23581.553) | 31506.346(14537.092) | 36117.296(25965.865) | 31705.54(14638.937) |
| FA (24:0) (lignoceric acid) | 9002.74(3316.901) | 9144.823(3857.725) | 8846.907(3295.393) | 9154.153(3881.644) |
| FA (24:1) (nervonic acid) | 3404.095(1062.601) | 3216.118(871.277) | 3325.956(1013.565) | 3211.83(866.174) |
| Gal-Gal-Cer (d18:1/16:0)/Lactosylceramide (d18:1/16:0) | 37250.999(11809.088) | 37236.749(10987.775) | 38126.573(12337.769) | 37387.827(11064.047) |
| GlcCer (d38:1) | 3730.008(1287.974) | 3948.513(1065.45) | 3692.222(1311.485) | 3943.808(1066.52) |
| GlcCer (d40:1) - ESI(-) | 22073.89(7846.193) | 22731.523(6665.458) | 21941.486(7818.015) | 22781.953(6713.619) |
| GlcCer (d41:1) | 12088.153(4455.943) | 12743.895(4333.197) | 11970.157(4433.761) | 12764.296(4374.701) |
| GlcCer (d42:1) - ESI(-) | 25575.096(9070.161) | 26263.818(7929.532) | 25292.022(8684.437) | 26290.06(7978.255) |
| GlcCer (d42:2) - ESI(-) | 19888.456(7749.181) | 17973.465(6288.748) | 19006.652(6801.252) | 18012.501(6352.361) |
| GlcCer (d14:1(4E)/20:0(2OH)) | 6302.067(3492.07) | 6460.128(3125.454) | 6083.889(3036.22) | 6470.613(3160.733) |
| GlcCer (d34:1) | 7468.445(3751.415) | 8251.003(4419.557) | 7688.989(3880.188) | 8293.003(4453.335) |
| GlcCer (d40:1) - ESI(+) | 4100.143(1480.243) | 4234.688(1219.049) | 4209.927(1613.923) | 4254.942(1225.185) |

**Supplementary Table S1.** Sample mean and standard deviations of metabolites in ME/CFS patients, controls, and their subgroups.

| Metabolite | ME/CFS (n=106) | Control (n=91) | ME/CFS without sr-IBS (n-71) | Control without sr-IBS (n-88) |
|---|---|---|---|---|
| | mean (SD) | mean (SD) | mean (SD) | mean (SD) |
| GlcCer (d42:1) - ESI(+) | 17983.989(6028.82) | 18240.869(5395.163) | 18380.701(6457.292) | 18301.315(5441.569) |
| GlcCer (d42:2) - ESI(+) | 13729.159(5608.295) | 12051.673(4176.986) | 13667.879(6015.572) | 12118.9(4189.449) |
| Lactosylceramide (d18:1/24:1(15Z)) | 2755.733(1184.633) | 2537.724(815.608) | 2807.221(1233.763) | 2555.792(813.809) |
| LPC (14:0) - ESI(-) | 1816.114(1007.334) | 2045.03(925.463) | 1819.835(1054.556) | 2025.459(893.736) |
| LPC (16:0) - ESI(-) | 224092.722(72172.597) | 240419.333(71704.173) | 222259.354(76620.932) | 239765.838(71679.031) |
| LPC (16:1) - ESI(-) | 3256.883(1286.541) | 3465.425(1194.712) | 3186.752(1416.047) | 3436.145(1176.568) |
| LPC (17:1) | 2907.335(879.847) | 2996.073(1096.874) | 3045.115(902.533) | 2990.696(1108.783) |
| LPC (18:0) A - ESI(-) | 7418.389(3009.415) | 8077.783(2590.056) | 7345.942(3256.194) | 8075.065(2619.873) |
| LPC (18:0) B - ESI(-) | 99494.905(36134.374) | 108242.985(32910.725) | 98789.161(39113.631) | 108552.619(33214.707) |
| LPC (18:1) - ESI(-) | 47798.748(19375.801) | 51591.653(19211.667) | 46814.46(19406.559) | 51560.576(19387.13) |
| LPC (18:2) - ESI(-) | 60906.166(28020.858) | 71522.652(31446.609) | 59431.225(27583.488) | 71157.294(31707.303) |
| LPC (20:0) | 2602.173(1120.102) | 2667.429(891.909) | 2649.61(1193.281) | 2693.924(883.003) |
| LPC (20:1) - ESI(-) | 3013.633(1298.505) | 2917.096(1050.301) | 2986.298(1387.035) | 2923.053(1061.524) |
| LPC (20:1) - ESI(+) | 713.456(419.647) | 654.494(268.413) | 687.461(421.378) | 657.034(270.807) |
| LPC (20:2) - ESI(-) | 4059.133(1281.292) | 4450.809(1469.606) | 4092.816(1404.591) | 4465.83(1480.733) |
| LPC (20:2) - ESI(+) | 518.937(285.076) | 598.846(314.265) | 508.188(291.326) | 606.59(313.175) |
| LPC (20:3) - ESI(-) | 3582.326(1705.034) | 4021.253(1564.565) | 3554.185(1701.252) | 3996.738(1552.401) |
| LPC (22:4) | 1706.729(976.366) | 1894.898(1004.387) | 1846.79(1046.267) | 1898.401(1012.839) |
| LPC (22:5) - ESI(-) | 675.167(447.702) | 664.823(383.062) | 658.84(432.803) | 667.096(386.874) |
| LPC (22:6) | 18296.637(8748.95) | 17120.89(7390.865) | 18842.819(9702.32) | 17162.43(7444.474) |
| LPC (o-16:0) | 7726.959(3135.329) | 8366.675(3289.673) | 8201.817(3494.261) | 8401.351(3319.459) |
| LPC (p-16:0)/LPC (o-16:1) | 10752.235(4110.82) | 11778.247(4172.532) | 11259.487(4523.187) | 11823.53(4198.094) |
| LPC (p-18:0)/LPC (o-18:1) | 5609.615(1996.592) | 5649.593(2083.157) | 5692.896(2158.371) | 5677.214(2098.323) |
| LPC (14:0) - ESI(+) | 12225.274(5163.537) | 14568.798(5062.405) | 12517.868(5427.698) | 14524.041(5082.835) |
| LPC (15:0) | 12084.127(4819.914) | 12943.535(4361.419) | 12643.424(5011.723) | 12914.073(4397.571) |
| LPC (16:0) - ESI(+) | 1926106.937(412946.468) | 2025455.264(509547.986) | 1976669.593(444192.812) | 2027571.315(515206.445) |
| LPC (16:1) - ESI(+) | 18357.864(6537.325) | 19256.04(6089.078) | 18601.498(7542.127) | 19201.023(6139.414) |
| LPC (18:0) - ESI(+) | 597884.618(199525.827) | 642665.246(221367.761) | 615986.831(215521.358) | 646231.415(221509.646) |
| LPC (18:1) - ESI(+) | 267553.679(104625.2) | 280752.212(113394.873) | 271445.876(106754.861) | 281850.679(114394.293) |
| LPC (18:2) - ESI(+) | 453109.903(200728.084) | 547483.468(267853.354) | 458809.666(206629.29) | 544878.381(267813.143) |
| LPC (18:3) | 3781.745(1556.029) | 4762.878(2448.244) | 3797.375(1629.076) | 4780.82(2472.778) |
| LPC (20:3) - ESI(+) | 18658.988(7504.447) | 20830.978(7040.033) | 19454.931(8147.028) | 20755.738(7092.886) |
| LPC (20:4) | 92588.56(39171.901) | 99801.287(39143.693) | 97075.22(41447.985) | 100006.125(39387.62) |
| LPC (20:5) | 8554.056(10259.538) | 6693.724(5097.151) | 9211.575(11668.719) | 6757.878(5138.079) |
| LPC (22:5) - ESI(+) | 4133.499(2034.537) | 4006.158(1626.825) | 4363.375(2052.93) | 4016.991(1636.58) |
| LPE (16:0) | 3007.892(1492.945) | 3345.916(1755.537) | 3082.04(1618.03) | 3326.568(1741.116) |
| LPE (18:2) - ESI(-) | 4521.977(2417.216) | 5517.915(3099.215) | 4561.548(2543.135) | 5398.702(3009.274) |
| LPE (20:4) - ESI(-) | 3411.332(1579.491) | 3835.167(1358.006) | 3420.042(1601.323) | 3805.198(1358.863) |
| LPE (22:6) | 2511.314(1103.103) | 2584.869(1142.272) | 2485.081(1071.693) | 2561.784(1140.98) |
| LPE (18:0) | 2976.4(935.468) | 3330.697(987.882) | 3093.293(1012.022) | 3316.427(994.418) |
| LPE (18:2) - ESI(+) | 3940.782(1604.285) | 4738.299(2407.019) | 4018.716(1697.213) | 4635.05(2259.741) |
| LPE (20:4) - ESI(+) | 3205.181(1241.82) | 3617.274(1320.76) | 3270.867(1321.852) | 3577.859(1287.694) |
| PC (16:0/9:0(CHO)) | 4327.064(2722.879) | 4282.307(2603.353) | 4589.78(2660.987) | 4235.219(2523.464) |
| PC (32:0) - ESI(-) | 14490.961(3964.7) | 14558.291(3478.332) | 14243.818(3899.426) | 14492.211(3372.222) |
| PC (32:1) - ESI(-) | 14447.376(7522.552) | 14662.134(6394.392) | 14331.384(7979.761) | 14478.031(6097.206) |
| PC (32:2) - ESI(-) | 13463.361(6508.264) | 16204.369(5624.383) | 13176.148(6724.934) | 16078.818(5564.473) |
| PC (33:1) - ESI(-) | 2353.684(1083.204) | 2374.083(853.644) | 2368.37(1072.803) | 2357.216(835.239) |
| PC (33:2) - ESI(-) | 3860.198(1605.855) | 4336.55(1397.757) | 3790.563(1586.109) | 4295.957(1361.3) |

**Supplementary Table S1.** Sample mean and standard deviations of metabolites in ME/CFS patients, controls, and their subgroups.

| Metabolite | ME/CFS (n=106) | Control (n=91) | ME/CFS without sr-IBS (n-71) | Control without sr-IBS (n-88) |
|---|---|---|---|---|
| | mean (SD) | mean (SD) | mean (SD) | mean (SD) |
| PC (34:0) - ESI(-) | 8251.523(2184.754) | 8469.609(1811.655) | 8162.85(2256.349) | 8443.034(1803.548) |
| PC (34:2) - ESI(-) | 240630.097(60351.691) | 258867.547(56457.148) | 234017.313(61164.965) | 257587.052(55868.894) |
| PC (34:3) | 16244.273(6359.067) | 18088.554(6094.852) | 15419.188(6184.462) | 17986.817(6025.427) |
| PC (34:4) - ESI(-) | 3988.556(2296.794) | 4598.624(1731.583) | 3792.963(2208.225) | 4571.552(1709.372) |
| PC (35:1) - ESI(-) | 3093.959(1098.086) | 3051.722(977.026) | 3090.742(1032.773) | 3045.792(940.977) |
| PC (35:2) | 9207.837(2691.891) | 9716.991(2633.897) | 9074.31(2674.248) | 9663.598(2629.522) |
| PC (35:4) - ESI(-) | 2567.664(1136.268) | 2622.666(936.586) | 2574.127(1111.966) | 2619.452(910.694) |
| PC (36:1) - ESI(-) | 60700.134(17787.723) | 61602.252(15986.223) | 59845.797(18852.856) | 61560.21(16024.035) |
| PC (36:3) A - ESI(-) | 78001.315(26248.654) | 80601.946(20165.252) | 76641.547(27589.436) | 80028.564(19163.692) |
| PC (36:3) B - ESI(-) | 80122.729(25240.712) | 84455.963(19138.885) | 79288.754(26075.408) | 83856.926(18245.131) |
| PC (36:4) A - ESI(-) | 50001.555(19418.961) | 58881.412(19057.435) | 48211.317(20388.858) | 58636.399(19196.913) |
| PC (36:4) B - ESI(-) | 165608.498(45603.524) | 172562.505(36378.869) | 161093.155(45105.372) | 172103.845(36230.461) |
| PC (36:5) A | 2138.31(1339.816) | 2567.216(1605.787) | 1995.656(1373.916) | 2570.171(1623.883) |
| PC (36:5) B | 41857.83(35378.719) | 33407.87(22725.882) | 40052.72(36387.91) | 33637.215(22867.504) |
| PC (37:2) - ESI(-) | 1427.867(571.478) | 1549.892(656.208) | 1356.668(550.82) | 1550.999(663.634) |
| PC (37:3) | 19429.669(8642.077) | 20766.774(7555.42) | 19792.246(8796.838) | 20590.196(7539.777) |
| PC (37:4) - ESI(-) | 3610.43(1262.482) | 3600.739(1164.94) | 3532.53(1237.455) | 3598.039(1176.602) |
| PC (38:2) | 4724.469(1540.569) | 4816.371(1178.537) | 4591.821(1714.226) | 4814.026(1183.586) |
| PC (38:3) - ESI(-) | 29648.655(10260.002) | 31432.948(7745.999) | 29200.155(11217.103) | 31334.232(7693.854) |
| PC (38:4) A - ESI(-) | 57986.619(17566.019) | 60883.346(16516.825) | 56059.722(17810.736) | 60997.571(16671.957) |
| PC (38:5) A - ESI(-) | 19182.481(6029.043) | 19166.914(4819.422) | 18959.816(6263.795) | 19173.336(4833.043) |
| PC (38:5) B - ESI(-) | 12329.808(8869.88) | 10448.295(4369.029) | 12060.15(9605.348) | 10497.711(4388.176) |
| PC (38:6) - ESI(-) | 75564.438(34561.384) | 68827.018(26086.774) | 72807.158(34179.525) | 68997.723(26204.556) |
| PC (39:6) | 18191.534(9715.309) | 16532.441(6454.175) | 18918.135(10297.911) | 16606.293(6492.648) |
| PC (40:4) - ESI(-) | 3651.767(1838.864) | 3888.447(1301.818) | 3705.488(1955.064) | 3873.374(1300.13) |
| PC (40:5) A - ESI(-) | 4364.652(1888.948) | 4187.943(1433.041) | 4349.552(2003.557) | 4197.426(1440.474) |
| PC (40:5) B - ESI(-) | 1949.867(1019.491) | 2160.184(912.295) | 1979.401(1074.925) | 2152.268(909.921) |
| PC (40:6) B | 20657.541(8269.246) | 18554.405(6543.511) | 19980.918(8878.717) | 18577.33(6547.096) |
| PC (40:7) - ESI(-) | 2669.718(1393.762) | 2433.114(925.635) | 2555.608(1399.183) | 2443.043(933.383) |
| PC (40:8) - ESI(-) | 2630.788(1139.643) | 2734.932(900.85) | 2633.311(1250.357) | 2743.855(903.745) |
| PC (42:5) | 3139.766(1127.172) | 3024.694(1140.962) | 2966.434(1195.792) | 3037.599(1149.787) |
| PC (42:6) | 2237.693(1216.735) | 2297.879(1311.502) | 2279.388(1333.629) | 2323.614(1315.028) |
| PC (o-32:0) - ESI(-) | 2806.219(868.036) | 2847.252(794.141) | 2774.69(860.116) | 2831.858(770.697) |
| PC (p-32:0)/PC (o-32:1) - ESI(-) | 2338.644(927.886) | 2365.854(804.708) | 2303.7(928.299) | 2361.585(783.92) |
| PC (p-32:0)/PC (o-32:1) - ESI(+) | 29152.037(7872.188) | 30771.998(7341.47) | 29722.833(8485.24) | 30799.486(7424.032) |
| PC (p-34:1)/PC (o-34:2) A | 4520.84(2258.236) | 5300.943(2014.065) | 4404.968(2150.798) | 5290.747(2023.083) |
| PC (p-34:1)/PC (o-34:2) B | 2874.162(1241.363) | 2975.868(1271.407) | 2792.387(1096.742) | 2982.974(1282.056) |
| PC (p-34:2)/PC (o-34:3) - ESI(-) | 13080.94(4655.151) | 15147.291(4618.811) | 12669.044(4411.375) | 15150.242(4671.192) |
| PC (p-36:1)/PC (o-36:2) | 1579.393(1039.658) | 1842.803(957.813) | 1478.322(947.172) | 1829.295(960.205) |
| PC (p-36:3)/PC (o-36:4) - ESI(-) | 21432.678(7961.51) | 23318.595(6892.306) | 21046.155(8038.699) | 23313.681(6922.909) |
| PC (p-36:3)/PC (o-36:4) - ESI(+) | 227889.598(84207.01) | 254677.838(68833.936) | 234246.607(87202.856) | 256174.875(68897.848) |
| PC (p-36:4)/PC (o-36:5) - ESI(-) | 12136.93(5243.46) | 13380.29(4590.962) | 11751.521(5145.248) | 13413.925(4630.98) |
| PC (p-38:3)/PC (o-38:4) - ESI(-) | 7180.199(2581.694) | 8011.26(2278.842) | 7191.411(2419.954) | 8002.046(2279.346) |
| PC (p-38:4)/PC (o-38:5) A | 16408.903(5205.505) | 17597.828(4431.343) | 15768.113(4826.907) | 17610.51(4453.417) |
| PC (p-38:4)/PC (o-38:5) B | 2559.33(1097.47) | 2769.763(1152.827) | 2509.36(1016.701) | 2776.522(1164.309) |
| PC (p-38:5)/PC (o-38:6) | 1297.362(880.416) | 1291.415(783.351) | 1285.878(843.374) | 1311.131(780.672) |
| PC (p-40:1)/PC (o-40:2) | 1485.155(608.109) | 1292.088(473.435) | 1463.259(613.669) | 1293.224(473.413) |
| PC (p-40:3)/PC (o-40:4) | 1750.816(685.913) | 1868.47(638.755) | 1743.351(652.732) | 1875.269(640.5) |

**Supplementary Table S1.** Sample mean and standard deviations of metabolites in ME/CFS patients, controls, and their subgroups.

| Metabolite | ME/CFS (n=106) | Control (n=91) | ME/CFS without sr-IBS (n-71) | Control without sr-IBS (n-88) |
|---|---|---|---|---|
| | mean (SD) | mean (SD) | mean (SD) | mean (SD) |
| PC (p-40:4)/PC (o-40:5) - ESI(-) | 1855.515(641.097) | 1784.499(529.196) | 1806.243(603.923) | 1784.252(528.02) |
| PC (p-40:7)/PC (o-40:8) | 12784.77(9315.863) | 11294.973(6789.332) | 11685.467(9260.77) | 11384.19(6825.683) |
| PC (p-42:3)/PC (o-42:4) | 4785.842(1784.509) | 5178.399(1630.148) | 5041.944(1885.999) | 5200.635(1632.756) |
| PC (p-42:4)/PC (o-42:5) - ESI(-) | 2268.307(909.076) | 2270.498(870.57) | 2184.702(855.245) | 2261.696(864.096) |
| PC (p-42:5)/PC (o-42:6) | 618.687(355.575) | 610.366(274.685) | 575.455(326.259) | 615.095(273.578) |
| PC (p-44:4)/PC (o-44:5) - ESI(-) | 2845.694(1045.311) | 2893.365(908.463) | 2733.951(944.072) | 2883.804(896.937) |
| PC (p-44:5)/PC (o-44:6) | 3310.494(1275.999) | 3174.484(993.019) | 3243.368(1220.927) | 3191.614(997.8) |
| PC 34:4e | 6443.081(2515.863) | 7999.075(2933.128) | 6404.258(2554.366) | 8019.482(2940.252) |
| PC 38:7e | 16964.282(13526.498) | 15342.968(9446.371) | 16856.913(13156.423) | 15490.122(9504.09) |
| PC 40:5e | 12165.024(3482.196) | 12677.876(3009.874) | 12025.323(3199.489) | 12739.163(3013.606) |
| PC 40:6e | 22316.948(7430.313) | 21474.846(6189.986) | 22197.188(7784.299) | 21623.783(6175.042) |
| PC (28:0) | 3246.565(4662.849) | 3239.16(2348.658) | 3558.295(5489.584) | 3233.267(2374.737) |
| PC (30:0) | 158777.616(83572.094) | 179096.713(65617.314) | 165216.361(93427.652) | 179210.623(66049.201) |
| PC (31:0) | 10922.351(4521.628) | 11042.588(3052.892) | 11358.688(4823.418) | 11035.171(3032.178) |
| PC (31:1) | 1677.788(1455.465) | 1660.939(915.401) | 1775.221(1628.096) | 1645.448(908.327) |
| PC (32:0) - ESI(+) | 256268.716(58170.605) | 250135.791(53964.518) | 259144.909(62925.656) | 250462.669(54506.815) |
| PC (32:1) - ESI(+) | 239846.485(124170.713) | 249536.641(113064.066) | 245455.432(139736.749) | 248950.869(113280.227) |
| PC (32:2) - ESI(+) | 56756.644(28246.729) | 67703.831(25937.146) | 55700.803(30016.44) | 67635.726(26221.119) |
| PC (33:0) | 10952.596(5641.864) | 11592.978(3878.492) | 11689.558(5842.141) | 11572.727(3868.088) |
| PC (33:1) - ESI(+) | 53543.632(20928.978) | 53553.558(13337.174) | 55279.073(22414.831) | 53406.585(13204.347) |
| PC (33:2) - ESI(+) | 78531.369(27527.116) | 87270.676(25504.3) | 78022.252(29190.385) | 86943.817(25702.157) |
| PC (34:0) - ESI(+) | 56673.124(14226.497) | 56575.078(11243.201) | 57988.448(15118.945) | 56790.977(11270.355) |
| PC (34:1) | 4013338.964(720952.007) | 4067426.422(727466.087) | 4062709.877(769160.87) | 4072020.238(730603.307) |
| PC (34:2) - ESI(+) | 8298954.78(1436737.757) | 8880671.15(1333757.007) | 8201779.521(1557049.345) | 8871299.896(1346265.758) |
| PC (34:3) A | 97711.278(31702.813) | 106631.972(39043.209) | 93915.056(33247.85) | 106793.471(39461.44) |
| PC (34:3) B | 108771.059(36242.03) | 128990.918(46188.814) | 106249.745(39129.52) | 129387.848(46646.006) |
| PC (34:3) C | 86598.612(39702.386) | 108697.965(49708.994) | 86591.607(42147.668) | 109263.919(50139.319) |
| PC (34:4) - ESI(+) | 17218.357(9083.701) | 20014.966(7594.249) | 17373.707(9531.92) | 20065.065(7666.87) |
| PC (35:1) - ESI(+) | 46139.315(13080.522) | 45638.931(9232.36) | 47505.991(12892.848) | 45629.634(9303.389) |
| PC (35:2) A | 109536.799(36351.893) | 103212.896(30675.48) | 107047.548(35343.348) | 102815.658(30367.982) |
| PC (35:2) B | 131176.614(32124.982) | 137458.977(30973.043) | 131382.573(33447.818) | 137344.717(31226.014) |
| PC (35:3) | 40372.126(10564.132) | 44343.372(11336.707) | 39709.617(11171.443) | 44234.168(11390.527) |
| PC (35:4) - ESI(+) | 22778.31(8961.192) | 23974.006(6756.722) | 23385.755(9416.11) | 24010.791(6817.916) |
| PC (36:1) - ESI(+) | 656088.196(185659.526) | 660392.494(148989.452) | 662290.869(195630.993) | 663377.135(149374.666) |
| PC (36:2) | 4529060.48(909760.507) | 4996033.975(881985.183) | 4402650.34(946009.211) | 5002768.328(872906.035) |
| PC (36:3) A - ESI(+) | 1277000.345(445850.9) | 1366182.069(370360.325) | 1221268.785(455608.087) | 1369124.271(371511.618) |
| PC (36:3) B - ESI(+) | 1639032.954(502617.113) | 1716418.049(393778.108) | 1637555.054(557291.285) | 1714709.947(396710.519) |
| PC (36:4) A - ESI(+) | 344228.244(141889.724) | 433150.27(175314.159) | 330340.32(148234.03) | 432937.74(176243.894) |
| PC (36:4) B - ESI(+) | 254789.338(417790.093) | 206974.331(69274.833) | 253436.033(397790.263) | 206658.634(69438.458) |
| PC (36:4) C - ESI(+) | 3945315.162(980469.466) | 4037185.749(932394.375) | 4000440.116(994299.382) | 4055722.581(934890.426) |
| PC (36:5) C | 63676.772(293144.05) | 37777.207(75598.623) | 81673.798(354897.755) | 38098.298(76442.208) |
| PC (36:5) D | 466335.725(583640.353) | 328794.138(256617.495) | 482947.978(643937.257) | 332594.28(258248.921) |
| PC (36:5)A | 10087.21(5302.765) | 12082.737(6355.171) | 9555.815(5406.625) | 12156.017(6404.45) |
| PC (36:6) | 7517.927(4687.721) | 7514.303(3539.182) | 7413.234(4814.471) | 7549.236(3569.667) |
| PC (37:2) - ESI(+) | 12948.098(3497.278) | 14134.187(3647.086) | 12778.893(3447.731) | 14195.756(3645.96) |
| PC (37:4) - ESI(+) | 51981.159(19322.358) | 49132.105(16397.252) | 52237.315(19599.551) | 49370.279(16476.626) |
| PC (37:5) | 36908.954(34432.55) | 28967.669(14940.073) | 38452.496(37877.76) | 29236.369(15001.717) |
| PC (37:6) | 9891.772(5465.158) | 8696.041(4047.719) | 9893.467(5484.538) | 8736.468(4077.449) |

**Supplementary Table S1.** Sample mean and standard deviations of metabolites in ME/CFS patients, controls, and their subgroups.

| Metabolite | ME/CFS (n=106) | Control (n=91) | ME/CFS without sr-IBS (n-71) | Control without sr-IBS (n-88) |
|---|---|---|---|---|
| | mean (SD) | mean (SD) | mean (SD) | mean (SD) |
| PC (38:3) - ESI(+) | 545962.223(199887.214) | 581406.422(170804.445) | 541919.095(208893.159) | 582742.133(172543.347) |
| PC (38:4) A - ESI(+) | 129827.943(52570.47) | 130649.431(33178.708) | 128843.841(57666.719) | 130983.455(33483.185) |
| PC (38:4) B - ESI(+) | 138945.853(49865.612) | 150899.97(43419.782) | 145027.775(54570.879) | 151336.198(43779.847) |
| PC (38:4) C - ESI(+) | 2313829.998(565304.201) | 2391882.015(574272.595) | 2321739.659(589973.366) | 2406694.888(570510.436) |
| PC (38:5) A | 732724.247(176405.92) | 721011.877(165292.616) | 735715.185(170649.669) | 725519.057(164432.584) |
| PC (38:5) B - ESI(+) | 385628.57(330057.265) | 324021.431(136699.566) | 399379.583(365571.608) | 326647.318(137120.575) |
| PC (38:6) A - ESI(+) | 122703.865(38145.153) | 135842.09(35120.916) | 121779.409(39418.963) | 136259.293(35324.013) |
| PC (38:6) B - ESI(+) | 3683483.214(1066200.996) | 3373471.275(825187.655) | 3657273.591(1085634.511) | 3389893.16(826762.373) |
| PC (38:7) | 9067.129(4522.378) | 8494.519(3671.831) | 8767.372(4415.118) | 8543.985(3698.92) |
| PC (39:4) | 4951.364(1392.551) | 4980.401(1209.519) | 4979.359(1261.552) | 5022.52(1190.291) |
| PC (40:4) - ESI(+) | 40710.749(17107.156) | 43768.486(14761.039) | 43031.205(18895.324) | 43907.06(14894.586) |
| PC (40:5) A - ESI(+) | 160984.496(51414.716) | 153215.298(48927.121) | 163231.207(53799.49) | 154338.924(48914.87) |
| PC (40:5) B - ESI(+) | 39290.673(16831.51) | 43514.802(16159.023) | 40021.609(17513.652) | 43527.988(16344.456) |
| PC (40:6) A | 26324.155(8352.257) | 26286.775(7120.512) | 26474.383(8548.768) | 26446.792(7116.955) |
| PC (40:6)B | 523343.738(235330.416) | 448154.607(162581.361) | 511675.299(239359.315) | 451652.474(162754.752) |
| PC (40:7) A - ESI(+) | 14123.954(4521.843) | 15295.102(4113.716) | 14402.113(4851.312) | 15322.173(4155.479) |
| PC (40:7) B - ESI(+) | 294687.493(123648.862) | 260986.662(85720.284) | 280811.224(122348.869) | 262880.664(85775.009) |
| PC (40:8) - ESI(+) | 15239.14(4848.028) | 15297.801(4079.753) | 15498.392(5079.096) | 15378.659(4090.812) |
| PC (42:10) | 2726.931(1374.175) | 2509.831(1360.724) | 2763.544(1424.331) | 2520.709(1364.459) |
| PC (o-32:0) - ESI(+) | 33066.939(8835.009) | 34463.696(8408.236) | 33492.212(9809.329) | 34464.353(8502.972) |
| PC (o-34:0) | 4253.752(1544.24) | 4625.498(1473.237) | 4415.904(1695.302) | 4629.054(1484.252) |
| PC (p-32:1)/PC (o-32:2) | 7732.065(2525.44) | 8442.017(2078.799) | 7881.476(2808.747) | 8457.282(2099.061) |
| PC (p-34:1)/PC (o-34:2) | 90514.308(41518.544) | 112732.145(42037.713) | 91475.293(46037.028) | 113376.226(42264.092) |
| PC (p-34:2)/PC (o-34:3) - ESI(+) | 134516.098(48579.028) | 167161.547(53529.226) | 133658.768(49629.539) | 167938.748(53754.934) |
| PC (p-36:1)/PC (o-36:2) B | 4576.41(1860.937) | 4990.913(1761.819) | 4699.805(1853.413) | 5003.991(1775.361) |
| PC (p-36:2)/PC (o-36:3) | 18401.743(7473.236) | 22260.671(8493.763) | 18842.485(7779.36) | 22351.138(8529.12) |
| PC (p-36:4)/PC (o-36:5) - ESI(+) | 222255.821(73873.841) | 244403.223(62140.636) | 225724.835(75604.567) | 245587.683(62296.124) |
| PC (p-36:5)/PC (o-36:6) | 43163.98(47010.214) | 35793.982(29043.548) | 43410.782(44710.87) | 36198.809(29253.43) |
| PC (p-38:3)/PC (o-38:4) A - ESI(+) | 23415.263(7747.924) | 26268.221(7020.386) | 24191.48(8087.438) | 26348.532(7078.493) |
| PC (p-38:3)/PC (o-38:4) B - ESI(+) | 60681.832(24417.325) | 68924.225(21887.47) | 64483.935(25968.209) | 69292.563(22001.673) |
| PC (p-38:4)/PC (o-38:5) A | 244736.068(73932.864) | 253186.358(52211.505) | 242645.006(75267.53) | 254571.544(51956.652) |
| PC (p-38:4)/PC (o-38:5) B | 46348.51(17350.154) | 52251.201(14810.562) | 48112.295(17621.65) | 52460.469(14869.201) |
| PC (p-38:5)/PC (o-38:6) A | 47602.588(24906.045) | 44907.468(17837.495) | 46699.307(23405.806) | 45238.272(17905.308) |
| PC (p-38:5)/PC (o-38:6) B | 60693.148(18465.69) | 63136.031(15203.868) | 59728.294(17931.149) | 63395.498(15228.685) |
| PC (p-38:6)/PC (o-38:7) | 38202.501(19743.59) | 36368.564(13760.086) | 37586.507(18596.862) | 36636.147(13802.699) |
| PC (p-40:4)/PC (o-40:5) - ESI(+) | 17184.561(5389.491) | 17672.249(4621.409) | 17633.138(5716.339) | 17791.313(4606.074) |
| PC (p-40:6)/PC (o-40:7) A | 40200.774(19957.817) | 35888.168(13738.726) | 38247.611(19252.016) | 36145.669(13781.758) |
| PC (p-40:6)/PC (o-40:7) B | 13510.106(6889.457) | 13100.064(5544.826) | 13676.333(7015.166) | 13204.922(5564.569) |
| PC (p-42:4)/PC (o-42:5) - ESI(+) | 12900.718(4496.163) | 12896.037(3917.285) | 12934.468(4555.335) | 12967.171(3931.021) |
| PC (p-42:5)/PC (o-42:6) A | 10237.461(3726.694) | 10713.657(3758.542) | 10267.451(3866.404) | 10795.536(3761.98) |
| PC (p-42:5)/PC (o-42:6) B | 3974.188(1415.292) | 3956.085(1237.45) | 3945.197(1361.956) | 3972.173(1245.398) |
| PC (p-44:4)/PC (o-44:5) - ESI(+) | 14453.858(5455.893) | 14223.731(4628.827) | 14571.775(5549.4) | 14252.115(4673.244) |
| PE (34:1) | 1935.048(1193.975) | 1997.27(1266.931) | 1904.402(1183.166) | 1990.722(1274.003) |
| PE (34:2) - ESI(-) | 2982.653(1876.385) | 3405.593(2151.45) | 3017.52(1905.946) | 3354.244(2128.94) |
| PE (36:1) - ESI(-) | 418.862(181.396) | 445.435(181.825) | 421.311(174.493) | 448.61(182.184) |
| PE (36:2) | 7905.774(4135.38) | 8803.532(4225.603) | 7810.351(4304.96) | 8670.866(4173.423) |
| PE (36:3) | 2026.786(1248.378) | 2279.738(1430.26) | 2010.435(1307.713) | 2242.802(1425.386) |
| PE (36:4) - ESI(-) | 2941.499(1644.752) | 3164.472(1950.982) | 2992.312(1611.669) | 3167.268(1969.338) |

**Supplementary Table S1.** Sample mean and standard deviations of metabolites in ME/CFS patients, controls, and their subgroups.

| Metabolite | ME/CFS (n=106) | Control (n=91) | ME/CFS without sr-IBS (n-71) | Control without sr-IBS (n-88) |
|---|---|---|---|---|
| | mean (SD) | mean (SD) | mean (SD) | mean (SD) |
| PE (38:2) | 629.602(222.186) | 682.787(206.229) | 629.87(231.692) | 684.371(207.976) |
| PE (38:6) - ESI(-) | 7437.759(5142.646) | 7356.489(4624.489) | 7280.063(4362.178) | 7337.96(4644.972) |
| PE (p-34:1)/PE (o-34:2) - ESI(-) | 2025.04(785.489) | 2051.91(695.945) | 1984.699(772.235) | 2041.719(699.616) |
| PE (p-34:2)/PE (o-34:3) | 3040.334(1576.283) | 3609.46(1747.103) | 2975.33(1597.431) | 3620.102(1765.852) |
| PE (p-36:1)/PE (o-36:2) - ESI(-) | 2760.373(1320.05) | 3068.12(1351.566) | 2780.87(1340.981) | 3059.064(1365.731) |
| PE (p-36:2)/PE (o-36:3) - ESI(-) | 5957.994(2773.012) | 6972.062(2936.918) | 5945.7(2826.471) | 6966.226(2968.837) |
| PE (p-36:4)/PE (o-36:5) - ESI(-) | 15261.909(9626.399) | 15928.908(8581.977) | 14636.588(9054.541) | 15918.27(8632.217) |
| PE (p-36:5)/PE (o-36:6) | 2102.097(2962.828) | 1485.236(2041.939) | 1988.288(2983.894) | 1502.625(2061.638) |
| PE (p-38:3)/PE (o-38:4) | 1340.345(687.934) | 1513.805(782.147) | 1359.457(685.127) | 1510.431(785.885) |
| PE (p-38:5)/PE (o-38:6) - ESI(-) | 11786.825(6628.023) | 12149.489(5341.659) | 11138.708(6249.873) | 12145.504(5396.945) |
| PE (p-38:6)/PE (o-38:7) | 7532.92(4292.66) | 7188.888(3364.912) | 6953.498(3689.32) | 7182.481(3397.962) |
| PE (p-40:4)/PE (o-40:5) | 3399.555(1897.801) | 3410.027(1799.437) | 3470.705(1947.091) | 3406.803(1797.404) |
| PE (34:2) - ESI(+) | 8757.929(6410.569) | 9879.682(7371.827) | 8750.926(6451.795) | 9744.42(7401.492) |
| PE (36:1) - ESI(+) | 3379.71(1611.842) | 3316.561(1592.325) | 3420.384(1649.593) | 3271.575(1581.854) |
| PE (36:4) - ESI(+) | 14739.008(7468.332) | 15255.178(7705.241) | 15153.184(7642.129) | 15253.619(7780.05) |
| PE (38:4) | 39570.697(19154.594) | 40390.086(15772.1) | 40941.692(20312.303) | 40415.795(15919.094) |
| PE (38:6) - ESI(+) | 20257.66(12022.688) | 19884.696(11866.824) | 19885.235(11213.456) | 19983.065(11974.331) |
| PE (p-34:1)/PE (o-34:2) - ESI(+) | 2742.254(1098.637) | 2862.174(974.659) | 2779.13(1152.772) | 2867.889(984.373) |
| PE (p-36:1)/PE (o-36:2) - ESI(+) | 2454.413(1024.933) | 2762.367(992.448) | 2525.096(1064.668) | 2771.035(995.888) |
| PE (p-36:2)/PE (o-36:3) - ESI(+) | 8991.903(3876.705) | 11041.406(4021.531) | 9186.075(4121.604) | 11064.28(4038.724) |
| PE (p-36:4)/PE (o-36:5) - ESI(+) | 29345.071(18055.185) | 30532.635(14508.762) | 30067.743(18241.048) | 30699.183(14625.534) |
| PE (p-38:4)/PE (o-38:5) | 42614.164(20321.576) | 46103.9(17411.189) | 44872.887(21563.563) | 46295.619(17564.043) |
| PE (p-38:5)/PE (o-38:6) - ESI(+) | 24747.834(11095.999) | 26262.641(9101.812) | 24924.424(11359.644) | 26372.865(9176.145) |
| PE (p-40:4)/PE (o-40:5) A | 2896.209(2724.733) | 2989.973(2175.181) | 3265.189(3067.35) | 2999.456(2195.18) |
| PE (p-40:4)/PE (o-40:5) B | 3708.007(1951.993) | 3692.493(1528.458) | 3894.973(2144.397) | 3697.527(1544.524) |
| PE (p-40:5)/PE (o-40:6) | 6740.897(2550.286) | 6806.706(1987.436) | 7168.298(2779.074) | 6822.697(2006.254) |
| PG (34:0)/PG (17:0/17:0) | 76853.368(21921.357) | 85280.313(20224.124) | 77047.644(24486.356) | 85397.434(20410.267) |
| PI (38:4)/PI (18:0-20:4) | 120817.481(39400.359) | 124008.066(31642.823) | 117906.601(39165.698) | 124171.713(31804.055) |
| SM (d30:1) - ESI(-) | 1704.845(1134.453) | 1947.17(774.108) | 1777.806(1209.982) | 1946.128(781.97) |
| SM (d32:0) - ESI(-) | 1032.341(809.55) | 1064.49(657.551) | 1052.653(876.669) | 1068.301(658.928) |
| SM (d32:1) - ESI(-) | 44492.705(15861.723) | 49754.267(12913.605) | 44905.165(15643.054) | 49583.07(12911.672) |
| SM (d32:2) - ESI(-) | 3030.003(1334.652) | 3230.614(1251.27) | 2989.558(1337.605) | 3215.642(1261.181) |
| SM (d33:1) - ESI(-) | 22975.489(8246.125) | 25365.753(7028.514) | 23436.193(7659.792) | 25236.624(7029.047) |
| SM (d34:0) - ESI(-) | 4927.976(1784.951) | 5032.83(1672.907) | 4838.108(1884.771) | 5023.758(1690.769) |
| SM (d34:1) - ESI(-) | 155715.48(39317.352) | 163369.808(34296.98) | 154490.25(40753.285) | 162869.741(34351.86) |
| SM (d34:2) - ESI(-) | 57475.881(14166.69) | 60371.724(13403.338) | 56155.122(14732.064) | 60082.503(13418.706) |
| SM (d36:0) - ESI(-) | 1066.803(653.242) | 879.715(483.034) | 1002.499(622.425) | 882.868(486.728) |
| SM (d36:1) - ESI(-) | 36132.26(10486.124) | 36986.044(8862.176) | 36042.616(11045.31) | 36600.281(8385.189) |
| SM (d36:2) - ESI(-) | 5807.249(1805.715) | 5799.331(1630.41) | 5816.71(1986.338) | 5725.611(1571.265) |
| SM (d36:3) - ESI(-) | 1713.798(653.047) | 1804.421(563.54) | 1683.146(688.603) | 1796.262(567.166) |
| SM (d37:1) | 2411.418(1019.95) | 2404.494(818.507) | 2448.956(995.601) | 2381.08(789.395) |
| SM (d38:0) | 696.441(811.576) | 522.617(447.127) | 658.1(856.002) | 528.797(449.782) |
| SM (d38:2) - ESI(-) | 4790.522(1380.077) | 4974.002(1318.512) | 4734.478(1521.276) | 4944.864(1317.226) |
| SM (d39:1) - ESI(-) | 12841.047(4278.992) | 14353.601(3784.665) | 12836.383(4287.686) | 14266.495(3690.375) |
| SM (d39:2) | 12449.201(3511.981) | 12954.634(3279.589) | 12344.23(3273.819) | 12936.232(3308.728) |
| SM (d40:1) - ESI(-) | 46897.381(13275.565) | 50082.888(10695.858) | 46268.966(13896.047) | 49880.885(10511.163) |
| SM (d40:2) A - ESI(-) | 12273.795(4442.543) | 12319.223(4118.364) | 12053.207(4935.659) | 12228.38(4042.72) |
| SM (d40:2) B - ESI(-) | 17613.206(5158.79) | 18864.027(4350.959) | 17514.243(5594.939) | 18798.146(4345.106) |

**Supplementary Table S1.** Sample mean and standard deviations of metabolites in ME/CFS patients, controls, and their subgroups.

| Metabolite | ME/CFS (n=106) | Control (n=91) | ME/CFS without sr-IBS (n-71) | Control without sr-IBS (n-88) |
|---|---|---|---|---|
| | mean (SD) | mean (SD) | mean (SD) | mean (SD) |
| SM (d40:3) | 1079.194(398.684) | 1151.905(387.017) | 1035.02(427.889) | 1147.152(390.039) |
| SM (d41:1) - ESI(-) | 11075.677(11704.719) | 13579.641(12424.293) | 11680.563(11652.556) | 13317.97(12444.504) |
| SM (d41:2) - ESI(-) | 10157.654(3759.068) | 10335.005(4166.592) | 9996.252(3844.751) | 10305.284(4191.64) |
| SM (d42:0) - ESI(+) | 631.117(319.733) | 572.32(282.695) | 591.761(300.097) | 578.069(283.225) |
| SM (d42:0) - ESI(-) | 2435.513(802.742) | 2325.825(740.385) | 2419.733(779.856) | 2327.178(747.955) |
| SM (d42:1) - ESI(-) | 74113.422(22000.912) | 76751.556(18783.673) | 72052.032(22585.309) | 76502.198(18837.417) |
| SM (d42:3) - ESI(-) | 45369.2(13437.893) | 44941.003(10551.682) | 43401.392(13356.33) | 44737.621(10584.166) |
| SM (d43:1) - ESI(-) | 1610.475(804.178) | 1785.517(763.855) | 1553.823(723.228) | 1779.224(769.915) |
| SM (d43:2) - ESI(-) | 6637.877(3329.003) | 6677.809(2804.014) | 6466.335(3236.283) | 6642.201(2791.712) |
| SM (d44:2) | 3401.174(1320.178) | 2954.684(961.942) | 3336.859(1363.665) | 2948.611(972.12) |
| SM (d41:3) | 17542.807(4858.342) | 18495.055(4033.934) | 17704.279(4931.166) | 18515.458(4062.311) |
| SM (d30:1) - ESI(+) | 4627.855(3294.462) | 5081.766(2205.718) | 4775.897(3718.25) | 5097.459(2227.118) |
| SM (d32:0) - ESI(+) | 4527.417(2257.972) | 4461.163(1515.584) | 4607.612(2496.391) | 4486.19(1523.778) |
| SM (d32:1) - ESI(+) | 127487.665(50390.527) | 141586.567(42157.93) | 130331.981(48929.228) | 141768.868(42598.043) |
| SM (d32:2) - ESI(+) | 8419.355(3072.987) | 8850.662(2752.191) | 8297.594(2942.597) | 8858.692(2771.314) |
| SM (d33:1) - ESI(+) | 78897.543(30030.413) | 81545.929(24155.104) | 80879.826(28114.586) | 81511.539(24404.167) |
| SM (d34:0) - ESI(+) | 77128.042(21734.185) | 77407.547(19252.704) | 75967.563(22440.613) | 77700.326(19302.556) |
| SM (d34:1) - ESI(+) | 1815521.625(427328.564) | 1839477.475(326129.156) | 1829807.294(456742.429) | 1839714.162(327659.875) |
| SM (d34:2) - ESI(+) | 169414.754(41285.046) | 172230.478(39764.32) | 166715.327(39577.835) | 171784.036(39237.286) |
| SM (d36:0) - ESI(+) | 18743.046(8907.96) | 15332.243(6070.249) | 18501.059(8850.863) | 15325.744(6136.489) |
| SM (d36:1) - ESI(+) | 282139.394(75455.508) | 277277.338(58280.773) | 286608.787(80810.636) | 276534.328(58672.045) |
| SM (d36:2) - ESI(+) | 637435.743(182402.554) | 634824.46(155709.619) | 645217.266(198451.742) | 631941.432(155065.972) |
| SM (d36:3) - ESI(+) | 6491.66(2413.088) | 6629.303(2061.528) | 6504.584(2499.182) | 6616.681(2057.058) |
| SM (d38:1) | 224074.217(52840.443) | 238184.289(46405.144) | 224889.924(55064.028) | 237892.274(46805.773) |
| SM (d38:2) - ESI(+) | 102238.769(23254.677) | 103697.728(23384.632) | 101377.833(24030.293) | 103522.414(23455.802) |
| SM (d39:1) - ESI(+) | 55126.946(19385.078) | 60979.282(17058.805) | 55798.226(18703.686) | 61069.426(17222.623) |
| SM (d40:0) | 8519.977(3597.278) | 7725.238(2745.348) | 8245.94(3178.193) | 7762.177(2765.776) |
| SM (d40:1) - ESI(+) | 357194.17(87098.783) | 373303.076(69403.177) | 356136.642(95201.351) | 373610.276(70067.044) |
| SM (d40:2) A - ESI(+) | 192003.867(74381.653) | 206939.521(72862.228) | 192417.37(77642.359) | 206037.77(73321.35) |
| SM (d40:2) B - ESI(+) | 235158.05(59785.384) | 255298.203(57750.75) | 234598.41(62433.707) | 255277.345(58240.397) |
| SM (d41:1) - ESI(+) | 136526.853(38416.919) | 143873.721(36214.448) | 136536.489(37662.407) | 143998.308(36485.875) |
| SM (d41:2) A - ESI(+) | 41382.821(14921.589) | 40584.006(12105.041) | 41213.339(14198.137) | 40572.482(12244.699) |
| SM (d41:2) B - ESI(+) | 65492.772(20330.14) | 72077.791(21032.207) | 65452.643(18795.602) | 72141.771(21186.803) |
| SM (d42:1) - ESI(+) | 227228.943(65564.52) | 235888.054(53178.336) | 226271.686(69919.87) | 236161.01(53650.016) |
| SM (d42:2) | 552276.944(187216.547) | 490026.241(121909.07) | 533141.681(191035.12) | 489051.667(121451.141) |
| SM (d42:3) - ESI(+) | 360096.856(99212.364) | 348735.256(79546.208) | 351337.078(102485.414) | 347985.851(79000.104) |
| SM (d43:1) - ESI(+) | 6015.056(2273.428) | 6198.991(2041.865) | 6143.595(2380.371) | 6211.676(2063.558) |
| SM (d43:2) - ESI(+) | 14891.052(6807.646) | 13945.815(5754.744) | 14853.778(6738.376) | 14011.678(5787.209) |
| TG (55:6) | 10089.48(6268.964) | 8140.342(3509.246) | 10836.292(6923.778) | 8162.42(3544.691) |
| TAG (58:7)/TAG (18:1/18:1/22:5) | 17350.361(7968.934) | 16648.95(5845.672) | 18379.414(8761.489) | 16682.467(5895.803) |
| TG (60:12) | 6594.761(13689.356) | 3710.704(4070.071) | 7251.689(15923.376) | 3747.563(4109.64) |
| TG (53:5) | 10973.54(5251.608) | 10286.984(3942.7) | 10701.711(5371.822) | 10283.557(3984.935) |
| TG (60:11) | 8428.304(11674.572) | 5497.847(3845.608) | 9231.739(13322.028) | 5529.571(3884.05) |
| TG (40:0) | 6969.903(22773.737) | 5975.792(11932.991) | 8766.071(27447.345) | 5516.076(11168.116) |
| TG (40:1) | 1931.434(4239.428) | 2087.013(3783.783) | 2330.745(5058.917) | 1968.038(3676.235) |
| TG (42:0) | 9234.522(21761.184) | 8844.978(15901.608) | 11245.866(25798.217) | 8330.903(15377.67) |
| TG (42:1) | 6797.568(17520.658) | 7739.975(16319.178) | 8371.053(20878.024) | 7245.426(15848.388) |
| TG (42:2) | 2571.971(5933.309) | 3829.423(10795.773) | 3024.04(6989.384) | 3574.733(10620.578) |

**Supplementary Table S1.** Sample mean and standard deviations of metabolites in ME/CFS patients, controls, and their subgroups.

| Metabolite | ME/CFS (n=106) | Control (n=91) | ME/CFS without sr-IBS (n-71) | Control without sr-IBS (n-88) |
|---|---|---|---|---|
| | mean (SD) | mean (SD) | mean (SD) | mean (SD) |
| TG (42:3) | 591.158(714.913) | 671.443(879.22) | 634.075(820.394) | 653.157(872.519) |
| TG (44:0) | 19087.048(32238.546) | 17886.319(25507.344) | 22032.526(36049.09) | 17298.772(25360.152) |
| TG (44:1) | 18788.092(29782.097) | 19478.318(28152.179) | 21422.802(33732.516) | 18739.362(27855.392) |
| TG (44:2) | 8060.679(14384.219) | 10064.354(23367.775) | 9291.989(16426.792) | 9627.643(23332.334) |
| TG (46:0) | 2913.683(2889.773) | 3210.711(2865.495) | 3263.752(3177.069) | 3137.386(2856.567) |
| TG (46:1) | 78988.174(99974.002) | 74842.169(86950.751) | 86568.493(106117.921) | 73084.458(87126.607) |
| TG (46:2) | 34399.438(37849.21) | 37666.18(47712.032) | 35611.907(37765.57) | 36780.009(47775.698) |
| TG (46:3) A | 601.482(563.369) | 733.247(1466.526) | 633.532(592.411) | 723.044(1481.594) |
| TG (46:3) B | 5520.005(7775.947) | 5107.884(7131.762) | 5695.095(8162.568) | 5062.03(7173.612) |
| TG (46:4) A | 1736.197(1706.675) | 2452.533(7382.425) | 1762.506(1820.052) | 2419.562(7461.742) |
| TG (46:4) B | 1520.942(1603.93) | 2160.803(6838.834) | 1542.796(1706.642) | 2126.776(6910.973) |
| TG (46:5) | 350.027(264.398) | 426.476(1213.469) | 333.912(252.928) | 427.762(1227.467) |
| TG (48:0) | 59633.947(40059.321) | 72255.424(51804.827) | 63645.305(44838.895) | 70978.459(51616.694) |
| TG (48:1) | 445353.036(404942.517) | 421062.403(327886.505) | 472991.329(429853.071) | 415044.497(326050.448) |
| TG (48:2) | 232753.491(212433.045) | 215309.555(160839.721) | 245026.006(228310.658) | 212704.831(161577.038) |
| TG (48:3) | 66257.783(45379.776) | 68608.011(52545.382) | 66098.171(46273.386) | 67099.49(51897.774) |
| TG (48:4) A | 14636.937(12229.537) | 16489.544(19911.708) | 13720.616(10557.492) | 16069.028(19760.771) |
| TG (48:4) B | 10569.309(10549.743) | 12439.978(18504.13) | 10642.263(9498.718) | 12501.835(18692.523) |
| TG (48:5) | 2394.932(2124.793) | 2607.435(2613.644) | 2220.184(1521.889) | 2584.862(2630.897) |
| TG (48:6) | 731.456(341.644) | 1120.169(4094.332) | 768.622(376.852) | 1126.843(4141.361) |
| TG (49:0) | 5475.884(3437.407) | 5547.724(2851.467) | 6051.479(3818.086) | 5464.309(2799.55) |
| TG (49:1) | 38039.35(31682.085) | 32431.609(18199.476) | 41285.123(34192.591) | 31982.711(17744.358) |
| TG (49:2) | 5347.71(2994.124) | 4908.392(3180.31) | 5728.394(3298.741) | 4879.799(3157.426) |
| TG (49:3) | 8782.798(5619.053) | 8058.518(3765.588) | 9036.136(6179.689) | 7963.304(3692.852) |
| TG (50:0) | 28294.36(17765.307) | 35864.991(22391.136) | 30391.144(18903.622) | 35660.291(22608.173) |
| TG (50:1) | 295740.881(378872.38) | 268871.222(259991.973) | 280015.863(359982.811) | 270562.524(262438.083) |
| TG (50:2) | 1473341.655(765417.549) | 1337335.537(606243.723) | 1496070.007(805949.723) | 1325579.751(598746.002) |
| TG (50:3) A | 599554.098(373888.427) | 538070.653(271710.362) | 600164.599(395106.976) | 535146.928(273168.136) |
| TG (50:3) B | 1463.989(2654.267) | 1979.068(6822.777) | 1763.535(3146.388) | 2011.907(6898.091) |
| TG (50:4) | 113705.603(61749.855) | 111373.426(49803.428) | 109116.643(60684.978) | 110796.918(50230.457) |
| TG (50:5) | 18164.523(11639.543) | 17487.784(9234.201) | 17215.323(11144.488) | 17459.143(9338.397) |
| TG (50:6) | 2907.741(1946.11) | 2679.302(1733.966) | 2888.373(1932.545) | 2676.27(1749.673) |
| TG (51:2) | 92658.55(49837.791) | 81298.301(33345.525) | 95995.942(52804.453) | 80607.855(32858.485) |
| TG (51:3) | 73828.122(33989.875) | 68061.653(24068.889) | 74772.231(36080.031) | 67624.149(24009.33) |
| TG (51:4) | 20600.271(10073.4) | 19733.727(8062.558) | 20408.26(10544.375) | 19674.057(8144.825) |
| TG (51:5) | 3397.515(1704.001) | 3356.461(1275.262) | 3369.705(1749.734) | 3379.77(1279.789) |
| TG (52:1) | 273344.812(258320.644) | 254133.337(220416.288) | 299612.616(284159.447) | 249925.333(217933.538) |
| TG (52:2) | 3924538.986(1182333.569) | 3636556.102(1122279.122) | 3897240.648(1199228.229) | 3618259.341(1114269.185) |
| TG (52:3) | 4416231.615(1252804.656) | 4180937.847(1101985.633) | 4352735.272(1257973.357) | 4163977.057(1105637.788) |
| TG (52:4) | 133068.448(44192.17) | 130479.658(39344.454) | 129212.885(42953.764) | 130221.218(39754.166) |
| TG (52:5) | 197791.574(102954.234) | 174359.995(84000.915) | 188522.527(102419.843) | 174809.025(84916.187) |
| TG (52:6) | 25178.432(15261.735) | 23719.028(10920.524) | 24829.566(14331.628) | 23815.35(11027.619) |
| TG (53:0) | 1583.982(681.915) | 1815.471(661.05) | 1653.076(772.232) | 1812.115(664.526) |
| TG (53:1) | 5936.774(4668.413) | 5776.819(3671.696) | 6579.165(5143.499) | 5657.963(3538.034) |
| TG (53:2) | 55792.745(24993.696) | 51317.499(21651.56) | 57210.703(25535.8) | 50865.024(21381.082) |
| TG (53:3) | 73056.718(28445.208) | 67703.814(23631.61) | 73975.978(29541.942) | 67458.574(23817.209) |
| TG (53:4) | 46389.39(19729.068) | 43856.196(15698.609) | 46487.513(21073.558) | 43726.327(15855.445) |
| TG (54:1) | 29942.881(33162.778) | 29095.13(22321.153) | 32980.657(38725.506) | 28604.443(22047.713) |

**Supplementary Table S1.** Sample mean and standard deviations of metabolites in ME/CFS patients, controls, and their subgroups.

| Metabolite | ME/CFS (n=106) | Control (n=91) | ME/CFS without sr-IBS (n-71) | Control without sr-IBS (n-88) |
|---|---|---|---|---|
| | mean (SD) | mean (SD) | mean (SD) | mean (SD) |
| TG (54:2) | 321515.5(253928.808) | 286249.499(201970.532) | 342510.274(287785.315) | 283770.114(202908.93) |
| TG (54:3) | 1412729.685(654867.912) | 1276837.39(524744.236) | 1418543.926(711952.268) | 1271328.969(528791.707) |
| TG (54:4) | 1525311.494(686168.793) | 1377217.291(578506.358) | 1497286.416(718331.673) | 1369710.413(579307.505) |
| TG (54:5) A | 886580.788(456096.063) | 802522.834(352287.756) | 859118.306(461089.103) | 799543.056(352238.975) |
| TG (54:5) B | 262000.181(111804.806) | 243734.88(92887.319) | 276603.512(117509.484) | 243850.79(93783.561) |
| TG (54:6) A | 199405.641(130496.852) | 176401.555(94248.554) | 189949.756(133588.756) | 176550.82(95020.055) |
| TG (54:6) B | 1358.531(998.464) | 1341.842(782.147) | 1267.351(943.621) | 1337.027(779.663) |
| TG (54:6) C | 156170.234(80514.484) | 137719.361(62448.4) | 164443.237(85300.433) | 138054.119(63101.913) |
| TG (54:7) A | 49122.361(41128.683) | 44915.907(27315.049) | 46148.658(42649.079) | 45158.064(27552.145) |
| TG (54:7) B | 43061.59(27306.858) | 34198.973(17471.92) | 44910.216(29325.904) | 34437.138(17594.818) |
| TG (54:8) | 7077.455(4680.131) | 6400.128(3571.251) | 6692.688(4452.706) | 6480.915(3565.906) |
| TG (54:9) | 965.383(952.873) | 800.15(740.888) | 954.254(976.091) | 812.865(744.458) |
| TG (55:1) | 2381.308(1476.992) | 2293.733(1193.428) | 2558.87(1643.717) | 2275.709(1190.663) |
| TG (55:2) | 3209.3(1454.698) | 3026.046(1288.231) | 3381.591(1556.957) | 2998.5(1282.234) |
| TG (56:1) | 8425.454(9238.656) | 8724.83(13724.801) | 8763.968(8320.807) | 8693.992(13861.114) |
| TG (56:10) | 3107.113(3323.17) | 2681.061(1812.641) | 3124.173(3634.468) | 2704.04(1827.021) |
| TG (56:2) | 12956.986(10569.613) | 12163.693(13180.75) | 13287.1(10099.284) | 12077.086(13305.387) |
| TG (56:3) | 30313.86(19743.679) | 27075.315(20697.58) | 30931.238(20382.621) | 26914.649(20902.266) |
| TG (56:4) | 52216.171(23781.081) | 49073.426(20687.965) | 51533.089(23278.094) | 48945.612(20887.754) |
| TG (56:5) A | 53791.601(25940.471) | 48027.297(18998.273) | 56305.333(27049.785) | 47916.754(19186.335) |
| TG (56:5) B | 56496.764(28288.186) | 50801.304(20794.293) | 59319.214(30578.439) | 50835.127(20990.189) |
| TG (56:5) C | 113050.899(43777.393) | 107344.31(43874.117) | 119231.419(47437.588) | 107123.484(44324.778) |
| TG (56:6) | 257540.767(82209.876) | 233475.447(74511.483) | 266989.082(87830.794) | 233954.485(75287.046) |
| TG (56:7) A | 847.239(514.565) | 783.159(401.886) | 872.146(560.019) | 784.776(405.716) |
| TG (56:7) B | 187264.223(134086.996) | 142003.825(96398.78) | 195785.76(145844.838) | 143108.577(97127.678) |
| TG (56:8) A | 48927.531(26901.231) | 42661.135(18087.989) | 50526.41(29262.934) | 42847.171(18199.922) |
| TG (56:8) B | 88914.046(72816.031) | 66492.66(53152.85) | 90886.292(77615.502) | 67041.347(53628.172) |
| TG (56:9) | 16134.849(11093.145) | 13921.157(6903.286) | 16389.574(11696.97) | 14029.73(6931.184) |
| TG (57:1) | 1925.193(905.225) | 1863.322(943.086) | 2052.23(993.706) | 1839.212(936.361) |
| TG (57:2) | 2561.985(1153.018) | 2479.474(1287.033) | 2624.876(1205.603) | 2467.196(1299.191) |
| TG (58:1) | 6517.314(7345.499) | 6747.301(10399.217) | 6592.938(5718.528) | 6753.942(10516.502) |
| TG (58:10) | 19441.101(14120.814) | 16598.368(8410.925) | 19887.224(14764.054) | 16693.484(8480.365) |
| TG (58:2) | 10622.347(16674.782) | 10834.266(23253.565) | 10431.712(12802.421) | 10856.796(23514.796) |
| TG (58:3) | 6147.029(9675.465) | 5932.174(10911.709) | 6045.156(6379.053) | 5963.783(11035.393) |
| TG (58:4) | 3243.624(2529.139) | 2899.892(2258.659) | 3302.313(2621.221) | 2902.309(2284.656) |
| TG (58:5) | 3750.517(1804.936) | 3824.305(1507.07) | 3880.535(1953.617) | 3827.191(1523.237) |
| TG (58:6) | 14653.583(5134.288) | 13533.44(3981.36) | 15097.802(5479.822) | 13527.491(4021.132) |
| TG (58:8) | 61621.544(53618.106) | 44303.591(28847.455) | 64513.107(56652.202) | 44689.771(29065.16) |
| TG (58:9) | 45461.573(35467.981) | 34450.345(20298.576) | 47193.161(37869.486) | 34669.934(20480.099) |
| TG (59:2) | 1491.779(1048.687) | 1415.968(980.975) | 1568.91(1122.262) | 1394.688(981.99) |
| TG (59:3) | 1482.159(789.741) | 1388.449(810.7) | 1535.439(841.044) | 1375.053(814.953) |
| TG (60:1) | 2548.677(2775.351) | 2609.795(3662.099) | 2503.608(1975.486) | 2618.535(3703.156) |
| TG (60:2) | 5106.678(6459.935) | 5391.177(12127.075) | 5543.014(7335.457) | 5404.857(12266.767) |
| TG (60:3) | 3356.443(4588.274) | 3498.227(6192.138) | 3203.246(3043.682) | 3529.114(6260.292) |
| TG (60:4) | 1479.004(1459.393) | 1489.448(1601.775) | 1466.126(1074.068) | 1490.622(1619.83) |
| TG (60:6) | 1058.884(479.188) | 956.092(409.713) | 1097.963(497.494) | 955.389(413.669) |
| TG (62:1) | 1025.712(1380.888) | 981.286(1057.831) | 948.483(779.416) | 979.935(1069.614) |
| TG (62:2) | 1814.769(2212.815) | 1781.426(2417.302) | 1743.47(1504.294) | 1779.345(2444.85) |

**Supplementary Table S1.** Sample mean and standard deviations of metabolites in ME/CFS patients, controls, and their subgroups.

| Metabolite | ME/CFS (n=106) | Control (n=91) | ME/CFS without sr-IBS (n-71) | Control without sr-IBS (n-88) |
|---|---|---|---|---|
| | mean (SD) | mean (SD) | mean (SD) | mean (SD) |
| TG (62:3) | 571.887(485.151) | 559.893(537.266) | 607.272(472.415) | 563.684(542.663) |
| TG (62:4) | 547.101(334.64) | 613.733(374.856) | 553.329(285.631) | 614.578(379.088) |
| TG (64:2) | 864.662(732.038) | 883.063(767.604) | 898.099(656.064) | 876.015(775.005) |
| TG (64:3) | 552.787(234.996) | 552.569(261.395) | 551.258(235.663) | 551.657(261.368) |
| **Oxylipins (OL)** | | | | |
| 10-nitrolinoleic acid | 0.186(0.36) | 0.149(0.221) | 0.164(0.299) | 0.152(0.224) |
| 10-nitrooleic acid | 5.022(4.039) | 4.734(2.95) | 5.109(4.537) | 4.796(2.976) |
| 11(12)-epoxy-5,8,14,17- eicosatetraenoic acid | 0.045(0.051) | 0.046(0.062) | 0.039(0.045) | 0.047(0.063) |
| 11,12-Dihydroxyicosa-5,8,14-trienoic acid | 0.729(0.365) | 0.798(0.415) | 0.749(0.395) | 0.797(0.415) |
| 11,12-Epoxyeicosa-5,8,14-trienoic acid | 0.298(0.235) | 0.25(0.225) | 0.305(0.239) | 0.247(0.221) |
| 11-Hydroxy-14,15-epoxyeicosatrienoic acid | 0.137(0.271) | 0.236(0.486) | 0.154(0.311) | 0.243(0.493) |
| 11-Hydroxy-arachidonic acid | 0.954(0.559) | 0.918(0.459) | 1.012(0.597) | 0.932(0.458) |
| 12(13)-epoxy-9,15-octadecadienoic acid | 0.131(0.14) | 0.155(0.255) | 0.146(0.149) | 0.155(0.258) |
| 12,13-dihydroxyoctadec-9-enoic acid | 3.714(2.623) | 4.151(2.308) | 3.81(2.822) | 4.104(2.326) |
| 12,13-dihydroxyoctadeca-9,15-dienoic acid | 0.107(0.148) | 0.111(0.152) | 0.106(0.149) | 0.114(0.154) |
| 12,13-epoxy-9-octadecenoic acid | 2.678(3.381) | 3.027(3.742) | 3.011(3.942) | 2.93(3.764) |
| 12-Hydroxy-5,8,10,14,17-eicosapentaenoic acid | 1.953(2.755) | 1.117(1.211) | 2.101(3.071) | 1.135(1.225) |
| 12-Hydroxy-5,8,10,14-eicosatetraenoic acid | 17.204(15.381) | 15.752(12.397) | 18.425(16.171) | 15.915(12.52) |
| 13-hydroxyoctadeca-9,11,15-trienoic acid | 0.935(0.885) | 0.869(0.593) | 0.995(1.017) | 0.848(0.589) |
| 13-Hydroxyoctadecadienoic acid | 19.586(7.961) | 19.825(7.897) | 19.316(8.205) | 19.491(7.649) |
| 13-ketooctadeca-9,11-dienoic acid | 11.779(7.152) | 12.915(7.243) | 11.225(7.239) | 12.965(7.347) |
| 14(15)-epoxy-5,8,11,17-eicosatetraenoic acid | 0.234(0.427) | 0.211(0.435) | 0.258(0.488) | 0.218(0.441) |
| 14,15-dihydroxyeicosa-5,8,11,17-tetraenoic acid | 0.445(0.453) | 0.503(0.551) | 0.434(0.453) | 0.512(0.558) |
| 14,15-dihydroxyeicosa-5,8,11-trienoic acid | 0.816(0.427) | 0.868(0.394) | 0.829(0.462) | 0.865(0.395) |
| 14-hydroxydocosa-4,7,10,12,16,19-hexaenoic acid | 13.05(14.814) | 10.01(9.899) | 13.681(15.635) | 10.17(9.989) |
| 15(16)-epoxy-9,12-octadecadienoic acid | 0.221(0.211) | 0.382(0.521) | 0.244(0.241) | 0.389(0.529) |
| 15,16-dihydroxyoctadeca-9,12-dienoic acid | 11.678(8.843) | 12.862(6.666) | 11.356(9.682) | 12.603(6.128) |
| 15-Deoxy-delta-12,14-Prostaglandin J2 | 0.043(0.036) | 0.05(0.053) | 0.043(0.04) | 0.05(0.054) |
| 15-hydroxyeicosa-5,8,11,13,17-pentaenoic acid | 0.387(0.417) | 0.288(0.303) | 0.416(0.473) | 0.294(0.306) |
| 15-hydroxyeicosa-5,8,11,13-tetraenoic acid | 3.04(1.536) | 2.905(1.282) | 3.109(1.576) | 2.925(1.285) |
| 15-ketoeicosa-5,8,11,13-tetraenoic acid | 0.159(0.211) | 0.167(0.276) | 0.151(0.228) | 0.171(0.279) |
| 15-Keto-prostaglandin E2 | 0.319(0.368) | 0.405(0.635) | 0.329(0.365) | 0.408(0.645) |
| 16(17)-epoxy-4,7,10,13,19-docosapentaenoic acid | 0.189(0.199) | 0.159(0.206) | 0.195(0.213) | 0.161(0.209) |
| 17,18-dihydroxyeicosa-5,8,11,14-tetraenoic acid | 6.057(5.778) | 4.785(3.299) | 5.77(4.594) | 4.834(3.34) |
| 17,18-Epoxy-5,8,11,14-eicosatetraenoic acid | 0.09(0.188) | 0.122(0.18) | 0.084(0.183) | 0.123(0.182) |
| 17-hydroxy-4,7,10,13,15,19-docosahexaenoic acid | 1.382(0.957) | 1.263(1.094) | 1.416(1.007) | 1.291(1.102) |
| 18-(3-ethyloxiran-2-yl)octadeca-4,7,10,13,16-pentaenoic acid | 0.406(0.383) | 0.403(0.512) | 0.382(0.393) | 0.408(0.519) |
| 19,20-dihydroxydocosa-4,7,10,13,16-pentaenoic acid | 1.929(1.019) | 1.91(1) | 1.897(1.084) | 1.934(1.007) |
| 20-Hydroxyarachidonic acid | 4.005(8.301) | 3.874(6.762) | 3.017(6.041) | 3.963(6.858) |
| 4-hydroxydocosa-5,7,10,13,16,19-hexaenoic acid | 0.763(0.83) | 0.673(0.657) | 0.854(0.96) | 0.686(0.661) |
| 5,15-dihydroxyeicosa-6,8,11,13-tetraenoic acid | 0.038(0.055) | 0.053(0.095) | 0.04(0.058) | 0.053(0.096) |
| 5,6,15-trihydroxyeicosa-7,9,11,13-tetraenoic acid | 0.157(0.194) | 0.207(0.32) | 0.163(0.194) | 0.211(0.325) |
| 5,6-dihydroxyeicosa-8,11,14-trienoic acid | 0.665(0.513) | 0.709(0.521) | 0.711(0.529) | 0.711(0.529) |
| 5-Hydroxy-6,8,11,14,17-eicosapentaenoic acid | 0.954(0.728) | 0.874(0.675) | 0.985(0.755) | 0.886(0.682) |
| 5-Hydroxy-6,8,11,14-eicosatetraenoic acid | 3.249(2.155) | 3.575(2.057) | 3.219(2.294) | 3.568(2.077) |
| 5-ketoeicosa-6,8,11,14-tetraenoic acid | 0.116(0.104) | 0.137(0.127) | 0.112(0.101) | 0.137(0.129) |
| 6-Ketoprostaglandin F1 alpha | 0.111(0.191) | 0.091(0.14) | 0.109(0.209) | 0.094(0.142) |
| 6-trans-Leukotriene B4 | 0.046(0.066) | 0.051(0.099) | 0.041(0.057) | 0.051(0.1) |

**Supplementary Table S1.** Sample mean and standard deviations of metabolites in ME/CFS patients, controls, and their subgroups.

| Metabolite | ME/CFS (n=106) | Control (n=91) | ME/CFS without sr-IBS (n-71) | Control without sr-IBS (n-88) |
|---|---|---|---|---|
| | mean (SD) | mean (SD) | mean (SD) | mean (SD) |
| 8,15-dihydroxyeicosa-5,9,11,13-tetraenoic acid | 0.121(0.272) | 0.115(0.202) | 0.12(0.307) | 0.117(0.205) |
| 8,9-dihydroxyeicosa-5,11,14-trienoic acid | 0.391(0.311) | 0.373(0.243) | 0.397(0.313) | 0.37(0.243) |
| 8,9-Epoxyeicosa-5,11,14-trienoic acid | 0.627(0.483) | 0.848(0.762) | 0.587(0.447) | 0.871(0.764) |
| 8-hydroxyeicosa-5,9,11,14-tetraenoic acid | 0.707(0.521) | 0.693(0.505) | 0.713(0.509) | 0.701(0.512) |
| 9(10)-epoxy-12,15-octadecadienoic acid | 1.238(2.543) | 1.495(3.452) | 1.369(3.033) | 1.518(3.506) |
| 9(10)-epoxy-12Z-octadecenoic acid | 0.778(0.586) | 0.789(0.455) | 0.783(0.656) | 0.775(0.453) |
| 9,10-dihydroxyoctadec-12-enoic acid | 3.994(2.996) | 4.527(2.771) | 3.997(3.128) | 4.483(2.803) |
| 9,10-dihydroxyoctadeca-12,15-dienoic acid | 0.195(0.127) | 0.233(0.156) | 0.201(0.139) | 0.234(0.156) |
| 9,10-Epoxystearic acid | 2.482(2.294) | 2.68(2.248) | 2.594(2.481) | 2.724(2.272) |
| 9,11,15-trihydroxy-5,13,1Z-prostatrienoic acid | 0.022(0.082) | 0.033(0.09) | 0.012(0.013) | 0.034(0.092) |
| 9,12,13-trihydroxyoctadec-10-enoic acid | 6.746(3.013) | 7.161(3.062) | 6.405(3.09) | 7.218(3.07) |
| 9-Hydroxy-5,7,11,14,17-icosapentaenoic acid | 0.259(0.225) | 0.218(0.148) | 0.258(0.231) | 0.222(0.149) |
| 9-hydroxyeicosa-5,7,11,14-tetraenoic acid | 0.366(0.266) | 0.386(0.267) | 0.361(0.261) | 0.388(0.268) |
| 9-Hydroxylinoleic acid | 9.759(4.207) | 10.407(3.998) | 9.531(4.066) | 10.248(3.945) |
| 9-hydroxyoctadeca-10,12,15-trienoic acid | 0.602(0.484) | 0.715(0.595) | 0.662(0.51) | 0.711(0.598) |
| 9-ketooctadeca-10,12-dienoic acid | 1.218(0.86) | 1.249(0.847) | 1.191(0.744) | 1.227(0.844) |
| 9-nitrooleic acid | 0.611(0.737) | 0.692(0.676) | 0.607(0.817) | 0.705(0.682) |
| 9S,10R-dihydroxy-stearic acid | 2.561(1.966) | 2.883(2.166) | 2.454(1.76) | 2.913(2.186) |
| Leukotriene B4 | 0.079(0.051) | 0.099(0.087) | 0.072(0.045) | 0.1(0.088) |
| Leukotriene B5 | 0.01(0.015) | 0.012(0.018) | 0.01(0.015) | 0.012(0.018) |
| Prostaglandin D2 | 0.336(0.565) | 0.54(0.959) | 0.318(0.552) | 0.525(0.939) |
| Prostaglandin E1 | 0.02(0.021) | 0.019(0.026) | 0.019(0.019) | 0.019(0.026) |
| Prostaglandin E2 | 0.057(0.126) | 0.052(0.08) | 0.057(0.128) | 0.054(0.081) |
| Prostaglandin E3 | 0.185(0.427) | 0.14(0.232) | 0.202(0.484) | 0.143(0.235) |
| Prostaglandin F2a | 0.83(0.921) | 0.952(0.717) | 0.637(0.59) | 0.963(0.723) |
| Resolvin D1 | 0.147(0.362) | 0.162(0.194) | 0.105(0.101) | 0.154(0.165) |
| Thromboxane B2 | 1.893(2.013) | 2.178(4.62) | 2.108(2.36) | 2.226(4.69) |
| trans-12,13-epoxy-11-oxo-trans-9-octadecenoic acid | 1.085(0.917) | 1.082(0.66) | 1.037(1.054) | 1.088(0.664) |

ME/CFS, myalgic encephalomyelitis/chronic fatigue syndrome; sr-IBS, self-reported physician diagnosed irritable bowel syndrome; SD, standard deviation

**Supplementary Table S2.** Regression and Bayesian estimates comparing all ME/CFS vs. all controls.

| Metabolite | Regression Model | ME/CFS vs. Controls | | | | | |
|---|---|---|---|---|---|---|---|
| | | Estimated Coefficient | 95% CI | p-value | FDR | BayesFactor | 95% HDI |
| **Primary Metabolites (PM)** | | | | | | | |
| succinic acid | Lognormal | 0.022 | (0.006, 0.038) | 0.009 | 0.177 | 1.216 | (0.006, 0.038) |
| arachidic acid | Lognormal | 0.024 | (0.004, 0.044) | 0.019 | 0.213 | 0.578 | (0.005, 0.045) |
| leucine | Lognormal | -0.010 | (-0.020, -0.001) | 0.035 | 0.287 | 0.312 | (-0.019, -0.001) |
| ornithine | Lognormal | -0.016 | (-0.032, -0.001) | 0.042 | 0.309 | 0.253 | (-0.032, -0.002) |
| alpha-ketoglutarate | Lognormal | 0.015 | (0.000, 0.031) | 0.056 | 0.361 | 0.211 | (0.000, 0.032) |
| mannitol | Lognormal | 0.029 | (-0.002, 0.060) | 0.067 | 0.396 | 0.187 | (-0.003, 0.059) |
| maleic acid | Lognormal | 0.026 | (-0.002, 0.053) | 0.075 | 0.409 | 0.169 | (-0.002, 0.054) |
| aminomalonate | Lognormal | 0.025 | (-0.003, 0.053) | 0.082 | 0.422 | 0.188 | (-0.004, 0.052) |
| tyrosine | Lognormal | -0.008 | (-0.016, 0.001) | 0.088 | 0.430 | 0.153 | (-0.016, 0.000) |
| glutaric acid | Lognormal | 0.017 | (-0.003, 0.037) | 0.100 | 0.461 | 0.138 | (-0.003, 0.036) |
| threonine | Lognormal | -0.010 | (-0.023, 0.003) | 0.146 | 0.551 | 0.093 | (-0.023, 0.003) |
| indole-3-propionic acid | Lognormal | -0.018 | (-0.043, 0.007) | 0.171 | 0.582 | 0.081 | (-0.042, 0.009) |
| benzoic acid | Lognormal | 0.011 | (-0.006, 0.028) | 0.207 | 0.624 | 0.081 | (-0.006, 0.028) |
| glycerol | Lognormal | 0.007 | (-0.004, 0.017) | 0.209 | 0.628 | 0.060 | (-0.004, 0.017) |
| glutamine | Lognormal | -0.012 | (-0.033, 0.008) | 0.235 | 0.667 | 0.066 | (-0.033, 0.007) |
| lysine | Lognormal | -0.015 | (-0.040, 0.011) | 0.262 | 0.697 | 0.054 | (-0.041, 0.010) |
| levoglucosan | Lognormal | -0.014 | (-0.038, 0.010) | 0.264 | 0.697 | 0.059 | (-0.037, 0.012) |
| pelargonic acid | Lognormal | 0.007 | (-0.006, 0.020) | 0.283 | 0.737 | 0.060 | (-0.005, 0.020) |
| 3-hydroxybutyric acid | Lognormal | 0.015 | (-0.014, 0.045) | 0.312 | 0.743 | 0.051 | (-0.013, 0.045) |
| threonic acid | Lognormal | 0.010 | (-0.009, 0.029) | 0.316 | 0.743 | 0.051 | (-0.009, 0.029) |
| urea | Lognormal | -0.005 | (-0.014, 0.005) | 0.320 | 0.743 | 0.056 | (-0.014, 0.005) |
| glucuronic acid | Lognormal | 0.010 | (-0.010, 0.030) | 0.325 | 0.746 | 0.055 | (-0.009, 0.032) |
| oxoproline | Lognormal | -0.003 | (-0.008, 0.003) | 0.335 | 0.748 | 0.049 | (-0.008, 0.003) |
| lysine | Lognormal | -0.009 | (-0.027, 0.009) | 0.341 | 0.754 | 0.049 | (-0.027, 0.009) |
| 2-deoxytetronic acid | Lognormal | 0.010 | (-0.011, 0.031) | 0.341 | 0.754 | 0.054 | (-0.011, 0.030) |
| pentadecanoic acid | Lognormal | 0.005 | (-0.005, 0.015) | 0.346 | 0.756 | 0.049 | (-0.005, 0.016) |
| phenylalanine | Lognormal | -0.005 | (-0.015, 0.005) | 0.350 | 0.760 | 0.049 | (-0.015, 0.006) |
| 2-hydroxybutanoic acid | Lognormal | 0.008 | (-0.009, 0.026) | 0.354 | 0.760 | 0.048 | (-0.009, 0.025) |
| beta-alanine | Lognormal | -0.010 | (-0.033, 0.012) | 0.363 | 0.764 | 0.048 | (-0.033, 0.012) |
| 2-ketoisocaproic acid | Lognormal | 0.005 | (-0.005, 0.015) | 0.368 | 0.764 | 0.046 | (-0.005, 0.014) |
| pyrrole-2-carboxylic acid | Lognormal | 0.010 | (-0.014, 0.034) | 0.393 | 0.780 | 0.048 | (-0.013, 0.034) |
| pseudo uridine | Lognormal | -0.005 | (-0.015, 0.006) | 0.395 | 0.781 | 0.050 | (-0.015, 0.006) |
| malic acid | Lognormal | 0.008 | (-0.012, 0.028) | 0.423 | 0.818 | 0.047 | (-0.011, 0.029) |
| citrulline | Lognormal | -0.005 | (-0.016, 0.007) | 0.429 | 0.822 | 0.044 | (-0.016, 0.007) |
| indole-3-lactate | Lognormal | -0.005 | (-0.019, 0.009) | 0.459 | 0.848 | 0.042 | (-0.019, 0.010) |
| maleimide | Lognormal | 0.007 | (-0.011, 0.025) | 0.463 | 0.848 | 0.041 | (-0.011, 0.026) |
| 1,2,4-benzenetriol | Lognormal | -0.013 | (-0.047, 0.021) | 0.463 | 0.848 | 0.047 | (-0.046, 0.021) |
| tryptophan | Lognormal | -0.006 | (-0.023, 0.011) | 0.470 | 0.853 | 0.040 | (-0.024, 0.010) |
| sucrose | Lognormal | 0.012 | (-0.021, 0.044) | 0.479 | 0.856 | 0.045 | (-0.019, 0.044) |
| fructose | Lognormal | -0.010 | (-0.037, 0.017) | 0.482 | 0.856 | 0.036 | (-0.038, 0.017) |
| salicylic acid | Lognormal | 0.010 | (-0.019, 0.039) | 0.487 | 0.856 | 0.042 | (-0.017, 0.039) |
| erythritol | Lognormal | 0.010 | (-0.019, 0.039) | 0.488 | 0.856 | 0.042 | (-0.019, 0.039) |
| alanine | Lognormal | -0.007 | (-0.026, 0.013) | 0.498 | 0.862 | 0.043 | (-0.027, 0.013) |
| quinic acid | Lognormal | -0.013 | (-0.052, 0.025) | 0.499 | 0.863 | 0.039 | (-0.050, 0.026) |
| isoleucine | Lognormal | -0.003 | (-0.014, 0.007) | 0.505 | 0.867 | 0.037 | (-0.013, 0.007) |
| glycerol-alpha-phosphate | Lognormal | 0.009 | (-0.017, 0.034) | 0.512 | 0.877 | 0.041 | (-0.016, 0.036) |

**Supplementary Table S2.** Regression and Bayesian estimates comparing all ME/CFS vs. all controls.

| Metabolite | Regression Model | ME/CFS vs. Controls | | | | | |
|---|---|---|---|---|---|---|---|
| | | Estimated Coefficient | 95% CI | p-value | FDR | BayesFactor | 95% HDI |
| glucose-1-phosphate | Lognormal | 0.005 | (-0.010, 0.020) | 0.525 | 0.883 | 0.038 | (-0.010, 0.020) |
| lactic acid | Lognormal | -0.005 | (-0.022, 0.011) | 0.525 | 0.883 | 0.039 | (-0.021, 0.012) |
| linoleic acid | Lognormal | 0.007 | (-0.015, 0.029) | 0.555 | 0.891 | 0.040 | (-0.016, 0.028) |
| gluconic acid | Lognormal | 0.004 | (-0.010, 0.019) | 0.558 | 0.893 | 0.040 | (-0.011, 0.019) |
| hydroxycarbamate NIST | Lognormal | -0.005 | (-0.024, 0.013) | 0.561 | 0.896 | 0.041 | (-0.022, 0.013) |
| uric acid | Lognormal | -0.005 | (-0.023, 0.013) | 0.597 | 0.914 | 0.038 | (-0.023, 0.013) |
| ribose | Lognormal | 0.004 | (-0.010, 0.017) | 0.597 | 0.914 | 0.038 | (-0.009, 0.018) |
| serine | Lognormal | -0.004 | (-0.018, 0.011) | 0.612 | 0.915 | 0.037 | (-0.018, 0.011) |
| stearic acid | Lognormal | -0.002 | (-0.009, 0.006) | 0.612 | 0.915 | 0.034 | (-0.009, 0.006) |
| lyxitol | Lognormal | -0.004 | (-0.020, 0.012) | 0.623 | 0.915 | 0.034 | (-0.019, 0.012) |
| glyceric acid | Lognormal | 0.005 | (-0.015, 0.025) | 0.625 | 0.915 | 0.036 | (-0.016, 0.024) |
| phthalic acid | Lognormal | 0.003 | (-0.011, 0.017) | 0.628 | 0.915 | 0.038 | (-0.010, 0.018) |
| phosphate | Lognormal | -0.004 | (-0.022, 0.013) | 0.629 | 0.915 | 0.037 | (-0.022, 0.013) |
| heptadecanoic acid | Lognormal | 0.004 | (-0.012, 0.019) | 0.631 | 0.915 | 0.038 | (-0.011, 0.020) |
| indole-3-acetate | Lognormal | 0.005 | (-0.015, 0.024) | 0.640 | 0.920 | 0.037 | (-0.013, 0.024) |
| conduritol-beta-expoxide | Lognormal | 0.007 | (-0.022, 0.035) | 0.649 | 0.925 | 0.038 | (-0.023, 0.033) |
| 4-hydroxybutyric acid | Lognormal | 0.004 | (-0.014, 0.023) | 0.651 | 0.925 | 0.040 | (-0.014, 0.022) |
| capric acid | Lognormal | -0.005 | (-0.029, 0.018) | 0.667 | 0.932 | 0.034 | (-0.029, 0.020) |
| glutamic acid | Lognormal | 0.003 | (-0.013, 0.020) | 0.683 | 0.932 | 0.033 | (-0.013, 0.021) |
| 2-hydroxyvaleric acid | Lognormal | 0.004 | (-0.014, 0.021) | 0.691 | 0.932 | 0.036 | (-0.015, 0.021) |
| methionine | Lognormal | -0.003 | (-0.021, 0.014) | 0.701 | 0.936 | 0.033 | (-0.021, 0.014) |
| fumaric acid | Lognormal | -0.003 | (-0.019, 0.013) | 0.709 | 0.936 | 0.036 | (-0.019, 0.014) |
| proline | Lognormal | -0.004 | (-0.026, 0.018) | 0.732 | 0.936 | 0.036 | (-0.026, 0.019) |
| creatinine | Lognormal | 0.003 | (-0.018, 0.025) | 0.747 | 0.936 | 0.033 | (-0.019, 0.023) |
| N-acetylornithine | Lognormal | -0.002 | (-0.013, 0.010) | 0.761 | 0.939 | 0.030 | (-0.013, 0.010) |
| threitol | Lognormal | 0.003 | (-0.014, 0.020) | 0.764 | 0.941 | 0.037 | (-0.014, 0.020) |
| adipic acid | Lognormal | -0.002 | (-0.021, 0.016) | 0.790 | 0.950 | 0.037 | (-0.021, 0.015) |
| myo-inositol | Lognormal | -0.001 | (-0.013, 0.010) | 0.803 | 0.950 | 0.037 | (-0.014, 0.011) |
| lauric acid | Lognormal | -0.003 | (-0.024, 0.018) | 0.805 | 0.950 | 0.031 | (-0.023, 0.018) |
| isopropylbenzene | Lognormal | 0.002 | (-0.016, 0.020) | 0.805 | 0.950 | 0.021 | (-0.015, 0.021) |
| isothreonic acid | Lognormal | 0.003 | (-0.021, 0.028) | 0.806 | 0.950 | 0.036 | (-0.021, 0.028) |
| behenic acid | Lognormal | -0.002 | (-0.016, 0.012) | 0.812 | 0.950 | 0.035 | (-0.016, 0.012) |
| ribonic acid | Lognormal | -0.002 | (-0.019, 0.015) | 0.822 | 0.950 | 0.033 | (-0.020, 0.015) |
| citric acid | Lognormal | -0.002 | (-0.018, 0.014) | 0.822 | 0.950 | 0.029 | (-0.017, 0.014) |
| 2-aminobutyric acid | Lognormal | -0.001 | (-0.014, 0.011) | 0.835 | 0.950 | 0.033 | (-0.014, 0.011) |
| pyruvic acid | Lognormal | -0.002 | (-0.017, 0.013) | 0.835 | 0.950 | 0.029 | (-0.017, 0.013) |
| palmitoleic acid | Lognormal | -0.003 | (-0.028, 0.023) | 0.840 | 0.950 | 0.035 | (-0.028, 0.023) |
| mannose | Lognormal | 0.001 | (-0.015, 0.017) | 0.866 | 0.963 | 0.034 | (-0.016, 0.017) |
| nicotinic acid | Lognormal | 0.003 | (-0.032, 0.037) | 0.868 | 0.963 | 0.028 | (-0.030, 0.038) |
| N-acetylputrescine | Lognormal | -0.001 | (-0.019, 0.016) | 0.883 | 0.966 | 0.032 | (-0.020, 0.016) |
| myristic acid | Lognormal | 0.001 | (-0.013, 0.015) | 0.887 | 0.968 | 0.037 | (-0.012, 0.016) |
| palmitic acid | Lognormal | -0.001 | (-0.008, 0.007) | 0.896 | 0.972 | 0.028 | (-0.008, 0.007) |
| 1-methylgalactose NIST | Lognormal | 0.002 | (-0.030, 0.034) | 0.897 | 0.973 | 0.033 | (-0.030, 0.035) |
| caprylic acid | Lognormal | 0.001 | (-0.012, 0.013) | 0.909 | 0.976 | 0.033 | (-0.012, 0.013) |
| creatine | Lognormal | 0.001 | (-0.022, 0.024) | 0.924 | 0.984 | 0.032 | (-0.022, 0.022) |
| trans-4-hydroxyproline | Lognormal | -0.001 | (-0.024, 0.021) | 0.925 | 0.984 | 0.032 | (-0.024, 0.020) |
| oxalic acid | Lognormal | -0.002 | (-0.045, 0.041) | 0.929 | 0.985 | 0.032 | (-0.045, 0.040) |

**Supplementary Table S2.** Regression and Bayesian estimates comparing all ME/CFS vs. all controls.

| Metabolite | Regression Model | ME/CFS vs. Controls | | | | | |
|---|---|---|---|---|---|---|---|
| | | Estimated Coefficient | 95% CI | p-value | FDR | BayesFactor | 95% HDI |
| valine | Lognormal | 0.000 | (-0.009, 0.009) | 0.930 | 0.985 | 0.032 | (-0.009, 0.008) |
| glucose | Lognormal | 0.000 | (-0.007, 0.007) | 0.936 | 0.987 | 0.031 | (-0.007, 0.007) |
| glycine | Lognormal | 0.000 | (-0.009, 0.009) | 0.938 | 0.987 | 0.030 | (-0.009, 0.009) |
| tagatose | Lognormal | 0.000 | (-0.017, 0.018) | 0.963 | 0.990 | 0.029 | (-0.018, 0.019) |
| glycolic acid | Lognormal | 0.000 | (-0.025, 0.026) | 0.980 | 0.992 | 0.033 | (-0.026, 0.025) |
| maltose | Lognormal | 0.000 | (-0.022, 0.022) | 0.988 | 0.994 | 0.031 | (-0.023, 0.022) |
| alloxanoic acid | Lognormal | 0.000 | (-0.030, 0.030) | 0.997 | 0.997 | 0.031 | (-0.030, 0.031) |
| **Biogenic Amines (BA)** | | | | | | | |
| Acetominophen | Lognormal | **0.068** | **(0.028, 0.108)** | **0.001** | **0.103** | **3.035** | **(0.026, 0.108)** |
| 2-Methylbutyryl-L-carnitine | Lognormal | -0.034 | (-0.055, -0.012) | 0.002 | 0.139 | 2.296 | (-0.055, -0.012) |
| Acyclovir | Lognormal | 0.112 | (0.038, 0.185) | 0.003 | 0.139 | 2.674 | (0.040, 0.185) |
| Caffeine | Lognormal | -0.083 | (-0.138, -0.028) | 0.003 | 0.139 | 2.445 | (-0.139, -0.030) |
| Guanine | Lognormal | 0.091 | (0.031, 0.152) | 0.003 | 0.139 | 2.203 | (0.031, 0.150) |
| Choline cation | Lognormal | -0.009 | (-0.015, -0.003) | 0.004 | 0.139 | 2.751 | (-0.015, -0.003) |
| Theobromine | Lognormal | -0.063 | (-0.108, -0.018) | 0.007 | 0.173 | 1.573 | (-0.107, -0.018) |
| Alprazolam | Lognormal | 0.063 | (0.016, 0.109) | 0.009 | 0.178 | 0.963 | (0.015, 0.109) |
| Piperine | Lognormal | -0.064 | (-0.115, -0.014) | 0.014 | 0.206 | 0.594 | (-0.112, -0.010) |
| Albendazole sulfoxide | Lognormal | 0.108 | (0.021, 0.194) | 0.015 | 0.208 | 0.673 | (0.024, 0.194) |
| Isopropylamine | Lognormal | -0.014 | (-0.024, -0.003) | 0.017 | 0.208 | 0.649 | (-0.024, -0.002) |
| 3-Methylglutarylcarnitine | Lognormal | -0.032 | (-0.059, -0.005) | 0.019 | 0.214 | 0.502 | (-0.057, -0.004) |
| Trigonelline | Lognormal | -0.044 | (-0.081, -0.006) | 0.023 | 0.225 | 0.552 | (-0.079, -0.006) |
| Ranitidine N-oxide | Lognormal | 0.055 | (0.006, 0.104) | 0.028 | 0.258 | 0.382 | (0.008, 0.103) |
| 7-Methylguanosine | Lognormal | -0.014 | (-0.027, -0.002) | 0.029 | 0.261 | 0.397 | (-0.026, -0.002) |
| H-Pro-Hyp-OH | Lognormal | 0.024 | (0.002, 0.045) | 0.031 | 0.268 | 0.362 | (0.004, 0.045) |
| 2,6-Diaminopimelic acid | Lognormal | 0.042 | (0.001, 0.084) | 0.049 | 0.341 | 0.222 | (-0.003, 0.081) |
| 4,5,7-Trihydroxyisoflavone | Lognormal | 0.082 | (0.001, 0.164) | 0.050 | 0.344 | 0.223 | (-0.001, 0.164) |
| SDMA | Lognormal | -0.009 | (-0.018, 0.000) | 0.055 | 0.361 | 0.191 | (-0.019, 0.000) |
| Methacholine cation | Lognormal | -0.018 | (-0.037, 0.001) | 0.058 | 0.367 | 0.213 | (-0.037, 0.000) |
| D-Turanose | Lognormal | -0.026 | (-0.054, 0.002) | 0.067 | 0.396 | 0.182 | (-0.052, 0.003) |
| H-gamma-glutamyl-glutamine | Lognormal | 0.023 | (-0.002, 0.047) | 0.068 | 0.397 | 0.156 | (0.000, 0.048) |
| Gabapentin | Lognormal | 0.047 | (-0.004, 0.099) | 0.074 | 0.409 | 0.164 | (-0.005, 0.097) |
| 3-Cysteinylacetaminophen | Lognormal | 0.081 | (-0.008, 0.169) | 0.075 | 0.409 | 0.157 | (-0.014, 0.161) |
| Betaine | Lognormal | -0.006 | (-0.012, 0.001) | 0.077 | 0.416 | 0.166 | (-0.012, 0.001) |
| Octanoylcarnitine | Lognormal | -0.017 | (-0.035, 0.002) | 0.077 | 0.416 | 0.134 | (-0.036, 0.002) |
| 3-Dehydrocarnitine | Lognormal | -0.013 | (-0.028, 0.002) | 0.084 | 0.430 | 0.129 | (-0.027, 0.002) |
| Lamotrigine; | Lognormal | 0.093 | (-0.013, 0.199) | 0.088 | 0.430 | 0.121 | (-0.011, 0.201) |
| Ranitidine | Lognormal | 0.035 | (-0.005, 0.075) | 0.088 | 0.430 | 0.140 | (-0.004, 0.074) |
| Kynurenine | Lognormal | -0.012 | (-0.026, 0.002) | 0.090 | 0.432 | 0.144 | (-0.026, 0.002) |
| Lauroyl-L-carnitine | Lognormal | -0.032 | (-0.070, 0.006) | 0.103 | 0.465 | 0.105 | (-0.070, 0.009) |
| Tri-2-ethylhexyl trimellitate | Lognormal | -0.038 | (-0.084, 0.008) | 0.107 | 0.468 | 0.132 | (-0.081, 0.008) |
| 4-Acetamidobutyric acid | Lognormal | -0.017 | (-0.037, 0.004) | 0.107 | 0.468 | 0.112 | (-0.038, 0.005) |
| Usnic acid | Lognormal | -0.036 | (-0.080, 0.008) | 0.108 | 0.471 | 0.125 | (-0.081, 0.007) |
| cis-10,11-Dihydroxy-10,11-dihydrocarbamazepine | Lognormal | -0.016 | (-0.036, 0.004) | 0.112 | 0.481 | 0.119 | (-0.036, 0.003) |
| Hydroxybupropion | Lognormal | -0.027 | (-0.059, 0.006) | 0.113 | 0.485 | 0.131 | (-0.060, 0.005) |
| p-Acetamidophenyl .beta.-D-glucuronide | Lognormal | 0.047 | (-0.011, 0.105) | 0.116 | 0.487 | 0.108 | (-0.016, 0.101) |
| (R)-Butyrylcarnitine | Lognormal | -0.016 | (-0.037, 0.005) | 0.135 | 0.531 | 0.087 | (-0.036, 0.005) |
| 3-Hydroxypyridine | Lognormal | -0.049 | (-0.113, 0.015) | 0.137 | 0.532 | 0.083 | (-0.113, 0.017) |

**Supplementary Table S2.** Regression and Bayesian estimates comparing all ME/CFS vs. all controls.

| Metabolite | Regression Model | ME/CFS vs. Controls | | | | | |
|---|---|---|---|---|---|---|---|
| | | Estimated Coefficient | 95% CI | p-value | FDR | BayesFactor | 95% HDI |
| L-Citrulline | Lognormal | -0.007 | (-0.016, 0.002) | 0.140 | 0.537 | 0.088 | (-0.017, 0.002) |
| N-Acetylhistidine | Lognormal | -0.015 | (-0.036, 0.005) | 0.150 | 0.554 | 0.091 | (-0.037, 0.005) |
| L-Tyrosine | Lognormal | -0.013 | (-0.031, 0.005) | 0.157 | 0.566 | 0.091 | (-0.030, 0.005) |
| Stachydrine | Lognormal | -0.025 | (-0.062, 0.011) | 0.169 | 0.582 | 0.087 | (-0.063, 0.010) |
| NEPSILON,NEPSILON,NEPSILON-TRIMETHYLLYSINE | Lognormal | -0.008 | (-0.020, 0.004) | 0.171 | 0.582 | 0.080 | (-0.021, 0.003) |
| Quetiapine | Lognormal | 0.017 | (-0.008, 0.042) | 0.178 | 0.591 | 0.081 | (-0.009, 0.040) |
| Linoleoylcarnitine | Lognormal | -0.022 | (-0.055, 0.010) | 0.179 | 0.591 | 0.078 | (-0.056, 0.009) |
| Trazodone | Lognormal | 0.027 | (-0.013, 0.067) | 0.189 | 0.599 | 0.078 | (-0.010, 0.071) |
| Urea | Lognormal | -0.006 | (-0.015, 0.003) | 0.196 | 0.606 | 0.066 | (-0.014, 0.004) |
| 2-Indolinone | Lognormal | -0.012 | (-0.031, 0.006) | 0.199 | 0.609 | 0.074 | (-0.031, 0.007) |
| N-.alpha.-Acetyl-L-arginine | Lognormal | -0.009 | (-0.023, 0.005) | 0.211 | 0.628 | 0.077 | (-0.023, 0.004) |
| Ergothioneine | Lognormal | -0.023 | (-0.058, 0.013) | 0.212 | 0.628 | 0.065 | (-0.057, 0.012) |
| Acetazolamide | Lognormal | -0.033 | (-0.085, 0.019) | 0.213 | 0.629 | 0.067 | (-0.086, 0.016) |
| 4'-Methyl-N-methylhexanophenone | Lognormal | -0.034 | (-0.087, 0.020) | 0.219 | 0.636 | 0.072 | (-0.086, 0.025) |
| Citrulline | Lognormal | -0.006 | (-0.016, 0.004) | 0.225 | 0.648 | 0.063 | (-0.015, 0.004) |
| Thr-Ile-Arg | Lognormal | -0.029 | (-0.076, 0.018) | 0.226 | 0.648 | 0.073 | (-0.076, 0.021) |
| L-Cystine | Lognormal | -0.013 | (-0.035, 0.009) | 0.235 | 0.667 | 0.046 | (-0.035, 0.008) |
| Ala-Ile | Lognormal | 0.011 | (-0.008, 0.030) | 0.238 | 0.671 | 0.064 | (-0.007, 0.031) |
| 7-Hydroxywarfarin | Lognormal | 0.018 | (-0.012, 0.048) | 0.247 | 0.685 | 0.066 | (-0.013, 0.048) |
| Prazepam | Lognormal | 0.024 | (-0.017, 0.065) | 0.248 | 0.685 | 0.052 | (-0.014, 0.070) |
| Fexofenadine | Lognormal | -0.015 | (-0.039, 0.010) | 0.248 | 0.685 | 0.067 | (-0.041, 0.010) |
| Testosterone | Lognormal | -0.019 | (-0.052, 0.013) | 0.250 | 0.687 | 0.060 | (-0.052, 0.014) |
| Pyrantel | Lognormal | -0.013 | (-0.035, 0.009) | 0.252 | 0.689 | 0.067 | (-0.035, 0.011) |
| Glutamic acid | Lognormal | 0.008 | (-0.006, 0.023) | 0.255 | 0.689 | 0.055 | (-0.006, 0.023) |
| Methioninesulfoxide | Lognormal | 0.011 | (-0.008, 0.030) | 0.257 | 0.692 | 0.057 | (-0.007, 0.030) |
| Creatinine | Lognormal | -0.003 | (-0.009, 0.002) | 0.273 | 0.721 | 0.050 | (-0.009, 0.002) |
| N.alpha.-Methyl-L-lysine | Lognormal | 0.019 | (-0.016, 0.054) | 0.281 | 0.735 | 0.057 | (-0.013, 0.055) |
| 1-Methylnicotinamide | Lognormal | -0.011 | (-0.030, 0.009) | 0.284 | 0.737 | 0.061 | (-0.030, 0.009) |
| Tyrosine | Lognormal | -0.007 | (-0.021, 0.006) | 0.286 | 0.737 | 0.059 | (-0.020, 0.005) |
| Ethiolat | Lognormal | -0.018 | (-0.051, 0.015) | 0.288 | 0.737 | 0.061 | (-0.052, 0.015) |
| Diazepam | Lognormal | 0.032 | (-0.027, 0.091) | 0.291 | 0.739 | 0.057 | (-0.028, 0.086) |
| L-Leucine, methyl ester | Lognormal | 0.018 | (-0.016, 0.052) | 0.293 | 0.739 | 0.047 | (-0.017, 0.051) |
| Guanidine | Lognormal | 0.010 | (-0.009, 0.029) | 0.293 | 0.739 | 0.055 | (-0.009, 0.028) |
| Ethylenediaminetetraacetic acid | Lognormal | 0.041 | (-0.036, 0.118) | 0.294 | 0.739 | 0.053 | (-0.035, 0.117) |
| Homoarginine; | Lognormal | 0.010 | (-0.008, 0.027) | 0.295 | 0.739 | 0.045 | (-0.007, 0.028) |
| Arginine | Lognormal | 0.007 | (-0.006, 0.019) | 0.302 | 0.740 | 0.052 | (-0.005, 0.020) |
| Bradykinin | Lognormal | 0.059 | (-0.053, 0.172) | 0.303 | 0.740 | 0.052 | (-0.053, 0.169) |
| Omeprazole sulfone N-oxide | Lognormal | 0.033 | (-0.030, 0.097) | 0.306 | 0.743 | 0.058 | (-0.024, 0.097) |
| 3-Methylxanthine | Lognormal | -0.022 | (-0.063, 0.020) | 0.310 | 0.743 | 0.058 | (-0.064, 0.019) |
| Hexanoyl-L-carnitine | Lognormal | -0.011 | (-0.031, 0.010) | 0.315 | 0.743 | 0.052 | (-0.032, 0.010) |
| Propionylcarnitine | Lognormal | -0.004 | (-0.013, 0.004) | 0.316 | 0.743 | 0.044 | (-0.013, 0.004) |
| Trimethylamine-N-oxide | Lognormal | 0.013 | (-0.012, 0.038) | 0.318 | 0.743 | 0.050 | (-0.011, 0.039) |
| Quetiapine sulfoxide | Lognormal | 0.016 | (-0.016, 0.048) | 0.322 | 0.743 | 0.054 | (-0.015, 0.048) |
| Indole-3-propionic acid | Lognormal | -0.014 | (-0.043, 0.014) | 0.327 | 0.747 | 0.052 | (-0.042, 0.016) |
| 3-Amino-1-propanol | Lognormal | 0.009 | (-0.009, 0.028) | 0.331 | 0.747 | 0.048 | (-0.010, 0.028) |
| Carnitine | Lognormal | -0.003 | (-0.010, 0.003) | 0.332 | 0.747 | 0.050 | (-0.010, 0.003) |
| 1-Acetyl-3-piperidinamine | Lognormal | 0.008 | (-0.009, 0.025) | 0.333 | 0.747 | 0.054 | (-0.008, 0.025) |

**Supplementary Table S2.** Regression and Bayesian estimates comparing all ME/CFS vs. all controls.

| Metabolite | Regression Model | ME/CFS vs. Controls | | | | | |
|---|---|---|---|---|---|---|---|
| | | Estimated Coefficient | 95% CI | p-value | FDR | BayesFactor | 95% HDI |
| 1-Monostearin | Lognormal | -0.017 | (-0.050, 0.017) | 0.335 | 0.748 | 0.049 | (-0.051, 0.017) |
| (3-Carboxypropyl)trimethylammonium cation | Lognormal | -0.005 | (-0.016, 0.006) | 0.344 | 0.756 | 0.049 | (-0.016, 0.006) |
| 1-Oleoyl-2-acetyl-sn-glycerol | Lognormal | -0.017 | (-0.053, 0.019) | 0.345 | 0.756 | 0.052 | (-0.054, 0.016) |
| 6-Methoxynaphthaleneacetic acid | Lognormal | -0.006 | (-0.018, 0.006) | 0.347 | 0.756 | 0.052 | (-0.018, 0.006) |
| N-Methylhistidine | Lognormal | -0.015 | (-0.047, 0.017) | 0.357 | 0.761 | 0.052 | (-0.048, 0.016) |
| D-.alpha.-Cyclohexylglycine | Lognormal | -0.013 | (-0.041, 0.015) | 0.365 | 0.764 | 0.048 | (-0.038, 0.018) |
| Histidine | Lognormal | -0.009 | (-0.030, 0.011) | 0.368 | 0.764 | 0.049 | (-0.031, 0.009) |
| 4-Pyridoxic acid; | Lognormal | 0.020 | (-0.024, 0.063) | 0.376 | 0.768 | 0.050 | (-0.022, 0.065) |
| 1-Oleoyl-sn-glycero-3-phosphoethanolamine | Lognormal | -0.015 | (-0.049, 0.019) | 0.376 | 0.768 | 0.045 | (-0.050, 0.017) |
| Oleoyl-L-carnitine | Lognormal | -0.016 | (-0.051, 0.019) | 0.377 | 0.768 | 0.048 | (-0.049, 0.021) |
| Norleucine | Lognormal | -0.005 | (-0.015, 0.006) | 0.380 | 0.768 | 0.047 | (-0.015, 0.006) |
| Lansoprazole | Lognormal | -0.042 | (-0.137, 0.053) | 0.384 | 0.771 | 0.045 | (-0.143, 0.048) |
| .beta.-Phenyl-.gamma.-aminobutyric acid | Lognormal | -0.008 | (-0.027, 0.010) | 0.384 | 0.771 | 0.050 | (-0.028, 0.011) |
| Topiramate | Lognormal | 0.013 | (-0.017, 0.043) | 0.399 | 0.786 | 0.043 | (-0.017, 0.041) |
| N-(3-(Aminomethyl)benzyl)acetamidine | Lognormal | -0.014 | (-0.045, 0.018) | 0.401 | 0.789 | 0.037 | (-0.044, 0.019) |
| Montelukast-1,2-diol | Lognormal | 0.030 | (-0.041, 0.100) | 0.412 | 0.808 | 0.044 | (-0.036, 0.102) |
| Omeprazole sulfone | Lognormal | 0.026 | (-0.037, 0.089) | 0.414 | 0.810 | 0.046 | (-0.042, 0.087) |
| Betonicine | Lognormal | -0.028 | (-0.094, 0.039) | 0.418 | 0.815 | 0.047 | (-0.095, 0.040) |
| Phenylacetyl-L-glutamine | Lognormal | -0.014 | (-0.046, 0.019) | 0.422 | 0.818 | 0.039 | (-0.043, 0.022) |
| Trimethoprim | Lognormal | -0.011 | (-0.039, 0.016) | 0.423 | 0.818 | 0.044 | (-0.038, 0.018) |
| Ticlopidine | Lognormal | -0.033 | (-0.115, 0.049) | 0.430 | 0.822 | 0.042 | (-0.121, 0.042) |
| alpha-Methylhistidine; | Lognormal | -0.018 | (-0.064, 0.028) | 0.435 | 0.828 | 0.046 | (-0.066, 0.027) |
| N-Acetyl-L-carnosine | Lognormal | -0.007 | (-0.026, 0.011) | 0.439 | 0.829 | 0.039 | (-0.025, 0.012) |
| Metoprolol acid | Lognormal | 0.021 | (-0.033, 0.074) | 0.449 | 0.845 | 0.045 | (-0.030, 0.076) |
| Phenylalanine | Lognormal | -0.003 | (-0.010, 0.005) | 0.455 | 0.848 | 0.040 | (-0.009, 0.005) |
| L-Threonine | Lognormal | -0.004 | (-0.015, 0.007) | 0.456 | 0.848 | 0.041 | (-0.015, 0.007) |
| Nudifloramide | Lognormal | -0.007 | (-0.025, 0.011) | 0.460 | 0.848 | 0.044 | (-0.025, 0.012) |
| Dexpanthenol | Lognormal | -0.011 | (-0.041, 0.019) | 0.462 | 0.848 | 0.043 | (-0.040, 0.019) |
| Aminodiphenylmethane | Lognormal | 0.013 | (-0.022, 0.047) | 0.469 | 0.853 | 0.041 | (-0.021, 0.046) |
| Coniferylaldehyde | Lognormal | 0.008 | (-0.014, 0.030) | 0.475 | 0.856 | 0.042 | (-0.014, 0.030) |
| 1-Stearoyl-2-arachidonoyl-sn-glycero-3-phosphoserine | Lognormal | -0.029 | (-0.109, 0.051) | 0.483 | 0.856 | 0.035 | (-0.111, 0.051) |
| 3,4-Dimethoxybenzaldehyde | Lognormal | 0.011 | (-0.020, 0.041) | 0.486 | 0.856 | 0.036 | (-0.018, 0.041) |
| 2-Amino-1-phenylethanol | Lognormal | -0.003 | (-0.010, 0.005) | 0.487 | 0.856 | 0.040 | (-0.010, 0.005) |
| 7.alpha.-Hydroxy-3-oxo-4-cholestenoic acid | Lognormal | -0.007 | (-0.028, 0.013) | 0.488 | 0.856 | 0.042 | (-0.027, 0.012) |
| N.epsilon.-Methyl-L-lysine | Lognormal | 0.010 | (-0.019, 0.039) | 0.490 | 0.856 | 0.043 | (-0.019, 0.039) |
| N-Acetyl-D-norleucine | Lognormal | 0.008 | (-0.014, 0.030) | 0.492 | 0.856 | 0.027 | (-0.015, 0.029) |
| Benthiavalicarb-isopropyl | Lognormal | -0.012 | (-0.049, 0.025) | 0.514 | 0.879 | 0.040 | (-0.050, 0.025) |
| Creatine | Lognormal | 0.005 | (-0.009, 0.019) | 0.518 | 0.882 | 0.035 | (-0.010, 0.019) |
| Albendazole | Lognormal | 0.009 | (-0.019, 0.037) | 0.528 | 0.886 | 0.043 | (-0.019, 0.036) |
| 4-Aminomethylcyclohexanecarboxylic acid; | Lognormal | -0.009 | (-0.037, 0.019) | 0.529 | 0.886 | 0.038 | (-0.036, 0.019) |
| Palmitamide | Lognormal | -0.012 | (-0.050, 0.026) | 0.533 | 0.890 | 0.044 | (-0.052, 0.027) |
| Ranitidine-S-oxide | Lognormal | 0.017 | (-0.038, 0.072) | 0.543 | 0.891 | 0.042 | (-0.038, 0.070) |
| D-erythro-Sphingosine-1-phosphate | Lognormal | -0.007 | (-0.030, 0.016) | 0.548 | 0.891 | 0.035 | (-0.031, 0.016) |
| Metoprolol | Lognormal | 0.013 | (-0.029, 0.054) | 0.553 | 0.891 | 0.039 | (-0.027, 0.058) |
| Tryptophan | Lognormal | 0.003 | (-0.007, 0.013) | 0.564 | 0.899 | 0.037 | (-0.007, 0.013) |
| 1-Methyladenosine A | Lognormal | -0.003 | (-0.012, 0.006) | 0.572 | 0.901 | 0.038 | (-0.011, 0.007) |
| rac-4-Sulfoxypropranolol | Lognormal | 0.008 | (-0.021, 0.037) | 0.579 | 0.907 | 0.039 | (-0.021, 0.036) |

**Supplementary Table S2.** Regression and Bayesian estimates comparing all ME/CFS vs. all controls.

| Metabolite | Regression Model | ME/CFS vs. Controls | | | | | |
|---|---|---|---|---|---|---|---|
| | | Estimated Coefficient | 95% CI | p-value | FDR | BayesFactor | 95% HDI |
| Modafinil acid | Lognormal | 0.017 | (-0.042, 0.075) | 0.581 | 0.907 | 0.041 | (-0.039, 0.077) |
| 5'-S-Methyl-5'-thioadenosine | Lognormal | -0.020 | (-0.092, 0.052) | 0.583 | 0.907 | 0.032 | (-0.093, 0.048) |
| Metformin | Lognormal | 0.009 | (-0.023, 0.041) | 0.587 | 0.909 | 0.038 | (-0.025, 0.041) |
| Pyridoxal | Lognormal | 0.006 | (-0.017, 0.030) | 0.589 | 0.911 | 0.039 | (-0.018, 0.029) |
| Serotonin | Lognormal | 0.005 | (-0.014, 0.024) | 0.590 | 0.911 | 0.033 | (-0.014, 0.025) |
| Theanine; | Lognormal | 0.009 | (-0.023, 0.040) | 0.591 | 0.911 | 0.033 | (-0.021, 0.043) |
| Irbesartan | Lognormal | 0.006 | (-0.016, 0.028) | 0.594 | 0.913 | 0.038 | (-0.015, 0.028) |
| 1-Palmitoyl-2-hydroxy-sn-glycero-3-phosphoethanolamine | Lognormal | -0.009 | (-0.044, 0.026) | 0.615 | 0.915 | 0.035 | (-0.045, 0.027) |
| 3-Hydroxyoleylcarnitine | Lognormal | 0.019 | (-0.054, 0.092) | 0.616 | 0.915 | 0.033 | (-0.054, 0.091) |
| Telmisartan | Lognormal | 0.007 | (-0.021, 0.035) | 0.617 | 0.915 | 0.034 | (-0.020, 0.035) |
| Ile-Glu-Arg | Lognormal | -0.016 | (-0.080, 0.047) | 0.619 | 0.915 | 0.039 | (-0.079, 0.048) |
| N8-Acetylspermidine | Lognormal | -0.003 | (-0.017, 0.010) | 0.619 | 0.915 | 0.032 | (-0.017, 0.010) |
| Codeine-6-.beta.-D-glucuronide | Lognormal | 0.007 | (-0.021, 0.035) | 0.621 | 0.915 | 0.039 | (-0.020, 0.035) |
| Tauroursodeoxycholic acid | Lognormal | 0.009 | (-0.025, 0.042) | 0.622 | 0.915 | 0.036 | (-0.024, 0.043) |
| 1,2-Dimethylimidazole | Lognormal | -0.005 | (-0.025, 0.015) | 0.630 | 0.915 | 0.041 | (-0.025, 0.015) |
| Hypoxanthine | Lognormal | -0.004 | (-0.020, 0.012) | 0.636 | 0.920 | 0.031 | (-0.021, 0.012) |
| DL-Indole-3-lactic acid | Lognormal | 0.003 | (-0.008, 0.013) | 0.640 | 0.920 | 0.036 | (-0.008, 0.013) |
| Sulfamethoxazole | Lognormal | -0.019 | (-0.100, 0.061) | 0.643 | 0.922 | 0.037 | (-0.101, 0.061) |
| Heptadecasphing-4-enine | Lognormal | 0.025 | (-0.084, 0.134) | 0.652 | 0.925 | 0.031 | (-0.084, 0.133) |
| Methylgallate | Lognormal | 0.018 | (-0.062, 0.098) | 0.652 | 0.925 | 0.037 | (-0.061, 0.099) |
| L-Cysteine-glutathione disulfide | Lognormal | -0.006 | (-0.034, 0.022) | 0.668 | 0.932 | 0.023 | (-0.034, 0.023) |
| Atenolol | Lognormal | -0.009 | (-0.050, 0.032) | 0.670 | 0.932 | 0.035 | (-0.048, 0.034) |
| 6-Hydroxyflavone | Lognormal | 0.008 | (-0.030, 0.047) | 0.672 | 0.932 | 0.034 | (-0.029, 0.050) |
| Pyridoxine; | Lognormal | 0.007 | (-0.026, 0.040) | 0.673 | 0.932 | 0.038 | (-0.028, 0.040) |
| 1-Stearoyl-2-arachidonoyl-sn-glycero-3-phospho-(1'-myo-inositol) | Lognormal | -0.019 | (-0.106, 0.068) | 0.675 | 0.932 | 0.034 | (-0.105, 0.063) |
| Ethyldiethanolamine | Lognormal | -0.006 | (-0.032, 0.021) | 0.679 | 0.932 | 0.033 | (-0.031, 0.021) |
| Proline | Lognormal | -0.005 | (-0.030, 0.020) | 0.681 | 0.932 | 0.036 | (-0.029, 0.022) |
| Glaucine | Lognormal | 0.004 | (-0.016, 0.025) | 0.683 | 0.932 | 0.035 | (-0.016, 0.025) |
| Betaine aldehyde cation | Lognormal | 0.010 | (-0.041, 0.062) | 0.689 | 0.932 | 0.032 | (-0.038, 0.063) |
| (2R)-3-Hydroxyisovaleroylcarnitine | Lognormal | -0.003 | (-0.021, 0.014) | 0.692 | 0.932 | 0.032 | (-0.020, 0.014) |
| Milnacipran | Lognormal | 0.007 | (-0.029, 0.042) | 0.700 | 0.936 | 0.033 | (-0.028, 0.044) |
| Carbamazepine | Lognormal | -0.010 | (-0.060, 0.041) | 0.709 | 0.936 | 0.036 | (-0.059, 0.042) |
| Biliverden | Lognormal | -0.004 | (-0.023, 0.016) | 0.709 | 0.936 | 0.035 | (-0.022, 0.016) |
| trans-3'-Hydroxycotinine | Lognormal | 0.006 | (-0.025, 0.037) | 0.721 | 0.936 | 0.033 | (-0.026, 0.036) |
| Adenosine | Lognormal | -0.005 | (-0.033, 0.023) | 0.721 | 0.936 | 0.032 | (-0.034, 0.022) |
| 3-Hydroxybutyrylcarnitine | Lognormal | -0.005 | (-0.033, 0.023) | 0.724 | 0.936 | 0.034 | (-0.032, 0.025) |
| 1-Acetyl-4-piperidinamine | Lognormal | 0.007 | (-0.031, 0.045) | 0.725 | 0.936 | 0.037 | (-0.032, 0.045) |
| 3-Pyridinemthanol | Lognormal | 0.004 | (-0.018, 0.026) | 0.726 | 0.936 | 0.036 | (-0.018, 0.026) |
| Avobenzone | Lognormal | -0.004 | (-0.028, 0.020) | 0.731 | 0.936 | 0.034 | (-0.029, 0.019) |
| Lysine | Lognormal | 0.002 | (-0.010, 0.014) | 0.733 | 0.936 | 0.025 | (-0.010, 0.014) |
| Triptolide | Lognormal | -0.004 | (-0.025, 0.018) | 0.738 | 0.936 | 0.034 | (-0.025, 0.017) |
| 1-Methyl-L-histidine | Lognormal | -0.007 | (-0.050, 0.036) | 0.742 | 0.936 | 0.036 | (-0.049, 0.040) |
| Androstan-3-ol-17-one 3-glucuronide | Lognormal | 0.005 | (-0.025, 0.035) | 0.743 | 0.936 | 0.034 | (-0.025, 0.033) |
| 1-Stearoyl-2-hydroxy-sn-glycero-3-phosphoethanolamine | Lognormal | 0.006 | (-0.030, 0.042) | 0.744 | 0.936 | 0.031 | (-0.031, 0.040) |
| Cotinine N-.beta.-D-glucuronide | Lognormal | 0.010 | (-0.052, 0.073) | 0.745 | 0.936 | 0.032 | (-0.053, 0.072) |
| R-(-)-O-Desmethylvenlafaxine | Lognormal | -0.007 | (-0.052, 0.037) | 0.746 | 0.936 | 0.036 | (-0.053, 0.036) |
| Meprobamate | Lognormal | 0.005 | (-0.026, 0.037) | 0.750 | 0.936 | 0.035 | (-0.024, 0.037) |

**Supplementary Table S2.** Regression and Bayesian estimates comparing all ME/CFS vs. all controls.

| Metabolite | Regression Model | ME/CFS vs. Controls | | | | | |
|---|---|---|---|---|---|---|---|
| | | Estimated Coefficient | 95% CI | p-value | FDR | BayesFactor | 95% HDI |
| Tapentadol-.beta.-D-glucuronide | Lognormal | -0.010 | (-0.068, 0.049) | 0.750 | 0.936 | 0.035 | (-0.067, 0.050) |
| Acetyl-DL-carnitine | Lognormal | 0.002 | (-0.012, 0.017) | 0.752 | 0.936 | 0.035 | (-0.013, 0.017) |
| 3-(1-Pyrazolyl)-alanine | Lognormal | 0.010 | (-0.054, 0.074) | 0.754 | 0.936 | 0.035 | (-0.051, 0.072) |
| Toradol | Lognormal | 0.006 | (-0.031, 0.043) | 0.755 | 0.936 | 0.035 | (-0.033, 0.042) |
| Pipecolic acid | Lognormal | -0.006 | (-0.044, 0.032) | 0.757 | 0.938 | 0.036 | (-0.045, 0.031) |
| Esomeprazole | Lognormal | 0.004 | (-0.025, 0.034) | 0.768 | 0.944 | 0.033 | (-0.024, 0.035) |
| Meloxicam | Lognormal | 0.016 | (-0.092, 0.125) | 0.769 | 0.944 | 0.036 | (-0.100, 0.123) |
| 3-Aminoquinoline | Lognormal | 0.006 | (-0.034, 0.046) | 0.775 | 0.947 | 0.025 | (-0.034, 0.048) |
| .epsilon.-Caprolactam | Lognormal | 0.002 | (-0.014, 0.019) | 0.775 | 0.947 | 0.020 | (-0.015, 0.019) |
| Scopoletin | Lognormal | -0.011 | (-0.087, 0.065) | 0.777 | 0.947 | 0.028 | (-0.087, 0.064) |
| threo-Dihydrobupropion | Lognormal | -0.004 | (-0.031, 0.023) | 0.779 | 0.949 | 0.033 | (-0.029, 0.023) |
| Benzophenone-3 | Lognormal | -0.008 | (-0.063, 0.047) | 0.782 | 0.949 | 0.035 | (-0.067, 0.045) |
| Pantothenic acid | Lognormal | 0.003 | (-0.019, 0.025) | 0.786 | 0.950 | 0.033 | (-0.018, 0.024) |
| 5'-S-Methylthioadenosine | Lognormal | -0.011 | (-0.089, 0.068) | 0.787 | 0.950 | 0.031 | (-0.088, 0.064) |
| Gly-Pro-Arg | Lognormal | -0.008 | (-0.072, 0.055) | 0.794 | 0.950 | 0.036 | (-0.075, 0.052) |
| Lauric acid diethanolamide | Lognormal | 0.006 | (-0.037, 0.048) | 0.797 | 0.950 | 0.030 | (-0.036, 0.046) |
| Temazepam | Lognormal | 0.005 | (-0.033, 0.042) | 0.803 | 0.950 | 0.032 | (-0.033, 0.040) |
| Ser-Tyr-Lys | Lognormal | -0.007 | (-0.065, 0.051) | 0.809 | 0.950 | 0.035 | (-0.065, 0.049) |
| Ondansetron | Lognormal | -0.003 | (-0.024, 0.019) | 0.816 | 0.950 | 0.034 | (-0.025, 0.019) |
| Glycocholic acid | Lognormal | -0.004 | (-0.040, 0.032) | 0.817 | 0.950 | 0.034 | (-0.041, 0.032) |
| N-Methylproline | Lognormal | -0.003 | (-0.027, 0.021) | 0.821 | 0.950 | 0.035 | (-0.028, 0.021) |
| Borrelidin | Lognormal | -0.005 | (-0.054, 0.043) | 0.824 | 0.950 | 0.033 | (-0.052, 0.044) |
| Isopentenyladenine | Lognormal | -0.001 | (-0.010, 0.008) | 0.826 | 0.950 | 0.033 | (-0.010, 0.008) |
| Naproxen | Lognormal | 0.005 | (-0.037, 0.047) | 0.827 | 0.950 | 0.035 | (-0.038, 0.049) |
| Urocanic acid; | Lognormal | -0.002 | (-0.020, 0.016) | 0.828 | 0.950 | 0.031 | (-0.020, 0.017) |
| N-Acetylalanine | Lognormal | 0.006 | (-0.052, 0.064) | 0.834 | 0.950 | 0.033 | (-0.048, 0.066) |
| Dinor-12-oxophytodienoic acid | Lognormal | -0.005 | (-0.050, 0.041) | 0.837 | 0.950 | 0.034 | (-0.053, 0.040) |
| Modafinil | Lognormal | 0.005 | (-0.041, 0.050) | 0.839 | 0.950 | 0.032 | (-0.040, 0.050) |
| Penciclovir | Lognormal | 0.002 | (-0.025, 0.030) | 0.862 | 0.963 | 0.028 | (-0.026, 0.029) |
| 2-Hydroxyibuprofen | Lognormal | -0.003 | (-0.036, 0.030) | 0.865 | 0.963 | 0.035 | (-0.036, 0.030) |
| D-Pyroglutamic acid | Lognormal | 0.000 | (-0.006, 0.005) | 0.867 | 0.963 | 0.031 | (-0.005, 0.005) |
| Matrine | Lognormal | 0.004 | (-0.041, 0.049) | 0.871 | 0.963 | 0.033 | (-0.041, 0.049) |
| Losartan | Lognormal | 0.003 | (-0.035, 0.042) | 0.874 | 0.963 | 0.036 | (-0.036, 0.040) |
| Ornithine | Lognormal | -0.001 | (-0.017, 0.015) | 0.875 | 0.963 | 0.025 | (-0.017, 0.014) |
| Mefenorex | Lognormal | -0.003 | (-0.039, 0.033) | 0.876 | 0.963 | 0.030 | (-0.040, 0.032) |
| N.epsilon.-Acetyl-L-lysine | Lognormal | -0.001 | (-0.016, 0.014) | 0.882 | 0.965 | 0.032 | (-0.016, 0.015) |
| Ezetimibe | Lognormal | 0.003 | (-0.033, 0.039) | 0.886 | 0.968 | 0.032 | (-0.033, 0.039) |
| Decanoyl-L-carnitine | Lognormal | -0.002 | (-0.030, 0.027) | 0.899 | 0.973 | 0.032 | (-0.030, 0.026) |
| Glutamine | Lognormal | 0.000 | (-0.006, 0.005) | 0.906 | 0.975 | 0.031 | (-0.006, 0.005) |
| Trileptal | Lognormal | -0.001 | (-0.024, 0.022) | 0.906 | 0.975 | 0.032 | (-0.026, 0.020) |
| Diphenhydramine | Lognormal | -0.001 | (-0.023, 0.020) | 0.910 | 0.976 | 0.031 | (-0.021, 0.020) |
| Atorvastatin | Lognormal | -0.001 | (-0.019, 0.017) | 0.911 | 0.976 | 0.033 | (-0.020, 0.017) |
| 1-Phenylpyrrolidine | Lognormal | 0.001 | (-0.027, 0.030) | 0.919 | 0.982 | 0.030 | (-0.027, 0.030) |
| Isoleucine | Lognormal | -0.001 | (-0.011, 0.010) | 0.921 | 0.982 | 0.033 | (-0.012, 0.010) |
| Pantoprazole | Lognormal | 0.003 | (-0.053, 0.059) | 0.922 | 0.982 | 0.032 | (-0.053, 0.062) |
| D-Fructose | Lognormal | 0.000 | (-0.007, 0.007) | 0.926 | 0.984 | 0.029 | (-0.007, 0.007) |
| L-Carnitine | Lognormal | 0.000 | (-0.011, 0.010) | 0.941 | 0.987 | 0.032 | (-0.010, 0.010) |

**Supplementary Table S2.** Regression and Bayesian estimates comparing all ME/CFS vs. all controls.

| Metabolite | Regression Model | ME/CFS vs. Controls | | | | | |
|---|---|---|---|---|---|---|---|
| | | Estimated Coefficient | 95% CI | p-value | FDR | BayesFactor | 95% HDI |
| Moxonidine | Lognormal | 0.001 | (-0.025, 0.026) | 0.954 | 0.990 | 0.034 | (-0.025, 0.027) |
| Phenylacetylglutamine | Lognormal | 0.001 | (-0.025, 0.026) | 0.957 | 0.990 | 0.033 | (-0.025, 0.027) |
| 2,2-Bishydroxymethyl]-2,2',2''-nitrilotriethanol | Lognormal | -0.001 | (-0.028, 0.026) | 0.957 | 0.990 | 0.030 | (-0.028, 0.026) |
| 4-Fluoro-.alpha.-pyrrolidinobutiophenone | Lognormal | 0.001 | (-0.022, 0.023) | 0.959 | 0.990 | 0.037 | (-0.024, 0.024) |
| 2,2',2''-Nitrilotriethanol | Lognormal | -0.001 | (-0.025, 0.023) | 0.961 | 0.990 | 0.032 | (-0.025, 0.024) |
| 3,5-Dihydroxyphenylglycine | Lognormal | -0.001 | (-0.029, 0.028) | 0.962 | 0.990 | 0.029 | (-0.029, 0.028) |
| Methionine | Lognormal | 0.000 | (-0.009, 0.010) | 0.971 | 0.990 | 0.034 | (-0.010, 0.009) |
| Glycodeoxycholic acid | Lognormal | 0.001 | (-0.038, 0.039) | 0.974 | 0.990 | 0.028 | (-0.038, 0.039) |
| Levocetirizine; | Lognormal | 0.001 | (-0.070, 0.071) | 0.984 | 0.993 | 0.035 | (-0.070, 0.067) |
| Cyclo(Leu-Pro) | Lognormal | 0.000 | (-0.026, 0.025) | 0.989 | 0.994 | 0.033 | (-0.027, 0.025) |
| **Complex Lipids (CL)** | | | | | | | |
| PC (p-34:2)/PC (o-34:3) - ESI(+) | Lognormal | **-0.018** | **(-0.027, -0.009)** | **0.000** | **0.062** | **44.620** | **(-0.027, -0.009)** |
| PC (p-34:1)/PC (o-34:2) | Lognormal | **-0.021** | **(-0.032, -0.010)** | **0.000** | **0.062** | **178.678** | **(-0.032, -0.011)** |
| PE (p-36:2)/PE (o-36:3) - ESI(+) | Lognormal | **-0.028** | **(-0.043, -0.013)** | **0.000** | **0.074** | **20.935** | **(-0.042, -0.013)** |
| PC (36:2) | Lognormal | **-0.007** | **(-0.011, -0.003)** | **0.000** | **0.074** | **11.241** | **(-0.011, -0.003)** |
| PC (36:4) A - ESI(+) | Lognormal | **-0.018** | **(-0.028, -0.008)** | **0.000** | **0.074** | **8.134** | **(-0.028, -0.007)** |
| PC (p-36:1)/PC (o-36:2) | Lognormal | **-0.055** | **(-0.086, -0.024)** | **0.001** | **0.074** | **11.555** | **(-0.087, -0.025)** |
| PC (p-34:2)/PC (o-34:3) - ESI(-) | Lognormal | **-0.020** | **(-0.032, -0.009)** | **0.001** | **0.074** | **12.281** | **(-0.031, -0.009)** |
| PC (p-36:4)/PC (o-36:5) - ESI(-) | Lognormal | **-0.021** | **(-0.034, -0.009)** | **0.001** | **0.103** | **7.046** | **(-0.033, -0.009)** |
| PC (36:4) A - ESI(-) | Lognormal | **-0.019** | **(-0.031, -0.008)** | **0.001** | **0.103** | **4.032** | **(-0.031, -0.008)** |
| PC (p-34:1)/PC (o-34:2) A | Lognormal | **-0.027** | **(-0.044, -0.011)** | **0.002** | **0.125** | **5.655** | **(-0.044, -0.011)** |
| PE (p-34:2)/PE (o-34:3) | Lognormal | **-0.037** | **(-0.060, -0.014)** | **0.002** | **0.126** | **5.662** | **(-0.060, -0.015)** |
| PC (32:2) - ESI(-) | Lognormal | **-0.027** | **(-0.043, -0.010)** | **0.002** | **0.135** | **7.389** | **(-0.044, -0.010)** |
| LPC (18:2) - ESI(-) | Lognormal | **-0.019** | **(-0.032, -0.007)** | **0.003** | **0.139** | **4.102** | **(-0.031, -0.006)** |
| PC 34:4e | Lognormal | **-0.022** | **(-0.036, -0.008)** | **0.003** | **0.139** | **4.327** | **(-0.035, -0.008)** |
| PC (34:2) - ESI(+) | Lognormal | -0.005 | (-0.008, -0.002) | 0.003 | 0.139 | 2.039 | (-0.008, -0.001) |
| SM (d39:1) - ESI(-) | Lognormal | -0.017 | (-0.029, -0.006) | 0.004 | 0.139 | 2.833 | (-0.029, -0.006) |
| PE (p-36:2)/PE (o-36:3) - ESI(-) | Lognormal | -0.029 | (-0.048, -0.010) | 0.004 | 0.139 | 2.505 | (-0.048, -0.010) |
| PC (p-36:3)/PC (o-36:4) - ESI(-) | Lognormal | -0.016 | (-0.026, -0.005) | 0.004 | 0.139 | 2.209 | (-0.026, -0.006) |
| SM (d40:1) - ESI(-) | Lognormal | -0.012 | (-0.020, -0.004) | 0.004 | 0.143 | 2.357 | (-0.020, -0.004) |
| PC (34:2) - ESI(-) | Lognormal | -0.009 | (-0.016, -0.003) | 0.005 | 0.171 | 1.878 | (-0.016, -0.003) |
| PE (p-36:4)/PE (o-36:5) - ESI(-) | Lognormal | -0.023 | (-0.039, -0.007) | 0.005 | 0.171 | 1.112 | (-0.038, -0.005) |
| LPC (18:3) | Lognormal | -0.025 | (-0.043, -0.008) | 0.006 | 0.172 | 1.596 | (-0.044, -0.009) |
| SM (d32:1) - ESI(-) | Lognormal | -0.015 | (-0.025, -0.004) | 0.006 | 0.172 | 1.401 | (-0.025, -0.004) |
| LPC (18:2) - ESI(+) | Lognormal | -0.015 | (-0.025, -0.004) | 0.007 | 0.173 | 1.455 | (-0.024, -0.004) |
| Ceramide (d42:1) - ESI (+) | Lognormal | -0.009 | (-0.015, -0.003) | 0.007 | 0.173 | 1.584 | (-0.016, -0.003) |
| LPE (20:4) - ESI(-) | Lognormal | -0.024 | (-0.040, -0.007) | 0.007 | 0.173 | 1.313 | (-0.041, -0.007) |
| SM (d40:3) | Lognormal | -0.025 | (-0.043, -0.007) | 0.007 | 0.173 | 1.222 | (-0.045, -0.008) |
| Ceramide (d41:1) - ESI(-) | Lognormal | -0.014 | (-0.025, -0.004) | 0.007 | 0.173 | 1.295 | (-0.024, -0.003) |
| LPC (14:0) - ESI(+) | Lognormal | -0.021 | (-0.036, -0.006) | 0.008 | 0.177 | 1.300 | (-0.036, -0.006) |
| PC (34:4) - ESI(-) | Lognormal | -0.033 | (-0.056, -0.009) | 0.008 | 0.177 | 0.952 | (-0.054, -0.006) |
| PE (p-38:5)/PE (o-38:6) - ESI(-) | Lognormal | -0.021 | (-0.036, -0.006) | 0.008 | 0.177 | 1.045 | (-0.036, -0.005) |
| PC (p-38:3)/PC (o-38:4) - ESI(-) | Lognormal | -0.016 | (-0.028, -0.004) | 0.008 | 0.177 | 1.287 | (-0.028, -0.004) |
| LPE (18:2) - ESI(-) | Lognormal | -0.028 | (-0.049, -0.007) | 0.008 | 0.177 | 1.008 | (-0.049, -0.008) |
| Ceramide (d42:1) - ESI(-) | Lognormal | -0.012 | (-0.020, -0.003) | 0.009 | 0.177 | 1.534 | (-0.020, -0.003) |
| LPC (18:0) A - ESI(-) | Lognormal | -0.019 | (-0.033, -0.005) | 0.009 | 0.177 | 1.172 | (-0.032, -0.005) |
| LPC (16:0) - ESI(-) | Lognormal | -0.010 | (-0.018, -0.003) | 0.010 | 0.185 | 0.725 | (-0.018, -0.002) |

**Supplementary Table S2.** Regression and Bayesian estimates comparing all ME/CFS vs. all controls.

| Metabolite | Regression Model | ME/CFS vs. Controls | | | | | |
|---|---|---|---|---|---|---|---|
| | | Estimated Coefficient | 95% CI | p-value | FDR | BayesFactor | 95% HDI |
| LPE (18:2) - ESI(+) | Lognormal | -0.022 | (-0.039, -0.005) | 0.010 | 0.185 | 1.241 | (-0.039, -0.007) |
| LPC (20:3) - ESI(-) | Lognormal | -0.024 | (-0.043, -0.006) | 0.010 | 0.185 | 0.931 | (-0.042, -0.006) |
| PC (34:3) | Lognormal | -0.016 | (-0.029, -0.004) | 0.010 | 0.187 | 0.795 | (-0.029, -0.004) |
| CE (18:3) | Lognormal | -0.018 | (-0.032, -0.004) | 0.011 | 0.188 | 0.818 | (-0.032, -0.004) |
| PC (p-38:4)/PC (o-38:5) A | Lognormal | -0.012 | (-0.021, -0.003) | 0.011 | 0.188 | 0.669 | (-0.021, -0.002) |
| SM (d33:1) - ESI(-) | Lognormal | -0.015 | (-0.026, -0.003) | 0.012 | 0.188 | 1.010 | (-0.026, -0.004) |
| SM (d36:3) - ESI(-) | Lognormal | -0.020 | (-0.036, -0.005) | 0.012 | 0.188 | 0.898 | (-0.035, -0.005) |
| TG (56:6) | Lognormal | 0.010 | (0.002, 0.017) | 0.012 | 0.188 | 0.715 | (0.001, 0.017) |
| PC (p-36:2)/PC (o-36:3) | Lognormal | -0.016 | (-0.028, -0.004) | 0.012 | 0.188 | 1.014 | (-0.028, -0.004) |
| SM (d43:1) - ESI(-) | Lognormal | -0.034 | (-0.060, -0.008) | 0.012 | 0.188 | 0.948 | (-0.059, -0.008) |
| LPC (18:0) B - ESI(-) | Lognormal | -0.013 | (-0.023, -0.003) | 0.013 | 0.192 | 0.780 | (-0.023, -0.003) |
| Ceramide (d42:2) B - ESI (+) | Lognormal | -0.011 | (-0.020, -0.002) | 0.013 | 0.192 | 0.654 | (-0.020, -0.003) |
| TG (55:6) | Lognormal | 0.020 | (0.004, 0.036) | 0.014 | 0.206 | 0.592 | (0.004, 0.036) |
| SM (d40:2) B - ESI(-) | Lognormal | -0.012 | (-0.021, -0.002) | 0.015 | 0.208 | 0.846 | (-0.021, -0.003) |
| TG (53:0) | Lognormal | -0.018 | (-0.033, -0.004) | 0.015 | 0.208 | 0.624 | (-0.032, -0.003) |
| Ceramide (d39:1) | Lognormal | -0.021 | (-0.037, -0.004) | 0.016 | 0.208 | 0.560 | (-0.037, -0.003) |
| PC (33:2) - ESI(-) | Lognormal | -0.020 | (-0.036, -0.004) | 0.016 | 0.208 | 0.580 | (-0.037, -0.004) |
| CE (18:2) | Lognormal | -0.008 | (-0.015, -0.002) | 0.016 | 0.208 | 0.498 | (-0.015, -0.001) |
| Ceramide (d40:1) | Lognormal | -0.010 | (-0.018, -0.002) | 0.016 | 0.208 | 0.689 | (-0.018, -0.001) |
| PE (p-38:4)/PE (o-38:5) | Lognormal | -0.016 | (-0.029, -0.003) | 0.016 | 0.208 | 0.588 | (-0.029, -0.003) |
| SM (d34:2) - ESI(-) | Lognormal | -0.009 | (-0.016, -0.002) | 0.017 | 0.208 | 0.649 | (-0.015, -0.001) |
| PE (p-36:4)/PE (o-36:5) - ESI(+) | Lognormal | -0.017 | (-0.031, -0.003) | 0.017 | 0.208 | 0.522 | (-0.031, -0.004) |
| PC (34:3) B | Lognormal | -0.011 | (-0.020, -0.002) | 0.018 | 0.213 | 0.579 | (-0.021, -0.003) |
| PC (36:4) B - ESI(-) | Lognormal | -0.008 | (-0.015, -0.001) | 0.018 | 0.213 | 0.572 | (-0.015, -0.001) |
| PC (36:5) A | Lognormal | -0.040 | (-0.073, -0.007) | 0.018 | 0.213 | 0.685 | (-0.074, -0.009) |
| PC (38:4) A - ESI(-) | Lognormal | -0.010 | (-0.019, -0.002) | 0.019 | 0.213 | 0.478 | (-0.019, -0.002) |
| TG (50:0) | Lognormal | -0.024 | (-0.045, -0.004) | 0.019 | 0.214 | 0.547 | (-0.043, -0.003) |
| PE (p-36:1)/PE (o-36:2) - ESI(-) | Lognormal | -0.029 | (-0.053, -0.005) | 0.020 | 0.214 | 0.587 | (-0.051, -0.003) |
| PC (34:3) C | Lognormal | -0.016 | (-0.029, -0.003) | 0.020 | 0.214 | 0.586 | (-0.028, -0.003) |
| CE (20:2) | Lognormal | -0.023 | (-0.043, -0.004) | 0.020 | 0.214 | 0.490 | (-0.043, -0.003) |
| FA (16:1) (palmitoleic acid) | Lognormal | 0.016 | (0.003, 0.030) | 0.021 | 0.214 | 0.402 | (0.002, 0.030) |
| SM (d42:1) - ESI(-) | Lognormal | -0.010 | (-0.018, -0.002) | 0.021 | 0.214 | 0.498 | (-0.017, -0.001) |
| PC (p-36:3)/PC (o-36:4) - ESI(+) | Lognormal | -0.010 | (-0.018, -0.002) | 0.021 | 0.214 | 0.496 | (-0.018, -0.002) |
| PC (32:2) - ESI(+) | Lognormal | -0.017 | (-0.030, -0.003) | 0.021 | 0.214 | 0.446 | (-0.030, -0.003) |
| PE (p-38:5)/PE (o-38:6) - ESI(+) | Lognormal | -0.013 | (-0.024, -0.002) | 0.022 | 0.218 | 0.424 | (-0.024, -0.001) |
| PC (p-36:4)/PC (o-36:5) - ESI(+) | Lognormal | -0.009 | (-0.017, -0.001) | 0.024 | 0.234 | 0.438 | (-0.017, -0.001) |
| SM (d34:1) - ESI(-) | Lognormal | -0.007 | (-0.014, -0.001) | 0.024 | 0.239 | 0.407 | (-0.014, -0.001) |
| TG (58:8) | Lognormal | 0.020 | (0.003, 0.038) | 0.026 | 0.247 | 0.350 | (0.002, 0.038) |
| PE (p-36:1)/PE (o-36:2) - ESI(+) | Lognormal | -0.020 | (-0.037, -0.003) | 0.026 | 0.247 | 0.437 | (-0.037, -0.002) |
| PC (36:5)A | Lognormal | -0.020 | (-0.037, -0.002) | 0.027 | 0.256 | 0.349 | (-0.038, -0.002) |
| LPE (18:0) | Lognormal | -0.014 | (-0.026, -0.002) | 0.028 | 0.258 | 0.347 | (-0.026, -0.002) |
| TG (56:7) B | Lognormal | 0.018 | (0.002, 0.034) | 0.029 | 0.261 | 0.401 | (0.002, 0.033) |
| SM (d30:1) - ESI(-) | Lognormal | -0.029 | (-0.056, -0.003) | 0.029 | 0.261 | 0.377 | (-0.055, -0.004) |
| PE (p-40:4)/PE (o-40:5) A | Lognormal | -0.024 | (-0.045, -0.002) | 0.031 | 0.268 | 0.236 | (-0.046, -0.003) |
| PC (p-38:3)/PC (o-38:4) A - ESI(+) | Lognormal | -0.010 | (-0.020, -0.001) | 0.033 | 0.282 | 0.312 | (-0.020, -0.001) |
| TG (58:9) | Lognormal | 0.018 | (0.002, 0.035) | 0.033 | 0.282 | 0.310 | (0.001, 0.034) |
| PC (40:8) - ESI(-) | Lognormal | -0.016 | (-0.030, -0.001) | 0.035 | 0.287 | 0.255 | (-0.031, -0.001) |

**Supplementary Table S2.** Regression and Bayesian estimates comparing all ME/CFS vs. all controls.

| Metabolite | Regression Model | ME/CFS vs. Controls | | | | | |
|---|---|---|---|---|---|---|---|
| | | Estimated Coefficient | 95% CI | p-value | FDR | BayesFactor | 95% HDI |
| PC (o-34:0) | Lognormal | -0.014 | (-0.027, -0.001) | 0.035 | 0.288 | 0.313 | (-0.027, -0.001) |
| LPC (20:3) - ESI(+) | Lognormal | -0.013 | (-0.025, -0.001) | 0.036 | 0.288 | 0.266 | (-0.025, 0.000) |
| PC (38:6) A - ESI(+) | Lognormal | -0.008 | (-0.016, -0.001) | 0.036 | 0.288 | 0.283 | (-0.017, -0.001) |
| PC (38:3) - ESI(-) | Lognormal | -0.012 | (-0.023, -0.001) | 0.036 | 0.288 | 0.315 | (-0.022, 0.000) |
| TG (54:7) B | Lognormal | 0.016 | (0.001, 0.032) | 0.037 | 0.292 | 0.332 | (0.000, 0.031) |
| LPC (14:0) - ESI(-) | Lognormal | -0.027 | (-0.053, -0.002) | 0.038 | 0.296 | 0.304 | (-0.053, -0.002) |
| TG (62:4) | Lognormal | -0.028 | (-0.055, -0.002) | 0.039 | 0.298 | 0.303 | (-0.054, 0.000) |
| LPC (16:1) - ESI(-) | Lognormal | -0.017 | (-0.032, -0.001) | 0.039 | 0.298 | 0.312 | (-0.033, -0.001) |
| PC (p-38:4)/PC (o-38:5) B | Lognormal | -0.011 | (-0.021, -0.001) | 0.039 | 0.298 | 0.294 | (-0.021, -0.001) |
| PC (34:0) - ESI(-) | Lognormal | -0.009 | (-0.019, 0.000) | 0.041 | 0.304 | 0.291 | (-0.019, -0.001) |
| SM (d38:2) - ESI(-) | Lognormal | -0.011 | (-0.022, 0.000) | 0.042 | 0.309 | 0.283 | (-0.023, -0.001) |
| PE (36:2) | Lognormal | -0.020 | (-0.040, -0.001) | 0.042 | 0.309 | 0.232 | (-0.041, -0.001) |
| PI (38:4)/PI (18:0-20:4) | Lognormal | -0.008 | (-0.016, 0.000) | 0.044 | 0.319 | 0.285 | (-0.016, 0.000) |
| PC (34:4) - ESI(+) | Lognormal | -0.017 | (-0.033, 0.000) | 0.046 | 0.327 | 0.282 | (-0.033, 0.000) |
| PC (p-38:3)/PC (o-38:4) B - ESI(+) | Lognormal | -0.010 | (-0.020, 0.000) | 0.046 | 0.330 | 0.239 | (-0.020, 0.000) |
| AC (10:1) | Lognormal | -0.020 | (-0.039, 0.000) | 0.048 | 0.336 | 0.266 | (-0.039, 0.000) |
| TG (54:6) C | Lognormal | 0.010 | (0.000, 0.021) | 0.051 | 0.348 | 0.186 | (0.000, 0.021) |
| SM (d40:2) B - ESI(+) | Lognormal | -0.006 | (-0.012, 0.000) | 0.051 | 0.348 | 0.227 | (-0.012, 0.000) |
| PE (p-34:1)/PE (o-34:2) - ESI(+) | Lognormal | -0.014 | (-0.029, 0.000) | 0.051 | 0.348 | 0.161 | (-0.028, 0.001) |
| SM (d40:1) - ESI(+) | Lognormal | -0.006 | (-0.011, 0.000) | 0.053 | 0.351 | 0.231 | (-0.011, 0.000) |
| PC (36:3) B - ESI(-) | Lognormal | -0.008 | (-0.017, 0.000) | 0.055 | 0.359 | 0.193 | (-0.017, 0.000) |
| PC (36:3) A - ESI(+) | Lognormal | -0.007 | (-0.013, 0.000) | 0.057 | 0.365 | 0.211 | (-0.013, 0.001) |
| PC (p-32:1)/PC (o-32:2) | Lognormal | -0.009 | (-0.019, 0.000) | 0.058 | 0.367 | 0.191 | (-0.019, 0.000) |
| GlcCer (d42:2) - ESI(+) | Lognormal | 0.012 | (0.000, 0.024) | 0.059 | 0.367 | 0.190 | (-0.001, 0.024) |
| LPC (18:1) - ESI(-) | Lognormal | -0.011 | (-0.022, 0.000) | 0.059 | 0.367 | 0.211 | (-0.021, 0.001) |
| TG (56:8) B | Lognormal | 0.018 | (-0.001, 0.036) | 0.060 | 0.370 | 0.198 | (0.000, 0.037) |
| PC (37:5) | Lognormal | 0.016 | (-0.001, 0.032) | 0.061 | 0.374 | 0.213 | (-0.001, 0.031) |
| PC (p-38:4)/PC (o-38:5) B | Lognormal | -0.017 | (-0.034, 0.001) | 0.062 | 0.376 | 0.185 | (-0.034, 0.001) |
| SM (d38:1) | Lognormal | -0.006 | (-0.011, 0.000) | 0.063 | 0.376 | 0.147 | (-0.012, 0.000) |
| SM (d36:1) - ESI(-) | Lognormal | -0.008 | (-0.016, 0.001) | 0.067 | 0.396 | 0.211 | (-0.017, 0.000) |
| Ceramide (d43:1) | Lognormal | -0.033 | (-0.068, 0.002) | 0.069 | 0.399 | 0.182 | (-0.069, 0.003) |
| DG (38:6) | Lognormal | 0.015 | (-0.001, 0.031) | 0.070 | 0.404 | 0.169 | (-0.001, 0.030) |
| PE (p-40:4)/PE (o-40:5) B | Lognormal | -0.013 | (-0.026, 0.001) | 0.071 | 0.404 | 0.126 | (-0.026, 0.001) |
| TG (60:11) | Lognormal | 0.025 | (-0.002, 0.053) | 0.071 | 0.406 | 0.159 | (-0.004, 0.052) |
| TG (48:0) | Lognormal | -0.018 | (-0.038, 0.002) | 0.073 | 0.408 | 0.149 | (-0.038, 0.002) |
| PE (p-34:1)/PE (o-34:2) - ESI(-) | Lognormal | -0.014 | (-0.030, 0.001) | 0.073 | 0.408 | 0.163 | (-0.031, 0.001) |
| LPC (20:2) - ESI(+) | Lognormal | -0.028 | (-0.058, 0.002) | 0.073 | 0.408 | 0.159 | (-0.057, 0.003) |
| GlcCer (d38:1) | Lognormal | -0.012 | (-0.024, 0.001) | 0.073 | 0.408 | 0.170 | (-0.024, 0.001) |
| FA (18:1) (oleic acid) | Lognormal | 0.007 | (-0.001, 0.015) | 0.078 | 0.416 | 0.155 | (-0.001, 0.016) |
| PC (36:3) A - ESI(-) | Lognormal | -0.009 | (-0.018, 0.001) | 0.080 | 0.422 | 0.165 | (-0.018, 0.001) |
| LPC (p-16:0)/LPC (o-16:1) | Lognormal | -0.011 | (-0.023, 0.001) | 0.080 | 0.422 | 0.159 | (-0.023, 0.002) |
| LPC (20:2) - ESI(-) | Lognormal | -0.012 | (-0.025, 0.001) | 0.081 | 0.422 | 0.160 | (-0.025, 0.000) |
| PE (p-38:3)/PE (o-38:4) | Lognormal | -0.022 | (-0.048, 0.003) | 0.082 | 0.422 | 0.156 | (-0.048, 0.001) |
| Ceramide (d33:1) | Lognormal | -0.011 | (-0.023, 0.001) | 0.084 | 0.430 | 0.146 | (-0.023, 0.002) |
| PC (40:5) B - ESI(-) | Lognormal | -0.020 | (-0.042, 0.003) | 0.085 | 0.430 | 0.145 | (-0.042, 0.004) |
| PC (38:6) B - ESI(+) | Lognormal | 0.005 | (-0.001, 0.011) | 0.086 | 0.430 | 0.121 | (-0.001, 0.011) |
| PC (33:2) - ESI(+) | Lognormal | -0.008 | (-0.017, 0.001) | 0.087 | 0.430 | 0.121 | (-0.017, 0.001) |

**Supplementary Table S2.** Regression and Bayesian estimates comparing all ME/CFS vs. all controls.

| Metabolite | Regression Model | ME/CFS vs. Controls | | | | | |
|---|---|---|---|---|---|---|---|
| | | Estimated Coefficient | 95% CI | p-value | FDR | BayesFactor | 95% HDI |
| SM (d39:1) - ESI(+) | Lognormal | -0.008 | (-0.018, 0.001) | 0.089 | 0.430 | 0.121 | (-0.018, 0.002) |
| Ceramide (d41:1) - ESI (+) | Lognormal | -0.008 | (-0.018, 0.001) | 0.089 | 0.430 | 0.103 | (-0.018, 0.001) |
| FA (24:0) (lignoceric acid) | Lognormal | -0.010 | (-0.021, 0.001) | 0.091 | 0.434 | 0.132 | (-0.021, 0.001) |
| PC (40:6)B | Lognormal | 0.008 | (-0.001, 0.017) | 0.092 | 0.438 | 0.128 | (-0.001, 0.017) |
| AC (18:0) | Lognormal | -0.010 | (-0.022, 0.002) | 0.097 | 0.457 | 0.131 | (-0.023, 0.001) |
| LPE (16:0) | Lognormal | -0.018 | (-0.039, 0.003) | 0.098 | 0.460 | 0.130 | (-0.038, 0.004) |
| AC (14:2) | Lognormal | -0.019 | (-0.042, 0.004) | 0.099 | 0.461 | 0.126 | (-0.043, 0.004) |
| TG (46:0) | Lognormal | -0.028 | (-0.061, 0.005) | 0.100 | 0.461 | 0.131 | (-0.060, 0.005) |
| PC (40:5) A - ESI(+) | Lognormal | 0.007 | (-0.001, 0.015) | 0.101 | 0.461 | 0.142 | (0.000, 0.015) |
| Ceramide (d38:1) - ESI(+) | Lognormal | -0.008 | (-0.017, 0.001) | 0.102 | 0.463 | 0.123 | (-0.017, 0.002) |
| PE (36:3) | Lognormal | -0.024 | (-0.052, 0.005) | 0.103 | 0.465 | 0.113 | (-0.052, 0.004) |
| PE (p-38:6)/PE (o-38:7) | Lognormal | -0.015 | (-0.034, 0.003) | 0.104 | 0.466 | 0.131 | (-0.033, 0.004) |
| Ceramide (d38:1) - ESI(-) | Lognormal | -0.010 | (-0.023, 0.002) | 0.105 | 0.467 | 0.141 | (-0.023, 0.002) |
| SM (d43:2) - ESI(-) | Lognormal | -0.018 | (-0.040, 0.004) | 0.106 | 0.468 | 0.120 | (-0.039, 0.004) |
| Lactosylceramide (d18:1/24:1(15Z)) | Lognormal | 0.012 | (-0.003, 0.027) | 0.111 | 0.481 | 0.111 | (-0.002, 0.028) |
| PC (37:2) - ESI(+) | Lognormal | -0.007 | (-0.016, 0.002) | 0.115 | 0.486 | 0.120 | (-0.016, 0.001) |
| SM (d30:1) - ESI(+) | Lognormal | -0.016 | (-0.037, 0.004) | 0.115 | 0.486 | 0.119 | (-0.037, 0.006) |
| PC (36:5) D | Lognormal | 0.014 | (-0.003, 0.032) | 0.116 | 0.487 | 0.106 | (-0.003, 0.032) |
| AC (18:2) | Lognormal | -0.011 | (-0.025, 0.003) | 0.117 | 0.489 | 0.112 | (-0.026, 0.002) |
| PC (p-40:3)/PC (o-40:4) | Lognormal | -0.014 | (-0.031, 0.003) | 0.118 | 0.490 | 0.107 | (-0.030, 0.005) |
| SM (d42:1) - ESI(+) | Lognormal | -0.005 | (-0.012, 0.001) | 0.119 | 0.490 | 0.108 | (-0.012, 0.001) |
| PC (40:7) A - ESI(+) | Lognormal | -0.008 | (-0.018, 0.002) | 0.121 | 0.496 | 0.098 | (-0.018, 0.002) |
| PC (37:6) | Lognormal | 0.014 | (-0.004, 0.031) | 0.123 | 0.504 | 0.115 | (-0.004, 0.030) |
| SM (d32:1) - ESI(+) | Lognormal | -0.008 | (-0.018, 0.002) | 0.127 | 0.516 | 0.109 | (-0.018, 0.002) |
| TG (48:4) A | Lognormal | -0.021 | (-0.047, 0.006) | 0.128 | 0.519 | 0.104 | (-0.047, 0.006) |
| Ceramide (d32:1) | Lognormal | -0.011 | (-0.026, 0.003) | 0.129 | 0.521 | 0.101 | (-0.025, 0.005) |
| PC (40:4) - ESI(-) | Lognormal | -0.014 | (-0.032, 0.004) | 0.131 | 0.524 | 0.116 | (-0.032, 0.004) |
| FA (20:5) (eicosapentaenoic acid) | Lognormal | 0.019 | (-0.005, 0.042) | 0.131 | 0.526 | 0.105 | (-0.004, 0.044) |
| AC (12:0) | Lognormal | -0.015 | (-0.035, 0.005) | 0.133 | 0.528 | 0.102 | (-0.036, 0.004) |
| TG (56:5) B | Lognormal | 0.009 | (-0.003, 0.021) | 0.133 | 0.528 | 0.106 | (-0.003, 0.021) |
| TG (56:8) A | Lognormal | 0.010 | (-0.003, 0.023) | 0.135 | 0.531 | 0.096 | (-0.003, 0.023) |
| LPE (20:4) - ESI(+) | Lognormal | -0.012 | (-0.028, 0.004) | 0.136 | 0.531 | 0.110 | (-0.029, 0.002) |
| CE (20:3) | Lognormal | -0.008 | (-0.020, 0.003) | 0.140 | 0.537 | 0.101 | (-0.020, 0.003) |
| PC (30:0) | Lognormal | -0.010 | (-0.023, 0.003) | 0.140 | 0.537 | 0.111 | (-0.024, 0.003) |
| PC (p-40:1)/PC (o-40:2) | Lognormal | 0.013 | (-0.005, 0.031) | 0.144 | 0.547 | 0.096 | (-0.004, 0.032) |
| PE (p-40:4)/PE (o-40:5) | Lognormal | -0.012 | (-0.028, 0.004) | 0.144 | 0.547 | 0.076 | (-0.027, 0.005) |
| AC (12:1) | Lognormal | -0.014 | (-0.033, 0.005) | 0.145 | 0.547 | 0.105 | (-0.033, 0.004) |
| PC (35:1) - ESI(+) | Lognormal | 0.005 | (-0.002, 0.012) | 0.147 | 0.551 | 0.093 | (-0.002, 0.012) |
| SM (d36:0) - ESI(+) | Lognormal | 0.010 | (-0.004, 0.025) | 0.148 | 0.553 | 0.098 | (-0.004, 0.025) |
| LPC (20:4) | Lognormal | -0.008 | (-0.018, 0.003) | 0.150 | 0.554 | 0.086 | (-0.018, 0.002) |
| PC (38:2) | Lognormal | -0.009 | (-0.021, 0.003) | 0.150 | 0.554 | 0.105 | (-0.020, 0.004) |
| PE (38:2) | Lognormal | -0.013 | (-0.030, 0.005) | 0.155 | 0.564 | 0.086 | (-0.030, 0.005) |
| LPC (o-16:0) | Lognormal | -0.009 | (-0.022, 0.003) | 0.155 | 0.564 | 0.079 | (-0.021, 0.004) |
| SM (d34:0) - ESI(-) | Lognormal | -0.010 | (-0.023, 0.004) | 0.156 | 0.564 | 0.102 | (-0.024, 0.003) |
| TG (60:12) | Lognormal | 0.026 | (-0.010, 0.061) | 0.159 | 0.573 | 0.086 | (-0.010, 0.062) |
| FA (14:1) (physeteric acid) | Lognormal | 0.013 | (-0.005, 0.031) | 0.161 | 0.577 | 0.079 | (-0.005, 0.030) |
| TG (54:5) B | Lognormal | 0.007 | (-0.003, 0.016) | 0.163 | 0.580 | 0.083 | (-0.002, 0.016) |

**Supplementary Table S2.** Regression and Bayesian estimates comparing all ME/CFS vs. all controls.

| Metabolite | Regression Model | ME/CFS vs. Controls | | | | | |
|---|---|---|---|---|---|---|---|
| | | Estimated Coefficient | 95% CI | p-value | FDR | BayesFactor | 95% HDI |
| Cholesterol | Lognormal | -0.004 | (-0.010, 0.002) | 0.163 | 0.580 | 0.083 | (-0.010, 0.002) |
| PC (40:7) B - ESI(+) | Lognormal | 0.006 | (-0.003, 0.015) | 0.165 | 0.581 | 0.076 | (-0.003, 0.015) |
| PE (34:2) - ESI(-) | Lognormal | -0.018 | (-0.045, 0.008) | 0.168 | 0.582 | 0.088 | (-0.046, 0.008) |
| PC (35:3) | Lognormal | -0.005 | (-0.013, 0.002) | 0.168 | 0.582 | 0.079 | (-0.013, 0.002) |
| PC (40:5) B - ESI(+) | Lognormal | -0.009 | (-0.022, 0.004) | 0.170 | 0.582 | 0.085 | (-0.022, 0.004) |
| LPC (22:4) | Lognormal | -0.017 | (-0.040, 0.007) | 0.170 | 0.582 | 0.074 | (-0.038, 0.008) |
| LPC (16:0) - ESI(+) | Lognormal | -0.003 | (-0.008, 0.001) | 0.171 | 0.582 | 0.078 | (-0.008, 0.002) |
| LPE (22:6) | Lognormal | -0.012 | (-0.030, 0.005) | 0.172 | 0.582 | 0.071 | (-0.031, 0.005) |
| PC (42:10) | Lognormal | 0.014 | (-0.006, 0.034) | 0.172 | 0.582 | 0.078 | (-0.007, 0.034) |
| PC (p-32:0)/PC (o-32:1) - ESI(-) | Lognormal | -0.011 | (-0.026, 0.005) | 0.173 | 0.585 | 0.084 | (-0.025, 0.004) |
| AC (10:0) | Lognormal | -0.019 | (-0.047, 0.008) | 0.174 | 0.586 | 0.083 | (-0.046, 0.009) |
| TG (56:5) A | Lognormal | 0.009 | (-0.004, 0.021) | 0.179 | 0.591 | 0.086 | (-0.004, 0.022) |
| TG (46:2) | Lognormal | -0.021 | (-0.051, 0.009) | 0.179 | 0.591 | 0.083 | (-0.049, 0.011) |
| Ceramide (d42:2) B - ESI(-) | Lognormal | -0.012 | (-0.029, 0.005) | 0.179 | 0.591 | 0.088 | (-0.029, 0.005) |
| SM (d44:2) | Lognormal | 0.010 | (-0.004, 0.024) | 0.180 | 0.591 | 0.072 | (-0.005, 0.023) |
| DG (38:5) | Lognormal | 0.008 | (-0.004, 0.019) | 0.180 | 0.591 | 0.075 | (-0.003, 0.019) |
| FA (22:0) (behenic acid) | Lognormal | -0.008 | (-0.021, 0.004) | 0.182 | 0.593 | 0.073 | (-0.021, 0.004) |
| TG (58:6) | Lognormal | 0.007 | (-0.003, 0.018) | 0.182 | 0.593 | 0.087 | (-0.003, 0.018) |
| SM (d32:2) - ESI(-) | Lognormal | -0.013 | (-0.033, 0.006) | 0.184 | 0.596 | 0.086 | (-0.033, 0.006) |
| TG (46:4) A | Lognormal | -0.024 | (-0.060, 0.011) | 0.185 | 0.596 | 0.085 | (-0.060, 0.012) |
| TG (56:5) C | Lognormal | 0.007 | (-0.003, 0.018) | 0.186 | 0.599 | 0.085 | (-0.003, 0.018) |
| PE (34:1) | Lognormal | -0.019 | (-0.048, 0.009) | 0.189 | 0.599 | 0.076 | (-0.048, 0.010) |
| Ceramide (d34:1) - ESI(-) | Lognormal | -0.005 | (-0.013, 0.003) | 0.189 | 0.599 | 0.076 | (-0.013, 0.002) |
| LPC (18:0) - ESI(+) | Lognormal | -0.006 | (-0.014, 0.003) | 0.189 | 0.599 | 0.067 | (-0.014, 0.003) |
| TG (44:0) | Lognormal | -0.022 | (-0.055, 0.011) | 0.190 | 0.599 | 0.074 | (-0.055, 0.010) |
| PC (34:3) A | Lognormal | -0.006 | (-0.015, 0.003) | 0.191 | 0.599 | 0.074 | (-0.016, 0.003) |
| SM (d42:3) - ESI(-) | Lognormal | -0.005 | (-0.014, 0.003) | 0.191 | 0.599 | 0.075 | (-0.013, 0.003) |
| CE (20:5) | Lognormal | 0.015 | (-0.007, 0.036) | 0.192 | 0.599 | 0.078 | (-0.007, 0.036) |
| TG (42:0) | Lognormal | -0.023 | (-0.058, 0.012) | 0.193 | 0.599 | 0.075 | (-0.059, 0.012) |
| PC (p-38:5)/PC (o-38:6) B | Lognormal | -0.006 | (-0.014, 0.003) | 0.193 | 0.599 | 0.076 | (-0.014, 0.003) |
| TG (49:2) | Lognormal | 0.013 | (-0.007, 0.032) | 0.196 | 0.606 | 0.085 | (-0.006, 0.032) |
| PC (36:3) B - ESI(+) | Lognormal | -0.005 | (-0.013, 0.003) | 0.198 | 0.609 | 0.080 | (-0.013, 0.003) |
| PC (38:5) B - ESI(+) | Lognormal | 0.007 | (-0.004, 0.018) | 0.200 | 0.610 | 0.071 | (-0.003, 0.018) |
| PC (p-38:4)/PC (o-38:5) A | Lognormal | -0.004 | (-0.011, 0.002) | 0.201 | 0.610 | 0.072 | (-0.011, 0.002) |
| CE (14:0) | Lognormal | -0.022 | (-0.056, 0.012) | 0.203 | 0.616 | 0.080 | (-0.059, 0.010) |
| PC (39:6) | Lognormal | 0.010 | (-0.005, 0.025) | 0.206 | 0.622 | 0.063 | (-0.006, 0.025) |
| PC (37:2) - ESI(-) | Lognormal | -0.013 | (-0.034, 0.007) | 0.210 | 0.628 | 0.063 | (-0.033, 0.007) |
| PC (32:0) - ESI(-) | Lognormal | -0.005 | (-0.014, 0.003) | 0.214 | 0.631 | 0.072 | (-0.014, 0.003) |
| TG (54:1) | Lognormal | -0.013 | (-0.034, 0.008) | 0.218 | 0.636 | 0.069 | (-0.034, 0.009) |
| SM (d42:2) | Lognormal | 0.004 | (-0.003, 0.012) | 0.218 | 0.636 | 0.065 | (-0.003, 0.012) |
| SM (d36:2) - ESI(-) | Lognormal | -0.007 | (-0.019, 0.004) | 0.219 | 0.636 | 0.066 | (-0.020, 0.004) |
| PE (38:4) | Lognormal | -0.007 | (-0.017, 0.004) | 0.222 | 0.642 | 0.055 | (-0.018, 0.004) |
| TG (60:6) | Lognormal | 0.013 | (-0.008, 0.033) | 0.226 | 0.648 | 0.075 | (-0.008, 0.034) |
| PC (p-32:0)/PC (o-32:1) - ESI(+) | Lognormal | -0.005 | (-0.012, 0.003) | 0.234 | 0.667 | 0.059 | (-0.012, 0.003) |
| GlcCer (d14:1(4E)/20:0(2OH)) | Lognormal | -0.011 | (-0.030, 0.007) | 0.238 | 0.671 | 0.065 | (-0.030, 0.007) |
| GlcCer (d41:1) | Lognormal | -0.007 | (-0.020, 0.005) | 0.247 | 0.685 | 0.067 | (-0.019, 0.006) |
| PC (38:3) - ESI(+) | Lognormal | -0.005 | (-0.013, 0.003) | 0.247 | 0.685 | 0.062 | (-0.013, 0.004) |

**Supplementary Table S2.** Regression and Bayesian estimates comparing all ME/CFS vs. all controls.

| Metabolite | Regression Model | ME/CFS vs. Controls | | | | | |
|---|---|---|---|---|---|---|---|
| | | Estimated Coefficient | 95% CI | p-value | FDR | BayesFactor | 95% HDI |
| TG (40:1) | Lognormal | -0.023 | (-0.061, 0.016) | 0.247 | 0.685 | 0.061 | (-0.061, 0.018) |
| PC (35:2) | Lognormal | -0.007 | (-0.018, 0.005) | 0.248 | 0.685 | 0.064 | (-0.018, 0.005) |
| Ceramide (d44:1) | Lognormal | -0.014 | (-0.039, 0.010) | 0.251 | 0.688 | 0.055 | (-0.038, 0.010) |
| PC (38:5) A | Lognormal | 0.003 | (-0.002, 0.009) | 0.254 | 0.689 | 0.063 | (-0.002, 0.009) |
| SM (d32:0) - ESI(-) | Lognormal | -0.031 | (-0.085, 0.022) | 0.254 | 0.689 | 0.063 | (-0.085, 0.023) |
| TG (56:9) | Lognormal | 0.011 | (-0.008, 0.029) | 0.257 | 0.692 | 0.061 | (-0.008, 0.029) |
| TG (48:3) | Lognormal | -0.012 | (-0.034, 0.009) | 0.258 | 0.692 | 0.062 | (-0.034, 0.007) |
| TG (54:6) B | Lognormal | -0.019 | (-0.053, 0.014) | 0.262 | 0.697 | 0.068 | (-0.055, 0.012) |
| TG (48:5) | Lognormal | -0.018 | (-0.048, 0.013) | 0.264 | 0.697 | 0.063 | (-0.046, 0.014) |
| SM (d41:2) B - ESI(+) | Lognormal | -0.005 | (-0.013, 0.003) | 0.264 | 0.697 | 0.052 | (-0.013, 0.003) |
| TG (42:3) | Lognormal | -0.021 | (-0.058, 0.016) | 0.276 | 0.726 | 0.061 | (-0.055, 0.019) |
| PC 40:5e | Lognormal | -0.005 | (-0.014, 0.004) | 0.279 | 0.730 | 0.056 | (-0.014, 0.004) |
| TG (58:2) | Lognormal | -0.027 | (-0.075, 0.022) | 0.287 | 0.737 | 0.065 | (-0.077, 0.023) |
| PC (p-44:4)/PC (o-44:5) - ESI(-) | Lognormal | -0.007 | (-0.021, 0.006) | 0.288 | 0.737 | 0.053 | (-0.021, 0.006) |
| LPC (16:1) - ESI(+) | Lognormal | -0.006 | (-0.016, 0.005) | 0.288 | 0.737 | 0.060 | (-0.016, 0.005) |
| PC (p-42:3)/PC (o-42:4) | Lognormal | -0.006 | (-0.019, 0.006) | 0.294 | 0.739 | 0.053 | (-0.019, 0.005) |
| SM (d41:3) | Lognormal | -0.005 | (-0.013, 0.004) | 0.295 | 0.739 | 0.059 | (-0.014, 0.003) |
| PC (38:7) | Lognormal | 0.008 | (-0.007, 0.023) | 0.296 | 0.740 | 0.052 | (-0.006, 0.023) |
| TG (48:4) B | Lognormal | -0.019 | (-0.054, 0.016) | 0.298 | 0.740 | 0.057 | (-0.054, 0.015) |
| DG (36:4) B | Lognormal | 0.019 | (-0.016, 0.053) | 0.299 | 0.740 | 0.054 | (-0.015, 0.054) |
| Ceramide (d42:0) | Lognormal | -0.008 | (-0.022, 0.007) | 0.301 | 0.740 | 0.048 | (-0.022, 0.006) |
| PE (p-40:5)/PE (o-40:6) | Lognormal | -0.005 | (-0.014, 0.004) | 0.303 | 0.740 | 0.044 | (-0.014, 0.004) |
| PG (34:0)/PG (17:0/17:0) | Lognormal | -0.008 | (-0.024, 0.008) | 0.303 | 0.740 | 0.055 | (-0.025, 0.007) |
| FA (18:2) (linoleic acid) | Lognormal | 0.004 | (-0.004, 0.012) | 0.306 | 0.743 | 0.055 | (-0.003, 0.012) |
| FA (20:1) (eicosenoic acid) | Lognormal | 0.005 | (-0.005, 0.015) | 0.309 | 0.743 | 0.053 | (-0.005, 0.015) |
| TG (42:1) | Lognormal | -0.033 | (-0.098, 0.031) | 0.314 | 0.743 | 0.052 | (-0.098, 0.032) |
| DG (36:2) | Lognormal | 0.005 | (-0.005, 0.016) | 0.314 | 0.743 | 0.051 | (-0.005, 0.016) |
| TG (46:1) | Lognormal | -0.016 | (-0.048, 0.015) | 0.319 | 0.743 | 0.054 | (-0.048, 0.016) |
| GlcCer (d40:1) - ESI(-) | Lognormal | -0.006 | (-0.016, 0.005) | 0.320 | 0.743 | 0.055 | (-0.017, 0.005) |
| TG (50:4) | Lognormal | -0.007 | (-0.022, 0.007) | 0.322 | 0.743 | 0.049 | (-0.021, 0.007) |
| PC (28:0) | Lognormal | -0.019 | (-0.057, 0.019) | 0.323 | 0.743 | 0.057 | (-0.059, 0.016) |
| GlcCer (d42:2) - ESI(-) | Lognormal | 0.006 | (-0.006, 0.018) | 0.323 | 0.743 | 0.050 | (-0.006, 0.018) |
| TG (44:1) | Lognormal | -0.021 | (-0.062, 0.021) | 0.328 | 0.747 | 0.050 | (-0.064, 0.020) |
| PC (36:1) - ESI(-) | Lognormal | -0.004 | (-0.012, 0.004) | 0.332 | 0.747 | 0.047 | (-0.012, 0.004) |
| FA (18:3) (linolenic acid) | Lognormal | 0.006 | (-0.006, 0.018) | 0.333 | 0.747 | 0.050 | (-0.006, 0.018) |
| AC (8:0) | Lognormal | -0.014 | (-0.042, 0.014) | 0.333 | 0.747 | 0.054 | (-0.042, 0.016) |
| TG (54:7) A | Lognormal | -0.010 | (-0.031, 0.011) | 0.338 | 0.751 | 0.050 | (-0.033, 0.011) |
| Ceramide (d42:2) A - ESI(-) | Lognormal | -0.005 | (-0.014, 0.005) | 0.338 | 0.751 | 0.047 | (-0.014, 0.005) |
| PC (p-36:1)/PC (o-36:2) B | Lognormal | -0.007 | (-0.022, 0.008) | 0.346 | 0.756 | 0.048 | (-0.023, 0.007) |
| PE (36:4) - ESI(-) | Lognormal | -0.012 | (-0.038, 0.013) | 0.350 | 0.760 | 0.048 | (-0.040, 0.012) |
| TG (40:0) | Lognormal | -0.014 | (-0.044, 0.016) | 0.352 | 0.760 | 0.043 | (-0.044, 0.017) |
| TG (50:1) | Lognormal | -0.008 | (-0.025, 0.009) | 0.352 | 0.760 | 0.052 | (-0.025, 0.008) |
| TG (46:3) A | Lognormal | -0.019 | (-0.059, 0.021) | 0.353 | 0.760 | 0.053 | (-0.060, 0.022) |
| PC (p-42:5)/PC (o-42:6) A | Lognormal | -0.006 | (-0.018, 0.006) | 0.354 | 0.760 | 0.050 | (-0.017, 0.007) |
| SM (d41:1) - ESI(+) | Lognormal | -0.003 | (-0.011, 0.004) | 0.356 | 0.761 | 0.045 | (-0.011, 0.004) |
| PE (34:2) - ESI(+) | Lognormal | -0.031 | (-0.098, 0.035) | 0.361 | 0.764 | 0.052 | (-0.101, 0.034) |
| PC (38:4) B - ESI(+) | Lognormal | -0.004 | (-0.014, 0.005) | 0.362 | 0.764 | 0.048 | (-0.014, 0.005) |

**Supplementary Table S2.** Regression and Bayesian estimates comparing all ME/CFS vs. all controls.

| Metabolite | Regression Model | ME/CFS vs. Controls | | | | | |
|---|---|---|---|---|---|---|---|
| | | Estimated Coefficient | 95% CI | p-value | FDR | BayesFactor | 95% HDI |
| PC (o-32:0) - ESI(-) | Lognormal | -0.006 | (-0.018, 0.007) | 0.364 | 0.764 | 0.047 | (-0.018, 0.006) |
| TG (46:4) B | Lognormal | -0.020 | (-0.062, 0.023) | 0.365 | 0.764 | 0.047 | (-0.060, 0.023) |
| TG (64:3) | Lognormal | -0.010 | (-0.032, 0.012) | 0.366 | 0.764 | 0.053 | (-0.033, 0.011) |
| TG (42:2) | Lognormal | -0.037 | (-0.117, 0.043) | 0.370 | 0.767 | 0.048 | (-0.117, 0.045) |
| LPC (20:5) | Lognormal | 0.012 | (-0.014, 0.038) | 0.372 | 0.767 | 0.047 | (-0.013, 0.038) |
| PE (36:1) - ESI(+) | Lognormal | -0.010 | (-0.030, 0.011) | 0.372 | 0.767 | 0.047 | (-0.032, 0.011) |
| SM (d40:2) A - ESI(+) | Lognormal | -0.005 | (-0.016, 0.006) | 0.375 | 0.768 | 0.051 | (-0.017, 0.005) |
| TG (51:2) | Lognormal | 0.006 | (-0.008, 0.020) | 0.378 | 0.768 | 0.049 | (-0.007, 0.020) |
| PC (35:2) A | Lognormal | 0.004 | (-0.005, 0.013) | 0.379 | 0.768 | 0.047 | (-0.005, 0.012) |
| TG (53:3) | Lognormal | 0.005 | (-0.006, 0.016) | 0.381 | 0.768 | 0.049 | (-0.006, 0.016) |
| SM (d40:2) A - ESI(-) | Lognormal | -0.008 | (-0.025, 0.009) | 0.382 | 0.768 | 0.054 | (-0.024, 0.009) |
| TG (50:5) | Lognormal | -0.009 | (-0.029, 0.011) | 0.386 | 0.771 | 0.044 | (-0.030, 0.011) |
| CE (22:6) | Lognormal | 0.005 | (-0.006, 0.016) | 0.387 | 0.772 | 0.043 | (-0.006, 0.017) |
| GlcCer (d42:1) - ESI(-) | Lognormal | -0.005 | (-0.015, 0.006) | 0.388 | 0.772 | 0.047 | (-0.016, 0.005) |
| PC (o-32:0) - ESI(+) | Lognormal | -0.003 | (-0.011, 0.004) | 0.391 | 0.777 | 0.046 | (-0.011, 0.004) |
| PC (37:4) - ESI(-) | Lognormal | -0.006 | (-0.019, 0.008) | 0.396 | 0.781 | 0.043 | (-0.019, 0.007) |
| PC (36:5) C | Lognormal | 0.008 | (-0.011, 0.027) | 0.424 | 0.818 | 0.043 | (-0.010, 0.027) |
| TG (52:2) | Lognormal | 0.003 | (-0.004, 0.010) | 0.425 | 0.818 | 0.044 | (-0.004, 0.010) |
| AC (14:1) | Lognormal | -0.009 | (-0.030, 0.012) | 0.426 | 0.819 | 0.041 | (-0.029, 0.013) |
| SM (d39:2) | Lognormal | -0.003 | (-0.012, 0.005) | 0.431 | 0.822 | 0.040 | (-0.011, 0.005) |
| PC (32:1) - ESI(-) | Lognormal | -0.007 | (-0.023, 0.010) | 0.435 | 0.828 | 0.049 | (-0.024, 0.009) |
| FA (20:3) (homo-gamma-linolenic acid) | Lognormal | 0.004 | (-0.005, 0.012) | 0.437 | 0.829 | 0.039 | (-0.005, 0.013) |
| PC (40:4) - ESI(+) | Lognormal | -0.005 | (-0.016, 0.007) | 0.439 | 0.829 | 0.041 | (-0.016, 0.008) |
| TG (53:2) | Lognormal | 0.005 | (-0.008, 0.018) | 0.455 | 0.848 | 0.045 | (-0.009, 0.018) |
| PC (35:4) - ESI(-) | Lognormal | -0.007 | (-0.024, 0.011) | 0.456 | 0.848 | 0.039 | (-0.024, 0.010) |
| Ceramide (d34:0) | Lognormal | -0.005 | (-0.017, 0.008) | 0.458 | 0.848 | 0.036 | (-0.017, 0.008) |
| TG (58:10) | Lognormal | 0.007 | (-0.011, 0.025) | 0.458 | 0.848 | 0.041 | (-0.011, 0.025) |
| CE (22:2) | Lognormal | 0.013 | (-0.022, 0.048) | 0.459 | 0.848 | 0.041 | (-0.022, 0.047) |
| PC (34:1) | Lognormal | 0.001 | (-0.002, 0.005) | 0.465 | 0.849 | 0.041 | (-0.002, 0.005) |
| TG (58:4) | Lognormal | 0.008 | (-0.014, 0.030) | 0.478 | 0.856 | 0.043 | (-0.014, 0.031) |
| TG (52:4) | Lognormal | -0.003 | (-0.012, 0.006) | 0.479 | 0.856 | 0.040 | (-0.012, 0.006) |
| LPC (15:0) | Lognormal | -0.005 | (-0.018, 0.009) | 0.481 | 0.856 | 0.041 | (-0.018, 0.009) |
| SM (d37:1) | Lognormal | -0.007 | (-0.027, 0.013) | 0.481 | 0.856 | 0.043 | (-0.027, 0.013) |
| PC (37:3) | Lognormal | -0.005 | (-0.018, 0.008) | 0.486 | 0.856 | 0.042 | (-0.018, 0.008) |
| Ceramide (d42:2) A - ESI (+) | Lognormal | 0.003 | (-0.005, 0.010) | 0.487 | 0.856 | 0.038 | (-0.005, 0.010) |
| TG (48:1) | Lognormal | -0.007 | (-0.027, 0.013) | 0.489 | 0.856 | 0.039 | (-0.028, 0.013) |
| PC (33:0) | Lognormal | -0.007 | (-0.025, 0.012) | 0.489 | 0.856 | 0.038 | (-0.025, 0.012) |
| TG (56:3) | Lognormal | 0.007 | (-0.012, 0.025) | 0.489 | 0.856 | 0.041 | (-0.012, 0.025) |
| SM (d36:3) - ESI(+) | Lognormal | -0.004 | (-0.016, 0.008) | 0.493 | 0.857 | 0.037 | (-0.016, 0.007) |
| TG (56:7) A | Lognormal | 0.013 | (-0.025, 0.052) | 0.497 | 0.862 | 0.044 | (-0.024, 0.050) |
| SM (d43:1) - ESI(+) | Lognormal | -0.005 | (-0.019, 0.009) | 0.502 | 0.865 | 0.041 | (-0.019, 0.010) |
| DG (36:4) A | Lognormal | -0.006 | (-0.022, 0.011) | 0.504 | 0.867 | 0.042 | (-0.022, 0.011) |
| FA (20:4) (arachidonic acid) | Lognormal | -0.002 | (-0.009, 0.005) | 0.515 | 0.879 | 0.037 | (-0.009, 0.005) |
| TG (62:1) | Lognormal | -0.009 | (-0.037, 0.019) | 0.520 | 0.883 | 0.040 | (-0.039, 0.018) |
| TG (48:2) | Lognormal | -0.007 | (-0.028, 0.014) | 0.522 | 0.883 | 0.040 | (-0.029, 0.014) |
| TG (54:3) | Lognormal | 0.003 | (-0.007, 0.013) | 0.522 | 0.883 | 0.040 | (-0.007, 0.013) |
| PC (p-42:4)/PC (o-42:5) - ESI(-) | Lognormal | -0.006 | (-0.023, 0.012) | 0.523 | 0.883 | 0.039 | (-0.023, 0.012) |

**Supplementary Table S2.** Regression and Bayesian estimates comparing all ME/CFS vs. all controls.

| Metabolite | Regression Model | ME/CFS vs. Controls | | | | | |
|---|---|---|---|---|---|---|---|
| | | Estimated Coefficient | 95% CI | p-value | FDR | BayesFactor | 95% HDI |
| PC (35:4) - ESI(+) | Lognormal | -0.004 | (-0.015, 0.007) | 0.525 | 0.883 | 0.040 | (-0.014, 0.008) |
| TG (56:1) | Lognormal | -0.008 | (-0.033, 0.017) | 0.533 | 0.890 | 0.038 | (-0.033, 0.017) |
| PC (42:5) | Lognormal | 0.004 | (-0.009, 0.018) | 0.535 | 0.891 | 0.038 | (-0.009, 0.018) |
| PC (33:1) - ESI(+) | Lognormal | 0.003 | (-0.006, 0.012) | 0.537 | 0.891 | 0.040 | (-0.007, 0.012) |
| DG (32:0) | Lognormal | 0.005 | (-0.011, 0.021) | 0.539 | 0.891 | 0.040 | (-0.010, 0.021) |
| TG (59:2) | Lognormal | -0.007 | (-0.031, 0.016) | 0.540 | 0.891 | 0.040 | (-0.031, 0.017) |
| Ceramide (d40:0) | Lognormal | -0.008 | (-0.034, 0.018) | 0.542 | 0.891 | 0.034 | (-0.036, 0.016) |
| LPC (20:1) - ESI(+) | Lognormal | -0.008 | (-0.035, 0.018) | 0.543 | 0.891 | 0.040 | (-0.035, 0.020) |
| LPC (18:1) - ESI(+) | Lognormal | -0.003 | (-0.013, 0.007) | 0.545 | 0.891 | 0.038 | (-0.012, 0.008) |
| FA (15:0) (pentadecylic acid) | Lognormal | -0.003 | (-0.012, 0.006) | 0.546 | 0.891 | 0.038 | (-0.012, 0.005) |
| SM (d34:1) - ESI(+) | Lognormal | -0.001 | (-0.006, 0.003) | 0.546 | 0.891 | 0.042 | (-0.006, 0.003) |
| Ceramide (d36:1) - ESI(-) | Lognormal | -0.006 | (-0.025, 0.013) | 0.548 | 0.891 | 0.038 | (-0.025, 0.013) |
| PC (p-34:1)/PC (o-34:2) B | Lognormal | -0.006 | (-0.026, 0.014) | 0.551 | 0.891 | 0.039 | (-0.026, 0.013) |
| TG (49:0) | Lognormal | -0.006 | (-0.027, 0.014) | 0.553 | 0.891 | 0.039 | (-0.026, 0.015) |
| PC (32:0) - ESI(+) | Lognormal | 0.002 | (-0.004, 0.007) | 0.554 | 0.891 | 0.036 | (-0.003, 0.007) |
| PC (p-40:6)/PC (o-40:7) A | Lognormal | 0.004 | (-0.009, 0.016) | 0.554 | 0.891 | 0.032 | (-0.009, 0.015) |
| LPC (20:1) - ESI(-) | Lognormal | -0.005 | (-0.020, 0.011) | 0.555 | 0.891 | 0.040 | (-0.020, 0.011) |
| TG (53:1) | Lognormal | -0.008 | (-0.036, 0.019) | 0.565 | 0.899 | 0.036 | (-0.036, 0.019) |
| SM (d41:2) - ESI(-) | Lognormal | -0.004 | (-0.017, 0.010) | 0.571 | 0.901 | 0.036 | (-0.017, 0.010) |
| PC (p-42:5)/PC (o-42:6) | Lognormal | -0.008 | (-0.035, 0.019) | 0.572 | 0.901 | 0.038 | (-0.036, 0.019) |
| PC (38:5) A - ESI(-) | Lognormal | -0.003 | (-0.012, 0.007) | 0.572 | 0.901 | 0.041 | (-0.011, 0.007) |
| FA (20:2) (eicosadienoic acid) | Lognormal | 0.003 | (-0.008, 0.014) | 0.572 | 0.901 | 0.036 | (-0.007, 0.015) |
| PC (35:2) B | Lognormal | -0.002 | (-0.008, 0.004) | 0.579 | 0.907 | 0.034 | (-0.008, 0.005) |
| SM (d43:2) - ESI(+) | Lognormal | 0.004 | (-0.011, 0.020) | 0.583 | 0.907 | 0.039 | (-0.012, 0.019) |
| PC (38:4) C - ESI(+) | Lognormal | -0.002 | (-0.007, 0.004) | 0.586 | 0.909 | 0.038 | (-0.007, 0.004) |
| TG (46:3) B | Lognormal | -0.010 | (-0.045, 0.026) | 0.601 | 0.915 | 0.036 | (-0.046, 0.025) |
| AC (16:0) | Lognormal | -0.003 | (-0.013, 0.007) | 0.602 | 0.915 | 0.039 | (-0.013, 0.007) |
| GlcCer (d40:1) - ESI(+) | Lognormal | -0.003 | (-0.016, 0.009) | 0.605 | 0.915 | 0.034 | (-0.016, 0.009) |
| SM (d32:2) - ESI(+) | Lognormal | -0.003 | (-0.014, 0.008) | 0.606 | 0.915 | 0.034 | (-0.014, 0.008) |
| PC (39:4) | Lognormal | 0.003 | (-0.008, 0.013) | 0.607 | 0.915 | 0.035 | (-0.008, 0.013) |
| Ceramide (d34:1) - ESI(+) | Lognormal | 0.002 | (-0.006, 0.010) | 0.611 | 0.915 | 0.037 | (-0.006, 0.011) |
| PE (p-36:5)/PE (o-36:6) | Lognormal | -0.014 | (-0.068, 0.040) | 0.613 | 0.915 | 0.034 | (-0.064, 0.044) |
| TG (58:5) | Lognormal | -0.004 | (-0.022, 0.013) | 0.617 | 0.915 | 0.036 | (-0.022, 0.014) |
| TG (51:5) | Lognormal | -0.005 | (-0.023, 0.014) | 0.619 | 0.915 | 0.041 | (-0.022, 0.013) |
| FA (17:0) (margaric acid) | Lognormal | 0.002 | (-0.005, 0.008) | 0.623 | 0.915 | 0.037 | (-0.005, 0.008) |
| SM (d34:2) - ESI(+) | Lognormal | -0.001 | (-0.007, 0.004) | 0.623 | 0.915 | 0.034 | (-0.007, 0.004) |
| PC (p-44:4)/PC (o-44:5) - ESI(+) | Lognormal | 0.003 | (-0.009, 0.014) | 0.626 | 0.915 | 0.035 | (-0.009, 0.014) |
| FA (22:6) (docosahexaenoic acid) | Lognormal | 0.003 | (-0.011, 0.018) | 0.629 | 0.915 | 0.035 | (-0.009, 0.018) |
| TG (58:3) | Lognormal | 0.007 | (-0.020, 0.034) | 0.630 | 0.915 | 0.038 | (-0.021, 0.034) |
| PC (36:5) B | Lognormal | 0.005 | (-0.016, 0.027) | 0.630 | 0.915 | 0.039 | (-0.017, 0.027) |
| TG (60:1) | Lognormal | -0.007 | (-0.035, 0.021) | 0.634 | 0.918 | 0.038 | (-0.034, 0.023) |
| AC (18:1) | Lognormal | -0.003 | (-0.015, 0.009) | 0.639 | 0.920 | 0.034 | (-0.014, 0.009) |
| CE (18:0) | Lognormal | 0.005 | (-0.016, 0.025) | 0.644 | 0.922 | 0.036 | (-0.015, 0.026) |
| TG (54:2) | Lognormal | 0.003 | (-0.011, 0.017) | 0.647 | 0.924 | 0.036 | (-0.010, 0.018) |
| TG (52:1) | Lognormal | -0.004 | (-0.023, 0.014) | 0.647 | 0.924 | 0.035 | (-0.023, 0.015) |
| PC (p-38:6)/PC (o-38:7) | Lognormal | -0.003 | (-0.017, 0.010) | 0.655 | 0.927 | 0.034 | (-0.016, 0.012) |
| TG (50:2) | Lognormal | 0.002 | (-0.008, 0.013) | 0.663 | 0.932 | 0.032 | (-0.008, 0.013) |

**Supplementary Table S2.** Regression and Bayesian estimates comparing all ME/CFS vs. all controls.

| Metabolite | Regression Model | ME/CFS vs. Controls | | | | | |
|---|---|---|---|---|---|---|---|
| | | Estimated Coefficient | 95% CI | p-value | FDR | BayesFactor | 95% HDI |
| TG (57:1) | Lognormal | -0.004 | (-0.023, 0.014) | 0.664 | 0.932 | 0.036 | (-0.024, 0.013) |
| LPC (17:1) | Lognormal | 0.003 | (-0.011, 0.017) | 0.664 | 0.932 | 0.036 | (-0.010, 0.018) |
| PC (p-44:5)/PC (o-44:6) | Lognormal | 0.003 | (-0.011, 0.017) | 0.666 | 0.932 | 0.036 | (-0.010, 0.018) |
| SM (d34:0) - ESI(+) | Lognormal | -0.002 | (-0.009, 0.006) | 0.666 | 0.932 | 0.036 | (-0.009, 0.005) |
| PC (38:5) B - ESI(-) | Lognormal | 0.004 | (-0.013, 0.020) | 0.668 | 0.932 | 0.038 | (-0.012, 0.020) |
| LPC (p-18:0)/LPC (o-18:1) | Lognormal | -0.003 | (-0.017, 0.011) | 0.677 | 0.932 | 0.037 | (-0.017, 0.011) |
| PC (37:4) - ESI(+) | Lognormal | 0.003 | (-0.010, 0.015) | 0.679 | 0.932 | 0.037 | (-0.011, 0.014) |
| TG (54:8) | Lognormal | 0.005 | (-0.020, 0.031) | 0.681 | 0.932 | 0.037 | (-0.022, 0.029) |
| PC (40:6) B | Lognormal | 0.003 | (-0.010, 0.015) | 0.685 | 0.932 | 0.034 | (-0.009, 0.016) |
| TG (53:4) | Lognormal | 0.002 | (-0.010, 0.015) | 0.686 | 0.932 | 0.038 | (-0.010, 0.014) |
| LPC (20:0) | Lognormal | -0.003 | (-0.019, 0.013) | 0.686 | 0.932 | 0.035 | (-0.019, 0.014) |
| TG (58:1) | Lognormal | -0.005 | (-0.031, 0.021) | 0.690 | 0.932 | 0.035 | (-0.029, 0.023) |
| PE (36:4) - ESI(+) | Lognormal | -0.004 | (-0.025, 0.017) | 0.691 | 0.932 | 0.034 | (-0.025, 0.017) |
| PC (p-40:7)/PC (o-40:8) | Lognormal | -0.008 | (-0.046, 0.031) | 0.693 | 0.932 | 0.033 | (-0.049, 0.029) |
| Ceramide (d36:1) - ESI(+) | Lognormal | 0.002 | (-0.009, 0.014) | 0.695 | 0.933 | 0.036 | (-0.008, 0.014) |
| TG (54:4) | Lognormal | 0.002 | (-0.009, 0.013) | 0.695 | 0.933 | 0.036 | (-0.009, 0.012) |
| PC (31:1) | Lognormal | -0.007 | (-0.041, 0.028) | 0.698 | 0.935 | 0.037 | (-0.040, 0.029) |
| PE (38:6) - ESI(-) | Lognormal | -0.005 | (-0.031, 0.021) | 0.704 | 0.936 | 0.035 | (-0.029, 0.022) |
| LPC (22:5) - ESI(+) | Lognormal | 0.004 | (-0.015, 0.023) | 0.707 | 0.936 | 0.034 | (-0.014, 0.023) |
| PC (38:4) A - ESI(+) | Lognormal | -0.002 | (-0.011, 0.007) | 0.708 | 0.936 | 0.038 | (-0.011, 0.008) |
| TG (52:5) | Lognormal | 0.002 | (-0.010, 0.015) | 0.708 | 0.936 | 0.038 | (-0.011, 0.015) |
| PC (34:0) - ESI(+) | Lognormal | -0.001 | (-0.007, 0.004) | 0.708 | 0.936 | 0.030 | (-0.006, 0.005) |
| TG (51:3) | Lognormal | 0.002 | (-0.010, 0.014) | 0.714 | 0.936 | 0.034 | (-0.010, 0.015) |
| TG (59:3) | Lognormal | 0.004 | (-0.018, 0.026) | 0.717 | 0.936 | 0.035 | (-0.018, 0.024) |
| Gal-Gal-Cer (d18:1/16:0)/Lactosylceramide (d18:1/16:0) | Lognormal | 0.002 | (-0.007, 0.011) | 0.717 | 0.936 | 0.031 | (-0.007, 0.011) |
| PC 40:6e | Lognormal | 0.002 | (-0.008, 0.011) | 0.720 | 0.936 | 0.035 | (-0.008, 0.011) |
| TG (52:6) | Lognormal | -0.003 | (-0.022, 0.015) | 0.726 | 0.936 | 0.033 | (-0.021, 0.016) |
| TG (48:6) | Lognormal | 0.004 | (-0.019, 0.028) | 0.729 | 0.936 | 0.037 | (-0.018, 0.028) |
| DG (34:1) | Lognormal | -0.004 | (-0.028, 0.020) | 0.731 | 0.936 | 0.033 | (-0.027, 0.020) |
| FA (14:0) (myristic acid) | Lognormal | 0.001 | (-0.007, 0.010) | 0.732 | 0.936 | 0.036 | (-0.007, 0.010) |
| PE (36:1) - ESI(-) | Lognormal | -0.005 | (-0.031, 0.022) | 0.733 | 0.936 | 0.036 | (-0.030, 0.023) |
| PC (36:1) - ESI(+) | Lognormal | 0.001 | (-0.005, 0.007) | 0.733 | 0.936 | 0.036 | (-0.005, 0.007) |
| TAG (58:7)/TAG (18:1/18:1/22:5) | Lognormal | 0.002 | (-0.011, 0.015) | 0.734 | 0.936 | 0.035 | (-0.012, 0.014) |
| TG (50:3) B | Lognormal | -0.013 | (-0.088, 0.062) | 0.735 | 0.936 | 0.035 | (-0.084, 0.070) |
| TG (56:10) | Lognormal | 0.005 | (-0.026, 0.036) | 0.739 | 0.936 | 0.032 | (-0.027, 0.037) |
| SM (d41:2) A - ESI(+) | Lognormal | 0.002 | (-0.008, 0.012) | 0.741 | 0.936 | 0.033 | (-0.009, 0.012) |
| LPC (22:5) - ESI(-) | Lognormal | -0.005 | (-0.038, 0.027) | 0.741 | 0.936 | 0.031 | (-0.037, 0.026) |
| TG (54:9) | Lognormal | 0.009 | (-0.047, 0.066) | 0.742 | 0.936 | 0.033 | (-0.049, 0.064) |
| FA (20:3) (eicosatrienoic acid) | Lognormal | 0.002 | (-0.012, 0.017) | 0.747 | 0.936 | 0.037 | (-0.013, 0.016) |
| SM (d42:0) - ESI(-) | Lognormal | -0.002 | (-0.014, 0.010) | 0.749 | 0.936 | 0.031 | (-0.013, 0.011) |
| SM (d42:3) - ESI(+) | Lognormal | 0.001 | (-0.005, 0.008) | 0.753 | 0.936 | 0.029 | (-0.005, 0.008) |
| CE (20:4) | Lognormal | -0.001 | (-0.011, 0.008) | 0.759 | 0.939 | 0.033 | (-0.011, 0.007) |
| PC 38:7e | Lognormal | -0.002 | (-0.019, 0.014) | 0.776 | 0.947 | 0.035 | (-0.020, 0.014) |
| Ceramide (d40:2) | Lognormal | 0.006 | (-0.034, 0.046) | 0.781 | 0.949 | 0.037 | (-0.033, 0.046) |
| TG (51:4) | Lognormal | -0.002 | (-0.017, 0.013) | 0.786 | 0.950 | 0.035 | (-0.017, 0.013) |
| PC (p-40:4)/PC (o-40:5) - ESI(+) | Lognormal | -0.001 | (-0.010, 0.008) | 0.789 | 0.950 | 0.034 | (-0.010, 0.008) |
| TG (57:2) | Lognormal | -0.002 | (-0.018, 0.014) | 0.789 | 0.950 | 0.033 | (-0.018, 0.015) |

**Supplementary Table S2.** Regression and Bayesian estimates comparing all ME/CFS vs. all controls.

| Metabolite | Regression Model | ME/CFS vs. Controls | | | | | |
|---|---|---|---|---|---|---|---|
| | | Estimated Coefficient | 95% CI | p-value | FDR | BayesFactor | 95% HDI |
| DG (34:3) | Lognormal | 0.002 | (-0.016, 0.021) | 0.794 | 0.950 | 0.030 | (-0.015, 0.022) |
| TG (55:1) | Lognormal | -0.003 | (-0.024, 0.018) | 0.795 | 0.950 | 0.034 | (-0.024, 0.018) |
| TG (62:2) | Lognormal | -0.004 | (-0.035, 0.027) | 0.797 | 0.950 | 0.034 | (-0.034, 0.029) |
| SM (d38:0) | Lognormal | -0.008 | (-0.068, 0.052) | 0.799 | 0.950 | 0.034 | (-0.069, 0.053) |
| PC (36:6) | Lognormal | -0.002 | (-0.021, 0.016) | 0.801 | 0.950 | 0.031 | (-0.022, 0.015) |
| TG (54:6) A | Lognormal | -0.002 | (-0.018, 0.014) | 0.814 | 0.950 | 0.033 | (-0.018, 0.015) |
| CE (18:1) | Lognormal | -0.001 | (-0.014, 0.011) | 0.814 | 0.950 | 0.036 | (-0.014, 0.011) |
| Ceramide (d34:2) | Lognormal | -0.001 | (-0.012, 0.009) | 0.817 | 0.950 | 0.034 | (-0.012, 0.010) |
| SM (d40:0) | Lognormal | 0.001 | (-0.011, 0.014) | 0.821 | 0.950 | 0.034 | (-0.011, 0.014) |
| PC (36:4) C - ESI(+) | Lognormal | -0.001 | (-0.006, 0.005) | 0.824 | 0.950 | 0.034 | (-0.006, 0.005) |
| TG (50:6) | Lognormal | 0.003 | (-0.024, 0.030) | 0.826 | 0.950 | 0.032 | (-0.022, 0.030) |
| PC (40:8) - ESI(+) | Lognormal | -0.001 | (-0.010, 0.008) | 0.831 | 0.950 | 0.030 | (-0.010, 0.008) |
| DG (32:1) | Lognormal | -0.002 | (-0.025, 0.020) | 0.832 | 0.950 | 0.033 | (-0.025, 0.021) |
| SM (d33:1) - ESI(+) | Lognormal | -0.001 | (-0.011, 0.009) | 0.832 | 0.950 | 0.033 | (-0.010, 0.009) |
| TG (64:2) | Lognormal | -0.003 | (-0.034, 0.028) | 0.833 | 0.950 | 0.034 | (-0.035, 0.027) |
| PC (16:0/9:0(CHO)) | Lognormal | -0.002 | (-0.023, 0.018) | 0.834 | 0.950 | 0.028 | (-0.024, 0.016) |
| TG (46:5) | Lognormal | 0.005 | (-0.039, 0.048) | 0.835 | 0.950 | 0.035 | (-0.038, 0.048) |
| SM (d38:2) - ESI(+) | Lognormal | -0.001 | (-0.007, 0.006) | 0.837 | 0.950 | 0.032 | (-0.007, 0.006) |
| DG (36:5) | Lognormal | -0.002 | (-0.023, 0.018) | 0.839 | 0.950 | 0.031 | (-0.022, 0.019) |
| DG (34:2) | Lognormal | 0.001 | (-0.011, 0.013) | 0.843 | 0.953 | 0.028 | (-0.010, 0.013) |
| TG (55:2) | Lognormal | 0.002 | (-0.016, 0.019) | 0.844 | 0.953 | 0.027 | (-0.016, 0.019) |
| SM (d36:0) - ESI(-) | Lognormal | 0.003 | (-0.025, 0.030) | 0.849 | 0.956 | 0.035 | (-0.026, 0.030) |
| CE (16:0) | Lognormal | 0.003 | (-0.024, 0.029) | 0.850 | 0.957 | 0.033 | (-0.024, 0.030) |
| PC (35:1) - ESI(-) | Lognormal | 0.001 | (-0.013, 0.015) | 0.858 | 0.963 | 0.032 | (-0.013, 0.015) |
| TG (60:2) | Lognormal | -0.004 | (-0.044, 0.037) | 0.860 | 0.963 | 0.033 | (-0.047, 0.036) |
| FA (24:1) (nervonic acid) | Lognormal | 0.001 | (-0.009, 0.010) | 0.865 | 0.963 | 0.030 | (-0.009, 0.010) |
| PC (p-40:6)/PC (o-40:7) B | Lognormal | -0.001 | (-0.017, 0.014) | 0.869 | 0.963 | 0.031 | (-0.017, 0.014) |
| PC (40:7) - ESI(-) | Lognormal | -0.002 | (-0.020, 0.017) | 0.870 | 0.963 | 0.033 | (-0.019, 0.017) |
| TG (62:3) | Lognormal | -0.003 | (-0.038, 0.032) | 0.871 | 0.963 | 0.029 | (-0.037, 0.033) |
| LPC (22:6) | Lognormal | 0.001 | (-0.013, 0.015) | 0.872 | 0.963 | 0.030 | (-0.013, 0.016) |
| GlcCer (d42:1) - ESI(+) | Lognormal | 0.001 | (-0.009, 0.010) | 0.875 | 0.963 | 0.029 | (-0.009, 0.010) |
| DG (36:3) | Lognormal | 0.001 | (-0.010, 0.012) | 0.877 | 0.963 | 0.031 | (-0.010, 0.011) |
| PC (42:6) | Lognormal | 0.002 | (-0.023, 0.027) | 0.877 | 0.963 | 0.033 | (-0.022, 0.028) |
| PC (p-36:5)/PC (o-36:6) | Lognormal | 0.002 | (-0.019, 0.022) | 0.878 | 0.963 | 0.035 | (-0.019, 0.021) |
| TG (56:4) | Lognormal | 0.001 | (-0.012, 0.014) | 0.879 | 0.964 | 0.035 | (-0.012, 0.014) |
| TG (52:3) | Lognormal | 0.000 | (-0.006, 0.006) | 0.892 | 0.972 | 0.033 | (-0.006, 0.007) |
| FA (12:0) (lauric acid) | Lognormal | -0.001 | (-0.017, 0.015) | 0.893 | 0.972 | 0.031 | (-0.018, 0.015) |
| PC (p-38:5)/PC (o-38:6) A | Lognormal | -0.001 | (-0.015, 0.013) | 0.901 | 0.975 | 0.032 | (-0.015, 0.013) |
| CE (16:1) | Lognormal | 0.001 | (-0.013, 0.015) | 0.903 | 0.975 | 0.031 | (-0.013, 0.015) |
| SM (d41:1) - ESI(-) | Lognormal | 0.006 | (-0.090, 0.102) | 0.904 | 0.975 | 0.035 | (-0.092, 0.098) |
| PC (32:1) - ESI(+) | Lognormal | -0.001 | (-0.012, 0.011) | 0.911 | 0.976 | 0.032 | (-0.011, 0.010) |
| SM (d36:2) - ESI(+) | Lognormal | 0.000 | (-0.007, 0.006) | 0.921 | 0.982 | 0.033 | (-0.007, 0.006) |
| PC (40:6) A | Lognormal | -0.001 | (-0.012, 0.011) | 0.929 | 0.985 | 0.034 | (-0.012, 0.011) |
| TG (60:3) | Lognormal | 0.001 | (-0.029, 0.032) | 0.934 | 0.987 | 0.035 | (-0.031, 0.030) |
| GlcCer (d34:1) | Lognormal | -0.001 | (-0.022, 0.020) | 0.936 | 0.987 | 0.033 | (-0.021, 0.020) |
| TG (44:2) | Lognormal | -0.003 | (-0.077, 0.071) | 0.937 | 0.987 | 0.036 | (-0.079, 0.068) |
| DG (36:1) | Lognormal | 0.001 | (-0.016, 0.017) | 0.942 | 0.987 | 0.032 | (-0.015, 0.017) |

**Supplementary Table S2.** Regression and Bayesian estimates comparing all ME/CFS vs. all controls.

| Metabolite | Regression Model | ME/CFS vs. Controls | | | | | |
|---|---|---|---|---|---|---|---|
| | | Estimated Coefficient | 95% CI | p-value | FDR | BayesFactor | 95% HDI |
| SM (d32:0) - ESI(+) | Lognormal | 0.001 | (-0.015, 0.016) | 0.942 | 0.987 | 0.031 | (-0.014, 0.018) |
| PC (33:1) - ESI(-) | Lognormal | -0.001 | (-0.021, 0.020) | 0.942 | 0.987 | 0.035 | (-0.022, 0.022) |
| TG (50:3) A | Lognormal | 0.000 | (-0.013, 0.014) | 0.945 | 0.988 | 0.033 | (-0.013, 0.015) |
| PE (38:6) - ESI(+) | Lognormal | 0.001 | (-0.028, 0.030) | 0.949 | 0.990 | 0.033 | (-0.030, 0.029) |
| PC (p-40:4)/PC (o-40:5) - ESI(-) | Lognormal | 0.000 | (-0.013, 0.014) | 0.958 | 0.990 | 0.033 | (-0.013, 0.014) |
| TG (49:1) | Lognormal | 0.001 | (-0.021, 0.022) | 0.962 | 0.990 | 0.033 | (-0.020, 0.022) |
| PC (38:6) - ESI(-) | Lognormal | 0.000 | (-0.012, 0.012) | 0.965 | 0.990 | 0.034 | (-0.011, 0.012) |
| TG (56:2) | Lognormal | 0.000 | (-0.022, 0.021) | 0.967 | 0.990 | 0.032 | (-0.021, 0.021) |
| FA (10:0) (capric acid) | Lognormal | 0.000 | (-0.024, 0.025) | 0.969 | 0.990 | 0.029 | (-0.022, 0.027) |
| PC (31:0) | Lognormal | 0.000 | (-0.013, 0.012) | 0.970 | 0.990 | 0.033 | (-0.013, 0.013) |
| PC (p-38:5)/PC (o-38:6) | Lognormal | -0.001 | (-0.038, 0.036) | 0.972 | 0.990 | 0.036 | (-0.036, 0.034) |
| SM (d42:0) - ESI(+) | Lognormal | 0.000 | (-0.026, 0.025) | 0.973 | 0.990 | 0.032 | (-0.026, 0.025) |
| PC (36:4) B - ESI(+) | Lognormal | 0.000 | (-0.012, 0.013) | 0.974 | 0.990 | 0.032 | (-0.012, 0.013) |
| PC (40:5) A - ESI(-) | Lognormal | 0.000 | (-0.015, 0.015) | 0.975 | 0.990 | 0.031 | (-0.016, 0.015) |
| TG (49:3) | Lognormal | 0.000 | (-0.021, 0.021) | 0.977 | 0.992 | 0.030 | (-0.021, 0.022) |
| PC (p-42:5)/PC (o-42:6) B | Lognormal | 0.000 | (-0.014, 0.014) | 0.980 | 0.992 | 0.035 | (-0.014, 0.014) |
| TG (60:4) | Lognormal | 0.000 | (-0.030, 0.031) | 0.980 | 0.992 | 0.033 | (-0.030, 0.032) |
| TG (53:5) | Lognormal | 0.000 | (-0.015, 0.015) | 0.986 | 0.994 | 0.033 | (-0.016, 0.014) |
| TG (54:5) A | Lognormal | 0.000 | (-0.012, 0.012) | 0.990 | 0.994 | 0.031 | (-0.012, 0.012) |
| PC (p-42:4)/PC (o-42:5) - ESI(+) | Lognormal | 0.000 | (-0.011, 0.011) | 0.992 | 0.994 | 0.031 | (-0.011, 0.011) |
| SM (d36:1) - ESI(+) | Lognormal | 0.000 | (-0.006, 0.006) | 0.994 | 0.995 | 0.033 | (-0.007, 0.006) |
| **Oxylipins (OL)** | | | | | | | |
| Resolvin D1 | Gamma | **-0.528** | **(-0.846, -0.210)** | **0.002** | **0.134** | **6.635** | **(-0.797, -0.224)** |
| 12-Hydroxy-5,8,10,14,17-eicosapentaenoic acid | Gamma | 0.454 | (0.092, 0.819) | 0.011 | 0.188 | 3.134 | (0.169, 0.724) |
| 11,12-Epoxyeicosa-5,8,14-trienoic acid | Gamma | 0.317 | (0.056, 0.578) | 0.018 | 0.213 | 0.442 | (0.067, 0.588) |
| Prostaglandin F2a | Gamma | -0.293 | (-0.560, -0.026) | 0.026 | 0.247 | 0.239 | (-0.577, 0.001) |
| 9,11,15-trihydroxy-5,13,1Z-prostatrienoic acid | Gamma | -0.576 | (-1.135, -0.019) | 0.031 | 0.268 | 3.858 | (-0.932, -0.211) |
| 5-Hydroxy-6,8,11,14,17-eicosapentaenoic acid | Gamma | 0.245 | (0.010, 0.481) | 0.034 | 0.285 | 0.311 | (0.040, 0.468) |
| 8,9-Epoxyeicosa-5,11,14-trienoic acid | Gamma | -0.260 | (-0.520, 0.001) | 0.040 | 0.302 | 0.165 | (-0.529, -0.002) |
| 16(17)-epoxy-4,7,10,13,19-docosapentaenoic acid | Gamma | 0.343 | (-0.022, 0.709) | 0.052 | 0.348 | 0.257 | (0.012, 0.714) |
| 9-Hydroxy-5,7,11,14,17-icosapentaenoic acid | Gamma | 0.242 | (-0.015, 0.500) | 0.053 | 0.351 | 0.180 | (-0.002, 0.490) |
| 15-hydroxyeicosa-5,8,11,13,17-pentaenoic acid | Gamma | 0.323 | (-0.020, 0.669) | 0.057 | 0.365 | 0.228 | (-0.014, 0.645) |
| 17,18-Epoxy-5,8,11,14-eicosatetraenoic acid | Gamma | -0.420 | (-0.849, 0.007) | 0.060 | 0.368 | 0.576 | (-0.785, -0.075) |
| Leukotriene B4 | Gamma | -0.193 | (-0.411, 0.026) | 0.081 | 0.422 | 0.127 | (-0.389, 0.005) |
| 17,18-dihydroxyeicosa-5,8,11,14-tetraenoic acid | Lognormal | 0.209 | (-0.029, 0.447) | 0.086 | 0.430 | 0.132 | (-0.032, 0.439) |
| 15,16-dihydroxyoctadeca-9,12-dienoic acid | Lognormal | -0.157 | (-0.335, 0.022) | 0.087 | 0.430 | 0.106 | (-0.327, 0.033) |
| 11-Hydroxy-14,15-epoxyeicosatrienoic acid | Gamma | -0.402 | (-0.902, 0.104) | 0.099 | 0.461 | 0.248 | (-0.824, 0.010) |
| Prostaglandin D2 | Gamma | -0.407 | (-0.902, 0.091) | 0.105 | 0.467 | 0.309 | (-0.792, -0.008) |
| 13-ketooctadeca-9,11-dienoic acid | Lognormal | -0.199 | (-0.446, 0.047) | 0.115 | 0.486 | 0.109 | (-0.439, 0.057) |
| 4-hydroxydocosa-5,7,10,13,16,19-hexaenoic acid | Gamma | 0.221 | (-0.075, 0.518) | 0.137 | 0.532 | 0.089 | (-0.054, 0.531) |
| 15(16)-epoxy-9,12-octadecadienoic acid | Gamma | -0.264 | (-0.655, 0.129) | 0.155 | 0.564 | 0.124 | (-0.612, 0.056) |
| 9,10-dihydroxyoctadec-12-enoic acid | Lognormal | -0.134 | (-0.322, 0.054) | 0.164 | 0.581 | 0.069 | (-0.319, 0.057) |
| 9-Hydroxylinoleic acid | Lognormal | -0.078 | (-0.202, 0.045) | 0.215 | 0.634 | 0.064 | (-0.206, 0.039) |
| 15-Deoxy-delta-12,14-Prostaglandin J2 | Gamma | -0.146 | (-0.408, 0.116) | 0.296 | 0.740 | 0.048 | (-0.416, 0.119) |
| 5-ketoeicosa-6,8,11,14-tetraenoic acid | Gamma | -0.151 | (-0.447, 0.146) | 0.303 | 0.740 | 0.050 | (-0.471, 0.158) |
| 14(15)-epoxy-5,8,11,17-eicosatetraenoic acid | Gamma | 0.280 | (-0.334, 0.895) | 0.308 | 0.743 | 0.090 | (-0.208, 0.775) |
| 17-hydroxy-4,7,10,13,15,19-docosahexaenoic acid | Gamma | 0.118 | (-0.119, 0.357) | 0.309 | 0.743 | 0.038 | (-0.109, 0.376) |

**Supplementary Table S2.** Regression and Bayesian estimates comparing all ME/CFS vs. all controls.

| Metabolite | Regression Model | ME/CFS vs. Controls | | | | | |
|---|---|---|---|---|---|---|---|
| | | Estimated Coefficient | 95% CI | p-value | FDR | BayesFactor | 95% HDI |
| 15-Keto-prostaglandin E2 | Gamma | -0.211 | (-0.644, 0.224) | 0.317 | 0.743 | 0.067 | (-0.578, 0.151) |
| 12,13-dihydroxyoctadec-9-enoic acid | Lognormal | -0.090 | (-0.268, 0.087) | 0.321 | 0.743 | 0.046 | (-0.270, 0.083) |
| Prostaglandin E3 | Gamma | 0.280 | (-0.378, 0.944) | 0.330 | 0.747 | 0.095 | (-0.259, 0.853) |
| 10-nitrolinoleic acid | Gamma | 0.190 | (-0.251, 0.635) | 0.362 | 0.764 | 0.060 | (-0.168, 0.589) |
| 8,9-dihydroxyeicosa-5,11,14-trienoic acid | Gamma | 0.102 | (-0.118, 0.322) | 0.364 | 0.764 | 0.042 | (-0.120, 0.338) |
| Prostaglandin E1 | Gamma | 0.162 | (-0.203, 0.529) | 0.366 | 0.764 | 0.050 | (-0.166, 0.482) |
| 9,10-dihydroxyoctadeca-12,15-dienoic acid | Gamma | -0.090 | (-0.293, 0.114) | 0.380 | 0.768 | 0.031 | (-0.326, 0.130) |
| 15-ketoeicosa-5,8,11,13-tetraenoic acid | Gamma | 0.146 | (-0.212, 0.505) | 0.419 | 0.815 | 0.049 | (-0.145, 0.435) |
| Leukotriene B5 | Gamma | -0.198 | (-0.691, 0.295) | 0.443 | 0.834 | 0.066 | (-0.614, 0.227) |
| 12,13-epoxy-9-octadecenoic acid | Lognormal | -0.108 | (-0.392, 0.176) | 0.457 | 0.848 | 0.042 | (-0.368, 0.199) |
| 6-Ketoprostaglandin F1 alpha | Gamma | 0.143 | (-0.257, 0.544) | 0.468 | 0.853 | 0.048 | (-0.209, 0.484) |
| 9,12,13-trihydroxyoctadec-10-enoic acid | Lognormal | -0.045 | (-0.172, 0.083) | 0.491 | 0.856 | 0.035 | (-0.180, 0.068) |
| 5-Hydroxy-6,8,11,14-eicosatetraenoic acid | Lognormal | -0.058 | (-0.244, 0.127) | 0.538 | 0.891 | 0.034 | (-0.243, 0.138) |
| 8-hydroxyeicosa-5,9,11,14-tetraenoic acid | Gamma | 0.071 | (-0.163, 0.306) | 0.550 | 0.891 | 0.032 | (-0.167, 0.331) |
| 14,15-dihydroxyeicosa-5,8,11,17-tetraenoic acid | Gamma | -0.091 | (-0.413, 0.232) | 0.557 | 0.893 | 0.037 | (-0.396, 0.245) |
| 20-Hydroxyarachidonic acid | Lognormal | -0.100 | (-0.448, 0.247) | 0.572 | 0.901 | 0.031 | (-0.486, 0.237) |
| 12,13-dihydroxyoctadeca-9,15-dienoic acid | Gamma | -0.128 | (-0.587, 0.336) | 0.573 | 0.901 | 0.047 | (-0.457, 0.207) |
| 10-nitrooleic acid | Lognormal | 0.063 | (-0.160, 0.286) | 0.581 | 0.907 | 0.034 | (-0.151, 0.286) |
| 12(13)-epoxy-9,15-octadecadienoic acid | Gamma | -0.101 | (-0.485, 0.283) | 0.594 | 0.913 | 0.039 | (-0.397, 0.241) |
| 9-hydroxyoctadeca-10,12,15-trienoic acid | Gamma | -0.067 | (-0.315, 0.181) | 0.598 | 0.915 | 0.028 | (-0.302, 0.168) |
| 8,15-dihydroxyeicosa-5,9,11,13-tetraenoic acid | Gamma | -0.125 | (-0.625, 0.383) | 0.631 | 0.915 | 0.053 | (-0.544, 0.337) |
| 9,10-Epoxystearic acid | Lognormal | -0.071 | (-0.368, 0.225) | 0.638 | 0.920 | 0.036 | (-0.358, 0.245) |
| 19,20-dihydroxydocosa-4,7,10,13,16-pentaenoic acid | Lognormal | 0.035 | (-0.123, 0.193) | 0.663 | 0.932 | 0.033 | (-0.126, 0.179) |
| 5,6-dihydroxyeicosa-8,11,14-trienoic acid | Gamma | 0.047 | (-0.172, 0.265) | 0.671 | 0.932 | 0.025 | (-0.168, 0.262) |
| 18-(3-ethyloxiran-2-yl)octadeca-4,7,10,13,16-pentaenoic acid | Gamma | 0.074 | (-0.293, 0.441) | 0.673 | 0.932 | 0.034 | (-0.238, 0.384) |
| 6-trans-Leukotriene B4 | Gamma | -0.102 | (-0.621, 0.426) | 0.678 | 0.932 | 0.044 | (-0.566, 0.303) |
| 14,15-dihydroxyeicosa-5,8,11-trienoic acid | Gamma | -0.028 | (-0.164, 0.108) | 0.689 | 0.932 | 0.015 | (-0.169, 0.110) |
| 11,12-Dihydroxyicosa-5,8,14-trienoic acid | Gamma | -0.027 | (-0.173, 0.120) | 0.723 | 0.936 | 0.016 | (-0.167, 0.122) |
| 11-Hydroxy-arachidonic acid | Gamma | 0.031 | (-0.144, 0.205) | 0.725 | 0.936 | 0.021 | (-0.151, 0.219) |
| 5,6,15-trihydroxyeicosa-7,9,11,13-tetraenoic acid | Gamma | -0.055 | (-0.385, 0.277) | 0.745 | 0.936 | 0.031 | (-0.388, 0.235) |
| Prostaglandin E2 | Gamma | 0.080 | (-0.433, 0.596) | 0.750 | 0.936 | 0.041 | (-0.293, 0.422) |
| 9S,10R-dihydroxy-stearic acid | Lognormal | -0.041 | (-0.305, 0.223) | 0.761 | 0.939 | 0.031 | (-0.298, 0.227) |
| Thromboxane B2 | Lognormal | 0.036 | (-0.205, 0.277) | 0.771 | 0.945 | 0.034 | (-0.194, 0.279) |
| 9(10)-epoxy-12Z-octadecenoic acid | Gamma | -0.027 | (-0.235, 0.181) | 0.797 | 0.950 | 0.020 | (-0.199, 0.150) |
| 9-nitrooleic acid | Gamma | 0.032 | (-0.270, 0.337) | 0.828 | 0.950 | 0.031 | (-0.288, 0.326) |
| 9-ketooctadeca-10,12-dienoic acid | Gamma | -0.024 | (-0.240, 0.192) | 0.830 | 0.950 | 0.026 | (-0.271, 0.217) |
| 13-Hydroxyoctadecadienoic acid | Lognormal | -0.012 | (-0.128, 0.104) | 0.835 | 0.950 | 0.031 | (-0.120, 0.109) |
| trans-12,13-epoxy-11-oxo-trans-9-octadecenoic acid | Gamma | 0.019 | (-0.167, 0.205) | 0.840 | 0.950 | 0.022 | (-0.165, 0.226) |
| 5,15-dihydroxyeicosa-6,8,11,13-tetraenoic acid | Gamma | 0.029 | (-0.423, 0.482) | 0.894 | 0.972 | 0.039 | (-0.393, 0.433) |
| 9-hydroxyeicosa-5,7,11,14-tetraenoic acid | Gamma | -0.006 | (-0.226, 0.215) | 0.960 | 0.990 | 0.025 | (-0.249, 0.230) |
| 13-hydroxyoctadeca-9,11,15-trienoic acid | Gamma | 0.006 | (-0.232, 0.243) | 0.960 | 0.990 | 0.025 | (-0.229, 0.233) |
| 11(12)-epoxy-5,8,14,17- eicosatetraenoic acid | Gamma | -0.009 | (-0.419, 0.407) | 0.965 | 0.990 | 0.040 | (-0.385, 0.408) |
| 12-Hydroxy-5,8,10,14-eicosatetraenoic acid | Lognormal | -0.004 | (-0.215, 0.206) | 0.967 | 0.990 | 0.025 | (-0.211, 0.217) |
| 14-hydroxydocosa-4,7,10,12,16,19-hexaenoic acid | Lognormal | -0.006 | (-0.343, 0.332) | 0.974 | 0.990 | 0.022 | (-0.347, 0.318) |
| 15-hydroxyeicosa-5,8,11,13-tetraenoic acid | Lognormal | -0.002 | (-0.153, 0.150) | 0.983 | 0.993 | 0.029 | (-0.161, 0.141) |
| 9(10)-epoxy-12,15-octadecadienoic acid | Gamma | -0.003 | (-0.502, 0.499) | 0.989 | 0.994 | 0.037 | (-0.387, 0.361) |

**Supplementary Table S2.** Regression and Bayesian estimates comparing all ME/CFS vs. all controls.

| Metabolite | Regression Model | ME/CFS vs. Controls | | | | | |
|---|---|---|---|---|---|---|---|
| | | Estimated Coefficient | 95% CI | p-value | FDR | BayesFactor | 95% HDI |

ME/CFS, myalgic encephalomyelitis/chronic fatigue syndrome; CI, confidence interval; FDR, false discovery rate adjusted p-value; HD,: highest density credible intervals. Regression models were adjusted for age, sex, race/ethnicity, geographic/clinical site, season of sampling, body mass index, and self-reported irritable bowel syndrome. In PM, BA, and CL panels, estimated coefficients are interpreted as the differences in the mean values of log-log transformation of metabolite levels between cases and controls. In OL panel for lognormal regression, estimated coefficients are interpreted as the mean differences of log transformation of metabolite levels between two groups; for Gamma regression, estimated coefficients are interpreted as the log of fold change between two groups. Estimations in **bold** are significant. Criteria for significance: 1) FDR adjusted p-value from the regression model < 0.15, 2) BayesFactor >3, and 3) 95% highest density credible intervals not covering 0.

**Supplementary Table S3.** ChemRICH enrichment analysis comparing all ME/CFS vs. all controls

| Cluster name | Cluster size | p-values | Key compound | Altered | Increased | Decreased | Altered Ratio |
|---|---|---|---|---|---|---|---|
| plasmalogens | 24 | 4.7E-10 | PE (p-36:2)/PE (o-36:3) - ESI(+) | 16 | 0 | 12 | 0.7 |
| unsaturated phosphatidylcholines | 90 | 0.000000072 | PC (36:2) | 26 | 0 | 19 | 0.3 |
| sphingomyelins | 49 | 0.000047 | SM (d39:1) - ESI(-) | 18 | 0 | 14 | 0.4 |
| HEPE | 5 | 0.0001 | 12-Hydroxy-5,8,10,14,17-eicosapentaenoic acid | 4 | 2 | 0 | 0.8 |
| unsaturated phospholipid ethers | 42 | 0.00016 | PC (p-34:2)/PC (o-34:3) - ESI(+) | 17 | 0 | 12 | 0.4 |
| unsaturated ceramides | 21 | 0.00017 | Ceramide (d42:1) - ESI (+) | 9 | 0 | 6 | 0.4 |
| saturated lysophospholipids | 10 | 0.00075 | LPC (14:0) - ESI(+) | 5 | 0 | 5 | 0.5 |
| carnitines | 27 | 0.00089 | 2-Methylbutyryl-L-carnitine | 8 | 0 | 3 | 0.3 |
| exposome_food | 14 | 0.0041 | Caffeine | 7 | 1 | 5 | 0.5 |
| unsaturated lysophosphatidylcholines | 20 | 0.0044 | LPC (18:2) - ESI(-) | 9 | 0 | 5 | 0.4 |
| unsaturated lysophosphoethanolamines | 6 | 0.0051 | LPE (18:2) - ESI(+) | 3 | 0 | 3 | 0.5 |
| saturated TGs | 8 | 0.0052 | TG (53:0) | 3 | 0 | 2 | 0.4 |
| unsaturated long chain TGs | 41 | 0.011 | TG (56:6) | 10 | 7 | 1 | 0.2 |
| dicarboxylic acids | 9 | 0.013 | succinic acid | 5 | 2 | 0 | 0.6 |
| prostaglandins | 9 | 0.031 | Prostaglandin F2a | 2 | 0 | 2 | 0.2 |
| EpODE | 7 | 0.035 | 11,12-Epoxyeicosa-5,8,14-trienoic acid | 3 | 1 | 1 | 0.4 |
| unsaturated phosphatidylethanolamines | 13 | 0.063 | PE (36:2) | 1 | 0 | 1 | 0.08 |
| indoles | 8 | 0.11 | Kynurenine | 1 | 0 | 0 | 0.1 |
| amino acids, basic | 8 | 0.13 | Ornithine | 2 | 0 | 0 | 0.2 |
| OH-FA 22_6_1 | 3 | 0.13 | Resolvin D1 | 1 | 0 | 1 | 0.3 |
| cholesterol esters | 13 | 0.16 | CE (18:2) | 3 | 0 | 3 | 0.2 |
| saturated lysophosphoethanolamines | 4 | 0.25 | LPE (18:0) | 1 | 0 | 1 | 0.2 |
| unsaturated glucyosylceramides | 12 | 0.26 | GlcCeramide (d42:2) - ESI(+) | 2 | 0 | 0 | 0.2 |
| amino acids | 24 | 0.36 | 2,6-Diaminopimelic acid | 1 | 1 | 0 | 0.04 |
| amino acids, branched-chain | 3 | 0.49 | Leucine | 1 | 0 | 1 | 0.3 |
| unsaturated FFA | 15 | 0.57 | FA (16:1) (palmitoleic acid) | 2 | 1 | 0 | 0.1 |
| DiHETE | 8 | 0.57 | 17,18-dihydroxyeicosa-5,8,11,14-tetraenoic acid | 2 | 0 | 0 | 0.2 |
| sugar alcohols | 7 | 0.58 | Mannitol | 1 | 0 | 0 | 0.1 |
| drugs | 85 | 0.58 | Acetominophen | 10 | 5 | 0 | 0.1 |
| adenosine | 4 | 0.59 | 7-Methylguanosine | 1 | 0 | 1 | 0.2 |
| dipeptides | 16 | 0.65 | H-Pro-Hyp-OH | 2 | 1 | 0 | 0.1 |
| saturated FFA | 18 | 0.7 | Arachidic acid | 1 | 1 | 0 | 0.06 |
| saturated phosphatidylcholines | 9 | 0.73 | PC (34:0) - ESI(-) | 1 | 0 | 0 | 0.1 |
| diglycerides | 14 | 0.76 | DG (38:6) | 1 | 0 | 0 | 0.07 |
| amino acids, aromatic | 7 | 1 | Tyrosine | 0 | 0 | 0 | 0 |
| amino acids, cyclic | 6 | 1 | Histidine | 0 | 0 | 0 | 0 |
| amino alcohols | 3 | 1 | 3-Amino-1-propanol | 0 | 0 | 0 | 0 |
| benzene derivatives | 6 | 1 | Tri-2-ethylhexyl trimellitate | 0 | 0 | 0 | 0 |
| DiHETrE | 5 | 1 | 19,20-dihydroxydocosa-4,7,10,13,16-pentaenoic acid | 0 | 0 | 0 | 0 |
| DiHODE | 3 | 1 | 15,16-dihydroxyoctadeca-9,12-dienoic acid | 0 | 0 | 0 | 0 |
| DiHOME | 4 | 1 | 9S,10R-dihydroxy-stearic acid | 0 | 0 | 0 | 0 |

**Supplementary Table S3.** ChemRICH enrichment analysis comparing all ME/CFS vs. all controls

| Cluster name | Cluster size | p-values | Key compound | Altered | Increased | Decreased | Altered Ratio |
|---|---|---|---|---|---|---|---|
| EpETrE | 7 | 1 | 15(16)-epoxy-9,12-octadecadienoic acid | 0 | 0 | 0 | 0 |
| ethanolamines | 3 | 1 | Heptadecasphing-4-enine | 0 | 0 | 0 | 0 |
| guanidines | 3 | 1 | Guanidine | 0 | 0 | 0 | 0 |
| HETE | 7 | 1 | 8-hydroxyeicosa-5,9,11,14-tetraenoic acid | 0 | 0 | 0 | 0 |
| hexoses | 10 | 1 | Levoglucosan | 0 | 0 | 0 | 0 |
| histidine | 4 | 1 | Ergothioneine | 0 | 0 | 0 | 0 |
| HODE | 3 | 1 | 9-Hydroxylinoleic acid | 0 | 0 | 0 | 0 |
| hydroxybutyrates | 7 | 1 | Threonic acid | 0 | 0 | 0 | 0 |
| imidazoles | 3 | 1 | Creatinine | 0 | 0 | 0 | 0 |
| lipids | 3 | 1 | 1-Monostearin | 0 | 0 | 0 | 0 |
| monounsaturated long chain TGs | 16 | 1 | TG (54:1) | 0 | 0 | 0 | 0 |
| monounsaturated TGs | 18 | 1 | TG (46:2) | 0 | 0 | 0 | 0 |
| nitroFA | 3 | 1 | 10-nitrooleic acid | 0 | 0 | 0 | 0 |
| oxo-ODE | 5 | 1 | 5-ketoeicosa-6,8,11,14-tetraenoic acid | 0 | 0 | 0 | 0 |
| pyridines | 3 | 1 | 3-Hydroxypyridine | 0 | 0 | 0 | 0 |
| saturated ceramides | 4 | 1 | Ceramide (d42:0) | 0 | 0 | 0 | 0 |
| saturated sphingomyelins | 10 | 1 | SM (d36:0) - ESI(-) | 0 | 0 | 0 | 0 |
| sugar acids | 3 | 1 | Gluconic acid | 0 | 0 | 0 | 0 |
| unsaturated TGs | 27 | 1 | TG (48:4) A | 0 | 0 | 0 | 0 |
| xanthines | 3 | 1 | 3-Methylxanthine | 0 | 0 | 0 | 0 |

ME/CFS, myalgic encephalomyelitis/chronic fatigue syndrome.

**Supplementary Table S4.** Compound-level ChemRICH enrichment analysis comparing all ME/CFS vs. all controls.

| Compound name | Effect size | p-values | Edirection |
|---|---|---|---|
| **Plasmalogens** | | | |
| **PE (p-36:2)/PE (o-36:3) - ESI(+)** | **-0.028** | **0.00035** | **down** |
| **PE (p-34:2)/PE (o-34:3)** | **-0.036** | **0.0028** | **down** |
| **PE (p-36:2)/PE (o-36:3) - ESI(-)** | **-0.028** | **0.0056** | **down** |
| **PE (p-36:4)/PE (o-36:5) - ESI(-)** | **-0.022** | **0.0098** | **down** |
| **PE (p-38:5)/PE (o-38:6) - ESI(-)** | **-0.020** | **0.014** | **down** |
| **PE (p-38:4)/PE (o-38:5)** | **-0.015** | **0.022** | **down** |
| **PE (p-36:4)/PE (o-36:5) - ESI(+)** | **-0.016** | **0.023** | **down** |
| **PE (p-36:1)/PE (o-36:2) - ESI(+)** | **-0.020** | **0.026** | **down** |
| **PE (p-36:1)/PE (o-36:2) - ESI(-)** | **-0.028** | **0.027** | **down** |
| **PE (p-38:5)/PE (o-38:6) - ESI(+)** | **-0.013** | **0.029** | **down** |
| **PC (o-34:0)** | **-0.013** | **0.047** | **down** |
| **PE (p-40:4)/PE (o-40:5) A** | **-0.022** | **0.049** | **down** |
| PE (p-34:1)/PE (o-34:2) - ESI(+) | -0.014 | 0.061 | down |
| PE (p-40:4)/PE (o-40:5) B | -0.012 | 0.092 | down |
| PE (p-34:1)/PE (o-34:2) - ESI(-) | -0.014 | 0.096 | down |
| LPC (p-16:0)/LPC (o-16:1) | -0.010 | 0.097 | down |
| PE (p-38:6)/PE (o-38:7) | -0.015 | 0.11 | no change |
| PE (p-38:3) or PE (o-38:4) | -0.021 | 0.12 | no change |
| PE (p-40:4)/PE (o-40:5) | -0.011 | 0.21 | no change |
| PE(p-40:5)/PE(o-40:6) | -0.004 | 0.38 | no change |
| PC (o-32:0) - ESI(-) | -0.005 | 0.45 | no change |
| PC (o-32:0) - ESI(+) | -0.003 | 0.46 | no change |
| PE (p-36:5)/PE (o-36:6) | -0.013 | 0.63 | no change |
| LPC (p-18:0)/LPC (o-18:1) | -0.003 | 0.71 | no change |
| **Unsaturated Phosphatidylcholines** | | | |
| **PC (36:2)** | **-0.007** | **0.00028** | **down** |
| **PC (36:4) A - ESI(+)** | **-0.018** | **0.00039** | **down** |
| **PC (36:4) A - ESI(-)** | **-0.019** | **0.0014** | **down** |
| **PC (32:2) - ESI(-)** | **-0.027** | **0.002** | **down** |
| **PC (34:2) - ESI(+)** | **-0.005** | **0.0028** | **down** |
| **PC 34:4e** | **-0.021** | **0.0038** | **down** |
| **PC (34:2) - ESI(-)** | **-0.009** | **0.0065** | **down** |
| **PC (34:4) - ESI(-)** | **-0.032** | **0.0084** | **down** |
| **PC (34:3)** | **-0.016** | **0.013** | **down** |
| **PC (33:2) - ESI(-)** | **-0.019** | **0.019** | **down** |
| **PC(32:2)** | **-0.017** | **0.019** | **down** |
| **PC (34:3) B** | **-0.011** | **0.019** | **down** |
| **PC (36:5) A** | **-0.040** | **0.02** | **down** |
| **PC (34:3) C** | **-0.015** | **0.022** | **down** |
| **PC (36:5) A** | **-0.020** | **0.026** | **down** |
| **PC (36:4) B - ESI(-)** | **-0.008** | **0.03** | **down** |
| **PC (38:4) A - ESI(-)** | **-0.010** | **0.03** | **down** |
| **PC (40:8) - ESI(-)** | **-0.015** | **0.04** | **down** |
| **PC (38:6) A - ESI(+)** | **-0.008** | **0.041** | **down** |
| **PC (34:4) - ESI(+)** | **-0.016** | **0.05** | **down** |
| PC (36:3) A - ESI(+) | -0.007 | 0.053 | down |
| PC (38:3) - ESI(-) | -0.011 | 0.054 | down |
| PC (37:5) | 0.016 | 0.058 | up |
| PC (40:5) A - ESI(+) | 0.007 | 0.084 | up |
| PC (33:2) - ESI(+) | -0.008 | 0.085 | down |

**Supplementary Table S4.** Compound-level ChemRICH enrichment analysis comparing all ME/CFS vs. all controls.

| Compound name | Effect size | p-values | Edirection |
|---|---|---|---|
| PC (36:3) B - ESI(-) | -0.008 | 0.085 | down |
| PC (36:3) A - ESI(-) | -0.008 | 0.1 | down |
| PC (38:6) B - ESI(+) | 0.005 | 0.1 | up |
| PC (40:6) B | 0.008 | 0.1 | up |
| PC (36:5) D | 0.014 | 0.11 | no change |
| PC (37:2) - ESI(+) | -0.007 | 0.11 | no change |
| PC (35:1) - ESI(+) | 0.005 | 0.12 | no change |
| PC (37:6) | 0.013 | 0.12 | no change |
| PC (40:5) B - ESI(-) | -0.018 | 0.13 | no change |
| PC (40:7) A - ESI(+) | -0.008 | 0.15 | no change |
| PC (38:2) | -0.009 | 0.17 | no change |
| PC (35:3) | -0.005 | 0.17 | no change |
| PC (42:10) | 0.014 | 0.17 | no change |
| PC (39:6) | 0.010 | 0.19 | no change |
| PC (36:3) B - ESI(+) | -0.005 | 0.19 | no change |
| PC (38:5) B - ESI(+) | 0.007 | 0.19 | no change |
| PC (40:4) - ESI(-) | -0.012 | 0.2 | no change |
| PC (34:3) A | -0.006 | 0.2 | no change |
| PC (40:7) B - ESI(+) | 0.006 | 0.21 | no change |
| PC (37:2) - ESI(-) | -0.013 | 0.22 | no change |
| PC (38:5) A | 0.004 | 0.22 | no change |
| PC (40:5) B - ESI(+) | -0.008 | 0.22 | no change |
| PC (35:2) | -0.006 | 0.26 | no change |
| PC (38:3) - ESI(+) | -0.005 | 0.27 | no change |
| PC (38:7) | 0.008 | 0.31 | no change |
| PC 40:5e | -0.004 | 0.33 | no change |
| PC (35:2) A | 0.004 | 0.4 | no change |
| PC (36:1) - ESI(-) | -0.004 | 0.41 | no change |
| PC (36:5) C | 0.008 | 0.42 | no change |
| PC (34:1) | 0.001 | 0.45 | no change |
| PC (37:4) - ESI(-) | -0.005 | 0.46 | no change |
| PC (38:4) B - ESI(+) | -0.004 | 0.46 | no change |
| PC (33:1) - ESI(+) | 0.003 | 0.48 | no change |
| PC (32:1) - ESI(-) | -0.006 | 0.49 | no change |
| PC (37:3) | -0.004 | 0.52 | no change |
| PC (35:4) - ESI(-) | -0.005 | 0.54 | no change |
| PC (42:5) | 0.004 | 0.54 | no change |
| PC (40:4) - ESI(+) | -0.004 | 0.54 | no change |
| PC (35:2) B | -0.002 | 0.56 | no change |
| PC (39:4) | 0.003 | 0.57 | no change |
| PC (35:4) - ESI(+) | -0.003 | 0.6 | no change |
| PC (36:5) B | 0.005 | 0.64 | no change |
| PC (37:4) - ESI(+) | 0.003 | 0.64 | no change |
| PC (38:5) B - ESI(-) | 0.004 | 0.65 | no change |
| PC (38:4) C - ESI(+) | -0.001 | 0.65 | no change |
| PC (p-42:5)/PC (o-42:6) | -0.006 | 0.66 | no change |
| PC 40:6e | 0.002 | 0.67 | no change |
| PC (40:6) B | 0.003 | 0.69 | no change |
| PC (36:1) - ESI(+) | 0.001 | 0.7 | no change |
| PC (38:5) A - ESI(-) | -0.002 | 0.71 | no change |
| PC (38:4) A - ESI(+) | -0.002 | 0.74 | no change |

**Supplementary Table S4.** Compound-level ChemRICH enrichment analysis comparing all ME/CFS vs. all controls.

| Compound name | Effect size | p-values | Edirection |
|---|---|---|---|
| PC (35:1) - ESI(-) | 0.002 | 0.76 | no change |
| PC (31:1) | -0.005 | 0.76 | no change |
| PC (36:6) | -0.003 | 0.77 | no change |
| PC 38:7e | -0.002 | 0.78 | no change |
| PC (42:6) | 0.003 | 0.83 | no change |
| PC (40:7) - ESI(-) | -0.002 | 0.86 | no change |
| PC (40:8) - ESI(+) | -0.001 | 0.88 | no change |
| PC (36:4) C - ESI(+) | 0.000 | 0.89 | no change |
| PC (40:5) A - ESI(-) | 0.001 | 0.91 | no change |
| PC (36:4) B - ESI(+) | 0.001 | 0.93 | no change |
| PC (32:1) - ESI(+) | 0.000 | 0.95 | no change |
| PC (38:6) - ESI(-) | 0.000 | 0.98 | no change |
| PC (33:1) - ESI(-) | 0.000 | 0.99 | no change |
| PC (40:6) A | 0.000 | 1 | no change |
| **Sphingomyelins** | | | |
| **SM (d39:1) - ESI(-)** | **-0.017** | **0.0038** | **down** |
| **SM (d40:1) - ESI(-)** | **-0.012** | **0.0053** | **down** |
| **SM (d32:1) - ESI(-)** | **-0.015** | **0.0065** | **down** |
| **SM (d40:3)** | **-0.025** | **0.008** | **down** |
| **SM (d33:1) - ESI(-)** | **-0.014** | **0.016** | **down** |
| **SM (d43:1) - ESI(-)** | **-0.033** | **0.017** | **down** |
| **SM (d36:3) - ESI(-)** | **-0.019** | **0.018** | **down** |
| **SM (d40:2) B - ESI(-)** | **-0.011** | **0.019** | **down** |
| **SM (d42:1) - ESI(-)** | **-0.009** | **0.024** | **down** |
| **SM (d34:2) - ESI(-)** | **-0.008** | **0.026** | **down** |
| **SM (d30:1) - ESI(-)** | **-0.029** | **0.029** | **down** |
| **SM (d34:1) - ESI(-)** | **-0.007** | **0.037** | **down** |
| **SM (d40:1) - ESI(+)** | **-0.006** | **0.046** | **down** |
| **SM (d40:2) B - ESI(+)** | **-0.006** | **0.048** | **down** |
| SM (d38:1) | -0.006 | 0.054 | down |
| SM (d38:2) - ESI(-) | -0.011 | 0.055 | down |
| SM (d39:1) - ESI(+) | -0.008 | 0.082 | down |
| SM (d36:1) - ESI(-) | -0.007 | 0.091 | down |
| SM (d42:1) - ESI(+) | -0.006 | 0.1 | down |
| SM (d30:1) - ESI(+) | -0.016 | 0.11 | no change |
| SM (d32:1) - ESI(-) | -0.008 | 0.13 | no change |
| SM (d43:2) - ESI(-) | -0.017 | 0.13 | no change |
| SM (d32:2) - ESI(-) | -0.013 | 0.18 | no change |
| SM (d44:2) | 0.009 | 0.21 | no change |
| SM (d42:3) - ESI(-) | -0.005 | 0.22 | no change |
| SM (d42:2) | 0.004 | 0.23 | no change |
| SM (d41:2) B - ESI(+) | -0.005 | 0.26 | no change |
| SM (d36:2) - ESI(-) | -0.006 | 0.3 | no change |
| SM (d41:1) - ESI(+) | -0.004 | 0.34 | no change |
| SM (d40:2) A - ESI(-) | -0.008 | 0.35 | no change |
| SM (d40:2) A - ESI(+) | -0.005 | 0.35 | no change |
| SM (d41:3) | -0.004 | 0.37 | no change |
| SM (d39:2) | -0.003 | 0.44 | no change |
| SM (d37:1) | -0.007 | 0.5 | no change |
| SM (d36:3) - ESI(+) | -0.004 | 0.53 | no change |
| D-erythro-Sphingosine-1-phosphate | -0.007 | 0.54 | no change |

**Supplementary Table S4.** Compound-level ChemRICH enrichment analysis comparing all ME/CFS vs. all controls.

| Compound name | Effect size | p-values | Edirection |
|---|---|---|---|
| SM (d43:1) - ESI(+) | -0.004 | 0.54 | no change |
| SM (d43:2) - ESI(+) | 0.005 | 0.56 | no change |
| SM (d34:1) - ESI(-) | -0.001 | 0.58 | no change |
| SM (d41:2) - ESI(-) | -0.004 | 0.59 | no change |
| SM (d32:2) - ESI(+) | -0.003 | 0.64 | no change |
| SM (d34:2) - ESI(-) | -0.001 | 0.65 | no change |
| SM (d41:2) A - ESI(+) | 0.002 | 0.72 | no change |
| SM (d42:3) - ESI(+) | 0.001 | 0.74 | no change |
| SM (d38:2) - ESI(+) | -0.001 | 0.85 | no change |
| SM (d41:1) - ESI(-) | 0.007 | 0.88 | no change |
| SM (d33:1) - ESI(+) | -0.001 | 0.89 | no change |
| SM (d36:2) - ESI(-) | 0.000 | 0.96 | no change |
| SM (d36:1) - ESI(-) | 0.000 | 0.99 | no change |
| **HEPE** | | | |
| **12-Hydroxy-5,8,10,14,17-eicosapentaenoic acid** | **0.440** | **0.015** | **up** |
| **5-Hydroxy-6,8,11,14,17-eicosapentaenoic acid** | **0.242** | **0.035** | **up** |
| 15-hydroxyeicosa-5,8,11,13,17-pentaenoic acid | 0.321 | 0.058 | up |
| 9-Hydroxy-5,7,11,14,17-icosapentaenoic acid | 0.222 | 0.069 | up |
| 4-hydroxydocosa-5,7,10,13,16,19-hexaenoic acid | 0.213 | 0.16 | no change |
| **Unsaturated Phospholipid Ethers** | | | |
| **PC (p-34:2)/PC (o-34:3) - ESI(+)** | **-0.018** | **0.000084** | **down** |
| **PC (p-34:1)/PC (o-34:2)** | **-0.021** | **0.00015** | **down** |
| **PC (p-34:2)/PC (o-34:3) - ESI(-)** | **-0.020** | **0.00069** | **down** |
| **PC (p-36:1)/PC (o-36:2)** | **-0.054** | **0.00075** | **down** |
| **PC (p-36:4)/PC (o-36:5) - ESI(-)** | **-0.020** | **0.0017** | **down** |
| **PC (p-34:1)/PC (o-34:2) A** | **-0.026** | **0.0024** | **down** |
| **PC (p-36:3)/PC (o-36:4) - ESI(-)** | **-0.015** | **0.0081** | **down** |
| **PC(p-36:2)/PC(o-36:3)** | **-0.016** | **0.013** | **down** |
| **PC (p-38:3) or PC (o-38:4)** | **-0.015** | **0.014** | **down** |
| **PC (p-38:4) or PC (o-38:5) A** | **-0.011** | **0.019** | **down** |
| **PC(p-36:4)/PC(o-36:5)** | **-0.009** | **0.031** | **down** |
| **PC (p-36:3)/PC (o-36:4) - ESI(+)** | **-0.009** | **0.031** | **down** |
| PC (p-38:4)/PC (o-38:5) B | -0.010 | 0.056 | down |
| PC (p-32:1)/PC (o-32:2) | -0.009 | 0.06 | down |
| PC (p-38:3)/PC (o-38:4) A - ESI(+) | -0.009 | 0.061 | down |
| PC (p-38:3)/PC (o-38:4) B - ESI(+) | -0.010 | 0.069 | down |
| PC (p-38:4)/PC (o-38:5) B | -0.016 | 0.082 | down |
| PC (p-40:1)/PC (o-40:2) | 0.014 | 0.13 | no change |
| PC (p-40:3)/PC (o-40:4) | -0.012 | 0.16 | no change |
| PC (p-38:4)/PC (o-38:5) A | -0.004 | 0.22 | no change |
| PC (p-38:5)/PC (o-38:6) B | -0.005 | 0.22 | no change |
| PC (p-32:0)/PC (o-32:1) - ESI(-) | -0.010 | 0.22 | no change |
| PC (p-32:0)/PC (o-32:1) - ESI(+) | -0.004 | 0.26 | no change |
| PC (p-44:4)/PC (o-44:5) - ESI(-) | -0.007 | 0.33 | no change |
| PC (p-42:3)/PC (o-42:4) | -0.006 | 0.34 | no change |
| PC (p-36:1)/PC (o-36:2) B | -0.007 | 0.38 | no change |
| PC (p-42:5)/PC (o-42:6) A | -0.005 | 0.39 | no change |
| PC (p-42:4)/PC (o-42:5) - ESI(-) | -0.005 | 0.57 | no change |
| PC (p-40:6)/PC (o-40:7) A | 0.003 | 0.58 | no change |
| PC (p-34:1)/PC (o-34:2) B | -0.005 | 0.6 | no change |
| PC (p-44:4)/PC (o-44:5) - ESI(+) | 0.003 | 0.61 | no change |

**Supplementary Table S4.** Compound-level ChemRICH enrichment analysis comparing all ME/CFS vs. all controls.

| Compound name | Effect size | p-values | Edirection |
|---|---|---|---|
| PC (p-38:6)/PC (o-38:7) | -0.003 | 0.65 | no change |
| PC (p-44:5)/PC (o-44:6) | 0.003 | 0.67 | no change |
| PC (p-40:7)/PC (o-40:8) | -0.008 | 0.68 | no change |
| PC (p-40:4)/PC (o-40:5) - ESI(+) | -0.001 | 0.87 | no change |
| PC (p-40:4)/PC (o-40:5) - ESI(-) | 0.001 | 0.87 | no change |
| PC (p-36:5)/PC (o-36:6) | 0.001 | 0.89 | no change |
| PC (p-40:6)/PC (o-40:7) B | -0.001 | 0.89 | no change |
| PC (p-38:5)/PC (o-38:6) A | -0.001 | 0.91 | no change |
| PC (p-38:5)/PC (o-38:6) | -0.001 | 0.94 | no change |
| PC (p-42:4)/PC (o-42:5) - ESI(+) | 0.000 | 0.96 | no change |
| PC (p-42:5)/PC (o-42:6) B | 0.000 | 0.96 | no change |
| **Unsaturated Ceramides** | | | |
| **Ceramide (d42:1) - ESI (+)** | **-0.009** | **0.0056** | **down** |
| **Ceramide (d41:1) - ESI(-)** | **-0.014** | **0.01** | **down** |
| **Ceramide (d42:2) B - ESI (+)** | **-0.011** | **0.011** | **down** |
| **Ceramide (d42:1) - ESI(-)** | **-0.011** | **0.011** | **down** |
| **Ceramide (d40:1)** | **-0.010** | **0.014** | **down** |
| **Ceramide (d39:1)** | **-0.020** | **0.018** | **down** |
| Ceramide (d43:1) | -0.032 | 0.078 | down |
| Ceramide (d41:1) - ESI (+) | -0.008 | 0.089 | down |
| Ceramide (d38:1) - ESI(+) | -0.008 | 0.096 | down |
| Ceramide (d33:1) | -0.010 | 0.12 | no change |
| Ceramide (d38:1) - ESI(-) | -0.010 | 0.12 | no change |
| Ceramide (d32:1) | -0.011 | 0.13 | no change |
| Ceramide (d42:2) B - ESI(-) | -0.011 | 0.2 | no change |
| Ceramide (d34:1) - ESI(-) | -0.005 | 0.24 | no change |
| Ceramide (d44:1) | -0.014 | 0.26 | no change |
| Ceramide (d42:2) A - ESI(-) | -0.004 | 0.39 | no change |
| Ceramide (d42:2) A - ESI (+) | 0.003 | 0.49 | no change |
| Ceramide (d34:1) - ESI(+) | 0.002 | 0.59 | no change |
| Ceramide (d36:1) - ESI(-) | -0.005 | 0.62 | no change |
| Ceramide (d36:1) - ESI(+) | 0.002 | 0.67 | no change |
| Ceramide (d34:2) | -0.001 | 0.9 | no change |
| **Saturated Lysophospholipids** | | | |
| **LPC (14:0) - ESI(+)** | **-0.021** | **0.0081** | **down** |
| **LPC (18:0) A - ESI(-)** | **-0.018** | **0.014** | **down** |
| **LPC (16:0) - ESI(-)** | **-0.010** | **0.017** | **down** |
| **LPC (18:0) B - ESI(-)** | **-0.012** | **0.019** | **down** |
| **LPC (14:0) - ESI(-)** | **-0.026** | **0.046** | **down** |
| LPC (16:0) - ESI(+) | -0.003 | 0.19 | no change |
| LPC (o-16:0) | -0.008 | 0.2 | no change |
| LPC (18:0) - ESI(+) | -0.006 | 0.2 | no change |
| LPC (15:0) | -0.004 | 0.55 | no change |
| LPC (20:0) | -0.004 | 0.65 | no change |
| **Carnitines** | | | |
| **2-Methylbutyryl-L-carnitine** | **-0.035** | **0.0023** | **down** |
| **3-Methylglutarylcarnitine** | **-0.033** | **0.018** | **down** |
| **AC (10:1)** | **-0.020** | **0.042** | **down** |
| Octanoylcarnitine | -0.017 | 0.07 | down |
| AC (14:2) | -0.020 | 0.085 | down |
| 3-Dehydrocarnitine | -0.013 | 0.089 | down |

**Supplementary Table S4.** Compound-level ChemRICH enrichment analysis comparing all ME/CFS vs. all controls.

| Compound name | Effect size | p-values | Edirection |
|---|---|---|---|
| AC (18:0) | -0.010 | 0.092 | down |
| Lauroyl-L-carnitine | -0.033 | 0.092 | down |
| AC (18:2) | -0.012 | 0.1 | down |
| (R)-Butyrylcarnitine | -0.017 | 0.12 | no change |
| AC (12:0) | -0.016 | 0.12 | no change |
| AC (12:1) | -0.015 | 0.13 | no change |
| AC (10:0) | -0.020 | 0.16 | no change |
| Linoleoylcarnitine | -0.024 | 0.16 | no change |
| Hexanoyl-L-carnitine | -0.011 | 0.28 | no change |
| AC (8:0) | -0.014 | 0.32 | no change |
| Propionylcarnitine | -0.004 | 0.32 | no change |
| Oleoyl-L-carnitine | -0.017 | 0.35 | no change |
| (3-Carboxypropyl)trimethylammonium cation | -0.005 | 0.37 | no change |
| AC (14:1) | -0.010 | 0.38 | no change |
| AC (16:0) | -0.003 | 0.58 | no change |
| AC (18:1) | -0.003 | 0.58 | no change |
| 3-Hydroxyoleylcarnitine | 0.018 | 0.63 | no change |
| 3-Hydroxybutyrylcarnitine | -0.006 | 0.66 | no change |
| (2R)-3-Hydroxyisovaleroylcarnitine | -0.004 | 0.68 | no change |
| Acetyl-DL-carnitine | 0.002 | 0.82 | no change |
| Decanoyl-L-carnitine | -0.003 | 0.85 | no change |
| **Food Exposome** | | | |
| **Caffeine** | **-0.084** | **0.0031** | **down** |
| **Choline** | **-0.009** | **0.0035** | **down** |
| **Theobromine** | **-0.064** | **0.0059** | **down** |
| **Piperine** | **-0.067** | **0.012** | **down** |
| **Trigonelline** | **-0.044** | **0.02** | **down** |
| **4,5,7-Trihydroxyisoflavone** | **0.083** | **0.047** | **up** |
| Betaine | -0.006 | 0.075 | down |
| Stachydrine | -0.025 | 0.17 | no change |
| Coniferylaldehyde | 0.008 | 0.51 | no change |
| Theanine; | 0.009 | 0.58 | no change |
| Methylgallate | 0.018 | 0.66 | no change |
| 6-Hydroxyflavone | 0.008 | 0.68 | no change |
| Betaine aldehyde cation | 0.008 | 0.75 | no change |
| Scopoletin | -0.012 | 0.76 | no change |
| **Unsaturated Lysophosphatidylcholines** | | | |
| **LPC (18:2) - ESI(-)** | **-0.019** | **0.004** | **down** |
| **LPC (18:3)** | **-0.025** | **0.006** | **down** |
| **LPC (18:2) - ESI(+)** | **-0.014** | **0.0067** | **down** |
| **LPC (20:3) - ESI(-)** | **-0.023** | **0.017** | **down** |
| **LPC (20:3) - ESI(+)** | **-0.012** | **0.045** | **down** |
| LPC (16:1) - ESI(-) | -0.016 | 0.057 | down |
| LPC (20:2) - ESI(-) | -0.027 | 0.077 | down |
| LPC (20:2) - ESI(+) | -0.012 | 0.081 | down |
| LPC (18:1) - ESI(-) | -0.010 | 0.082 | down |
| LPC (20:4) | -0.007 | 0.19 | no change |
| LPC (22:4) | -0.015 | 0.23 | no change |
| LPC (16:1) - ESI(+) | -0.005 | 0.33 | no change |
| LPC (20:5) | 0.012 | 0.36 | no change |
| LPC (20:1) - ESI(+) | -0.005 | 0.53 | no change |

**Supplementary Table S4.** Compound-level ChemRICH enrichment analysis comparing all ME/CFS vs. all controls.

| Compound name | Effect size | p-values | Edirection |
|---|---|---|---|
| LPC (20:1) - ESI(-) | -0.008 | 0.55 | no change |
| LPC (18:1) - ESI(+) | -0.003 | 0.55 | no change |
| LPC (17:1) | 0.004 | 0.61 | no change |
| LPC (22:5) - ESI(+) | 0.005 | 0.63 | no change |
| LPC (22:5) - ESI(-) | -0.004 | 0.8 | no change |
| LPC (22:6) | 0.001 | 0.88 | no change |
| **Unsaturated Lysophosphoethanolamines** | | | |
| **LPE (18:2) - ESI(+)** | **-0.022** | **0.01** | **down** |
| **LPE (18:2) - ESI(-)** | **-0.027** | **0.011** | **down** |
| **LPE (20:4) - ESI(-)** | **-0.022** | **0.011** | **down** |
| LPE (22:6) | -0.012 | 0.17 | no change |
| LPE (20:4) - ESI(+) | -0.011 | 0.17 | no change |
| 1-Oleoyl-sn-glycero-3-phosphoethanolamine | -0.015 | 0.38 | no change |
| **Saturated Triglycerides** | | | |
| **TG (53:0)** | **-0.018** | **0.018** | **down** |
| **TG (50:0)** | **-0.024** | **0.021** | **down** |
| TG (48:0) | -0.018 | 0.08 | down |
| TG (46:0) | -0.027 | 0.11 | no change |
| TG (42:0) | -0.022 | 0.2 | no change |
| TG (44:0) | -0.021 | 0.21 | no change |
| TG (40:0) | -0.014 | 0.37 | no change |
| TG (49:0) | -0.005 | 0.6 | no change |
| **Unsaturated Long-chain Triglycerides** | | | |
| **TG (56:6)** | **0.010** | **0.01** | **up** |
| **TG (55:6)** | **0.021** | **0.012** | **up** |
| **TG (58:8)** | **0.019** | **0.032** | **up** |
| **TG (62:4)** | **-0.029** | **0.034** | **down** |
| **TG (56:7) B** | **0.018** | **0.034** | **up** |
| **TG (54:7) B** | **0.017** | **0.035** | **up** |
| **TG (58:9)** | **0.017** | **0.042** | **up** |
| **TG (54:6) C** | **0.011** | **0.043** | **up** |
| TG (60:11) | 0.025 | 0.072 | up |
| TG (56:8) B | 0.017 | 0.074 | up |
| TG (56:5) B | 0.009 | 0.12 | no change |
| TG (56:8) A | 0.010 | 0.13 | no change |
| TG (54:5) B | 0.007 | 0.14 | no change |
| TG (56:5) A | 0.009 | 0.17 | no change |
| TG (58:6) | 0.007 | 0.17 | no change |
| TG (56:5) C | 0.007 | 0.18 | no change |
| TG (60:12) | 0.025 | 0.18 | no change |
| TG (60:6) | 0.013 | 0.22 | no change |
| TG (54:6) B | -0.021 | 0.24 | no change |
| TG (56:9) | 0.011 | 0.26 | no change |
| TG (54:7) A | -0.010 | 0.33 | no change |
| TG (64:3) | -0.010 | 0.37 | no change |
| TG (58:10) | 0.006 | 0.48 | no change |
| TG (58:4) | 0.008 | 0.48 | no change |
| TG (56:7) A | 0.014 | 0.49 | no change |
| TG (56:3) | 0.006 | 0.5 | no change |
| TG (54:3) | 0.003 | 0.52 | no change |
| TG (58:5) | -0.005 | 0.61 | no change |

**Supplementary Table S4.** Compound-level ChemRICH enrichment analysis comparing all ME/CFS vs. all controls.

| Compound name | Effect size | p-values | Edirection |
|---|---|---|---|
| TG (58:3) | 0.006 | 0.64 | no change |
| TG (54:8) | 0.006 | 0.67 | no change |
| TG (54:4) | 0.002 | 0.71 | no change |
| TG (59:3) | 0.004 | 0.71 | no change |
| TG (58:7)/TG (18:1/18:1/22:5) | 0.002 | 0.72 | no change |
| TG (56:10) | 0.005 | 0.74 | no change |
| TG (54:9) | 0.009 | 0.75 | no change |
| TG (54:6) A | -0.002 | 0.8 | no change |
| TG (62:3) | -0.003 | 0.88 | no change |
| TG (56:4) | 0.001 | 0.91 | no change |
| TG (60:3) | 0.001 | 0.93 | no change |
| TG (54:5) A | 0.000 | 0.97 | no change |
| TG (60:4) | 0.000 | 0.99 | no change |
| Dicarboxylic Acids | | | |
| **succinic acid** | **0.022** | **0.007** | **up** |
| **alpha ketoglutarate** | **0.016** | **0.048** | **up** |
| maleic acid | 0.026 | 0.065 | up |
| aminomalonate | 0.026 | 0.067 | up |
| glutaric acid | 0.017 | 0.088 | up |
| malic acid | 0.009 | 0.38 | no change |
| fumaric acid | -0.003 | 0.74 | no change |
| adipic acid | -0.002 | 0.79 | no change |
| oxalic acid | -0.003 | 0.91 | no change |
| Prostaglandins | | | |
| **Prostaglandin F2a** | **-0.293** | **0.025** | **down** |
| **9,11,15-trihydroxy-5,13,1Z-prostatrienoic acid** | **-0.572** | **0.042** | **down** |
| 15-Deoxy-delta-12,14-Prostaglandin J2 | -0.125 | 0.39 | no change |
| Prostaglandin D2 | -0.395 | 0.13 | no change |
| 15-Keto-prostaglandin E2 | -0.156 | 0.45 | no change |
| Prostaglandin E1 | 0.161 | 0.37 | no change |
| Prostaglandin E2 | 0.066 | 0.82 | no change |
| Prostaglandin E3 | 0.270 | 0.37 | no change |
| 6-Ketoprostaglandin F1 alpha | 0.169 | 0.4 | no change |
| EpODE | | | |
| **11,12-Epoxyeicosa-5,8,14-trienoic acid** | **0.304** | **0.021** | **up** |
| **8,9-Epoxyeicosa-5,11,14-trienoic acid** | **-0.259** | **0.04** | **down** |
| 17,18-Epoxy-5,8,11,14-eicosatetraenoic acid | -0.424 | 0.055 | down |
| 11-Hydroxy-14,15-epoxyeicosatrienoic acid | -0.385 | 0.11 | no change |
| 14(15)-epoxy-5,8,11,17-eicosatetraenoic acid | 0.278 | 0.31 | no change |
| Thromboxane B2 | 0.069 | 0.75 | no change |
| 11(12)-epoxy-5,8,14,17- eicosatetraenoic acid | 0.006 | 0.98 | no change |

ME/CFS, myalgic encephalomyelitis/chronic fatigue syndrome. Analytes in bold have p-values < 0.05.

**Supplementary Table S5.** ChemRICH enrichment analysis comparing female ME/CFS vs. female controls

| Cluster name | Cluster size | p-values | Key compound | Altered | Increased | Decreased | Altered Ratio |
|---|---|---|---|---|---|---|---|
| plasmalogens | 24 | 4.7E-10 | PE (p-36:2)/PE (o-36:3) - ESI(+) | 16 | 0 | 12 | 0.7 |
| unsaturated phosphatidylcholines | 90 | 0.000000072 | PC (36:2) | 26 | 0 | 19 | 0.3 |
| HEPE | 5 | 0.00000054 | 5-Hydroxy-6,8,11,14,17-eicosapentaenoic acid | 5 | 4 | 0 | 1 |
| sphingomyelins | 49 | 0.000047 | SM (d39:1) - ESI(-) | 18 | 0 | 14 | 0.4 |
| unsaturated phospholipid ethers | 42 | 0.00016 | PC (p-34:2)/PC (o-34:3) - ESI(+) | 17 | 0 | 12 | 0.4 |
| unsaturated ceramides | 21 | 0.00017 | Ceramide (d42:1) - ESI (+) | 9 | 0 | 6 | 0.4 |
| saturated lysophospholipids | 10 | 0.00075 | LPC (14:0) - ESI(+) | 5 | 0 | 5 | 0.5 |
| carnitines | 27 | 0.00089 | 2-Methylbutyryl-L-carnitine | 8 | 0 | 3 | 0.3 |
| exposome food | 14 | 0.0041 | Caffeine | 7 | 1 | 5 | 0.5 |
| unsaturated lysophosphatidylcholines | 20 | 0.0044 | LPC (18:2) - ESI(-) | 9 | 0 | 5 | 0.4 |
| unsaturated lysophosphoethanolamines | 6 | 0.0051 | LPE (18:2) - ESI(+) | 3 | 0 | 3 | 0.5 |
| saturated TGs | 8 | 0.0052 | TG (53:0) | 3 | 0 | 2 | 0.4 |
| unsaturated long chain TGs | 41 | 0.011 | TG (56:6) | 10 | 7 | 1 | 0.2 |
| dicarboxylic acids | 9 | 0.013 | Succinic acid | 5 | 2 | 0 | 0.6 |
| EpODE | 7 | 0.017 | 17,18-Epoxy-5,8,11,14-eicosatetraenoic acid | 4 | 0 | 0 | 0.6 |
| prostaglandins | 9 | 0.031 | Prostaglandin F2a | 2 | 0 | 2 | 0.2 |
| unsaturated phosphatidylethanolamines | 13 | 0.063 | PE (36:2) | 1 | 0 | 1 | 0.08 |
| indoles | 8 | 0.11 | Kynurenine | 1 | 0 | 0 | 0.1 |
| amino acids, basic | 8 | 0.13 | Ornithine | 2 | 0 | 0 | 0.2 |
| DiHOME | 4 | 0.14 | 9,10-dihydroxyoctadec-12-enoic acid | 1 | 0 | 0 | 0.2 |
| DiHODE | 3 | 0.16 | 15,16-dihydroxyoctadeca-9,12-dienoic acid | 1 | 0 | 1 | 0.3 |
| cholesterol esters | 13 | 0.16 | CE (18:2) | 3 | 0 | 3 | 0.2 |
| saturated lysophosphoethanolamines | 4 | 0.25 | LPE (18:0) | 1 | 0 | 1 | 0.2 |
| unsaturated glucyosylceramides | 12 | 0.26 | GlcCer (d42:2) - ESI(+) | 2 | 0 | 0 | 0.2 |
| amino acids | 24 | 0.36 | 2,6-Diaminopimelic acid | 1 | 1 | 0 | 0.04 |
| OH-FA_22_6_1 | 3 | 0.42 | Resolvin D1 | 1 | 0 | 1 | 0.3 |
| amino acids, branched-chain | 3 | 0.49 | Leucine | 1 | 0 | 1 | 0.3 |
| DiHETE | 8 | 0.51 | 17,18-dihydroxyeicosa-5,8,11,14-tetraenoic acid | 2 | 0 | 0 | 0.2 |
| unsaturated FFA | 15 | 0.57 | FA (16:1) (palmitoleic acid) | 2 | 1 | 0 | 0.1 |
| sugar alcohols | 7 | 0.58 | Mannitol | 1 | 0 | 0 | 0.1 |
| drugs | 85 | 0.58 | Acetominophen | 10 | 5 | 0 | 0.1 |
| adenosine | 4 | 0.59 | 7-Methylguanosine | 1 | 0 | 1 | 0.2 |
| dipeptides | 16 | 0.65 | H-Pro-Hyp-OH | 2 | 1 | 0 | 0.1 |
| saturated FFA | 18 | 0.7 | Arachidic acid | 1 | 1 | 0 | 0.06 |
| saturated phosphatidylcholines | 9 | 0.73 | PC (34:0) - ESI(-) | 1 | 0 | 0 | 0.1 |
| diglycerides | 14 | 0.76 | DG (38:6) | 1 | 0 | 0 | 0.07 |
| HETE | 7 | 0.93 | 8-hydroxyeicosa-5,9,11,14-tetraenoic acid | 1 | 0 | 0 | 0.1 |
| amino acids, aromatic | 7 | 1 | Tyrosine | 0 | 0 | 0 | 0 |
| amino acids, cyclic | 6 | 1 | Histidine | 0 | 0 | 0 | 0 |
| amino alcohols | 3 | 1 | 3-Amino-1-propanol | 0 | 0 | 0 | 0 |
| benzene derivatives | 6 | 1 | Tri-2-ethylhexyl trimellitate | 0 | 0 | 0 | 0 |
| DiHETrE | 5 | 1 | 8,9-dihydroxyeicosa-5,11,14-trienoic acid | 0 | 0 | 0 | 0 |
| EpETrE | 7 | 1 | 15(16)-epoxy-9,12-octadecadienoic acid | 0 | 0 | 0 | 0 |
| ethanolamines | 3 | 1 | Heptadecasphing-4-enine | 0 | 0 | 0 | 0 |
| guanidines | 3 | 1 | Guanidine | 0 | 0 | 0 | 0 |
| hexoses | 10 | 1 | Levoglucosan | 0 | 0 | 0 | 0 |
| histidine | 4 | 1 | Ergothioneine | 0 | 0 | 0 | 0 |
| HODE | 3 | 1 | 13-hydroxyoctadeca-9,11,15-trienoic acid | 0 | 0 | 0 | 0 |
| hydroxybutyrates | 7 | 1 | Threonic acid | 0 | 0 | 0 | 0 |

**Supplementary Table S5.** ChemRICH enrichment analysis comparing female ME/CFS vs. female controls

| Cluster name | Cluster size | p-values | Key compound | Altered | Increased | Decreased | Altered Ratio |
|---|---|---|---|---|---|---|---|
| imidazoles | 3 | 1 | Creatinine | 0 | 0 | 0 | 0 |
| lipids | 3 | 1 | 1-Monostearin | 0 | 0 | 0 | 0 |
| monounsaturated long chain TGs | 16 | 1 | TG (54:1) | 0 | 0 | 0 | 0 |
| monounsaturated TGs | 18 | 1 | TG (46:2) | 0 | 0 | 0 | 0 |
| nitroFA | 3 | 1 | 10-nitrooleic acid | 0 | 0 | 0 | 0 |
| oxo-ODE | 5 | 1 | 15-ketoeicosa-5,8,11,13-tetraenoic acid | 0 | 0 | 0 | 0 |
| pyridines | 3 | 1 | 3-Hydroxypyridine | 0 | 0 | 0 | 0 |
| saturated ceramides | 4 | 1 | Ceramide (d42:0) | 0 | 0 | 0 | 0 |
| saturated sphingomyelins | 10 | 1 | SM (d36:0) - ESI(-) | 0 | 0 | 0 | 0 |
| sugar acids | 3 | 1 | Gluconic acid | 0 | 0 | 0 | 0 |
| unsaturated TGs | 27 | 1 | TG (48:4) A | 0 | 0 | 0 | 0 |
| xanthines | 3 | 1 | 3-Methylxanthine | 0 | 0 | 0 | 0 |

ME/CFS, myalgic encephalomyelitis/chronic fatigue syndrome.

**Supplementary Table S6.** Regression and Bayesian estimates comparing sex-stratified ME/CFS vs. controls.

| Metabolite | Regression Model | Female ME/CFS vs. Female Controls | | | | | |
|---|---|---|---|---|---|---|---|
| | | Estimated Coefficient | 95% CI | p-value | FDR | BayesFactor | 95% HDI |
| **Primary Metabolites (PM)** | | | | | | | |
| ornithine | Lognormal | -0.027 | (-0.046, -0.008) | 0.005 | 0.180 | 1.549 | (-0.046, -0.008) |
| tyrosine | Lognormal | -0.012 | (-0.022, -0.001) | 0.032 | 0.315 | 0.463 | (-0.022, -0.001) |
| threonine | Lognormal | -0.018 | (-0.035, -0.001) | 0.040 | 0.326 | 0.351 | (-0.035, -0.001) |
| succinic acid | Lognormal | 0.020 | (0.000, 0.041) | 0.050 | 0.373 | 0.302 | (0.000, 0.041) |
| lysine | Lognormal | -0.021 | (-0.043, 0.000) | 0.057 | 0.383 | 0.263 | (-0.043, 0.000) |
| phenylalanine | Lognormal | -0.012 | (-0.025, 0.000) | 0.060 | 0.390 | 0.259 | (-0.025, 0.000) |
| fructose | Lognormal | -0.030 | (-0.061, 0.001) | 0.061 | 0.390 | 0.247 | (-0.063, -0.001) |
| tagatose | Lognormal | -0.018 | (-0.037, 0.001) | 0.061 | 0.392 | 0.217 | (-0.038, 0.000) |
| leucine | Lognormal | -0.011 | (-0.023, 0.001) | 0.063 | 0.399 | 0.254 | (-0.024, 0.000) |
| glutamine | Lognormal | -0.024 | (-0.051, 0.002) | 0.072 | 0.419 | 0.209 | (-0.051, 0.001) |
| methionine | Lognormal | -0.019 | (-0.040, 0.002) | 0.080 | 0.443 | 0.175 | (-0.039, 0.002) |
| mannitol | Lognormal | 0.034 | (-0.004, 0.073) | 0.085 | 0.444 | 0.198 | (-0.003, 0.075) |
| pelargonic acid | Lognormal | 0.014 | (-0.002, 0.029) | 0.089 | 0.451 | 0.163 | (-0.002, 0.029) |
| arachidic acid | Lognormal | 0.019 | (-0.004, 0.042) | 0.109 | 0.492 | 0.145 | (-0.004, 0.042) |
| N-acetylputrescine | Lognormal | -0.014 | (-0.034, 0.005) | 0.146 | 0.535 | 0.107 | (-0.033, 0.006) |
| creatinine | Lognormal | -0.018 | (-0.042, 0.007) | 0.159 | 0.559 | 0.099 | (-0.042, 0.007) |
| urea | Lognormal | -0.007 | (-0.018, 0.003) | 0.177 | 0.591 | 0.105 | (-0.017, 0.003) |
| citrulline | Lognormal | -0.010 | (-0.025, 0.005) | 0.190 | 0.601 | 0.088 | (-0.024, 0.007) |
| erythritol | Lognormal | 0.020 | (-0.010, 0.051) | 0.193 | 0.605 | 0.092 | (-0.008, 0.052) |
| 2-deoxytetronic acid | Lognormal | 0.016 | (-0.009, 0.040) | 0.211 | 0.610 | 0.081 | (-0.007, 0.042) |
| maleimide | Lognormal | 0.014 | (-0.008, 0.037) | 0.214 | 0.610 | 0.081 | (-0.008, 0.036) |
| 2-hydroxybutanoic acid | Lognormal | 0.013 | (-0.009, 0.036) | 0.236 | 0.633 | 0.075 | (-0.009, 0.036) |
| oxoproline | Lognormal | -0.004 | (-0.012, 0.003) | 0.247 | 0.636 | 0.078 | (-0.012, 0.004) |
| 2-ketoisocaproic acid | Lognormal | 0.008 | (-0.005, 0.021) | 0.247 | 0.636 | 0.073 | (-0.005, 0.021) |
| glucose | Lognormal | -0.005 | (-0.013, 0.003) | 0.248 | 0.636 | 0.070 | (-0.012, 0.004) |
| indole-3-propionic acid | Lognormal | -0.017 | (-0.045, 0.012) | 0.254 | 0.637 | 0.074 | (-0.043, 0.013) |
| glutaric acid | Lognormal | 0.014 | (-0.011, 0.039) | 0.273 | 0.650 | 0.074 | (-0.012, 0.038) |
| alpha-ketoglutarate | Lognormal | 0.010 | (-0.008, 0.029) | 0.286 | 0.657 | 0.068 | (-0.009, 0.029) |
| tryptophan | Lognormal | -0.011 | (-0.031, 0.010) | 0.305 | 0.676 | 0.069 | (-0.031, 0.011) |
| conduritol-beta-expoxide | Lognormal | 0.018 | (-0.016, 0.051) | 0.308 | 0.676 | 0.070 | (-0.015, 0.053) |
| sucrose | Lognormal | 0.020 | (-0.018, 0.057) | 0.313 | 0.676 | 0.070 | (-0.018, 0.058) |
| linoleic acid | Lognormal | 0.013 | (-0.013, 0.040) | 0.329 | 0.685 | 0.062 | (-0.012, 0.041) |
| alanine | Lognormal | -0.011 | (-0.033, 0.012) | 0.344 | 0.691 | 0.065 | (-0.033, 0.012) |
| lysine | Lognormal | -0.013 | (-0.041, 0.015) | 0.364 | 0.704 | 0.061 | (-0.043, 0.015) |
| isoleucine | Lognormal | -0.005 | (-0.017, 0.006) | 0.374 | 0.711 | 0.053 | (-0.017, 0.006) |
| pseudo uridine | Lognormal | -0.006 | (-0.019, 0.007) | 0.393 | 0.728 | 0.055 | (-0.019, 0.007) |
| hydroxycarbamate NIST | Lognormal | -0.008 | (-0.031, 0.014) | 0.461 | 0.775 | 0.052 | (-0.032, 0.014) |
| levoglucosan | Lognormal | -0.011 | (-0.040, 0.019) | 0.480 | 0.789 | 0.053 | (-0.039, 0.018) |
| mannose | Lognormal | -0.007 | (-0.027, 0.013) | 0.483 | 0.791 | 0.048 | (-0.027, 0.013) |
| glycerol | Lognormal | 0.004 | (-0.008, 0.017) | 0.499 | 0.806 | 0.041 | (-0.008, 0.018) |
| indole-3-lactate | Lognormal | -0.006 | (-0.023, 0.011) | 0.512 | 0.810 | 0.050 | (-0.023, 0.011) |
| serine | Lognormal | -0.006 | (-0.024, 0.012) | 0.518 | 0.811 | 0.051 | (-0.024, 0.014) |
| beta-alanine | Lognormal | -0.008 | (-0.034, 0.017) | 0.521 | 0.813 | 0.049 | (-0.035, 0.018) |
| quinic acid | Lognormal | -0.014 | (-0.058, 0.030) | 0.527 | 0.813 | 0.041 | (-0.057, 0.029) |
| glycerol-alpha-phosphate | Lognormal | 0.010 | (-0.021, 0.041) | 0.529 | 0.813 | 0.047 | (-0.019, 0.042) |
| aminomalonate | Lognormal | 0.011 | (-0.023, 0.045) | 0.535 | 0.813 | 0.045 | (-0.023, 0.043) |

**Supplementary Table S6.** Regression and Bayesian estimates comparing sex-stratified ME/CFS vs. controls.

| Metabolite | Regression Model | Female ME/CFS vs. Female Controls | | | | | |
|---|---|---|---|---|---|---|---|
| | | Estimated Coefficient | 95% CI | p-value | FDR | BayesFactor | 95% HDI |
| heptadecanoic acid | Lognormal | 0.006 | (-0.014, 0.026) | 0.542 | 0.821 | 0.045 | (-0.015, 0.025) |
| phosphate | Lognormal | 0.003 | (-0.007, 0.014) | 0.544 | 0.822 | 0.038 | (-0.007, 0.014) |
| isothreonic acid | Lognormal | -0.009 | (-0.037, 0.020) | 0.551 | 0.829 | 0.049 | (-0.036, 0.020) |
| citric acid | Lognormal | -0.006 | (-0.027, 0.015) | 0.560 | 0.836 | 0.038 | (-0.028, 0.014) |
| behenic acid | Lognormal | -0.005 | (-0.023, 0.013) | 0.572 | 0.847 | 0.053 | (-0.022, 0.014) |
| palmitic acid | Lognormal | 0.003 | (-0.007, 0.012) | 0.576 | 0.847 | 0.045 | (-0.007, 0.012) |
| ribose | Lognormal | -0.005 | (-0.021, 0.012) | 0.579 | 0.847 | 0.050 | (-0.020, 0.013) |
| maltose | Lognormal | -0.007 | (-0.033, 0.019) | 0.593 | 0.858 | 0.045 | (-0.033, 0.020) |
| pentadecanoic acid | Lognormal | -0.003 | (-0.012, 0.007) | 0.610 | 0.865 | 0.041 | (-0.013, 0.007) |
| benzoic acid | Lognormal | 0.006 | (-0.016, 0.027) | 0.611 | 0.865 | 0.047 | (-0.018, 0.026) |
| 3-hydroxybutyric acid | Lognormal | 0.009 | (-0.027, 0.046) | 0.618 | 0.872 | 0.048 | (-0.025, 0.049) |
| creatine | Lognormal | -0.007 | (-0.037, 0.022) | 0.626 | 0.873 | 0.046 | (-0.037, 0.023) |
| fumaric acid | Lognormal | -0.005 | (-0.027, 0.016) | 0.631 | 0.874 | 0.045 | (-0.028, 0.016) |
| myristic acid | Lognormal | 0.004 | (-0.013, 0.021) | 0.637 | 0.878 | 0.046 | (-0.012, 0.022) |
| glycolic acid | Lognormal | 0.008 | (-0.026, 0.043) | 0.639 | 0.879 | 0.045 | (-0.023, 0.045) |
| 2-hydroxyvaleric acid | Lognormal | 0.005 | (-0.016, 0.026) | 0.642 | 0.881 | 0.051 | (-0.017, 0.025) |
| N-acetylornithine | Lognormal | -0.003 | (-0.018, 0.011) | 0.648 | 0.887 | 0.038 | (-0.018, 0.012) |
| malic acid | Lognormal | -0.005 | (-0.025, 0.016) | 0.660 | 0.895 | 0.043 | (-0.025, 0.016) |
| alloxanoic acid | Lognormal | 0.008 | (-0.030, 0.046) | 0.678 | 0.904 | 0.046 | (-0.033, 0.045) |
| 4-hydroxybutyric acid | Lognormal | 0.005 | (-0.019, 0.028) | 0.689 | 0.906 | 0.048 | (-0.019, 0.028) |
| threonic acid | Lognormal | 0.005 | (-0.020, 0.030) | 0.701 | 0.908 | 0.041 | (-0.021, 0.030) |
| gluconic acid | Lognormal | -0.003 | (-0.020, 0.014) | 0.707 | 0.913 | 0.040 | (-0.020, 0.015) |
| isopropylbenzene | Lognormal | 0.004 | (-0.017, 0.025) | 0.710 | 0.913 | 0.027 | (-0.018, 0.025) |
| 1,2,4-benzenetriol | Lognormal | -0.008 | (-0.051, 0.035) | 0.713 | 0.913 | 0.045 | (-0.053, 0.033) |
| lauric acid | Lognormal | 0.005 | (-0.020, 0.029) | 0.720 | 0.920 | 0.040 | (-0.020, 0.029) |
| pyruvic acid | Lognormal | 0.003 | (-0.013, 0.018) | 0.727 | 0.927 | 0.034 | (-0.013, 0.018) |
| salicylic acid | Lognormal | 0.006 | (-0.029, 0.042) | 0.731 | 0.927 | 0.043 | (-0.028, 0.045) |
| pyrrole-2-carboxylic acid | Lognormal | 0.005 | (-0.022, 0.031) | 0.734 | 0.928 | 0.041 | (-0.024, 0.032) |
| stearic acid | Lognormal | 0.002 | (-0.008, 0.011) | 0.734 | 0.928 | 0.038 | (-0.008, 0.011) |
| lactic acid | Lognormal | -0.004 | (-0.025, 0.017) | 0.739 | 0.928 | 0.042 | (-0.025, 0.017) |
| glucose-1-phosphate | Lognormal | 0.003 | (-0.016, 0.022) | 0.749 | 0.928 | 0.040 | (-0.015, 0.023) |
| palmitoleic acid | Lognormal | -0.005 | (-0.038, 0.028) | 0.755 | 0.931 | 0.042 | (-0.039, 0.026) |
| lyxitol | Lognormal | -0.003 | (-0.020, 0.015) | 0.756 | 0.931 | 0.041 | (-0.019, 0.015) |
| phthalic acid | Lognormal | 0.003 | (-0.015, 0.020) | 0.764 | 0.931 | 0.040 | (-0.013, 0.021) |
| glucuronic acid | Lognormal | -0.004 | (-0.027, 0.020) | 0.767 | 0.931 | 0.045 | (-0.026, 0.021) |
| threitol | Lognormal | 0.003 | (-0.017, 0.023) | 0.775 | 0.936 | 0.039 | (-0.016, 0.023) |
| capric acid | Lognormal | -0.004 | (-0.033, 0.025) | 0.780 | 0.936 | 0.038 | (-0.034, 0.024) |
| valine | Lognormal | 0.002 | (-0.010, 0.013) | 0.782 | 0.936 | 0.038 | (-0.009, 0.013) |
| glyceric acid | Lognormal | 0.003 | (-0.022, 0.029) | 0.790 | 0.936 | 0.042 | (-0.021, 0.029) |
| glutamic acid | Lognormal | -0.003 | (-0.024, 0.019) | 0.806 | 0.947 | 0.042 | (-0.025, 0.019) |
| trans-4-hydroxyproline | Lognormal | -0.003 | (-0.031, 0.024) | 0.822 | 0.952 | 0.039 | (-0.033, 0.024) |
| uric acid | Lognormal | -0.002 | (-0.025, 0.020) | 0.830 | 0.953 | 0.042 | (-0.024, 0.021) |
| adipic acid | Lognormal | -0.002 | (-0.025, 0.021) | 0.850 | 0.964 | 0.043 | (-0.025, 0.021) |
| 1-methylgalactose NIST | Lognormal | -0.004 | (-0.041, 0.034) | 0.854 | 0.964 | 0.041 | (-0.039, 0.034) |
| myo-inositol | Lognormal | 0.001 | (-0.011, 0.014) | 0.864 | 0.966 | 0.041 | (-0.012, 0.013) |
| maleic acid | Lognormal | 0.002 | (-0.026, 0.031) | 0.866 | 0.966 | 0.036 | (-0.026, 0.031) |
| indole-3-acetate | Lognormal | 0.002 | (-0.023, 0.026) | 0.887 | 0.970 | 0.037 | (-0.023, 0.026) |

**Supplementary Table S6.** Regression and Bayesian estimates comparing sex-stratified ME/CFS vs. controls.

| Metabolite | Regression Model | Female ME/CFS vs. Female Controls | | | | | |
|---|---|---|---|---|---|---|---|
| | | Estimated Coefficient | 95% CI | p-value | FDR | BayesFactor | 95% HDI |
| proline | Lognormal | -0.002 | (-0.028, 0.025) | 0.900 | 0.973 | 0.041 | (-0.028, 0.025) |
| 2-aminobutyric acid | Lognormal | 0.001 | (-0.014, 0.016) | 0.907 | 0.976 | 0.041 | (-0.014, 0.016) |
| oxalic acid | Lognormal | 0.002 | (-0.054, 0.059) | 0.933 | 0.990 | 0.041 | (-0.057, 0.059) |
| caprylic acid | Lognormal | 0.000 | (-0.014, 0.013) | 0.964 | 0.994 | 0.037 | (-0.013, 0.013) |
| glycine | Lognormal | 0.000 | (-0.012, 0.012) | 0.987 | 0.995 | 0.040 | (-0.011, 0.013) |
| ribonic acid | Lognormal | 0.000 | (-0.020, 0.020) | 0.993 | 0.997 | 0.039 | (-0.020, 0.021) |
| nicotinic acid | Lognormal | 0.000 | (-0.041, 0.042) | 0.995 | 0.997 | 0.032 | (-0.043, 0.040) |
| **Biogenic Amines (BA)** | | | | | | | |
| Alprazolam | Lognormal | **0.081** | **(0.030, 0.132)** | **0.002** | **0.121** | **3.486** | **(0.030, 0.130)** |
| Acyclovir | Lognormal | **0.152** | **(0.057, 0.247)** | **0.002** | **0.121** | **3.179** | **(0.056, 0.249)** |
| Linoleoylcarnitine | Lognormal | -0.052 | (-0.085, -0.018) | 0.003 | 0.140 | 2.103 | (-0.086, -0.018) |
| Guanine | Lognormal | 0.117 | (0.040, 0.194) | 0.004 | 0.160 | 2.962 | (0.042, 0.196) |
| Caffeine | Lognormal | -0.093 | (-0.156, -0.030) | 0.004 | 0.169 | 2.442 | (-0.152, -0.028) |
| 2-Methylbutyryl-L-carnitine | Lognormal | -0.039 | (-0.068, -0.010) | 0.009 | 0.206 | 1.009 | (-0.067, -0.010) |
| Acetominophen | Lognormal | 0.064 | (0.016, 0.113) | 0.010 | 0.211 | 1.172 | (0.019, 0.116) |
| Theobromine | Lognormal | -0.066 | (-0.117, -0.015) | 0.012 | 0.225 | 0.940 | (-0.115, -0.012) |
| Tri-2-ethylhexyl trimellitate | Lognormal | -0.058 | (-0.108, -0.008) | 0.025 | 0.291 | 0.511 | (-0.109, -0.009) |
| 7-Methylguanosine | Lognormal | -0.017 | (-0.032, -0.002) | 0.026 | 0.300 | 0.456 | (-0.032, -0.002) |
| Usnic acid | Lognormal | -0.056 | (-0.105, -0.007) | 0.028 | 0.312 | 0.440 | (-0.105, -0.007) |
| Choline cation | Lognormal | -0.008 | (-0.015, -0.001) | 0.031 | 0.315 | 0.411 | (-0.015, -0.001) |
| Piperine | Lognormal | -0.064 | (-0.123, -0.005) | 0.035 | 0.320 | 0.376 | (-0.120, -0.002) |
| Trigonelline | Lognormal | -0.048 | (-0.092, -0.004) | 0.036 | 0.320 | 0.300 | (-0.090, -0.001) |
| Oleoyl-L-carnitine | Lognormal | -0.041 | (-0.079, -0.003) | 0.036 | 0.320 | 0.363 | (-0.080, -0.005) |
| Kynurenine | Lognormal | -0.018 | (-0.034, -0.001) | 0.038 | 0.322 | 0.346 | (-0.034, -0.002) |
| Isopropylamine | Lognormal | -0.014 | (-0.027, -0.001) | 0.043 | 0.338 | 0.310 | (-0.026, 0.000) |
| Octanoylcarnitine | Lognormal | -0.023 | (-0.047, 0.000) | 0.053 | 0.379 | 0.227 | (-0.046, 0.001) |
| D-erythro-Sphingosine-1-phosphate | Lognormal | -0.024 | (-0.048, 0.000) | 0.054 | 0.379 | 0.251 | (-0.048, 0.000) |
| 1-Stearoyl-2-arachidonoyl-sn-glycero-3-phosphoserine | Lognormal | -0.081 | (-0.168, 0.005) | 0.067 | 0.413 | 0.184 | (-0.173, 0.000) |
| Albendazole sulfoxide | Lognormal | 0.100 | (-0.007, 0.207) | 0.068 | 0.414 | 0.236 | (-0.002, 0.205) |
| Ranitidine N-oxide | Lognormal | 0.052 | (-0.004, 0.107) | 0.070 | 0.416 | 0.204 | (-0.002, 0.109) |
| H-Pro-Hyp-OH | Lognormal | 0.025 | (-0.002, 0.052) | 0.074 | 0.428 | 0.202 | (-0.002, 0.052) |
| 4-Acetamidobutyric acid | Lognormal | -0.022 | (-0.047, 0.002) | 0.079 | 0.442 | 0.198 | (-0.048, 0.003) |
| 3-Methylglutarylcarnitine | Lognormal | -0.022 | (-0.046, 0.003) | 0.081 | 0.444 | 0.217 | (-0.045, 0.002) |
| 4,5,7-Trihydroxyisoflavone | Lognormal | 0.087 | (-0.010, 0.185) | 0.081 | 0.444 | 0.191 | (-0.005, 0.193) |
| 7-Hydroxywarfarin | Lognormal | 0.034 | (-0.004, 0.072) | 0.082 | 0.444 | 0.180 | (-0.005, 0.073) |
| 1-Monostearin | Lognormal | -0.032 | (-0.068, 0.004) | 0.084 | 0.444 | 0.172 | (-0.068, 0.004) |
| SDMA | Lognormal | -0.010 | (-0.021, 0.002) | 0.093 | 0.457 | 0.161 | (-0.021, 0.001) |
| Ergothioneine | Lognormal | -0.036 | (-0.078, 0.006) | 0.098 | 0.465 | 0.159 | (-0.078, 0.006) |
| 1-Oleoyl-2-acetyl-sn-glycerol | Lognormal | -0.034 | (-0.075, 0.007) | 0.110 | 0.493 | 0.126 | (-0.079, 0.008) |
| L-Citrulline | Lognormal | -0.009 | (-0.021, 0.002) | 0.112 | 0.494 | 0.163 | (-0.020, 0.002) |
| Pantoprazole | Lognormal | 0.051 | (-0.015, 0.116) | 0.131 | 0.521 | 0.118 | (-0.014, 0.114) |
| Phenylalanine | Lognormal | -0.007 | (-0.016, 0.002) | 0.133 | 0.521 | 0.113 | (-0.016, 0.002) |
| D-Turanose | Lognormal | -0.023 | (-0.053, 0.007) | 0.134 | 0.521 | 0.114 | (-0.052, 0.007) |
| Trazodone | Lognormal | 0.042 | (-0.013, 0.096) | 0.138 | 0.529 | 0.120 | (-0.012, 0.095) |
| 2,6-Diaminopimelic acid | Lognormal | 0.037 | (-0.012, 0.086) | 0.140 | 0.532 | 0.111 | (-0.011, 0.085) |
| N-.alpha.-Acetyl-L-arginine | Lognormal | -0.012 | (-0.028, 0.004) | 0.141 | 0.533 | 0.119 | (-0.029, 0.004) |
| 2-Amino-1-phenylethanol | Lognormal | -0.007 | (-0.016, 0.002) | 0.143 | 0.535 | 0.117 | (-0.016, 0.002) |

**Supplementary Table S6.** Regression and Bayesian estimates comparing sex-stratified ME/CFS vs. controls.

| Metabolite | Regression Model | Female ME/CFS vs. Female Controls | | | | | |
|---|---|---|---|---|---|---|---|
| | | Estimated Coefficient | 95% CI | p-value | FDR | BayesFactor | 95% HDI |
| Modafinil acid | Lognormal | 0.051 | (-0.017, 0.119) | 0.143 | 0.535 | 0.125 | (-0.018, 0.119) |
| Decanoyl-L-carnitine | Lognormal | -0.021 | (-0.049, 0.007) | 0.147 | 0.535 | 0.109 | (-0.050, 0.010) |
| 3-Amino-1-propanol | Lognormal | 0.016 | (-0.006, 0.039) | 0.150 | 0.543 | 0.107 | (-0.006, 0.040) |
| 1-Oleoyl-sn-glycero-3-phosphoethanolamine | Lognormal | -0.027 | (-0.063, 0.010) | 0.155 | 0.555 | 0.114 | (-0.063, 0.013) |
| H-gamma-glutamyl-glutamine | Lognormal | 0.020 | (-0.008, 0.049) | 0.159 | 0.559 | 0.089 | (-0.009, 0.049) |
| Lauroyl-L-carnitine | Lognormal | -0.032 | (-0.076, 0.012) | 0.160 | 0.561 | 0.092 | (-0.076, 0.015) |
| L-Leucine, methyl ester | Lognormal | 0.025 | (-0.010, 0.060) | 0.160 | 0.561 | 0.091 | (-0.013, 0.059) |
| Glutamic acid | Lognormal | 0.011 | (-0.005, 0.028) | 0.168 | 0.584 | 0.082 | (-0.005, 0.027) |
| Citrulline | Lognormal | -0.008 | (-0.021, 0.004) | 0.176 | 0.591 | 0.100 | (-0.022, 0.003) |
| Trimethylamine-N-oxide | Lognormal | 0.019 | (-0.009, 0.048) | 0.176 | 0.591 | 0.085 | (-0.010, 0.047) |
| 3-Hydroxypyridine | Lognormal | -0.053 | (-0.132, 0.025) | 0.183 | 0.596 | 0.075 | (-0.133, 0.022) |
| Losartan | Lognormal | -0.027 | (-0.066, 0.013) | 0.184 | 0.596 | 0.105 | (-0.067, 0.014) |
| Lamotrigine; | Lognormal | 0.091 | (-0.044, 0.225) | 0.189 | 0.601 | 0.093 | (-0.045, 0.235) |
| Gabapentin | Lognormal | 0.040 | (-0.020, 0.101) | 0.197 | 0.605 | 0.086 | (-0.021, 0.097) |
| Pyridoxine; | Lognormal | 0.027 | (-0.014, 0.068) | 0.202 | 0.610 | 0.076 | (-0.016, 0.069) |
| N-Acetylhistidine | Lognormal | -0.017 | (-0.043, 0.009) | 0.203 | 0.610 | 0.090 | (-0.043, 0.008) |
| Quetiapine | Lognormal | 0.017 | (-0.009, 0.044) | 0.203 | 0.610 | 0.092 | (-0.010, 0.042) |
| N-Methylhistidine | Lognormal | -0.024 | (-0.062, 0.013) | 0.206 | 0.610 | 0.087 | (-0.060, 0.015) |
| .beta.-Phenyl-.gamma.-aminobutyric acid | Lognormal | -0.013 | (-0.033, 0.007) | 0.208 | 0.610 | 0.087 | (-0.032, 0.008) |
| Testosterone | Lognormal | -0.025 | (-0.063, 0.014) | 0.210 | 0.610 | 0.077 | (-0.062, 0.012) |
| Thr-Ile-Arg | Lognormal | -0.031 | (-0.080, 0.017) | 0.211 | 0.610 | 0.086 | (-0.078, 0.018) |
| Dexpanthenol | Lognormal | -0.023 | (-0.058, 0.013) | 0.212 | 0.610 | 0.070 | (-0.056, 0.015) |
| 1-Stearoyl-2-arachidonoyl-sn-glycero-3-phospho-(1'-myo-inositol) | Lognormal | -0.058 | (-0.150, 0.034) | 0.220 | 0.621 | 0.091 | (-0.144, 0.039) |
| Creatinine | Lognormal | -0.004 | (-0.011, 0.003) | 0.225 | 0.624 | 0.072 | (-0.012, 0.003) |
| p-Acetamidophenyl .beta.-D-glucuronide | Lognormal | 0.038 | (-0.023, 0.098) | 0.229 | 0.624 | 0.086 | (-0.025, 0.096) |
| Ranitidine | Lognormal | 0.028 | (-0.017, 0.073) | 0.230 | 0.624 | 0.084 | (-0.019, 0.073) |
| (R)-Butyrylcarnitine | Lognormal | -0.017 | (-0.045, 0.011) | 0.231 | 0.624 | 0.078 | (-0.046, 0.011) |
| Glycocholic acid | Lognormal | -0.023 | (-0.062, 0.016) | 0.245 | 0.636 | 0.068 | (-0.061, 0.018) |
| N-Acetyl-D-norleucine | Lognormal | 0.015 | (-0.010, 0.040) | 0.246 | 0.636 | 0.046 | (-0.012, 0.038) |
| N.alpha.-Methyl-L-lysine | Lognormal | 0.023 | (-0.016, 0.062) | 0.250 | 0.637 | 0.079 | (-0.017, 0.062) |
| Betaine | Lognormal | -0.005 | (-0.012, 0.003) | 0.252 | 0.637 | 0.077 | (-0.012, 0.003) |
| Norleucine | Lognormal | -0.006 | (-0.017, 0.005) | 0.252 | 0.637 | 0.071 | (-0.018, 0.006) |
| Nudifloramide | Lognormal | -0.013 | (-0.035, 0.009) | 0.258 | 0.642 | 0.073 | (-0.034, 0.010) |
| 1-Palmitoyl-2-hydroxy-sn-glycero-3-phosphoethanolamine | Lognormal | -0.022 | (-0.060, 0.016) | 0.261 | 0.642 | 0.078 | (-0.059, 0.016) |
| Fexofenadine | Lognormal | -0.018 | (-0.049, 0.013) | 0.263 | 0.642 | 0.073 | (-0.048, 0.015) |
| Hexanoyl-L-carnitine | Lognormal | -0.015 | (-0.042, 0.011) | 0.263 | 0.642 | 0.069 | (-0.042, 0.011) |
| cis-10,11-Dihydroxy-10,11-dihydrocarbamazepine | Lognormal | -0.015 | (-0.042, 0.011) | 0.264 | 0.642 | 0.073 | (-0.042, 0.012) |
| Acetazolamide | Lognormal | -0.025 | (-0.069, 0.019) | 0.264 | 0.642 | 0.074 | (-0.068, 0.017) |
| 1-Acetyl-3-piperidinamine | Lognormal | 0.011 | (-0.009, 0.031) | 0.271 | 0.649 | 0.072 | (-0.009, 0.031) |
| Ala-Ile | Lognormal | 0.013 | (-0.010, 0.037) | 0.272 | 0.650 | 0.070 | (-0.010, 0.037) |
| Quetiapine sulfoxide | Lognormal | 0.019 | (-0.015, 0.052) | 0.276 | 0.651 | 0.068 | (-0.015, 0.052) |
| Tryptophan | Lognormal | 0.007 | (-0.006, 0.019) | 0.283 | 0.657 | 0.067 | (-0.005, 0.020) |
| Prazepam | Lognormal | 0.028 | (-0.023, 0.080) | 0.285 | 0.657 | 0.064 | (-0.021, 0.079) |
| Omeprazole sulfone N-oxide | Lognormal | 0.042 | (-0.035, 0.119) | 0.287 | 0.657 | 0.067 | (-0.037, 0.127) |
| Montelukast-1,2-diol | Lognormal | 0.044 | (-0.037, 0.125) | 0.287 | 0.657 | 0.070 | (-0.038, 0.129) |
| Diazepam | Lognormal | 0.038 | (-0.032, 0.108) | 0.288 | 0.657 | 0.067 | (-0.027, 0.114) |
| Irbesartan | Lognormal | 0.017 | (-0.014, 0.047) | 0.289 | 0.657 | 0.067 | (-0.014, 0.047) |

**Supplementary Table S6.** Regression and Bayesian estimates comparing sex-stratified ME/CFS vs. controls.

| Metabolite | Regression Model | Female ME/CFS vs. Female Controls | | | | | |
|---|---|---|---|---|---|---|---|
| | | Estimated Coefficient | 95% CI | p-value | FDR | BayesFactor | 95% HDI |
| Betonicine | Lognormal | -0.043 | (-0.122, 0.037) | 0.293 | 0.662 | 0.070 | (-0.128, 0.035) |
| Adenosine | Lognormal | -0.017 | (-0.048, 0.015) | 0.306 | 0.676 | 0.064 | (-0.049, 0.017) |
| Ethiolat | Lognormal | -0.021 | (-0.062, 0.019) | 0.309 | 0.676 | 0.065 | (-0.059, 0.021) |
| Toradol | Lognormal | 0.025 | (-0.023, 0.072) | 0.310 | 0.676 | 0.069 | (-0.022, 0.071) |
| NEPSILON,NEPSILON,NEPSILON-TRIMETHYLLYSINE | Lognormal | -0.007 | (-0.020, 0.006) | 0.314 | 0.676 | 0.054 | (-0.020, 0.007) |
| Histidine | Lognormal | -0.013 | (-0.040, 0.013) | 0.320 | 0.684 | 0.068 | (-0.040, 0.014) |
| Codeine-6-.beta.-D-glucuronide | Lognormal | 0.019 | (-0.018, 0.056) | 0.325 | 0.684 | 0.068 | (-0.020, 0.056) |
| Ethyldiethanolamine | Lognormal | -0.016 | (-0.049, 0.016) | 0.325 | 0.684 | 0.057 | (-0.050, 0.015) |
| 6-Hydroxyflavone | Lognormal | 0.023 | (-0.022, 0.068) | 0.325 | 0.684 | 0.056 | (-0.025, 0.065) |
| Omeprazole sulfone | Lognormal | 0.037 | (-0.039, 0.113) | 0.340 | 0.691 | 0.064 | (-0.039, 0.109) |
| L-Tyrosine | Lognormal | -0.011 | (-0.034, 0.012) | 0.342 | 0.691 | 0.063 | (-0.034, 0.012) |
| Aminodiphenylmethane | Lognormal | 0.020 | (-0.021, 0.060) | 0.345 | 0.691 | 0.065 | (-0.021, 0.061) |
| 1-Methylnicotinamide | Lognormal | -0.011 | (-0.033, 0.011) | 0.345 | 0.691 | 0.057 | (-0.035, 0.011) |
| Phenylacetyl-L-glutamine | Lognormal | -0.020 | (-0.062, 0.022) | 0.354 | 0.701 | 0.054 | (-0.061, 0.022) |
| Tauroursodeoxycholic acid | Lognormal | 0.018 | (-0.021, 0.057) | 0.360 | 0.704 | 0.063 | (-0.021, 0.060) |
| Tyrosine | Lognormal | -0.008 | (-0.024, 0.009) | 0.360 | 0.704 | 0.053 | (-0.025, 0.008) |
| N-(3-(Aminomethyl)benzyl)acetamidine | Lognormal | -0.017 | (-0.053, 0.019) | 0.360 | 0.704 | 0.047 | (-0.055, 0.021) |
| 3-Dehydrocarnitine | Lognormal | -0.008 | (-0.026, 0.009) | 0.363 | 0.704 | 0.050 | (-0.025, 0.010) |
| 3-Hydroxyoleylcarnitine | Lognormal | 0.039 | (-0.045, 0.124) | 0.363 | 0.704 | 0.060 | (-0.042, 0.127) |
| Pyrantel | Lognormal | -0.014 | (-0.043, 0.016) | 0.369 | 0.709 | 0.062 | (-0.042, 0.017) |
| Ile-Glu-Arg | Lognormal | -0.031 | (-0.100, 0.038) | 0.380 | 0.713 | 0.059 | (-0.098, 0.042) |
| Moxonidine | Lognormal | -0.013 | (-0.043, 0.016) | 0.380 | 0.713 | 0.060 | (-0.042, 0.016) |
| Ornithine | Lognormal | -0.008 | (-0.027, 0.010) | 0.380 | 0.713 | 0.045 | (-0.028, 0.009) |
| L-Threonine | Lognormal | -0.006 | (-0.020, 0.008) | 0.385 | 0.721 | 0.061 | (-0.019, 0.008) |
| 3-Methylxanthine | Lognormal | -0.021 | (-0.069, 0.027) | 0.387 | 0.722 | 0.055 | (-0.071, 0.027) |
| (3-Carboxypropyl)trimethylammonium cation | Lognormal | -0.005 | (-0.017, 0.007) | 0.401 | 0.733 | 0.052 | (-0.017, 0.006) |
| Hydroxybupropion | Lognormal | -0.017 | (-0.057, 0.023) | 0.404 | 0.733 | 0.063 | (-0.058, 0.022) |
| Glaucine | Lognormal | 0.011 | (-0.016, 0.038) | 0.414 | 0.739 | 0.054 | (-0.017, 0.035) |
| L-Cystine | Lognormal | -0.010 | (-0.036, 0.015) | 0.417 | 0.739 | 0.039 | (-0.036, 0.015) |
| Methacholine cation | Lognormal | -0.009 | (-0.032, 0.013) | 0.421 | 0.739 | 0.051 | (-0.032, 0.015) |
| Urea | Lognormal | -0.005 | (-0.016, 0.006) | 0.423 | 0.739 | 0.048 | (-0.015, 0.006) |
| 4-Fluoro-.alpha.-pyrrolidinobutiophenone | Lognormal | 0.011 | (-0.016, 0.038) | 0.432 | 0.753 | 0.054 | (-0.017, 0.039) |
| Benthiavalicarb-isopropyl | Lognormal | -0.017 | (-0.062, 0.027) | 0.444 | 0.763 | 0.052 | (-0.064, 0.027) |
| Modafinil | Lognormal | 0.021 | (-0.033, 0.074) | 0.447 | 0.765 | 0.054 | (-0.032, 0.074) |
| Palmitamide | Lognormal | -0.018 | (-0.066, 0.029) | 0.452 | 0.768 | 0.058 | (-0.068, 0.030) |
| alpha-Methylhistidine; | Lognormal | -0.020 | (-0.073, 0.033) | 0.455 | 0.771 | 0.052 | (-0.075, 0.034) |
| Ranitidine-S-oxide | Lognormal | -0.023 | (-0.084, 0.038) | 0.459 | 0.775 | 0.049 | (-0.086, 0.039) |
| Metoprolol acid | Lognormal | 0.021 | (-0.034, 0.075) | 0.462 | 0.776 | 0.049 | (-0.031, 0.076) |
| 7.alpha.-Hydroxy-3-oxo-4-cholestenoic acid | Lognormal | -0.009 | (-0.031, 0.014) | 0.467 | 0.781 | 0.054 | (-0.031, 0.015) |
| 5'-S-Methyl-5'-thioadenosine | Lognormal | -0.031 | (-0.113, 0.052) | 0.470 | 0.781 | 0.045 | (-0.119, 0.044) |
| Stachydrine | Lognormal | -0.016 | (-0.059, 0.027) | 0.470 | 0.781 | 0.051 | (-0.061, 0.026) |
| Glycodeoxycholic acid | Lognormal | -0.016 | (-0.058, 0.027) | 0.474 | 0.784 | 0.047 | (-0.056, 0.031) |
| Carnitine | Lognormal | -0.003 | (-0.011, 0.005) | 0.488 | 0.796 | 0.047 | (-0.011, 0.005) |
| Topiramate | Lognormal | 0.013 | (-0.024, 0.051) | 0.491 | 0.798 | 0.050 | (-0.026, 0.051) |
| Meprobamate | Lognormal | 0.012 | (-0.022, 0.045) | 0.504 | 0.806 | 0.052 | (-0.023, 0.044) |
| DL-Indole-3-lactic acid | Lognormal | 0.005 | (-0.009, 0.018) | 0.506 | 0.806 | 0.052 | (-0.009, 0.018) |
| Ethylenediaminetetraacetic acid | Lognormal | 0.031 | (-0.060, 0.122) | 0.507 | 0.806 | 0.047 | (-0.064, 0.123) |

**Supplementary Table S6.** Regression and Bayesian estimates comparing sex-stratified ME/CFS vs. controls.

| Metabolite | Regression Model | Female ME/CFS vs. Female Controls | | | | | |
|---|---|---|---|---|---|---|---|
| | | Estimated Coefficient | 95% CI | p-value | FDR | BayesFactor | 95% HDI |
| Gly-Pro-Arg | Lognormal | -0.022 | (-0.087, 0.043) | 0.507 | 0.806 | 0.049 | (-0.086, 0.041) |
| Pipecolic acid | Lognormal | -0.016 | (-0.065, 0.032) | 0.510 | 0.808 | 0.046 | (-0.063, 0.031) |
| L-Cysteine-glutathione disulfide | Lognormal | -0.011 | (-0.045, 0.022) | 0.518 | 0.811 | 0.031 | (-0.043, 0.024) |
| 3-Cysteinylacetaminophen | Lognormal | 0.034 | (-0.069, 0.138) | 0.518 | 0.811 | 0.046 | (-0.067, 0.138) |
| Ezetimibe | Lognormal | 0.015 | (-0.030, 0.060) | 0.523 | 0.813 | 0.048 | (-0.032, 0.057) |
| 1-Methyl-L-histidine | Lognormal | -0.016 | (-0.063, 0.032) | 0.525 | 0.813 | 0.046 | (-0.063, 0.031) |
| Carbamazepine | Lognormal | -0.020 | (-0.081, 0.041) | 0.529 | 0.813 | 0.051 | (-0.078, 0.044) |
| 5'-S-Methylthioadenosine | Lognormal | -0.030 | (-0.122, 0.063) | 0.530 | 0.813 | 0.041 | (-0.121, 0.067) |
| Dinor-12-oxophytodienoic acid | Lognormal | -0.016 | (-0.067, 0.035) | 0.532 | 0.813 | 0.047 | (-0.070, 0.033) |
| Atorvastatin | Lognormal | 0.007 | (-0.016, 0.031) | 0.536 | 0.814 | 0.052 | (-0.016, 0.030) |
| N-Methylproline | Lognormal | -0.009 | (-0.036, 0.019) | 0.543 | 0.822 | 0.046 | (-0.036, 0.019) |
| 4'-Methyl-N-methylhexanophenone | Lognormal | -0.019 | (-0.082, 0.044) | 0.548 | 0.826 | 0.046 | (-0.079, 0.046) |
| trans-3'-Hydroxycotinine | Lognormal | 0.010 | (-0.023, 0.043) | 0.556 | 0.834 | 0.047 | (-0.024, 0.042) |
| Guanidine | Lognormal | 0.007 | (-0.017, 0.031) | 0.573 | 0.847 | 0.045 | (-0.017, 0.030) |
| Telmisartan | Lognormal | 0.009 | (-0.026, 0.045) | 0.597 | 0.859 | 0.040 | (-0.027, 0.043) |
| Theanine; | Lognormal | 0.010 | (-0.028, 0.049) | 0.601 | 0.861 | 0.038 | (-0.028, 0.052) |
| Methylgallate | Lognormal | 0.026 | (-0.072, 0.124) | 0.607 | 0.865 | 0.043 | (-0.074, 0.123) |
| Avobenzone | Lognormal | 0.007 | (-0.020, 0.033) | 0.613 | 0.867 | 0.047 | (-0.019, 0.034) |
| 3,4-Dimethoxybenzaldehyde | Lognormal | 0.009 | (-0.026, 0.044) | 0.622 | 0.872 | 0.037 | (-0.027, 0.043) |
| Trimethoprim | Lognormal | -0.009 | (-0.046, 0.028) | 0.624 | 0.872 | 0.045 | (-0.045, 0.028) |
| Ticlopidine | Lognormal | -0.024 | (-0.123, 0.074) | 0.632 | 0.874 | 0.043 | (-0.123, 0.075) |
| N.epsilon.-Acetyl-L-lysine | Lognormal | -0.005 | (-0.024, 0.015) | 0.634 | 0.876 | 0.046 | (-0.024, 0.015) |
| 2,2-Bishydroxymethyl]-2,2',2''-nitrilotriethanol | Lognormal | -0.007 | (-0.039, 0.024) | 0.645 | 0.884 | 0.044 | (-0.040, 0.025) |
| Betaine aldehyde cation | Lognormal | 0.014 | (-0.050, 0.078) | 0.661 | 0.895 | 0.041 | (-0.052, 0.075) |
| Pantothenic acid | Lognormal | -0.006 | (-0.031, 0.020) | 0.661 | 0.895 | 0.043 | (-0.030, 0.021) |
| D-Pyroglutamic acid | Lognormal | -0.001 | (-0.008, 0.005) | 0.670 | 0.900 | 0.043 | (-0.008, 0.005) |
| 2-Indolinone | Lognormal | -0.005 | (-0.028, 0.018) | 0.682 | 0.905 | 0.043 | (-0.028, 0.020) |
| Pyridoxal | Lognormal | 0.006 | (-0.024, 0.037) | 0.685 | 0.906 | 0.041 | (-0.022, 0.037) |
| (2R)-3-Hydroxyisovaleroylcarnitine | Lognormal | -0.004 | (-0.023, 0.015) | 0.686 | 0.906 | 0.041 | (-0.023, 0.015) |
| Arginine | Lognormal | 0.003 | (-0.013, 0.019) | 0.689 | 0.906 | 0.039 | (-0.014, 0.018) |
| Metoprolol | Lognormal | 0.009 | (-0.037, 0.056) | 0.696 | 0.907 | 0.045 | (-0.039, 0.056) |
| N-Acetyl-L-carnosine | Lognormal | -0.004 | (-0.026, 0.018) | 0.698 | 0.908 | 0.043 | (-0.025, 0.018) |
| 3-Hydroxybutyrylcarnitine | Lognormal | -0.006 | (-0.040, 0.027) | 0.710 | 0.913 | 0.043 | (-0.038, 0.026) |
| 4-Aminomethylcyclohexanecarboxylic acid; | Lognormal | -0.006 | (-0.036, 0.025) | 0.712 | 0.913 | 0.037 | (-0.035, 0.026) |
| D-.alpha.-Cyclohexylglycine | Lognormal | -0.006 | (-0.038, 0.026) | 0.713 | 0.913 | 0.044 | (-0.039, 0.027) |
| Urocanic acid; | Lognormal | -0.004 | (-0.026, 0.018) | 0.729 | 0.927 | 0.041 | (-0.025, 0.019) |
| L-Carnitine | Lognormal | -0.002 | (-0.015, 0.010) | 0.742 | 0.928 | 0.042 | (-0.014, 0.011) |
| Metformin | Lognormal | 0.007 | (-0.036, 0.051) | 0.743 | 0.928 | 0.042 | (-0.038, 0.049) |
| Cotinine N-.beta.-D-glucuronide | Lognormal | 0.012 | (-0.058, 0.081) | 0.746 | 0.928 | 0.044 | (-0.056, 0.083) |
| Glutamine | Lognormal | -0.001 | (-0.008, 0.006) | 0.750 | 0.928 | 0.040 | (-0.008, 0.006) |
| Heptadecasphing-4-enine | Lognormal | 0.021 | (-0.107, 0.149) | 0.750 | 0.928 | 0.039 | (-0.106, 0.150) |
| Ser-Tyr-Lys | Lognormal | -0.010 | (-0.072, 0.052) | 0.759 | 0.931 | 0.042 | (-0.075, 0.050) |
| 3-Aminoquinoline | Lognormal | -0.007 | (-0.054, 0.039) | 0.762 | 0.931 | 0.031 | (-0.056, 0.039) |
| Biliverden | Lognormal | -0.003 | (-0.026, 0.019) | 0.763 | 0.931 | 0.039 | (-0.026, 0.018) |
| Borrelidin | Lognormal | 0.008 | (-0.048, 0.065) | 0.769 | 0.931 | 0.041 | (-0.047, 0.063) |
| Creatine | Lognormal | -0.003 | (-0.019, 0.014) | 0.769 | 0.931 | 0.045 | (-0.019, 0.015) |
| Isopentenyladenine | Lognormal | -0.001 | (-0.012, 0.009) | 0.784 | 0.936 | 0.042 | (-0.012, 0.008) |

**Supplementary Table S6.** Regression and Bayesian estimates comparing sex-stratified ME/CFS vs. controls.

| Metabolite | Regression Model | Female ME/CFS vs. Female Controls | | | | | |
|---|---|---|---|---|---|---|---|
| | | Estimated Coefficient | 95% CI | p-value | FDR | BayesFactor | 95% HDI |
| Indole-3-propionic acid | Lognormal | -0.004 | (-0.036, 0.027) | 0.786 | 0.936 | 0.040 | (-0.035, 0.027) |
| D-Fructose | Lognormal | 0.001 | (-0.007, 0.009) | 0.790 | 0.936 | 0.034 | (-0.007, 0.008) |
| rac-4-Sulfoxypropranolol | Lognormal | 0.004 | (-0.028, 0.036) | 0.799 | 0.945 | 0.040 | (-0.025, 0.038) |
| 3-Pyridinemthanol | Lognormal | 0.004 | (-0.024, 0.032) | 0.800 | 0.945 | 0.042 | (-0.025, 0.031) |
| Levocetirizine; | Lognormal | 0.010 | (-0.074, 0.095) | 0.810 | 0.948 | 0.042 | (-0.074, 0.097) |
| .epsilon.-Caprolactam | Lognormal | 0.002 | (-0.017, 0.021) | 0.813 | 0.949 | 0.024 | (-0.016, 0.021) |
| N-Acetylalanine | Lognormal | 0.008 | (-0.064, 0.080) | 0.825 | 0.952 | 0.042 | (-0.062, 0.081) |
| 1,2-Dimethylimidazole | Lognormal | -0.003 | (-0.029, 0.023) | 0.827 | 0.952 | 0.041 | (-0.028, 0.022) |
| R-(-)-O-Desmethylvenlafaxine | Lognormal | -0.006 | (-0.060, 0.048) | 0.833 | 0.955 | 0.041 | (-0.059, 0.046) |
| Naproxen | Lognormal | -0.005 | (-0.052, 0.042) | 0.841 | 0.960 | 0.041 | (-0.049, 0.043) |
| N8-Acetylspermidine | Lognormal | -0.002 | (-0.017, 0.014) | 0.843 | 0.960 | 0.035 | (-0.016, 0.014) |
| 1-Stearoyl-2-hydroxy-sn-glycero-3-phosphoethanolamine | Lognormal | -0.004 | (-0.040, 0.033) | 0.848 | 0.963 | 0.037 | (-0.040, 0.034) |
| Propionylcarnitine | Lognormal | -0.001 | (-0.011, 0.009) | 0.851 | 0.964 | 0.035 | (-0.011, 0.010) |
| Matrine | Lognormal | -0.005 | (-0.057, 0.047) | 0.856 | 0.964 | 0.039 | (-0.058, 0.046) |
| Trileptal | Lognormal | -0.003 | (-0.034, 0.028) | 0.858 | 0.964 | 0.039 | (-0.031, 0.030) |
| Androstan-3-ol-17-one 3-glucuronide | Lognormal | 0.003 | (-0.032, 0.038) | 0.858 | 0.964 | 0.039 | (-0.032, 0.039) |
| Serotonin | Lognormal | 0.002 | (-0.019, 0.023) | 0.860 | 0.964 | 0.035 | (-0.018, 0.022) |
| Lansoprazole | Lognormal | 0.010 | (-0.106, 0.126) | 0.862 | 0.965 | 0.041 | (-0.102, 0.131) |
| Atenolol | Lognormal | -0.003 | (-0.044, 0.037) | 0.870 | 0.966 | 0.040 | (-0.043, 0.035) |
| 1-Acetyl-4-piperidinamine | Lognormal | 0.004 | (-0.041, 0.049) | 0.870 | 0.966 | 0.042 | (-0.043, 0.047) |
| Homoarginine; | Lognormal | 0.002 | (-0.020, 0.023) | 0.871 | 0.966 | 0.031 | (-0.021, 0.023) |
| 1-Phenylpyrrolidine | Lognormal | 0.003 | (-0.033, 0.039) | 0.875 | 0.969 | 0.040 | (-0.032, 0.038) |
| Acetyl-DL-carnitine | Lognormal | 0.001 | (-0.016, 0.019) | 0.878 | 0.970 | 0.039 | (-0.016, 0.019) |
| 2-Hydroxyibuprofen | Lognormal | 0.003 | (-0.033, 0.038) | 0.881 | 0.970 | 0.039 | (-0.032, 0.039) |
| Milnacipran | Lognormal | 0.003 | (-0.039, 0.045) | 0.882 | 0.970 | 0.039 | (-0.039, 0.044) |
| Triptolide | Lognormal | 0.002 | (-0.025, 0.029) | 0.887 | 0.970 | 0.036 | (-0.025, 0.028) |
| 2,2',2''-Nitrilotriethanol | Lognormal | 0.002 | (-0.029, 0.033) | 0.889 | 0.971 | 0.039 | (-0.032, 0.031) |
| Albendazole | Lognormal | 0.003 | (-0.035, 0.041) | 0.893 | 0.973 | 0.040 | (-0.037, 0.042) |
| Scopoletin | Lognormal | -0.006 | (-0.096, 0.084) | 0.894 | 0.973 | 0.031 | (-0.095, 0.087) |
| Mefenorex | Lognormal | 0.002 | (-0.034, 0.039) | 0.901 | 0.973 | 0.034 | (-0.032, 0.041) |
| Diphenhydramine | Lognormal | -0.002 | (-0.028, 0.025) | 0.901 | 0.973 | 0.040 | (-0.028, 0.023) |
| 3-(1-Pyrazolyl)-alanine | Lognormal | 0.005 | (-0.071, 0.080) | 0.905 | 0.975 | 0.038 | (-0.075, 0.079) |
| Methionine | Lognormal | -0.001 | (-0.012, 0.011) | 0.919 | 0.982 | 0.038 | (-0.013, 0.011) |
| Hypoxanthine | Lognormal | -0.001 | (-0.020, 0.019) | 0.935 | 0.990 | 0.034 | (-0.021, 0.019) |
| Tapentadol-.beta.-D-glucuronide | Lognormal | 0.002 | (-0.056, 0.061) | 0.943 | 0.994 | 0.041 | (-0.059, 0.059) |
| Methioninesulfoxide | Lognormal | 0.001 | (-0.022, 0.024) | 0.949 | 0.994 | 0.037 | (-0.022, 0.024) |
| Ondansetron | Lognormal | 0.001 | (-0.027, 0.029) | 0.952 | 0.994 | 0.037 | (-0.029, 0.027) |
| threo-Dihydrobupropion | Lognormal | 0.001 | (-0.032, 0.034) | 0.953 | 0.994 | 0.040 | (-0.031, 0.035) |
| Isoleucine | Lognormal | 0.000 | (-0.012, 0.011) | 0.954 | 0.994 | 0.038 | (-0.012, 0.011) |
| Esomeprazole | Lognormal | 0.001 | (-0.038, 0.040) | 0.955 | 0.994 | 0.042 | (-0.041, 0.038) |
| Meloxicam | Lognormal | -0.003 | (-0.123, 0.116) | 0.957 | 0.994 | 0.042 | (-0.123, 0.118) |
| 3,5-Dihydroxyphenylglycine | Lognormal | 0.001 | (-0.034, 0.036) | 0.959 | 0.994 | 0.036 | (-0.036, 0.034) |
| Temazepam | Lognormal | 0.001 | (-0.042, 0.044) | 0.960 | 0.994 | 0.039 | (-0.040, 0.044) |
| Lauric acid diethanolamide | Lognormal | -0.001 | (-0.048, 0.046) | 0.961 | 0.994 | 0.041 | (-0.049, 0.047) |
| 4-Pyridoxic acid; | Lognormal | 0.001 | (-0.051, 0.053) | 0.962 | 0.994 | 0.037 | (-0.050, 0.053) |
| Phenylacetylglutamine | Lognormal | 0.001 | (-0.030, 0.031) | 0.970 | 0.994 | 0.035 | (-0.031, 0.031) |
| Penciclovir | Lognormal | -0.001 | (-0.035, 0.034) | 0.971 | 0.994 | 0.034 | (-0.036, 0.032) |

**Supplementary Table S6.** Regression and Bayesian estimates comparing sex-stratified ME/CFS vs. controls.

| Metabolite | Regression Model | Female ME/CFS vs. Female Controls | | | | | |
|---|---|---|---|---|---|---|---|
| | | Estimated Coefficient | 95% CI | p-value | FDR | BayesFactor | 95% HDI |
| 6-Methoxynaphthaleneacetic acid | Lognormal | 0.000 | (-0.013, 0.012) | 0.974 | 0.994 | 0.037 | (-0.013, 0.012) |
| 1-Methyladenosine A | Lognormal | 0.000 | (-0.011, 0.011) | 0.977 | 0.995 | 0.038 | (-0.011, 0.010) |
| Coniferylaldehyde | Lognormal | 0.000 | (-0.027, 0.027) | 0.983 | 0.995 | 0.041 | (-0.029, 0.026) |
| Bradykinin | Lognormal | 0.001 | (-0.113, 0.115) | 0.984 | 0.995 | 0.038 | (-0.113, 0.112) |
| Lysine | Lognormal | 0.000 | (-0.015, 0.016) | 0.984 | 0.995 | 0.032 | (-0.016, 0.016) |
| Benzophenone-3 | Lognormal | 0.001 | (-0.071, 0.072) | 0.987 | 0.995 | 0.040 | (-0.069, 0.074) |
| Sulfamethoxazole | Lognormal | -0.001 | (-0.102, 0.101) | 0.990 | 0.997 | 0.040 | (-0.111, 0.097) |
| Cyclo(Leu-Pro) | Lognormal | 0.000 | (-0.030, 0.029) | 0.991 | 0.997 | 0.040 | (-0.032, 0.027) |
| N.epsilon.-Methyl-L-lysine | Lognormal | 0.000 | (-0.034, 0.034) | 0.993 | 0.997 | 0.036 | (-0.034, 0.035) |
| Proline | Lognormal | 0.000 | (-0.034, 0.034) | 1.000 | 1.000 | 0.039 | (-0.035, 0.036) |
| **Complex Lipids (CL)** | | | | | | | |
| PC (p-34:2)/PC (o-34:3) - ESI(+) | Lognormal | **-0.022** | **(-0.032, -0.011)** | **0.000** | **0.048** | **36.107** | **(-0.032, -0.012)** |
| PE (p-36:2)/PE (o-36:3) - ESI(+) | Lognormal | **-0.033** | **(-0.049, -0.017)** | **0.000** | **0.048** | **24.602** | **(-0.049, -0.016)** |
| PC (p-34:1)/PC (o-34:2) | Lognormal | **-0.025** | **(-0.038, -0.012)** | **0.000** | **0.054** | **26.013** | **(-0.038, -0.012)** |
| PC (36:4) A - ESI(+) | Lognormal | **-0.022** | **(-0.033, -0.010)** | **0.000** | **0.054** | **8.061** | **(-0.034, -0.010)** |
| PC (36:2) | Lognormal | **-0.009** | **(-0.013, -0.004)** | **0.000** | **0.054** | **14.972** | **(-0.013, -0.004)** |
| PC 34:4e | Lognormal | **-0.029** | **(-0.045, -0.014)** | **0.000** | **0.054** | **13.135** | **(-0.044, -0.012)** |
| PC (36:4) A - ESI(-) | Lognormal | **-0.025** | **(-0.038, -0.011)** | **0.000** | **0.054** | **11.432** | **(-0.037, -0.010)** |
| PC (34:2) - ESI(+) | Lognormal | **-0.006** | **(-0.010, -0.003)** | **0.000** | **0.054** | **6.913** | **(-0.010, -0.003)** |
| PC (p-34:2)/PC (o-34:3) - ESI(-) | Lognormal | **-0.025** | **(-0.038, -0.011)** | **0.001** | **0.054** | **7.542** | **(-0.038, -0.010)** |
| PC (p-36:4)/PC (o-36:5) - ESI(-) | Lognormal | **-0.025** | **(-0.040, -0.011)** | **0.001** | **0.064** | **7.044** | **(-0.040, -0.011)** |
| PC (p-34:1)/PC (o-34:2) A | Lognormal | **-0.035** | **(-0.055, -0.015)** | **0.001** | **0.064** | **6.285** | **(-0.057, -0.015)** |
| PC (p-36:1)/PC (o-36:2) | Lognormal | **-0.067** | **(-0.106, -0.029)** | **0.001** | **0.064** | **5.458** | **(-0.107, -0.030)** |
| PE (p-34:2)/PE (o-34:3) | Lognormal | **-0.042** | **(-0.066, -0.018)** | **0.001** | **0.064** | **6.155** | **(-0.067, -0.020)** |
| SM (d40:3) | Lognormal | **-0.035** | **(-0.055, -0.014)** | **0.001** | **0.064** | **6.392** | **(-0.055, -0.015)** |
| PE (p-36:2)/PE (o-36:3) - ESI(-) | Lognormal | -0.032 | (-0.052, -0.012) | 0.002 | 0.125 | 2.051 | (-0.052, -0.011) |
| PE (p-36:4)/PE (o-36:5) - ESI(-) | Lognormal | -0.026 | (-0.044, -0.009) | 0.003 | 0.157 | 2.676 | (-0.044, -0.009) |
| CE (18:2) | Lognormal | -0.012 | (-0.019, -0.004) | 0.004 | 0.169 | 1.882 | (-0.020, -0.003) |
| PC (36:5) A | Lognormal | -0.060 | (-0.100, -0.019) | 0.005 | 0.169 | 2.389 | (-0.100, -0.017) |
| LPC (18:2) - ESI(-) | Lognormal | -0.022 | (-0.036, -0.007) | 0.005 | 0.169 | 2.295 | (-0.037, -0.007) |
| PC (34:2) - ESI(-) | Lognormal | -0.011 | (-0.018, -0.003) | 0.005 | 0.169 | 3.075 | (-0.017, -0.003) |
| PC (p-36:2)/PC (o-36:3) | Lognormal | -0.020 | (-0.034, -0.006) | 0.006 | 0.183 | 1.539 | (-0.034, -0.006) |
| TG (56:6) | Lognormal | 0.012 | (0.004, 0.021) | 0.006 | 0.183 | 2.004 | (0.004, 0.021) |
| PC (p-36:3)/PC (o-36:4) - ESI(-) | Lognormal | -0.017 | (-0.029, -0.005) | 0.007 | 0.192 | 1.692 | (-0.029, -0.004) |
| TG (55:6) | Lognormal | 0.026 | (0.008, 0.045) | 0.007 | 0.192 | 1.601 | (0.007, 0.044) |
| PC (36:5)A | Lognormal | -0.029 | (-0.050, -0.008) | 0.007 | 0.195 | 1.405 | (-0.049, -0.008) |
| SM (d34:2) - ESI(-) | Lognormal | -0.011 | (-0.018, -0.003) | 0.008 | 0.202 | 1.323 | (-0.019, -0.003) |
| SM (d34:1) - ESI(-) | Lognormal | -0.009 | (-0.016, -0.003) | 0.008 | 0.202 | 1.173 | (-0.017, -0.003) |
| LPE (18:2) - ESI(-) | Lognormal | -0.032 | (-0.055, -0.009) | 0.008 | 0.202 | 1.353 | (-0.057, -0.010) |
| TG (54:6) C | Lognormal | 0.015 | (0.004, 0.027) | 0.009 | 0.202 | 1.126 | (0.004, 0.027) |
| PC (34:3) | Lognormal | -0.019 | (-0.033, -0.005) | 0.009 | 0.202 | 1.163 | (-0.033, -0.005) |
| PE (p-38:5)/PE (o-38:6) - ESI(-) | Lognormal | -0.023 | (-0.040, -0.006) | 0.009 | 0.202 | 1.093 | (-0.041, -0.007) |
| LPE (18:2) - ESI(+) | Lognormal | -0.027 | (-0.046, -0.007) | 0.009 | 0.206 | 1.090 | (-0.046, -0.005) |
| SM (d38:2) - ESI(-) | Lognormal | -0.016 | (-0.028, -0.004) | 0.010 | 0.211 | 1.159 | (-0.028, -0.004) |
| LPC (18:2) - ESI(+) | Lognormal | -0.016 | (-0.028, -0.004) | 0.011 | 0.218 | 0.698 | (-0.029, -0.003) |
| PE (p-36:1)/PE (o-36:2) - ESI(-) | Lognormal | -0.028 | (-0.049, -0.007) | 0.011 | 0.218 | 0.873 | (-0.049, -0.007) |
| SM (d39:1) - ESI(-) | Lognormal | -0.016 | (-0.029, -0.004) | 0.011 | 0.218 | 1.051 | (-0.028, -0.004) |

**Supplementary Table S6.** Regression and Bayesian estimates comparing sex-stratified ME/CFS vs. controls.

| Metabolite | Regression Model | Female ME/CFS vs. Female Controls | | | | | |
|---|---|---|---|---|---|---|---|
| | | Estimated Coefficient | 95% CI | p-value | FDR | BayesFactor | 95% HDI |
| PC (32:2) - ESI(-) | Lognormal | -0.023 | (-0.041, -0.006) | 0.011 | 0.218 | 0.909 | (-0.041, -0.005) |
| PC (p-38:4)/PC (o-38:5) A | Lognormal | -0.014 | (-0.025, -0.003) | 0.012 | 0.218 | 0.854 | (-0.024, -0.003) |
| TG (54:7) B | Lognormal | 0.022 | (0.005, 0.040) | 0.013 | 0.225 | 1.036 | (0.006, 0.040) |
| PC (33:2) - ESI(+) | Lognormal | -0.013 | (-0.024, -0.003) | 0.013 | 0.225 | 0.837 | (-0.024, -0.003) |
| AC (10:1) | Lognormal | -0.029 | (-0.052, -0.006) | 0.013 | 0.225 | 1.070 | (-0.051, -0.007) |
| PC (34:3) B | Lognormal | -0.013 | (-0.024, -0.003) | 0.014 | 0.229 | 0.846 | (-0.023, -0.003) |
| LPC (18:3) | Lognormal | -0.025 | (-0.045, -0.005) | 0.014 | 0.229 | 0.729 | (-0.046, -0.006) |
| PE (p-38:4)/PE (o-38:5) | Lognormal | -0.015 | (-0.027, -0.003) | 0.015 | 0.239 | 0.663 | (-0.027, -0.003) |
| CE (20:2) | Lognormal | -0.030 | (-0.054, -0.006) | 0.016 | 0.240 | 0.750 | (-0.055, -0.007) |
| SM (d40:1) - ESI(-) | Lognormal | -0.011 | (-0.020, -0.002) | 0.016 | 0.242 | 0.682 | (-0.020, -0.002) |
| PC (o-34:0) | Lognormal | -0.019 | (-0.035, -0.004) | 0.017 | 0.248 | 0.675 | (-0.035, -0.004) |
| PC (35:2) | Lognormal | -0.015 | (-0.027, -0.003) | 0.017 | 0.249 | 0.672 | (-0.027, -0.002) |
| Ceramide (d42:1) - ESI (+) | Lognormal | -0.008 | (-0.015, -0.002) | 0.019 | 0.256 | 0.671 | (-0.016, -0.002) |
| FA (16:1) (palmitoleic acid) | Lognormal | 0.018 | (0.003, 0.033) | 0.019 | 0.256 | 0.598 | (0.003, 0.033) |
| PE (p-36:4)/PE (o-36:5) - ESI(+) | Lognormal | -0.017 | (-0.032, -0.003) | 0.019 | 0.256 | 0.515 | (-0.031, -0.003) |
| PC (32:2) - ESI(+) | Lognormal | -0.018 | (-0.033, -0.003) | 0.020 | 0.266 | 0.646 | (-0.033, -0.003) |
| TG (56:7) B | Lognormal | 0.024 | (0.004, 0.045) | 0.021 | 0.274 | 0.582 | (0.005, 0.046) |
| PC (p-38:3)/PC (o-38:4) - ESI(-) | Lognormal | -0.016 | (-0.029, -0.003) | 0.022 | 0.274 | 0.526 | (-0.030, -0.003) |
| SM (d40:2) B - ESI(+) | Lognormal | -0.008 | (-0.015, -0.001) | 0.022 | 0.274 | 0.652 | (-0.015, -0.002) |
| PE (p-40:4)/PE (o-40:5) A | Lognormal | -0.029 | (-0.053, -0.004) | 0.022 | 0.274 | 0.494 | (-0.053, -0.004) |
| TG (54:5) B | Lognormal | 0.013 | (0.002, 0.023) | 0.022 | 0.274 | 0.575 | (0.002, 0.023) |
| SM (d40:2) B - ESI(-) | Lognormal | -0.012 | (-0.023, -0.002) | 0.022 | 0.274 | 0.661 | (-0.023, -0.002) |
| TG (58:8) | Lognormal | 0.024 | (0.003, 0.046) | 0.025 | 0.291 | 0.522 | (0.003, 0.045) |
| PC (35:3) | Lognormal | -0.010 | (-0.019, -0.001) | 0.025 | 0.291 | 0.513 | (-0.019, -0.001) |
| PC (p-36:4)/PC (o-36:5) - ESI(+) | Lognormal | -0.010 | (-0.019, -0.001) | 0.025 | 0.291 | 0.592 | (-0.019, -0.002) |
| TG (56:5) B | Lognormal | 0.016 | (0.002, 0.029) | 0.028 | 0.312 | 0.405 | (0.002, 0.030) |
| TG (53:0) | Lognormal | -0.020 | (-0.037, -0.002) | 0.029 | 0.315 | 0.487 | (-0.038, -0.002) |
| SM (d33:1) - ESI(-) | Lognormal | -0.015 | (-0.028, -0.002) | 0.029 | 0.315 | 0.371 | (-0.029, -0.001) |
| PC (p-32:1)/PC (o-32:2) | Lognormal | -0.012 | (-0.023, -0.001) | 0.031 | 0.315 | 0.413 | (-0.023, -0.001) |
| TG (62:4) | Lognormal | -0.034 | (-0.064, -0.003) | 0.031 | 0.315 | 0.492 | (-0.063, -0.003) |
| PE (36:2) | Lognormal | -0.022 | (-0.042, -0.002) | 0.031 | 0.315 | 0.386 | (-0.042, -0.002) |
| PE (p-36:1)/PE (o-36:2) - ESI(+) | Lognormal | -0.021 | (-0.040, -0.002) | 0.031 | 0.315 | 0.392 | (-0.040, -0.002) |
| PE (p-34:1)/PE (o-34:2) - ESI(-) | Lognormal | -0.019 | (-0.036, -0.002) | 0.032 | 0.315 | 0.372 | (-0.036, -0.002) |
| TG (50:0) | Lognormal | -0.026 | (-0.050, -0.003) | 0.032 | 0.315 | 0.405 | (-0.048, -0.001) |
| SM (d32:1) - ESI(-) | Lognormal | -0.012 | (-0.023, -0.001) | 0.033 | 0.317 | 0.347 | (-0.023, -0.001) |
| LPE (20:4) - ESI(-) | Lognormal | -0.021 | (-0.040, -0.002) | 0.033 | 0.317 | 0.414 | (-0.040, -0.001) |
| PE (p-38:5)/PE (o-38:6) - ESI(+) | Lognormal | -0.013 | (-0.025, -0.001) | 0.033 | 0.317 | 0.381 | (-0.025, -0.001) |
| SM (d42:3) - ESI(-) | Lognormal | -0.009 | (-0.018, -0.001) | 0.034 | 0.320 | 0.349 | (-0.018, 0.000) |
| Ceramide (d41:1) - ESI(-) | Lognormal | -0.012 | (-0.024, -0.001) | 0.036 | 0.320 | 0.392 | (-0.024, -0.001) |
| PC (p-32:0)/PC (o-32:1) - ESI(+) | Lognormal | -0.009 | (-0.017, -0.001) | 0.036 | 0.320 | 0.355 | (-0.017, -0.001) |
| PC (36:4) B - ESI(-) | Lognormal | -0.008 | (-0.015, -0.001) | 0.037 | 0.322 | 0.357 | (-0.015, 0.000) |
| DG (38:5) | Lognormal | 0.015 | (0.001, 0.028) | 0.037 | 0.322 | 0.342 | (0.001, 0.028) |
| DG (38:6) | Lognormal | 0.020 | (0.001, 0.038) | 0.038 | 0.322 | 0.300 | (0.001, 0.038) |
| Ceramide (d42:2) B - ESI (+) | Lognormal | -0.010 | (-0.020, -0.001) | 0.039 | 0.324 | 0.338 | (-0.020, -0.001) |
| PC (34:3) C | Lognormal | -0.017 | (-0.033, -0.001) | 0.039 | 0.326 | 0.309 | (-0.032, -0.002) |
| TG (58:9) | Lognormal | 0.021 | (0.001, 0.042) | 0.042 | 0.337 | 0.285 | (0.000, 0.042) |
| PE (p-34:1)/PE (o-34:2) - ESI(+) | Lognormal | -0.016 | (-0.031, -0.001) | 0.042 | 0.338 | 0.310 | (-0.031, 0.000) |

**Supplementary Table S6.** Regression and Bayesian estimates comparing sex-stratified ME/CFS vs. controls.

| Metabolite | Regression Model | Female ME/CFS vs. Female Controls | | | | | |
|---|---|---|---|---|---|---|---|
| | | Estimated Coefficient | 95% CI | p-value | FDR | BayesFactor | 95% HDI |
| SM (d42:1) - ESI(-) | Lognormal | -0.009 | (-0.018, 0.000) | 0.043 | 0.338 | 0.304 | (-0.018, -0.001) |
| AC (18:2) | Lognormal | -0.018 | (-0.034, -0.001) | 0.043 | 0.338 | 0.292 | (-0.034, -0.001) |
| Ceramide (d42:1) - ESI(-) | Lognormal | -0.010 | (-0.019, 0.000) | 0.048 | 0.365 | 0.294 | (-0.020, -0.001) |
| Ceramide (d34:1) - ESI(-) | Lognormal | -0.008 | (-0.017, 0.000) | 0.048 | 0.365 | 0.255 | (-0.018, 0.000) |
| PC (p-38:4)/PC (o-38:5) B | Lognormal | -0.012 | (-0.023, 0.000) | 0.048 | 0.365 | 0.300 | (-0.024, -0.001) |
| CE (18:3) | Lognormal | -0.015 | (-0.030, 0.000) | 0.050 | 0.373 | 0.289 | (-0.030, 0.000) |
| AC (14:2) | Lognormal | -0.027 | (-0.054, 0.000) | 0.050 | 0.373 | 0.294 | (-0.056, -0.001) |
| PC (40:5) A - ESI(+) | Lognormal | 0.010 | (0.000, 0.019) | 0.052 | 0.376 | 0.263 | (0.000, 0.019) |
| PC (33:2) - ESI(-) | Lognormal | -0.018 | (-0.037, 0.000) | 0.052 | 0.376 | 0.258 | (-0.035, 0.002) |
| PC (37:2) - ESI(+) | Lognormal | -0.010 | (-0.021, 0.000) | 0.052 | 0.376 | 0.286 | (-0.020, 0.000) |
| LPC (14:0) - ESI(+) | Lognormal | -0.017 | (-0.034, 0.000) | 0.054 | 0.379 | 0.266 | (-0.034, 0.001) |
| TG (54:6) B | Lognormal | -0.039 | (-0.078, 0.000) | 0.054 | 0.379 | 0.232 | (-0.077, 0.003) |
| SM (d36:3) - ESI(-) | Lognormal | -0.018 | (-0.037, 0.000) | 0.056 | 0.380 | 0.234 | (-0.038, 0.000) |
| FA (18:1) (oleic acid) | Lognormal | 0.009 | (0.000, 0.017) | 0.056 | 0.380 | 0.229 | (0.000, 0.017) |
| PC (p-36:3)/PC (o-36:4) - ESI(+) | Lognormal | -0.009 | (-0.019, 0.000) | 0.057 | 0.380 | 0.248 | (-0.019, 0.000) |
| FA (14:1) (physeteric acid) | Lognormal | 0.019 | (0.000, 0.038) | 0.058 | 0.383 | 0.238 | (0.000, 0.038) |
| PC (34:0) - ESI(-) | Lognormal | -0.009 | (-0.019, 0.000) | 0.060 | 0.390 | 0.227 | (-0.019, 0.001) |
| Ceramide (d40:1) | Lognormal | -0.008 | (-0.016, 0.000) | 0.064 | 0.399 | 0.230 | (-0.016, 0.001) |
| SM (d36:0) - ESI(+) | Lognormal | 0.015 | (-0.001, 0.030) | 0.064 | 0.399 | 0.238 | (-0.001, 0.030) |
| LPC (16:0) - ESI(-) | Lognormal | -0.009 | (-0.018, 0.000) | 0.064 | 0.399 | 0.221 | (-0.017, 0.001) |
| PE (p-40:5)/PE (o-40:6) | Lognormal | -0.010 | (-0.021, 0.001) | 0.066 | 0.407 | 0.213 | (-0.022, 0.000) |
| LPC (18:0) A - ESI(-) | Lognormal | -0.015 | (-0.032, 0.001) | 0.069 | 0.414 | 0.220 | (-0.031, 0.002) |
| PC (38:4) A - ESI(-) | Lognormal | -0.009 | (-0.019, 0.001) | 0.069 | 0.416 | 0.227 | (-0.019, 0.001) |
| TG (54:7) A | Lognormal | -0.023 | (-0.048, 0.002) | 0.070 | 0.416 | 0.198 | (-0.048, 0.001) |
| TG (56:8) B | Lognormal | 0.022 | (-0.002, 0.045) | 0.072 | 0.419 | 0.190 | (-0.001, 0.046) |
| PE (36:3) | Lognormal | -0.027 | (-0.057, 0.002) | 0.075 | 0.429 | 0.228 | (-0.056, 0.003) |
| PC (37:4) - ESI(+) | Lognormal | 0.011 | (-0.001, 0.024) | 0.076 | 0.430 | 0.209 | (-0.001, 0.024) |
| PC (34:4) - ESI(-) | Lognormal | -0.024 | (-0.051, 0.002) | 0.077 | 0.434 | 0.191 | (-0.052, 0.000) |
| SM (d36:1) - ESI(-) | Lognormal | -0.008 | (-0.017, 0.001) | 0.077 | 0.435 | 0.176 | (-0.016, 0.001) |
| AC (12:1) | Lognormal | -0.020 | (-0.043, 0.002) | 0.080 | 0.443 | 0.205 | (-0.042, 0.001) |
| Cholesterol | Lognormal | -0.006 | (-0.014, 0.001) | 0.082 | 0.444 | 0.195 | (-0.014, 0.001) |
| PC (p-38:3)/PC (o-38:4) A - ESI(+) | Lognormal | -0.010 | (-0.021, 0.001) | 0.083 | 0.444 | 0.180 | (-0.021, 0.001) |
| PC (p-38:3)/PC (o-38:4) B - ESI(+) | Lognormal | -0.010 | (-0.022, 0.001) | 0.086 | 0.444 | 0.158 | (-0.021, 0.002) |
| AC (10:0) | Lognormal | -0.030 | (-0.064, 0.004) | 0.086 | 0.444 | 0.139 | (-0.067, 0.004) |
| TG (60:11) | Lognormal | 0.030 | (-0.004, 0.065) | 0.086 | 0.444 | 0.176 | (-0.005, 0.064) |
| PC (p-38:4)/PC (o-38:5) B | Lognormal | -0.018 | (-0.038, 0.003) | 0.089 | 0.451 | 0.185 | (-0.037, 0.003) |
| TG (52:4) | Lognormal | -0.009 | (-0.020, 0.001) | 0.092 | 0.457 | 0.177 | (-0.019, 0.001) |
| LPC (18:0) B - ESI(-) | Lognormal | -0.010 | (-0.022, 0.002) | 0.092 | 0.457 | 0.133 | (-0.022, 0.001) |
| SM (d41:2) B - ESI(+) | Lognormal | -0.008 | (-0.017, 0.001) | 0.092 | 0.457 | 0.151 | (-0.017, 0.001) |
| TG (58:6) | Lognormal | 0.011 | (-0.002, 0.023) | 0.093 | 0.457 | 0.176 | (-0.001, 0.024) |
| PC (p-36:1)/PC (o-36:2) B | Lognormal | -0.014 | (-0.031, 0.002) | 0.093 | 0.457 | 0.158 | (-0.031, 0.002) |
| PC (37:5) | Lognormal | 0.017 | (-0.003, 0.037) | 0.094 | 0.457 | 0.154 | (-0.004, 0.038) |
| PC (36:5) D | Lognormal | 0.018 | (-0.003, 0.040) | 0.095 | 0.461 | 0.150 | (-0.004, 0.039) |
| SM (d40:1) - ESI(+) | Lognormal | -0.005 | (-0.011, 0.001) | 0.096 | 0.464 | 0.165 | (-0.011, 0.001) |
| PC (35:2) B | Lognormal | -0.006 | (-0.013, 0.001) | 0.097 | 0.465 | 0.177 | (-0.014, 0.001) |
| PC (36:3) A - ESI(+) | Lognormal | -0.007 | (-0.014, 0.001) | 0.097 | 0.465 | 0.161 | (-0.015, 0.001) |
| PC (p-32:0)/PC (o-32:1) - ESI(-) | Lognormal | -0.014 | (-0.030, 0.002) | 0.099 | 0.465 | 0.153 | (-0.031, 0.002) |

**Supplementary Table S6.** Regression and Bayesian estimates comparing sex-stratified ME/CFS vs. controls.

| Metabolite | Regression Model | Female ME/CFS vs. Female Controls | | | | | |
|---|---|---|---|---|---|---|---|
| | | Estimated Coefficient | 95% CI | p-value | FDR | BayesFactor | 95% HDI |
| SM (d43:1) - ESI(-) | Lognormal | -0.024 | (-0.053, 0.004) | 0.100 | 0.471 | 0.149 | (-0.053, 0.006) |
| PC (40:6)B | Lognormal | 0.010 | (-0.002, 0.021) | 0.103 | 0.480 | 0.153 | (-0.002, 0.022) |
| TG (48:0) | Lognormal | -0.019 | (-0.041, 0.004) | 0.103 | 0.480 | 0.153 | (-0.040, 0.005) |
| SM (d42:0) - ESI(+) | Lognormal | 0.023 | (-0.005, 0.051) | 0.108 | 0.492 | 0.139 | (-0.006, 0.051) |
| SM (d30:1) - ESI(-) | Lognormal | -0.023 | (-0.051, 0.005) | 0.109 | 0.492 | 0.154 | (-0.050, 0.005) |
| PC (36:3) B - ESI(-) | Lognormal | -0.008 | (-0.017, 0.002) | 0.109 | 0.492 | 0.129 | (-0.016, 0.002) |
| Ceramide (d32:1) | Lognormal | -0.012 | (-0.027, 0.003) | 0.109 | 0.492 | 0.141 | (-0.026, 0.003) |
| PE (p-40:4)/PE (o-40:5) B | Lognormal | -0.013 | (-0.028, 0.003) | 0.110 | 0.493 | 0.106 | (-0.027, 0.004) |
| DG (36:4) B | Lognormal | 0.034 | (-0.008, 0.076) | 0.112 | 0.494 | 0.145 | (-0.008, 0.077) |
| TG (48:4) A | Lognormal | -0.023 | (-0.052, 0.005) | 0.114 | 0.501 | 0.131 | (-0.054, 0.005) |
| PC (34:3) A | Lognormal | -0.008 | (-0.018, 0.002) | 0.114 | 0.501 | 0.135 | (-0.018, 0.002) |
| PC (35:1) - ESI(+) | Lognormal | 0.007 | (-0.002, 0.015) | 0.115 | 0.501 | 0.130 | (-0.002, 0.014) |
| LPC (p-16:0)/LPC (o-16:1) | Lognormal | -0.011 | (-0.025, 0.003) | 0.116 | 0.502 | 0.134 | (-0.026, 0.002) |
| PC (p-38:5)/PC (o-38:6) B | Lognormal | -0.007 | (-0.017, 0.002) | 0.120 | 0.510 | 0.138 | (-0.017, 0.002) |
| TG (49:2) | Lognormal | 0.017 | (-0.004, 0.039) | 0.121 | 0.512 | 0.128 | (-0.005, 0.038) |
| LPC (20:3) - ESI(-) | Lognormal | -0.017 | (-0.038, 0.004) | 0.121 | 0.512 | 0.127 | (-0.039, 0.004) |
| Ceramide (d41:1) - ESI (+) | Lognormal | -0.008 | (-0.018, 0.002) | 0.122 | 0.512 | 0.135 | (-0.018, 0.002) |
| AC (12:0) | Lognormal | -0.018 | (-0.041, 0.005) | 0.123 | 0.513 | 0.124 | (-0.041, 0.005) |
| PC (36:3) A - ESI(-) | Lognormal | -0.008 | (-0.019, 0.002) | 0.124 | 0.515 | 0.119 | (-0.018, 0.002) |
| FA (20:3) (homo-gamma-linolenic acid) | Lognormal | 0.008 | (-0.002, 0.018) | 0.124 | 0.515 | 0.125 | (-0.002, 0.017) |
| PC (42:10) | Lognormal | 0.019 | (-0.005, 0.043) | 0.126 | 0.519 | 0.133 | (-0.006, 0.042) |
| Lactosylceramide (d18:1/24:1(15Z)) | Lognormal | 0.013 | (-0.004, 0.030) | 0.127 | 0.520 | 0.125 | (-0.004, 0.030) |
| TG (60:6) | Lognormal | 0.019 | (-0.005, 0.043) | 0.129 | 0.521 | 0.110 | (-0.006, 0.042) |
| PE (p-40:4)/PE (o-40:5) | Lognormal | -0.014 | (-0.031, 0.004) | 0.130 | 0.521 | 0.093 | (-0.030, 0.004) |
| PC (38:5) A | Lognormal | 0.005 | (-0.001, 0.011) | 0.130 | 0.521 | 0.125 | (-0.001, 0.012) |
| PC (34:1) | Lognormal | 0.003 | (-0.001, 0.007) | 0.130 | 0.521 | 0.109 | (-0.001, 0.007) |
| TG (56:5) A | Lognormal | 0.012 | (-0.003, 0.027) | 0.131 | 0.521 | 0.135 | (-0.003, 0.027) |
| DG (36:4) A | Lognormal | -0.015 | (-0.035, 0.004) | 0.132 | 0.521 | 0.118 | (-0.035, 0.004) |
| AC (8:0) | Lognormal | -0.026 | (-0.060, 0.008) | 0.133 | 0.521 | 0.123 | (-0.059, 0.008) |
| Ceramide (d39:1) | Lognormal | -0.014 | (-0.032, 0.004) | 0.133 | 0.521 | 0.120 | (-0.032, 0.004) |
| PC (38:6) A - ESI(+) | Lognormal | -0.007 | (-0.016, 0.002) | 0.134 | 0.521 | 0.117 | (-0.016, 0.002) |
| PC (38:5) B - ESI(+) | Lognormal | 0.010 | (-0.003, 0.024) | 0.135 | 0.523 | 0.120 | (-0.002, 0.024) |
| SM (d40:0) | Lognormal | 0.011 | (-0.003, 0.025) | 0.138 | 0.529 | 0.102 | (-0.004, 0.025) |
| PC (o-32:0) - ESI(+) | Lognormal | -0.006 | (-0.015, 0.002) | 0.144 | 0.535 | 0.110 | (-0.015, 0.002) |
| PE (p-38:3)/PE (o-38:4) | Lognormal | -0.022 | (-0.052, 0.007) | 0.145 | 0.535 | 0.119 | (-0.053, 0.007) |
| TG (56:9) | Lognormal | 0.016 | (-0.005, 0.037) | 0.145 | 0.535 | 0.116 | (-0.006, 0.038) |
| TG (50:4) | Lognormal | -0.012 | (-0.028, 0.004) | 0.147 | 0.535 | 0.114 | (-0.028, 0.004) |
| PC (p-34:1)/PC (o-34:2) B | Lognormal | -0.016 | (-0.039, 0.006) | 0.150 | 0.543 | 0.118 | (-0.041, 0.005) |
| SM (d34:0) - ESI(-) | Lognormal | -0.011 | (-0.027, 0.004) | 0.154 | 0.551 | 0.096 | (-0.025, 0.005) |
| PE (p-38:6)/PE (o-38:7) | Lognormal | -0.015 | (-0.035, 0.006) | 0.163 | 0.568 | 0.091 | (-0.034, 0.007) |
| SM (d38:1) | Lognormal | -0.004 | (-0.011, 0.002) | 0.170 | 0.589 | 0.100 | (-0.010, 0.002) |
| FA (24:0) (lignoceric acid) | Lognormal | -0.009 | (-0.021, 0.004) | 0.172 | 0.589 | 0.102 | (-0.022, 0.003) |
| SM (d34:1) - ESI(+) | Lognormal | -0.004 | (-0.009, 0.002) | 0.173 | 0.591 | 0.094 | (-0.008, 0.001) |
| CE (20:3) | Lognormal | -0.009 | (-0.021, 0.004) | 0.174 | 0.591 | 0.097 | (-0.021, 0.004) |
| LPC (20:2) - ESI(+) | Lognormal | -0.025 | (-0.061, 0.011) | 0.175 | 0.591 | 0.106 | (-0.061, 0.012) |
| PC (42:6) | Lognormal | 0.018 | (-0.008, 0.044) | 0.177 | 0.591 | 0.099 | (-0.009, 0.042) |
| DG (32:0) | Lognormal | 0.012 | (-0.005, 0.029) | 0.179 | 0.593 | 0.094 | (-0.007, 0.028) |

**Supplementary Table S6.** Regression and Bayesian estimates comparing sex-stratified ME/CFS vs. controls.

| Metabolite | Regression Model | Female ME/CFS vs. Female Controls | | | | | |
|---|---|---|---|---|---|---|---|
| | | Estimated Coefficient | 95% CI | p-value | FDR | BayesFactor | 95% HDI |
| CE (20:5) | Lognormal | 0.018 | (-0.008, 0.045) | 0.181 | 0.596 | 0.094 | (-0.009, 0.045) |
| PC (o-32:0) - ESI(-) | Lognormal | -0.010 | (-0.024, 0.004) | 0.182 | 0.596 | 0.088 | (-0.024, 0.005) |
| PC (32:0) - ESI(-) | Lognormal | -0.006 | (-0.016, 0.003) | 0.182 | 0.596 | 0.078 | (-0.016, 0.003) |
| PC (40:8) - ESI(-) | Lognormal | -0.012 | (-0.029, 0.005) | 0.183 | 0.596 | 0.085 | (-0.027, 0.006) |
| Ceramide (d38:1) - ESI(+) | Lognormal | -0.007 | (-0.016, 0.003) | 0.184 | 0.596 | 0.088 | (-0.015, 0.004) |
| Ceramide (d43:1) | Lognormal | -0.019 | (-0.046, 0.009) | 0.189 | 0.601 | 0.092 | (-0.049, 0.008) |
| SM (d42:1) - ESI(+) | Lognormal | -0.005 | (-0.013, 0.003) | 0.190 | 0.601 | 0.098 | (-0.013, 0.003) |
| PC (37:2) - ESI(-) | Lognormal | -0.017 | (-0.041, 0.008) | 0.190 | 0.601 | 0.085 | (-0.042, 0.008) |
| PC (38:3) - ESI(-) | Lognormal | -0.008 | (-0.019, 0.004) | 0.193 | 0.605 | 0.087 | (-0.020, 0.004) |
| SM (d39:2) | Lognormal | -0.006 | (-0.015, 0.003) | 0.195 | 0.605 | 0.092 | (-0.015, 0.003) |
| SM (d30:1) - ESI(+) | Lognormal | -0.014 | (-0.035, 0.007) | 0.195 | 0.605 | 0.102 | (-0.036, 0.007) |
| LPC (o-16:0) | Lognormal | -0.010 | (-0.025, 0.005) | 0.196 | 0.605 | 0.088 | (-0.025, 0.006) |
| LPC (20:2) - ESI(-) | Lognormal | -0.009 | (-0.024, 0.005) | 0.197 | 0.605 | 0.091 | (-0.023, 0.006) |
| LPC (18:1) - ESI(-) | Lognormal | -0.009 | (-0.022, 0.005) | 0.197 | 0.605 | 0.085 | (-0.022, 0.005) |
| LPC (20:3) - ESI(+) | Lognormal | -0.009 | (-0.022, 0.004) | 0.198 | 0.605 | 0.084 | (-0.022, 0.004) |
| PC (38:6) B - ESI(+) | Lognormal | 0.005 | (-0.002, 0.012) | 0.198 | 0.605 | 0.091 | (-0.002, 0.012) |
| FA (20:3) (eicosatrienoic acid) | Lognormal | 0.011 | (-0.006, 0.029) | 0.203 | 0.610 | 0.081 | (-0.006, 0.029) |
| PC (p-44:4)/PC (o-44:5) - ESI(-) | Lognormal | -0.010 | (-0.026, 0.005) | 0.203 | 0.610 | 0.082 | (-0.025, 0.005) |
| CE (14:0) | Lognormal | -0.025 | (-0.064, 0.014) | 0.205 | 0.610 | 0.091 | (-0.063, 0.014) |
| TG (56:8) A | Lognormal | 0.010 | (-0.005, 0.024) | 0.207 | 0.610 | 0.085 | (-0.005, 0.025) |
| Ceramide (d33:1) | Lognormal | -0.009 | (-0.024, 0.005) | 0.208 | 0.610 | 0.085 | (-0.023, 0.005) |
| TG (46:0) | Lognormal | -0.024 | (-0.062, 0.014) | 0.211 | 0.610 | 0.082 | (-0.062, 0.015) |
| PE (38:6) - ESI(+) | Lognormal | 0.021 | (-0.012, 0.055) | 0.213 | 0.610 | 0.083 | (-0.010, 0.056) |
| SM (d34:2) - ESI(+) | Lognormal | -0.004 | (-0.011, 0.002) | 0.213 | 0.610 | 0.086 | (-0.011, 0.003) |
| AC (18:0) | Lognormal | -0.009 | (-0.023, 0.005) | 0.214 | 0.610 | 0.083 | (-0.023, 0.005) |
| TG (51:2) | Lognormal | 0.010 | (-0.006, 0.025) | 0.217 | 0.615 | 0.085 | (-0.006, 0.025) |
| LPC (14:0) - ESI(-) | Lognormal | -0.019 | (-0.049, 0.011) | 0.218 | 0.617 | 0.083 | (-0.050, 0.011) |
| TG (54:6) A | Lognormal | -0.012 | (-0.030, 0.007) | 0.222 | 0.622 | 0.077 | (-0.031, 0.006) |
| FA (22:0) (behenic acid) | Lognormal | -0.009 | (-0.022, 0.005) | 0.223 | 0.623 | 0.072 | (-0.022, 0.005) |
| Ceramide (d42:2) B - ESI(-) | Lognormal | -0.014 | (-0.035, 0.008) | 0.224 | 0.624 | 0.086 | (-0.037, 0.007) |
| SM (d39:1) - ESI(+) | Lognormal | -0.007 | (-0.017, 0.004) | 0.226 | 0.624 | 0.083 | (-0.017, 0.004) |
| TG (50:5) | Lognormal | -0.014 | (-0.037, 0.009) | 0.226 | 0.624 | 0.090 | (-0.036, 0.008) |
| TG (46:4) A | Lognormal | -0.025 | (-0.066, 0.016) | 0.230 | 0.624 | 0.075 | (-0.068, 0.013) |
| GlcCer (d42:2) - ESI(+) | Lognormal | 0.009 | (-0.005, 0.023) | 0.230 | 0.624 | 0.084 | (-0.005, 0.024) |
| LPC (16:1) - ESI(-) | Lognormal | -0.011 | (-0.029, 0.007) | 0.231 | 0.624 | 0.079 | (-0.030, 0.007) |
| PC (35:2) A | Lognormal | 0.006 | (-0.004, 0.016) | 0.233 | 0.629 | 0.069 | (-0.004, 0.016) |
| TG (51:4) | Lognormal | -0.010 | (-0.025, 0.006) | 0.236 | 0.633 | 0.077 | (-0.026, 0.005) |
| PC (38:2) | Lognormal | -0.008 | (-0.022, 0.005) | 0.237 | 0.634 | 0.073 | (-0.022, 0.004) |
| PC 40:5e | Lognormal | -0.006 | (-0.017, 0.004) | 0.239 | 0.636 | 0.076 | (-0.016, 0.005) |
| PC (37:4) - ESI(-) | Lognormal | -0.010 | (-0.025, 0.006) | 0.240 | 0.636 | 0.080 | (-0.026, 0.005) |
| SM (d36:3) - ESI(+) | Lognormal | -0.008 | (-0.022, 0.006) | 0.241 | 0.636 | 0.073 | (-0.022, 0.005) |
| PE (34:2) - ESI(-) | Lognormal | -0.017 | (-0.047, 0.012) | 0.242 | 0.636 | 0.084 | (-0.047, 0.011) |
| TG (48:5) | Lognormal | -0.021 | (-0.055, 0.014) | 0.243 | 0.636 | 0.071 | (-0.055, 0.014) |
| GlcCer (d38:1) | Lognormal | -0.008 | (-0.022, 0.006) | 0.244 | 0.636 | 0.069 | (-0.022, 0.005) |
| PC (p-42:4)/PC (o-42:5) - ESI(-) | Lognormal | -0.013 | (-0.034, 0.009) | 0.244 | 0.636 | 0.075 | (-0.035, 0.009) |
| TG (54:9) | Lognormal | 0.039 | (-0.026, 0.104) | 0.245 | 0.636 | 0.065 | (-0.027, 0.101) |
| LPE (18:0) | Lognormal | -0.009 | (-0.024, 0.006) | 0.251 | 0.637 | 0.069 | (-0.023, 0.007) |

**Supplementary Table S6.** Regression and Bayesian estimates comparing sex-stratified ME/CFS vs. controls.

| Metabolite | Regression Model | Female ME/CFS vs. Female Controls | | | | | |
|---|---|---|---|---|---|---|---|
| | | Estimated Coefficient | 95% CI | p-value | FDR | BayesFactor | 95% HDI |
| TG (58:4) | Lognormal | 0.015 | (-0.010, 0.039) | 0.252 | 0.637 | 0.085 | (-0.012, 0.039) |
| PC (34:4) - ESI(+) | Lognormal | -0.010 | (-0.028, 0.007) | 0.254 | 0.637 | 0.071 | (-0.027, 0.007) |
| TG (42:0) | Lognormal | -0.023 | (-0.063, 0.017) | 0.257 | 0.642 | 0.074 | (-0.062, 0.015) |
| TG (53:2) | Lognormal | 0.008 | (-0.006, 0.022) | 0.258 | 0.642 | 0.079 | (-0.006, 0.022) |
| PC (p-38:4)/PC (o-38:5) A | Lognormal | -0.004 | (-0.012, 0.003) | 0.259 | 0.642 | 0.066 | (-0.012, 0.003) |
| TG (46:3) A | Lognormal | -0.026 | (-0.071, 0.019) | 0.262 | 0.642 | 0.069 | (-0.070, 0.017) |
| TG (56:5) C | Lognormal | 0.007 | (-0.006, 0.020) | 0.265 | 0.642 | 0.068 | (-0.005, 0.020) |
| PC (36:1) - ESI(+) | Lognormal | 0.004 | (-0.003, 0.011) | 0.265 | 0.642 | 0.073 | (-0.003, 0.010) |
| FA (20:1) (eicosenoic acid) | Lognormal | 0.006 | (-0.005, 0.017) | 0.266 | 0.644 | 0.079 | (-0.005, 0.017) |
| PC (37:6) | Lognormal | 0.012 | (-0.009, 0.033) | 0.268 | 0.647 | 0.075 | (-0.010, 0.032) |
| LPE (22:6) | Lognormal | -0.012 | (-0.032, 0.009) | 0.270 | 0.649 | 0.069 | (-0.033, 0.009) |
| SM (d40:2) A - ESI(+) | Lognormal | -0.008 | (-0.021, 0.006) | 0.272 | 0.650 | 0.077 | (-0.021, 0.006) |
| LPC (20:5) | Lognormal | 0.018 | (-0.014, 0.049) | 0.274 | 0.651 | 0.071 | (-0.011, 0.051) |
| SM (d36:2) - ESI(-) | Lognormal | -0.007 | (-0.020, 0.006) | 0.279 | 0.655 | 0.073 | (-0.020, 0.007) |
| DG (36:5) | Lognormal | -0.014 | (-0.038, 0.011) | 0.279 | 0.655 | 0.071 | (-0.037, 0.012) |
| PC (42:5) | Lognormal | 0.009 | (-0.007, 0.024) | 0.282 | 0.657 | 0.074 | (-0.007, 0.024) |
| SM (d44:2) | Lognormal | 0.008 | (-0.007, 0.024) | 0.286 | 0.657 | 0.065 | (-0.007, 0.023) |
| LPE (16:0) | Lognormal | -0.013 | (-0.037, 0.011) | 0.286 | 0.657 | 0.067 | (-0.039, 0.011) |
| SM (d32:1) - ESI(+) | Lognormal | -0.006 | (-0.017, 0.005) | 0.287 | 0.657 | 0.065 | (-0.018, 0.005) |
| SM (d41:3) | Lognormal | -0.005 | (-0.015, 0.004) | 0.289 | 0.657 | 0.069 | (-0.015, 0.004) |
| TG (42:1) | Lognormal | -0.036 | (-0.101, 0.030) | 0.289 | 0.657 | 0.062 | (-0.099, 0.030) |
| TG (60:12) | Lognormal | 0.024 | (-0.020, 0.068) | 0.291 | 0.659 | 0.068 | (-0.020, 0.070) |
| PC (p-40:3)/PC (o-40:4) | Lognormal | -0.011 | (-0.031, 0.009) | 0.297 | 0.668 | 0.063 | (-0.030, 0.010) |
| PC (40:4) - ESI(-) | Lognormal | -0.011 | (-0.032, 0.010) | 0.298 | 0.668 | 0.062 | (-0.033, 0.010) |
| FA (20:5) (eicosapentaenoic acid) | Lognormal | 0.015 | (-0.014, 0.044) | 0.299 | 0.668 | 0.063 | (-0.011, 0.048) |
| DG (36:2) | Lognormal | 0.007 | (-0.006, 0.020) | 0.305 | 0.676 | 0.067 | (-0.007, 0.019) |
| AC (14:1) | Lognormal | -0.013 | (-0.038, 0.012) | 0.309 | 0.676 | 0.063 | (-0.039, 0.010) |
| SM (d40:2) A - ESI(-) | Lognormal | -0.009 | (-0.027, 0.009) | 0.312 | 0.676 | 0.067 | (-0.027, 0.009) |
| DG (36:1) | Lognormal | 0.009 | (-0.009, 0.027) | 0.312 | 0.676 | 0.064 | (-0.009, 0.027) |
| LPC (17:1) | Lognormal | 0.009 | (-0.008, 0.025) | 0.314 | 0.676 | 0.063 | (-0.007, 0.026) |
| PG (34:0)/PG (17:0/17:0) | Lognormal | -0.011 | (-0.032, 0.010) | 0.314 | 0.676 | 0.061 | (-0.032, 0.010) |
| SM (d43:2) - ESI(-) | Lognormal | -0.011 | (-0.032, 0.010) | 0.314 | 0.676 | 0.063 | (-0.034, 0.010) |
| SM (d36:0) - ESI(-) | Lognormal | 0.016 | (-0.015, 0.048) | 0.316 | 0.678 | 0.063 | (-0.016, 0.048) |
| SM (d38:0) | Lognormal | 0.036 | (-0.035, 0.108) | 0.321 | 0.684 | 0.065 | (-0.034, 0.109) |
| TG (42:3) | Lognormal | -0.021 | (-0.062, 0.020) | 0.323 | 0.684 | 0.055 | (-0.064, 0.019) |
| Ceramide (d44:1) | Lognormal | -0.014 | (-0.040, 0.013) | 0.325 | 0.684 | 0.068 | (-0.039, 0.015) |
| PC (36:3) B - ESI(+) | Lognormal | -0.003 | (-0.010, 0.003) | 0.326 | 0.684 | 0.062 | (-0.011, 0.003) |
| TG (40:1) | Lognormal | -0.022 | (-0.066, 0.022) | 0.327 | 0.684 | 0.058 | (-0.066, 0.023) |
| Ceramide (d38:1) - ESI(-) | Lognormal | -0.007 | (-0.020, 0.007) | 0.327 | 0.684 | 0.060 | (-0.021, 0.006) |
| PC (33:1) - ESI(+) | Lognormal | 0.006 | (-0.006, 0.017) | 0.333 | 0.690 | 0.061 | (-0.005, 0.016) |
| TG (48:3) | Lognormal | -0.011 | (-0.034, 0.011) | 0.333 | 0.690 | 0.059 | (-0.034, 0.011) |
| Ceramide (d42:2) A - ESI(-) | Lognormal | -0.005 | (-0.015, 0.005) | 0.333 | 0.690 | 0.063 | (-0.015, 0.005) |
| SM (d32:2) - ESI(+) | Lognormal | -0.006 | (-0.019, 0.006) | 0.335 | 0.691 | 0.061 | (-0.019, 0.006) |
| TG (44:0) | Lognormal | -0.018 | (-0.054, 0.018) | 0.336 | 0.691 | 0.057 | (-0.056, 0.019) |
| TG (54:5) A | Lognormal | -0.007 | (-0.021, 0.007) | 0.337 | 0.691 | 0.058 | (-0.020, 0.007) |
| TG (52:2) | Lognormal | 0.004 | (-0.004, 0.012) | 0.337 | 0.691 | 0.062 | (-0.004, 0.012) |
| PI (38:4)/PI (18:0-20:4) | Lognormal | -0.004 | (-0.013, 0.004) | 0.340 | 0.691 | 0.061 | (-0.012, 0.004) |

**Supplementary Table S6.** Regression and Bayesian estimates comparing sex-stratified ME/CFS vs. controls.

| Metabolite | Regression Model | Female ME/CFS vs. Female Controls | | | | | |
|---|---|---|---|---|---|---|---|
| | | Estimated Coefficient | 95% CI | p-value | FDR | BayesFactor | 95% HDI |
| PC (37:3) | Lognormal | -0.007 | (-0.022, 0.008) | 0.340 | 0.691 | 0.064 | (-0.022, 0.007) |
| TG (56:3) | Lognormal | 0.010 | (-0.011, 0.031) | 0.342 | 0.691 | 0.068 | (-0.011, 0.031) |
| SM (d32:2) - ESI(-) | Lognormal | -0.010 | (-0.029, 0.010) | 0.343 | 0.691 | 0.059 | (-0.028, 0.011) |
| TG (46:4) B | Lognormal | -0.024 | (-0.074, 0.025) | 0.343 | 0.691 | 0.060 | (-0.078, 0.022) |
| TG (46:2) | Lognormal | -0.016 | (-0.049, 0.017) | 0.345 | 0.691 | 0.058 | (-0.050, 0.016) |
| PC (p-40:1)/PC (o-40:2) | Lognormal | 0.011 | (-0.011, 0.033) | 0.350 | 0.697 | 0.061 | (-0.014, 0.031) |
| PE (34:1) | Lognormal | -0.015 | (-0.046, 0.016) | 0.354 | 0.701 | 0.061 | (-0.049, 0.016) |
| SM (d41:1) - ESI(+) | Lognormal | -0.004 | (-0.012, 0.004) | 0.355 | 0.701 | 0.062 | (-0.012, 0.005) |
| SM (d38:2) - ESI(+) | Lognormal | -0.003 | (-0.010, 0.004) | 0.355 | 0.701 | 0.059 | (-0.011, 0.004) |
| LPC (15:0) | Lognormal | -0.007 | (-0.021, 0.008) | 0.361 | 0.704 | 0.052 | (-0.021, 0.008) |
| PC (p-40:7)/PC (o-40:8) | Lognormal | -0.020 | (-0.062, 0.022) | 0.363 | 0.704 | 0.060 | (-0.061, 0.023) |
| PC (p-42:3)/PC (o-42:4) | Lognormal | -0.007 | (-0.021, 0.008) | 0.363 | 0.704 | 0.053 | (-0.021, 0.008) |
| PE (36:4) - ESI(+) | Lognormal | 0.010 | (-0.012, 0.033) | 0.366 | 0.705 | 0.057 | (-0.013, 0.033) |
| SM (d42:3) - ESI(+) | Lognormal | -0.003 | (-0.010, 0.004) | 0.366 | 0.705 | 0.061 | (-0.011, 0.004) |
| GlcCer (d41:1) | Lognormal | -0.006 | (-0.020, 0.007) | 0.369 | 0.709 | 0.053 | (-0.020, 0.008) |
| TG (58:3) | Lognormal | 0.014 | (-0.016, 0.043) | 0.371 | 0.711 | 0.058 | (-0.018, 0.041) |
| TG (48:4) B | Lognormal | -0.019 | (-0.060, 0.022) | 0.375 | 0.711 | 0.055 | (-0.059, 0.022) |
| PC (39:6) | Lognormal | 0.008 | (-0.010, 0.026) | 0.376 | 0.711 | 0.057 | (-0.010, 0.026) |
| PE (p-36:5)/PE (o-36:6) | Lognormal | -0.030 | (-0.096, 0.036) | 0.376 | 0.711 | 0.055 | (-0.098, 0.035) |
| PC (36:4) B - ESI(+) | Lognormal | 0.007 | (-0.008, 0.022) | 0.377 | 0.711 | 0.058 | (-0.008, 0.023) |
| TG (54:1) | Lognormal | -0.010 | (-0.032, 0.012) | 0.384 | 0.719 | 0.053 | (-0.030, 0.014) |
| FA (18:2) (linoleic acid) | Lognormal | 0.004 | (-0.005, 0.012) | 0.392 | 0.728 | 0.051 | (-0.005, 0.012) |
| LPE (20:4) - ESI(+) | Lognormal | -0.008 | (-0.026, 0.010) | 0.393 | 0.728 | 0.058 | (-0.025, 0.012) |
| PC (p-42:5)/PC (o-42:6) A | Lognormal | -0.006 | (-0.020, 0.008) | 0.398 | 0.733 | 0.051 | (-0.020, 0.007) |
| LPC (16:1) - ESI(+) | Lognormal | -0.005 | (-0.016, 0.007) | 0.402 | 0.733 | 0.054 | (-0.016, 0.007) |
| PC (40:7) B - ESI(+) | Lognormal | 0.005 | (-0.006, 0.015) | 0.403 | 0.733 | 0.054 | (-0.006, 0.015) |
| GlcCer (d40:1) - ESI(-) | Lognormal | -0.005 | (-0.018, 0.007) | 0.404 | 0.733 | 0.052 | (-0.018, 0.007) |
| PE (38:2) | Lognormal | -0.008 | (-0.028, 0.011) | 0.405 | 0.733 | 0.052 | (-0.028, 0.012) |
| PC (36:5) C | Lognormal | 0.009 | (-0.013, 0.031) | 0.405 | 0.733 | 0.059 | (-0.013, 0.032) |
| TG (55:2) | Lognormal | 0.008 | (-0.011, 0.028) | 0.405 | 0.733 | 0.056 | (-0.011, 0.027) |
| TG (51:5) | Lognormal | -0.008 | (-0.027, 0.011) | 0.409 | 0.737 | 0.057 | (-0.026, 0.011) |
| GlcCer (d42:1) - ESI(-) | Lognormal | -0.005 | (-0.017, 0.007) | 0.410 | 0.737 | 0.052 | (-0.017, 0.007) |
| TG (54:8) | Lognormal | 0.013 | (-0.017, 0.042) | 0.410 | 0.737 | 0.053 | (-0.017, 0.042) |
| PC (40:7) A - ESI(+) | Lognormal | -0.005 | (-0.016, 0.006) | 0.414 | 0.739 | 0.048 | (-0.017, 0.007) |
| TG (50:1) | Lognormal | 0.007 | (-0.010, 0.025) | 0.418 | 0.739 | 0.053 | (-0.010, 0.026) |
| TG (54:2) | Lognormal | 0.006 | (-0.009, 0.021) | 0.418 | 0.739 | 0.050 | (-0.008, 0.021) |
| AC (18:1) | Lognormal | -0.006 | (-0.019, 0.008) | 0.419 | 0.739 | 0.050 | (-0.018, 0.009) |
| TG (40:0) | Lognormal | -0.014 | (-0.048, 0.020) | 0.419 | 0.739 | 0.056 | (-0.048, 0.022) |
| SM (d42:0) - ESI(-) | Lognormal | 0.006 | (-0.008, 0.020) | 0.423 | 0.739 | 0.048 | (-0.008, 0.021) |
| TG (58:10) | Lognormal | 0.009 | (-0.013, 0.031) | 0.425 | 0.741 | 0.053 | (-0.012, 0.031) |
| PC (39:4) | Lognormal | 0.005 | (-0.007, 0.017) | 0.433 | 0.753 | 0.050 | (-0.007, 0.017) |
| LPC (16:0) - ESI(+) | Lognormal | -0.002 | (-0.008, 0.004) | 0.437 | 0.757 | 0.050 | (-0.008, 0.004) |
| TG (53:5) | Lognormal | -0.007 | (-0.023, 0.010) | 0.437 | 0.757 | 0.054 | (-0.024, 0.011) |
| SM (d36:2) - ESI(+) | Lognormal | -0.003 | (-0.010, 0.004) | 0.438 | 0.757 | 0.056 | (-0.010, 0.004) |
| Ceramide (d34:1) - ESI(+) | Lognormal | -0.004 | (-0.014, 0.006) | 0.445 | 0.763 | 0.048 | (-0.014, 0.006) |
| Ceramide (d34:2) | Lognormal | -0.005 | (-0.016, 0.007) | 0.448 | 0.765 | 0.045 | (-0.015, 0.008) |
| TG (56:10) | Lognormal | 0.014 | (-0.022, 0.051) | 0.452 | 0.768 | 0.053 | (-0.024, 0.050) |

**Supplementary Table S6.** Regression and Bayesian estimates comparing sex-stratified ME/CFS vs. controls.

| Metabolite | Regression Model | Female ME/CFS vs. Female Controls | | | | | |
|---|---|---|---|---|---|---|---|
| | | Estimated Coefficient | 95% CI | p-value | FDR | BayesFactor | 95% HDI |
| PC (30:0) | Lognormal | -0.006 | (-0.020, 0.009) | 0.457 | 0.772 | 0.050 | (-0.020, 0.009) |
| TG (59:3) | Lognormal | 0.009 | (-0.016, 0.034) | 0.466 | 0.781 | 0.047 | (-0.016, 0.035) |
| LPC (20:4) | Lognormal | -0.004 | (-0.015, 0.007) | 0.469 | 0.781 | 0.053 | (-0.016, 0.007) |
| TAG (58:7)/TAG (18:1/18:1/22:5) | Lognormal | 0.006 | (-0.010, 0.022) | 0.471 | 0.783 | 0.050 | (-0.011, 0.021) |
| GlcCer (d14:1(4E)/20:0(2OH)) | Lognormal | -0.008 | (-0.030, 0.014) | 0.473 | 0.783 | 0.051 | (-0.030, 0.015) |
| TG (50:2) | Lognormal | 0.005 | (-0.008, 0.017) | 0.475 | 0.784 | 0.049 | (-0.008, 0.017) |
| TG (56:7) A | Lognormal | 0.016 | (-0.027, 0.059) | 0.477 | 0.787 | 0.051 | (-0.028, 0.059) |
| TG (60:4) | Lognormal | 0.012 | (-0.022, 0.046) | 0.495 | 0.804 | 0.045 | (-0.024, 0.045) |
| CE (18:1) | Lognormal | -0.005 | (-0.019, 0.009) | 0.497 | 0.806 | 0.050 | (-0.019, 0.009) |
| TG (46:3) B | Lognormal | -0.013 | (-0.052, 0.025) | 0.499 | 0.806 | 0.051 | (-0.052, 0.026) |
| GlcCer (d42:2) - ESI(-) | Lognormal | 0.005 | (-0.009, 0.018) | 0.502 | 0.806 | 0.045 | (-0.008, 0.018) |
| PE (36:1) - ESI(-) | Lognormal | 0.010 | (-0.019, 0.039) | 0.504 | 0.806 | 0.049 | (-0.019, 0.039) |
| LPC (18:0) - ESI(+) | Lognormal | -0.004 | (-0.014, 0.007) | 0.504 | 0.806 | 0.047 | (-0.014, 0.007) |
| TG (64:2) | Lognormal | 0.012 | (-0.023, 0.047) | 0.505 | 0.806 | 0.051 | (-0.023, 0.048) |
| LPC (22:5) - ESI(+) | Lognormal | 0.008 | (-0.015, 0.030) | 0.507 | 0.806 | 0.049 | (-0.015, 0.030) |
| SM (d41:1) - ESI(-) | Lognormal | -0.035 | (-0.141, 0.071) | 0.515 | 0.811 | 0.052 | (-0.136, 0.074) |
| TG (60:2) | Lognormal | 0.015 | (-0.031, 0.062) | 0.516 | 0.811 | 0.046 | (-0.032, 0.063) |
| PC (40:5) B - ESI(-) | Lognormal | -0.008 | (-0.033, 0.017) | 0.518 | 0.811 | 0.044 | (-0.034, 0.018) |
| TG (56:2) | Lognormal | 0.008 | (-0.015, 0.030) | 0.519 | 0.811 | 0.054 | (-0.018, 0.030) |
| FA (20:2) (eicosadienoic acid) | Lognormal | 0.004 | (-0.008, 0.015) | 0.523 | 0.813 | 0.050 | (-0.009, 0.015) |
| AC (16:0) | Lognormal | -0.004 | (-0.015, 0.008) | 0.527 | 0.813 | 0.047 | (-0.015, 0.007) |
| DG (32:1) | Lognormal | 0.008 | (-0.018, 0.034) | 0.527 | 0.813 | 0.044 | (-0.017, 0.035) |
| PC (p-40:6)/PC (o-40:7) B | Lognormal | -0.006 | (-0.024, 0.012) | 0.532 | 0.813 | 0.042 | (-0.024, 0.012) |
| FA (14:0) (myristic acid) | Lognormal | 0.003 | (-0.006, 0.012) | 0.533 | 0.813 | 0.046 | (-0.006, 0.013) |
| TG (64:3) | Lognormal | -0.008 | (-0.033, 0.017) | 0.534 | 0.813 | 0.043 | (-0.034, 0.016) |
| PC (p-38:5)/PC (o-38:6) | Lognormal | 0.013 | (-0.028, 0.053) | 0.535 | 0.813 | 0.047 | (-0.029, 0.052) |
| PC (32:0) - ESI(+) | Lognormal | 0.002 | (-0.004, 0.007) | 0.551 | 0.829 | 0.044 | (-0.004, 0.008) |
| TG (62:3) | Lognormal | 0.013 | (-0.030, 0.056) | 0.558 | 0.836 | 0.049 | (-0.029, 0.056) |
| PC (35:4) - ESI(-) | Lognormal | -0.006 | (-0.025, 0.014) | 0.559 | 0.836 | 0.042 | (-0.026, 0.012) |
| TG (49:1) | Lognormal | 0.007 | (-0.017, 0.031) | 0.563 | 0.838 | 0.047 | (-0.018, 0.031) |
| PC (33:1) - ESI(-) | Lognormal | 0.007 | (-0.016, 0.030) | 0.563 | 0.838 | 0.050 | (-0.017, 0.029) |
| PC (36:1) - ESI(-) | Lognormal | -0.003 | (-0.012, 0.007) | 0.568 | 0.844 | 0.045 | (-0.013, 0.006) |
| SM (d43:2) - ESI(+) | Lognormal | 0.005 | (-0.013, 0.023) | 0.575 | 0.847 | 0.046 | (-0.014, 0.023) |
| SM (d32:0) - ESI(+) | Lognormal | 0.005 | (-0.013, 0.023) | 0.577 | 0.847 | 0.047 | (-0.013, 0.023) |
| PC (p-38:6)/PC (o-38:7) | Lognormal | -0.005 | (-0.020, 0.011) | 0.578 | 0.847 | 0.044 | (-0.020, 0.012) |
| PE (38:6) - ESI(-) | Lognormal | 0.008 | (-0.021, 0.037) | 0.578 | 0.847 | 0.045 | (-0.020, 0.036) |
| PC (32:1) - ESI(+) | Lognormal | 0.004 | (-0.009, 0.016) | 0.578 | 0.847 | 0.044 | (-0.009, 0.016) |
| PC (p-40:6)/PC (o-40:7) A | Lognormal | 0.004 | (-0.010, 0.018) | 0.581 | 0.849 | 0.044 | (-0.010, 0.019) |
| SM (d34:0) - ESI(+) | Lognormal | -0.002 | (-0.011, 0.006) | 0.586 | 0.853 | 0.044 | (-0.011, 0.005) |
| PC (38:5) A - ESI(-) | Lognormal | -0.003 | (-0.013, 0.007) | 0.592 | 0.858 | 0.042 | (-0.013, 0.008) |
| CE (16:1) | Lognormal | 0.004 | (-0.010, 0.018) | 0.593 | 0.858 | 0.043 | (-0.010, 0.017) |
| Ceramide (d40:0) | Lognormal | 0.008 | (-0.020, 0.036) | 0.593 | 0.858 | 0.038 | (-0.020, 0.035) |
| PC (p-42:5)/PC (o-42:6) B | Lognormal | -0.005 | (-0.021, 0.012) | 0.594 | 0.858 | 0.044 | (-0.022, 0.013) |
| Ceramide (d42:0) | Lognormal | 0.004 | (-0.011, 0.020) | 0.596 | 0.859 | 0.042 | (-0.012, 0.020) |
| LPC (22:4) | Lognormal | -0.007 | (-0.034, 0.019) | 0.598 | 0.859 | 0.044 | (-0.034, 0.019) |
| PC (35:1) - ESI(-) | Lognormal | 0.004 | (-0.012, 0.021) | 0.599 | 0.859 | 0.039 | (-0.012, 0.020) |
| TG (57:2) | Lognormal | 0.005 | (-0.013, 0.023) | 0.608 | 0.865 | 0.047 | (-0.013, 0.023) |

**Supplementary Table S6.** Regression and Bayesian estimates comparing sex-stratified ME/CFS vs. controls.

| Metabolite | Regression Model | Female ME/CFS vs. Female Controls | | | | | |
|---|---|---|---|---|---|---|---|
| | | Estimated Coefficient | 95% CI | p-value | FDR | BayesFactor | 95% HDI |
| PC (40:6) B | Lognormal | 0.004 | (-0.011, 0.018) | 0.609 | 0.865 | 0.042 | (-0.010, 0.018) |
| PC (40:5) B - ESI(+) | Lognormal | -0.004 | (-0.018, 0.011) | 0.616 | 0.870 | 0.040 | (-0.018, 0.011) |
| TG (42:2) | Lognormal | -0.024 | (-0.118, 0.071) | 0.621 | 0.872 | 0.044 | (-0.115, 0.068) |
| CE (22:6) | Lognormal | 0.003 | (-0.010, 0.016) | 0.622 | 0.872 | 0.041 | (-0.009, 0.017) |
| TG (52:3) | Lognormal | -0.002 | (-0.009, 0.005) | 0.623 | 0.872 | 0.047 | (-0.009, 0.005) |
| PC (38:7) | Lognormal | 0.004 | (-0.013, 0.022) | 0.627 | 0.873 | 0.046 | (-0.013, 0.021) |
| TG (60:3) | Lognormal | 0.008 | (-0.026, 0.043) | 0.627 | 0.873 | 0.046 | (-0.025, 0.044) |
| CE (22:2) | Lognormal | -0.010 | (-0.052, 0.031) | 0.629 | 0.873 | 0.043 | (-0.055, 0.030) |
| FA (22:6) (docosahexaenoic acid) | Lognormal | 0.004 | (-0.013, 0.021) | 0.629 | 0.873 | 0.043 | (-0.014, 0.020) |
| PC (36:4) C - ESI(+) | Lognormal | 0.001 | (-0.004, 0.007) | 0.639 | 0.879 | 0.042 | (-0.004, 0.007) |
| DG (34:1) | Lognormal | -0.007 | (-0.036, 0.023) | 0.650 | 0.889 | 0.044 | (-0.036, 0.023) |
| PE (36:1) - ESI(+) | Lognormal | -0.005 | (-0.029, 0.018) | 0.658 | 0.895 | 0.039 | (-0.030, 0.017) |
| TG (53:1) | Lognormal | 0.007 | (-0.024, 0.037) | 0.660 | 0.895 | 0.041 | (-0.023, 0.039) |
| TG (46:1) | Lognormal | -0.008 | (-0.043, 0.027) | 0.661 | 0.895 | 0.039 | (-0.045, 0.025) |
| PC (35:4) - ESI(+) | Lognormal | -0.003 | (-0.015, 0.010) | 0.664 | 0.897 | 0.040 | (-0.016, 0.009) |
| SM (d42:2) | Lognormal | 0.002 | (-0.006, 0.010) | 0.665 | 0.897 | 0.042 | (-0.006, 0.010) |
| DG (36:3) | Lognormal | -0.003 | (-0.016, 0.010) | 0.671 | 0.900 | 0.043 | (-0.016, 0.010) |
| SM (d37:1) | Lognormal | -0.004 | (-0.023, 0.015) | 0.672 | 0.900 | 0.044 | (-0.023, 0.014) |
| LPC (20:1) - ESI(-) | Lognormal | -0.004 | (-0.022, 0.014) | 0.672 | 0.900 | 0.046 | (-0.023, 0.015) |
| TG (53:3) | Lognormal | 0.002 | (-0.009, 0.014) | 0.678 | 0.904 | 0.044 | (-0.009, 0.014) |
| TG (52:5) | Lognormal | -0.003 | (-0.017, 0.011) | 0.681 | 0.905 | 0.041 | (-0.016, 0.012) |
| TG (48:6) | Lognormal | 0.006 | (-0.023, 0.036) | 0.681 | 0.905 | 0.045 | (-0.023, 0.036) |
| TG (54:4) | Lognormal | -0.003 | (-0.015, 0.010) | 0.687 | 0.906 | 0.042 | (-0.014, 0.010) |
| GlcCer (d34:1) | Lognormal | -0.005 | (-0.032, 0.021) | 0.688 | 0.906 | 0.043 | (-0.033, 0.021) |
| SM (d41:2) - ESI(-) | Lognormal | -0.003 | (-0.018, 0.012) | 0.693 | 0.907 | 0.044 | (-0.019, 0.011) |
| PC (p-40:4)/PC (o-40:5) - ESI(+) | Lognormal | -0.002 | (-0.013, 0.009) | 0.694 | 0.907 | 0.039 | (-0.013, 0.008) |
| SM (d32:0) - ESI(-) | Lognormal | -0.013 | (-0.076, 0.051) | 0.696 | 0.907 | 0.044 | (-0.075, 0.055) |
| LPC (22:6) | Lognormal | 0.003 | (-0.013, 0.020) | 0.696 | 0.907 | 0.040 | (-0.014, 0.020) |
| PC 38:7e | Lognormal | -0.004 | (-0.024, 0.016) | 0.699 | 0.908 | 0.040 | (-0.024, 0.014) |
| LPC (20:1) - ESI(+) | Lognormal | -0.006 | (-0.038, 0.026) | 0.712 | 0.913 | 0.040 | (-0.038, 0.027) |
| TG (50:6) | Lognormal | 0.006 | (-0.025, 0.037) | 0.712 | 0.913 | 0.041 | (-0.025, 0.036) |
| TG (55:1) | Lognormal | 0.005 | (-0.020, 0.029) | 0.714 | 0.914 | 0.041 | (-0.020, 0.030) |
| FA (15:0) (pentadecylic acid) | Lognormal | -0.002 | (-0.012, 0.008) | 0.728 | 0.927 | 0.038 | (-0.012, 0.009) |
| TG (44:1) | Lognormal | -0.008 | (-0.054, 0.038) | 0.731 | 0.927 | 0.041 | (-0.056, 0.041) |
| PC (28:0) | Lognormal | -0.008 | (-0.052, 0.037) | 0.733 | 0.928 | 0.042 | (-0.050, 0.038) |
| TG (59:2) | Lognormal | -0.005 | (-0.031, 0.022) | 0.736 | 0.928 | 0.045 | (-0.030, 0.023) |
| PC (38:3) - ESI(+) | Lognormal | -0.002 | (-0.010, 0.007) | 0.739 | 0.928 | 0.036 | (-0.010, 0.007) |
| PC (p-42:4)/PC (o-42:5) - ESI(+) | Lognormal | -0.002 | (-0.015, 0.011) | 0.739 | 0.928 | 0.036 | (-0.014, 0.011) |
| Ceramide (d36:1) - ESI(-) | Lognormal | -0.004 | (-0.025, 0.018) | 0.744 | 0.928 | 0.042 | (-0.025, 0.018) |
| TG (62:1) | Lognormal | -0.005 | (-0.037, 0.027) | 0.745 | 0.928 | 0.042 | (-0.038, 0.026) |
| PC (33:0) | Lognormal | -0.004 | (-0.026, 0.018) | 0.748 | 0.928 | 0.041 | (-0.026, 0.018) |
| TG (52:6) | Lognormal | 0.003 | (-0.018, 0.025) | 0.749 | 0.928 | 0.041 | (-0.018, 0.024) |
| TG (46:5) | Lognormal | -0.008 | (-0.057, 0.042) | 0.762 | 0.931 | 0.043 | (-0.060, 0.040) |
| FA (17:0) (margaric acid) | Lognormal | 0.001 | (-0.006, 0.009) | 0.765 | 0.931 | 0.040 | (-0.007, 0.008) |
| TG (58:1) | Lognormal | 0.005 | (-0.025, 0.034) | 0.765 | 0.931 | 0.042 | (-0.025, 0.034) |
| SM (d33:1) - ESI(+) | Lognormal | -0.002 | (-0.013, 0.009) | 0.767 | 0.931 | 0.044 | (-0.013, 0.009) |
| PC (36:5) B | Lognormal | 0.004 | (-0.022, 0.030) | 0.776 | 0.936 | 0.038 | (-0.020, 0.030) |

**Supplementary Table S6.** Regression and Bayesian estimates comparing sex-stratified ME/CFS vs. controls.

| Metabolite | Regression Model | Female ME/CFS vs. Female Controls | | | | | |
|---|---|---|---|---|---|---|---|
| | | Estimated Coefficient | 95% CI | p-value | FDR | BayesFactor | 95% HDI |
| TG (54:3) | Lognormal | 0.002 | (-0.010, 0.013) | 0.778 | 0.936 | 0.042 | (-0.009, 0.013) |
| FA (12:0) (lauric acid) | Lognormal | -0.003 | (-0.022, 0.016) | 0.779 | 0.936 | 0.041 | (-0.022, 0.016) |
| CE (16:0) | Lognormal | -0.004 | (-0.035, 0.026) | 0.784 | 0.936 | 0.037 | (-0.036, 0.025) |
| PE (38:4) | Lognormal | -0.002 | (-0.013, 0.010) | 0.785 | 0.936 | 0.034 | (-0.014, 0.011) |
| TG (53:4) | Lognormal | -0.002 | (-0.015, 0.011) | 0.787 | 0.936 | 0.041 | (-0.016, 0.011) |
| TG (60:1) | Lognormal | 0.004 | (-0.028, 0.037) | 0.788 | 0.936 | 0.040 | (-0.028, 0.036) |
| DG (34:3) | Lognormal | -0.003 | (-0.026, 0.019) | 0.789 | 0.936 | 0.037 | (-0.026, 0.020) |
| PC (38:5) B - ESI(-) | Lognormal | 0.003 | (-0.017, 0.022) | 0.792 | 0.937 | 0.041 | (-0.018, 0.022) |
| FA (18:3) (linolenic acid) | Lognormal | 0.002 | (-0.011, 0.014) | 0.801 | 0.945 | 0.042 | (-0.012, 0.015) |
| GlcCer (d42:1) - ESI(+) | Lognormal | 0.001 | (-0.010, 0.013) | 0.804 | 0.946 | 0.036 | (-0.010, 0.012) |
| PC (31:1) | Lognormal | -0.005 | (-0.043, 0.033) | 0.807 | 0.947 | 0.040 | (-0.042, 0.033) |
| PC (38:6) - ESI(-) | Lognormal | 0.002 | (-0.012, 0.015) | 0.810 | 0.948 | 0.038 | (-0.011, 0.015) |
| PC (p-44:4)/PC (o-44:5) - ESI(+) | Lognormal | -0.002 | (-0.016, 0.012) | 0.811 | 0.948 | 0.039 | (-0.016, 0.013) |
| CE (18:0) | Lognormal | -0.003 | (-0.028, 0.022) | 0.811 | 0.948 | 0.036 | (-0.028, 0.020) |
| PC (40:6) A | Lognormal | 0.001 | (-0.010, 0.013) | 0.818 | 0.952 | 0.039 | (-0.011, 0.013) |
| GlcCer (d40:1) - ESI(+) | Lognormal | -0.002 | (-0.016, 0.013) | 0.818 | 0.952 | 0.039 | (-0.016, 0.012) |
| Ceramide (d34:0) | Lognormal | -0.002 | (-0.015, 0.012) | 0.820 | 0.952 | 0.031 | (-0.014, 0.011) |
| TG (57:1) | Lognormal | 0.002 | (-0.019, 0.023) | 0.823 | 0.952 | 0.038 | (-0.019, 0.024) |
| PC (16:0/9:0(CHO)) | Lognormal | -0.003 | (-0.025, 0.020) | 0.826 | 0.952 | 0.032 | (-0.025, 0.021) |
| PC (38:4) A - ESI(+) | Lognormal | -0.001 | (-0.012, 0.010) | 0.826 | 0.952 | 0.041 | (-0.012, 0.010) |
| TG (52:1) | Lognormal | 0.002 | (-0.018, 0.023) | 0.827 | 0.952 | 0.037 | (-0.018, 0.022) |
| TG (49:3) | Lognormal | 0.003 | (-0.021, 0.026) | 0.830 | 0.953 | 0.042 | (-0.021, 0.026) |
| Ceramide (d40:2) | Lognormal | 0.005 | (-0.043, 0.053) | 0.830 | 0.953 | 0.043 | (-0.041, 0.055) |
| PC (p-42:5)/PC (o-42:6) | Lognormal | -0.004 | (-0.037, 0.030) | 0.835 | 0.956 | 0.041 | (-0.037, 0.030) |
| TG (56:1) | Lognormal | -0.003 | (-0.031, 0.025) | 0.839 | 0.959 | 0.039 | (-0.031, 0.026) |
| CE (20:4) | Lognormal | 0.001 | (-0.009, 0.011) | 0.843 | 0.960 | 0.041 | (-0.008, 0.011) |
| TG (49:0) | Lognormal | -0.002 | (-0.025, 0.021) | 0.847 | 0.963 | 0.038 | (-0.025, 0.023) |
| PC (40:5) A - ESI(-) | Lognormal | -0.002 | (-0.019, 0.016) | 0.852 | 0.964 | 0.040 | (-0.019, 0.016) |
| SM (d36:1) - ESI(+) | Lognormal | -0.001 | (-0.007, 0.006) | 0.856 | 0.964 | 0.043 | (-0.007, 0.006) |
| TG (58:5) | Lognormal | 0.002 | (-0.019, 0.022) | 0.857 | 0.964 | 0.041 | (-0.018, 0.023) |
| PC (38:4) C - ESI(+) | Lognormal | 0.001 | (-0.006, 0.007) | 0.870 | 0.966 | 0.039 | (-0.006, 0.007) |
| LPC (20:0) | Lognormal | -0.002 | (-0.021, 0.018) | 0.876 | 0.969 | 0.040 | (-0.021, 0.018) |
| PC (40:7) - ESI(-) | Lognormal | -0.002 | (-0.023, 0.020) | 0.879 | 0.970 | 0.040 | (-0.022, 0.021) |
| DG (34:2) | Lognormal | 0.001 | (-0.013, 0.015) | 0.879 | 0.970 | 0.039 | (-0.014, 0.015) |
| LPC (p-18:0)/LPC (o-18:1) | Lognormal | -0.001 | (-0.018, 0.016) | 0.882 | 0.970 | 0.038 | (-0.018, 0.016) |
| TG (58:2) | Lognormal | 0.003 | (-0.039, 0.045) | 0.886 | 0.970 | 0.040 | (-0.038, 0.047) |
| PC (31:0) | Lognormal | 0.001 | (-0.014, 0.016) | 0.894 | 0.973 | 0.036 | (-0.015, 0.014) |
| PC (40:8) - ESI(+) | Lognormal | 0.001 | (-0.010, 0.011) | 0.895 | 0.973 | 0.037 | (-0.011, 0.010) |
| PC (p-38:5)/PC (o-38:6) A | Lognormal | -0.001 | (-0.018, 0.015) | 0.897 | 0.973 | 0.039 | (-0.017, 0.015) |
| TG (62:2) | Lognormal | 0.002 | (-0.036, 0.040) | 0.901 | 0.973 | 0.040 | (-0.035, 0.040) |
| PE (34:2) - ESI(+) | Lognormal | -0.005 | (-0.083, 0.074) | 0.909 | 0.977 | 0.038 | (-0.090, 0.068) |
| TG (50:3) A | Lognormal | 0.001 | (-0.014, 0.016) | 0.910 | 0.977 | 0.037 | (-0.015, 0.016) |
| TG (51:3) | Lognormal | 0.001 | (-0.012, 0.014) | 0.915 | 0.981 | 0.039 | (-0.012, 0.014) |
| PC (p-40:4)/PC (o-40:5) - ESI(-) | Lognormal | -0.001 | (-0.017, 0.015) | 0.916 | 0.981 | 0.040 | (-0.017, 0.016) |
| PC (36:6) | Lognormal | 0.001 | (-0.020, 0.022) | 0.919 | 0.982 | 0.039 | (-0.019, 0.023) |
| FA (10:0) (capric acid) | Lognormal | 0.001 | (-0.028, 0.031) | 0.920 | 0.982 | 0.037 | (-0.026, 0.032) |
| Ceramide (d42:2) A - ESI (+) | Lognormal | 0.000 | (-0.009, 0.008) | 0.925 | 0.986 | 0.034 | (-0.008, 0.008) |

**Supplementary Table S6.** Regression and Bayesian estimates comparing sex-stratified ME/CFS vs. controls.

| Metabolite | Regression Model | Female ME/CFS vs. Female Controls | | | | | |
|---|---|---|---|---|---|---|---|
| | | Estimated Coefficient | 95% CI | p-value | FDR | BayesFactor | 95% HDI |
| Gal-Gal-Cer (d18:1/16:0)/Lactosylceramide (d18:1/16:0) | Lognormal | -0.001 | (-0.012, 0.011) | 0.927 | 0.986 | 0.036 | (-0.011, 0.011) |
| PE (36:4) - ESI(-) | Lognormal | -0.001 | (-0.030, 0.028) | 0.927 | 0.986 | 0.034 | (-0.031, 0.028) |
| LPC (18:1) - ESI(+) | Lognormal | -0.001 | (-0.012, 0.011) | 0.930 | 0.988 | 0.040 | (-0.012, 0.011) |
| PC (p-36:5)/PC (o-36:6) | Lognormal | 0.001 | (-0.023, 0.025) | 0.940 | 0.994 | 0.041 | (-0.023, 0.026) |
| TG (56:4) | Lognormal | 0.000 | (-0.014, 0.015) | 0.952 | 0.994 | 0.042 | (-0.014, 0.015) |
| PC 40:6e | Lognormal | 0.000 | (-0.011, 0.011) | 0.956 | 0.994 | 0.037 | (-0.012, 0.010) |
| PC (p-44:5)/PC (o-44:6) | Lognormal | 0.000 | (-0.017, 0.016) | 0.956 | 0.994 | 0.038 | (-0.018, 0.016) |
| SM (d41:2) A - ESI(+) | Lognormal | 0.000 | (-0.012, 0.011) | 0.956 | 0.994 | 0.041 | (-0.012, 0.012) |
| TG (50:3) B | Lognormal | -0.002 | (-0.099, 0.094) | 0.963 | 0.994 | 0.036 | (-0.101, 0.091) |
| TG (44:2) | Lognormal | -0.002 | (-0.090, 0.086) | 0.967 | 0.994 | 0.040 | (-0.096, 0.082) |
| TG (48:1) | Lognormal | 0.000 | (-0.023, 0.022) | 0.968 | 0.994 | 0.038 | (-0.023, 0.021) |
| SM (d43:1) - ESI(+) | Lognormal | 0.000 | (-0.015, 0.015) | 0.969 | 0.994 | 0.040 | (-0.015, 0.015) |
| LPC (22:5) - ESI(-) | Lognormal | -0.001 | (-0.040, 0.039) | 0.970 | 0.994 | 0.040 | (-0.040, 0.037) |
| TG (48:2) | Lognormal | 0.000 | (-0.024, 0.023) | 0.971 | 0.994 | 0.038 | (-0.026, 0.022) |
| FA (24:1) (nervonic acid) | Lognormal | 0.000 | (-0.011, 0.011) | 0.972 | 0.994 | 0.037 | (-0.010, 0.011) |
| FA (20:4) (arachidonic acid) | Lognormal | 0.000 | (-0.008, 0.009) | 0.973 | 0.994 | 0.039 | (-0.008, 0.009) |
| PC (40:4) - ESI(+) | Lognormal | 0.000 | (-0.014, 0.013) | 0.976 | 0.995 | 0.039 | (-0.015, 0.012) |
| PC (32:1) - ESI(-) | Lognormal | 0.000 | (-0.018, 0.018) | 0.979 | 0.995 | 0.039 | (-0.018, 0.017) |
| PC (38:4) B - ESI(+) | Lognormal | 0.000 | (-0.010, 0.010) | 0.982 | 0.995 | 0.033 | (-0.010, 0.010) |
| PC (34:0) - ESI(+) | Lognormal | 0.000 | (-0.006, 0.006) | 0.985 | 0.995 | 0.034 | (-0.006, 0.006) |
| Ceramide (d36:1) - ESI(+) | Lognormal | 0.000 | (-0.012, 0.012) | 0.998 | 0.999 | 0.036 | (-0.012, 0.013) |
| **Oxylipins (OL)** | | | | | | | |
| Resolvin D1 | Gamma | -0.513 | (-0.871, -0.156) | 0.006 | 0.183 | 1.811 | (-0.857, -0.173) |
| 5-Hydroxy-6,8,11,14,17-eicosapentaenoic acid | Gamma | 0.330 | (0.072, 0.587) | 0.013 | 0.225 | 0.520 | (0.069, 0.597) |
| 9-nitrooleic acid | Gamma | 0.409 | (0.089, 0.729) | 0.013 | 0.225 | 0.588 | (0.082, 0.778) |
| 15,16-dihydroxyoctadeca-9,12-dienoic acid | Lognormal | -0.272 | (-0.487, -0.057) | 0.014 | 0.229 | 0.691 | (-0.464, -0.041) |
| 15-hydroxyeicosa-5,8,11,13,17-pentaenoic acid | Gamma | 0.506 | (0.101, 0.911) | 0.016 | 0.239 | 0.788 | (0.113, 0.947) |
| 12-Hydroxy-5,8,10,14,17-eicosapentaenoic acid | Gamma | 0.514 | (0.080, 0.948) | 0.022 | 0.274 | 2.049 | (0.156, 0.855) |
| Prostaglandin F2a | Gamma | -0.338 | (-0.647, -0.029) | 0.034 | 0.320 | 0.228 | (-0.688, -0.005) |
| Prostaglandin D2 | Gamma | -0.619 | (-1.188, -0.050) | 0.035 | 0.320 | 1.241 | (-1.107, -0.160) |
| 4-hydroxydocosa-5,7,10,13,16,19-hexaenoic acid | Gamma | 0.373 | (0.021, 0.725) | 0.040 | 0.326 | 0.245 | (-0.001, 0.726) |
| 9-Hydroxy-5,7,11,14,17-icosapentaenoic acid | Gamma | 0.267 | (-0.004, 0.538) | 0.056 | 0.380 | 0.147 | (-0.027, 0.566) |
| 8,9-Epoxyeicosa-5,11,14-trienoic acid | Gamma | -0.292 | (-0.590, 0.005) | 0.056 | 0.380 | 0.129 | (-0.632, 0.052) |
| 9,10-dihydroxyoctadec-12-enoic acid | Lognormal | -0.228 | (-0.461, 0.006) | 0.058 | 0.383 | 0.245 | (-0.458, 0.005) |
| 11,12-Epoxyeicosa-5,8,14-trienoic acid | Gamma | 0.298 | (-0.024, 0.620) | 0.072 | 0.419 | 0.148 | (-0.034, 0.640) |
| 11(12)-epoxy-5,8,14,17- eicosatetraenoic acid | Gamma | 0.413 | (-0.052, 0.878) | 0.084 | 0.444 | 0.220 | (-0.096, 0.893) |
| 8-hydroxyeicosa-5,9,11,14-tetraenoic acid | Gamma | 0.235 | (-0.034, 0.504) | 0.089 | 0.451 | 0.116 | (-0.060, 0.550) |
| 6-Ketoprostaglandin F1 alpha | Gamma | 0.362 | (-0.075, 0.799) | 0.107 | 0.492 | 0.182 | (-0.039, 0.786) |
| 15-ketoeicosa-5,8,11,13-tetraenoic acid | Gamma | 0.344 | (-0.082, 0.770) | 0.116 | 0.502 | 0.229 | (-0.011, 0.688) |
| 17,18-Epoxy-5,8,11,14-eicosatetraenoic acid | Gamma | -0.398 | (-0.893, 0.098) | 0.118 | 0.506 | 0.201 | (-0.833, 0.038) |
| 17,18-dihydroxyeicosa-5,8,11,14-tetraenoic acid | Lognormal | 0.220 | (-0.060, 0.501) | 0.126 | 0.519 | 0.134 | (-0.064, 0.489) |
| 15-Deoxy-delta-12,14-Prostaglandin J2 | Gamma | -0.232 | (-0.539, 0.074) | 0.140 | 0.532 | 0.096 | (-0.547, 0.071) |
| 15(16)-epoxy-9,12-octadecadienoic acid | Gamma | -0.315 | (-0.739, 0.108) | 0.146 | 0.535 | 0.107 | (-0.706, 0.090) |
| 9,11,15-trihydroxy-5,13,1Z-prostatrienoic acid | Gamma | -0.459 | (-1.084, 0.166) | 0.153 | 0.549 | 0.400 | (-0.941, -0.071) |
| 9,10-dihydroxyoctadeca-12,15-dienoic acid | Gamma | -0.178 | (-0.423, 0.067) | 0.157 | 0.556 | 0.060 | (-0.460, 0.107) |
| 9(10)-epoxy-12Z-octadecenoic acid | Gamma | -0.151 | (-0.368, 0.065) | 0.172 | 0.589 | 0.067 | (-0.352, 0.050) |
| 9(10)-epoxy-12,15-octadecadienoic acid | Gamma | 0.393 | (-0.177, 0.963) | 0.179 | 0.593 | 0.198 | (-0.095, 0.887) |

**Supplementary Table S6.** Regression and Bayesian estimates comparing sex-stratified ME/CFS vs. controls.

| Metabolite | Regression Model | Female ME/CFS vs. Female Controls | | | | | |
|---|---|---|---|---|---|---|---|
| | | Estimated Coefficient | 95% CI | p-value | FDR | BayesFactor | 95% HDI |
| 10-nitrooleic acid | Lognormal | 0.179 | (-0.085, 0.442) | 0.186 | 0.598 | 0.087 | (-0.084, 0.456) |
| 11-Hydroxy-arachidonic acid | Gamma | 0.131 | (-0.071, 0.333) | 0.206 | 0.610 | 0.049 | (-0.087, 0.356) |
| 6-trans-Leukotriene B4 | Gamma | -0.385 | (-0.979, 0.210) | 0.207 | 0.610 | 0.147 | (-0.814, 0.142) |
| 8,9-dihydroxyeicosa-5,11,14-trienoic acid | Gamma | 0.159 | (-0.095, 0.414) | 0.221 | 0.621 | 0.056 | (-0.087, 0.423) |
| 13-hydroxyoctadeca-9,11,15-trienoic acid | Gamma | -0.149 | (-0.395, 0.098) | 0.240 | 0.636 | 0.049 | (-0.418, 0.116) |
| 12(13)-epoxy-9,15-octadecadienoic acid | Gamma | -0.256 | (-0.694, 0.181) | 0.253 | 0.637 | 0.092 | (-0.622, 0.100) |
| Leukotriene B5 | Gamma | -0.334 | (-0.931, 0.263) | 0.275 | 0.651 | 0.121 | (-0.852, 0.152) |
| 12,13-dihydroxyoctadec-9-enoic acid | Lognormal | -0.106 | (-0.305, 0.093) | 0.298 | 0.668 | 0.062 | (-0.293, 0.093) |
| 9-Hydroxylinoleic acid | Lognormal | -0.077 | (-0.226, 0.072) | 0.310 | 0.676 | 0.058 | (-0.230, 0.071) |
| 13-ketooctadeca-9,11-dienoic acid | Lognormal | -0.160 | (-0.469, 0.148) | 0.310 | 0.676 | 0.057 | (-0.464, 0.149) |
| 16(17)-epoxy-4,7,10,13,19-docosapentaenoic acid | Gamma | 0.194 | (-0.189, 0.577) | 0.322 | 0.684 | 0.069 | (-0.182, 0.560) |
| 5,6-dihydroxyeicosa-8,11,14-trienoic acid | Gamma | 0.122 | (-0.132, 0.375) | 0.348 | 0.694 | 0.043 | (-0.131, 0.383) |
| 17-hydroxy-4,7,10,13,15,19-docosahexaenoic acid | Gamma | 0.116 | (-0.139, 0.371) | 0.375 | 0.711 | 0.043 | (-0.193, 0.408) |
| 9,12,13-trihydroxyoctadec-10-enoic acid | Lognormal | -0.064 | (-0.211, 0.083) | 0.394 | 0.728 | 0.046 | (-0.208, 0.086) |
| 10-nitrolinoleic acid | Gamma | 0.199 | (-0.265, 0.663) | 0.402 | 0.733 | 0.069 | (-0.258, 0.612) |
| Prostaglandin E2 | Gamma | 0.260 | (-0.350, 0.871) | 0.405 | 0.733 | 0.086 | (-0.208, 0.727) |
| 9-hydroxyeicosa-5,7,11,14-tetraenoic acid | Gamma | 0.108 | (-0.149, 0.365) | 0.412 | 0.739 | 0.040 | (-0.184, 0.391) |
| Prostaglandin E1 | Gamma | 0.178 | (-0.250, 0.605) | 0.417 | 0.739 | 0.054 | (-0.175, 0.594) |
| 8,15-dihydroxyeicosa-5,9,11,13-tetraenoic acid | Gamma | -0.253 | (-0.866, 0.359) | 0.419 | 0.739 | 0.077 | (-0.782, 0.286) |
| 9-hydroxyoctadeca-10,12,15-trienoic acid | Gamma | -0.127 | (-0.436, 0.182) | 0.422 | 0.739 | 0.043 | (-0.425, 0.157) |
| 14(15)-epoxy-5,8,11,17-eicosatetraenoic acid | Gamma | 0.237 | (-0.367, 0.841) | 0.443 | 0.763 | 0.083 | (-0.382, 0.869) |
| 9,10-Epoxystearic acid | Lognormal | 0.132 | (-0.205, 0.469) | 0.445 | 0.763 | 0.049 | (-0.205, 0.450) |
| 5,6,15-trihydroxyeicosa-7,9,11,13-tetraenoic acid | Gamma | 0.153 | (-0.243, 0.548) | 0.450 | 0.767 | 0.053 | (-0.199, 0.538) |
| 12,13-epoxy-9-octadecenoic acid | Lognormal | -0.118 | (-0.430, 0.194) | 0.460 | 0.775 | 0.052 | (-0.410, 0.212) |
| Thromboxane B2 | Lognormal | 0.106 | (-0.188, 0.399) | 0.481 | 0.789 | 0.048 | (-0.197, 0.382) |
| 20-Hydroxyarachidonic acid | Lognormal | -0.155 | (-0.587, 0.276) | 0.482 | 0.789 | 0.039 | (-0.588, 0.270) |
| Leukotriene B4 | Gamma | -0.080 | (-0.307, 0.147) | 0.490 | 0.798 | 0.028 | (-0.284, 0.151) |
| 13-Hydroxyoctadecadienoic acid | Lognormal | -0.038 | (-0.173, 0.097) | 0.583 | 0.850 | 0.043 | (-0.170, 0.105) |
| 9S,10R-dihydroxy-stearic acid | Lognormal | -0.080 | (-0.380, 0.221) | 0.605 | 0.865 | 0.039 | (-0.366, 0.220) |
| 5-ketoeicosa-6,8,11,14-tetraenoic acid | Gamma | -0.088 | (-0.424, 0.247) | 0.607 | 0.865 | 0.043 | (-0.460, 0.285) |
| Prostaglandin E3 | Gamma | 0.133 | (-0.470, 0.735) | 0.667 | 0.899 | 0.077 | (-0.422, 0.914) |
| 5,15-dihydroxyeicosa-6,8,11,13-tetraenoic acid | Gamma | -0.107 | (-0.605, 0.392) | 0.676 | 0.904 | 0.049 | (-0.588, 0.364) |
| 12,13-dihydroxyoctadeca-9,15-dienoic acid | Gamma | -0.112 | (-0.662, 0.438) | 0.690 | 0.906 | 0.048 | (-0.504, 0.308) |
| 14-hydroxydocosa-4,7,10,12,16,19-hexaenoic acid | Lognormal | 0.079 | (-0.309, 0.466) | 0.691 | 0.906 | 0.032 | (-0.293, 0.465) |
| trans-12,13-epoxy-11-oxo-trans-9-octadecenoic acid | Gamma | -0.031 | (-0.217, 0.156) | 0.748 | 0.928 | 0.025 | (-0.251, 0.187) |
| 14,15-dihydroxyeicosa-5,8,11,17-tetraenoic acid | Gamma | 0.053 | (-0.275, 0.380) | 0.753 | 0.930 | 0.041 | (-0.315, 0.424) |
| 19,20-dihydroxydocosa-4,7,10,13,16-pentaenoic acid | Lognormal | 0.028 | (-0.150, 0.206) | 0.760 | 0.931 | 0.038 | (-0.146, 0.200) |
| 5-Hydroxy-6,8,11,14-eicosatetraenoic acid | Lognormal | 0.032 | (-0.180, 0.244) | 0.766 | 0.931 | 0.034 | (-0.183, 0.242) |
| 14,15-dihydroxyeicosa-5,8,11-trienoic acid | Gamma | -0.014 | (-0.171, 0.144) | 0.865 | 0.966 | 0.016 | (-0.166, 0.139) |
| 18-(3-ethyloxiran-2-yl)octadeca-4,7,10,13,16-pentaenoic acid | Gamma | -0.028 | (-0.398, 0.343) | 0.884 | 0.970 | 0.037 | (-0.380, 0.314) |
| 15-hydroxyeicosa-5,8,11,13-tetraenoic acid | Lognormal | 0.011 | (-0.162, 0.184) | 0.902 | 0.973 | 0.035 | (-0.167, 0.185) |
| 12-Hydroxy-5,8,10,14-eicosatetraenoic acid | Lognormal | -0.008 | (-0.244, 0.227) | 0.945 | 0.994 | 0.027 | (-0.240, 0.239) |
| 11-Hydroxy-14,15-epoxyeicosatrienoic acid | Gamma | 0.020 | (-0.560, 0.600) | 0.946 | 0.994 | 0.055 | (-0.517, 0.562) |
| 9-ketooctadeca-10,12-dienoic acid | Gamma | 0.006 | (-0.260, 0.273) | 0.962 | 0.994 | 0.030 | (-0.282, 0.324) |
| 15-Keto-prostaglandin E2 | Gamma | 0.013 | (-0.529, 0.554) | 0.963 | 0.994 | 0.045 | (-0.445, 0.476) |
| 11,12-Dihydroxyicosa-5,8,14-trienoic acid | Gamma | 0.002 | (-0.168, 0.172) | 0.983 | 0.995 | 0.016 | (-0.157, 0.169) |

**Supplementary Table S6.** Regression and Bayesian estimates comparing sex-stratified ME/CFS vs. controls.

| Metabolite | Regression Model | Male ME/CFS vs. Male Controls | | | | | |
|---|---|---|---|---|---|---|---|
| | | Estimated Coefficient | 95% CI | p-value | FDR | BayesFactor | 95% HDI |
| **Primary Metabolites (PM)** | | | | | | | |
| ornithine | Lognormal | 0.002 | (-0.027, 0.032) | 0.884 | 0.975 | 0.058 | (-0.028, 0.033) |
| tyrosine | Lognormal | 0.005 | (-0.011, 0.021) | 0.528 | 0.899 | 0.072 | (-0.010, 0.021) |
| threonine | Lognormal | 0.005 | (-0.017, 0.027) | 0.672 | 0.928 | 0.069 | (-0.016, 0.028) |
| succinic acid | Lognormal | 0.016 | (-0.014, 0.045) | 0.306 | 0.864 | 0.122 | (-0.015, 0.045) |
| lysine | Lognormal | 0.020 | (-0.017, 0.056) | 0.302 | 0.864 | 0.104 | (-0.019, 0.056) |
| phenylalanine | Lognormal | 0.014 | (-0.003, 0.032) | 0.119 | 0.864 | 0.219 | (-0.003, 0.033) |
| fructose | Lognormal | 0.033 | (-0.024, 0.090) | 0.262 | 0.864 | 0.113 | (-0.026, 0.092) |
| tagatose | Lognormal | 0.038 | (-0.001, 0.077) | 0.065 | 0.864 | 0.305 | (-0.004, 0.080) |
| leucine | Lognormal | -0.002 | (-0.019, 0.015) | 0.796 | 0.956 | 0.067 | (-0.020, 0.016) |
| glutamine | Lognormal | 0.016 | (-0.016, 0.047) | 0.343 | 0.864 | 0.101 | (-0.019, 0.047) |
| methionine | Lognormal | 0.032 | (0.004, 0.060) | 0.029 | 0.864 | 0.716 | (0.005, 0.062) |
| mannitol | Lognormal | 0.004 | (-0.049, 0.057) | 0.886 | 0.975 | 0.061 | (-0.054, 0.057) |
| pelargonic acid | Lognormal | -0.014 | (-0.034, 0.007) | 0.205 | 0.864 | 0.143 | (-0.036, 0.006) |
| arachidic acid | Lognormal | 0.029 | (-0.010, 0.068) | 0.147 | 0.864 | 0.175 | (-0.011, 0.069) |
| N-acetylputrescine | Lognormal | 0.023 | (-0.016, 0.062) | 0.250 | 0.864 | 0.125 | (-0.016, 0.067) |
| creatinine | Lognormal | 0.057 | (0.013, 0.102) | 0.016 | 0.864 | 1.482 | (0.012, 0.100) |
| urea | Lognormal | 0.002 | (-0.020, 0.024) | 0.837 | 0.962 | 0.067 | (-0.021, 0.024) |
| citrulline | Lognormal | 0.006 | (-0.011, 0.023) | 0.472 | 0.884 | 0.072 | (-0.011, 0.024) |
| erythritol | Lognormal | -0.035 | (-0.103, 0.034) | 0.327 | 0.864 | 0.105 | (-0.106, 0.038) |
| 2-deoxytetronic acid | Lognormal | 0.010 | (-0.037, 0.058) | 0.672 | 0.928 | 0.073 | (-0.036, 0.060) |
| maleimide | Lognormal | -0.021 | (-0.054, 0.011) | 0.207 | 0.864 | 0.136 | (-0.053, 0.013) |
| 2-hydroxybutanoic acid | Lognormal | 0.010 | (-0.021, 0.040) | 0.538 | 0.902 | 0.081 | (-0.021, 0.042) |
| oxoproline | Lognormal | -0.002 | (-0.009, 0.005) | 0.619 | 0.928 | 0.066 | (-0.009, 0.006) |
| 2-ketoisocaproic acid | Lognormal | -0.003 | (-0.020, 0.014) | 0.715 | 0.939 | 0.068 | (-0.020, 0.014) |
| glucose | Lognormal | 0.009 | (-0.006, 0.024) | 0.245 | 0.864 | 0.122 | (-0.007, 0.023) |
| indole-3-propionic acid | Lognormal | -0.040 | (-0.098, 0.018) | 0.180 | 0.864 | 0.158 | (-0.100, 0.016) |
| glutaric acid | Lognormal | 0.007 | (-0.026, 0.041) | 0.670 | 0.928 | 0.069 | (-0.028, 0.041) |
| alpha-ketoglutarate | Lognormal | 0.026 | (-0.007, 0.059) | 0.128 | 0.864 | 0.247 | (-0.007, 0.060) |
| tryptophan | Lognormal | 0.001 | (-0.029, 0.031) | 0.943 | 0.994 | 0.067 | (-0.028, 0.033) |
| conduritol-beta-expoxide | Lognormal | -0.028 | (-0.085, 0.030) | 0.353 | 0.869 | 0.109 | (-0.088, 0.033) |
| sucrose | Lognormal | -0.024 | (-0.087, 0.040) | 0.468 | 0.884 | 0.082 | (-0.089, 0.040) |
| linoleic acid | Lognormal | -0.008 | (-0.052, 0.036) | 0.726 | 0.945 | 0.072 | (-0.055, 0.035) |
| alanine | Lognormal | 0.006 | (-0.037, 0.049) | 0.793 | 0.956 | 0.066 | (-0.037, 0.050) |
| lysine | Lognormal | -0.033 | (-0.092, 0.026) | 0.280 | 0.864 | 0.111 | (-0.099, 0.026) |
| isoleucine | Lognormal | 0.007 | (-0.014, 0.028) | 0.511 | 0.899 | 0.088 | (-0.015, 0.028) |
| pseudo uridine | Lognormal | -0.008 | (-0.027, 0.011) | 0.428 | 0.873 | 0.096 | (-0.027, 0.012) |
| hydroxycarbamate NIST | Lognormal | -0.016 | (-0.047, 0.014) | 0.302 | 0.864 | 0.111 | (-0.050, 0.013) |
| levoglucosan | Lognormal | -0.045 | (-0.091, 0.000) | 0.057 | 0.864 | 0.415 | (-0.091, 0.003) |
| mannose | Lognormal | 0.027 | (-0.002, 0.056) | 0.077 | 0.864 | 0.454 | (-0.003, 0.059) |
| glycerol | Lognormal | 0.005 | (-0.012, 0.022) | 0.579 | 0.923 | 0.063 | (-0.013, 0.023) |
| indole-3-lactate | Lognormal | -0.023 | (-0.049, 0.003) | 0.092 | 0.864 | 0.283 | (-0.048, 0.004) |
| serine | Lognormal | 0.006 | (-0.019, 0.032) | 0.628 | 0.928 | 0.084 | (-0.020, 0.034) |
| beta-alanine | Lognormal | -0.022 | (-0.073, 0.028) | 0.394 | 0.873 | 0.095 | (-0.078, 0.027) |
| quinic acid | Lognormal | -0.052 | (-0.121, 0.018) | 0.154 | 0.864 | 0.145 | (-0.125, 0.020) |
| glycerol-alpha-phosphate | Lognormal | 0.006 | (-0.045, 0.058) | 0.814 | 0.956 | 0.072 | (-0.049, 0.060) |
| aminomalonate | Lognormal | 0.049 | (-0.003, 0.102) | 0.072 | 0.864 | 0.322 | (-0.005, 0.100) |

**Supplementary Table S6.** Regression and Bayesian estimates comparing sex-stratified ME/CFS vs. controls.

| Metabolite | Regression Model | Male ME/CFS vs. Male Controls | | | | | |
|---|---|---|---|---|---|---|---|
| | | Estimated Coefficient | 95% CI | p-value | FDR | BayesFactor | 95% HDI |
| heptadecanoic acid | Lognormal | -0.004 | (-0.027, 0.020) | 0.766 | 0.956 | 0.077 | (-0.029, 0.020) |
| phosphate | Lognormal | -0.022 | (-0.084, 0.040) | 0.493 | 0.894 | 0.086 | (-0.087, 0.043) |
| isothreonic acid | Lognormal | 0.030 | (-0.020, 0.081) | 0.248 | 0.864 | 0.120 | (-0.019, 0.084) |
| citric acid | Lognormal | 0.003 | (-0.016, 0.023) | 0.732 | 0.949 | 0.059 | (-0.018, 0.022) |
| behenic acid | Lognormal | -0.004 | (-0.027, 0.018) | 0.708 | 0.937 | 0.069 | (-0.028, 0.018) |
| palmitic acid | Lognormal | -0.009 | (-0.023, 0.004) | 0.190 | 0.864 | 0.149 | (-0.023, 0.004) |
| ribose | Lognormal | 0.021 | (-0.002, 0.044) | 0.088 | 0.864 | 0.286 | (-0.004, 0.043) |
| maltose | Lognormal | 0.015 | (-0.031, 0.060) | 0.536 | 0.902 | 0.073 | (-0.032, 0.062) |
| pentadecanoic acid | Lognormal | 0.018 | (-0.013, 0.048) | 0.260 | 0.864 | 0.137 | (-0.013, 0.049) |
| benzoic acid | Lognormal | 0.022 | (-0.011, 0.054) | 0.195 | 0.864 | 0.162 | (-0.011, 0.055) |
| 3-hydroxybutyric acid | Lognormal | 0.020 | (-0.033, 0.073) | 0.466 | 0.884 | 0.081 | (-0.037, 0.072) |
| creatine | Lognormal | 0.028 | (-0.012, 0.068) | 0.176 | 0.864 | 0.172 | (-0.014, 0.071) |
| fumaric acid | Lognormal | -0.006 | (-0.027, 0.015) | 0.564 | 0.908 | 0.074 | (-0.027, 0.016) |
| myristic acid | Lognormal | -0.008 | (-0.034, 0.018) | 0.540 | 0.902 | 0.087 | (-0.033, 0.021) |
| glycolic acid | Lognormal | -0.012 | (-0.035, 0.011) | 0.314 | 0.864 | 0.099 | (-0.035, 0.012) |
| 2-hydroxyvaleric acid | Lognormal | -0.002 | (-0.033, 0.028) | 0.891 | 0.976 | 0.066 | (-0.033, 0.031) |
| N-acetylornithine | Lognormal | -0.004 | (-0.023, 0.015) | 0.711 | 0.939 | 0.064 | (-0.022, 0.017) |
| malic acid | Lognormal | 0.024 | (-0.029, 0.077) | 0.382 | 0.873 | 0.097 | (-0.030, 0.079) |
| alloxanoic acid | Lognormal | -0.024 | (-0.070, 0.023) | 0.319 | 0.864 | 0.091 | (-0.069, 0.026) |
| 4-hydroxybutyric acid | Lognormal | -0.013 | (-0.043, 0.018) | 0.419 | 0.873 | 0.093 | (-0.045, 0.017) |
| threonic acid | Lognormal | 0.017 | (-0.011, 0.044) | 0.235 | 0.864 | 0.126 | (-0.010, 0.044) |
| gluconic acid | Lognormal | 0.013 | (-0.017, 0.043) | 0.408 | 0.873 | 0.092 | (-0.018, 0.043) |
| isopropylbenzene | Lognormal | 0.001 | (-0.032, 0.034) | 0.962 | 0.996 | 0.040 | (-0.033, 0.035) |
| 1,2,4-benzenetriol | Lognormal | -0.050 | (-0.103, 0.004) | 0.075 | 0.864 | 0.292 | (-0.106, 0.004) |
| lauric acid | Lognormal | -0.024 | (-0.066, 0.019) | 0.286 | 0.864 | 0.107 | (-0.066, 0.021) |
| pyruvic acid | Lognormal | -0.016 | (-0.055, 0.024) | 0.450 | 0.884 | 0.093 | (-0.057, 0.024) |
| salicylic acid | Lognormal | 0.005 | (-0.048, 0.059) | 0.849 | 0.966 | 0.070 | (-0.045, 0.062) |
| pyrrole-2-carboxylic acid | Lognormal | 0.019 | (-0.030, 0.068) | 0.460 | 0.884 | 0.068 | (-0.033, 0.065) |
| stearic acid | Lognormal | -0.010 | (-0.024, 0.004) | 0.160 | 0.864 | 0.168 | (-0.025, 0.004) |
| lactic acid | Lognormal | -0.008 | (-0.036, 0.020) | 0.600 | 0.926 | 0.067 | (-0.037, 0.021) |
| glucose-1-phosphate | Lognormal | 0.000 | (-0.023, 0.023) | 0.987 | 0.996 | 0.059 | (-0.024, 0.022) |
| palmitoleic acid | Lognormal | -0.003 | (-0.041, 0.034) | 0.863 | 0.968 | 0.074 | (-0.043, 0.033) |
| lyxitol | Lognormal | -0.023 | (-0.058, 0.012) | 0.213 | 0.864 | 0.142 | (-0.057, 0.014) |
| phthalic acid | Lognormal | -0.005 | (-0.030, 0.019) | 0.668 | 0.928 | 0.067 | (-0.030, 0.019) |
| glucuronic acid | Lognormal | 0.036 | (-0.007, 0.078) | 0.110 | 0.864 | 0.240 | (-0.008, 0.079) |
| threitol | Lognormal | -0.023 | (-0.055, 0.010) | 0.177 | 0.864 | 0.170 | (-0.056, 0.010) |
| capric acid | Lognormal | -0.024 | (-0.070, 0.021) | 0.302 | 0.864 | 0.106 | (-0.068, 0.024) |
| valine | Lognormal | -0.002 | (-0.019, 0.015) | 0.842 | 0.962 | 0.068 | (-0.019, 0.014) |
| glyceric acid | Lognormal | -0.001 | (-0.030, 0.028) | 0.938 | 0.993 | 0.059 | (-0.030, 0.030) |
| glutamic acid | Lognormal | 0.024 | (-0.002, 0.051) | 0.080 | 0.864 | 0.334 | (-0.001, 0.052) |
| trans-4-hydroxyproline | Lognormal | -0.016 | (-0.055, 0.024) | 0.445 | 0.883 | 0.092 | (-0.057, 0.024) |
| uric acid | Lognormal | -0.009 | (-0.042, 0.024) | 0.606 | 0.926 | 0.079 | (-0.041, 0.027) |
| adipic acid | Lognormal | -0.019 | (-0.051, 0.012) | 0.243 | 0.864 | 0.133 | (-0.050, 0.015) |
| 1-methylgalactose NIST | Lognormal | 0.012 | (-0.057, 0.081) | 0.743 | 0.949 | 0.071 | (-0.059, 0.080) |
| myo-inositol | Lognormal | -0.015 | (-0.044, 0.014) | 0.311 | 0.864 | 0.115 | (-0.044, 0.016) |
| maleic acid | Lognormal | 0.075 | (0.000, 0.150) | 0.058 | 0.864 | 0.483 | (0.002, 0.152) |
| indole-3-acetate | Lognormal | 0.004 | (-0.026, 0.034) | 0.799 | 0.956 | 0.065 | (-0.028, 0.035) |

**Supplementary Table S6.** Regression and Bayesian estimates comparing sex-stratified ME/CFS vs. controls.

| Metabolite | Regression Model | Male ME/CFS vs. Male Controls | | | | | |
|---|---|---|---|---|---|---|---|
| | | Estimated Coefficient | 95% CI | p-value | FDR | BayesFactor | 95% HDI |
| proline | Lognormal | -0.008 | (-0.051, 0.036) | 0.736 | 0.949 | 0.066 | (-0.052, 0.036) |
| 2-aminobutyric acid | Lognormal | -0.002 | (-0.027, 0.023) | 0.854 | 0.966 | 0.064 | (-0.028, 0.024) |
| oxalic acid | Lognormal | 0.007 | (-0.052, 0.067) | 0.811 | 0.956 | 0.064 | (-0.055, 0.066) |
| caprylic acid | Lognormal | -0.002 | (-0.029, 0.026) | 0.894 | 0.976 | 0.070 | (-0.029, 0.026) |
| glycine | Lognormal | -0.001 | (-0.015, 0.012) | 0.865 | 0.968 | 0.063 | (-0.013, 0.014) |
| ribonic acid | Lognormal | -0.008 | (-0.042, 0.025) | 0.627 | 0.928 | 0.085 | (-0.044, 0.028) |
| nicotinic acid | Lognormal | 0.017 | (-0.055, 0.089) | 0.648 | 0.928 | 0.070 | (-0.052, 0.096) |
| Biogenic Amines (BA) | | | | | | | |
| Alprazolam | Lognormal | 0.017 | (-0.090, 0.123) | 0.761 | 0.956 | 0.068 | (-0.100, 0.118) |
| Acyclovir | Lognormal | 0.013 | (-0.093, 0.120) | 0.806 | 0.956 | 0.066 | (-0.093, 0.117) |
| Linoleoylcarnitine | Lognormal | 0.046 | (-0.037, 0.129) | 0.284 | 0.864 | 0.118 | (-0.039, 0.131) |
| Guanine | Lognormal | 0.032 | (-0.064, 0.127) | 0.518 | 0.899 | 0.073 | (-0.067, 0.127) |
| Caffeine | Lognormal | -0.106 | (-0.210, -0.002) | 0.052 | 0.864 | 0.381 | (-0.211, -0.003) |
| 2-Methylbutyryl-L-carnitine | Lognormal | -0.016 | (-0.039, 0.007) | 0.187 | 0.864 | 0.138 | (-0.041, 0.006) |
| Acetominophen | Lognormal | 0.064 | (-0.020, 0.148) | 0.143 | 0.864 | 0.200 | (-0.023, 0.153) |
| Theobromine | Lognormal | -0.086 | (-0.174, 0.003) | 0.065 | 0.864 | 0.339 | (-0.173, 0.006) |
| Tri-2-ethylhexyl trimellitate | Lognormal | 0.014 | (-0.098, 0.126) | 0.806 | 0.956 | 0.073 | (-0.112, 0.127) |
| 7-Methylguanosine | Lognormal | -0.015 | (-0.042, 0.012) | 0.294 | 0.864 | 0.109 | (-0.041, 0.012) |
| Usnic acid | Lognormal | 0.028 | (-0.073, 0.129) | 0.593 | 0.926 | 0.075 | (-0.077, 0.128) |
| Choline cation | Lognormal | -0.009 | (-0.021, 0.002) | 0.116 | 0.864 | 0.183 | (-0.021, 0.002) |
| Piperine | Lognormal | -0.090 | (-0.190, 0.010) | 0.086 | 0.864 | 0.239 | (-0.182, 0.024) |
| Trigonelline | Lognormal | -0.069 | (-0.136, -0.003) | 0.048 | 0.864 | 0.370 | (-0.137, 0.000) |
| Oleoyl-L-carnitine | Lognormal | 0.049 | (-0.034, 0.132) | 0.256 | 0.864 | 0.134 | (-0.035, 0.130) |
| Kynurenine | Lognormal | 0.001 | (-0.027, 0.029) | 0.949 | 0.996 | 0.062 | (-0.026, 0.032) |
| Isopropylamine | Lognormal | -0.017 | (-0.038, 0.004) | 0.124 | 0.864 | 0.196 | (-0.037, 0.004) |
| Octanoylcarnitine | Lognormal | 0.002 | (-0.029, 0.033) | 0.910 | 0.978 | 0.059 | (-0.030, 0.033) |
| D-erythro-Sphingosine-1-phosphate | Lognormal | 0.043 | (-0.016, 0.102) | 0.159 | 0.864 | 0.195 | (-0.018, 0.101) |
| 1-Stearoyl-2-arachidonoyl-sn-glycero-3-phosphoserine | Lognormal | 0.110 | (-0.086, 0.307) | 0.278 | 0.864 | 0.097 | (-0.093, 0.297) |
| Albendazole sulfoxide | Lognormal | 0.099 | (-0.058, 0.256) | 0.224 | 0.864 | 0.136 | (-0.066, 0.257) |
| Ranitidine N-oxide | Lognormal | 0.072 | (-0.037, 0.180) | 0.203 | 0.864 | 0.148 | (-0.033, 0.186) |
| H-Pro-Hyp-OH | Lognormal | 0.018 | (-0.019, 0.056) | 0.343 | 0.864 | 0.101 | (-0.021, 0.057) |
| 4-Acetamidobutyric acid | Lognormal | -0.018 | (-0.057, 0.022) | 0.393 | 0.873 | 0.096 | (-0.060, 0.021) |
| 3-Methylglutarylcarnitine | Lognormal | -0.056 | (-0.134, 0.021) | 0.161 | 0.864 | 0.178 | (-0.131, 0.026) |
| 4,5,7-Trihydroxyisoflavone | Lognormal | 0.067 | (-0.094, 0.228) | 0.420 | 0.873 | 0.091 | (-0.101, 0.229) |
| 7-Hydroxywarfarin | Lognormal | -0.015 | (-0.069, 0.039) | 0.582 | 0.924 | 0.074 | (-0.071, 0.038) |
| 1-Monostearin | Lognormal | 0.044 | (-0.038, 0.125) | 0.299 | 0.864 | 0.112 | (-0.041, 0.126) |
| SDMA | Lognormal | -0.004 | (-0.022, 0.014) | 0.661 | 0.928 | 0.063 | (-0.023, 0.015) |
| Ergothioneine | Lognormal | -0.005 | (-0.076, 0.066) | 0.890 | 0.976 | 0.061 | (-0.083, 0.062) |
| 1-Oleoyl-2-acetyl-sn-glycerol | Lognormal | 0.028 | (-0.053, 0.110) | 0.497 | 0.896 | 0.085 | (-0.056, 0.116) |
| L-Citrulline | Lognormal | 0.001 | (-0.017, 0.019) | 0.906 | 0.977 | 0.067 | (-0.017, 0.020) |
| Pantoprazole | Lognormal | -0.088 | (-0.200, 0.024) | 0.130 | 0.864 | 0.208 | (-0.194, 0.030) |
| Phenylalanine | Lognormal | 0.009 | (-0.004, 0.022) | 0.164 | 0.864 | 0.164 | (-0.003, 0.023) |
| D-Turanose | Lognormal | -0.028 | (-0.093, 0.036) | 0.399 | 0.873 | 0.092 | (-0.092, 0.041) |
| Trazodone | Lognormal | -0.003 | (-0.041, 0.035) | 0.866 | 0.968 | 0.058 | (-0.042, 0.035) |
| 2,6-Diaminopimelic acid | Lognormal | 0.058 | (-0.030, 0.146) | 0.207 | 0.864 | 0.146 | (-0.032, 0.148) |
| N-.alpha.-Acetyl-L-arginine | Lognormal | -0.003 | (-0.032, 0.026) | 0.832 | 0.962 | 0.072 | (-0.034, 0.026) |
| 2-Amino-1-phenylethanol | Lognormal | 0.009 | (-0.004, 0.022) | 0.179 | 0.864 | 0.146 | (-0.005, 0.022) |

**Supplementary Table S6.** Regression and Bayesian estimates comparing sex-stratified ME/CFS vs. controls.

| Metabolite | Regression Model | Male ME/CFS vs. Male Controls | | | | | |
|---|---|---|---|---|---|---|---|
| | | Estimated Coefficient | 95% CI | p-value | FDR | BayesFactor | 95% HDI |
| Modafinil acid | Lognormal | -0.085 | (-0.197, 0.028) | 0.149 | 0.864 | 0.185 | (-0.203, 0.024) |
| Decanoyl-L-carnitine | Lognormal | 0.054 | (-0.022, 0.130) | 0.172 | 0.864 | 0.161 | (-0.026, 0.135) |
| 3-Amino-1-propanol | Lognormal | -0.021 | (-0.057, 0.015) | 0.254 | 0.864 | 0.125 | (-0.060, 0.014) |
| 1-Oleoyl-sn-glycero-3-phosphoethanolamine | Lognormal | 0.002 | (-0.082, 0.086) | 0.965 | 0.996 | 0.064 | (-0.088, 0.082) |
| H-gamma-glutamyl-glutamine | Lognormal | 0.046 | (-0.007, 0.098) | 0.098 | 0.864 | 0.261 | (-0.006, 0.101) |
| Lauroyl-L-carnitine | Lognormal | -0.030 | (-0.114, 0.054) | 0.494 | 0.896 | 0.077 | (-0.107, 0.062) |
| L-Leucine, methyl ester | Lognormal | 0.031 | (-0.054, 0.115) | 0.482 | 0.884 | 0.082 | (-0.051, 0.123) |
| Glutamic acid | Lognormal | 0.022 | (-0.007, 0.050) | 0.140 | 0.864 | 0.199 | (-0.008, 0.049) |
| Citrulline | Lognormal | 0.002 | (-0.017, 0.022) | 0.819 | 0.957 | 0.068 | (-0.017, 0.023) |
| Trimethylamine-N-oxide | Lognormal | -0.020 | (-0.076, 0.035) | 0.480 | 0.884 | 0.078 | (-0.073, 0.037) |
| 3-Hydroxypyridine | Lognormal | -0.066 | (-0.170, 0.038) | 0.223 | 0.864 | 0.089 | (-0.176, 0.041) |
| Losartan | Lognormal | 0.076 | (-0.024, 0.176) | 0.143 | 0.864 | 0.239 | (-0.021, 0.181) |
| Lamotrigine; | Lognormal | 0.105 | (-0.080, 0.289) | 0.275 | 0.864 | 0.126 | (-0.088, 0.296) |
| Gabapentin | Lognormal | 0.044 | (-0.063, 0.152) | 0.425 | 0.873 | 0.097 | (-0.063, 0.154) |
| Pyridoxine; | Lognormal | -0.036 | (-0.090, 0.018) | 0.203 | 0.864 | 0.134 | (-0.089, 0.018) |
| N-Acetylhistidine | Lognormal | -0.006 | (-0.043, 0.032) | 0.771 | 0.956 | 0.060 | (-0.044, 0.031) |
| Quetiapine | Lognormal | 0.015 | (-0.050, 0.080) | 0.658 | 0.928 | 0.077 | (-0.052, 0.081) |
| N-Methylhistidine | Lognormal | 0.004 | (-0.065, 0.073) | 0.908 | 0.977 | 0.069 | (-0.068, 0.074) |
| .beta.-Phenyl-.gamma.-aminobutyric acid | Lognormal | -0.010 | (-0.055, 0.035) | 0.661 | 0.928 | 0.068 | (-0.054, 0.036) |
| Testosterone | Lognormal | -0.019 | (-0.087, 0.049) | 0.596 | 0.926 | 0.068 | (-0.088, 0.050) |
| Thr-Ile-Arg | Lognormal | -0.010 | (-0.120, 0.099) | 0.854 | 0.966 | 0.067 | (-0.122, 0.100) |
| Dexpanthenol | Lognormal | 0.030 | (-0.026, 0.086) | 0.296 | 0.864 | 0.099 | (-0.025, 0.086) |
| 1-Stearoyl-2-arachidonoyl-sn-glycero-3-phospho-(1'-myo-inositol) | Lognormal | 0.090 | (-0.131, 0.312) | 0.429 | 0.873 | 0.091 | (-0.137, 0.320) |
| Creatinine | Lognormal | 0.003 | (-0.006, 0.011) | 0.536 | 0.902 | 0.056 | (-0.006, 0.012) |
| p-Acetamidophenyl .beta.-D-glucuronide | Lognormal | 0.036 | (-0.114, 0.185) | 0.642 | 0.928 | 0.079 | (-0.115, 0.200) |
| Ranitidine | Lognormal | 0.064 | (-0.027, 0.154) | 0.176 | 0.864 | 0.167 | (-0.030, 0.161) |
| (R)-Butyrylcarnitine | Lognormal | -0.009 | (-0.029, 0.011) | 0.393 | 0.873 | 0.080 | (-0.030, 0.012) |
| Glycocholic acid | Lognormal | 0.030 | (-0.057, 0.117) | 0.505 | 0.899 | 0.088 | (-0.057, 0.119) |
| N-Acetyl-D-norleucine | Lognormal | -0.006 | (-0.051, 0.039) | 0.797 | 0.956 | 0.052 | (-0.052, 0.043) |
| N.alpha.-Methyl-L-lysine | Lognormal | -0.014 | (-0.084, 0.057) | 0.706 | 0.937 | 0.067 | (-0.087, 0.058) |
| Betaine | Lognormal | -0.008 | (-0.018, 0.002) | 0.146 | 0.864 | 0.217 | (-0.018, 0.003) |
| Norleucine | Lognormal | 0.000 | (-0.026, 0.026) | 0.980 | 0.996 | 0.068 | (-0.026, 0.026) |
| Nudifloramide | Lognormal | 0.007 | (-0.027, 0.041) | 0.688 | 0.930 | 0.067 | (-0.027, 0.042) |
| 1-Palmitoyl-2-hydroxy-sn-glycero-3-phosphoethanolamine | Lognormal | 0.024 | (-0.065, 0.112) | 0.600 | 0.926 | 0.080 | (-0.061, 0.117) |
| Fexofenadine | Lognormal | -0.001 | (-0.044, 0.043) | 0.979 | 0.996 | 0.065 | (-0.046, 0.041) |
| Hexanoyl-L-carnitine | Lognormal | 0.009 | (-0.023, 0.041) | 0.583 | 0.924 | 0.064 | (-0.023, 0.043) |
| cis-10,11-Dihydroxy-10,11-dihydrocarbamazepine | Lognormal | -0.013 | (-0.039, 0.012) | 0.316 | 0.864 | 0.103 | (-0.039, 0.013) |
| Acetazolamide | Lognormal | -0.025 | (-0.188, 0.137) | 0.763 | 0.956 | 0.066 | (-0.185, 0.148) |
| 1-Acetyl-3-piperidinamine | Lognormal | 0.001 | (-0.032, 0.034) | 0.943 | 0.994 | 0.059 | (-0.031, 0.035) |
| Ala-Ile | Lognormal | 0.009 | (-0.023, 0.040) | 0.582 | 0.924 | 0.060 | (-0.025, 0.041) |
| Quetiapine sulfoxide | Lognormal | 0.007 | (-0.081, 0.095) | 0.877 | 0.972 | 0.072 | (-0.084, 0.093) |
| Tryptophan | Lognormal | -0.005 | (-0.024, 0.013) | 0.575 | 0.920 | 0.079 | (-0.024, 0.013) |
| Prazepam | Lognormal | 0.019 | (-0.054, 0.092) | 0.613 | 0.928 | 0.072 | (-0.053, 0.091) |
| Omeprazole sulfone N-oxide | Lognormal | -0.002 | (-0.133, 0.130) | 0.979 | 0.996 | 0.069 | (-0.136, 0.135) |
| Montelukast-1,2-diol | Lognormal | -0.019 | (-0.182, 0.143) | 0.817 | 0.957 | 0.068 | (-0.193, 0.142) |
| Diazepam | Lognormal | 0.051 | (-0.063, 0.165) | 0.384 | 0.873 | 0.084 | (-0.077, 0.164) |
| Irbesartan | Lognormal | -0.020 | (-0.037, -0.002) | 0.032 | 0.864 | 0.700 | (-0.037, -0.001) |

**Supplementary Table S6.** Regression and Bayesian estimates comparing sex-stratified ME/CFS vs. controls.

| Metabolite | Regression Model | Male ME/CFS vs. Male Controls | | | | | |
|---|---|---|---|---|---|---|---|
| | | Estimated Coefficient | 95% CI | p-value | FDR | BayesFactor | 95% HDI |
| Betonicine | Lognormal | -0.058 | (-0.186, 0.069) | 0.376 | 0.873 | 0.095 | (-0.184, 0.068) |
| Adenosine | Lognormal | 0.043 | (-0.018, 0.105) | 0.175 | 0.864 | 0.144 | (-0.020, 0.108) |
| Ethiolat | Lognormal | -0.009 | (-0.074, 0.056) | 0.790 | 0.956 | 0.068 | (-0.077, 0.058) |
| Toradol | Lognormal | -0.038 | (-0.097, 0.022) | 0.223 | 0.864 | 0.142 | (-0.099, 0.022) |
| NEPSILON,NEPSILON,NEPSILON-TRIMETHYLLYSINE | Lognormal | -0.002 | (-0.030, 0.026) | 0.870 | 0.968 | 0.063 | (-0.031, 0.026) |
| Histidine | Lognormal | 0.011 | (-0.019, 0.041) | 0.468 | 0.884 | 0.081 | (-0.018, 0.042) |
| Codeine-6-.beta.-D-glucuronide | Lognormal | -0.011 | (-0.049, 0.027) | 0.559 | 0.905 | 0.071 | (-0.052, 0.026) |
| Ethyldiethanolamine | Lognormal | 0.030 | (-0.007, 0.068) | 0.115 | 0.864 | 0.200 | (-0.009, 0.067) |
| 6-Hydroxyflavone | Lognormal | -0.033 | (-0.113, 0.048) | 0.432 | 0.876 | 0.091 | (-0.118, 0.046) |
| Omeprazole sulfone | Lognormal | -0.001 | (-0.133, 0.131) | 0.985 | 0.996 | 0.072 | (-0.138, 0.131) |
| L-Tyrosine | Lognormal | -0.016 | (-0.043, 0.010) | 0.236 | 0.864 | 0.132 | (-0.042, 0.011) |
| Aminodiphenylmethane | Lognormal | -0.017 | (-0.089, 0.056) | 0.657 | 0.928 | 0.070 | (-0.089, 0.059) |
| 1-Methylnicotinamide | Lognormal | -0.015 | (-0.056, 0.027) | 0.495 | 0.896 | 0.086 | (-0.056, 0.028) |
| Phenylacetyl-L-glutamine | Lognormal | 0.019 | (-0.034, 0.072) | 0.484 | 0.884 | 0.077 | (-0.034, 0.076) |
| Tauroursodeoxycholic acid | Lognormal | -0.011 | (-0.085, 0.062) | 0.762 | 0.956 | 0.063 | (-0.088, 0.068) |
| Tyrosine | Lognormal | -0.007 | (-0.030, 0.016) | 0.558 | 0.905 | 0.077 | (-0.031, 0.015) |
| N-(3-(Aminomethyl)benzyl)acetamidine | Lognormal | 0.015 | (-0.055, 0.084) | 0.683 | 0.929 | 0.060 | (-0.055, 0.086) |
| 3-Dehydrocarnitine | Lognormal | -0.028 | (-0.057, 0.001) | 0.065 | 0.864 | 0.303 | (-0.058, 0.005) |
| 3-Hydroxyoleylcarnitine | Lognormal | -0.012 | (-0.174, 0.150) | 0.885 | 0.975 | 0.066 | (-0.176, 0.153) |
| Pyrantel | Lognormal | -0.006 | (-0.032, 0.021) | 0.669 | 0.928 | 0.061 | (-0.034, 0.020) |
| Ile-Glu-Arg | Lognormal | 0.043 | (-0.097, 0.184) | 0.551 | 0.904 | 0.078 | (-0.101, 0.180) |
| Moxonidine | Lognormal | 0.053 | (0.002, 0.105) | 0.050 | 0.864 | 0.466 | (-0.001, 0.103) |
| Ornithine | Lognormal | 0.020 | (-0.014, 0.053) | 0.259 | 0.864 | 0.097 | (-0.016, 0.053) |
| L-Threonine | Lognormal | 0.002 | (-0.018, 0.021) | 0.854 | 0.966 | 0.063 | (-0.018, 0.022) |
| 3-Methylxanthine | Lognormal | -0.044 | (-0.130, 0.042) | 0.320 | 0.864 | 0.114 | (-0.140, 0.040) |
| (3-Carboxypropyl)trimethylammonium cation | Lognormal | -0.006 | (-0.034, 0.023) | 0.691 | 0.930 | 0.073 | (-0.035, 0.024) |
| Hydroxybupropion | Lognormal | -0.031 | (-0.094, 0.032) | 0.338 | 0.864 | 0.098 | (-0.097, 0.027) |
| Glaucine | Lognormal | -0.007 | (-0.035, 0.021) | 0.617 | 0.928 | 0.069 | (-0.035, 0.022) |
| L-Cystine | Lognormal | 0.001 | (-0.047, 0.048) | 0.980 | 0.996 | 0.051 | (-0.048, 0.047) |
| Methacholine cation | Lognormal | -0.032 | (-0.067, 0.003) | 0.076 | 0.864 | 0.327 | (-0.069, 0.004) |
| Urea | Lognormal | -0.007 | (-0.024, 0.009) | 0.395 | 0.873 | 0.081 | (-0.025, 0.010) |
| 4-Fluoro-.alpha.-pyrrolidinobutiophenone | Lognormal | -0.010 | (-0.055, 0.036) | 0.674 | 0.928 | 0.077 | (-0.057, 0.035) |
| Benthiavalicarb-isopropyl | Lognormal | -0.002 | (-0.080, 0.076) | 0.961 | 0.996 | 0.070 | (-0.084, 0.077) |
| Modafinil | Lognormal | -0.043 | (-0.127, 0.041) | 0.318 | 0.864 | 0.101 | (-0.118, 0.043) |
| Palmitamide | Lognormal | -0.002 | (-0.069, 0.065) | 0.955 | 0.996 | 0.068 | (-0.070, 0.065) |
| alpha-Methylhistidine; | Lognormal | 0.013 | (-0.079, 0.105) | 0.777 | 0.956 | 0.062 | (-0.079, 0.109) |
| Ranitidine-S-oxide | Lognormal | 0.079 | (-0.039, 0.198) | 0.198 | 0.864 | 0.144 | (-0.049, 0.197) |
| Metoprolol acid | Lognormal | 0.062 | (-0.068, 0.192) | 0.356 | 0.871 | 0.089 | (-0.069, 0.195) |
| 7.alpha.-Hydroxy-3-oxo-4-cholestenoic acid | Lognormal | -0.007 | (-0.055, 0.042) | 0.781 | 0.956 | 0.073 | (-0.054, 0.045) |
| 5'-S-Methyl-5'-thioadenosine | Lognormal | 0.011 | (-0.143, 0.166) | 0.887 | 0.975 | 0.057 | (-0.156, 0.159) |
| Stachydrine | Lognormal | -0.084 | (-0.154, -0.014) | 0.023 | 0.864 | 0.954 | (-0.154, -0.016) |
| Glycodeoxycholic acid | Lognormal | 0.027 | (-0.061, 0.116) | 0.548 | 0.904 | 0.074 | (-0.060, 0.117) |
| Carnitine | Lognormal | -0.003 | (-0.017, 0.010) | 0.655 | 0.928 | 0.064 | (-0.017, 0.011) |
| Topiramate | Lognormal | 0.000 | (-0.053, 0.052) | 0.987 | 0.996 | 0.064 | (-0.052, 0.054) |
| Meprobamate | Lognormal | 0.000 | (-0.075, 0.075) | 0.999 | 1.000 | 0.068 | (-0.074, 0.079) |
| DL-Indole-3-lactic acid | Lognormal | 0.001 | (-0.017, 0.019) | 0.934 | 0.992 | 0.061 | (-0.017, 0.019) |
| Ethylenediaminetetraacetic acid | Lognormal | 0.068 | (-0.084, 0.220) | 0.384 | 0.873 | 0.100 | (-0.093, 0.210) |

**Supplementary Table S6.** Regression and Bayesian estimates comparing sex-stratified ME/CFS vs. controls.

| Metabolite | Regression Model | Male ME/CFS vs. Male Controls | | | | | |
|---|---|---|---|---|---|---|---|
| | | Estimated Coefficient | 95% CI | p-value | FDR | BayesFactor | 95% HDI |
| Gly-Pro-Arg | Lognormal | 0.048 | (-0.104, 0.199) | 0.542 | 0.902 | 0.079 | (-0.107, 0.190) |
| Pipecolic acid | Lognormal | -0.004 | (-0.059, 0.051) | 0.887 | 0.975 | 0.052 | (-0.062, 0.051) |
| L-Cysteine-glutathione disulfide | Lognormal | 0.024 | (-0.033, 0.081) | 0.418 | 0.873 | 0.066 | (-0.038, 0.083) |
| 3-Cysteinylacetaminophen | Lognormal | 0.216 | (0.030, 0.401) | 0.028 | 0.864 | 0.718 | (0.033, 0.408) |
| Ezetimibe | Lognormal | -0.028 | (-0.090, 0.034) | 0.378 | 0.873 | 0.093 | (-0.091, 0.038) |
| 1-Methyl-L-histidine | Lognormal | 0.032 | (-0.066, 0.131) | 0.526 | 0.899 | 0.086 | (-0.065, 0.135) |
| Carbamazepine | Lognormal | 0.048 | (-0.035, 0.132) | 0.265 | 0.864 | 0.114 | (-0.037, 0.135) |
| 5'-S-Methylthioadenosine | Lognormal | 0.052 | (-0.112, 0.215) | 0.540 | 0.902 | 0.068 | (-0.106, 0.218) |
| Dinor-12-oxophytodienoic acid | Lognormal | 0.029 | (-0.077, 0.135) | 0.596 | 0.926 | 0.075 | (-0.076, 0.134) |
| Atorvastatin | Lognormal | -0.016 | (-0.047, 0.015) | 0.315 | 0.864 | 0.097 | (-0.048, 0.016) |
| N-Methylproline | Lognormal | -0.011 | (-0.065, 0.043) | 0.699 | 0.935 | 0.064 | (-0.066, 0.042) |
| 4'-Methyl-N-methylhexanophenone | Lognormal | -0.119 | (-0.229, -0.009) | 0.040 | 0.864 | 0.665 | (-0.229, -0.014) |
| trans-3'-Hydroxycotinine | Lognormal | -0.013 | (-0.080, 0.053) | 0.699 | 0.935 | 0.057 | (-0.084, 0.056) |
| Guanidine | Lognormal | 0.009 | (-0.022, 0.039) | 0.572 | 0.917 | 0.076 | (-0.022, 0.040) |
| Telmisartan | Lognormal | 0.000 | (-0.032, 0.033) | 0.993 | 0.997 | 0.055 | (-0.034, 0.032) |
| Theanine; | Lognormal | 0.018 | (-0.037, 0.073) | 0.528 | 0.899 | 0.068 | (-0.038, 0.072) |
| Methylgallate | Lognormal | -0.008 | (-0.159, 0.143) | 0.918 | 0.983 | 0.068 | (-0.170, 0.141) |
| Avobenzone | Lognormal | -0.020 | (-0.067, 0.028) | 0.420 | 0.873 | 0.071 | (-0.062, 0.033) |
| 3,4-Dimethoxybenzaldehyde | Lognormal | 0.014 | (-0.049, 0.078) | 0.661 | 0.928 | 0.064 | (-0.053, 0.078) |
| Trimethoprim | Lognormal | -0.015 | (-0.047, 0.017) | 0.369 | 0.873 | 0.095 | (-0.048, 0.019) |
| Ticlopidine | Lognormal | -0.073 | (-0.235, 0.090) | 0.386 | 0.873 | 0.091 | (-0.232, 0.108) |
| N.epsilon.-Acetyl-L-lysine | Lognormal | 0.008 | (-0.016, 0.033) | 0.506 | 0.899 | 0.086 | (-0.017, 0.034) |
| 2,2-Bishydroxymethyl]-2,2',2''-nitrilotriethanol | Lognormal | 0.023 | (-0.038, 0.083) | 0.468 | 0.884 | 0.086 | (-0.041, 0.084) |
| Betaine aldehyde cation | Lognormal | -0.007 | (-0.087, 0.073) | 0.868 | 0.968 | 0.048 | (-0.090, 0.073) |
| Pantothenic acid | Lognormal | 0.036 | (-0.006, 0.079) | 0.102 | 0.864 | 0.243 | (-0.007, 0.081) |
| D-Pyroglutamic acid | Lognormal | 0.007 | (-0.001, 0.014) | 0.085 | 0.864 | 0.274 | (-0.001, 0.015) |
| 2-Indolinone | Lognormal | -0.023 | (-0.055, 0.009) | 0.161 | 0.864 | 0.153 | (-0.054, 0.010) |
| Pyridoxal | Lognormal | 0.006 | (-0.029, 0.042) | 0.724 | 0.944 | 0.070 | (-0.028, 0.044) |
| (2R)-3-Hydroxyisovaleroylcarnitine | Lognormal | 0.001 | (-0.036, 0.038) | 0.958 | 0.996 | 0.054 | (-0.036, 0.039) |
| Arginine | Lognormal | 0.015 | (-0.004, 0.035) | 0.134 | 0.864 | 0.163 | (-0.006, 0.034) |
| Metoprolol | Lognormal | 0.046 | (-0.049, 0.141) | 0.350 | 0.864 | 0.102 | (-0.049, 0.150) |
| N-Acetyl-L-carnosine | Lognormal | -0.001 | (-0.038, 0.035) | 0.942 | 0.994 | 0.063 | (-0.042, 0.034) |
| 3-Hydroxybutyrylcarnitine | Lognormal | 0.011 | (-0.043, 0.066) | 0.692 | 0.930 | 0.075 | (-0.043, 0.065) |
| 4-Aminomethylcyclohexanecarboxylic acid; | Lognormal | 0.012 | (-0.051, 0.076) | 0.703 | 0.937 | 0.067 | (-0.050, 0.076) |
| D-.alpha.-Cyclohexylglycine | Lognormal | -0.029 | (-0.089, 0.031) | 0.350 | 0.864 | 0.092 | (-0.091, 0.031) |
| Urocanic acid; | Lognormal | 0.010 | (-0.023, 0.043) | 0.557 | 0.905 | 0.068 | (-0.023, 0.044) |
| L-Carnitine | Lognormal | 0.007 | (-0.012, 0.026) | 0.473 | 0.884 | 0.078 | (-0.012, 0.027) |
| Metformin | Lognormal | 0.005 | (-0.031, 0.041) | 0.778 | 0.956 | 0.055 | (-0.029, 0.041) |
| Cotinine N-.beta.-D-glucuronide | Lognormal | -0.009 | (-0.141, 0.123) | 0.894 | 0.976 | 0.052 | (-0.142, 0.125) |
| Glutamine | Lognormal | 0.007 | (-0.001, 0.016) | 0.099 | 0.864 | 0.269 | (-0.001, 0.016) |
| Heptadecasphing-4-enine | Lognormal | 0.059 | (-0.168, 0.287) | 0.613 | 0.928 | 0.065 | (-0.180, 0.284) |
| Ser-Tyr-Lys | Lognormal | 0.005 | (-0.120, 0.129) | 0.941 | 0.994 | 0.060 | (-0.120, 0.130) |
| 3-Aminoquinoline | Lognormal | 0.030 | (-0.060, 0.119) | 0.518 | 0.899 | 0.071 | (-0.070, 0.115) |
| Biliverden | Lognormal | 0.009 | (-0.031, 0.049) | 0.661 | 0.928 | 0.069 | (-0.031, 0.049) |
| Borrelidin | Lognormal | -0.027 | (-0.127, 0.073) | 0.606 | 0.926 | 0.076 | (-0.132, 0.081) |
| Creatine | Lognormal | 0.028 | (0.001, 0.056) | 0.050 | 0.864 | 0.447 | (0.000, 0.057) |
| Isopentenyladenine | Lognormal | 0.005 | (-0.013, 0.022) | 0.592 | 0.926 | 0.078 | (-0.012, 0.023) |

**Supplementary Table S6.** Regression and Bayesian estimates comparing sex-stratified ME/CFS vs. controls.

| Metabolite | Regression Model | Male ME/CFS vs. Male Controls | | | | | |
|---|---|---|---|---|---|---|---|
| | | Estimated Coefficient | 95% CI | p-value | FDR | BayesFactor | 95% HDI |
| Indole-3-propionic acid | Lognormal | -0.048 | (-0.114, 0.018) | 0.163 | 0.864 | 0.175 | (-0.118, 0.017) |
| D-Fructose | Lognormal | 0.004 | (-0.012, 0.019) | 0.647 | 0.928 | 0.072 | (-0.012, 0.019) |
| rac-4-Sulfoxypropranolol | Lognormal | 0.020 | (-0.049, 0.088) | 0.577 | 0.921 | 0.079 | (-0.047, 0.091) |
| 3-Pyridinemthanol | Lognormal | -0.004 | (-0.037, 0.030) | 0.828 | 0.962 | 0.062 | (-0.038, 0.029) |
| Levocetirizine; | Lognormal | 0.002 | (-0.140, 0.144) | 0.981 | 0.996 | 0.071 | (-0.150, 0.136) |
| .epsilon.-Caprolactam | Lognormal | 0.009 | (-0.030, 0.047) | 0.653 | 0.928 | 0.055 | (-0.030, 0.048) |
| N-Acetylalanine | Lognormal | 0.016 | (-0.093, 0.125) | 0.780 | 0.956 | 0.075 | (-0.095, 0.129) |
| 1,2-Dimethylimidazole | Lognormal | -0.011 | (-0.042, 0.019) | 0.461 | 0.884 | 0.081 | (-0.044, 0.016) |
| R-(-)-O-Desmethylvenlafaxine | Lognormal | 0.000 | (-0.089, 0.089) | 1.000 | 1.000 | 0.065 | (-0.088, 0.095) |
| Naproxen | Lognormal | 0.027 | (-0.074, 0.129) | 0.601 | 0.926 | 0.082 | (-0.076, 0.132) |
| N8-Acetylspermidine | Lognormal | -0.002 | (-0.032, 0.028) | 0.899 | 0.976 | 0.063 | (-0.032, 0.029) |
| 1-Stearoyl-2-hydroxy-sn-glycero-3-phosphoethanolamine | Lognormal | 0.032 | (-0.065, 0.130) | 0.517 | 0.899 | 0.080 | (-0.065, 0.133) |
| Propionylcarnitine | Lognormal | -0.008 | (-0.025, 0.008) | 0.339 | 0.864 | 0.088 | (-0.026, 0.010) |
| Matrine | Lognormal | 0.038 | (-0.060, 0.137) | 0.449 | 0.884 | 0.087 | (-0.059, 0.139) |
| Trileptal | Lognormal | 0.006 | (-0.023, 0.035) | 0.677 | 0.929 | 0.065 | (-0.024, 0.038) |
| Androstan-3-ol-17-one 3-glucuronide | Lognormal | 0.005 | (-0.052, 0.062) | 0.861 | 0.967 | 0.059 | (-0.049, 0.066) |
| Serotonin | Lognormal | 0.012 | (-0.026, 0.050) | 0.543 | 0.902 | 0.053 | (-0.028, 0.049) |
| Lansoprazole | Lognormal | -0.165 | (-0.330, 0.000) | 0.056 | 0.864 | 0.390 | (-0.332, 0.010) |
| Atenolol | Lognormal | 0.018 | (-0.087, 0.123) | 0.739 | 0.949 | 0.067 | (-0.096, 0.121) |
| 1-Acetyl-4-piperidinamine | Lognormal | 0.035 | (-0.042, 0.113) | 0.376 | 0.873 | 0.095 | (-0.043, 0.116) |
| Homoarginine; | Lognormal | 0.037 | (0.005, 0.069) | 0.027 | 0.864 | 0.545 | (0.003, 0.070) |
| 1-Phenylpyrrolidine | Lognormal | 0.003 | (-0.047, 0.053) | 0.909 | 0.977 | 0.062 | (-0.047, 0.055) |
| Acetyl-DL-carnitine | Lognormal | 0.007 | (-0.021, 0.034) | 0.624 | 0.928 | 0.068 | (-0.022, 0.035) |
| 2-Hydroxyibuprofen | Lognormal | -0.022 | (-0.092, 0.047) | 0.531 | 0.899 | 0.068 | (-0.096, 0.048) |
| Milnacipran | Lognormal | 0.031 | (-0.042, 0.103) | 0.413 | 0.873 | 0.081 | (-0.047, 0.102) |
| Triptolide | Lognormal | -0.018 | (-0.049, 0.013) | 0.254 | 0.864 | 0.124 | (-0.052, 0.012) |
| 2,2',2''-Nitrilotriethanol | Lognormal | 0.007 | (-0.023, 0.036) | 0.657 | 0.928 | 0.069 | (-0.023, 0.038) |
| Albendazole | Lognormal | 0.017 | (-0.015, 0.048) | 0.307 | 0.864 | 0.091 | (-0.015, 0.050) |
| Scopoletin | Lognormal | -0.045 | (-0.204, 0.115) | 0.588 | 0.926 | 0.065 | (-0.210, 0.109) |
| Mefenorex | Lognormal | 0.010 | (-0.084, 0.105) | 0.835 | 0.962 | 0.061 | (-0.088, 0.105) |
| Diphenhydramine | Lognormal | -0.003 | (-0.036, 0.030) | 0.859 | 0.967 | 0.054 | (-0.036, 0.030) |
| 3-(1-Pyrazolyl)-alanine | Lognormal | 0.010 | (-0.107, 0.126) | 0.871 | 0.968 | 0.064 | (-0.101, 0.136) |
| Methionine | Lognormal | 0.008 | (-0.010, 0.025) | 0.389 | 0.873 | 0.094 | (-0.010, 0.026) |
| Hypoxanthine | Lognormal | -0.002 | (-0.033, 0.028) | 0.887 | 0.975 | 0.047 | (-0.033, 0.029) |
| Tapentadol-.beta.-D-glucuronide | Lognormal | -0.053 | (-0.214, 0.108) | 0.523 | 0.899 | 0.091 | (-0.219, 0.120) |
| Methioninesulfoxide | Lognormal | 0.038 | (0.004, 0.072) | 0.036 | 0.864 | 0.652 | (0.001, 0.069) |
| Ondansetron | Lognormal | -0.005 | (-0.028, 0.018) | 0.668 | 0.928 | 0.061 | (-0.028, 0.018) |
| threo-Dihydrobupropion | Lognormal | -0.011 | (-0.061, 0.040) | 0.681 | 0.929 | 0.065 | (-0.060, 0.041) |
| Isoleucine | Lognormal | 0.006 | (-0.019, 0.030) | 0.651 | 0.928 | 0.070 | (-0.020, 0.030) |
| Esomeprazole | Lognormal | 0.010 | (-0.034, 0.055) | 0.653 | 0.928 | 0.074 | (-0.035, 0.058) |
| Meloxicam | Lognormal | 0.078 | (-0.181, 0.337) | 0.560 | 0.905 | 0.082 | (-0.189, 0.345) |
| 3,5-Dihydroxyphenylglycine | Lognormal | 0.007 | (-0.044, 0.057) | 0.798 | 0.956 | 0.062 | (-0.041, 0.061) |
| Temazepam | Lognormal | 0.026 | (-0.052, 0.103) | 0.521 | 0.899 | 0.083 | (-0.059, 0.099) |
| Lauric acid diethanolamide | Lognormal | 0.022 | (-0.078, 0.122) | 0.665 | 0.928 | 0.072 | (-0.080, 0.126) |
| 4-Pyridoxic acid; | Lognormal | 0.050 | (-0.035, 0.134) | 0.260 | 0.864 | 0.124 | (-0.037, 0.138) |
| Phenylacetylglutamine | Lognormal | 0.016 | (-0.034, 0.065) | 0.539 | 0.902 | 0.071 | (-0.039, 0.065) |
| Penciclovir | Lognormal | 0.001 | (-0.041, 0.043) | 0.963 | 0.996 | 0.056 | (-0.040, 0.045) |

**Supplementary Table S6.** Regression and Bayesian estimates comparing sex-stratified ME/CFS vs. controls.

| Metabolite | Regression Model | Male ME/CFS vs. Male Controls | | | | | |
|---|---|---|---|---|---|---|---|
| | | Estimated Coefficient | 95% CI | p-value | FDR | BayesFactor | 95% HDI |
| 6-Methoxynaphthaleneacetic acid | Lognormal | -0.018 | (-0.050, 0.014) | 0.280 | 0.864 | 0.119 | (-0.051, 0.017) |
| 1-Methyladenosine A | Lognormal | -0.003 | (-0.019, 0.013) | 0.695 | 0.932 | 0.064 | (-0.020, 0.012) |
| Coniferylaldehyde | Lognormal | 0.020 | (-0.018, 0.058) | 0.303 | 0.864 | 0.107 | (-0.020, 0.060) |
| Bradykinin | Lognormal | 0.258 | (-0.018, 0.535) | 0.075 | 0.864 | 0.301 | (-0.004, 0.560) |
| Lysine | Lognormal | 0.010 | (-0.009, 0.029) | 0.300 | 0.864 | 0.075 | (-0.010, 0.029) |
| Benzophenone-3 | Lognormal | -0.023 | (-0.107, 0.061) | 0.597 | 0.926 | 0.080 | (-0.110, 0.058) |
| Sulfamethoxazole | Lognormal | -0.066 | (-0.204, 0.071) | 0.350 | 0.864 | 0.102 | (-0.210, 0.067) |
| Cyclo(Leu-Pro) | Lognormal | -0.001 | (-0.054, 0.052) | 0.971 | 0.996 | 0.066 | (-0.051, 0.055) |
| N.epsilon.-Methyl-L-lysine | Lognormal | 0.030 | (-0.029, 0.088) | 0.328 | 0.864 | 0.102 | (-0.032, 0.086) |
| Proline | Lognormal | -0.017 | (-0.039, 0.006) | 0.164 | 0.864 | 0.177 | (-0.038, 0.009) |
| **Complex Lipids (CL)** | | | | | | | |
| PC (p-34:2)/PC (o-34:3) - ESI(+) | Lognormal | -0.010 | (-0.031, 0.011) | 0.343 | 0.864 | 0.106 | (-0.031, 0.012) |
| PE (p-36:2)/PE (o-36:3) - ESI(+) | Lognormal | -0.020 | (-0.059, 0.018) | 0.310 | 0.864 | 0.117 | (-0.062, 0.018) |
| PC (p-34:1)/PC (o-34:2) | Lognormal | -0.009 | (-0.028, 0.010) | 0.365 | 0.873 | 0.091 | (-0.030, 0.011) |
| PC (36:4) A - ESI(+) | Lognormal | -0.015 | (-0.034, 0.004) | 0.128 | 0.864 | 0.174 | (-0.033, 0.005) |
| PC (36:2) | Lognormal | -0.004 | (-0.011, 0.003) | 0.271 | 0.864 | 0.106 | (-0.011, 0.004) |
| PC 34:4e | Lognormal | -0.007 | (-0.040, 0.025) | 0.666 | 0.928 | 0.073 | (-0.039, 0.026) |
| PC (36:4) A - ESI(-) | Lognormal | -0.014 | (-0.035, 0.006) | 0.181 | 0.864 | 0.151 | (-0.035, 0.007) |
| PC (34:2) - ESI(+) | Lognormal | 0.000 | (-0.006, 0.006) | 0.900 | 0.976 | 0.061 | (-0.006, 0.007) |
| PC (p-34:2)/PC (o-34:3) - ESI(-) | Lognormal | -0.016 | (-0.038, 0.005) | 0.144 | 0.864 | 0.169 | (-0.037, 0.007) |
| PC (p-36:4)/PC (o-36:5) - ESI(-) | Lognormal | -0.019 | (-0.046, 0.008) | 0.176 | 0.864 | 0.169 | (-0.046, 0.010) |
| PC (p-34:1)/PC (o-34:2) A | Lognormal | -0.013 | (-0.042, 0.015) | 0.363 | 0.873 | 0.107 | (-0.043, 0.014) |
| PC (p-36:1)/PC (o-36:2) | Lognormal | -0.020 | (-0.061, 0.021) | 0.348 | 0.864 | 0.082 | (-0.060, 0.025) |
| PE (p-34:2)/PE (o-34:3) | Lognormal | -0.032 | (-0.090, 0.027) | 0.291 | 0.864 | 0.119 | (-0.091, 0.026) |
| SM (d40:3) | Lognormal | -0.002 | (-0.040, 0.037) | 0.933 | 0.992 | 0.060 | (-0.042, 0.036) |
| PE (p-36:2)/PE (o-36:3) - ESI(-) | Lognormal | -0.028 | (-0.077, 0.020) | 0.259 | 0.864 | 0.138 | (-0.078, 0.021) |
| PE (p-36:4)/PE (o-36:5) - ESI(-) | Lognormal | -0.021 | (-0.060, 0.017) | 0.282 | 0.864 | 0.118 | (-0.062, 0.017) |
| CE (18:2) | Lognormal | 0.003 | (-0.010, 0.017) | 0.637 | 0.928 | 0.065 | (-0.010, 0.018) |
| PC (36:5) A | Lognormal | -0.020 | (-0.075, 0.035) | 0.480 | 0.884 | 0.076 | (-0.078, 0.033) |
| LPC (18:2) - ESI(-) | Lognormal | -0.022 | (-0.043, -0.001) | 0.048 | 0.864 | 0.426 | (-0.043, -0.001) |
| PC (34:2) - ESI(-) | Lognormal | -0.011 | (-0.025, 0.004) | 0.162 | 0.864 | 0.187 | (-0.026, 0.005) |
| PC (p-36:2)/PC (o-36:3) | Lognormal | -0.007 | (-0.033, 0.020) | 0.636 | 0.928 | 0.076 | (-0.033, 0.021) |
| TG (56:6) | Lognormal | 0.004 | (-0.011, 0.019) | 0.615 | 0.928 | 0.073 | (-0.013, 0.019) |
| PC (p-36:3)/PC (o-36:4) - ESI(-) | Lognormal | -0.016 | (-0.037, 0.005) | 0.145 | 0.864 | 0.204 | (-0.038, 0.005) |
| TG (55:6) | Lognormal | -0.001 | (-0.032, 0.029) | 0.936 | 0.992 | 0.065 | (-0.033, 0.030) |
| PC (36:5)A | Lognormal | -0.018 | (-0.050, 0.013) | 0.258 | 0.864 | 0.102 | (-0.051, 0.014) |
| SM (d34:2) - ESI(-) | Lognormal | -0.006 | (-0.023, 0.010) | 0.460 | 0.884 | 0.092 | (-0.024, 0.010) |
| SM (d34:1) - ESI(-) | Lognormal | -0.006 | (-0.021, 0.009) | 0.441 | 0.881 | 0.093 | (-0.020, 0.009) |
| LPE (18:2) - ESI(-) | Lognormal | -0.033 | (-0.074, 0.008) | 0.119 | 0.864 | 0.194 | (-0.073, 0.010) |
| TG (54:6) C | Lognormal | -0.006 | (-0.031, 0.020) | 0.672 | 0.928 | 0.069 | (-0.032, 0.021) |
| PC (34:3) | Lognormal | -0.018 | (-0.046, 0.009) | 0.199 | 0.864 | 0.178 | (-0.048, 0.010) |
| PE (p-38:5)/PE (o-38:6) - ESI(-) | Lognormal | -0.025 | (-0.061, 0.011) | 0.185 | 0.864 | 0.142 | (-0.059, 0.015) |
| LPE (18:2) - ESI(+) | Lognormal | -0.022 | (-0.053, 0.009) | 0.176 | 0.864 | 0.164 | (-0.054, 0.009) |
| SM (d38:2) - ESI(-) | Lognormal | -0.005 | (-0.028, 0.018) | 0.678 | 0.929 | 0.068 | (-0.027, 0.019) |
| LPC (18:2) - ESI(+) | Lognormal | -0.016 | (-0.037, 0.004) | 0.123 | 0.864 | 0.176 | (-0.039, 0.004) |
| PE (p-36:1)/PE (o-36:2) - ESI(-) | Lognormal | -0.035 | (-0.113, 0.043) | 0.390 | 0.873 | 0.112 | (-0.116, 0.044) |
| SM (d39:1) - ESI(-) | Lognormal | -0.023 | (-0.052, 0.006) | 0.124 | 0.864 | 0.221 | (-0.052, 0.007) |

**Supplementary Table S6.** Regression and Bayesian estimates comparing sex-stratified ME/CFS vs. controls.

| Metabolite | Regression Model | Male ME/CFS vs. Male Controls | | | | | |
|---|---|---|---|---|---|---|---|
| | | Estimated Coefficient | 95% CI | p-value | FDR | BayesFactor | 95% HDI |
| PC (32:2) - ESI(-) | Lognormal | -0.041 | (-0.084, 0.001) | 0.065 | 0.864 | 0.438 | (-0.085, 0.000) |
| PC (p-38:4)/PC (o-38:5) A | Lognormal | -0.012 | (-0.030, 0.005) | 0.174 | 0.864 | 0.173 | (-0.030, 0.005) |
| TG (54:7) B | Lognormal | -0.012 | (-0.045, 0.021) | 0.471 | 0.884 | 0.091 | (-0.046, 0.023) |
| PC (33:2) - ESI(+) | Lognormal | 0.009 | (-0.009, 0.026) | 0.346 | 0.864 | 0.102 | (-0.009, 0.027) |
| AC (10:1) | Lognormal | 0.007 | (-0.032, 0.047) | 0.716 | 0.939 | 0.076 | (-0.030, 0.049) |
| PC (34:3) B | Lognormal | -0.011 | (-0.031, 0.010) | 0.309 | 0.864 | 0.119 | (-0.032, 0.011) |
| LPC (18:3) | Lognormal | -0.041 | (-0.081, 0.000) | 0.056 | 0.864 | 0.451 | (-0.084, 0.000) |
| PE (p-38:4)/PE (o-38:5) | Lognormal | -0.016 | (-0.055, 0.024) | 0.441 | 0.881 | 0.085 | (-0.057, 0.025) |
| CE (20:2) | Lognormal | -0.012 | (-0.046, 0.023) | 0.506 | 0.899 | 0.075 | (-0.047, 0.025) |
| SM (d40:1) - ESI(-) | Lognormal | -0.016 | (-0.036, 0.005) | 0.136 | 0.864 | 0.205 | (-0.036, 0.005) |
| PC (o-34:0) | Lognormal | 0.004 | (-0.021, 0.030) | 0.748 | 0.951 | 0.062 | (-0.021, 0.030) |
| PC (35:2) | Lognormal | 0.003 | (-0.019, 0.025) | 0.788 | 0.956 | 0.061 | (-0.019, 0.027) |
| Ceramide (d42:1) - ESI (+) | Lognormal | -0.007 | (-0.022, 0.009) | 0.418 | 0.873 | 0.098 | (-0.022, 0.010) |
| FA (16:1) (palmitoleic acid) | Lognormal | 0.020 | (-0.010, 0.050) | 0.200 | 0.864 | 0.133 | (-0.010, 0.052) |
| PE (p-36:4)/PE (o-36:5) - ESI(+) | Lognormal | -0.011 | (-0.047, 0.024) | 0.531 | 0.899 | 0.071 | (-0.048, 0.024) |
| PC (32:2) - ESI(+) | Lognormal | -0.016 | (-0.049, 0.018) | 0.364 | 0.873 | 0.106 | (-0.048, 0.021) |
| TG (56:7) B | Lognormal | 0.003 | (-0.024, 0.030) | 0.832 | 0.962 | 0.067 | (-0.025, 0.030) |
| PC (p-38:3)/PC (o-38:4) - ESI(-) | Lognormal | -0.020 | (-0.046, 0.006) | 0.144 | 0.864 | 0.194 | (-0.046, 0.006) |
| SM (d40:2) B - ESI(+) | Lognormal | 0.002 | (-0.011, 0.015) | 0.778 | 0.956 | 0.068 | (-0.011, 0.015) |
| PE (p-40:4)/PE (o-40:5) A | Lognormal | -0.005 | (-0.054, 0.045) | 0.856 | 0.966 | 0.044 | (-0.053, 0.046) |
| TG (54:5) B | Lognormal | -0.009 | (-0.028, 0.011) | 0.395 | 0.873 | 0.094 | (-0.028, 0.012) |
| SM (d40:2) B - ESI(-) | Lognormal | -0.010 | (-0.031, 0.011) | 0.347 | 0.864 | 0.108 | (-0.031, 0.013) |
| TG (58:8) | Lognormal | 0.006 | (-0.029, 0.040) | 0.745 | 0.949 | 0.064 | (-0.030, 0.041) |
| PC (35:3) | Lognormal | 0.002 | (-0.012, 0.015) | 0.803 | 0.956 | 0.054 | (-0.013, 0.015) |
| PC (p-36:4)/PC (o-36:5) - ESI(+) | Lognormal | -0.005 | (-0.024, 0.015) | 0.657 | 0.928 | 0.082 | (-0.024, 0.017) |
| TG (56:5) B | Lognormal | -0.004 | (-0.028, 0.021) | 0.765 | 0.956 | 0.071 | (-0.030, 0.021) |
| TG (53:0) | Lognormal | -0.015 | (-0.045, 0.016) | 0.357 | 0.872 | 0.108 | (-0.049, 0.015) |
| SM (d33:1) - ESI(-) | Lognormal | -0.015 | (-0.040, 0.009) | 0.216 | 0.864 | 0.159 | (-0.040, 0.008) |
| PC (p-32:1)/PC (o-32:2) | Lognormal | -0.005 | (-0.027, 0.016) | 0.616 | 0.928 | 0.069 | (-0.027, 0.016) |
| TG (62:4) | Lognormal | -0.011 | (-0.067, 0.044) | 0.691 | 0.930 | 0.074 | (-0.066, 0.050) |
| PE (36:2) | Lognormal | -0.026 | (-0.078, 0.027) | 0.346 | 0.864 | 0.105 | (-0.079, 0.029) |
| PE (p-36:1)/PE (o-36:2) - ESI(+) | Lognormal | -0.019 | (-0.063, 0.024) | 0.392 | 0.873 | 0.104 | (-0.066, 0.024) |
| PE (p-34:1)/PE (o-34:2) - ESI(-) | Lognormal | -0.014 | (-0.054, 0.025) | 0.480 | 0.884 | 0.090 | (-0.059, 0.022) |
| TG (50:0) | Lognormal | -0.007 | (-0.049, 0.035) | 0.741 | 0.949 | 0.068 | (-0.050, 0.035) |
| SM (d32:1) - ESI(-) | Lognormal | -0.024 | (-0.050, 0.002) | 0.074 | 0.864 | 0.385 | (-0.051, 0.003) |
| LPE (20:4) - ESI(-) | Lognormal | -0.039 | (-0.076, -0.003) | 0.042 | 0.864 | 0.715 | (-0.076, -0.004) |
| PE (p-38:5)/PE (o-38:6) - ESI(+) | Lognormal | -0.010 | (-0.038, 0.017) | 0.464 | 0.884 | 0.077 | (-0.038, 0.017) |
| SM (d42:3) - ESI(-) | Lognormal | -0.001 | (-0.019, 0.018) | 0.949 | 0.996 | 0.063 | (-0.021, 0.017) |
| Ceramide (d41:1) - ESI(-) | Lognormal | -0.024 | (-0.049, 0.002) | 0.076 | 0.864 | 0.331 | (-0.050, 0.002) |
| PC (p-32:0)/PC (o-32:1) - ESI(+) | Lognormal | 0.010 | (-0.009, 0.028) | 0.304 | 0.864 | 0.117 | (-0.010, 0.028) |
| PC (36:4) B - ESI(-) | Lognormal | -0.011 | (-0.027, 0.005) | 0.170 | 0.864 | 0.178 | (-0.028, 0.004) |
| DG (38:5) | Lognormal | -0.010 | (-0.031, 0.010) | 0.330 | 0.864 | 0.103 | (-0.032, 0.011) |
| DG (38:6) | Lognormal | 0.000 | (-0.032, 0.031) | 0.987 | 0.996 | 0.061 | (-0.031, 0.036) |
| Ceramide (d42:2) B - ESI (+) | Lognormal | -0.010 | (-0.029, 0.009) | 0.311 | 0.864 | 0.112 | (-0.031, 0.008) |
| PC (34:3) C | Lognormal | -0.020 | (-0.044, 0.004) | 0.105 | 0.864 | 0.277 | (-0.045, 0.003) |
| TG (58:9) | Lognormal | 0.006 | (-0.025, 0.036) | 0.714 | 0.939 | 0.068 | (-0.028, 0.035) |
| PE (p-34:1)/PE (o-34:2) - ESI(+) | Lognormal | -0.012 | (-0.050, 0.025) | 0.518 | 0.899 | 0.085 | (-0.051, 0.029) |

**Supplementary Table S6.** Regression and Bayesian estimates comparing sex-stratified ME/CFS vs. controls.

| Metabolite | Regression Model | Male ME/CFS vs. Male Controls | | | | | |
|---|---|---|---|---|---|---|---|
| | | Estimated Coefficient | 95% CI | p-value | FDR | BayesFactor | 95% HDI |
| SM (d42:1) - ESI(-) | Lognormal | -0.012 | (-0.032, 0.008) | 0.231 | 0.864 | 0.132 | (-0.033, 0.007) |
| AC (18:2) | Lognormal | 0.007 | (-0.020, 0.033) | 0.627 | 0.928 | 0.080 | (-0.020, 0.035) |
| Ceramide (d42:1) - ESI(-) | Lognormal | -0.017 | (-0.038, 0.004) | 0.125 | 0.864 | 0.259 | (-0.039, 0.005) |
| Ceramide (d34:1) - ESI(-) | Lognormal | 0.002 | (-0.018, 0.022) | 0.830 | 0.962 | 0.071 | (-0.019, 0.021) |
| PC (p-38:4)/PC (o-38:5) B | Lognormal | -0.007 | (-0.030, 0.017) | 0.584 | 0.924 | 0.082 | (-0.032, 0.016) |
| CE (18:3) | Lognormal | -0.026 | (-0.063, 0.011) | 0.172 | 0.864 | 0.196 | (-0.066, 0.010) |
| AC (14:2) | Lognormal | 0.000 | (-0.050, 0.049) | 0.992 | 0.997 | 0.072 | (-0.048, 0.051) |
| PC (40:5) A - ESI(+) | Lognormal | -0.002 | (-0.018, 0.014) | 0.792 | 0.956 | 0.069 | (-0.017, 0.014) |
| PC (33:2) - ESI(-) | Lognormal | -0.028 | (-0.065, 0.008) | 0.132 | 0.864 | 0.222 | (-0.067, 0.008) |
| PC (37:2) - ESI(+) | Lognormal | 0.000 | (-0.015, 0.016) | 0.956 | 0.996 | 0.058 | (-0.016, 0.016) |
| LPC (14:0) - ESI(+) | Lognormal | -0.036 | (-0.070, -0.002) | 0.046 | 0.864 | 0.578 | (-0.072, -0.003) |
| TG (54:6) B | Lognormal | 0.030 | (-0.043, 0.103) | 0.425 | 0.873 | 0.096 | (-0.042, 0.105) |
| SM (d36:3) - ESI(-) | Lognormal | -0.036 | (-0.063, -0.008) | 0.015 | 0.864 | 1.197 | (-0.064, -0.008) |
| FA (18:1) (oleic acid) | Lognormal | 0.009 | (-0.011, 0.028) | 0.377 | 0.873 | 0.108 | (-0.010, 0.030) |
| PC (p-36:3)/PC (o-36:4) - ESI(+) | Lognormal | -0.007 | (-0.025, 0.010) | 0.413 | 0.873 | 0.099 | (-0.026, 0.010) |
| FA (14:1) (physeteric acid) | Lognormal | -0.008 | (-0.053, 0.037) | 0.739 | 0.949 | 0.070 | (-0.056, 0.037) |
| PC (34:0) - ESI(-) | Lognormal | -0.010 | (-0.033, 0.012) | 0.381 | 0.873 | 0.097 | (-0.034, 0.012) |
| Ceramide (d40:1) | Lognormal | -0.010 | (-0.031, 0.011) | 0.370 | 0.873 | 0.108 | (-0.030, 0.012) |
| SM (d36:0) - ESI(+) | Lognormal | 0.008 | (-0.024, 0.040) | 0.633 | 0.928 | 0.066 | (-0.024, 0.041) |
| LPC (16:0) - ESI(-) | Lognormal | -0.021 | (-0.038, -0.003) | 0.025 | 0.864 | 0.751 | (-0.038, -0.002) |
| PE (p-40:5)/PE (o-40:6) | Lognormal | 0.006 | (-0.013, 0.025) | 0.529 | 0.899 | 0.056 | (-0.014, 0.025) |
| LPC (18:0) A - ESI(-) | Lognormal | -0.035 | (-0.062, -0.008) | 0.014 | 0.864 | 1.869 | (-0.062, -0.007) |
| PC (38:4) A - ESI(-) | Lognormal | -0.014 | (-0.033, 0.004) | 0.132 | 0.864 | 0.233 | (-0.033, 0.005) |
| TG (54:7) A | Lognormal | 0.013 | (-0.030, 0.055) | 0.570 | 0.915 | 0.078 | (-0.031, 0.055) |
| TG (56:8) B | Lognormal | 0.007 | (-0.024, 0.038) | 0.665 | 0.928 | 0.068 | (-0.025, 0.038) |
| PE (36:3) | Lognormal | -0.030 | (-0.103, 0.043) | 0.427 | 0.873 | 0.094 | (-0.106, 0.044) |
| PC (37:4) - ESI(+) | Lognormal | -0.018 | (-0.051, 0.016) | 0.316 | 0.864 | 0.112 | (-0.053, 0.016) |
| PC (34:4) - ESI(-) | Lognormal | -0.061 | (-0.117, -0.006) | 0.036 | 0.864 | 0.735 | (-0.117, -0.004) |
| SM (d36:1) - ESI(-) | Lognormal | -0.014 | (-0.035, 0.008) | 0.219 | 0.864 | 0.143 | (-0.036, 0.007) |
| AC (12:1) | Lognormal | 0.004 | (-0.034, 0.041) | 0.851 | 0.966 | 0.069 | (-0.036, 0.043) |
| Cholesterol | Lognormal | 0.005 | (-0.004, 0.014) | 0.269 | 0.864 | 0.101 | (-0.004, 0.013) |
| PC (p-38:3)/PC (o-38:4) A - ESI(+) | Lognormal | -0.007 | (-0.028, 0.013) | 0.498 | 0.897 | 0.081 | (-0.029, 0.013) |
| PC (p-38:3)/PC (o-38:4) B - ESI(+) | Lognormal | -0.006 | (-0.028, 0.016) | 0.620 | 0.928 | 0.074 | (-0.027, 0.017) |
| AC (10:0) | Lognormal | 0.008 | (-0.043, 0.059) | 0.752 | 0.952 | 0.069 | (-0.047, 0.059) |
| TG (60:11) | Lognormal | 0.007 | (-0.040, 0.054) | 0.778 | 0.956 | 0.068 | (-0.042, 0.057) |
| PC (p-38:4)/PC (o-38:5) B | Lognormal | -0.026 | (-0.063, 0.010) | 0.170 | 0.864 | 0.179 | (-0.064, 0.009) |
| TG (52:4) | Lognormal | 0.014 | (-0.005, 0.033) | 0.167 | 0.864 | 0.191 | (-0.006, 0.034) |
| LPC (18:0) B - ESI(-) | Lognormal | -0.025 | (-0.044, -0.005) | 0.017 | 0.864 | 1.323 | (-0.044, -0.004) |
| SM (d41:2) B - ESI(+) | Lognormal | 0.003 | (-0.016, 0.022) | 0.759 | 0.956 | 0.071 | (-0.017, 0.021) |
| TG (58:6) | Lognormal | 0.001 | (-0.020, 0.023) | 0.907 | 0.977 | 0.062 | (-0.021, 0.023) |
| PC (p-36:1)/PC (o-36:2) B | Lognormal | 0.007 | (-0.025, 0.038) | 0.674 | 0.928 | 0.072 | (-0.025, 0.039) |
| PC (37:5) | Lognormal | 0.001 | (-0.027, 0.030) | 0.930 | 0.991 | 0.065 | (-0.027, 0.029) |
| PC (36:5) D | Lognormal | -0.011 | (-0.042, 0.020) | 0.484 | 0.884 | 0.086 | (-0.042, 0.021) |
| SM (d40:1) - ESI(+) | Lognormal | -0.003 | (-0.016, 0.010) | 0.667 | 0.928 | 0.069 | (-0.017, 0.010) |
| PC (35:2) B | Lognormal | 0.008 | (-0.005, 0.021) | 0.217 | 0.864 | 0.126 | (-0.005, 0.020) |
| PC (36:3) A - ESI(+) | Lognormal | -0.009 | (-0.025, 0.006) | 0.248 | 0.864 | 0.128 | (-0.028, 0.005) |
| PC (p-32:0)/PC (o-32:1) - ESI(-) | Lognormal | -0.009 | (-0.046, 0.029) | 0.651 | 0.928 | 0.080 | (-0.045, 0.032) |

**Supplementary Table S6.** Regression and Bayesian estimates comparing sex-stratified ME/CFS vs. controls.

| Metabolite | Regression Model | Male ME/CFS vs. Male Controls | | | | | |
|---|---|---|---|---|---|---|---|
| | | Estimated Coefficient | 95% CI | p-value | FDR | BayesFactor | 95% HDI |
| SM (d43:1) - ESI(-) | Lognormal | -0.046 | (-0.110, 0.019) | 0.172 | 0.864 | 0.165 | (-0.113, 0.015) |
| PC (40:6)B | Lognormal | 0.006 | (-0.011, 0.022) | 0.525 | 0.899 | 0.082 | (-0.011, 0.023) |
| TG (48:0) | Lognormal | -0.019 | (-0.064, 0.027) | 0.430 | 0.873 | 0.094 | (-0.062, 0.030) |
| SM (d42:0) - ESI(+) | Lognormal | -0.068 | (-0.126, -0.011) | 0.025 | 0.864 | 0.799 | (-0.128, -0.007) |
| SM (d30:1) - ESI(-) | Lognormal | -0.051 | (-0.117, 0.015) | 0.138 | 0.864 | 0.210 | (-0.116, 0.019) |
| PC (36:3) B - ESI(-) | Lognormal | -0.013 | (-0.032, 0.006) | 0.201 | 0.864 | 0.167 | (-0.032, 0.007) |
| Ceramide (d32:1) | Lognormal | -0.005 | (-0.044, 0.035) | 0.811 | 0.956 | 0.073 | (-0.046, 0.034) |
| PE (p-40:4)/PE (o-40:5) B | Lognormal | -0.016 | (-0.049, 0.017) | 0.346 | 0.864 | 0.073 | (-0.048, 0.017) |
| DG (36:4) B | Lognormal | -0.021 | (-0.091, 0.048) | 0.551 | 0.904 | 0.077 | (-0.087, 0.054) |
| TG (48:4) A | Lognormal | -0.033 | (-0.093, 0.027) | 0.283 | 0.864 | 0.133 | (-0.096, 0.029) |
| PC (34:3) A | Lognormal | -0.007 | (-0.031, 0.017) | 0.564 | 0.908 | 0.087 | (-0.033, 0.017) |
| PC (35:1) - ESI(+) | Lognormal | 0.002 | (-0.014, 0.018) | 0.814 | 0.956 | 0.073 | (-0.014, 0.017) |
| LPC (p-16:0)/LPC (o-16:1) | Lognormal | -0.013 | (-0.041, 0.015) | 0.379 | 0.873 | 0.103 | (-0.044, 0.015) |
| PC (p-38:5)/PC (o-38:6) B | Lognormal | 0.000 | (-0.022, 0.021) | 0.969 | 0.996 | 0.071 | (-0.024, 0.020) |
| TG (49:2) | Lognormal | -0.008 | (-0.053, 0.036) | 0.715 | 0.939 | 0.077 | (-0.052, 0.038) |
| LPC (20:3) - ESI(-) | Lognormal | -0.046 | (-0.080, -0.011) | 0.013 | 0.864 | 1.640 | (-0.082, -0.010) |
| Ceramide (d41:1) - ESI (+) | Lognormal | -0.011 | (-0.036, 0.014) | 0.385 | 0.873 | 0.100 | (-0.036, 0.015) |
| AC (12:0) | Lognormal | -0.006 | (-0.051, 0.040) | 0.807 | 0.956 | 0.074 | (-0.058, 0.038) |
| PC (36:3) A - ESI(-) | Lognormal | -0.011 | (-0.032, 0.010) | 0.310 | 0.864 | 0.119 | (-0.031, 0.013) |
| FA (20:3) (homo-gamma-linolenic acid) | Lognormal | -0.005 | (-0.026, 0.016) | 0.634 | 0.928 | 0.070 | (-0.026, 0.016) |
| PC (42:10) | Lognormal | -0.013 | (-0.054, 0.028) | 0.530 | 0.899 | 0.077 | (-0.056, 0.029) |
| Lactosylceramide (d18:1/24:1(15Z)) | Lognormal | 0.011 | (-0.025, 0.048) | 0.553 | 0.904 | 0.078 | (-0.025, 0.049) |
| TG (60:6) | Lognormal | -0.007 | (-0.047, 0.033) | 0.735 | 0.949 | 0.066 | (-0.048, 0.033) |
| PE (p-40:4)/PE (o-40:5) | Lognormal | -0.016 | (-0.055, 0.023) | 0.437 | 0.881 | 0.070 | (-0.053, 0.029) |
| PC (38:5) A | Lognormal | -0.003 | (-0.016, 0.010) | 0.648 | 0.928 | 0.076 | (-0.017, 0.011) |
| PC (34:1) | Lognormal | -0.003 | (-0.012, 0.006) | 0.553 | 0.904 | 0.081 | (-0.013, 0.006) |
| TG (56:5) A | Lognormal | 0.001 | (-0.026, 0.027) | 0.953 | 0.996 | 0.077 | (-0.026, 0.026) |
| DG (36:4) A | Lognormal | 0.021 | (-0.012, 0.054) | 0.222 | 0.864 | 0.136 | (-0.012, 0.053) |
| AC (8:0) | Lognormal | 0.016 | (-0.041, 0.072) | 0.591 | 0.926 | 0.074 | (-0.042, 0.071) |
| Ceramide (d39:1) | Lognormal | -0.038 | (-0.080, 0.004) | 0.082 | 0.864 | 0.296 | (-0.078, 0.006) |
| PC (38:6) A - ESI(+) | Lognormal | -0.018 | (-0.035, -0.001) | 0.039 | 0.864 | 0.649 | (-0.035, -0.001) |
| PC (38:5) B - ESI(+) | Lognormal | -0.006 | (-0.024, 0.012) | 0.509 | 0.899 | 0.085 | (-0.023, 0.013) |
| SM (d40:0) | Lognormal | -0.013 | (-0.037, 0.010) | 0.275 | 0.864 | 0.107 | (-0.037, 0.012) |
| PC (o-32:0) - ESI(+) | Lognormal | 0.011 | (-0.005, 0.027) | 0.196 | 0.864 | 0.163 | (-0.007, 0.027) |
| PE (p-38:3)/PE (o-38:4) | Lognormal | -0.038 | (-0.088, 0.011) | 0.139 | 0.864 | 0.226 | (-0.091, 0.013) |
| TG (56:9) | Lognormal | -0.015 | (-0.055, 0.025) | 0.474 | 0.884 | 0.094 | (-0.056, 0.025) |
| TG (50:4) | Lognormal | -0.002 | (-0.037, 0.034) | 0.930 | 0.991 | 0.077 | (-0.039, 0.032) |
| PC (p-34:1)/PC (o-34:2) B | Lognormal | 0.012 | (-0.031, 0.054) | 0.597 | 0.926 | 0.070 | (-0.032, 0.055) |
| SM (d34:0) - ESI(-) | Lognormal | -0.010 | (-0.039, 0.018) | 0.475 | 0.884 | 0.086 | (-0.040, 0.019) |
| PE (p-38:6)/PE (o-38:7) | Lognormal | -0.028 | (-0.075, 0.019) | 0.252 | 0.864 | 0.133 | (-0.076, 0.019) |
| SM (d38:1) | Lognormal | -0.006 | (-0.020, 0.008) | 0.417 | 0.873 | 0.093 | (-0.020, 0.008) |
| FA (24:0) (lignoceric acid) | Lognormal | -0.011 | (-0.037, 0.016) | 0.429 | 0.873 | 0.090 | (-0.038, 0.017) |
| SM (d34:1) - ESI(+) | Lognormal | 0.006 | (-0.004, 0.017) | 0.220 | 0.864 | 0.141 | (-0.004, 0.017) |
| CE (20:3) | Lognormal | 0.000 | (-0.027, 0.026) | 0.975 | 0.996 | 0.066 | (-0.028, 0.027) |
| LPC (20:2) - ESI(+) | Lognormal | -0.039 | (-0.097, 0.018) | 0.188 | 0.864 | 0.162 | (-0.093, 0.022) |
| PC (42:6) | Lognormal | -0.031 | (-0.093, 0.032) | 0.340 | 0.864 | 0.110 | (-0.092, 0.035) |
| DG (32:0) | Lognormal | -0.005 | (-0.040, 0.030) | 0.773 | 0.956 | 0.066 | (-0.039, 0.033) |

**Supplementary Table S6.** Regression and Bayesian estimates comparing sex-stratified ME/CFS vs. controls.

| Metabolite | Regression Model | Male ME/CFS vs. Male Controls | | | | | |
|---|---|---|---|---|---|---|---|
| | | Estimated Coefficient | 95% CI | p-value | FDR | BayesFactor | 95% HDI |
| CE (20:5) | Lognormal | -0.006 | (-0.046, 0.034) | 0.765 | 0.956 | 0.068 | (-0.050, 0.033) |
| PC (o-32:0) - ESI(-) | Lognormal | 0.002 | (-0.025, 0.028) | 0.896 | 0.976 | 0.065 | (-0.024, 0.029) |
| PC (32:0) - ESI(-) | Lognormal | -0.006 | (-0.025, 0.013) | 0.561 | 0.906 | 0.075 | (-0.026, 0.014) |
| PC (40:8) - ESI(-) | Lognormal | -0.038 | (-0.066, -0.010) | 0.011 | 0.864 | 1.471 | (-0.066, -0.008) |
| Ceramide (d38:1) - ESI(+) | Lognormal | -0.003 | (-0.025, 0.019) | 0.783 | 0.956 | 0.071 | (-0.027, 0.019) |
| Ceramide (d43:1) | Lognormal | -0.066 | (-0.183, 0.052) | 0.281 | 0.864 | 0.134 | (-0.186, 0.060) |
| SM (d42:1) - ESI(+) | Lognormal | -0.001 | (-0.014, 0.012) | 0.904 | 0.977 | 0.070 | (-0.015, 0.013) |
| PC (37:2) - ESI(-) | Lognormal | -0.010 | (-0.049, 0.028) | 0.598 | 0.926 | 0.074 | (-0.051, 0.028) |
| PC (38:3) - ESI(-) | Lognormal | -0.021 | (-0.047, 0.005) | 0.122 | 0.864 | 0.224 | (-0.048, 0.005) |
| SM (d39:2) | Lognormal | 0.003 | (-0.017, 0.023) | 0.772 | 0.956 | 0.065 | (-0.017, 0.024) |
| SM (d30:1) - ESI(+) | Lognormal | -0.027 | (-0.079, 0.025) | 0.316 | 0.864 | 0.112 | (-0.081, 0.028) |
| LPC (o-16:0) | Lognormal | -0.006 | (-0.033, 0.020) | 0.640 | 0.928 | 0.070 | (-0.033, 0.023) |
| LPC (20:2) - ESI(-) | Lognormal | -0.022 | (-0.053, 0.010) | 0.185 | 0.864 | 0.154 | (-0.053, 0.011) |
| LPC (18:1) - ESI(-) | Lognormal | -0.028 | (-0.048, -0.008) | 0.011 | 0.864 | 1.679 | (-0.048, -0.006) |
| LPC (20:3) - ESI(+) | Lognormal | -0.023 | (-0.048, 0.002) | 0.085 | 0.864 | 0.293 | (-0.049, 0.003) |
| PC (38:6) B - ESI(+) | Lognormal | 0.007 | (-0.003, 0.018) | 0.195 | 0.864 | 0.149 | (-0.004, 0.018) |
| FA (20:3) (eicosatrienoic acid) | Lognormal | -0.018 | (-0.050, 0.015) | 0.293 | 0.864 | 0.126 | (-0.052, 0.014) |
| PC (p-44:4)/PC (o-44:5) - ESI(-) | Lognormal | -0.006 | (-0.032, 0.020) | 0.643 | 0.928 | 0.072 | (-0.031, 0.020) |
| CE (14:0) | Lognormal | 0.002 | (-0.072, 0.075) | 0.968 | 0.996 | 0.060 | (-0.072, 0.082) |
| TG (56:8) A | Lognormal | 0.005 | (-0.024, 0.035) | 0.721 | 0.943 | 0.075 | (-0.025, 0.036) |
| Ceramide (d33:1) | Lognormal | -0.012 | (-0.040, 0.016) | 0.394 | 0.873 | 0.106 | (-0.041, 0.015) |
| TG (46:0) | Lognormal | -0.042 | (-0.114, 0.030) | 0.258 | 0.864 | 0.129 | (-0.117, 0.029) |
| PE (38:6) - ESI(+) | Lognormal | -0.049 | (-0.113, 0.014) | 0.134 | 0.864 | 0.191 | (-0.113, 0.015) |
| SM (d34:2) - ESI(+) | Lognormal | 0.008 | (-0.005, 0.020) | 0.232 | 0.864 | 0.131 | (-0.004, 0.021) |
| AC (18:0) | Lognormal | -0.009 | (-0.035, 0.018) | 0.519 | 0.899 | 0.101 | (-0.036, 0.019) |
| TG (51:2) | Lognormal | -0.003 | (-0.036, 0.031) | 0.865 | 0.968 | 0.069 | (-0.035, 0.033) |
| LPC (14:0) - ESI(-) | Lognormal | -0.058 | (-0.110, -0.005) | 0.038 | 0.864 | 0.720 | (-0.110, -0.005) |
| TG (54:6) A | Lognormal | 0.018 | (-0.017, 0.053) | 0.311 | 0.864 | 0.125 | (-0.019, 0.053) |
| FA (22:0) (behenic acid) | Lognormal | -0.006 | (-0.037, 0.025) | 0.685 | 0.929 | 0.071 | (-0.038, 0.026) |
| Ceramide (d42:2) B - ESI(-) | Lognormal | -0.007 | (-0.035, 0.020) | 0.615 | 0.928 | 0.079 | (-0.036, 0.020) |
| SM (d39:1) - ESI(+) | Lognormal | -0.012 | (-0.035, 0.010) | 0.279 | 0.864 | 0.114 | (-0.034, 0.011) |
| TG (50:5) | Lognormal | -0.009 | (-0.058, 0.040) | 0.708 | 0.937 | 0.079 | (-0.060, 0.039) |
| TG (46:4) A | Lognormal | -0.045 | (-0.125, 0.035) | 0.280 | 0.864 | 0.139 | (-0.128, 0.030) |
| GlcCer (d42:2) - ESI(+) | Lognormal | 0.022 | (-0.006, 0.050) | 0.130 | 0.864 | 0.232 | (-0.006, 0.050) |
| LPC (16:1) - ESI(-) | Lognormal | -0.043 | (-0.076, -0.010) | 0.015 | 0.864 | 1.339 | (-0.075, -0.006) |
| PC (35:2) A | Lognormal | -0.004 | (-0.021, 0.013) | 0.651 | 0.928 | 0.072 | (-0.021, 0.014) |
| TG (51:4) | Lognormal | 0.016 | (-0.021, 0.052) | 0.408 | 0.873 | 0.101 | (-0.021, 0.052) |
| PC (38:2) | Lognormal | -0.012 | (-0.038, 0.014) | 0.370 | 0.873 | 0.098 | (-0.037, 0.017) |
| PC 40:5e | Lognormal | 0.001 | (-0.017, 0.020) | 0.881 | 0.975 | 0.073 | (-0.018, 0.021) |
| PC (37:4) - ESI(-) | Lognormal | 0.000 | (-0.026, 0.027) | 0.979 | 0.996 | 0.065 | (-0.026, 0.028) |
| SM (d36:3) - ESI(+) | Lognormal | 0.004 | (-0.019, 0.027) | 0.719 | 0.942 | 0.066 | (-0.019, 0.028) |
| PE (34:2) - ESI(-) | Lognormal | -0.037 | (-0.096, 0.022) | 0.223 | 0.864 | 0.139 | (-0.103, 0.020) |
| TG (48:5) | Lognormal | -0.035 | (-0.104, 0.033) | 0.321 | 0.864 | 0.119 | (-0.109, 0.032) |
| GlcCer (d38:1) | Lognormal | -0.027 | (-0.052, -0.003) | 0.035 | 0.864 | 0.650 | (-0.054, -0.004) |
| PC (p-42:4)/PC (o-42:5) - ESI(-) | Lognormal | 0.007 | (-0.024, 0.038) | 0.658 | 0.928 | 0.073 | (-0.024, 0.040) |
| TG (54:9) | Lognormal | -0.093 | (-0.213, 0.026) | 0.133 | 0.864 | 0.203 | (-0.210, 0.025) |
| LPE (18:0) | Lognormal | -0.028 | (-0.049, -0.006) | 0.017 | 0.864 | 1.366 | (-0.047, -0.004) |

**Supplementary Table S6.** Regression and Bayesian estimates comparing sex-stratified ME/CFS vs. controls.

| Metabolite | Regression Model | Male ME/CFS vs. Male Controls | | | | | |
|---|---|---|---|---|---|---|---|
| | | Estimated Coefficient | 95% CI | p-value | FDR | BayesFactor | 95% HDI |
| TG (58:4) | Lognormal | -0.012 | (-0.065, 0.040) | 0.653 | 0.928 | 0.085 | (-0.066, 0.041) |
| PC (34:4) - ESI(+) | Lognormal | -0.035 | (-0.077, 0.006) | 0.103 | 0.864 | 0.272 | (-0.078, 0.005) |
| TG (42:0) | Lognormal | -0.037 | (-0.112, 0.038) | 0.343 | 0.864 | 0.103 | (-0.110, 0.044) |
| TG (53:2) | Lognormal | -0.003 | (-0.035, 0.029) | 0.869 | 0.968 | 0.071 | (-0.034, 0.031) |
| PC (p-38:4)/PC (o-38:5) A | Lognormal | -0.002 | (-0.017, 0.014) | 0.849 | 0.966 | 0.081 | (-0.018, 0.014) |
| TG (46:3) A | Lognormal | -0.026 | (-0.120, 0.068) | 0.596 | 0.926 | 0.088 | (-0.124, 0.065) |
| TG (56:5) C | Lognormal | 0.009 | (-0.011, 0.028) | 0.405 | 0.873 | 0.093 | (-0.013, 0.028) |
| PC (36:1) - ESI(+) | Lognormal | -0.009 | (-0.020, 0.001) | 0.084 | 0.864 | 0.261 | (-0.019, 0.002) |
| FA (20:1) (eicosenoic acid) | Lognormal | 0.011 | (-0.012, 0.034) | 0.340 | 0.864 | 0.098 | (-0.011, 0.034) |
| PC (37:6) | Lognormal | 0.017 | (-0.017, 0.051) | 0.323 | 0.864 | 0.124 | (-0.016, 0.054) |
| LPE (22:6) | Lognormal | -0.027 | (-0.061, 0.007) | 0.131 | 0.864 | 0.220 | (-0.061, 0.010) |
| SM (d40:2) A - ESI(+) | Lognormal | 0.003 | (-0.018, 0.024) | 0.765 | 0.956 | 0.072 | (-0.017, 0.025) |
| LPC (20:5) | Lognormal | -0.024 | (-0.069, 0.021) | 0.296 | 0.864 | 0.132 | (-0.074, 0.021) |
| SM (d36:2) - ESI(-) | Lognormal | -0.014 | (-0.041, 0.012) | 0.292 | 0.864 | 0.124 | (-0.043, 0.011) |
| DG (36:5) | Lognormal | 0.026 | (-0.014, 0.067) | 0.214 | 0.864 | 0.152 | (-0.012, 0.065) |
| PC (42:5) | Lognormal | 0.001 | (-0.031, 0.032) | 0.961 | 0.996 | 0.073 | (-0.030, 0.034) |
| SM (d44:2) | Lognormal | 0.014 | (-0.015, 0.044) | 0.346 | 0.864 | 0.082 | (-0.016, 0.046) |
| LPE (16:0) | Lognormal | -0.045 | (-0.090, -0.001) | 0.054 | 0.864 | 0.421 | (-0.095, -0.001) |
| SM (d32:1) - ESI(+) | Lognormal | -0.013 | (-0.037, 0.011) | 0.288 | 0.864 | 0.121 | (-0.039, 0.012) |
| SM (d41:3) | Lognormal | 0.000 | (-0.022, 0.021) | 0.979 | 0.996 | 0.070 | (-0.021, 0.023) |
| TG (42:1) | Lognormal | -0.061 | (-0.230, 0.108) | 0.483 | 0.884 | 0.088 | (-0.242, 0.101) |
| TG (60:12) | Lognormal | 0.026 | (-0.034, 0.085) | 0.400 | 0.873 | 0.081 | (-0.039, 0.086) |
| PC (p-40:3)/PC (o-40:4) | Lognormal | -0.024 | (-0.057, 0.009) | 0.168 | 0.864 | 0.179 | (-0.060, 0.010) |
| PC (40:4) - ESI(-) | Lognormal | -0.019 | (-0.056, 0.018) | 0.317 | 0.864 | 0.109 | (-0.057, 0.018) |
| FA (20:5) (eicosapentaenoic acid) | Lognormal | 0.018 | (-0.026, 0.063) | 0.419 | 0.873 | 0.105 | (-0.027, 0.066) |
| DG (36:2) | Lognormal | 0.003 | (-0.018, 0.024) | 0.789 | 0.956 | 0.069 | (-0.018, 0.025) |
| AC (14:1) | Lognormal | 0.005 | (-0.040, 0.051) | 0.819 | 0.957 | 0.073 | (-0.041, 0.052) |
| SM (d40:2) A - ESI(-) | Lognormal | -0.017 | (-0.062, 0.028) | 0.460 | 0.884 | 0.089 | (-0.062, 0.026) |
| DG (36:1) | Lognormal | -0.024 | (-0.064, 0.015) | 0.230 | 0.864 | 0.138 | (-0.066, 0.015) |
| LPC (17:1) | Lognormal | -0.018 | (-0.047, 0.010) | 0.214 | 0.864 | 0.162 | (-0.047, 0.014) |
| PG (34:0)/PG (17:0/17:0) | Lognormal | -0.001 | (-0.013, 0.012) | 0.900 | 0.976 | 0.061 | (-0.013, 0.013) |
| SM (d43:2) - ESI(-) | Lognormal | -0.029 | (-0.091, 0.034) | 0.373 | 0.873 | 0.098 | (-0.096, 0.034) |
| SM (d36:0) - ESI(-) | Lognormal | -0.042 | (-0.104, 0.020) | 0.196 | 0.864 | 0.159 | (-0.111, 0.018) |
| SM (d38:0) | Lognormal | -0.149 | (-0.266, -0.032) | 0.017 | 0.864 | 1.419 | (-0.263, -0.030) |
| TG (42:3) | Lognormal | -0.032 | (-0.120, 0.056) | 0.477 | 0.884 | 0.087 | (-0.121, 0.057) |
| Ceramide (d44:1) | Lognormal | -0.023 | (-0.083, 0.038) | 0.468 | 0.884 | 0.093 | (-0.081, 0.041) |
| PC (36:3) B - ESI(+) | Lognormal | -0.006 | (-0.031, 0.020) | 0.668 | 0.928 | 0.081 | (-0.033, 0.019) |
| TG (40:1) | Lognormal | -0.041 | (-0.122, 0.040) | 0.327 | 0.864 | 0.111 | (-0.126, 0.049) |
| Ceramide (d38:1) - ESI(-) | Lognormal | -0.018 | (-0.050, 0.013) | 0.263 | 0.864 | 0.115 | (-0.050, 0.014) |
| PC (33:1) - ESI(+) | Lognormal | -0.002 | (-0.023, 0.018) | 0.842 | 0.962 | 0.071 | (-0.023, 0.019) |
| TG (48:3) | Lognormal | -0.028 | (-0.080, 0.024) | 0.296 | 0.864 | 0.124 | (-0.086, 0.021) |
| Ceramide (d42:2) A - ESI(-) | Lognormal | -0.010 | (-0.033, 0.013) | 0.414 | 0.873 | 0.084 | (-0.033, 0.014) |
| SM (d32:2) - ESI(+) | Lognormal | 0.004 | (-0.021, 0.029) | 0.741 | 0.949 | 0.063 | (-0.020, 0.030) |
| TG (44:0) | Lognormal | -0.046 | (-0.118, 0.026) | 0.223 | 0.864 | 0.147 | (-0.120, 0.030) |
| TG (54:5) A | Lognormal | 0.017 | (-0.009, 0.043) | 0.216 | 0.864 | 0.147 | (-0.012, 0.042) |
| TG (52:2) | Lognormal | -0.002 | (-0.017, 0.012) | 0.743 | 0.949 | 0.070 | (-0.016, 0.014) |
| PI (38:4)/PI (18:0-20:4) | Lognormal | -0.021 | (-0.042, 0.000) | 0.054 | 0.864 | 0.447 | (-0.043, 0.000) |

**Supplementary Table S6.** Regression and Bayesian estimates comparing sex-stratified ME/CFS vs. controls.

| Metabolite | Regression Model | Male ME/CFS vs. Male Controls | | | | | |
|---|---|---|---|---|---|---|---|
| | | Estimated Coefficient | 95% CI | p-value | FDR | BayesFactor | 95% HDI |
| PC (37:3) | Lognormal | 0.007 | (-0.023, 0.038) | 0.649 | 0.928 | 0.078 | (-0.024, 0.039) |
| TG (56:3) | Lognormal | 0.000 | (-0.044, 0.043) | 0.987 | 0.996 | 0.074 | (-0.043, 0.046) |
| SM (d32:2) - ESI(-) | Lognormal | -0.027 | (-0.078, 0.024) | 0.307 | 0.864 | 0.119 | (-0.078, 0.024) |
| TG (46:4) B | Lognormal | -0.036 | (-0.125, 0.054) | 0.439 | 0.881 | 0.107 | (-0.123, 0.058) |
| TG (46:2) | Lognormal | -0.048 | (-0.118, 0.022) | 0.189 | 0.864 | 0.189 | (-0.119, 0.021) |
| PC (p-40:1)/PC (o-40:2) | Lognormal | 0.018 | (-0.015, 0.051) | 0.285 | 0.864 | 0.103 | (-0.016, 0.051) |
| PE (34:1) | Lognormal | -0.049 | (-0.114, 0.016) | 0.146 | 0.864 | 0.174 | (-0.115, 0.020) |
| SM (d41:1) - ESI(+) | Lognormal | 0.000 | (-0.018, 0.017) | 0.969 | 0.996 | 0.064 | (-0.017, 0.017) |
| SM (d38:2) - ESI(+) | Lognormal | 0.009 | (-0.006, 0.024) | 0.243 | 0.864 | 0.148 | (-0.006, 0.024) |
| LPC (15:0) | Lognormal | 0.001 | (-0.033, 0.035) | 0.955 | 0.996 | 0.072 | (-0.034, 0.036) |
| PC (p-40:7)/PC (o-40:8) | Lognormal | -0.002 | (-0.092, 0.089) | 0.973 | 0.996 | 0.065 | (-0.093, 0.092) |
| PC (p-42:3)/PC (o-42:4) | Lognormal | 0.000 | (-0.023, 0.023) | 0.989 | 0.997 | 0.061 | (-0.022, 0.024) |
| PE (36:4) - ESI(+) | Lognormal | -0.040 | (-0.090, 0.011) | 0.134 | 0.864 | 0.204 | (-0.094, 0.010) |
| SM (d42:3) - ESI(+) | Lognormal | 0.013 | (-0.003, 0.029) | 0.130 | 0.864 | 0.231 | (-0.004, 0.029) |
| GlcCer (d41:1) | Lognormal | -0.021 | (-0.048, 0.006) | 0.139 | 0.864 | 0.198 | (-0.048, 0.007) |
| TG (58:3) | Lognormal | -0.007 | (-0.073, 0.059) | 0.837 | 0.962 | 0.070 | (-0.077, 0.058) |
| TG (48:4) B | Lognormal | -0.046 | (-0.117, 0.025) | 0.211 | 0.864 | 0.153 | (-0.119, 0.027) |
| PC (39:6) | Lognormal | 0.017 | (-0.012, 0.046) | 0.268 | 0.864 | 0.130 | (-0.013, 0.047) |
| PE (p-36:5)/PE (o-36:6) | Lognormal | -0.001 | (-0.109, 0.108) | 0.992 | 0.997 | 0.062 | (-0.107, 0.119) |
| PC (36:4) B - ESI(+) | Lognormal | -0.022 | (-0.046, 0.003) | 0.098 | 0.864 | 0.253 | (-0.046, 0.006) |
| TG (54:1) | Lognormal | -0.024 | (-0.080, 0.032) | 0.402 | 0.873 | 0.097 | (-0.081, 0.032) |
| FA (18:2) (linoleic acid) | Lognormal | 0.011 | (-0.009, 0.031) | 0.289 | 0.864 | 0.121 | (-0.010, 0.032) |
| LPE (20:4) - ESI(+) | Lognormal | -0.023 | (-0.055, 0.009) | 0.164 | 0.864 | 0.165 | (-0.055, 0.011) |
| PC (p-42:5)/PC (o-42:6) A | Lognormal | 0.000 | (-0.029, 0.028) | 0.974 | 0.996 | 0.071 | (-0.030, 0.029) |
| LPC (16:1) - ESI(+) | Lognormal | -0.008 | (-0.032, 0.016) | 0.528 | 0.899 | 0.084 | (-0.032, 0.017) |
| PC (40:7) B - ESI(+) | Lognormal | 0.008 | (-0.012, 0.028) | 0.452 | 0.884 | 0.086 | (-0.012, 0.028) |
| GlcCer (d40:1) - ESI(-) | Lognormal | -0.012 | (-0.034, 0.010) | 0.284 | 0.864 | 0.113 | (-0.034, 0.011) |
| PE (38:2) | Lognormal | -0.032 | (-0.070, 0.006) | 0.104 | 0.864 | 0.275 | (-0.069, 0.007) |
| PC (36:5) C | Lognormal | 0.009 | (-0.035, 0.052) | 0.702 | 0.937 | 0.077 | (-0.039, 0.052) |
| TG (55:2) | Lognormal | -0.013 | (-0.055, 0.029) | 0.549 | 0.904 | 0.083 | (-0.053, 0.032) |
| TG (51:5) | Lognormal | -0.007 | (-0.056, 0.042) | 0.791 | 0.956 | 0.071 | (-0.055, 0.047) |
| GlcCer (d42:1) - ESI(-) | Lognormal | -0.010 | (-0.032, 0.012) | 0.362 | 0.873 | 0.100 | (-0.032, 0.014) |
| TG (54:8) | Lognormal | -0.024 | (-0.078, 0.031) | 0.405 | 0.873 | 0.104 | (-0.080, 0.034) |
| PC (40:7) A - ESI(+) | Lognormal | -0.018 | (-0.039, 0.003) | 0.095 | 0.864 | 0.245 | (-0.040, 0.003) |
| TG (50:1) | Lognormal | -0.053 | (-0.094, -0.012) | 0.015 | 0.864 | 1.192 | (-0.096, -0.011) |
| TG (54:2) | Lognormal | -0.005 | (-0.042, 0.032) | 0.794 | 0.956 | 0.070 | (-0.040, 0.033) |
| AC (18:1) | Lognormal | 0.008 | (-0.016, 0.033) | 0.508 | 0.899 | 0.083 | (-0.017, 0.032) |
| TG (40:0) | Lognormal | -0.023 | (-0.091, 0.045) | 0.509 | 0.899 | 0.085 | (-0.097, 0.043) |
| SM (d42:0) - ESI(-) | Lognormal | -0.012 | (-0.033, 0.008) | 0.231 | 0.864 | 0.109 | (-0.033, 0.009) |
| TG (58:10) | Lognormal | -0.008 | (-0.043, 0.026) | 0.638 | 0.928 | 0.075 | (-0.042, 0.028) |
| PC (39:4) | Lognormal | 0.003 | (-0.021, 0.026) | 0.825 | 0.962 | 0.067 | (-0.021, 0.027) |
| LPC (16:0) - ESI(+) | Lognormal | -0.006 | (-0.017, 0.004) | 0.221 | 0.864 | 0.152 | (-0.017, 0.004) |
| TG (53:5) | Lognormal | 0.012 | (-0.024, 0.047) | 0.520 | 0.899 | 0.080 | (-0.026, 0.048) |
| SM (d36:2) - ESI(+) | Lognormal | 0.009 | (-0.007, 0.025) | 0.273 | 0.864 | 0.121 | (-0.008, 0.025) |
| Ceramide (d34:1) - ESI(+) | Lognormal | 0.027 | (0.012, 0.042) | 0.001 | 0.452 | 9.566 | (0.012, 0.042) |
| Ceramide (d34:2) | Lognormal | 0.008 | (-0.018, 0.033) | 0.548 | 0.904 | 0.083 | (-0.019, 0.034) |
| TG (56:10) | Lognormal | -0.041 | (-0.103, 0.021) | 0.202 | 0.864 | 0.157 | (-0.103, 0.025) |

**Supplementary Table S6.** Regression and Bayesian estimates comparing sex-stratified ME/CFS vs. controls.

| Metabolite | Regression Model | Male ME/CFS vs. Male Controls | | | | | |
|---|---|---|---|---|---|---|---|
| | | Estimated Coefficient | 95% CI | p-value | FDR | BayesFactor | 95% HDI |
| PC (30:0) | Lognormal | -0.023 | (-0.053, 0.008) | 0.155 | 0.864 | 0.186 | (-0.054, 0.008) |
| TG (59:3) | Lognormal | -0.019 | (-0.069, 0.032) | 0.472 | 0.884 | 0.092 | (-0.072, 0.031) |
| LPC (20:4) | Lognormal | -0.018 | (-0.045, 0.008) | 0.173 | 0.864 | 0.167 | (-0.046, 0.007) |
| TAG (58:7)/TAG (18:1/18:1/22:5) | Lognormal | -0.006 | (-0.030, 0.017) | 0.605 | 0.926 | 0.085 | (-0.031, 0.018) |
| GlcCer (d14:1(4E)/20:0(2OH)) | Lognormal | -0.029 | (-0.066, 0.008) | 0.132 | 0.864 | 0.201 | (-0.066, 0.007) |
| TG (50:2) | Lognormal | -0.004 | (-0.028, 0.020) | 0.749 | 0.951 | 0.071 | (-0.029, 0.021) |
| TG (56:7) A | Lognormal | 0.009 | (-0.068, 0.086) | 0.813 | 0.956 | 0.063 | (-0.065, 0.090) |
| TG (60:4) | Lognormal | -0.028 | (-0.100, 0.045) | 0.458 | 0.884 | 0.091 | (-0.107, 0.046) |
| CE (18:1) | Lognormal | 0.008 | (-0.020, 0.035) | 0.587 | 0.926 | 0.069 | (-0.019, 0.037) |
| TG (46:3) B | Lognormal | -0.011 | (-0.097, 0.074) | 0.793 | 0.956 | 0.073 | (-0.093, 0.083) |
| GlcCer (d42:2) - ESI(-) | Lognormal | 0.001 | (-0.026, 0.028) | 0.920 | 0.983 | 0.061 | (-0.027, 0.029) |
| PE (36:1) - ESI(-) | Lognormal | -0.062 | (-0.123, -0.001) | 0.053 | 0.864 | 0.447 | (-0.127, -0.001) |
| LPC (18:0) - ESI(+) | Lognormal | -0.013 | (-0.029, 0.002) | 0.106 | 0.864 | 0.273 | (-0.028, 0.004) |
| TG (64:2) | Lognormal | -0.047 | (-0.116, 0.021) | 0.184 | 0.864 | 0.178 | (-0.115, 0.022) |
| LPC (22:5) - ESI(+) | Lognormal | -0.018 | (-0.051, 0.016) | 0.305 | 0.864 | 0.109 | (-0.050, 0.017) |
| SM (d41:1) - ESI(-) | Lognormal | 0.117 | (-0.110, 0.343) | 0.320 | 0.864 | 0.125 | (-0.121, 0.349) |
| TG (60:2) | Lognormal | -0.063 | (-0.154, 0.029) | 0.186 | 0.864 | 0.198 | (-0.150, 0.031) |
| PC (40:5) B - ESI(-) | Lognormal | -0.049 | (-0.102, 0.004) | 0.075 | 0.864 | 0.375 | (-0.101, 0.005) |
| TG (56:2) | Lognormal | -0.020 | (-0.075, 0.035) | 0.478 | 0.884 | 0.095 | (-0.075, 0.039) |
| FA (20:2) (eicosadienoic acid) | Lognormal | 0.010 | (-0.017, 0.036) | 0.483 | 0.884 | 0.095 | (-0.017, 0.037) |
| AC (16:0) | Lognormal | 0.005 | (-0.018, 0.028) | 0.677 | 0.929 | 0.084 | (-0.019, 0.028) |
| DG (32:1) | Lognormal | -0.034 | (-0.081, 0.013) | 0.167 | 0.864 | 0.170 | (-0.082, 0.015) |
| PC (p-40:6)/PC (o-40:7) B | Lognormal | 0.008 | (-0.025, 0.040) | 0.642 | 0.928 | 0.077 | (-0.027, 0.038) |
| FA (14:0) (myristic acid) | Lognormal | 0.001 | (-0.018, 0.020) | 0.950 | 0.996 | 0.072 | (-0.019, 0.019) |
| TG (64:3) | Lognormal | -0.006 | (-0.055, 0.042) | 0.805 | 0.956 | 0.070 | (-0.054, 0.044) |
| PC (p-38:5)/PC (o-38:6) | Lognormal | -0.055 | (-0.142, 0.031) | 0.216 | 0.864 | 0.169 | (-0.149, 0.031) |
| PC (32:0) - ESI(+) | Lognormal | 0.004 | (-0.008, 0.017) | 0.511 | 0.899 | 0.079 | (-0.008, 0.017) |
| TG (62:3) | Lognormal | -0.050 | (-0.113, 0.012) | 0.124 | 0.864 | 0.237 | (-0.112, 0.017) |
| PC (35:4) - ESI(-) | Lognormal | -0.012 | (-0.050, 0.025) | 0.522 | 0.899 | 0.085 | (-0.050, 0.027) |
| TG (49:1) | Lognormal | -0.019 | (-0.070, 0.032) | 0.472 | 0.884 | 0.085 | (-0.072, 0.032) |
| PC (33:1) - ESI(-) | Lognormal | -0.024 | (-0.074, 0.025) | 0.342 | 0.864 | 0.111 | (-0.075, 0.028) |
| PC (36:1) - ESI(-) | Lognormal | -0.013 | (-0.030, 0.004) | 0.153 | 0.864 | 0.208 | (-0.031, 0.005) |
| SM (d43:2) - ESI(+) | Lognormal | 0.007 | (-0.028, 0.042) | 0.689 | 0.930 | 0.079 | (-0.028, 0.043) |
| SM (d32:0) - ESI(+) | Lognormal | -0.012 | (-0.045, 0.020) | 0.466 | 0.884 | 0.084 | (-0.048, 0.019) |
| PC (p-38:6)/PC (o-38:7) | Lognormal | -0.001 | (-0.030, 0.029) | 0.958 | 0.996 | 0.068 | (-0.029, 0.030) |
| PE (38:6) - ESI(-) | Lognormal | -0.053 | (-0.110, 0.005) | 0.079 | 0.864 | 0.309 | (-0.116, 0.005) |
| PC (32:1) - ESI(+) | Lognormal | -0.012 | (-0.040, 0.016) | 0.416 | 0.873 | 0.100 | (-0.042, 0.017) |
| PC (p-40:6)/PC (o-40:7) A | Lognormal | 0.002 | (-0.025, 0.029) | 0.896 | 0.976 | 0.070 | (-0.024, 0.033) |
| SM (d34:0) - ESI(+) | Lognormal | 0.002 | (-0.016, 0.020) | 0.811 | 0.956 | 0.066 | (-0.016, 0.021) |
| PC (38:5) A - ESI(-) | Lognormal | -0.010 | (-0.031, 0.011) | 0.356 | 0.871 | 0.101 | (-0.031, 0.012) |
| CE (16:1) | Lognormal | -0.003 | (-0.041, 0.034) | 0.859 | 0.967 | 0.070 | (-0.042, 0.037) |
| Ceramide (d40:0) | Lognormal | -0.052 | (-0.117, 0.012) | 0.120 | 0.864 | 0.231 | (-0.119, 0.015) |
| PC (p-42:5)/PC (o-42:6) B | Lognormal | 0.016 | (-0.012, 0.044) | 0.260 | 0.864 | 0.121 | (-0.013, 0.045) |
| Ceramide (d42:0) | Lognormal | -0.042 | (-0.078, -0.005) | 0.033 | 0.864 | 0.768 | (-0.079, -0.005) |
| LPC (22:4) | Lognormal | -0.033 | (-0.088, 0.021) | 0.241 | 0.864 | 0.112 | (-0.092, 0.019) |
| PC (35:1) - ESI(-) | Lognormal | -0.015 | (-0.043, 0.013) | 0.303 | 0.864 | 0.107 | (-0.044, 0.014) |
| TG (57:2) | Lognormal | -0.023 | (-0.062, 0.016) | 0.263 | 0.864 | 0.131 | (-0.065, 0.016) |

**Supplementary Table S6.** Regression and Bayesian estimates comparing sex-stratified ME/CFS vs. controls.

| Metabolite | Regression Model | Male ME/CFS vs. Male Controls | | | | | |
|---|---|---|---|---|---|---|---|
| | | Estimated Coefficient | 95% CI | p-value | FDR | BayesFactor | 95% HDI |
| PC (40:6) B | Lognormal | -0.005 | (-0.032, 0.022) | 0.704 | 0.937 | 0.074 | (-0.034, 0.023) |
| PC (40:5) B - ESI(+) | Lognormal | -0.012 | (-0.042, 0.017) | 0.422 | 0.873 | 0.091 | (-0.043, 0.017) |
| TG (42:2) | Lognormal | -0.086 | (-0.241, 0.069) | 0.282 | 0.864 | 0.121 | (-0.241, 0.070) |
| CE (22:6) | Lognormal | 0.012 | (-0.011, 0.035) | 0.323 | 0.864 | 0.107 | (-0.010, 0.035) |
| TG (52:3) | Lognormal | 0.007 | (-0.006, 0.021) | 0.268 | 0.864 | 0.130 | (-0.005, 0.021) |
| PC (38:7) | Lognormal | 0.012 | (-0.018, 0.043) | 0.422 | 0.873 | 0.099 | (-0.020, 0.045) |
| TG (60:3) | Lognormal | -0.010 | (-0.085, 0.065) | 0.803 | 0.956 | 0.076 | (-0.090, 0.067) |
| CE (22:2) | Lognormal | 0.098 | (0.035, 0.161) | 0.004 | 0.864 | 3.006 | (0.032, 0.164) |
| FA (22:6) (docosahexaenoic acid) | Lognormal | -0.003 | (-0.031, 0.025) | 0.815 | 0.956 | 0.067 | (-0.033, 0.026) |
| PC (36:4) C - ESI(+) | Lognormal | -0.004 | (-0.016, 0.009) | 0.545 | 0.903 | 0.087 | (-0.017, 0.009) |
| DG (34:1) | Lognormal | -0.004 | (-0.046, 0.038) | 0.845 | 0.963 | 0.063 | (-0.045, 0.042) |
| PE (36:1) - ESI(+) | Lognormal | -0.028 | (-0.074, 0.018) | 0.234 | 0.864 | 0.144 | (-0.073, 0.019) |
| TG (53:1) | Lognormal | -0.046 | (-0.114, 0.022) | 0.196 | 0.864 | 0.168 | (-0.118, 0.025) |
| TG (46:1) | Lognormal | -0.052 | (-0.122, 0.017) | 0.146 | 0.864 | 0.192 | (-0.127, 0.017) |
| PC (35:4) - ESI(+) | Lognormal | -0.002 | (-0.029, 0.024) | 0.856 | 0.966 | 0.075 | (-0.030, 0.023) |
| SM (d42:2) | Lognormal | 0.013 | (-0.004, 0.029) | 0.138 | 0.864 | 0.205 | (-0.004, 0.030) |
| DG (36:3) | Lognormal | 0.011 | (-0.009, 0.032) | 0.280 | 0.864 | 0.131 | (-0.010, 0.031) |
| SM (d37:1) | Lognormal | -0.024 | (-0.082, 0.033) | 0.406 | 0.873 | 0.089 | (-0.082, 0.030) |
| LPC (20:1) - ESI(-) | Lognormal | -0.012 | (-0.045, 0.020) | 0.460 | 0.884 | 0.091 | (-0.044, 0.022) |
| TG (53:3) | Lognormal | 0.012 | (-0.016, 0.039) | 0.419 | 0.873 | 0.098 | (-0.015, 0.040) |
| TG (52:5) | Lognormal | 0.011 | (-0.019, 0.040) | 0.486 | 0.884 | 0.083 | (-0.018, 0.042) |
| TG (48:6) | Lognormal | -0.020 | (-0.058, 0.019) | 0.321 | 0.864 | 0.124 | (-0.060, 0.018) |
| TG (54:4) | Lognormal | 0.015 | (-0.009, 0.039) | 0.233 | 0.864 | 0.143 | (-0.009, 0.039) |
| GlcCer (d34:1) | Lognormal | 0.018 | (-0.012, 0.047) | 0.252 | 0.864 | 0.123 | (-0.013, 0.048) |
| SM (d41:2) - ESI(-) | Lognormal | -0.015 | (-0.045, 0.015) | 0.328 | 0.864 | 0.116 | (-0.044, 0.017) |
| PC (p-40:4)/PC (o-40:5) - ESI(+) | Lognormal | 0.003 | (-0.015, 0.022) | 0.745 | 0.949 | 0.072 | (-0.016, 0.022) |
| SM (d32:0) - ESI(-) | Lognormal | -0.067 | (-0.180, 0.046) | 0.255 | 0.864 | 0.137 | (-0.181, 0.048) |
| LPC (22:6) | Lognormal | -0.009 | (-0.037, 0.019) | 0.526 | 0.899 | 0.083 | (-0.037, 0.018) |
| PC 38:7e | Lognormal | -0.005 | (-0.041, 0.031) | 0.799 | 0.956 | 0.072 | (-0.041, 0.034) |
| LPC (20:1) - ESI(+) | Lognormal | -0.025 | (-0.074, 0.025) | 0.335 | 0.864 | 0.094 | (-0.074, 0.026) |
| TG (50:6) | Lognormal | -0.034 | (-0.088, 0.019) | 0.218 | 0.864 | 0.151 | (-0.090, 0.021) |
| TG (55:1) | Lognormal | -0.023 | (-0.069, 0.024) | 0.345 | 0.864 | 0.104 | (-0.071, 0.025) |
| FA (15:0) (pentadecylic acid) | Lognormal | -0.004 | (-0.025, 0.016) | 0.684 | 0.929 | 0.080 | (-0.026, 0.016) |
| TG (44:1) | Lognormal | -0.074 | (-0.162, 0.013) | 0.105 | 0.864 | 0.275 | (-0.170, 0.014) |
| PC (28:0) | Lognormal | -0.064 | (-0.138, 0.011) | 0.101 | 0.864 | 0.249 | (-0.139, 0.015) |
| TG (59:2) | Lognormal | -0.013 | (-0.070, 0.044) | 0.653 | 0.928 | 0.081 | (-0.070, 0.047) |
| PC (38:3) - ESI(+) | Lognormal | -0.009 | (-0.027, 0.010) | 0.363 | 0.873 | 0.094 | (-0.027, 0.010) |
| PC (p-42:4)/PC (o-42:5) - ESI(+) | Lognormal | 0.009 | (-0.013, 0.032) | 0.429 | 0.873 | 0.091 | (-0.013, 0.033) |
| Ceramide (d36:1) - ESI(-) | Lognormal | -0.010 | (-0.054, 0.034) | 0.659 | 0.928 | 0.073 | (-0.054, 0.035) |
| TG (62:1) | Lognormal | -0.010 | (-0.073, 0.052) | 0.752 | 0.952 | 0.071 | (-0.075, 0.054) |
| PC (33:0) | Lognormal | -0.004 | (-0.045, 0.037) | 0.842 | 0.962 | 0.071 | (-0.043, 0.040) |
| TG (52:6) | Lognormal | -0.030 | (-0.072, 0.012) | 0.172 | 0.864 | 0.178 | (-0.073, 0.012) |
| TG (46:5) | Lognormal | 0.008 | (-0.086, 0.102) | 0.868 | 0.968 | 0.069 | (-0.087, 0.107) |
| FA (17:0) (margaric acid) | Lognormal | 0.003 | (-0.008, 0.015) | 0.585 | 0.925 | 0.074 | (-0.008, 0.015) |
| TG (58:1) | Lognormal | -0.034 | (-0.094, 0.026) | 0.273 | 0.864 | 0.137 | (-0.099, 0.027) |
| SM (d33:1) - ESI(+) | Lognormal | 0.002 | (-0.019, 0.024) | 0.826 | 0.962 | 0.070 | (-0.019, 0.025) |
| PC (36:5) B | Lognormal | -0.006 | (-0.049, 0.037) | 0.797 | 0.956 | 0.066 | (-0.052, 0.038) |

**Supplementary Table S6.** Regression and Bayesian estimates comparing sex-stratified ME/CFS vs. controls.

| Metabolite | Regression Model | Male ME/CFS vs. Male Controls | | | | | |
|---|---|---|---|---|---|---|---|
| | | Estimated Coefficient | 95% CI | p-value | FDR | BayesFactor | 95% HDI |
| TG (54:3) | Lognormal | 0.006 | (-0.018, 0.031) | 0.603 | 0.926 | 0.088 | (-0.019, 0.029) |
| FA (12:0) (lauric acid) | Lognormal | -0.001 | (-0.035, 0.032) | 0.932 | 0.992 | 0.074 | (-0.036, 0.034) |
| CE (16:0) | Lognormal | 0.029 | (-0.021, 0.079) | 0.268 | 0.864 | 0.103 | (-0.025, 0.078) |
| PE (38:4) | Lognormal | -0.017 | (-0.041, 0.008) | 0.195 | 0.864 | 0.118 | (-0.041, 0.008) |
| TG (53:4) | Lognormal | 0.012 | (-0.018, 0.042) | 0.443 | 0.881 | 0.089 | (-0.018, 0.044) |
| TG (60:1) | Lognormal | -0.030 | (-0.091, 0.031) | 0.342 | 0.864 | 0.109 | (-0.092, 0.033) |
| DG (34:3) | Lognormal | 0.016 | (-0.022, 0.054) | 0.413 | 0.873 | 0.084 | (-0.023, 0.055) |
| PC (38:5) B - ESI(-) | Lognormal | -0.004 | (-0.038, 0.031) | 0.840 | 0.962 | 0.071 | (-0.039, 0.034) |
| FA (18:3) (linolenic acid) | Lognormal | 0.020 | (-0.010, 0.050) | 0.195 | 0.864 | 0.161 | (-0.011, 0.050) |
| GlcCer (d42:1) - ESI(+) | Lognormal | 0.001 | (-0.018, 0.020) | 0.919 | 0.983 | 0.060 | (-0.019, 0.021) |
| PC (31:1) | Lognormal | -0.002 | (-0.087, 0.083) | 0.963 | 0.996 | 0.072 | (-0.082, 0.088) |
| PC (38:6) - ESI(-) | Lognormal | -0.011 | (-0.036, 0.015) | 0.405 | 0.873 | 0.090 | (-0.035, 0.017) |
| PC (p-44:4)/PC (o-44:5) - ESI(+) | Lognormal | 0.019 | (-0.005, 0.042) | 0.129 | 0.864 | 0.231 | (-0.004, 0.044) |
| CE (18:0) | Lognormal | 0.042 | (0.010, 0.074) | 0.014 | 0.864 | 1.498 | (0.010, 0.076) |
| PC (40:6) A | Lognormal | -0.006 | (-0.036, 0.024) | 0.707 | 0.937 | 0.070 | (-0.037, 0.026) |
| GlcCer (d40:1) - ESI(+) | Lognormal | -0.006 | (-0.034, 0.022) | 0.674 | 0.928 | 0.077 | (-0.036, 0.021) |
| Ceramide (d34:0) | Lognormal | -0.016 | (-0.046, 0.014) | 0.308 | 0.864 | 0.091 | (-0.050, 0.012) |
| TG (57:1) | Lognormal | -0.026 | (-0.070, 0.018) | 0.248 | 0.864 | 0.131 | (-0.070, 0.018) |
| PC (16:0/9:0(CHO)) | Lognormal | -0.005 | (-0.053, 0.043) | 0.840 | 0.962 | 0.057 | (-0.056, 0.042) |
| PC (38:4) A - ESI(+) | Lognormal | -0.002 | (-0.020, 0.015) | 0.786 | 0.956 | 0.070 | (-0.021, 0.016) |
| TG (52:1) | Lognormal | -0.023 | (-0.069, 0.024) | 0.343 | 0.864 | 0.106 | (-0.070, 0.023) |
| TG (49:3) | Lognormal | -0.008 | (-0.058, 0.043) | 0.771 | 0.956 | 0.071 | (-0.060, 0.044) |
| Ceramide (d40:2) | Lognormal | -0.011 | (-0.090, 0.068) | 0.787 | 0.956 | 0.071 | (-0.092, 0.070) |
| PC (p-42:5)/PC (o-42:6) | Lognormal | -0.023 | (-0.076, 0.030) | 0.399 | 0.873 | 0.110 | (-0.076, 0.033) |
| TG (56:1) | Lognormal | -0.020 | (-0.081, 0.040) | 0.511 | 0.899 | 0.082 | (-0.077, 0.046) |
| CE (20:4) | Lognormal | -0.004 | (-0.028, 0.020) | 0.766 | 0.956 | 0.079 | (-0.028, 0.022) |
| TG (49:0) | Lognormal | -0.014 | (-0.060, 0.032) | 0.556 | 0.905 | 0.091 | (-0.060, 0.034) |
| PC (40:5) A - ESI(-) | Lognormal | -0.002 | (-0.035, 0.031) | 0.902 | 0.976 | 0.068 | (-0.038, 0.031) |
| SM (d36:1) - ESI(+) | Lognormal | 0.006 | (-0.011, 0.023) | 0.526 | 0.899 | 0.083 | (-0.011, 0.023) |
| TG (58:5) | Lognormal | -0.020 | (-0.058, 0.019) | 0.321 | 0.864 | 0.123 | (-0.057, 0.020) |
| PC (38:4) C - ESI(+) | Lognormal | -0.004 | (-0.017, 0.009) | 0.508 | 0.899 | 0.087 | (-0.017, 0.010) |
| LPC (20:0) | Lognormal | -0.013 | (-0.046, 0.020) | 0.442 | 0.881 | 0.084 | (-0.047, 0.022) |
| PC (40:7) - ESI(-) | Lognormal | -0.012 | (-0.050, 0.025) | 0.525 | 0.899 | 0.079 | (-0.049, 0.026) |
| DG (34:2) | Lognormal | 0.003 | (-0.020, 0.026) | 0.797 | 0.956 | 0.062 | (-0.022, 0.026) |
| LPC (p-18:0)/LPC (o-18:1) | Lognormal | -0.006 | (-0.032, 0.020) | 0.637 | 0.928 | 0.082 | (-0.031, 0.021) |
| TG (58:2) | Lognormal | -0.132 | (-0.277, 0.013) | 0.083 | 0.864 | 0.342 | (-0.285, 0.021) |
| PC (31:0) | Lognormal | 0.001 | (-0.025, 0.027) | 0.917 | 0.983 | 0.073 | (-0.024, 0.028) |
| PC (40:8) - ESI(+) | Lognormal | -0.011 | (-0.029, 0.008) | 0.262 | 0.864 | 0.109 | (-0.030, 0.009) |
| PC (p-38:5)/PC (o-38:6) A | Lognormal | 0.000 | (-0.027, 0.026) | 0.973 | 0.996 | 0.065 | (-0.028, 0.026) |
| TG (62:2) | Lognormal | -0.024 | (-0.081, 0.033) | 0.417 | 0.873 | 0.102 | (-0.086, 0.032) |
| PE (34:2) - ESI(+) | Lognormal | -0.119 | (-0.260, 0.022) | 0.107 | 0.864 | 0.292 | (-0.268, 0.020) |
| TG (50:3) A | Lognormal | -0.004 | (-0.037, 0.028) | 0.801 | 0.956 | 0.076 | (-0.036, 0.029) |
| TG (51:3) | Lognormal | 0.008 | (-0.024, 0.039) | 0.639 | 0.928 | 0.078 | (-0.025, 0.039) |
| PC (p-40:4)/PC (o-40:5) - ESI(-) | Lognormal | 0.001 | (-0.022, 0.025) | 0.908 | 0.977 | 0.065 | (-0.022, 0.026) |
| PC (36:6) | Lognormal | -0.015 | (-0.056, 0.026) | 0.481 | 0.884 | 0.093 | (-0.058, 0.026) |
| FA (10:0) (capric acid) | Lognormal | -0.005 | (-0.052, 0.041) | 0.824 | 0.962 | 0.056 | (-0.054, 0.042) |
| Ceramide (d42:2) A - ESI (+) | Lognormal | 0.012 | (-0.006, 0.030) | 0.198 | 0.864 | 0.165 | (-0.007, 0.030) |

**Supplementary Table S6.** Regression and Bayesian estimates comparing sex-stratified ME/CFS vs. controls.

| Metabolite | Regression Model | Male ME/CFS vs. Male Controls | | | | | |
|---|---|---|---|---|---|---|---|
| | | Estimated Coefficient | 95% CI | p-value | FDR | BayesFactor | 95% HDI |
| Gal-Gal-Cer (d18:1/16:0)/Lactosylceramide (d18:1/16:0) | Lognormal | 0.012 | (-0.002, 0.026) | 0.101 | 0.864 | 0.277 | (-0.002, 0.026) |
| PE (36:4) - ESI(-) | Lognormal | -0.047 | (-0.103, 0.010) | 0.112 | 0.864 | 0.234 | (-0.106, 0.008) |
| LPC (18:1) - ESI(+) | Lognormal | -0.016 | (-0.038, 0.005) | 0.148 | 0.864 | 0.203 | (-0.038, 0.005) |
| PC (p-36:5)/PC (o-36:6) | Lognormal | -0.008 | (-0.049, 0.034) | 0.711 | 0.939 | 0.069 | (-0.051, 0.033) |
| TG (56:4) | Lognormal | 0.006 | (-0.022, 0.034) | 0.681 | 0.929 | 0.084 | (-0.023, 0.036) |
| PC 40:6e | Lognormal | 0.007 | (-0.013, 0.027) | 0.503 | 0.899 | 0.085 | (-0.013, 0.027) |
| PC (p-44:5)/PC (o-44:6) | Lognormal | 0.016 | (-0.013, 0.044) | 0.299 | 0.864 | 0.107 | (-0.014, 0.046) |
| SM (d41:2) A - ESI(+) | Lognormal | 0.007 | (-0.016, 0.031) | 0.543 | 0.902 | 0.083 | (-0.015, 0.032) |
| TG (50:3) B | Lognormal | -0.047 | (-0.150, 0.057) | 0.381 | 0.873 | 0.116 | (-0.150, 0.067) |
| TG (44:2) | Lognormal | -0.027 | (-0.177, 0.123) | 0.724 | 0.944 | 0.077 | (-0.187, 0.123) |
| TG (48:1) | Lognormal | -0.030 | (-0.075, 0.016) | 0.206 | 0.864 | 0.172 | (-0.074, 0.018) |
| SM (d43:1) - ESI(+) | Lognormal | -0.001 | (-0.035, 0.033) | 0.958 | 0.996 | 0.061 | (-0.036, 0.033) |
| LPC (22:5) - ESI(-) | Lognormal | -0.038 | (-0.096, 0.020) | 0.203 | 0.864 | 0.158 | (-0.099, 0.019) |
| TG (48:2) | Lognormal | -0.034 | (-0.084, 0.017) | 0.197 | 0.864 | 0.166 | (-0.083, 0.017) |
| FA (24:1) (nervonic acid) | Lognormal | -0.002 | (-0.022, 0.018) | 0.841 | 0.962 | 0.052 | (-0.024, 0.018) |
| FA (20:4) (arachidonic acid) | Lognormal | -0.010 | (-0.023, 0.004) | 0.176 | 0.864 | 0.140 | (-0.023, 0.004) |
| PC (40:4) - ESI(+) | Lognormal | -0.009 | (-0.033, 0.015) | 0.452 | 0.884 | 0.087 | (-0.032, 0.017) |
| PC (32:1) - ESI(-) | Lognormal | -0.029 | (-0.070, 0.013) | 0.185 | 0.864 | 0.176 | (-0.071, 0.013) |
| PC (38:4) B - ESI(+) | Lognormal | -0.010 | (-0.033, 0.013) | 0.407 | 0.873 | 0.091 | (-0.034, 0.014) |
| PC (34:0) - ESI(+) | Lognormal | -0.002 | (-0.014, 0.010) | 0.736 | 0.949 | 0.058 | (-0.015, 0.010) |
| Ceramide (d36:1) - ESI(+) | Lognormal | 0.020 | (-0.007, 0.046) | 0.163 | 0.864 | 0.176 | (-0.009, 0.046) |
| **Oxylipins (OL)** | | | | | | | |
| Resolvin D1 | Gamma | -0.282 | (-0.991, 0.426) | 0.439 | 0.881 | 0.099 | (-1.004, 0.410) |
| 5-Hydroxy-6,8,11,14,17-eicosapentaenoic acid | Gamma | 0.055 | (-0.345, 0.455) | 0.788 | 0.956 | 0.043 | (-0.386, 0.454) |
| 9-nitrooleic acid | Gamma | -0.634 | (-1.110, -0.158) | 0.013 | 0.864 | 0.503 | (-1.279, -0.003) |
| 15,16-dihydroxyoctadeca-9,12-dienoic acid | Lognormal | 0.072 | (-0.280, 0.425) | 0.690 | 0.930 | 0.059 | (-0.277, 0.445) |
| 15-hydroxyeicosa-5,8,11,13,17-pentaenoic acid | Gamma | -0.180 | (-0.668, 0.308) | 0.474 | 0.884 | 0.070 | (-0.747, 0.395) |
| 12-Hydroxy-5,8,10,14,17-eicosapentaenoic acid | Gamma | 0.205 | (-0.315, 0.725) | 0.444 | 0.882 | 0.071 | (-0.349, 0.790) |
| Prostaglandin F2a | Gamma | -0.150 | (-0.625, 0.326) | 0.540 | 0.902 | 0.067 | (-0.729, 0.514) |
| Prostaglandin D2 | Gamma | 0.617 | (-0.488, 1.722) | 0.280 | 0.864 | 0.121 | (-0.708, 1.337) |
| 4-hydroxydocosa-5,7,10,13,16,19-hexaenoic acid | Gamma | -0.065 | (-0.654, 0.523) | 0.829 | 0.962 | 0.054 | (-0.677, 0.511) |
| 9-Hydroxy-5,7,11,14,17-icosapentaenoic acid | Gamma | 0.189 | (-0.285, 0.664) | 0.438 | 0.881 | 0.067 | (-0.364, 0.689) |
| 8,9-Epoxyeicosa-5,11,14-trienoic acid | Gamma | -0.037 | (-0.486, 0.412) | 0.873 | 0.969 | 0.054 | (-0.543, 0.484) |
| 9,10-dihydroxyoctadec-12-enoic acid | Lognormal | 0.060 | (-0.279, 0.398) | 0.732 | 0.949 | 0.060 | (-0.302, 0.392) |
| 11,12-Epoxyeicosa-5,8,14-trienoic acid | Gamma | 0.426 | (-0.006, 0.859) | 0.061 | 0.864 | 0.228 | (-0.035, 0.906) |
| 11(12)-epoxy-5,8,14,17- eicosatetraenoic acid | Gamma | -1.314 | (-1.984, -0.645) | 0.000 | 0.374 | 5.610 | (-2.063, -0.433) |
| 8-hydroxyeicosa-5,9,11,14-tetraenoic acid | Gamma | -0.330 | (-0.848, 0.188) | 0.219 | 0.864 | 0.114 | (-0.950, 0.267) |
| 6-Ketoprostaglandin F1 alpha | Gamma | -0.620 | (-1.324, 0.084) | 0.092 | 0.864 | 0.326 | (-1.336, 0.037) |
| 15-ketoeicosa-5,8,11,13-tetraenoic acid | Gamma | -0.217 | (-0.800, 0.366) | 0.470 | 0.884 | 0.080 | (-0.779, 0.402) |
| 17,18-Epoxy-5,8,11,14-eicosatetraenoic acid | Gamma | -0.820 | (-1.753, 0.113) | 0.093 | 0.864 | 0.394 | (-1.788, 0.186) |
| 17,18-dihydroxyeicosa-5,8,11,14-tetraenoic acid | Lognormal | 0.108 | (-0.398, 0.613) | 0.679 | 0.929 | 0.064 | (-0.381, 0.661) |
| 15-Deoxy-delta-12,14-Prostaglandin J2 | Gamma | -0.091 | (-0.626, 0.445) | 0.742 | 0.949 | 0.075 | (-0.784, 0.559) |
| 15(16)-epoxy-9,12-octadecadienoic acid | Gamma | -0.043 | (-0.655, 0.570) | 0.892 | 0.976 | 0.059 | (-0.660, 0.592) |
| 9,11,15-trihydroxy-5,13,1Z-prostatrienoic acid | Gamma | -0.734 | (-1.454, -0.014) | 0.053 | 0.864 | 0.650 | (-1.489, 0.011) |
| 9,10-dihydroxyoctadeca-12,15-dienoic acid | Gamma | 0.307 | (0.011, 0.645) | 0.054 | 0.864 | 1.568 | (-0.009, 0.549) |
| 9(10)-epoxy-12Z-octadecenoic acid | Gamma | 0.214 | (-0.237, 0.665) | 0.358 | 0.872 | 0.067 | (-0.204, 0.639) |
| 9(10)-epoxy-12,15-octadecadienoic acid | Gamma | -0.277 | (-0.847, 0.294) | 0.347 | 0.864 | 0.099 | (-1.019, 0.338) |

**Supplementary Table S6.** Regression and Bayesian estimates comparing sex-stratified ME/CFS vs. controls.

| Metabolite | Regression Model | Male ME/CFS vs. Male Controls | | | | | |
|---|---|---|---|---|---|---|---|
| | | Estimated Coefficient | 95% CI | p-value | FDR | BayesFactor | 95% HDI |
| 10-nitrooleic acid | Lognormal | -0.127 | (-0.610, 0.356) | 0.610 | 0.928 | 0.073 | (-0.658, 0.357) |
| 11-Hydroxy-arachidonic acid | Gamma | -0.095 | (-0.449, 0.259) | 0.602 | 0.926 | 0.047 | (-0.481, 0.322) |
| 6-trans-Leukotriene B4 | Gamma | 0.781 | (0.124, 1.438) | 0.025 | 0.864 | 0.581 | (-0.017, 1.491) |
| 8,9-dihydroxyeicosa-5,11,14-trienoic acid | Gamma | -0.006 | (-0.492, 0.480) | 0.981 | 0.996 | 0.058 | (-0.593, 0.572) |
| 13-hydroxyoctadeca-9,11,15-trienoic acid | Gamma | 0.230 | (-0.238, 0.699) | 0.340 | 0.864 | 0.072 | (-0.204, 0.730) |
| 12(13)-epoxy-9,15-octadecadienoic acid | Gamma | 0.500 | (-0.032, 1.031) | 0.073 | 0.864 | 0.177 | (-0.226, 1.167) |
| Leukotriene B5 | Gamma | 0.384 | (-0.498, 1.265) | 0.399 | 0.873 | 0.144 | (-0.857, 1.507) |
| 12,13-dihydroxyoctadec-9-enoic acid | Lognormal | -0.178 | (-0.573, 0.217) | 0.382 | 0.873 | 0.084 | (-0.598, 0.227) |
| 9-Hydroxylinoleic acid | Lognormal | 0.000 | (-0.241, 0.241) | 0.998 | 1.000 | 0.059 | (-0.242, 0.255) |
| 13-ketooctadeca-9,11-dienoic acid | Lognormal | -0.252 | (-0.698, 0.195) | 0.276 | 0.864 | 0.106 | (-0.716, 0.199) |
| 16(17)-epoxy-4,7,10,13,19-docosapentaenoic acid | Gamma | 0.271 | (-0.365, 0.906) | 0.409 | 0.873 | 0.096 | (-0.510, 1.015) |
| 5,6-dihydroxyeicosa-8,11,14-trienoic acid | Gamma | 0.001 | (-0.445, 0.448) | 0.995 | 0.998 | 0.051 | (-0.503, 0.503) |
| 17-hydroxy-4,7,10,13,15,19-docosahexaenoic acid | Gamma | 0.217 | (-0.247, 0.680) | 0.365 | 0.873 | 0.071 | (-0.310, 0.716) |
| 9,12,13-trihydroxyoctadec-10-enoic acid | Lognormal | 0.070 | (-0.198, 0.338) | 0.613 | 0.928 | 0.061 | (-0.189, 0.339) |
| 10-nitrolinoleic acid | Gamma | 0.386 | (-0.311, 1.083) | 0.284 | 0.864 | 0.119 | (-0.445, 1.141) |
| Prostaglandin E2 | Gamma | -0.221 | (-0.798, 0.357) | 0.458 | 0.884 | 0.082 | (-0.815, 0.386) |
| 9-hydroxyeicosa-5,7,11,14-tetraenoic acid | Gamma | -0.232 | (-0.656, 0.192) | 0.290 | 0.864 | 0.082 | (-0.691, 0.264) |
| Prostaglandin E1 | Gamma | 0.309 | (-0.320, 0.939) | 0.341 | 0.864 | 0.099 | (-0.335, 0.935) |
| 8,15-dihydroxyeicosa-5,9,11,13-tetraenoic acid | Gamma | -0.350 | (-1.137, 0.437) | 0.388 | 0.873 | 0.123 | (-1.374, 0.779) |
| 9-hydroxyoctadeca-10,12,15-trienoic acid | Gamma | 0.091 | (-0.289, 0.471) | 0.643 | 0.928 | 0.048 | (-0.347, 0.551) |
| 14(15)-epoxy-5,8,11,17-eicosatetraenoic acid | Gamma | 0.809 | (-0.142, 1.760) | 0.103 | 0.864 | 0.435 | (-0.145, 1.677) |
| 9,10-Epoxystearic acid | Lognormal | -0.591 | (-1.261, 0.078) | 0.091 | 0.864 | 0.269 | (-1.271, 0.138) |
| 5,6,15-trihydroxyeicosa-7,9,11,13-tetraenoic acid | Gamma | -0.583 | (-1.229, 0.063) | 0.085 | 0.864 | 0.365 | (-1.263, 0.038) |
| 12,13-epoxy-9-octadecenoic acid | Lognormal | -0.291 | (-0.946, 0.363) | 0.388 | 0.873 | 0.093 | (-0.986, 0.373) |
| Thromboxane B2 | Lognormal | 0.031 | (-0.432, 0.494) | 0.897 | 0.976 | 0.063 | (-0.441, 0.513) |
| 20-Hydroxyarachidonic acid | Lognormal | 0.208 | (-0.372, 0.788) | 0.486 | 0.884 | 0.070 | (-0.352, 0.823) |
| Leukotriene B4 | Gamma | -0.324 | (-0.760, 0.111) | 0.152 | 0.864 | 0.120 | (-0.787, 0.147) |
| 13-Hydroxyoctadecadienoic acid | Lognormal | 0.011 | (-0.251, 0.273) | 0.935 | 0.992 | 0.068 | (-0.253, 0.279) |
| 9S,10R-dihydroxy-stearic acid | Lognormal | 0.064 | (-0.467, 0.594) | 0.815 | 0.956 | 0.068 | (-0.475, 0.619) |
| 5-ketoeicosa-6,8,11,14-tetraenoic acid | Gamma | -0.282 | (-0.848, 0.284) | 0.335 | 0.864 | 0.107 | (-0.975, 0.426) |
| Prostaglandin E3 | Gamma | 0.969 | (0.038, 1.901) | 0.048 | 0.864 | 0.431 | (-0.267, 2.086) |
| 5,15-dihydroxyeicosa-6,8,11,13-tetraenoic acid | Gamma | 0.444 | (-0.426, 1.314) | 0.323 | 0.864 | 0.138 | (-0.464, 1.207) |
| 12,13-dihydroxyoctadeca-9,15-dienoic acid | Gamma | -0.100 | (-0.765, 0.566) | 0.771 | 0.956 | 0.073 | (-0.765, 0.580) |
| 14-hydroxydocosa-4,7,10,12,16,19-hexaenoic acid | Lognormal | -0.129 | (-0.919, 0.662) | 0.751 | 0.952 | 0.054 | (-0.947, 0.690) |
| trans-12,13-epoxy-11-oxo-trans-9-octadecenoic acid | Gamma | 0.112 | (-0.336, 0.560) | 0.628 | 0.928 | 0.053 | (-0.390, 0.593) |
| 14,15-dihydroxyeicosa-5,8,11,17-tetraenoic acid | Gamma | -0.563 | (-1.200, 0.075) | 0.091 | 0.864 | 0.267 | (-1.276, 0.116) |
| 19,20-dihydroxydocosa-4,7,10,13,16-pentaenoic acid | Lognormal | 0.105 | (-0.241, 0.452) | 0.555 | 0.905 | 0.062 | (-0.259, 0.435) |
| 5-Hydroxy-6,8,11,14-eicosatetraenoic acid | Lognormal | -0.179 | (-0.596, 0.238) | 0.405 | 0.873 | 0.074 | (-0.580, 0.229) |
| 14,15-dihydroxyeicosa-5,8,11-trienoic acid | Gamma | 0.031 | (-0.256, 0.330) | 0.839 | 0.962 | 0.067 | (-0.295, 0.359) |
| 18-(3-ethyloxiran-2-yl)octadeca-4,7,10,13,16-pentaenoic acid | Gamma | 0.354 | (-0.262, 0.969) | 0.267 | 0.864 | 0.112 | (-0.291, 1.021) |
| 15-hydroxyeicosa-5,8,11,13-tetraenoic acid | Lognormal | 0.071 | (-0.267, 0.409) | 0.682 | 0.929 | 0.061 | (-0.283, 0.397) |
| 12-Hydroxy-5,8,10,14-eicosatetraenoic acid | Lognormal | 0.087 | (-0.400, 0.574) | 0.728 | 0.947 | 0.052 | (-0.414, 0.575) |
| 11-Hydroxy-14,15-epoxyeicosatrienoic acid | Gamma | -0.642 | (-1.380, 0.096) | 0.096 | 0.864 | 0.337 | (-1.450, 0.103) |
| 9-ketooctadeca-10,12-dienoic acid | Gamma | -0.187 | (-0.539, 0.162) | 0.278 | 0.864 | 0.092 | (-0.720, 0.184) |
| 15-Keto-prostaglandin E2 | Gamma | -0.673 | (-1.265, -0.081) | 0.032 | 0.864 | 0.515 | (-1.369, 0.054) |
| 11,12-Dihydroxyicosa-5,8,14-trienoic acid | Gamma | -0.038 | (-0.345, 0.268) | 0.807 | 0.956 | 0.040 | (-0.417, 0.352) |

**Supplementary Table S6.** Regression and Bayesian estimates comparing sex-stratified ME/CFS vs. controls.

| Metabolite | Regression Model | Male ME/CFS vs. Male Controls | | | | | |
|---|---|---|---|---|---|---|---|
| | | Estimated Coefficient | 95% CI | p-value | FDR | BayesFactor | 95% HDI |

ME/CFS, myalgic encephalomyelitis/chronic fatigue syndrome; CI, confidence interval; FDR, false discovery rate adjusted p-value; HDI, highest density credible intervals. Regression models were adjusted for age, race/ethnicity, geographic/clinical site, season of sampling, body mass index, and self-reported irritable bowel syndrome. In PM, BA, and CL panels, estimated coefficients are interpreted as the differences in the mean values of log-log transformation of metabolite levels between cases and controls. In OL panel for lognormal regression, estimated coefficients are interpreted as the mean differences of log transformation of metabolite levels between two groups; for Gamma regression, estimated coefficients are interpreted as the log of fold change between two groups. Estimations in **bold** are significant. Criteria for significance: 1) FDR adjusted p-value from the regression model < 0.15, 2) BayesFactor > 3, and 3) 95% highest density credible intervals not covering 0.

**Supplementary Table S7.** Regression and Bayesian estimates comparing ME/CFS without sr-IBS vs. controls without sr-IBS.

| Metabolite | Regression Model | ME/CFS without sr-IBS vs. Controls without sr-IBS | | | | | |
|---|---|---|---|---|---|---|---|
| | | Estimated Coefficient | 95% CI | p-value | FDR | BayesFactor | 95% HDI |
| **Primary Metabolites (PM)** | | | | | | | |
| mannitol | Lognormal | 0.039 | (0.006, 0.072) | 0.021 | 0.327 | 0.495 | (0.005, 0.072) |
| succinic acid | Lognormal | 0.020 | (0.002, 0.038) | 0.027 | 0.352 | 0.403 | (0.001, 0.038) |
| arachidic acid | Lognormal | 0.023 | (0.002, 0.044) | 0.034 | 0.362 | 0.353 | (0.004, 0.046) |
| ornithine | Lognormal | -0.016 | (-0.032, 0.000) | 0.046 | 0.383 | 0.247 | (-0.032, -0.001) |
| leucine | Lognormal | -0.010 | (-0.021, 0.000) | 0.050 | 0.397 | 0.242 | (-0.020, 0.000) |
| maleic acid | Lognormal | 0.031 | (0.000, 0.062) | 0.054 | 0.402 | 0.235 | (-0.002, 0.061) |
| threonine | Lognormal | -0.013 | (-0.028, 0.002) | 0.098 | 0.522 | 0.146 | (-0.027, 0.003) |
| glucuronic acid | Lognormal | 0.017 | (-0.004, 0.039) | 0.109 | 0.553 | 0.141 | (-0.003, 0.039) |
| 2-hydroxybutanoic acid | Lognormal | 0.014 | (-0.004, 0.032) | 0.118 | 0.560 | 0.115 | (-0.005, 0.032) |
| alpha-ketoglutarate | Lognormal | 0.013 | (-0.003, 0.029) | 0.119 | 0.560 | 0.106 | (-0.003, 0.030) |
| tyrosine | Lognormal | -0.007 | (-0.015, 0.002) | 0.148 | 0.628 | 0.099 | (-0.016, 0.002) |
| pentadecanoic acid | Lognormal | 0.009 | (-0.003, 0.020) | 0.151 | 0.635 | 0.089 | (-0.003, 0.020) |
| glutaric acid | Lognormal | 0.015 | (-0.006, 0.037) | 0.160 | 0.648 | 0.097 | (-0.006, 0.037) |
| 3-hydroxybutyric acid | Lognormal | 0.023 | (-0.009, 0.055) | 0.167 | 0.658 | 0.082 | (-0.010, 0.054) |
| indole-3-propionic acid | Lognormal | -0.019 | (-0.046, 0.008) | 0.170 | 0.659 | 0.088 | (-0.047, 0.007) |
| levoglucosan | Lognormal | -0.017 | (-0.043, 0.008) | 0.191 | 0.678 | 0.087 | (-0.043, 0.007) |
| ribose | Lognormal | 0.009 | (-0.005, 0.023) | 0.215 | 0.698 | 0.085 | (-0.006, 0.023) |
| 1,2,4-benzenetriol | Lognormal | -0.021 | (-0.055, 0.014) | 0.245 | 0.724 | 0.071 | (-0.056, 0.013) |
| 2-ketoisocaproic acid | Lognormal | 0.007 | (-0.005, 0.018) | 0.250 | 0.724 | 0.059 | (-0.005, 0.018) |
| urea | Lognormal | -0.006 | (-0.016, 0.004) | 0.255 | 0.724 | 0.067 | (-0.015, 0.004) |
| aminomalonate | Lognormal | 0.017 | (-0.012, 0.045) | 0.263 | 0.726 | 0.068 | (-0.014, 0.045) |
| indole-3-lactate | Lognormal | -0.008 | (-0.023, 0.007) | 0.285 | 0.741 | 0.061 | (-0.022, 0.008) |
| glutamine | Lognormal | -0.011 | (-0.032, 0.010) | 0.310 | 0.757 | 0.054 | (-0.033, 0.010) |
| pyrrole-2-carboxylic acid | Lognormal | 0.013 | (-0.012, 0.039) | 0.314 | 0.757 | 0.059 | (-0.011, 0.039) |
| glucose-1-phosphate | Lognormal | 0.008 | (-0.008, 0.025) | 0.314 | 0.757 | 0.057 | (-0.007, 0.026) |
| oxoproline | Lognormal | -0.003 | (-0.010, 0.003) | 0.321 | 0.758 | 0.059 | (-0.009, 0.003) |
| citrulline | Lognormal | -0.006 | (-0.018, 0.006) | 0.324 | 0.760 | 0.054 | (-0.018, 0.006) |
| erythritol | Lognormal | 0.016 | (-0.016, 0.047) | 0.327 | 0.761 | 0.059 | (-0.015, 0.050) |
| pseudo uridine | Lognormal | -0.006 | (-0.017, 0.006) | 0.328 | 0.763 | 0.056 | (-0.017, 0.006) |
| glycerol | Lognormal | 0.006 | (-0.006, 0.017) | 0.332 | 0.764 | 0.048 | (-0.006, 0.017) |
| benzoic acid | Lognormal | 0.009 | (-0.010, 0.028) | 0.345 | 0.772 | 0.058 | (-0.010, 0.028) |
| malic acid | Lognormal | 0.010 | (-0.011, 0.032) | 0.352 | 0.784 | 0.052 | (-0.012, 0.030) |
| beta-alanine | Lognormal | -0.011 | (-0.036, 0.013) | 0.364 | 0.805 | 0.055 | (-0.037, 0.011) |
| 2-deoxytetronic acid | Lognormal | 0.010 | (-0.012, 0.032) | 0.384 | 0.825 | 0.052 | (-0.012, 0.033) |
| proline | Lognormal | -0.009 | (-0.032, 0.013) | 0.407 | 0.834 | 0.048 | (-0.031, 0.013) |
| phenylalanine | Lognormal | -0.005 | (-0.015, 0.006) | 0.414 | 0.834 | 0.044 | (-0.015, 0.006) |
| pelargonic acid | Lognormal | 0.006 | (-0.008, 0.020) | 0.421 | 0.834 | 0.047 | (-0.008, 0.019) |
| isoleucine | Lognormal | -0.004 | (-0.016, 0.007) | 0.428 | 0.838 | 0.048 | (-0.015, 0.007) |
| tagatose | Lognormal | 0.007 | (-0.011, 0.026) | 0.437 | 0.846 | 0.042 | (-0.012, 0.026) |
| 4-hydroxybutyric acid | Lognormal | 0.008 | (-0.012, 0.027) | 0.439 | 0.848 | 0.049 | (-0.012, 0.027) |
| 2-hydroxyvaleric acid | Lognormal | 0.007 | (-0.011, 0.025) | 0.456 | 0.855 | 0.045 | (-0.012, 0.026) |
| glycine | Lognormal | -0.003 | (-0.013, 0.006) | 0.488 | 0.879 | 0.042 | (-0.013, 0.006) |
| lysine | Lognormal | -0.010 | (-0.038, 0.018) | 0.491 | 0.879 | 0.040 | (-0.036, 0.019) |
| sucrose | Lognormal | 0.012 | (-0.022, 0.046) | 0.501 | 0.890 | 0.046 | (-0.023, 0.048) |
| glycerol-alpha-phosphate | Lognormal | 0.009 | (-0.018, 0.036) | 0.519 | 0.896 | 0.044 | (-0.020, 0.035) |
| myo-inositol | Lognormal | -0.004 | (-0.017, 0.008) | 0.520 | 0.896 | 0.044 | (-0.016, 0.008) |

**Supplementary Table S7.** Regression and Bayesian estimates comparing ME/CFS without sr-IBS vs. controls without sr-IBS.

| Metabolite | Regression Model | ME/CFS without sr-IBS vs. Controls without sr-IBS | | | | | |
|---|---|---|---|---|---|---|---|
| | | Estimated Coefficient | 95% CI | p-value | FDR | BayesFactor | 95% HDI |
| phosphate | Lognormal | -0.007 | (-0.027, 0.014) | 0.521 | 0.896 | 0.040 | (-0.028, 0.014) |
| lactic acid | Lognormal | -0.005 | (-0.022, 0.011) | 0.530 | 0.896 | 0.040 | (-0.021, 0.012) |
| indole-3-acetate | Lognormal | 0.006 | (-0.014, 0.027) | 0.542 | 0.898 | 0.042 | (-0.014, 0.026) |
| lysine | Lognormal | -0.005 | (-0.023, 0.013) | 0.563 | 0.909 | 0.037 | (-0.024, 0.011) |
| glucose | Lognormal | 0.002 | (-0.005, 0.010) | 0.568 | 0.914 | 0.038 | (-0.006, 0.010) |
| tryptophan | Lognormal | -0.005 | (-0.022, 0.012) | 0.571 | 0.914 | 0.036 | (-0.022, 0.011) |
| palmitoleic acid | Lognormal | -0.008 | (-0.034, 0.019) | 0.573 | 0.914 | 0.042 | (-0.035, 0.018) |
| stearic acid | Lognormal | -0.002 | (-0.011, 0.006) | 0.578 | 0.916 | 0.038 | (-0.010, 0.006) |
| alanine | Lognormal | -0.006 | (-0.029, 0.016) | 0.584 | 0.919 | 0.043 | (-0.031, 0.015) |
| hydroxycarbamate NIST | Lognormal | -0.005 | (-0.025, 0.014) | 0.590 | 0.924 | 0.039 | (-0.026, 0.014) |
| methionine | Lognormal | -0.005 | (-0.022, 0.013) | 0.611 | 0.933 | 0.039 | (-0.024, 0.013) |
| heptadecanoic acid | Lognormal | 0.004 | (-0.013, 0.021) | 0.614 | 0.933 | 0.037 | (-0.012, 0.020) |
| lyxitol | Lognormal | -0.004 | (-0.020, 0.012) | 0.619 | 0.934 | 0.041 | (-0.019, 0.011) |
| linoleic acid | Lognormal | 0.006 | (-0.018, 0.029) | 0.634 | 0.938 | 0.040 | (-0.019, 0.028) |
| trans-4-hydroxyproline | Lognormal | -0.005 | (-0.028, 0.018) | 0.653 | 0.938 | 0.036 | (-0.027, 0.018) |
| maleimide | Lognormal | 0.004 | (-0.015, 0.024) | 0.663 | 0.938 | 0.035 | (-0.016, 0.022) |
| glyceric acid | Lognormal | 0.004 | (-0.015, 0.024) | 0.667 | 0.938 | 0.036 | (-0.016, 0.024) |
| quinic acid | Lognormal | -0.009 | (-0.050, 0.032) | 0.672 | 0.938 | 0.038 | (-0.050, 0.031) |
| ribonic acid | Lognormal | -0.004 | (-0.023, 0.015) | 0.689 | 0.942 | 0.037 | (-0.024, 0.015) |
| mannose | Lognormal | -0.004 | (-0.021, 0.014) | 0.695 | 0.945 | 0.038 | (-0.020, 0.015) |
| adipic acid | Lognormal | -0.004 | (-0.023, 0.015) | 0.701 | 0.945 | 0.036 | (-0.023, 0.015) |
| threonic acid | Lognormal | 0.004 | (-0.017, 0.025) | 0.701 | 0.945 | 0.039 | (-0.018, 0.025) |
| fructose | Lognormal | -0.005 | (-0.034, 0.023) | 0.709 | 0.946 | 0.035 | (-0.033, 0.021) |
| glutamic acid | Lognormal | 0.003 | (-0.015, 0.022) | 0.725 | 0.948 | 0.034 | (-0.015, 0.022) |
| serine | Lognormal | -0.003 | (-0.017, 0.012) | 0.727 | 0.948 | 0.037 | (-0.017, 0.012) |
| gluconic acid | Lognormal | 0.003 | (-0.013, 0.018) | 0.747 | 0.955 | 0.037 | (-0.014, 0.017) |
| conduritol-beta-expoxide | Lognormal | 0.005 | (-0.026, 0.036) | 0.755 | 0.955 | 0.038 | (-0.028, 0.034) |
| myristic acid | Lognormal | 0.002 | (-0.012, 0.017) | 0.759 | 0.955 | 0.039 | (-0.013, 0.017) |
| behenic acid | Lognormal | -0.002 | (-0.019, 0.014) | 0.768 | 0.955 | 0.037 | (-0.019, 0.013) |
| phthalic acid | Lognormal | 0.002 | (-0.013, 0.018) | 0.769 | 0.955 | 0.037 | (-0.013, 0.016) |
| nicotinic acid | Lognormal | -0.005 | (-0.042, 0.033) | 0.798 | 0.960 | 0.032 | (-0.042, 0.034) |
| maltose | Lognormal | 0.002 | (-0.022, 0.027) | 0.847 | 0.981 | 0.033 | (-0.022, 0.028) |
| isopropylbenzene | Lognormal | 0.002 | (-0.017, 0.021) | 0.850 | 0.981 | 0.023 | (-0.017, 0.020) |
| citric acid | Lognormal | 0.001 | (-0.014, 0.017) | 0.858 | 0.981 | 0.031 | (-0.014, 0.017) |
| isothreonic acid | Lognormal | -0.002 | (-0.030, 0.025) | 0.859 | 0.981 | 0.035 | (-0.031, 0.023) |
| uric acid | Lognormal | -0.002 | (-0.020, 0.017) | 0.860 | 0.981 | 0.035 | (-0.021, 0.017) |
| palmitic acid | Lognormal | -0.001 | (-0.009, 0.008) | 0.872 | 0.984 | 0.034 | (-0.009, 0.008) |
| 2-aminobutyric acid | Lognormal | -0.001 | (-0.015, 0.013) | 0.880 | 0.984 | 0.036 | (-0.015, 0.013) |
| glycolic acid | Lognormal | 0.002 | (-0.027, 0.031) | 0.887 | 0.984 | 0.035 | (-0.025, 0.033) |
| salicylic acid | Lognormal | 0.002 | (-0.028, 0.032) | 0.893 | 0.984 | 0.035 | (-0.027, 0.032) |
| capric acid | Lognormal | 0.002 | (-0.023, 0.026) | 0.904 | 0.984 | 0.034 | (-0.022, 0.027) |
| caprylic acid | Lognormal | 0.001 | (-0.013, 0.015) | 0.907 | 0.984 | 0.034 | (-0.012, 0.015) |
| N-acetylputrescine | Lognormal | -0.001 | (-0.020, 0.018) | 0.914 | 0.988 | 0.035 | (-0.020, 0.019) |
| creatinine | Lognormal | 0.001 | (-0.022, 0.025) | 0.918 | 0.989 | 0.033 | (-0.022, 0.025) |
| lauric acid | Lognormal | 0.001 | (-0.022, 0.024) | 0.932 | 0.990 | 0.032 | (-0.022, 0.023) |
| threitol | Lognormal | 0.001 | (-0.017, 0.018) | 0.945 | 0.994 | 0.031 | (-0.016, 0.019) |
| fumaric acid | Lognormal | 0.000 | (-0.019, 0.018) | 0.962 | 0.998 | 0.034 | (-0.019, 0.017) |

**Supplementary Table S7.** Regression and Bayesian estimates comparing ME/CFS without sr-IBS vs. controls without sr-IBS.

| Metabolite | Regression Model | ME/CFS without sr-IBS vs. Controls without sr-IBS | | | | | |
|---|---|---|---|---|---|---|---|
| | | Estimated Coefficient | 95% CI | p-value | FDR | BayesFactor | 95% HDI |
| valine | Lognormal | 0.000 | (-0.010, 0.010) | 0.973 | 0.998 | 0.035 | (-0.010, 0.010) |
| creatine | Lognormal | 0.000 | (-0.025, 0.025) | 0.978 | 0.998 | 0.035 | (-0.025, 0.025) |
| 1-methylgalactose NIST | Lognormal | 0.000 | (-0.035, 0.034) | 0.979 | 0.998 | 0.033 | (-0.035, 0.034) |
| alloxanoic acid | Lognormal | 0.000 | (-0.031, 0.030) | 0.980 | 0.998 | 0.032 | (-0.032, 0.030) |
| pyruvic acid | Lognormal | 0.000 | (-0.017, 0.016) | 0.983 | 0.998 | 0.031 | (-0.017, 0.016) |
| oxalic acid | Lognormal | 0.000 | (-0.041, 0.040) | 0.986 | 0.998 | 0.036 | (-0.039, 0.042) |
| N-acetylornithine | Lognormal | 0.000 | (-0.013, 0.012) | 0.988 | 0.998 | 0.034 | (-0.012, 0.012) |
| **Biogenic Amines (BA)** | | | | | | | |
| Acetominophen | Lognormal | 0.066 | (0.023, 0.109) | 0.003 | 0.291 | 2.586 | (0.024, 0.109) |
| Acyclovir | Lognormal | 0.104 | (0.031, 0.177) | 0.006 | 0.310 | 1.688 | (0.034, 0.180) |
| Guanine | Lognormal | 0.087 | (0.026, 0.148) | 0.006 | 0.310 | 1.280 | (0.022, 0.147) |
| 2-Methylbutyryl-L-carnitine | Lognormal | -0.035 | (-0.059, -0.010) | 0.006 | 0.310 | 1.361 | (-0.060, -0.011) |
| Alprazolam | Lognormal | 0.068 | (0.018, 0.119) | 0.008 | 0.310 | 0.850 | (0.015, 0.118) |
| SDMA | Lognormal | -0.013 | (-0.022, -0.003) | 0.011 | 0.310 | 0.900 | (-0.022, -0.003) |
| Choline cation | Lognormal | -0.008 | (-0.014, -0.002) | 0.015 | 0.310 | 0.591 | (-0.014, -0.001) |
| L-Citrulline | Lognormal | -0.012 | (-0.021, -0.002) | 0.017 | 0.310 | 0.620 | (-0.022, -0.002) |
| Ranitidine N-oxide | Lognormal | 0.061 | (0.011, 0.111) | 0.019 | 0.325 | 0.680 | (0.011, 0.110) |
| Albendazole sulfoxide | Lognormal | 0.105 | (0.014, 0.195) | 0.024 | 0.339 | 0.458 | (0.015, 0.197) |
| Caffeine | Lognormal | -0.064 | (-0.120, -0.008) | 0.026 | 0.351 | 0.416 | (-0.119, -0.008) |
| Citrulline | Lognormal | -0.011 | (-0.021, -0.001) | 0.032 | 0.352 | 0.398 | (-0.021, 0.000) |
| Piperine | Lognormal | -0.053 | (-0.102, -0.005) | 0.033 | 0.362 | 0.383 | (-0.103, -0.006) |
| Gabapentin | Lognormal | 0.048 | (0.003, 0.093) | 0.037 | 0.369 | 0.322 | (0.004, 0.093) |
| 4-Acetamidobutyric acid | Lognormal | -0.021 | (-0.041, -0.001) | 0.040 | 0.373 | 0.339 | (-0.040, -0.001) |
| 3-Methylglutarylcarnitine | Lognormal | -0.031 | (-0.060, -0.002) | 0.040 | 0.373 | 0.349 | (-0.059, -0.002) |
| Methacholine cation | Lognormal | -0.017 | (-0.034, -0.001) | 0.041 | 0.374 | 0.276 | (-0.033, -0.001) |
| 3-Cysteinylacetaminophen | Lognormal | 0.094 | (0.004, 0.184) | 0.042 | 0.376 | 0.314 | (0.006, 0.186) |
| Theobromine | Lognormal | -0.044 | (-0.087, -0.002) | 0.042 | 0.376 | 0.247 | (-0.088, -0.001) |
| D-Turanose | Lognormal | -0.029 | (-0.057, -0.001) | 0.044 | 0.381 | 0.288 | (-0.058, -0.001) |
| N-Acetylhistidine | Lognormal | -0.021 | (-0.041, 0.000) | 0.049 | 0.395 | 0.211 | (-0.041, 0.000) |
| Metformin | Lognormal | 0.030 | (0.000, 0.060) | 0.053 | 0.402 | 0.229 | (0.001, 0.061) |
| Isopropylamine | Lognormal | -0.011 | (-0.023, 0.000) | 0.055 | 0.402 | 0.232 | (-0.023, 0.000) |
| L-Cystine | Lognormal | -0.022 | (-0.045, 0.000) | 0.056 | 0.402 | 0.160 | (-0.044, 0.000) |
| Ranitidine | Lognormal | 0.041 | (-0.001, 0.082) | 0.057 | 0.405 | 0.205 | (-0.002, 0.083) |
| Betaine | Lognormal | -0.006 | (-0.012, 0.000) | 0.066 | 0.426 | 0.191 | (-0.012, 0.000) |
| 4,5,7-Trihydroxyisoflavone | Lognormal | 0.082 | (-0.008, 0.172) | 0.077 | 0.473 | 0.164 | (-0.006, 0.175) |
| p-Acetamidophenyl .beta.-D-glucuronide | Lognormal | 0.054 | (-0.006, 0.115) | 0.078 | 0.475 | 0.161 | (-0.008, 0.113) |
| Quetiapine | Lognormal | 0.024 | (-0.003, 0.050) | 0.079 | 0.478 | 0.165 | (-0.001, 0.050) |
| rac-4-Sulfoxypropranolol | Lognormal | 0.026 | (-0.003, 0.054) | 0.080 | 0.479 | 0.178 | (-0.001, 0.056) |
| 7-Methylguanosine | Lognormal | -0.012 | (-0.026, 0.001) | 0.082 | 0.479 | 0.165 | (-0.027, 0.001) |
| Stachydrine | Lognormal | -0.032 | (-0.068, 0.005) | 0.094 | 0.516 | 0.138 | (-0.068, 0.007) |
| Trazodone | Lognormal | 0.031 | (-0.005, 0.067) | 0.096 | 0.520 | 0.141 | (-0.005, 0.066) |
| Trigonelline | Lognormal | -0.033 | (-0.073, 0.006) | 0.098 | 0.522 | 0.122 | (-0.074, 0.006) |
| NEPSILON,NEPSILON,NEPSILON-TRIMETHYLLYSINE | Lognormal | -0.011 | (-0.024, 0.002) | 0.101 | 0.533 | 0.114 | (-0.023, 0.003) |
| (R)-Butyrylcarnitine | Lognormal | -0.016 | (-0.035, 0.003) | 0.111 | 0.557 | 0.115 | (-0.034, 0.004) |
| Lamotrigine; | Lognormal | 0.085 | (-0.020, 0.190) | 0.114 | 0.557 | 0.124 | (-0.016, 0.190) |
| Tri-2-ethylhexyl trimellitate | Lognormal | -0.035 | (-0.080, 0.010) | 0.126 | 0.579 | 0.095 | (-0.079, 0.010) |
| N-.alpha.-Acetyl-L-arginine | Lognormal | -0.011 | (-0.025, 0.003) | 0.134 | 0.605 | 0.113 | (-0.024, 0.004) |

**Supplementary Table S7.** Regression and Bayesian estimates comparing ME/CFS without sr-IBS vs. controls without sr-IBS.

| Metabolite | Regression Model | ME/CFS without sr-IBS vs. Controls without sr-IBS | | | | | |
|---|---|---|---|---|---|---|---|
| | | Estimated Coefficient | 95% CI | p-value | FDR | BayesFactor | 95% HDI |
| cis-10,11-Dihydroxy-10,11-dihydrocarbamazepine | Lognormal | -0.017 | (-0.038, 0.005) | 0.139 | 0.612 | 0.122 | (-0.039, 0.006) |
| 3,4-Dimethoxybenzaldehyde | Lognormal | 0.023 | (-0.007, 0.053) | 0.141 | 0.612 | 0.081 | (-0.007, 0.052) |
| 2,6-Diaminopimelic acid | Lognormal | 0.031 | (-0.011, 0.074) | 0.151 | 0.635 | 0.097 | (-0.015, 0.074) |
| Guanidine | Lognormal | 0.013 | (-0.005, 0.032) | 0.154 | 0.641 | 0.086 | (-0.005, 0.032) |
| 4'-Methyl-N-methylhexanophenone | Lognormal | -0.038 | (-0.091, 0.014) | 0.157 | 0.646 | 0.102 | (-0.090, 0.014) |
| Quetiapine sulfoxide | Lognormal | 0.024 | (-0.009, 0.058) | 0.161 | 0.648 | 0.088 | (-0.010, 0.058) |
| Kynurenine | Lognormal | -0.011 | (-0.025, 0.004) | 0.163 | 0.650 | 0.073 | (-0.025, 0.004) |
| Metoprolol acid | Lognormal | 0.036 | (-0.015, 0.087) | 0.169 | 0.659 | 0.079 | (-0.014, 0.090) |
| 3-Dehydrocarnitine | Lognormal | -0.010 | (-0.025, 0.004) | 0.171 | 0.659 | 0.075 | (-0.024, 0.005) |
| L-Tyrosine | Lognormal | -0.013 | (-0.031, 0.006) | 0.177 | 0.664 | 0.083 | (-0.031, 0.006) |
| Ergothioneine | Lognormal | -0.027 | (-0.066, 0.012) | 0.180 | 0.665 | 0.080 | (-0.063, 0.014) |
| Pantoprazole | Lognormal | 0.037 | (-0.017, 0.092) | 0.182 | 0.667 | 0.088 | (-0.017, 0.095) |
| Losartan | Lognormal | 0.024 | (-0.012, 0.061) | 0.193 | 0.678 | 0.075 | (-0.011, 0.063) |
| 7-Hydroxywarfarin | Lognormal | 0.021 | (-0.011, 0.054) | 0.200 | 0.687 | 0.076 | (-0.013, 0.052) |
| H-Pro-Hyp-OH | Lognormal | 0.014 | (-0.008, 0.036) | 0.203 | 0.687 | 0.078 | (-0.008, 0.036) |
| Octanoylcarnitine | Lognormal | -0.012 | (-0.031, 0.007) | 0.205 | 0.691 | 0.059 | (-0.032, 0.005) |
| Naproxen | Lognormal | 0.026 | (-0.014, 0.067) | 0.208 | 0.692 | 0.071 | (-0.017, 0.065) |
| 1-Stearoyl-2-hydroxy-sn-glycero-3-phosphoethanolamine | Lognormal | 0.022 | (-0.013, 0.056) | 0.217 | 0.698 | 0.072 | (-0.008, 0.059) |
| Bradykinin | Lognormal | 0.076 | (-0.044, 0.196) | 0.217 | 0.698 | 0.072 | (-0.040, 0.198) |
| Ticlopidine | Lognormal | -0.051 | (-0.132, 0.030) | 0.218 | 0.698 | 0.071 | (-0.127, 0.036) |
| 2-Indolinone | Lognormal | -0.013 | (-0.033, 0.007) | 0.220 | 0.701 | 0.075 | (-0.033, 0.007) |
| Coniferylaldehyde | Lognormal | 0.014 | (-0.008, 0.036) | 0.226 | 0.715 | 0.067 | (-0.007, 0.036) |
| Topiramate | Lognormal | 0.016 | (-0.010, 0.041) | 0.229 | 0.716 | 0.066 | (-0.008, 0.042) |
| alpha-Methylhistidine; | Lognormal | -0.029 | (-0.077, 0.018) | 0.229 | 0.716 | 0.072 | (-0.080, 0.016) |
| Omeprazole sulfone N-oxide | Lognormal | 0.039 | (-0.025, 0.103) | 0.229 | 0.716 | 0.075 | (-0.022, 0.107) |
| Histidine | Lognormal | -0.012 | (-0.031, 0.008) | 0.232 | 0.716 | 0.080 | (-0.031, 0.006) |
| Propionylcarnitine | Lognormal | -0.006 | (-0.015, 0.004) | 0.232 | 0.716 | 0.061 | (-0.015, 0.004) |
| Betaine aldehyde cation | Lognormal | 0.028 | (-0.018, 0.075) | 0.239 | 0.724 | 0.059 | (-0.021, 0.073) |
| Prazepam | Lognormal | 0.022 | (-0.015, 0.059) | 0.251 | 0.724 | 0.055 | (-0.017, 0.058) |
| Methylgallate | Lognormal | 0.045 | (-0.032, 0.122) | 0.251 | 0.724 | 0.073 | (-0.027, 0.129) |
| Albendazole | Lognormal | 0.011 | (-0.008, 0.031) | 0.252 | 0.724 | 0.061 | (-0.008, 0.031) |
| Heptadecasphing-4-enine | Lognormal | 0.063 | (-0.045, 0.171) | 0.254 | 0.724 | 0.058 | (-0.045, 0.171) |
| 1-Acetyl-3-piperidinamine | Lognormal | 0.009 | (-0.007, 0.026) | 0.261 | 0.726 | 0.060 | (-0.007, 0.026) |
| Indole-3-propionic acid | Lognormal | -0.018 | (-0.050, 0.014) | 0.263 | 0.726 | 0.064 | (-0.048, 0.014) |
| Acetazolamide | Lognormal | -0.028 | (-0.078, 0.021) | 0.264 | 0.726 | 0.066 | (-0.075, 0.024) |
| Phenylacetyl-L-glutamine | Lognormal | -0.021 | (-0.058, 0.016) | 0.267 | 0.729 | 0.061 | (-0.057, 0.016) |
| Tyrosine | Lognormal | -0.008 | (-0.022, 0.006) | 0.270 | 0.731 | 0.066 | (-0.022, 0.006) |
| Omeprazole sulfone | Lognormal | 0.034 | (-0.026, 0.094) | 0.271 | 0.731 | 0.066 | (-0.023, 0.098) |
| N.alpha.-Methyl-L-lysine | Lognormal | 0.021 | (-0.016, 0.059) | 0.271 | 0.731 | 0.064 | (-0.017, 0.059) |
| .epsilon.-Caprolactam | Lognormal | 0.009 | (-0.008, 0.027) | 0.284 | 0.739 | 0.038 | (-0.008, 0.026) |
| Hydroxybupropion | Lognormal | -0.015 | (-0.043, 0.013) | 0.296 | 0.752 | 0.060 | (-0.045, 0.013) |
| Diazepam | Lognormal | 0.034 | (-0.030, 0.099) | 0.300 | 0.753 | 0.054 | (-0.032, 0.097) |
| Milnacipran | Lognormal | 0.019 | (-0.017, 0.055) | 0.301 | 0.753 | 0.057 | (-0.019, 0.054) |
| DL-Indole-3-lactic acid | Lognormal | 0.006 | (-0.005, 0.017) | 0.301 | 0.753 | 0.058 | (-0.005, 0.017) |
| Metoprolol | Lognormal | 0.021 | (-0.019, 0.062) | 0.302 | 0.753 | 0.056 | (-0.020, 0.061) |
| 3,5-Dihydroxyphenylglycine | Lognormal | -0.015 | (-0.043, 0.013) | 0.307 | 0.757 | 0.049 | (-0.043, 0.014) |
| Testosterone | Lognormal | -0.017 | (-0.051, 0.016) | 0.311 | 0.757 | 0.055 | (-0.050, 0.017) |

**Supplementary Table S7.** Regression and Bayesian estimates comparing ME/CFS without sr-IBS vs. controls without sr-IBS.

| Metabolite | Regression Model | ME/CFS without sr-IBS vs. Controls without sr-IBS | | | | | |
|---|---|---|---|---|---|---|---|
| | | Estimated Coefficient | 95% CI | p-value | FDR | BayesFactor | 95% HDI |
| N-Acetyl-L-carnosine | Lognormal | -0.010 | (-0.030, 0.010) | 0.311 | 0.757 | 0.048 | (-0.030, 0.010) |
| L-Cysteine-glutathione disulfide | Lognormal | -0.015 | (-0.043, 0.014) | 0.314 | 0.757 | 0.039 | (-0.043, 0.013) |
| Betonicine | Lognormal | -0.034 | (-0.102, 0.033) | 0.319 | 0.757 | 0.054 | (-0.105, 0.031) |
| Cyclo(Leu-Pro) | Lognormal | 0.012 | (-0.012, 0.036) | 0.321 | 0.758 | 0.049 | (-0.011, 0.037) |
| 3-Hydroxypyridine | Lognormal | -0.034 | (-0.102, 0.034) | 0.331 | 0.764 | 0.046 | (-0.106, 0.032) |
| Ornithine | Lognormal | -0.008 | (-0.025, 0.008) | 0.334 | 0.764 | 0.042 | (-0.025, 0.008) |
| Homoarginine; | Lognormal | 0.009 | (-0.010, 0.028) | 0.339 | 0.765 | 0.045 | (-0.009, 0.028) |
| N-Methylhistidine | Lognormal | -0.016 | (-0.049, 0.017) | 0.352 | 0.784 | 0.053 | (-0.050, 0.018) |
| H-gamma-glutamyl-glutamine | Lognormal | 0.011 | (-0.013, 0.036) | 0.372 | 0.819 | 0.039 | (-0.014, 0.036) |
| Serotonin | Lognormal | 0.009 | (-0.011, 0.030) | 0.378 | 0.820 | 0.041 | (-0.010, 0.030) |
| L-Threonine | Lognormal | -0.005 | (-0.017, 0.006) | 0.389 | 0.825 | 0.055 | (-0.016, 0.007) |
| Trimethylamine-N-oxide | Lognormal | 0.012 | (-0.015, 0.038) | 0.389 | 0.825 | 0.047 | (-0.014, 0.038) |
| Matrine | Lognormal | 0.020 | (-0.025, 0.064) | 0.390 | 0.825 | 0.050 | (-0.030, 0.060) |
| (3-Carboxypropyl)trimethylammonium cation | Lognormal | -0.005 | (-0.017, 0.007) | 0.392 | 0.827 | 0.047 | (-0.017, 0.006) |
| Glaucine | Lognormal | 0.009 | (-0.012, 0.031) | 0.396 | 0.832 | 0.050 | (-0.014, 0.031) |
| Androstan-3-ol-17-one 3-glucuronide | Lognormal | 0.013 | (-0.018, 0.044) | 0.400 | 0.832 | 0.045 | (-0.017, 0.044) |
| Ala-Ile | Lognormal | 0.008 | (-0.011, 0.027) | 0.405 | 0.834 | 0.045 | (-0.012, 0.027) |
| Lauroyl-L-carnitine | Lognormal | -0.015 | (-0.051, 0.021) | 0.407 | 0.834 | 0.052 | (-0.050, 0.023) |
| N8-Acetylspermidine | Lognormal | -0.006 | (-0.019, 0.008) | 0.410 | 0.834 | 0.046 | (-0.019, 0.008) |
| Urea | Lognormal | -0.004 | (-0.013, 0.005) | 0.412 | 0.834 | 0.042 | (-0.013, 0.006) |
| 3-Hydroxyoleylcarnitine | Lognormal | 0.032 | (-0.045, 0.108) | 0.419 | 0.834 | 0.046 | (-0.046, 0.109) |
| Codeine-6-.beta.-D-glucuronide | Lognormal | 0.012 | (-0.017, 0.042) | 0.422 | 0.834 | 0.052 | (-0.017, 0.041) |
| Fexofenadine | Lognormal | -0.010 | (-0.035, 0.015) | 0.429 | 0.838 | 0.047 | (-0.035, 0.016) |
| 3-Pyridinemthanol | Lognormal | 0.010 | (-0.014, 0.034) | 0.431 | 0.838 | 0.049 | (-0.015, 0.034) |
| 4-Pyridoxic acid; | Lognormal | 0.017 | (-0.027, 0.062) | 0.444 | 0.854 | 0.048 | (-0.030, 0.060) |
| N-Methylproline | Lognormal | -0.010 | (-0.035, 0.015) | 0.449 | 0.854 | 0.048 | (-0.034, 0.016) |
| Tryptophan | Lognormal | 0.004 | (-0.006, 0.014) | 0.456 | 0.855 | 0.039 | (-0.007, 0.014) |
| D-Fructose | Lognormal | 0.003 | (-0.004, 0.010) | 0.459 | 0.855 | 0.037 | (-0.005, 0.010) |
| Pyrantel | Lognormal | -0.009 | (-0.033, 0.015) | 0.459 | 0.855 | 0.043 | (-0.031, 0.015) |
| Aminodiphenylmethane | Lognormal | 0.014 | (-0.023, 0.052) | 0.461 | 0.856 | 0.043 | (-0.021, 0.051) |
| Linoleoylcarnitine | Lognormal | -0.012 | (-0.046, 0.021) | 0.470 | 0.863 | 0.042 | (-0.045, 0.021) |
| Hexanoyl-L-carnitine | Lognormal | -0.008 | (-0.030, 0.014) | 0.475 | 0.863 | 0.041 | (-0.030, 0.014) |
| Penciclovir | Lognormal | 0.007 | (-0.013, 0.028) | 0.482 | 0.873 | 0.038 | (-0.013, 0.028) |
| 1-Methyl-L-histidine | Lognormal | -0.015 | (-0.056, 0.027) | 0.490 | 0.879 | 0.045 | (-0.056, 0.029) |
| Trimethoprim | Lognormal | -0.011 | (-0.041, 0.020) | 0.492 | 0.879 | 0.048 | (-0.043, 0.020) |
| Meprobamate | Lognormal | 0.011 | (-0.020, 0.041) | 0.501 | 0.890 | 0.045 | (-0.020, 0.041) |
| 6-Methoxynaphthaleneacetic acid | Lognormal | -0.004 | (-0.017, 0.008) | 0.507 | 0.894 | 0.044 | (-0.018, 0.009) |
| L-Leucine, methyl ester | Lognormal | 0.012 | (-0.023, 0.046) | 0.508 | 0.894 | 0.042 | (-0.021, 0.048) |
| Esomeprazole | Lognormal | 0.010 | (-0.020, 0.041) | 0.517 | 0.896 | 0.039 | (-0.022, 0.038) |
| Temazepam | Lognormal | 0.013 | (-0.028, 0.054) | 0.521 | 0.896 | 0.041 | (-0.026, 0.058) |
| 3-Amino-1-propanol | Lognormal | 0.006 | (-0.013, 0.026) | 0.523 | 0.896 | 0.045 | (-0.013, 0.026) |
| Methioninesulfoxide | Lognormal | 0.006 | (-0.013, 0.025) | 0.528 | 0.896 | 0.041 | (-0.013, 0.025) |
| Thr-Ile-Arg | Lognormal | -0.015 | (-0.061, 0.031) | 0.529 | 0.896 | 0.044 | (-0.064, 0.031) |
| Modafinil acid | Lognormal | 0.018 | (-0.039, 0.075) | 0.534 | 0.896 | 0.041 | (-0.037, 0.075) |
| Borrelidin | Lognormal | -0.015 | (-0.063, 0.033) | 0.537 | 0.896 | 0.043 | (-0.061, 0.035) |
| Irbesartan | Lognormal | 0.008 | (-0.017, 0.032) | 0.540 | 0.898 | 0.043 | (-0.015, 0.032) |
| Proline | Lognormal | 0.004 | (-0.009, 0.018) | 0.542 | 0.898 | 0.041 | (-0.010, 0.017) |

**Supplementary Table S7.** Regression and Bayesian estimates comparing ME/CFS without sr-IBS vs. controls without sr-IBS.

| Metabolite | Regression Model | ME/CFS without sr-IBS vs. Controls without sr-IBS | | | | | |
|---|---|---|---|---|---|---|---|
| | | Estimated Coefficient | 95% CI | p-value | FDR | BayesFactor | 95% HDI |
| 4-Fluoro-.alpha.-pyrrolidinobutiophenone | Lognormal | 0.007 | (-0.016, 0.030) | 0.558 | 0.908 | 0.041 | (-0.015, 0.030) |
| Glutamic acid | Lognormal | 0.005 | (-0.011, 0.020) | 0.560 | 0.909 | 0.035 | (-0.010, 0.019) |
| Carnitine | Lognormal | -0.002 | (-0.009, 0.005) | 0.563 | 0.909 | 0.036 | (-0.009, 0.005) |
| N.epsilon.-Methyl-L-lysine | Lognormal | 0.009 | (-0.022, 0.039) | 0.565 | 0.911 | 0.040 | (-0.022, 0.039) |
| Lauric acid diethanolamide | Lognormal | 0.012 | (-0.030, 0.055) | 0.572 | 0.914 | 0.041 | (-0.034, 0.052) |
| 1-Phenylpyrrolidine | Lognormal | 0.007 | (-0.018, 0.033) | 0.573 | 0.914 | 0.039 | (-0.019, 0.032) |
| Ezetimibe | Lognormal | 0.011 | (-0.027, 0.048) | 0.583 | 0.919 | 0.040 | (-0.031, 0.048) |
| Ethylenediaminetetraacetic acid | Lognormal | 0.022 | (-0.058, 0.102) | 0.594 | 0.928 | 0.037 | (-0.055, 0.107) |
| Dinor-12-oxophytodienoic acid | Lognormal | 0.012 | (-0.034, 0.057) | 0.609 | 0.933 | 0.042 | (-0.036, 0.055) |
| Montelukast-1,2-diol | Lognormal | 0.019 | (-0.057, 0.095) | 0.622 | 0.934 | 0.037 | (-0.055, 0.096) |
| 1-Methylnicotinamide | Lognormal | -0.005 | (-0.024, 0.015) | 0.624 | 0.934 | 0.039 | (-0.024, 0.015) |
| Ethiolat | Lognormal | -0.008 | (-0.042, 0.025) | 0.624 | 0.934 | 0.039 | (-0.041, 0.025) |
| trans-3'-Hydroxycotinine | Lognormal | 0.008 | (-0.027, 0.043) | 0.642 | 0.938 | 0.037 | (-0.027, 0.043) |
| Usnic acid | Lognormal | -0.009 | (-0.049, 0.030) | 0.644 | 0.938 | 0.041 | (-0.051, 0.030) |
| 1-Methyladenosine A | Lognormal | -0.002 | (-0.012, 0.007) | 0.644 | 0.938 | 0.035 | (-0.012, 0.008) |
| Telmisartan | Lognormal | 0.007 | (-0.023, 0.038) | 0.645 | 0.938 | 0.033 | (-0.022, 0.039) |
| Ser-Tyr-Lys | Lognormal | 0.014 | (-0.044, 0.071) | 0.646 | 0.938 | 0.037 | (-0.047, 0.072) |
| 5'-S-Methylthioadenosine | Lognormal | 0.018 | (-0.058, 0.094) | 0.649 | 0.938 | 0.032 | (-0.056, 0.093) |
| Decanoyl-L-carnitine | Lognormal | 0.007 | (-0.023, 0.037) | 0.653 | 0.938 | 0.038 | (-0.025, 0.038) |
| Biliverden | Lognormal | -0.004 | (-0.024, 0.015) | 0.657 | 0.938 | 0.038 | (-0.023, 0.016) |
| threo-Dihydrobupropion | Lognormal | 0.005 | (-0.018, 0.029) | 0.658 | 0.938 | 0.035 | (-0.018, 0.029) |
| N-(3-(Aminomethyl)benzyl)acetamidine | Lognormal | -0.008 | (-0.043, 0.027) | 0.660 | 0.938 | 0.031 | (-0.044, 0.025) |
| 1-Monostearin | Lognormal | -0.008 | (-0.043, 0.028) | 0.664 | 0.938 | 0.037 | (-0.042, 0.028) |
| Nudifloramide | Lognormal | -0.004 | (-0.020, 0.013) | 0.665 | 0.938 | 0.039 | (-0.022, 0.013) |
| Arginine | Lognormal | 0.003 | (-0.010, 0.016) | 0.669 | 0.938 | 0.032 | (-0.010, 0.016) |
| D-.alpha.-Cyclohexylglycine | Lognormal | -0.006 | (-0.034, 0.022) | 0.676 | 0.938 | 0.036 | (-0.032, 0.023) |
| N-Acetyl-D-norleucine | Lognormal | 0.005 | (-0.018, 0.028) | 0.677 | 0.938 | 0.022 | (-0.019, 0.026) |
| Meloxicam | Lognormal | 0.024 | (-0.089, 0.136) | 0.682 | 0.939 | 0.040 | (-0.091, 0.142) |
| Carbamazepine | Lognormal | -0.011 | (-0.064, 0.042) | 0.684 | 0.939 | 0.037 | (-0.066, 0.044) |
| Acetyl-DL-carnitine | Lognormal | 0.003 | (-0.012, 0.018) | 0.689 | 0.942 | 0.038 | (-0.012, 0.019) |
| Cotinine N-.beta.-D-glucuronide | Lognormal | 0.014 | (-0.054, 0.082) | 0.689 | 0.942 | 0.035 | (-0.058, 0.082) |
| Ranitidine-S-oxide | Lognormal | 0.012 | (-0.047, 0.071) | 0.693 | 0.945 | 0.035 | (-0.047, 0.072) |
| 2,2-Bishydroxymethyl]-2,2',2''-nitrilotriethanol | Lognormal | 0.006 | (-0.023, 0.035) | 0.694 | 0.945 | 0.037 | (-0.023, 0.034) |
| D-Pyroglutamic acid | Lognormal | -0.001 | (-0.007, 0.004) | 0.701 | 0.945 | 0.038 | (-0.007, 0.004) |
| Dexpanthenol | Lognormal | -0.006 | (-0.038, 0.025) | 0.701 | 0.945 | 0.036 | (-0.038, 0.026) |
| Lysine | Lognormal | -0.002 | (-0.013, 0.009) | 0.704 | 0.946 | 0.025 | (-0.014, 0.009) |
| Tapentadol-.beta.-D-glucuronide | Lognormal | -0.012 | (-0.072, 0.049) | 0.707 | 0.946 | 0.039 | (-0.074, 0.048) |
| (2R)-3-Hydroxyisovaleroylcarnitine | Lognormal | -0.003 | (-0.019, 0.013) | 0.715 | 0.948 | 0.035 | (-0.019, 0.014) |
| 1,2-Dimethylimidazole | Lognormal | 0.004 | (-0.016, 0.023) | 0.718 | 0.948 | 0.035 | (-0.016, 0.024) |
| N.epsilon.-Acetyl-L-lysine | Lognormal | -0.003 | (-0.017, 0.012) | 0.719 | 0.948 | 0.037 | (-0.018, 0.012) |
| D-erythro-Sphingosine-1-phosphate | Lognormal | -0.004 | (-0.028, 0.019) | 0.727 | 0.948 | 0.037 | (-0.028, 0.018) |
| 6-Hydroxyflavone | Lognormal | 0.007 | (-0.033, 0.047) | 0.727 | 0.948 | 0.034 | (-0.035, 0.045) |
| Toradol | Lognormal | 0.007 | (-0.033, 0.047) | 0.729 | 0.948 | 0.035 | (-0.031, 0.046) |
| .beta.-Phenyl-.gamma.-aminobutyric acid | Lognormal | -0.003 | (-0.023, 0.016) | 0.731 | 0.948 | 0.033 | (-0.023, 0.017) |
| Sulfamethoxazole | Lognormal | -0.015 | (-0.103, 0.072) | 0.731 | 0.948 | 0.035 | (-0.101, 0.073) |
| Levocetirizine; | Lognormal | 0.012 | (-0.059, 0.084) | 0.736 | 0.952 | 0.039 | (-0.058, 0.085) |
| Norleucine | Lognormal | -0.002 | (-0.013, 0.009) | 0.741 | 0.954 | 0.037 | (-0.013, 0.010) |

**Supplementary Table S7.** Regression and Bayesian estimates comparing ME/CFS without sr-IBS vs. controls without sr-IBS.

| Metabolite | Regression Model | ME/CFS without sr-IBS vs. Controls without sr-IBS | | | | | |
|---|---|---|---|---|---|---|---|
| | | Estimated Coefficient | 95% CI | p-value | FDR | BayesFactor | 95% HDI |
| 3-Methylxanthine | Lognormal | -0.007 | (-0.048, 0.035) | 0.754 | 0.955 | 0.034 | (-0.051, 0.034) |
| Atenolol | Lognormal | 0.006 | (-0.030, 0.041) | 0.756 | 0.955 | 0.036 | (-0.028, 0.042) |
| Phenylacetylglutamine | Lognormal | -0.004 | (-0.033, 0.024) | 0.766 | 0.955 | 0.033 | (-0.034, 0.023) |
| 1-Acetyl-4-piperidinamine | Lognormal | 0.006 | (-0.033, 0.044) | 0.769 | 0.955 | 0.035 | (-0.034, 0.044) |
| Urocanic acid; | Lognormal | -0.003 | (-0.021, 0.016) | 0.770 | 0.955 | 0.035 | (-0.021, 0.016) |
| Phenylalanine | Lognormal | -0.001 | (-0.009, 0.007) | 0.771 | 0.955 | 0.032 | (-0.010, 0.006) |
| Modafinil | Lognormal | 0.007 | (-0.038, 0.051) | 0.773 | 0.955 | 0.036 | (-0.041, 0.049) |
| 4-Aminomethylcyclohexanecarboxylic acid; | Lognormal | 0.004 | (-0.023, 0.031) | 0.779 | 0.958 | 0.031 | (-0.024, 0.032) |
| Theanine; | Lognormal | -0.004 | (-0.036, 0.027) | 0.783 | 0.958 | 0.032 | (-0.034, 0.029) |
| Triptolide | Lognormal | -0.003 | (-0.024, 0.018) | 0.783 | 0.958 | 0.036 | (-0.024, 0.019) |
| 3-(1-Pyrazolyl)-alanine | Lognormal | 0.010 | (-0.059, 0.078) | 0.784 | 0.958 | 0.035 | (-0.054, 0.076) |
| Pyridoxal | Lognormal | 0.003 | (-0.019, 0.025) | 0.790 | 0.958 | 0.035 | (-0.019, 0.025) |
| Glycodeoxycholic acid | Lognormal | 0.005 | (-0.034, 0.044) | 0.790 | 0.958 | 0.034 | (-0.036, 0.042) |
| 2-Amino-1-phenylethanol | Lognormal | -0.001 | (-0.009, 0.007) | 0.791 | 0.958 | 0.035 | (-0.009, 0.008) |
| Creatinine | Lognormal | -0.001 | (-0.007, 0.005) | 0.792 | 0.958 | 0.031 | (-0.007, 0.005) |
| 3-Hydroxybutyrylcarnitine | Lognormal | -0.004 | (-0.033, 0.026) | 0.798 | 0.960 | 0.036 | (-0.033, 0.024) |
| 1-Oleoyl-2-acetyl-sn-glycerol | Lognormal | -0.005 | (-0.041, 0.032) | 0.801 | 0.961 | 0.036 | (-0.041, 0.032) |
| Glutamine | Lognormal | -0.001 | (-0.006, 0.005) | 0.801 | 0.961 | 0.034 | (-0.007, 0.005) |
| 7.alpha.-Hydroxy-3-oxo-4-cholestenoic acid | Lognormal | -0.003 | (-0.023, 0.018) | 0.806 | 0.963 | 0.033 | (-0.024, 0.017) |
| Ondansetron | Lognormal | 0.002 | (-0.014, 0.018) | 0.811 | 0.963 | 0.031 | (-0.015, 0.018) |
| Pyridoxine; | Lognormal | 0.004 | (-0.029, 0.037) | 0.811 | 0.963 | 0.036 | (-0.030, 0.036) |
| Isoleucine | Lognormal | 0.001 | (-0.010, 0.012) | 0.818 | 0.968 | 0.034 | (-0.009, 0.012) |
| L-Carnitine | Lognormal | -0.001 | (-0.012, 0.010) | 0.824 | 0.972 | 0.034 | (-0.012, 0.010) |
| Pantothenic acid | Lognormal | 0.002 | (-0.020, 0.024) | 0.830 | 0.976 | 0.038 | (-0.018, 0.026) |
| Atorvastatin | Lognormal | 0.002 | (-0.017, 0.021) | 0.860 | 0.981 | 0.035 | (-0.018, 0.021) |
| R-(-)-O-Desmethylvenlafaxine | Lognormal | -0.004 | (-0.048, 0.040) | 0.861 | 0.981 | 0.036 | (-0.049, 0.038) |
| Diphenhydramine | Lognormal | -0.002 | (-0.024, 0.020) | 0.862 | 0.981 | 0.034 | (-0.025, 0.019) |
| Trileptal | Lognormal | 0.002 | (-0.023, 0.028) | 0.862 | 0.981 | 0.036 | (-0.025, 0.027) |
| Avobenzone | Lognormal | 0.002 | (-0.022, 0.026) | 0.862 | 0.981 | 0.032 | (-0.022, 0.026) |
| Gly-Pro-Arg | Lognormal | 0.006 | (-0.059, 0.070) | 0.862 | 0.981 | 0.038 | (-0.057, 0.067) |
| Oleoyl-L-carnitine | Lognormal | -0.003 | (-0.038, 0.032) | 0.862 | 0.981 | 0.036 | (-0.038, 0.032) |
| Tauroursodeoxycholic acid | Lognormal | 0.003 | (-0.030, 0.036) | 0.865 | 0.981 | 0.034 | (-0.031, 0.036) |
| 2,2',2''-Nitrilotriethanol | Lognormal | 0.002 | (-0.024, 0.028) | 0.870 | 0.984 | 0.036 | (-0.023, 0.029) |
| Methionine | Lognormal | 0.001 | (-0.009, 0.011) | 0.872 | 0.984 | 0.034 | (-0.010, 0.011) |
| 2-Hydroxyibuprofen | Lognormal | -0.003 | (-0.036, 0.030) | 0.878 | 0.984 | 0.037 | (-0.037, 0.029) |
| 1-Stearoyl-2-arachidonoyl-sn-glycero-3-phospho-(1'-myo-inositol) | Lognormal | 0.007 | (-0.082, 0.095) | 0.882 | 0.984 | 0.035 | (-0.081, 0.098) |
| Palmitamide | Lognormal | 0.003 | (-0.034, 0.040) | 0.884 | 0.984 | 0.037 | (-0.033, 0.040) |
| Lansoprazole | Lognormal | -0.007 | (-0.104, 0.090) | 0.886 | 0.984 | 0.035 | (-0.109, 0.089) |
| 1-Palmitoyl-2-hydroxy-sn-glycero-3-phosphoethanolamine | Lognormal | 0.003 | (-0.035, 0.040) | 0.889 | 0.984 | 0.029 | (-0.034, 0.039) |
| 1-Oleoyl-sn-glycero-3-phosphoethanolamine | Lognormal | -0.002 | (-0.039, 0.034) | 0.894 | 0.984 | 0.032 | (-0.038, 0.034) |
| Isopentenyladenine | Lognormal | -0.001 | (-0.010, 0.009) | 0.894 | 0.984 | 0.035 | (-0.010, 0.009) |
| 1-Stearoyl-2-arachidonoyl-sn-glycero-3-phosphoserine | Lognormal | -0.005 | (-0.087, 0.077) | 0.900 | 0.984 | 0.028 | (-0.084, 0.082) |
| Creatine | Lognormal | 0.001 | (-0.014, 0.016) | 0.905 | 0.984 | 0.036 | (-0.013, 0.017) |
| 3-Aminoquinoline | Lognormal | 0.002 | (-0.040, 0.045) | 0.910 | 0.985 | 0.027 | (-0.040, 0.043) |
| 5'-S-Methyl-5'-thioadenosine | Lognormal | 0.003 | (-0.063, 0.069) | 0.925 | 0.990 | 0.028 | (-0.060, 0.073) |
| Hypoxanthine | Lognormal | 0.001 | (-0.016, 0.018) | 0.932 | 0.990 | 0.032 | (-0.017, 0.017) |
| Glycocholic acid | Lognormal | 0.001 | (-0.034, 0.037) | 0.935 | 0.991 | 0.032 | (-0.033, 0.037) |

**Supplementary Table S7.** Regression and Bayesian estimates comparing ME/CFS without sr-IBS vs. controls without sr-IBS.

| Metabolite | Regression Model | ME/CFS without sr-IBS vs. Controls without sr-IBS | | | | | |
|---|---|---|---|---|---|---|---|
| | | Estimated Coefficient | 95% CI | p-value | FDR | BayesFactor | 95% HDI |
| Ile-Glu-Arg | Lognormal | -0.002 | (-0.068, 0.063) | 0.941 | 0.991 | 0.035 | (-0.066, 0.064) |
| Moxonidine | Lognormal | 0.001 | (-0.025, 0.027) | 0.952 | 0.997 | 0.035 | (-0.026, 0.027) |
| N-Acetylalanine | Lognormal | 0.002 | (-0.060, 0.064) | 0.956 | 0.998 | 0.033 | (-0.057, 0.062) |
| Adenosine | Lognormal | 0.001 | (-0.028, 0.030) | 0.965 | 0.998 | 0.035 | (-0.029, 0.031) |
| Pipecolic acid | Lognormal | 0.001 | (-0.040, 0.041) | 0.968 | 0.998 | 0.032 | (-0.044, 0.041) |
| Scopoletin | Lognormal | 0.001 | (-0.074, 0.076) | 0.972 | 0.998 | 0.025 | (-0.073, 0.078) |
| Benthiavalicarb-isopropyl | Lognormal | 0.000 | (-0.036, 0.035) | 0.982 | 0.998 | 0.033 | (-0.036, 0.034) |
| Mefenorex | Lognormal | 0.000 | (-0.039, 0.038) | 0.983 | 0.998 | 0.029 | (-0.039, 0.039) |
| Ethyldiethanolamine | Lognormal | 0.000 | (-0.027, 0.027) | 0.991 | 0.998 | 0.035 | (-0.028, 0.026) |
| Benzophenone-3 | Lognormal | 0.000 | (-0.059, 0.059) | 0.999 | 0.999 | 0.032 | (-0.057, 0.059) |
| **Complex Lipids (CL)** | | | | | | | |
| PC (p-34:2)/PC (o-34:3) - ESI(+) | Lognormal | **-0.019** | **(-0.028, -0.010)** | **0.000** | **0.076** | **16.961** | **(-0.029, -0.010)** |
| PC (36:2) | Lognormal | **-0.008** | **(-0.012, -0.004)** | **0.000** | **0.076** | **8.626** | **(-0.012, -0.004)** |
| PC (p-34:1)/PC (o-34:2) | Lognormal | **-0.022** | **(-0.033, -0.010)** | **0.000** | **0.076** | **30.007** | **(-0.034, -0.010)** |
| PE (p-36:2)/PE (o-36:3) - ESI(+) | Lognormal | **-0.029** | **(-0.045, -0.013)** | **0.000** | **0.081** | **6.915** | **(-0.044, -0.013)** |
| PC (p-36:4)/PC (o-36:5) - ESI(-) | Lognormal | -0.020 | (-0.032, -0.008) | 0.001 | 0.218 | 6.200 | (-0.031, -0.008) |
| PC (p-36:1)/PC (o-36:2) | Lognormal | -0.052 | (-0.084, -0.020) | 0.002 | 0.218 | 4.636 | (-0.085, -0.022) |
| PC (36:4) A - ESI(+) | Lognormal | -0.017 | (-0.028, -0.007) | 0.002 | 0.218 | 3.025 | (-0.027, -0.007) |
| PC (p-34:2)/PC (o-34:3) - ESI(-) | Lognormal | -0.019 | (-0.030, -0.007) | 0.002 | 0.248 | 3.502 | (-0.031, -0.007) |
| PC 34:4e | Lognormal | -0.023 | (-0.037, -0.008) | 0.003 | 0.248 | 3.559 | (-0.037, -0.009) |
| PE (p-34:2)/PE (o-34:3) | Lognormal | -0.036 | (-0.060, -0.012) | 0.004 | 0.310 | 2.620 | (-0.060, -0.013) |
| PC (36:4) A - ESI(-) | Lognormal | -0.018 | (-0.030, -0.006) | 0.005 | 0.310 | 2.130 | (-0.030, -0.006) |
| PC (p-34:1)/PC (o-34:2) A | Lognormal | -0.025 | (-0.042, -0.008) | 0.005 | 0.310 | 1.458 | (-0.042, -0.008) |
| PC (32:2) - ESI(-) | Lognormal | -0.024 | (-0.040, -0.007) | 0.006 | 0.310 | 1.525 | (-0.041, -0.007) |
| PC (34:4) - ESI(-) | Lognormal | -0.030 | (-0.053, -0.008) | 0.009 | 0.310 | 1.080 | (-0.055, -0.008) |
| TG (62:4) | Lognormal | -0.035 | (-0.061, -0.009) | 0.010 | 0.310 | 0.863 | (-0.061, -0.007) |
| PE (p-36:4)/PE (o-36:5) - ESI(+) | Lognormal | -0.018 | (-0.031, -0.004) | 0.010 | 0.310 | 0.778 | (-0.031, -0.005) |
| LPC (18:3) | Lognormal | -0.024 | (-0.043, -0.006) | 0.010 | 0.310 | 0.976 | (-0.042, -0.006) |
| SM (d43:1) - ESI(-) | Lognormal | -0.032 | (-0.056, -0.008) | 0.011 | 0.310 | 0.889 | (-0.058, -0.008) |
| LPC (14:0) - ESI(+) | Lognormal | -0.021 | (-0.036, -0.005) | 0.011 | 0.310 | 0.909 | (-0.036, -0.005) |
| PE (p-36:4)/PE (o-36:5) - ESI(-) | Lognormal | -0.020 | (-0.035, -0.005) | 0.012 | 0.310 | 0.614 | (-0.035, -0.004) |
| PE (p-36:2)/PE (o-36:3) - ESI(-) | Lognormal | -0.026 | (-0.046, -0.006) | 0.012 | 0.310 | 0.805 | (-0.046, -0.005) |
| PC (p-36:2)/PC (o-36:3) | Lognormal | -0.016 | (-0.029, -0.004) | 0.012 | 0.310 | 0.903 | (-0.029, -0.004) |
| PC (p-36:3)/PC (o-36:4) - ESI(-) | Lognormal | -0.013 | (-0.024, -0.003) | 0.012 | 0.310 | 0.736 | (-0.023, -0.002) |
| LPC (18:0) B - ESI(-) | Lognormal | -0.014 | (-0.025, -0.003) | 0.013 | 0.310 | 0.830 | (-0.025, -0.003) |
| LPC (18:0) A - ESI(-) | Lognormal | -0.019 | (-0.034, -0.004) | 0.013 | 0.310 | 0.880 | (-0.035, -0.005) |
| SM (d40:3) | Lognormal | -0.026 | (-0.046, -0.006) | 0.013 | 0.310 | 1.103 | (-0.045, -0.007) |
| SM (d39:1) - ESI(-) | Lognormal | -0.015 | (-0.027, -0.003) | 0.013 | 0.310 | 0.723 | (-0.027, -0.003) |
| PC (p-36:4)/PC (o-36:5) - ESI(+) | Lognormal | -0.010 | (-0.018, -0.002) | 0.014 | 0.310 | 0.750 | (-0.018, -0.002) |
| PE (p-38:5)/PE (o-38:6) - ESI(-) | Lognormal | -0.019 | (-0.034, -0.004) | 0.014 | 0.310 | 0.951 | (-0.034, -0.005) |
| PC (p-36:3)/PC (o-36:4) - ESI(+) | Lognormal | -0.011 | (-0.019, -0.002) | 0.015 | 0.310 | 0.697 | (-0.019, -0.002) |
| SM (d40:1) - ESI(-) | Lognormal | -0.011 | (-0.020, -0.002) | 0.015 | 0.310 | 0.745 | (-0.019, -0.002) |
| Ceramide (d42:1) - ESI (+) | Lognormal | -0.009 | (-0.016, -0.002) | 0.015 | 0.310 | 0.947 | (-0.015, -0.002) |
| LPC (18:2) - ESI(-) | Lognormal | -0.017 | (-0.030, -0.003) | 0.016 | 0.310 | 0.720 | (-0.030, -0.003) |
| SM (d32:1) - ESI(-) | Lognormal | -0.013 | (-0.024, -0.003) | 0.016 | 0.310 | 0.795 | (-0.023, -0.002) |
| PE (p-38:5)/PE (o-38:6) - ESI(+) | Lognormal | -0.014 | (-0.026, -0.003) | 0.016 | 0.310 | 0.660 | (-0.026, -0.003) |
| CE (18:3) | Lognormal | -0.018 | (-0.033, -0.003) | 0.017 | 0.310 | 0.690 | (-0.032, -0.003) |

**Supplementary Table S7.** Regression and Bayesian estimates comparing ME/CFS without sr-IBS vs. controls without sr-IBS.

| Metabolite | Regression Model | ME/CFS without sr-IBS vs. Controls without sr-IBS | | | | | |
|---|---|---|---|---|---|---|---|
| | | Estimated Coefficient | 95% CI | p-value | FDR | BayesFactor | 95% HDI |
| Ceramide (d42:2) B - ESI (+) | Lognormal | -0.011 | (-0.020, -0.002) | 0.018 | 0.311 | 0.514 | (-0.020, -0.002) |
| PC (p-38:4)/PC (o-38:5) A | Lognormal | -0.011 | (-0.021, -0.002) | 0.019 | 0.327 | 0.453 | (-0.021, -0.002) |
| Ceramide (d42:1) - ESI(-) | Lognormal | -0.011 | (-0.019, -0.002) | 0.020 | 0.327 | 0.496 | (-0.019, -0.002) |
| PE (p-38:4)/PE (o-38:5) | Lognormal | -0.016 | (-0.030, -0.003) | 0.020 | 0.327 | 0.567 | (-0.029, -0.003) |
| LPC (16:0) - ESI(-) | Lognormal | -0.010 | (-0.018, -0.002) | 0.021 | 0.327 | 0.530 | (-0.018, -0.002) |
| Ceramide (d41:1) - ESI(-) | Lognormal | -0.013 | (-0.023, -0.002) | 0.021 | 0.327 | 0.480 | (-0.024, -0.002) |
| PC (34:2) - ESI(+) | Lognormal | -0.004 | (-0.007, -0.001) | 0.021 | 0.327 | 0.553 | (-0.007, -0.001) |
| LPC (18:2) - ESI(+) | Lognormal | -0.013 | (-0.024, -0.002) | 0.022 | 0.327 | 0.460 | (-0.024, -0.002) |
| PC (34:2) - ESI(-) | Lognormal | -0.008 | (-0.015, -0.001) | 0.022 | 0.327 | 0.578 | (-0.015, -0.001) |
| PE (p-36:1)/PE (o-36:2) - ESI(+) | Lognormal | -0.020 | (-0.038, -0.003) | 0.023 | 0.336 | 0.558 | (-0.038, -0.003) |
| CE (20:2) | Lognormal | -0.022 | (-0.042, -0.003) | 0.024 | 0.339 | 0.437 | (-0.043, -0.004) |
| LPC (20:2) - ESI(+) | Lognormal | -0.037 | (-0.069, -0.005) | 0.026 | 0.351 | 0.455 | (-0.067, -0.004) |
| PC (34:3) | Lognormal | -0.014 | (-0.027, -0.002) | 0.026 | 0.351 | 0.383 | (-0.028, -0.002) |
| PC (p-38:4)/PC (o-38:5) B | Lognormal | -0.012 | (-0.022, -0.001) | 0.028 | 0.352 | 0.413 | (-0.022, -0.001) |
| Ceramide (d40:1) | Lognormal | -0.010 | (-0.018, -0.001) | 0.028 | 0.352 | 0.391 | (-0.018, -0.001) |
| TG (50:0) | Lognormal | -0.024 | (-0.046, -0.003) | 0.029 | 0.352 | 0.431 | (-0.045, -0.003) |
| LPE (20:4) - ESI(-) | Lognormal | -0.019 | (-0.036, -0.002) | 0.030 | 0.352 | 0.323 | (-0.037, -0.002) |
| TG (53:0) | Lognormal | -0.018 | (-0.033, -0.002) | 0.030 | 0.352 | 0.403 | (-0.033, -0.002) |
| LPC (20:3) - ESI(-) | Lognormal | -0.021 | (-0.041, -0.002) | 0.030 | 0.352 | 0.362 | (-0.041, -0.002) |
| PC (p-38:3)/PC (o-38:4) - ESI(-) | Lognormal | -0.013 | (-0.025, -0.001) | 0.030 | 0.352 | 0.353 | (-0.025, -0.002) |
| TG (56:6) | Lognormal | 0.009 | (0.001, 0.017) | 0.030 | 0.352 | 0.364 | (0.001, 0.018) |
| SM (d36:3) - ESI(-) | Lognormal | -0.019 | (-0.035, -0.002) | 0.031 | 0.352 | 0.355 | (-0.036, -0.001) |
| PC (38:4) A - ESI(-) | Lognormal | -0.010 | (-0.019, -0.001) | 0.032 | 0.352 | 0.376 | (-0.018, -0.001) |
| Ceramide (d39:1) | Lognormal | -0.019 | (-0.035, -0.002) | 0.032 | 0.352 | 0.394 | (-0.035, -0.002) |
| PC (36:5) A | Lognormal | -0.037 | (-0.072, -0.003) | 0.034 | 0.362 | 0.322 | (-0.071, -0.001) |
| CE (18:2) | Lognormal | -0.008 | (-0.015, -0.001) | 0.034 | 0.362 | 0.331 | (-0.015, 0.000) |
| PC (32:2) - ESI(+) | Lognormal | -0.016 | (-0.031, -0.001) | 0.035 | 0.368 | 0.321 | (-0.031, -0.001) |
| PC (34:3) B | Lognormal | -0.011 | (-0.020, -0.001) | 0.036 | 0.369 | 0.316 | (-0.020, -0.001) |
| PC (34:3) C | Lognormal | -0.015 | (-0.029, -0.001) | 0.037 | 0.369 | 0.386 | (-0.029, -0.001) |
| PC (36:5)A | Lognormal | -0.020 | (-0.038, -0.001) | 0.038 | 0.373 | 0.310 | (-0.038, -0.001) |
| TG (55:6) | Lognormal | 0.018 | (0.001, 0.035) | 0.039 | 0.373 | 0.259 | (-0.001, 0.035) |
| SM (d30:1) - ESI(-) | Lognormal | -0.028 | (-0.055, -0.002) | 0.039 | 0.373 | 0.285 | (-0.056, -0.002) |
| PC (p-38:3)/PC (o-38:4) A - ESI(+) | Lognormal | -0.010 | (-0.020, -0.001) | 0.039 | 0.373 | 0.269 | (-0.020, -0.001) |
| SM (d33:1) - ESI(-) | Lognormal | -0.011 | (-0.022, -0.001) | 0.040 | 0.373 | 0.322 | (-0.023, -0.001) |
| LPE (18:2) - ESI(-) | Lognormal | -0.022 | (-0.044, -0.001) | 0.044 | 0.381 | 0.298 | (-0.045, -0.002) |
| Ceramide (d43:1) | Lognormal | -0.029 | (-0.057, -0.001) | 0.044 | 0.381 | 0.241 | (-0.056, 0.001) |
| SM (d40:2) B - ESI(-) | Lognormal | -0.010 | (-0.020, 0.000) | 0.045 | 0.382 | 0.318 | (-0.020, -0.001) |
| PC (33:2) - ESI(-) | Lognormal | -0.016 | (-0.032, 0.000) | 0.045 | 0.382 | 0.253 | (-0.032, 0.000) |
| PI (38:4)/PI (18:0-20:4) | Lognormal | -0.009 | (-0.018, 0.000) | 0.046 | 0.383 | 0.302 | (-0.018, 0.000) |
| SM (d42:1) - ESI(-) | Lognormal | -0.009 | (-0.017, 0.000) | 0.049 | 0.395 | 0.253 | (-0.017, 0.000) |
| LPE (18:0) | Lognormal | -0.013 | (-0.027, 0.000) | 0.049 | 0.395 | 0.278 | (-0.026, 0.000) |
| PC (37:2) - ESI(+) | Lognormal | -0.009 | (-0.018, 0.000) | 0.050 | 0.397 | 0.250 | (-0.019, -0.001) |
| LPC (14:0) - ESI(-) | Lognormal | -0.026 | (-0.052, 0.000) | 0.052 | 0.402 | 0.211 | (-0.050, 0.001) |
| SM (d34:2) - ESI(-) | Lognormal | -0.008 | (-0.015, 0.000) | 0.052 | 0.402 | 0.269 | (-0.015, 0.000) |
| TG (58:8) | Lognormal | 0.018 | (0.000, 0.036) | 0.053 | 0.402 | 0.236 | (0.001, 0.036) |
| TG (58:9) | Lognormal | 0.017 | (0.000, 0.034) | 0.054 | 0.402 | 0.203 | (0.000, 0.034) |
| PC (36:3) A - ESI(+) | Lognormal | -0.007 | (-0.014, 0.000) | 0.055 | 0.402 | 0.218 | (-0.014, 0.000) |

**Supplementary Table S7.** Regression and Bayesian estimates comparing ME/CFS without sr-IBS vs. controls without sr-IBS.

| Metabolite | Regression Model | ME/CFS without sr-IBS vs. Controls without sr-IBS | | | | | |
|---|---|---|---|---|---|---|---|
| | | Estimated Coefficient | 95% CI | p-value | FDR | BayesFactor | 95% HDI |
| PC (38:6) A - ESI(+) | Lognormal | -0.008 | (-0.016, 0.000) | 0.056 | 0.402 | 0.223 | (-0.016, 0.000) |
| PE (p-34:1)/PE (o-34:2) - ESI(+) | Lognormal | -0.015 | (-0.029, 0.000) | 0.057 | 0.405 | 0.205 | (-0.030, 0.001) |
| PE (p-36:1)/PE (o-36:2) - ESI(-) | Lognormal | -0.026 | (-0.052, 0.001) | 0.058 | 0.405 | 0.219 | (-0.051, 0.000) |
| SM (d40:1) - ESI(+) | Lognormal | -0.006 | (-0.012, 0.000) | 0.059 | 0.405 | 0.249 | (-0.011, 0.000) |
| PC (p-38:3)/PC (o-38:4) B - ESI(+) | Lognormal | -0.010 | (-0.021, 0.000) | 0.059 | 0.405 | 0.197 | (-0.021, 0.000) |
| TG (56:7) B | Lognormal | 0.016 | (-0.001, 0.033) | 0.059 | 0.405 | 0.210 | (-0.001, 0.033) |
| PC (o-34:0) | Lognormal | -0.014 | (-0.028, 0.000) | 0.059 | 0.405 | 0.235 | (-0.028, -0.001) |
| AC (10:1) | Lognormal | -0.020 | (-0.040, 0.001) | 0.060 | 0.407 | 0.237 | (-0.041, -0.001) |
| LPE (18:2) - ESI(+) | Lognormal | -0.017 | (-0.034, 0.001) | 0.061 | 0.408 | 0.233 | (-0.035, -0.001) |
| PC (36:4) B - ESI(-) | Lognormal | -0.006 | (-0.013, 0.000) | 0.062 | 0.414 | 0.213 | (-0.014, 0.000) |
| SM (d40:2) B - ESI(+) | Lognormal | -0.006 | (-0.013, 0.000) | 0.064 | 0.424 | 0.188 | (-0.012, 0.001) |
| PC (34:4) - ESI(+) | Lognormal | -0.016 | (-0.033, 0.001) | 0.065 | 0.424 | 0.197 | (-0.033, 0.001) |
| FA (16:1) (palmitoleic acid) | Lognormal | 0.014 | (-0.001, 0.028) | 0.066 | 0.426 | 0.187 | (-0.001, 0.027) |
| PC (40:8) - ESI(-) | Lognormal | -0.014 | (-0.030, 0.001) | 0.070 | 0.447 | 0.171 | (-0.030, 0.001) |
| TG (60:11) | Lognormal | 0.026 | (-0.002, 0.055) | 0.071 | 0.449 | 0.159 | (-0.004, 0.055) |
| PC (38:3) - ESI(-) | Lognormal | -0.011 | (-0.023, 0.001) | 0.073 | 0.460 | 0.185 | (-0.022, 0.001) |
| LPC (p-16:0)/LPC (o-16:1) | Lognormal | -0.012 | (-0.025, 0.001) | 0.075 | 0.471 | 0.173 | (-0.025, 0.001) |
| SM (d39:1) - ESI(+) | Lognormal | -0.009 | (-0.019, 0.001) | 0.076 | 0.472 | 0.163 | (-0.019, 0.001) |
| LPC (16:1) - ESI(-) | Lognormal | -0.016 | (-0.033, 0.002) | 0.077 | 0.473 | 0.175 | (-0.033, 0.001) |
| TG (54:7) B | Lognormal | 0.015 | (-0.002, 0.031) | 0.081 | 0.479 | 0.176 | (-0.001, 0.030) |
| PE (p-40:4)/PE (o-40:5) A | Lognormal | -0.019 | (-0.040, 0.002) | 0.082 | 0.479 | 0.111 | (-0.041, 0.003) |
| PC (p-32:1)/PC (o-32:2) | Lognormal | -0.009 | (-0.020, 0.001) | 0.083 | 0.480 | 0.152 | (-0.020, 0.001) |
| SM (d30:1) - ESI(+) | Lognormal | -0.019 | (-0.040, 0.002) | 0.084 | 0.486 | 0.157 | (-0.040, 0.002) |
| PE (36:2) | Lognormal | -0.018 | (-0.039, 0.002) | 0.085 | 0.488 | 0.163 | (-0.038, 0.002) |
| PC (37:5) | Lognormal | 0.014 | (-0.002, 0.030) | 0.087 | 0.494 | 0.165 | (-0.001, 0.031) |
| GlcCer (d42:2) - ESI(+) | Lognormal | 0.011 | (-0.002, 0.024) | 0.087 | 0.494 | 0.143 | (-0.001, 0.024) |
| PE (p-40:4)/PE (o-40:5) B | Lognormal | -0.011 | (-0.024, 0.002) | 0.092 | 0.515 | 0.115 | (-0.025, 0.001) |
| LPC (18:0) - ESI(+) | Lognormal | -0.008 | (-0.017, 0.001) | 0.093 | 0.516 | 0.162 | (-0.017, 0.001) |
| DG (38:6) | Lognormal | 0.014 | (-0.002, 0.031) | 0.094 | 0.516 | 0.138 | (-0.001, 0.031) |
| SM (d34:1) - ESI(-) | Lognormal | -0.006 | (-0.012, 0.001) | 0.095 | 0.516 | 0.128 | (-0.012, 0.001) |
| PC (p-38:4)/PC (o-38:5) B | Lognormal | -0.015 | (-0.032, 0.002) | 0.095 | 0.516 | 0.148 | (-0.032, 0.002) |
| TG (54:6) C | Lognormal | 0.009 | (-0.002, 0.021) | 0.098 | 0.522 | 0.124 | (-0.002, 0.020) |
| LPC (20:2) - ESI(-) | Lognormal | -0.012 | (-0.026, 0.002) | 0.102 | 0.537 | 0.123 | (-0.027, 0.002) |
| LPC (20:3) - ESI(+) | Lognormal | -0.011 | (-0.023, 0.002) | 0.103 | 0.537 | 0.117 | (-0.024, 0.002) |
| SM (d42:1) - ESI(+) | Lognormal | -0.006 | (-0.013, 0.001) | 0.104 | 0.542 | 0.138 | (-0.013, 0.001) |
| PC (35:1) - ESI(+) | Lognormal | 0.006 | (-0.001, 0.013) | 0.107 | 0.546 | 0.129 | (-0.001, 0.013) |
| FA (24:0) (lignoceric acid) | Lognormal | -0.009 | (-0.021, 0.002) | 0.107 | 0.546 | 0.126 | (-0.021, 0.002) |
| LPC (18:1) - ESI(-) | Lognormal | -0.010 | (-0.022, 0.002) | 0.111 | 0.557 | 0.123 | (-0.022, 0.002) |
| PC (p-38:4)/PC (o-38:5) A | Lognormal | -0.006 | (-0.013, 0.001) | 0.112 | 0.557 | 0.120 | (-0.013, 0.001) |
| SM (d32:1) - ESI(+) | Lognormal | -0.008 | (-0.019, 0.002) | 0.113 | 0.557 | 0.134 | (-0.019, 0.001) |
| Ceramide (d33:1) | Lognormal | -0.010 | (-0.023, 0.002) | 0.114 | 0.557 | 0.121 | (-0.023, 0.002) |
| SM (d38:2) - ESI(-) | Lognormal | -0.010 | (-0.022, 0.002) | 0.116 | 0.560 | 0.115 | (-0.022, 0.002) |
| TG (56:8) B | Lognormal | 0.015 | (-0.004, 0.035) | 0.116 | 0.560 | 0.109 | (-0.004, 0.035) |
| PC (34:0) - ESI(-) | Lognormal | -0.008 | (-0.017, 0.002) | 0.119 | 0.560 | 0.126 | (-0.017, 0.002) |
| PC (p-38:5)/PC (o-38:6) B | Lognormal | -0.007 | (-0.015, 0.002) | 0.119 | 0.560 | 0.114 | (-0.015, 0.002) |
| Ceramide (d38:1) - ESI(+) | Lognormal | -0.007 | (-0.016, 0.002) | 0.120 | 0.560 | 0.108 | (-0.016, 0.002) |
| PC (p-40:1)/PC (o-40:2) | Lognormal | 0.015 | (-0.004, 0.033) | 0.120 | 0.560 | 0.101 | (-0.004, 0.033) |

**Supplementary Table S7.** Regression and Bayesian estimates comparing ME/CFS without sr-IBS vs. controls without sr-IBS.

| Metabolite | Regression Model | ME/CFS without sr-IBS vs. Controls without sr-IBS | | | | | |
|---|---|---|---|---|---|---|---|
| | | Estimated Coefficient | 95% CI | p-value | FDR | BayesFactor | 95% HDI |
| PC (p-40:3)/PC (o-40:4) | Lognormal | -0.013 | (-0.030, 0.003) | 0.121 | 0.560 | 0.126 | (-0.031, 0.002) |
| SM (d38:1) | Lognormal | -0.005 | (-0.011, 0.001) | 0.123 | 0.568 | 0.124 | (-0.012, 0.001) |
| Ceramide (d41:1) - ESI (+) | Lognormal | -0.008 | (-0.018, 0.002) | 0.130 | 0.594 | 0.105 | (-0.018, 0.002) |
| TG (48:4) A | Lognormal | -0.020 | (-0.046, 0.006) | 0.133 | 0.605 | 0.100 | (-0.046, 0.006) |
| PC 40:5e | Lognormal | -0.007 | (-0.016, 0.002) | 0.134 | 0.605 | 0.100 | (-0.016, 0.002) |
| SM (d36:0) - ESI(+) | Lognormal | 0.012 | (-0.004, 0.027) | 0.138 | 0.612 | 0.109 | (-0.004, 0.027) |
| AC (14:2) | Lognormal | -0.018 | (-0.041, 0.006) | 0.139 | 0.612 | 0.106 | (-0.040, 0.006) |
| PC (33:2) - ESI(+) | Lognormal | -0.007 | (-0.016, 0.002) | 0.140 | 0.612 | 0.084 | (-0.016, 0.002) |
| PC (38:6) B - ESI(+) | Lognormal | 0.004 | (-0.001, 0.010) | 0.140 | 0.612 | 0.100 | (-0.002, 0.010) |
| PC (36:5) D | Lognormal | 0.013 | (-0.004, 0.031) | 0.143 | 0.619 | 0.089 | (-0.004, 0.031) |
| LPE (16:0) | Lognormal | -0.016 | (-0.038, 0.006) | 0.145 | 0.622 | 0.102 | (-0.038, 0.005) |
| TG (48:0) | Lognormal | -0.016 | (-0.037, 0.005) | 0.148 | 0.628 | 0.096 | (-0.037, 0.004) |
| SM (d44:2) | Lognormal | 0.011 | (-0.004, 0.026) | 0.151 | 0.635 | 0.091 | (-0.005, 0.025) |
| TG (56:5) A | Lognormal | 0.010 | (-0.004, 0.023) | 0.154 | 0.641 | 0.094 | (-0.003, 0.023) |
| SM (d43:2) - ESI(-) | Lognormal | -0.016 | (-0.038, 0.006) | 0.157 | 0.646 | 0.094 | (-0.038, 0.006) |
| PC (36:3) B - ESI(-) | Lognormal | -0.006 | (-0.015, 0.002) | 0.159 | 0.648 | 0.091 | (-0.015, 0.002) |
| TG (56:5) B | Lognormal | 0.009 | (-0.003, 0.021) | 0.160 | 0.648 | 0.091 | (-0.003, 0.021) |
| PC (37:6) | Lognormal | 0.012 | (-0.005, 0.029) | 0.162 | 0.650 | 0.089 | (-0.004, 0.029) |
| Ceramide (d42:2) B - ESI(-) | Lognormal | -0.010 | (-0.024, 0.004) | 0.164 | 0.652 | 0.093 | (-0.024, 0.004) |
| FA (20:5) (eicosapentaenoic acid) | Lognormal | 0.018 | (-0.007, 0.043) | 0.168 | 0.659 | 0.082 | (-0.006, 0.043) |
| PC (40:5) B - ESI(-) | Lognormal | -0.016 | (-0.038, 0.007) | 0.169 | 0.659 | 0.081 | (-0.038, 0.007) |
| PC (36:3) A - ESI(-) | Lognormal | -0.007 | (-0.017, 0.003) | 0.171 | 0.659 | 0.097 | (-0.017, 0.003) |
| PE (p-34:1)/PE (o-34:2) - ESI(-) | Lognormal | -0.012 | (-0.028, 0.005) | 0.173 | 0.661 | 0.089 | (-0.028, 0.006) |
| Ceramide (d38:1) - ESI(-) | Lognormal | -0.009 | (-0.022, 0.004) | 0.176 | 0.664 | 0.088 | (-0.021, 0.005) |
| LPC (o-16:0) | Lognormal | -0.010 | (-0.024, 0.004) | 0.176 | 0.664 | 0.080 | (-0.023, 0.005) |
| TG (46:0) | Lognormal | -0.024 | (-0.059, 0.011) | 0.178 | 0.664 | 0.083 | (-0.061, 0.008) |
| PE (p-38:6)/PE (o-38:7) | Lognormal | -0.013 | (-0.031, 0.006) | 0.179 | 0.664 | 0.083 | (-0.030, 0.006) |
| FA (18:1) (oleic acid) | Lognormal | 0.006 | (-0.003, 0.015) | 0.180 | 0.665 | 0.092 | (-0.003, 0.014) |
| TG (60:6) | Lognormal | 0.015 | (-0.007, 0.036) | 0.185 | 0.675 | 0.080 | (-0.009, 0.035) |
| TG (58:6) | Lognormal | 0.008 | (-0.004, 0.019) | 0.187 | 0.678 | 0.073 | (-0.004, 0.019) |
| TG (49:2) | Lognormal | 0.014 | (-0.007, 0.035) | 0.188 | 0.678 | 0.085 | (-0.006, 0.035) |
| PC (40:7) A - ESI(+) | Lognormal | -0.007 | (-0.018, 0.003) | 0.189 | 0.678 | 0.081 | (-0.018, 0.004) |
| SM (d42:2) | Lognormal | 0.005 | (-0.002, 0.012) | 0.189 | 0.678 | 0.070 | (-0.002, 0.012) |
| PE (36:3) | Lognormal | -0.020 | (-0.049, 0.010) | 0.190 | 0.678 | 0.089 | (-0.049, 0.011) |
| PC (40:5) B - ESI(+) | Lognormal | -0.009 | (-0.023, 0.004) | 0.192 | 0.678 | 0.079 | (-0.023, 0.005) |
| PC (38:3) - ESI(+) | Lognormal | -0.006 | (-0.015, 0.003) | 0.193 | 0.678 | 0.080 | (-0.015, 0.003) |
| GlcCer (d14:1(4E)/20:0(2OH)) | Lognormal | -0.013 | (-0.032, 0.006) | 0.193 | 0.678 | 0.078 | (-0.033, 0.005) |
| PE (38:2) | Lognormal | -0.012 | (-0.031, 0.006) | 0.194 | 0.678 | 0.081 | (-0.031, 0.006) |
| PC (42:10) | Lognormal | 0.013 | (-0.007, 0.033) | 0.196 | 0.682 | 0.069 | (-0.006, 0.032) |
| TG (46:2) | Lognormal | -0.020 | (-0.051, 0.010) | 0.197 | 0.684 | 0.082 | (-0.051, 0.012) |
| LPC (20:4) | Lognormal | -0.007 | (-0.018, 0.004) | 0.200 | 0.687 | 0.073 | (-0.019, 0.004) |
| LPC (22:4) | Lognormal | -0.016 | (-0.041, 0.009) | 0.201 | 0.687 | 0.079 | (-0.042, 0.008) |
| PC (p-42:5)/PC (o-42:6) A | Lognormal | -0.008 | (-0.021, 0.004) | 0.202 | 0.687 | 0.078 | (-0.021, 0.005) |
| PC (30:0) | Lognormal | -0.009 | (-0.023, 0.005) | 0.202 | 0.687 | 0.069 | (-0.024, 0.004) |
| AC (12:0) | Lognormal | -0.013 | (-0.033, 0.007) | 0.206 | 0.691 | 0.080 | (-0.034, 0.006) |
| PC (37:2) - ESI(-) | Lognormal | -0.014 | (-0.036, 0.008) | 0.207 | 0.692 | 0.083 | (-0.037, 0.008) |
| FA (14:1) (physeteric acid) | Lognormal | 0.012 | (-0.007, 0.030) | 0.208 | 0.692 | 0.074 | (-0.006, 0.031) |

**Supplementary Table S7.** Regression and Bayesian estimates comparing ME/CFS without sr-IBS vs. controls without sr-IBS.

| Metabolite | Regression Model | ME/CFS without sr-IBS vs. Controls without sr-IBS | | | | | |
|---|---|---|---|---|---|---|---|
| | | Estimated Coefficient | 95% CI | p-value | FDR | BayesFactor | 95% HDI |
| TG (56:8) A | Lognormal | 0.009 | (-0.005, 0.023) | 0.210 | 0.696 | 0.071 | (-0.006, 0.022) |
| CE (20:3) | Lognormal | -0.007 | (-0.019, 0.004) | 0.211 | 0.696 | 0.074 | (-0.019, 0.005) |
| PE (p-38:3)/PE (o-38:4) | Lognormal | -0.016 | (-0.041, 0.009) | 0.212 | 0.698 | 0.072 | (-0.042, 0.008) |
| Ceramide (d44:1) | Lognormal | -0.017 | (-0.043, 0.010) | 0.215 | 0.698 | 0.070 | (-0.042, 0.011) |
| TG (60:12) | Lognormal | 0.024 | (-0.014, 0.061) | 0.215 | 0.698 | 0.068 | (-0.013, 0.059) |
| SM (d41:2) B - ESI(+) | Lognormal | -0.005 | (-0.014, 0.003) | 0.217 | 0.698 | 0.074 | (-0.014, 0.003) |
| TG (56:5) C | Lognormal | 0.007 | (-0.004, 0.019) | 0.219 | 0.698 | 0.074 | (-0.004, 0.020) |
| LPC (16:0) - ESI(+) | Lognormal | -0.003 | (-0.009, 0.002) | 0.224 | 0.710 | 0.071 | (-0.009, 0.002) |
| PE (p-40:4)/PE (o-40:5) | Lognormal | -0.010 | (-0.026, 0.006) | 0.230 | 0.716 | 0.052 | (-0.027, 0.005) |
| Ceramide (d32:1) | Lognormal | -0.009 | (-0.025, 0.006) | 0.232 | 0.716 | 0.067 | (-0.024, 0.006) |
| AC (12:1) | Lognormal | -0.012 | (-0.031, 0.008) | 0.237 | 0.724 | 0.068 | (-0.032, 0.007) |
| CE (20:5) | Lognormal | 0.013 | (-0.009, 0.035) | 0.238 | 0.724 | 0.060 | (-0.008, 0.035) |
| TG (58:2) | Lognormal | -0.030 | (-0.080, 0.020) | 0.243 | 0.724 | 0.075 | (-0.079, 0.021) |
| TG (54:5) B | Lognormal | 0.006 | (-0.004, 0.016) | 0.243 | 0.724 | 0.065 | (-0.005, 0.015) |
| PE (34:1) | Lognormal | -0.018 | (-0.049, 0.013) | 0.247 | 0.724 | 0.063 | (-0.048, 0.014) |
| PC (38:2) | Lognormal | -0.008 | (-0.021, 0.005) | 0.248 | 0.724 | 0.069 | (-0.021, 0.005) |
| PC (40:5) A - ESI(+) | Lognormal | 0.005 | (-0.003, 0.013) | 0.249 | 0.724 | 0.073 | (-0.004, 0.014) |
| Lactosylceramide (d18:1/24:1(15Z)) | Lognormal | 0.009 | (-0.006, 0.025) | 0.250 | 0.724 | 0.059 | (-0.006, 0.025) |
| GlcCer (d38:1) | Lognormal | -0.008 | (-0.021, 0.005) | 0.251 | 0.724 | 0.069 | (-0.021, 0.005) |
| PC (38:5) B - ESI(+) | Lognormal | 0.006 | (-0.004, 0.017) | 0.252 | 0.724 | 0.063 | (-0.005, 0.017) |
| SM (d41:3) | Lognormal | -0.005 | (-0.014, 0.004) | 0.254 | 0.724 | 0.068 | (-0.014, 0.004) |
| TG (46:4) A | Lognormal | -0.022 | (-0.058, 0.015) | 0.255 | 0.724 | 0.065 | (-0.057, 0.017) |
| TG (54:6) B | Lognormal | -0.020 | (-0.054, 0.014) | 0.255 | 0.724 | 0.070 | (-0.053, 0.015) |
| TG (54:7) A | Lognormal | -0.012 | (-0.034, 0.009) | 0.261 | 0.726 | 0.063 | (-0.034, 0.009) |
| PC (40:6)B | Lognormal | 0.005 | (-0.004, 0.015) | 0.261 | 0.726 | 0.061 | (-0.004, 0.014) |
| AC (10:0) | Lognormal | -0.017 | (-0.047, 0.013) | 0.262 | 0.726 | 0.067 | (-0.048, 0.013) |
| SM (d32:2) - ESI(-) | Lognormal | -0.012 | (-0.033, 0.009) | 0.262 | 0.726 | 0.070 | (-0.033, 0.009) |
| TG (48:5) | Lognormal | -0.017 | (-0.047, 0.013) | 0.264 | 0.726 | 0.063 | (-0.047, 0.012) |
| PE (38:4) | Lognormal | -0.006 | (-0.017, 0.005) | 0.264 | 0.726 | 0.047 | (-0.017, 0.005) |
| AC (18:0) | Lognormal | -0.007 | (-0.020, 0.005) | 0.265 | 0.726 | 0.055 | (-0.019, 0.005) |
| DG (38:5) | Lognormal | 0.007 | (-0.005, 0.019) | 0.271 | 0.731 | 0.055 | (-0.005, 0.019) |
| PC (35:3) | Lognormal | -0.004 | (-0.012, 0.003) | 0.272 | 0.731 | 0.060 | (-0.012, 0.003) |
| TG (44:0) | Lognormal | -0.019 | (-0.054, 0.015) | 0.273 | 0.731 | 0.062 | (-0.055, 0.013) |
| LPE (20:4) - ESI(+) | Lognormal | -0.009 | (-0.024, 0.007) | 0.273 | 0.731 | 0.057 | (-0.025, 0.006) |
| PC (p-44:4)/PC (o-44:5) - ESI(-) | Lognormal | -0.008 | (-0.022, 0.006) | 0.276 | 0.731 | 0.060 | (-0.023, 0.006) |
| FA (17:0) (margaric acid) | Lognormal | 0.003 | (-0.003, 0.009) | 0.276 | 0.731 | 0.059 | (-0.003, 0.010) |
| PC (p-42:3)/PC (o-42:4) | Lognormal | -0.007 | (-0.020, 0.006) | 0.276 | 0.731 | 0.055 | (-0.019, 0.006) |
| TG (48:3) | Lognormal | -0.012 | (-0.033, 0.010) | 0.278 | 0.733 | 0.061 | (-0.034, 0.009) |
| PC (p-42:5)/PC (o-42:6) | Lognormal | -0.015 | (-0.043, 0.012) | 0.279 | 0.733 | 0.066 | (-0.042, 0.014) |
| PC (40:4) - ESI(-) | Lognormal | -0.010 | (-0.029, 0.008) | 0.279 | 0.733 | 0.063 | (-0.030, 0.007) |
| TG (48:4) B | Lognormal | -0.021 | (-0.058, 0.017) | 0.280 | 0.733 | 0.068 | (-0.062, 0.016) |
| AC (18:2) | Lognormal | -0.008 | (-0.023, 0.007) | 0.282 | 0.737 | 0.059 | (-0.022, 0.006) |
| FA (22:0) (behenic acid) | Lognormal | -0.007 | (-0.019, 0.006) | 0.289 | 0.748 | 0.051 | (-0.020, 0.006) |
| PC (34:1) | Lognormal | 0.002 | (-0.002, 0.006) | 0.291 | 0.750 | 0.057 | (-0.002, 0.006) |
| CE (14:0) | Lognormal | -0.019 | (-0.055, 0.016) | 0.291 | 0.750 | 0.057 | (-0.055, 0.018) |
| SM (d41:1) - ESI(+) | Lognormal | -0.004 | (-0.012, 0.003) | 0.294 | 0.752 | 0.060 | (-0.012, 0.003) |
| GlcCer (d42:2) - ESI(-) | Lognormal | 0.007 | (-0.006, 0.019) | 0.296 | 0.752 | 0.053 | (-0.005, 0.019) |

**Supplementary Table S7.** Regression and Bayesian estimates comparing ME/CFS without sr-IBS vs. controls without sr-IBS.

| Metabolite | Regression Model | ME/CFS without sr-IBS vs. Controls without sr-IBS | | | | | |
|---|---|---|---|---|---|---|---|
| | | Estimated Coefficient | 95% CI | p-value | FDR | BayesFactor | 95% HDI |
| SM (d32:0) - ESI(-) | Lognormal | -0.031 | (-0.089, 0.027) | 0.296 | 0.752 | 0.066 | (-0.090, 0.027) |
| PC (36:3) B - ESI(+) | Lognormal | -0.005 | (-0.014, 0.004) | 0.296 | 0.752 | 0.059 | (-0.013, 0.004) |
| TG (54:1) | Lognormal | -0.012 | (-0.035, 0.011) | 0.301 | 0.753 | 0.054 | (-0.035, 0.010) |
| PC (34:3) A | Lognormal | -0.005 | (-0.015, 0.005) | 0.302 | 0.753 | 0.055 | (-0.015, 0.005) |
| TG (42:0) | Lognormal | -0.019 | (-0.056, 0.017) | 0.305 | 0.757 | 0.060 | (-0.054, 0.019) |
| SM (d42:3) - ESI(-) | Lognormal | -0.005 | (-0.013, 0.004) | 0.306 | 0.757 | 0.054 | (-0.013, 0.004) |
| PE (34:2) - ESI(-) | Lognormal | -0.014 | (-0.042, 0.013) | 0.306 | 0.757 | 0.060 | (-0.041, 0.013) |
| TG (50:4) | Lognormal | -0.008 | (-0.022, 0.007) | 0.310 | 0.757 | 0.060 | (-0.023, 0.006) |
| PC (39:6) | Lognormal | 0.008 | (-0.007, 0.023) | 0.311 | 0.757 | 0.050 | (-0.008, 0.023) |
| FA (20:1) (eicosenoic acid) | Lognormal | 0.006 | (-0.005, 0.016) | 0.314 | 0.757 | 0.056 | (-0.005, 0.017) |
| FA (20:3) (homo-gamma-linolenic acid) | Lognormal | 0.005 | (-0.005, 0.014) | 0.317 | 0.757 | 0.051 | (-0.005, 0.014) |
| DG (36:2) | Lognormal | 0.006 | (-0.005, 0.016) | 0.318 | 0.757 | 0.051 | (-0.006, 0.016) |
| PC (p-32:0)/PC (o-32:1) - ESI(-) | Lognormal | -0.008 | (-0.023, 0.007) | 0.323 | 0.760 | 0.055 | (-0.024, 0.008) |
| CE (22:2) | Lognormal | 0.018 | (-0.018, 0.054) | 0.325 | 0.760 | 0.058 | (-0.019, 0.053) |
| Ceramide (d42:0) | Lognormal | -0.008 | (-0.023, 0.008) | 0.325 | 0.760 | 0.049 | (-0.024, 0.006) |
| TG (50:5) | Lognormal | -0.010 | (-0.031, 0.010) | 0.331 | 0.764 | 0.053 | (-0.032, 0.010) |
| Cholesterol | Lognormal | -0.003 | (-0.009, 0.003) | 0.333 | 0.764 | 0.051 | (-0.009, 0.003) |
| Ceramide (d34:1) - ESI(+) | Lognormal | 0.004 | (-0.004, 0.013) | 0.334 | 0.764 | 0.054 | (-0.004, 0.013) |
| FA (10:0) (capric acid) | Lognormal | 0.012 | (-0.013, 0.037) | 0.335 | 0.764 | 0.051 | (-0.012, 0.038) |
| SM (d34:0) - ESI(-) | Lognormal | -0.007 | (-0.022, 0.007) | 0.336 | 0.764 | 0.054 | (-0.023, 0.007) |
| SM (d36:1) - ESI(-) | Lognormal | -0.004 | (-0.014, 0.005) | 0.336 | 0.764 | 0.057 | (-0.013, 0.004) |
| TG (42:2) | Lognormal | -0.041 | (-0.125, 0.043) | 0.338 | 0.764 | 0.049 | (-0.128, 0.043) |
| TG (51:5) | Lognormal | -0.009 | (-0.028, 0.010) | 0.338 | 0.764 | 0.051 | (-0.027, 0.010) |
| DG (32:0) | Lognormal | 0.008 | (-0.009, 0.025) | 0.340 | 0.765 | 0.056 | (-0.009, 0.026) |
| PC (40:7) B - ESI(+) | Lognormal | 0.004 | (-0.005, 0.014) | 0.343 | 0.769 | 0.051 | (-0.005, 0.013) |
| FA (12:0) (lauric acid) | Lognormal | 0.008 | (-0.009, 0.024) | 0.361 | 0.802 | 0.051 | (-0.009, 0.023) |
| PC (28:0) | Lognormal | -0.018 | (-0.058, 0.021) | 0.364 | 0.805 | 0.051 | (-0.059, 0.021) |
| PC (38:5) A | Lognormal | 0.003 | (-0.003, 0.008) | 0.376 | 0.820 | 0.052 | (-0.003, 0.009) |
| TG (46:1) | Lognormal | -0.015 | (-0.048, 0.018) | 0.376 | 0.820 | 0.047 | (-0.047, 0.016) |
| TG (42:1) | Lognormal | -0.031 | (-0.098, 0.037) | 0.376 | 0.820 | 0.049 | (-0.098, 0.038) |
| TG (44:1) | Lognormal | -0.020 | (-0.063, 0.024) | 0.379 | 0.820 | 0.052 | (-0.066, 0.024) |
| Ceramide (d42:2) A - ESI (+) | Lognormal | 0.004 | (-0.004, 0.012) | 0.379 | 0.820 | 0.046 | (-0.005, 0.011) |
| TG (46:3) A | Lognormal | -0.019 | (-0.061, 0.023) | 0.380 | 0.820 | 0.049 | (-0.058, 0.025) |
| SM (d43:1) - ESI(+) | Lognormal | -0.007 | (-0.022, 0.009) | 0.381 | 0.820 | 0.053 | (-0.022, 0.009) |
| LPC (16:1) - ESI(+) | Lognormal | -0.005 | (-0.016, 0.006) | 0.381 | 0.820 | 0.052 | (-0.017, 0.006) |
| PC (p-36:1)/PC (o-36:2) B | Lognormal | -0.006 | (-0.021, 0.008) | 0.389 | 0.825 | 0.048 | (-0.022, 0.008) |
| TG (50:1) | Lognormal | -0.008 | (-0.025, 0.010) | 0.390 | 0.825 | 0.049 | (-0.026, 0.009) |
| TG (42:3) | Lognormal | -0.017 | (-0.056, 0.022) | 0.390 | 0.825 | 0.050 | (-0.055, 0.024) |
| TG (52:2) | Lognormal | 0.003 | (-0.004, 0.010) | 0.393 | 0.827 | 0.045 | (-0.004, 0.010) |
| LPE (22:6) | Lognormal | -0.008 | (-0.025, 0.010) | 0.397 | 0.832 | 0.049 | (-0.025, 0.011) |
| FA (14:0) (myristic acid) | Lognormal | 0.004 | (-0.005, 0.012) | 0.398 | 0.832 | 0.048 | (-0.005, 0.012) |
| SM (d40:2) A - ESI(+) | Lognormal | -0.005 | (-0.017, 0.007) | 0.400 | 0.832 | 0.048 | (-0.017, 0.007) |
| PC (38:4) C - ESI(+) | Lognormal | -0.002 | (-0.008, 0.003) | 0.403 | 0.834 | 0.048 | (-0.009, 0.003) |
| AC (8:0) | Lognormal | -0.013 | (-0.043, 0.017) | 0.404 | 0.834 | 0.048 | (-0.044, 0.015) |
| SM (d39:2) | Lognormal | -0.004 | (-0.013, 0.005) | 0.407 | 0.834 | 0.040 | (-0.012, 0.005) |
| PC (p-32:0)/PC (o-32:1) - ESI(+) | Lognormal | -0.003 | (-0.011, 0.005) | 0.410 | 0.834 | 0.043 | (-0.011, 0.004) |
| PC (38:4) B - ESI(+) | Lognormal | -0.004 | (-0.014, 0.006) | 0.413 | 0.834 | 0.045 | (-0.014, 0.006) |

**Supplementary Table S7.** Regression and Bayesian estimates comparing ME/CFS without sr-IBS vs. controls without sr-IBS.

| Metabolite | Regression Model | ME/CFS without sr-IBS vs. Controls without sr-IBS | | | | | |
|---|---|---|---|---|---|---|---|
| | | Estimated Coefficient | 95% CI | p-value | FDR | BayesFactor | 95% HDI |
| PC (p-38:6)/PC (o-38:7) | Lognormal | -0.005 | (-0.019, 0.008) | 0.414 | 0.834 | 0.043 | (-0.019, 0.008) |
| PE (p-40:5)/PE (o-40:6) | Lognormal | -0.004 | (-0.014, 0.006) | 0.415 | 0.834 | 0.037 | (-0.014, 0.005) |
| TG (40:1) | Lognormal | -0.017 | (-0.057, 0.024) | 0.418 | 0.834 | 0.049 | (-0.057, 0.025) |
| LPC (20:5) | Lognormal | 0.011 | (-0.016, 0.039) | 0.420 | 0.834 | 0.047 | (-0.017, 0.039) |
| PC (33:1) - ESI(+) | Lognormal | 0.004 | (-0.006, 0.014) | 0.420 | 0.834 | 0.046 | (-0.005, 0.015) |
| TG (51:2) | Lognormal | 0.006 | (-0.008, 0.020) | 0.425 | 0.837 | 0.051 | (-0.009, 0.020) |
| PE (36:4) - ESI(-) | Lognormal | -0.011 | (-0.038, 0.016) | 0.425 | 0.837 | 0.045 | (-0.038, 0.017) |
| PC (36:5) C | Lognormal | 0.009 | (-0.013, 0.030) | 0.427 | 0.838 | 0.042 | (-0.012, 0.030) |
| TG (64:3) | Lognormal | -0.009 | (-0.031, 0.013) | 0.428 | 0.838 | 0.044 | (-0.029, 0.015) |
| GlcCer (d41:1) | Lognormal | -0.005 | (-0.018, 0.008) | 0.431 | 0.838 | 0.047 | (-0.018, 0.007) |
| TG (56:9) | Lognormal | 0.008 | (-0.012, 0.027) | 0.432 | 0.838 | 0.048 | (-0.011, 0.028) |
| SM (d40:2) A - ESI(-) | Lognormal | -0.008 | (-0.027, 0.012) | 0.448 | 0.854 | 0.045 | (-0.027, 0.013) |
| PC (32:0) - ESI(+) | Lognormal | 0.002 | (-0.003, 0.008) | 0.448 | 0.854 | 0.041 | (-0.003, 0.008) |
| LPC (20:0) | Lognormal | -0.007 | (-0.023, 0.010) | 0.448 | 0.854 | 0.045 | (-0.023, 0.011) |
| SM (d32:2) - ESI(+) | Lognormal | -0.004 | (-0.016, 0.007) | 0.455 | 0.855 | 0.040 | (-0.016, 0.008) |
| PC (35:2) A | Lognormal | 0.003 | (-0.005, 0.012) | 0.456 | 0.855 | 0.046 | (-0.005, 0.012) |
| TG (52:6) | Lognormal | -0.007 | (-0.026, 0.011) | 0.456 | 0.855 | 0.047 | (-0.026, 0.012) |
| LPC (20:1) - ESI(-) | Lognormal | -0.006 | (-0.023, 0.010) | 0.456 | 0.855 | 0.045 | (-0.022, 0.011) |
| PC (40:4) - ESI(+) | Lognormal | -0.005 | (-0.017, 0.007) | 0.458 | 0.855 | 0.045 | (-0.017, 0.008) |
| PC (38:7) | Lognormal | 0.006 | (-0.010, 0.022) | 0.459 | 0.855 | 0.042 | (-0.010, 0.022) |
| CE (18:0) | Lognormal | 0.008 | (-0.013, 0.029) | 0.465 | 0.861 | 0.047 | (-0.013, 0.028) |
| PG (34:0)/PG (17:0/17:0) | Lognormal | -0.007 | (-0.025, 0.011) | 0.466 | 0.861 | 0.042 | (-0.025, 0.012) |
| TG (46:4) B | Lognormal | -0.016 | (-0.060, 0.027) | 0.469 | 0.863 | 0.045 | (-0.060, 0.027) |
| PC (32:0) - ESI(-) | Lognormal | -0.003 | (-0.012, 0.006) | 0.473 | 0.863 | 0.043 | (-0.011, 0.006) |
| TG (62:1) | Lognormal | -0.010 | (-0.038, 0.018) | 0.474 | 0.863 | 0.045 | (-0.039, 0.017) |
| PC (35:2) | Lognormal | -0.004 | (-0.016, 0.007) | 0.475 | 0.863 | 0.047 | (-0.016, 0.007) |
| Ceramide (d34:1) - ESI(-) | Lognormal | -0.003 | (-0.012, 0.005) | 0.475 | 0.863 | 0.043 | (-0.013, 0.004) |
| LPC (20:1) - ESI(+) | Lognormal | -0.010 | (-0.039, 0.018) | 0.475 | 0.863 | 0.046 | (-0.039, 0.018) |
| LPC (p-18:0)/LPC (o-18:1) | Lognormal | -0.005 | (-0.020, 0.010) | 0.475 | 0.863 | 0.045 | (-0.020, 0.010) |
| TG (52:4) | Lognormal | -0.003 | (-0.013, 0.006) | 0.490 | 0.879 | 0.045 | (-0.013, 0.006) |
| AC (14:1) | Lognormal | -0.008 | (-0.030, 0.014) | 0.490 | 0.879 | 0.043 | (-0.030, 0.014) |
| TG (53:3) | Lognormal | 0.004 | (-0.007, 0.015) | 0.497 | 0.887 | 0.044 | (-0.007, 0.015) |
| TG (53:2) | Lognormal | 0.005 | (-0.009, 0.018) | 0.504 | 0.892 | 0.043 | (-0.010, 0.018) |
| PC (33:0) | Lognormal | -0.006 | (-0.024, 0.012) | 0.509 | 0.894 | 0.043 | (-0.023, 0.012) |
| Ceramide (d42:2) A - ESI(-) | Lognormal | -0.003 | (-0.013, 0.007) | 0.514 | 0.896 | 0.039 | (-0.014, 0.007) |
| FA (18:2) (linoleic acid) | Lognormal | 0.003 | (-0.006, 0.011) | 0.514 | 0.896 | 0.038 | (-0.006, 0.012) |
| DG (36:4) A | Lognormal | -0.006 | (-0.022, 0.011) | 0.516 | 0.896 | 0.040 | (-0.022, 0.011) |
| GlcCer (d40:1) - ESI(-) | Lognormal | -0.004 | (-0.015, 0.008) | 0.518 | 0.896 | 0.041 | (-0.015, 0.008) |
| PE (34:2) - ESI(+) | Lognormal | -0.022 | (-0.088, 0.044) | 0.522 | 0.896 | 0.040 | (-0.091, 0.044) |
| TG (60:1) | Lognormal | -0.009 | (-0.038, 0.019) | 0.523 | 0.896 | 0.044 | (-0.039, 0.019) |
| LPC (15:0) | Lognormal | -0.004 | (-0.018, 0.009) | 0.528 | 0.896 | 0.042 | (-0.018, 0.009) |
| SM (d36:3) - ESI(+) | Lognormal | -0.004 | (-0.016, 0.008) | 0.528 | 0.896 | 0.039 | (-0.016, 0.008) |
| CE (22:6) | Lognormal | 0.004 | (-0.008, 0.015) | 0.529 | 0.896 | 0.040 | (-0.008, 0.015) |
| TG (56:3) | Lognormal | 0.006 | (-0.013, 0.026) | 0.530 | 0.896 | 0.041 | (-0.014, 0.026) |
| FA (20:4) (arachidonic acid) | Lognormal | -0.002 | (-0.009, 0.005) | 0.533 | 0.896 | 0.038 | (-0.009, 0.005) |
| PC (p-40:7)/PC (o-40:8) | Lognormal | -0.013 | (-0.054, 0.028) | 0.533 | 0.896 | 0.039 | (-0.053, 0.029) |
| FA (22:6) (docosahexaenoic acid) | Lognormal | 0.005 | (-0.010, 0.019) | 0.535 | 0.896 | 0.041 | (-0.010, 0.019) |

**Supplementary Table S7.** Regression and Bayesian estimates comparing ME/CFS without sr-IBS vs. controls without sr-IBS.

| Metabolite | Regression Model | ME/CFS without sr-IBS vs. Controls without sr-IBS | | | | | |
|---|---|---|---|---|---|---|---|
| | | Estimated Coefficient | 95% CI | p-value | FDR | BayesFactor | 95% HDI |
| TG (40:0) | Lognormal | -0.010 | (-0.042, 0.022) | 0.536 | 0.896 | 0.041 | (-0.042, 0.021) |
| Ceramide (d36:1) - ESI(+) | Lognormal | 0.004 | (-0.008, 0.015) | 0.537 | 0.896 | 0.039 | (-0.008, 0.015) |
| PC (36:1) - ESI(-) | Lognormal | -0.003 | (-0.012, 0.006) | 0.541 | 0.898 | 0.042 | (-0.012, 0.005) |
| TG (56:1) | Lognormal | -0.008 | (-0.034, 0.018) | 0.545 | 0.899 | 0.042 | (-0.034, 0.018) |
| TG (48:2) | Lognormal | -0.007 | (-0.029, 0.015) | 0.545 | 0.899 | 0.039 | (-0.031, 0.015) |
| LPC (18:1) - ESI(+) | Lognormal | -0.003 | (-0.014, 0.008) | 0.550 | 0.905 | 0.044 | (-0.014, 0.008) |
| Ceramide (d34:0) | Lognormal | -0.004 | (-0.017, 0.009) | 0.551 | 0.905 | 0.034 | (-0.017, 0.008) |
| PE (36:1) - ESI(+) | Lognormal | -0.007 | (-0.029, 0.015) | 0.553 | 0.907 | 0.039 | (-0.029, 0.015) |
| PC (o-32:0) - ESI(+) | Lognormal | -0.002 | (-0.010, 0.006) | 0.557 | 0.908 | 0.038 | (-0.011, 0.005) |
| PC (37:4) - ESI(-) | Lognormal | -0.004 | (-0.018, 0.010) | 0.558 | 0.908 | 0.042 | (-0.018, 0.010) |
| TG (48:1) | Lognormal | -0.006 | (-0.027, 0.015) | 0.558 | 0.908 | 0.039 | (-0.028, 0.014) |
| TG (58:10) | Lognormal | 0.006 | (-0.013, 0.024) | 0.563 | 0.909 | 0.042 | (-0.012, 0.026) |
| PC (35:4) - ESI(+) | Lognormal | -0.003 | (-0.014, 0.008) | 0.575 | 0.914 | 0.038 | (-0.014, 0.007) |
| TG (53:1) | Lognormal | -0.008 | (-0.037, 0.021) | 0.578 | 0.916 | 0.040 | (-0.037, 0.020) |
| PC (p-42:4)/PC (o-42:5) - ESI(-) | Lognormal | -0.005 | (-0.024, 0.013) | 0.581 | 0.917 | 0.038 | (-0.024, 0.013) |
| PC 38:7e | Lognormal | -0.005 | (-0.021, 0.012) | 0.585 | 0.920 | 0.037 | (-0.022, 0.011) |
| FA (20:2) (eicosadienoic acid) | Lognormal | 0.003 | (-0.009, 0.015) | 0.596 | 0.930 | 0.042 | (-0.009, 0.015) |
| TG (54:3) | Lognormal | 0.003 | (-0.008, 0.013) | 0.598 | 0.931 | 0.042 | (-0.008, 0.014) |
| LPC (17:1) | Lognormal | 0.004 | (-0.011, 0.019) | 0.599 | 0.931 | 0.039 | (-0.010, 0.020) |
| PC (35:2) B | Lognormal | -0.002 | (-0.008, 0.005) | 0.600 | 0.931 | 0.035 | (-0.009, 0.005) |
| DG (36:4) B | Lognormal | 0.010 | (-0.027, 0.047) | 0.600 | 0.931 | 0.040 | (-0.026, 0.048) |
| TG (58:4) | Lognormal | 0.006 | (-0.017, 0.030) | 0.605 | 0.933 | 0.041 | (-0.018, 0.031) |
| TG (59:3) | Lognormal | 0.006 | (-0.017, 0.029) | 0.605 | 0.933 | 0.037 | (-0.017, 0.029) |
| GlcCer (d42:1) - ESI(-) | Lognormal | -0.003 | (-0.014, 0.008) | 0.609 | 0.933 | 0.036 | (-0.013, 0.008) |
| CE (16:0) | Lognormal | 0.007 | (-0.020, 0.034) | 0.613 | 0.933 | 0.034 | (-0.019, 0.034) |
| SM (d34:2) - ESI(+) | Lognormal | -0.002 | (-0.008, 0.005) | 0.613 | 0.933 | 0.036 | (-0.008, 0.005) |
| TG (56:7) A | Lognormal | 0.011 | (-0.031, 0.052) | 0.619 | 0.934 | 0.041 | (-0.034, 0.049) |
| PC (p-40:4)/PC (o-40:5) - ESI(+) | Lognormal | -0.002 | (-0.012, 0.007) | 0.619 | 0.934 | 0.037 | (-0.012, 0.007) |
| SM (d34:0) - ESI(+) | Lognormal | -0.002 | (-0.010, 0.006) | 0.620 | 0.934 | 0.035 | (-0.010, 0.006) |
| PC (35:4) - ESI(-) | Lognormal | -0.004 | (-0.020, 0.012) | 0.624 | 0.934 | 0.039 | (-0.020, 0.012) |
| PC (34:0) - ESI(+) | Lognormal | -0.001 | (-0.007, 0.004) | 0.625 | 0.934 | 0.032 | (-0.007, 0.004) |
| TG (48:6) | Lognormal | 0.006 | (-0.019, 0.032) | 0.631 | 0.938 | 0.039 | (-0.020, 0.031) |
| PC (p-44:4)/PC (o-44:5) - ESI(+) | Lognormal | 0.003 | (-0.009, 0.015) | 0.636 | 0.938 | 0.039 | (-0.009, 0.015) |
| PC (p-40:6)/PC (o-40:7) B | Lognormal | -0.004 | (-0.019, 0.011) | 0.637 | 0.938 | 0.036 | (-0.019, 0.012) |
| SM (d34:1) - ESI(+) | Lognormal | -0.001 | (-0.006, 0.004) | 0.638 | 0.938 | 0.034 | (-0.006, 0.004) |
| PC (35:1) - ESI(-) | Lognormal | 0.003 | (-0.011, 0.018) | 0.639 | 0.938 | 0.037 | (-0.010, 0.018) |
| GlcCer (d40:1) - ESI(+) | Lognormal | -0.003 | (-0.016, 0.010) | 0.641 | 0.938 | 0.036 | (-0.017, 0.009) |
| Ceramide (d40:2) | Lognormal | 0.010 | (-0.032, 0.052) | 0.646 | 0.938 | 0.038 | (-0.030, 0.051) |
| TG (54:2) | Lognormal | 0.004 | (-0.011, 0.019) | 0.647 | 0.938 | 0.040 | (-0.011, 0.019) |
| DG (34:2) | Lognormal | 0.003 | (-0.009, 0.014) | 0.648 | 0.938 | 0.033 | (-0.009, 0.014) |
| SM (d42:0) - ESI(+) | Lognormal | -0.006 | (-0.034, 0.021) | 0.648 | 0.938 | 0.037 | (-0.033, 0.022) |
| PC (33:1) - ESI(-) | Lognormal | 0.005 | (-0.016, 0.025) | 0.650 | 0.938 | 0.037 | (-0.015, 0.027) |
| TG (58:1) | Lognormal | -0.006 | (-0.032, 0.020) | 0.651 | 0.938 | 0.035 | (-0.030, 0.021) |
| PC (36:5) B | Lognormal | 0.005 | (-0.017, 0.028) | 0.651 | 0.938 | 0.038 | (-0.016, 0.027) |
| FA (18:3) (linolenic acid) | Lognormal | 0.003 | (-0.010, 0.016) | 0.662 | 0.938 | 0.040 | (-0.011, 0.015) |
| PC (16:0/9:0(CHO)) | Lognormal | 0.005 | (-0.016, 0.026) | 0.662 | 0.938 | 0.031 | (-0.015, 0.026) |
| PC (38:4) A - ESI(+) | Lognormal | -0.002 | (-0.012, 0.008) | 0.662 | 0.938 | 0.038 | (-0.012, 0.008) |

**Supplementary Table S7.** Regression and Bayesian estimates comparing ME/CFS without sr-IBS vs. controls without sr-IBS.

| Metabolite | Regression Model | ME/CFS without sr-IBS vs. Controls without sr-IBS | | | | | |
|---|---|---|---|---|---|---|---|
| | | Estimated Coefficient | 95% CI | p-value | FDR | BayesFactor | 95% HDI |
| FA (24:1) (nervonic acid) | Lognormal | 0.002 | (-0.008, 0.012) | 0.664 | 0.938 | 0.034 | (-0.008, 0.012) |
| TG (50:2) | Lognormal | 0.003 | (-0.009, 0.014) | 0.667 | 0.938 | 0.036 | (-0.008, 0.014) |
| SM (d38:0) | Lognormal | -0.014 | (-0.077, 0.049) | 0.668 | 0.938 | 0.036 | (-0.077, 0.048) |
| SM (d41:1) - ESI(-) | Lognormal | 0.023 | (-0.082, 0.127) | 0.669 | 0.938 | 0.039 | (-0.081, 0.129) |
| TG (58:5) | Lognormal | -0.004 | (-0.022, 0.014) | 0.670 | 0.938 | 0.036 | (-0.021, 0.014) |
| PC (38:5) B - ESI(-) | Lognormal | 0.004 | (-0.013, 0.021) | 0.671 | 0.938 | 0.038 | (-0.013, 0.021) |
| TG (49:0) | Lognormal | -0.005 | (-0.026, 0.017) | 0.675 | 0.938 | 0.037 | (-0.025, 0.017) |
| SM (d36:2) - ESI(-) | Lognormal | -0.003 | (-0.015, 0.010) | 0.676 | 0.938 | 0.038 | (-0.015, 0.010) |
| TG (46:3) B | Lognormal | -0.008 | (-0.044, 0.029) | 0.677 | 0.938 | 0.037 | (-0.042, 0.030) |
| PE (p-36:5)/PE (o-36:6) | Lognormal | -0.011 | (-0.065, 0.042) | 0.678 | 0.938 | 0.037 | (-0.063, 0.043) |
| PC (36:6) | Lognormal | -0.004 | (-0.022, 0.014) | 0.680 | 0.939 | 0.034 | (-0.021, 0.015) |
| PC (p-38:5)/PC (o-38:6) A | Lognormal | -0.003 | (-0.016, 0.011) | 0.682 | 0.939 | 0.035 | (-0.016, 0.011) |
| PC (40:8) - ESI(+) | Lognormal | -0.002 | (-0.011, 0.007) | 0.684 | 0.939 | 0.033 | (-0.011, 0.007) |
| PC (40:6) B | Lognormal | 0.003 | (-0.011, 0.016) | 0.700 | 0.945 | 0.040 | (-0.011, 0.016) |
| TG (51:4) | Lognormal | -0.003 | (-0.018, 0.012) | 0.701 | 0.945 | 0.036 | (-0.019, 0.013) |
| FA (15:0) (pentadecylic acid) | Lognormal | -0.002 | (-0.010, 0.007) | 0.705 | 0.946 | 0.033 | (-0.010, 0.007) |
| SM (d36:1) - ESI(+) | Lognormal | 0.001 | (-0.006, 0.008) | 0.706 | 0.946 | 0.038 | (-0.005, 0.009) |
| TG (54:6) A | Lognormal | -0.003 | (-0.020, 0.014) | 0.715 | 0.948 | 0.036 | (-0.019, 0.014) |
| Ceramide (d40:0) | Lognormal | -0.005 | (-0.034, 0.023) | 0.718 | 0.948 | 0.030 | (-0.034, 0.023) |
| TG (58:3) | Lognormal | 0.005 | (-0.023, 0.033) | 0.722 | 0.948 | 0.036 | (-0.023, 0.033) |
| PC (37:4) - ESI(+) | Lognormal | 0.002 | (-0.010, 0.015) | 0.722 | 0.948 | 0.035 | (-0.010, 0.014) |
| PC (38:5) A - ESI(-) | Lognormal | -0.002 | (-0.011, 0.008) | 0.725 | 0.948 | 0.037 | (-0.011, 0.008) |
| PC (32:1) - ESI(-) | Lognormal | -0.003 | (-0.020, 0.014) | 0.728 | 0.948 | 0.035 | (-0.020, 0.015) |
| Gal-Gal-Cer (d18:1/16:0)/Lactosylceramide (d18:1/16:0) | Lognormal | 0.002 | (-0.007, 0.011) | 0.728 | 0.948 | 0.033 | (-0.007, 0.011) |
| SM (d43:2) - ESI(+) | Lognormal | 0.003 | (-0.014, 0.019) | 0.729 | 0.948 | 0.037 | (-0.014, 0.020) |
| LPC (22:5) - ESI(+) | Lognormal | 0.003 | (-0.016, 0.023) | 0.730 | 0.948 | 0.038 | (-0.016, 0.023) |
| CE (16:1) | Lognormal | 0.003 | (-0.012, 0.017) | 0.739 | 0.954 | 0.033 | (-0.011, 0.017) |
| TG (62:2) | Lognormal | -0.005 | (-0.035, 0.025) | 0.742 | 0.955 | 0.035 | (-0.036, 0.025) |
| SM (d42:3) - ESI(+) | Lognormal | 0.001 | (-0.006, 0.008) | 0.745 | 0.955 | 0.036 | (-0.006, 0.008) |
| TG (62:3) | Lognormal | -0.006 | (-0.042, 0.030) | 0.747 | 0.955 | 0.035 | (-0.043, 0.031) |
| TG (51:3) | Lognormal | 0.002 | (-0.010, 0.015) | 0.751 | 0.955 | 0.032 | (-0.011, 0.015) |
| TG (52:1) | Lognormal | -0.003 | (-0.023, 0.017) | 0.753 | 0.955 | 0.036 | (-0.025, 0.015) |
| DG (36:1) | Lognormal | 0.003 | (-0.014, 0.020) | 0.755 | 0.955 | 0.036 | (-0.014, 0.019) |
| PE (36:4) - ESI(+) | Lognormal | -0.004 | (-0.026, 0.019) | 0.756 | 0.955 | 0.033 | (-0.026, 0.018) |
| SM (d42:0) - ESI(-) | Lognormal | -0.002 | (-0.015, 0.011) | 0.756 | 0.955 | 0.033 | (-0.015, 0.011) |
| SM (d41:2) A - ESI(+) | Lognormal | 0.002 | (-0.009, 0.012) | 0.759 | 0.955 | 0.031 | (-0.008, 0.013) |
| PC (37:3) | Lognormal | -0.002 | (-0.016, 0.011) | 0.759 | 0.955 | 0.037 | (-0.016, 0.012) |
| SM (d37:1) | Lognormal | -0.003 | (-0.022, 0.016) | 0.759 | 0.955 | 0.037 | (-0.022, 0.016) |
| DG (34:1) | Lognormal | -0.004 | (-0.029, 0.021) | 0.767 | 0.955 | 0.032 | (-0.030, 0.021) |
| TG (54:4) | Lognormal | 0.002 | (-0.010, 0.013) | 0.769 | 0.955 | 0.033 | (-0.009, 0.014) |
| TG (54:8) | Lognormal | -0.004 | (-0.029, 0.021) | 0.771 | 0.955 | 0.039 | (-0.028, 0.021) |
| PC (p-42:4)/PC (o-42:5) - ESI(+) | Lognormal | -0.002 | (-0.013, 0.010) | 0.772 | 0.955 | 0.032 | (-0.012, 0.010) |
| SM (d33:1) - ESI(+) | Lognormal | -0.001 | (-0.011, 0.008) | 0.773 | 0.955 | 0.036 | (-0.011, 0.009) |
| PC (p-38:5)/PC (o-38:6) | Lognormal | -0.005 | (-0.043, 0.032) | 0.778 | 0.958 | 0.039 | (-0.041, 0.033) |
| PC (42:5) | Lognormal | 0.002 | (-0.013, 0.017) | 0.780 | 0.958 | 0.035 | (-0.013, 0.018) |
| CE (20:4) | Lognormal | -0.001 | (-0.011, 0.008) | 0.785 | 0.958 | 0.038 | (-0.011, 0.008) |
| TG (59:2) | Lognormal | -0.003 | (-0.028, 0.021) | 0.785 | 0.958 | 0.033 | (-0.030, 0.019) |

**Supplementary Table S7.** Regression and Bayesian estimates comparing ME/CFS without sr-IBS vs. controls without sr-IBS.

| Metabolite | Regression Model | ME/CFS without sr-IBS vs. Controls without sr-IBS | | | | | |
|---|---|---|---|---|---|---|---|
| | | Estimated Coefficient | 95% CI | p-value | FDR | BayesFactor | 95% HDI |
| SM (d36:2) - ESI(+) | Lognormal | 0.001 | (-0.006, 0.008) | 0.791 | 0.958 | 0.034 | (-0.006, 0.008) |
| DG (34:3) | Lognormal | 0.003 | (-0.016, 0.022) | 0.795 | 0.959 | 0.031 | (-0.016, 0.022) |
| PC (o-32:0) - ESI(-) | Lognormal | -0.002 | (-0.014, 0.011) | 0.795 | 0.959 | 0.035 | (-0.015, 0.011) |
| LPC (22:5) - ESI(-) | Lognormal | -0.004 | (-0.039, 0.030) | 0.802 | 0.961 | 0.031 | (-0.036, 0.032) |
| GlcCer (d42:1) - ESI(+) | Lognormal | 0.001 | (-0.009, 0.011) | 0.804 | 0.963 | 0.032 | (-0.008, 0.012) |
| DG (36:5) | Lognormal | -0.003 | (-0.023, 0.018) | 0.806 | 0.963 | 0.033 | (-0.024, 0.019) |
| TG (52:5) | Lognormal | 0.002 | (-0.011, 0.014) | 0.812 | 0.963 | 0.034 | (-0.011, 0.014) |
| TG (53:4) | Lognormal | 0.002 | (-0.011, 0.014) | 0.815 | 0.967 | 0.034 | (-0.013, 0.014) |
| TG (50:3) B | Lognormal | -0.009 | (-0.088, 0.069) | 0.819 | 0.968 | 0.038 | (-0.088, 0.067) |
| PC (40:6) A | Lognormal | -0.001 | (-0.013, 0.010) | 0.823 | 0.972 | 0.035 | (-0.013, 0.009) |
| TG (57:2) | Lognormal | -0.002 | (-0.019, 0.015) | 0.828 | 0.975 | 0.033 | (-0.019, 0.014) |
| TG (60:2) | Lognormal | -0.004 | (-0.046, 0.037) | 0.834 | 0.979 | 0.033 | (-0.044, 0.036) |
| PC (40:7) - ESI(-) | Lognormal | -0.002 | (-0.021, 0.017) | 0.835 | 0.979 | 0.038 | (-0.021, 0.018) |
| SM (d41:2) - ESI(-) | Lognormal | -0.002 | (-0.016, 0.013) | 0.838 | 0.981 | 0.034 | (-0.016, 0.013) |
| PE (36:1) - ESI(-) | Lognormal | -0.003 | (-0.027, 0.022) | 0.843 | 0.981 | 0.033 | (-0.027, 0.024) |
| TG (46:5) | Lognormal | 0.005 | (-0.041, 0.051) | 0.843 | 0.981 | 0.038 | (-0.039, 0.050) |
| PC (p-34:1)/PC (o-34:2) B | Lognormal | -0.002 | (-0.021, 0.018) | 0.846 | 0.981 | 0.033 | (-0.022, 0.017) |
| TG (53:5) | Lognormal | -0.002 | (-0.017, 0.014) | 0.849 | 0.981 | 0.035 | (-0.017, 0.015) |
| PC (p-44:5)/PC (o-44:6) | Lognormal | 0.001 | (-0.013, 0.016) | 0.849 | 0.981 | 0.033 | (-0.013, 0.015) |
| DG (36:3) | Lognormal | 0.001 | (-0.010, 0.012) | 0.852 | 0.981 | 0.031 | (-0.010, 0.011) |
| PC (42:6) | Lognormal | -0.003 | (-0.030, 0.024) | 0.853 | 0.981 | 0.032 | (-0.028, 0.025) |
| TG (56:10) | Lognormal | 0.003 | (-0.029, 0.035) | 0.854 | 0.981 | 0.033 | (-0.027, 0.037) |
| Ceramide (d34:2) | Lognormal | -0.001 | (-0.013, 0.010) | 0.854 | 0.981 | 0.035 | (-0.013, 0.010) |
| PE (38:6) - ESI(-) | Lognormal | -0.002 | (-0.029, 0.024) | 0.861 | 0.981 | 0.035 | (-0.028, 0.025) |
| TG (52:3) | Lognormal | 0.001 | (-0.006, 0.007) | 0.864 | 0.981 | 0.034 | (-0.006, 0.007) |
| TAG (58:7)/TAG (18:1/18:1/22:5) | Lognormal | 0.001 | (-0.013, 0.015) | 0.881 | 0.984 | 0.038 | (-0.013, 0.014) |
| PE (38:6) - ESI(+) | Lognormal | -0.002 | (-0.034, 0.030) | 0.882 | 0.984 | 0.033 | (-0.034, 0.031) |
| PC (38:6) - ESI(-) | Lognormal | 0.001 | (-0.011, 0.013) | 0.884 | 0.984 | 0.034 | (-0.011, 0.013) |
| PC (36:4) C - ESI(+) | Lognormal | 0.000 | (-0.006, 0.005) | 0.884 | 0.984 | 0.036 | (-0.006, 0.005) |
| TG (55:2) | Lognormal | 0.001 | (-0.017, 0.020) | 0.889 | 0.984 | 0.036 | (-0.017, 0.020) |
| Ceramide (d36:1) - ESI(-) | Lognormal | -0.001 | (-0.021, 0.018) | 0.890 | 0.984 | 0.034 | (-0.021, 0.018) |
| PC (36:4) B - ESI(+) | Lognormal | 0.001 | (-0.012, 0.014) | 0.892 | 0.984 | 0.033 | (-0.012, 0.014) |
| SM (d36:0) - ESI(-) | Lognormal | 0.002 | (-0.028, 0.032) | 0.893 | 0.984 | 0.035 | (-0.028, 0.032) |
| TG (54:5) A | Lognormal | -0.001 | (-0.013, 0.012) | 0.896 | 0.984 | 0.033 | (-0.013, 0.012) |
| PC (p-40:4)/PC (o-40:5) - ESI(-) | Lognormal | 0.001 | (-0.013, 0.014) | 0.901 | 0.984 | 0.034 | (-0.014, 0.014) |
| GlcCer (d34:1) | Lognormal | 0.001 | (-0.020, 0.023) | 0.902 | 0.984 | 0.038 | (-0.021, 0.023) |
| TG (60:3) | Lognormal | -0.002 | (-0.033, 0.029) | 0.905 | 0.984 | 0.035 | (-0.033, 0.031) |
| TG (60:4) | Lognormal | -0.002 | (-0.034, 0.030) | 0.905 | 0.984 | 0.036 | (-0.035, 0.029) |
| PC (p-40:6)/PC (o-40:7) A | Lognormal | 0.001 | (-0.012, 0.013) | 0.914 | 0.988 | 0.034 | (-0.011, 0.013) |
| TG (56:4) | Lognormal | 0.001 | (-0.013, 0.014) | 0.923 | 0.990 | 0.035 | (-0.012, 0.015) |
| TG (49:1) | Lognormal | 0.001 | (-0.021, 0.023) | 0.926 | 0.990 | 0.032 | (-0.021, 0.024) |
| CE (18:1) | Lognormal | -0.001 | (-0.013, 0.012) | 0.927 | 0.990 | 0.033 | (-0.012, 0.013) |
| LPC (22:6) | Lognormal | 0.001 | (-0.014, 0.015) | 0.929 | 0.990 | 0.033 | (-0.014, 0.016) |
| FA (20:3) (eicosatrienoic acid) | Lognormal | 0.001 | (-0.015, 0.016) | 0.929 | 0.990 | 0.033 | (-0.014, 0.016) |
| SM (d32:0) - ESI(+) | Lognormal | -0.001 | (-0.017, 0.016) | 0.930 | 0.990 | 0.032 | (-0.017, 0.016) |
| AC (18:1) | Lognormal | 0.001 | (-0.011, 0.012) | 0.931 | 0.990 | 0.034 | (-0.012, 0.013) |
| PC (36:1) - ESI(+) | Lognormal | 0.000 | (-0.006, 0.006) | 0.932 | 0.990 | 0.032 | (-0.006, 0.006) |

**Supplementary Table S7.** Regression and Bayesian estimates comparing ME/CFS without sr-IBS vs. controls without sr-IBS.

| Metabolite | Regression Model | ME/CFS without sr-IBS vs. Controls without sr-IBS | | | | | |
|---|---|---|---|---|---|---|---|
| | | Estimated Coefficient | 95% CI | p-value | FDR | BayesFactor | 95% HDI |
| PC (39:4) | Lognormal | 0.000 | (-0.011, 0.010) | 0.937 | 0.991 | 0.033 | (-0.012, 0.010) |
| PC (40:5) A - ESI(-) | Lognormal | -0.001 | (-0.017, 0.016) | 0.938 | 0.991 | 0.035 | (-0.017, 0.015) |
| PC (31:0) | Lognormal | 0.000 | (-0.012, 0.013) | 0.946 | 0.994 | 0.035 | (-0.012, 0.013) |
| PC (32:1) - ESI(+) | Lognormal | 0.000 | (-0.012, 0.013) | 0.947 | 0.994 | 0.035 | (-0.012, 0.012) |
| SM (d40:0) | Lognormal | 0.000 | (-0.013, 0.013) | 0.957 | 0.998 | 0.033 | (-0.014, 0.012) |
| PC (p-42:5)/PC (o-42:6) B | Lognormal | 0.000 | (-0.015, 0.014) | 0.961 | 0.998 | 0.035 | (-0.016, 0.014) |
| TG (64:2) | Lognormal | -0.001 | (-0.031, 0.030) | 0.964 | 0.998 | 0.036 | (-0.027, 0.032) |
| PC 40:6e | Lognormal | 0.000 | (-0.010, 0.010) | 0.968 | 0.998 | 0.032 | (-0.010, 0.010) |
| TG (50:3) A | Lognormal | 0.000 | (-0.014, 0.014) | 0.976 | 0.998 | 0.031 | (-0.014, 0.015) |
| TG (49:3) | Lognormal | 0.000 | (-0.022, 0.022) | 0.977 | 0.998 | 0.033 | (-0.023, 0.021) |
| TG (55:1) | Lognormal | 0.000 | (-0.022, 0.022) | 0.978 | 0.998 | 0.035 | (-0.022, 0.022) |
| AC (16:0) | Lognormal | 0.000 | (-0.010, 0.011) | 0.978 | 0.998 | 0.032 | (-0.010, 0.011) |
| DG (32:1) | Lognormal | 0.000 | (-0.023, 0.023) | 0.981 | 0.998 | 0.030 | (-0.024, 0.022) |
| TG (57:1) | Lognormal | 0.000 | (-0.020, 0.019) | 0.983 | 0.998 | 0.033 | (-0.019, 0.019) |
| TG (44:2) | Lognormal | 0.001 | (-0.078, 0.079) | 0.987 | 0.998 | 0.035 | (-0.080, 0.079) |
| TG (54:9) | Lognormal | 0.000 | (-0.060, 0.059) | 0.988 | 0.998 | 0.033 | (-0.061, 0.059) |
| PC (p-36:5)/PC (o-36:6) | Lognormal | 0.000 | (-0.020, 0.020) | 0.989 | 0.998 | 0.034 | (-0.021, 0.020) |
| TG (56:2) | Lognormal | 0.000 | (-0.022, 0.022) | 0.992 | 0.998 | 0.035 | (-0.024, 0.023) |
| TG (50:6) | Lognormal | 0.000 | (-0.027, 0.028) | 0.996 | 0.998 | 0.034 | (-0.028, 0.027) |
| PC (31:1) | Lognormal | 0.000 | (-0.036, 0.036) | 0.996 | 0.998 | 0.034 | (-0.036, 0.037) |
| SM (d38:2) - ESI(+) | Lognormal | 0.000 | (-0.007, 0.007) | 0.997 | 0.998 | 0.033 | (-0.007, 0.007) |
| **Oxylipins (OL)** | | | | | | | |
| 11,12-Epoxyeicosa-5,8,14-trienoic acid | Gamma | 0.383 | (0.102, 0.666) | 0.007 | 0.310 | 1.128 | (0.108, 0.672) |
| 8,9-Epoxyeicosa-5,11,14-trienoic acid | Gamma | -0.351 | (-0.614, -0.086) | 0.008 | 0.310 | 0.680 | (-0.631, -0.086) |
| Prostaglandin F2a | Gamma | -0.348 | (-0.637, -0.058) | 0.016 | 0.310 | 0.294 | (-0.670, -0.013) |
| Resolvin D1 | Gamma | -0.379 | (-0.683, -0.073) | 0.016 | 0.310 | 0.597 | (-0.674, -0.055) |
| 12-Hydroxy-5,8,10,14,17-eicosapentaenoic acid | Gamma | 0.451 | (0.077, 0.828) | 0.017 | 0.310 | 3.271 | (0.158, 0.720) |
| 9,11,15-trihydroxy-5,13,1Z-prostatrienoic acid | Gamma | -0.520 | (-0.990, -0.049) | 0.022 | 0.327 | 2.494 | (-0.880, -0.175) |
| Leukotriene B4 | Gamma | -0.246 | (-0.473, -0.018) | 0.037 | 0.369 | 0.332 | (-0.437, -0.028) |
| 11-Hydroxy-14,15-epoxyeicosatrienoic acid | Gamma | -0.525 | (-1.040, -0.005) | 0.041 | 0.375 | 0.705 | (-0.971, -0.107) |
| 5-Hydroxy-6,8,11,14,17-eicosapentaenoic acid | Gamma | 0.236 | (-0.009, 0.483) | 0.052 | 0.402 | 0.189 | (0.010, 0.468) |
| 17,18-Epoxy-5,8,11,14-eicosatetraenoic acid | Gamma | -0.428 | (-0.874, 0.022) | 0.064 | 0.424 | 0.460 | (-0.810, -0.045) |
| 16(17)-epoxy-4,7,10,13,19-docosapentaenoic acid | Gamma | 0.324 | (-0.067, 0.717) | 0.082 | 0.479 | 0.166 | (-0.053, 0.691) |
| 9-Hydroxy-5,7,11,14,17-icosapentaenoic acid | Gamma | 0.205 | (-0.051, 0.462) | 0.105 | 0.542 | 0.092 | (-0.043, 0.464) |
| 8,9-dihydroxyeicosa-5,11,14-trienoic acid | Gamma | 0.181 | (-0.046, 0.410) | 0.114 | 0.557 | 0.073 | (-0.061, 0.416) |
| 17,18-dihydroxyeicosa-5,8,11,14-tetraenoic acid | Lognormal | 0.194 | (-0.060, 0.448) | 0.137 | 0.612 | 0.100 | (-0.050, 0.453) |
| 15-hydroxyeicosa-5,8,11,13,17-pentaenoic acid | Gamma | 0.268 | (-0.097, 0.637) | 0.145 | 0.622 | 0.112 | (-0.060, 0.626) |
| 13-ketooctadeca-9,11-dienoic acid | Lognormal | -0.196 | (-0.471, 0.079) | 0.165 | 0.653 | 0.079 | (-0.476, 0.077) |
| 5-ketoeicosa-6,8,11,14-tetraenoic acid | Gamma | -0.198 | (-0.499, 0.105) | 0.177 | 0.664 | 0.067 | (-0.535, 0.150) |
| Prostaglandin D2 | Gamma | -0.350 | (-0.884, 0.197) | 0.177 | 0.664 | 0.145 | (-0.784, 0.097) |
| 4-hydroxydocosa-5,7,10,13,16,19-hexaenoic acid | Gamma | 0.187 | (-0.125, 0.501) | 0.236 | 0.724 | 0.065 | (-0.126, 0.519) |
| 15,16-dihydroxyoctadeca-9,12-dienoic acid | Lognormal | -0.110 | (-0.294, 0.074) | 0.244 | 0.724 | 0.056 | (-0.297, 0.077) |
| 15(16)-epoxy-9,12-octadecadienoic acid | Gamma | -0.242 | (-0.687, 0.210) | 0.248 | 0.724 | 0.080 | (-0.602, 0.135) |
| 9,10-dihydroxyoctadec-12-enoic acid | Lognormal | -0.116 | (-0.316, 0.083) | 0.255 | 0.724 | 0.050 | (-0.313, 0.084) |
| 9,12,13-trihydroxyoctadec-10-enoic acid | Lognormal | -0.071 | (-0.210, 0.068) | 0.318 | 0.757 | 0.047 | (-0.218, 0.073) |
| 15-Deoxy-delta-12,14-Prostaglandin J2 | Gamma | -0.158 | (-0.454, 0.140) | 0.319 | 0.757 | 0.052 | (-0.454, 0.138) |
| 9-Hydroxylinoleic acid | Lognormal | -0.057 | (-0.188, 0.073) | 0.389 | 0.825 | 0.043 | (-0.182, 0.079) |

**Supplementary Table S7.** Regression and Bayesian estimates comparing ME/CFS without sr-IBS vs. controls without sr-IBS.

| Metabolite | Regression Model | ME/CFS without sr-IBS vs. Controls without sr-IBS | | | | | |
|---|---|---|---|---|---|---|---|
| | | Estimated Coefficient | 95% CI | p-value | FDR | BayesFactor | 95% HDI |
| 5,6-dihydroxyeicosa-8,11,14-trienoic acid | Gamma | 0.094 | (-0.135, 0.324) | 0.412 | 0.834 | 0.033 | (-0.137, 0.326) |
| 15-Keto-prostaglandin E2 | Gamma | -0.191 | (-0.680, 0.306) | 0.415 | 0.834 | 0.064 | (-0.596, 0.226) |
| 14,15-dihydroxyeicosa-5,8,11,17-tetraenoic acid | Gamma | -0.138 | (-0.483, 0.212) | 0.420 | 0.834 | 0.046 | (-0.472, 0.202) |
| 11(12)-epoxy-5,8,14,17- eicosatetraenoic acid | Gamma | -0.162 | (-0.591, 0.273) | 0.447 | 0.854 | 0.057 | (-0.573, 0.259) |
| 12,13-dihydroxyoctadeca-9,15-dienoic acid | Gamma | -0.186 | (-0.659, 0.295) | 0.448 | 0.854 | 0.059 | (-0.568, 0.149) |
| Prostaglandin E1 | Gamma | 0.126 | (-0.239, 0.494) | 0.484 | 0.875 | 0.044 | (-0.188, 0.447) |
| 14(15)-epoxy-5,8,11,17-eicosatetraenoic acid | Gamma | 0.200 | (-0.465, 0.874) | 0.504 | 0.892 | 0.081 | (-0.368, 0.711) |
| 12,13-dihydroxyoctadec-9-enoic acid | Lognormal | -0.061 | (-0.251, 0.129) | 0.529 | 0.896 | 0.038 | (-0.258, 0.130) |
| 17-hydroxy-4,7,10,13,15,19-docosahexaenoic acid | Gamma | 0.075 | (-0.177, 0.328) | 0.551 | 0.905 | 0.033 | (-0.178, 0.358) |
| 10-nitrolinoleic acid | Gamma | 0.120 | (-0.319, 0.564) | 0.571 | 0.914 | 0.045 | (-0.253, 0.499) |
| Leukotriene B5 | Gamma | -0.150 | (-0.680, 0.385) | 0.577 | 0.916 | 0.057 | (-0.645, 0.303) |
| 20-Hydroxyarachidonic acid | Lognormal | -0.096 | (-0.447, 0.254) | 0.590 | 0.924 | 0.029 | (-0.436, 0.269) |
| 5,6,15-trihydroxyeicosa-7,9,11,13-tetraenoic acid | Gamma | -0.090 | (-0.424, 0.249) | 0.609 | 0.933 | 0.037 | (-0.397, 0.240) |
| 8-hydroxyeicosa-5,9,11,14-tetraenoic acid | Gamma | 0.064 | (-0.183, 0.312) | 0.613 | 0.933 | 0.030 | (-0.205, 0.329) |
| 9,10-Epoxystearic acid | Lognormal | -0.083 | (-0.405, 0.239) | 0.614 | 0.933 | 0.037 | (-0.399, 0.229) |
| 8,15-dihydroxyeicosa-5,9,11,13-tetraenoic acid | Gamma | -0.140 | (-0.660, 0.393) | 0.616 | 0.934 | 0.058 | (-0.614, 0.324) |
| 9,10-dihydroxyoctadeca-12,15-dienoic acid | Gamma | -0.051 | (-0.273, 0.173) | 0.647 | 0.938 | 0.026 | (-0.294, 0.186) |
| 15-ketoeicosa-5,8,11,13-tetraenoic acid | Gamma | 0.085 | (-0.307, 0.480) | 0.669 | 0.938 | 0.036 | (-0.224, 0.398) |
| 5-Hydroxy-6,8,11,14-eicosatetraenoic acid | Lognormal | -0.039 | (-0.241, 0.163) | 0.707 | 0.946 | 0.031 | (-0.238, 0.157) |
| 9S,10R-dihydroxy-stearic acid | Lognormal | -0.049 | (-0.326, 0.228) | 0.731 | 0.948 | 0.036 | (-0.323, 0.235) |
| 10-nitrooleic acid | Lognormal | 0.042 | (-0.202, 0.285) | 0.736 | 0.952 | 0.033 | (-0.209, 0.283) |
| 9-nitrooleic acid | Gamma | -0.049 | (-0.364, 0.270) | 0.760 | 0.955 | 0.031 | (-0.350, 0.269) |
| 9-hydroxyoctadeca-10,12,15-trienoic acid | Gamma | -0.039 | (-0.301, 0.224) | 0.763 | 0.955 | 0.025 | (-0.297, 0.216) |
| 18-(3-ethyloxiran-2-yl)octadeca-4,7,10,13,16-pentaenoic acid | Gamma | 0.049 | (-0.332, 0.433) | 0.786 | 0.958 | 0.035 | (-0.247, 0.406) |
| 19,20-dihydroxydocosa-4,7,10,13,16-pentaenoic acid | Lognormal | 0.024 | (-0.149, 0.196) | 0.788 | 0.958 | 0.031 | (-0.156, 0.188) |
| 12(13)-epoxy-9,15-octadecadienoic acid | Gamma | -0.049 | (-0.477, 0.384) | 0.811 | 0.963 | 0.038 | (-0.411, 0.289) |
| 9(10)-epoxy-12Z-octadecenoic acid | Gamma | 0.020 | (-0.210, 0.251) | 0.864 | 0.981 | 0.020 | (-0.173, 0.218) |
| Prostaglandin E3 | Gamma | 0.046 | (-0.612, 0.715) | 0.872 | 0.984 | 0.062 | (-0.556, 0.566) |
| 14,15-dihydroxyeicosa-5,8,11-trienoic acid | Gamma | -0.011 | (-0.158, 0.136) | 0.882 | 0.984 | 0.016 | (-0.153, 0.159) |
| 14-hydroxydocosa-4,7,10,12,16,19-hexaenoic acid | Lognormal | 0.022 | (-0.307, 0.350) | 0.897 | 0.984 | 0.022 | (-0.306, 0.338) |
| 11,12-Dihydroxyicosa-5,8,14-trienoic acid | Gamma | -0.010 | (-0.171, 0.151) | 0.902 | 0.984 | 0.016 | (-0.167, 0.151) |
| 5,15-dihydroxyeicosa-6,8,11,13-tetraenoic acid | Gamma | 0.028 | (-0.467, 0.531) | 0.907 | 0.984 | 0.049 | (-0.420, 0.508) |
| 9-ketooctadeca-10,12-dienoic acid | Gamma | -0.013 | (-0.235, 0.210) | 0.908 | 0.984 | 0.030 | (-0.247, 0.266) |
| 12,13-epoxy-9-octadecenoic acid | Lognormal | -0.017 | (-0.323, 0.290) | 0.916 | 0.988 | 0.034 | (-0.338, 0.269) |
| 9-hydroxyeicosa-5,7,11,14-tetraenoic acid | Gamma | -0.012 | (-0.242, 0.220) | 0.918 | 0.989 | 0.027 | (-0.267, 0.242) |
| 13-Hydroxyoctadecadienoic acid | Lognormal | -0.005 | (-0.127, 0.118) | 0.938 | 0.991 | 0.030 | (-0.128, 0.118) |
| Thromboxane B2 | Lognormal | -0.010 | (-0.275, 0.254) | 0.939 | 0.991 | 0.036 | (-0.288, 0.246) |
| 12-Hydroxy-5,8,10,14-eicosatetraenoic acid | Lognormal | 0.009 | (-0.212, 0.229) | 0.940 | 0.991 | 0.025 | (-0.229, 0.218) |
| 15-hydroxyeicosa-5,8,11,13-tetraenoic acid | Lognormal | -0.005 | (-0.166, 0.155) | 0.948 | 0.994 | 0.029 | (-0.172, 0.159) |
| 13-hydroxyoctadeca-9,11,15-trienoic acid | Gamma | 0.007 | (-0.239, 0.254) | 0.957 | 0.998 | 0.025 | (-0.240, 0.246) |
| trans-12,13-epoxy-11-oxo-trans-9-octadecenoic acid | Gamma | -0.005 | (-0.209, 0.200) | 0.960 | 0.998 | 0.020 | (-0.219, 0.197) |
| 6-Ketoprostaglandin F1 alpha | Gamma | 0.009 | (-0.398, 0.420) | 0.963 | 0.998 | 0.038 | (-0.342, 0.389) |
| 6-trans-Leukotriene B4 | Gamma | -0.004 | (-0.491, 0.490) | 0.987 | 0.998 | 0.038 | (-0.426, 0.452) |
| 9(10)-epoxy-12,15-octadecadienoic acid | Gamma | 0.003 | (-0.537, 0.549) | 0.990 | 0.998 | 0.041 | (-0.389, 0.422) |
| 11-Hydroxy-arachidonic acid | Gamma | 0.001 | (-0.178, 0.180) | 0.994 | 0.998 | 0.019 | (-0.188, 0.190) |
| Prostaglandin E2 | Gamma | 0.002 | (-0.528, 0.537) | 0.995 | 0.998 | 0.039 | (-0.380, 0.381) |

**Supplementary Table S7.** Regression and Bayesian estimates comparing ME/CFS without sr-IBS vs. controls without sr-IBS.

| Metabolite | Regression Model | ME/CFS without sr-IBS vs. Controls without sr-IBS | | | | | |
|---|---|---|---|---|---|---|---|
| | | Estimated Coefficient | 95% CI | p-value | FDR | BayesFactor | 95% HDI |

ME/CFS, myalgic encephalomyelitis/chronic fatigue syndrome; sr-IBS, self-reported physician diagnosed irritable bowl syndrome; CI, confidence interval; FDR, false discovery rate adjusted p-value; HDI, highest density credible intervals. Regression models were adjusted for age, sex, race/ethnicity, geographic/clinical site, season of sampling, and body mass index. In PM, BA, and CL panels, estimated coefficients are interpreted as the differences in the mean values of log-log transformation of metabolite levels between cases and controls. In OL panel for lognormal regression, estimated coefficients are interpreted as the mean differences of log transformation of metabolite levels between two groups; for Gamma regression, estimated coefficients are interpreted as the log of fold change between two groups. Estimations in **bold** are significant. Criteria for significance: 1) FDR adjusted p-vale from the regression model < 0.15, 2) BayesFactor >3, and 3) 95% highest density credible intervals not covering 0.

**Supplementary Table S8.** ChemRICH enrichment analysis comparing ME/CFS without sr-IBS vs. controls without sr-IBS.

| Cluster name | Cluster size | p-values | Key compound | Altered | Increased | Decreased | Altered Ratio |
|---|---|---|---|---|---|---|---|
| plasmalogens | 24 | 0.000000058 | PE (p-36:2)/PE (o-36:3) - ESI(+) | 15 | 0 | 9 | 0.6 |
| unsaturated phospholipid ethers | 42 | 0.000003 | PC (p-34:2)/PC (o-34:3) - ESI(+) | 17 | 0 | 14 | 0.4 |
| sphingomyelins | 49 | 0.000019 | SM (d43:1) - ESI(-) | 19 | 0 | 11 | 0.4 |
| unsaturated phosphatidylcholines | 90 | 0.000041 | PC (36:2) | 24 | 0 | 16 | 0.3 |
| exposome food | 14 | 0.00015 | Choline | 8 | 0 | 4 | 0.6 |
| unsaturated ceramides | 21 | 0.0002 | Ceramide (d42:1) - ESI (+) | 7 | 0 | 7 | 0.3 |
| saturated lysophospholipids | 10 | 0.00056 | LPC (14:0) - ESI(+) | 6 | 0 | 4 | 0.6 |
| HEPE | 5 | 0.00074 | 12-Hydroxy-5,8,10,14,17-eicosapentaenoic acid | 2 | 1 | 0 | 0.4 |
| unsaturated lysophosphatidylcholines | 20 | 0.0055 | LPC (18:3) | 6 | 0 | 5 | 0.3 |
| unsaturated long chain TGs | 41 | 0.012 | TG (62:4) | 9 | 2 | 1 | 0.2 |
| EpODE | 7 | 0.017 | 11,12-Epoxyeicosa-5,8,14-trienoic acid | 4 | 1 | 2 | 0.6 |
| carnitines | 27 | 0.028 | 2-Methylbutyryl-L-carnitine | 3 | 0 | 2 | 0.1 |
| saturated TGs | 8 | 0.029 | TG (50:0) | 2 | 0 | 2 | 0.2 |
| saturated lysophosphoethanolamines | 4 | 0.069 | LPE (18:0) | 1 | 0 | 1 | 0.2 |
| unsaturated lysophosphoethanolamines | 6 | 0.071 | LPE (20:4) - ESI(-) | 3 | 0 | 2 | 0.5 |
| drugs | 85 | 0.074 | Acetominophen | 15 | 7 | 2 | 0.2 |
| amino acids | 24 | 0.088 | L-Citrulline | 4 | 0 | 2 | 0.2 |
| unsaturated FFA | 15 | 0.094 | FA (16:1) (palmitoleic acid) | 1 | 0 | 0 | 0.07 |
| dicarboxylic acids | 9 | 0.14 | Succinic acid | 2 | 1 | 0 | 0.2 |
| cholesterol esters | 13 | 0.27 | CE (18:3) | 3 | 0 | 3 | 0.2 |
| dipeptides | 16 | 0.41 | N-Acetylhistidine | 1 | 0 | 1 | 0.06 |
| prostaglandins | 9 | 0.43 | Prostaglandin F2a | 2 | 0 | 2 | 0.2 |
| OH-FA_22_6_1 | 3 | 0.45 | Resolvin D1 | 1 | 0 | 1 | 0.3 |
| amino acids, basic | 8 | 0.46 | SDMA | 2 | 0 | 2 | 0.2 |
| unsaturated phosphatidylethanolamines | 13 | 0.48 | PE (36:2) | 1 | 0 | 0 | 0.08 |
| amino acids, branched-chain | 3 | 0.53 | Leucine | 1 | 0 | 1 | 0.3 |
| unsaturated glucyosylceramides | 12 | 0.65 | GlcCer (d42:2) - ESI(+) | 1 | 0 | 0 | 0.08 |
| DiHETE | 8 | 0.69 | Leukotriene B4 | 1 | 0 | 1 | 0.1 |
| adenosine | 4 | 0.73 | 7-Methylguanosine | 1 | 0 | 0 | 0.2 |
| sugar alcohols | 7 | 0.76 | Mannitol | 1 | 1 | 0 | 0.1 |
| saturated FFA | 18 | 0.76 | Arachidic acid | 1 | 1 | 0 | 0.06 |
| diglycerides | 14 | 0.81 | DG (38:6) | 1 | 0 | 0 | 0.07 |
| amino acids, aromatic | 7 | 1 | Tyrosine | 0 | 0 | 0 | 0 |
| amino acids, cyclic | 6 | 1 | Histidine | 0 | 0 | 0 | 0 |
| amino alcohols | 3 | 1 | 3-Amino-1-propanol | 0 | 0 | 0 | 0 |
| benzene derivatives | 6 | 1 | Tri-2-ethylhexyl trimellitate | 0 | 0 | 0 | 0 |
| DiHETrE | 5 | 1 | 8,9-dihydroxyeicosa-5,11,14-trienoic acid | 0 | 0 | 0 | 0 |
| DiHODE | 3 | 1 | 15,16-dihydroxyoctadeca-9,12-dienoic acid | 0 | 0 | 0 | 0 |
| DiHOME | 4 | 1 | 9,10-dihydroxyoctadec-12-enoic acid | 0 | 0 | 0 | 0 |
| EpETrE | 7 | 1 | 15(16)-epoxy-9,12-octadecadienoic acid | 0 | 0 | 0 | 0 |
| ethanolamines | 3 | 1 | Heptadecasphing-4-enine | 0 | 0 | 0 | 0 |
| guanidines | 3 | 1 | Guanidine | 0 | 0 | 0 | 0 |
| HETE | 7 | 1 | 20-Hydroxyarachidonic acid | 0 | 0 | 0 | 0 |
| hexoses | 10 | 1 | Levoglucosan | 0 | 0 | 0 | 0 |
| histidine | 4 | 1 | Ergothioneine | 0 | 0 | 0 | 0 |
| HODE | 3 | 1 | 9-Hydroxylinoleic acid | 0 | 0 | 0 | 0 |
| hydroxybutyrates | 7 | 1 | 2-hydroxybutanoic acid | 0 | 0 | 0 | 0 |
| imidazoles | 3 | 1 | 1,2-Dimethylimidazole | 0 | 0 | 0 | 0 |
| indoles | 8 | 1 | Kynurenine | 0 | 0 | 0 | 0 |

**Supplementary Table S8.** ChemRICH enrichment analysis comparing ME/CFS without sr-IBS vs. controls without sr-IBS.

| Cluster name | Cluster size | p-values | Key compound | Altered | Increased | Decreased | Altered Ratio |
|---|---|---|---|---|---|---|---|
| lipids | 3 | 1 | Dinor-12-oxophytodienoic acid | 0 | 0 | 0 | 0 |
| monounsaturated long chain TGs | 16 | 1 | TG (58:2) | 0 | 0 | 0 | 0 |
| monounsaturated TGs | 18 | 1 | TG (49:2) | 0 | 0 | 0 | 0 |
| nitroFA | 3 | 1 | 10-nitrolinoleic acid | 0 | 0 | 0 | 0 |
| oxo-ODE | 5 | 1 | 13-ketooctadeca-9,11-dienoic acid | 0 | 0 | 0 | 0 |
| pyridines | 3 | 1 | 3-Hydroxypyridine | 0 | 0 | 0 | 0 |
| saturated ceramides | 4 | 1 | Ceramide (d42:0) | 0 | 0 | 0 | 0 |
| saturated phosphatidylcholines | 9 | 1 | PC (34:0) - ESI(-) | 0 | 0 | 0 | 0 |
| saturated sphingomyelins | 10 | 1 | SM (d32:0) - ESI(-) | 0 | 0 | 0 | 0 |
| sugar acids | 3 | 1 | Glyceric acid | 0 | 0 | 0 | 0 |
| unsaturated TGs | 27 | 1 | TG (48:4) A | 0 | 0 | 0 | 0 |
| xanthines | 3 | 1 | 3-Methylxanthine | 0 | 0 | 0 | 0 |

ME/CFS, myalgic encephalomyelitis/chronic fatigue syndrome; sr-IBS, self-reported physican diagnosed irritable bowel syndrome.

**Supplementary Table S9.** AUROC values and associated 95% CIs predicting ME/CFS and ME/CFS subgroups.

| Predictive Modeling | | All Metabolites | | Metabolites with BF>1 | | Metabolites with BF>3 | |
|---|---|---|---|---|---|---|---|
| | | AUROC | 95% CI | AUROC | 95% CI | AUROC | 95% CI |
| ME/CFS vs. Controls | Lasso | 0.636 | (0.450, 0.822) | 0.656 | (0.477, 0.834) | 0.608 | (0.424, 0.793) |
| | Adaptive Lasso | 0.644 | (0.462, 0.827) | 0.575 | (0.388, 0.762) | 0.575 | (0.388, 0.762) |
| | Random Forests | 0.589 | (0.402, 0.776) | 0.656 | (0.478, 0.833) | 0.631 | (0.447, 0.814) |
| | XGBoost | 0.589 | (0.402, 0.776) | 0.567 | (0.377, 0.756) | 0.592 | (0.406, 0.777) |
| | Model Average | 0.589 | (0.402, 0.776) | 0.656 | (0.478, 0.833) | 0.633 | (0.451, 0.816) |
| Female ME/CFS vs. Female Controls | Lasso | 0.709 | (0.480, 0.938) | 0.794 | (0.612, 0.976) | 0.745 | (0.538, 0.953) |
| | Adaptive Lasso | 0.703 | (0.474, 0.932) | 0.721 | (0.489, 0.953) | 0.727 | (0.496, 0.959) |
| | Random Forests | 0.691 | (0.471, 0.911) | 0.673 | (0.447, 0.898) | 0.764 | (0.553, 0.975) |
| | XGBoost | 0.655 | (0.425, 0.884) | 0.624 | (0.387, 0.862) | 0.776 | (0.571, 0.980) |
| | Model Average | 0.679 | (0.455, 0.902) | 0.770 | (0.570, 0.969) | 0.739 | (0.527, 0.952) |
| ME/CFS without sr-IBS vs. Controls without sr-IBS | Lasso | 0.500 | (0.500, 0.500) | 0.810 | (0.654, 0.966) | 0.873 | (0.747, 0.999) |
| | Adaptive Lasso | 0.552 | (0.332, 0.772) | 0.679 | (0.453, 0.905) | 0.588 | (0.365, 0.812) |
| | Random Forests | 0.638 | (0.432, 0.844) | 0.862 | (0.733, 0.991) | 0.810 | (0.641, 0.979) |
| | XGBoost | 0.588 | (0.373, 0.804) | 0.738 | (0.544, 0.931) | 0.756 | (0.572, 0.939) |
| | Model Average | 0.638 | (0.432, 0.844) | 0.864 | (0.736, 0.993) | 0.837 | (0.675, 0.999) |

ME/CFS, myalgic encephalomyelitis/chronic fatigue syndrome; sr-IBS, self-reported physician diagnosed irritable bowel syndrome; BF, BayesFactor; AUROC, area under the receiver operating characteristic; CI, confidence interval. The predictive models were first trained in the 80% randomly-selected training set and were validated in the remaining 20% test set.
